# Methodological and Clinical Validation of TholdStormDX v0.0.1: An Advanced Stochastic Engine for the Optimization of Thresholds and Multimarker Panels Applied to Oncology

**DOI:** 10.64898/2026.04.24.26351692

**Authors:** Roberto Reinosa Fernández

## Abstract

**Introduction:** The translation of biomarkers into binary clinical decisions requires the determination of precise cut-off points. This study validates the **TholdStormDX v0.0.1** tool, a mathematical engine that employs Dual Annealing, 2- and 4-parameter logistic fitting, and vectorized Monte Carlo simulations for panel optimization under Boolean OR logic.

**Methods:** The tool was evaluated using datasets from four diagnostic domains (Pulmonary Nodules, Hepatocellular Carcinoma [HCC], Cervical Cancer, and Breast Cancer), along with a prognosis-oriented analytical context (Breast Cancer). Validation followed a strict workflow: characterization and selection of the best individual and combined thresholds in the Training (Train) and Validation (Val) sets, using the Test set in a completely independent manner, solely to assess the model’s performance and generalizability.

**Results:** The tool enabled precise derivation of cut-off points for both individual biomarkers and multivariable combinations. Evaluation on the Test set objectively demonstrated in which scenarios a single biomarker outperforms a complex panel, promoting clinical parsimony. For example, in Breast Cancer diagnosis, an individual predictor outperformed the optimized panel (Sensitivity: 0.953 / Specificity: 0.952 in Test); conversely, in Hepatocellular Carcinoma, the multivariable combination showed superior performance compared to the single marker (Sens: 0.707 / Spe: 0.718 in Test). Additionally, the self-auditing system effectively flagged metric degradation when noisy variables were included, preventing potential issues.

**Conclusion:** TholdStormDX v0.0.1 proves to be a robust and transparent bioinformatics platform for deriving clinical thresholds. Its main contribution lies in mitigating local minima and promoting clinical parsimony, enabling researchers to objectively identify when a single biomarker is sufficient and when a panel provides real added value. Furthermore, it transforms the problem of biological noise into a safety feature: by systematically warning about algorithmic instability, it prevents overfitting and ensures the clinical viability of medical decisions.

**Availability:** The software is free and distributed under the GNU GPLv3 license. TholdStormDX v0.0.1 is written in Python, and its source code is available at the following GitHub address: https://github.com/roberto117343/TholdStormDX.

## 1. Introduction

In the field of precision oncology, risk stratification through biomarkers and quantitative clinical features is fundamental. Whether for early diagnosis (e.g., Pulmonary Nodules [1], Hepatocellular Carcinoma [2], Breast Cancer [3], Cervical Cancer [4]) or prognostic assessment (e.g., long-term recurrence prediction in breast cancer) [5], the underlying mathematical challenge is the same: determining the operational cut-off point that maximizes clinical utility.

The literature shows that combinations of biomarkers can outperform individual predictors [1, 2]. However, conventional approaches for defining thresholds in multimarker panels often derive from static analyses on entire cohorts, which invariably leads to overfitting and catastrophic failure in real-world external validation.

To address these limitations, **TholdStormDX v0.0.1** has been developed. This software tool proposes a two-stage approach based on previous methodological frameworks [6, 7]: (1) individual mathematical modeling using global optimization algorithms (Dual Simulated Annealing) [8] to fit 2- and 4-parameter logistic functions, and (2) a vector-driven stochastic combinatorial optimization that evaluates millions of iterations under Boolean “OR” logic.

The aim of this study is to validate the mathematical architecture and clinical safety of TholdStormDX v0.0.1. To this end, while placing less emphasis on the biological selection of variables, the study focuses on subjecting the tool to a rigorous cross-domain stress test using datasets from four diagnostic scenarios and establishing the groundwork for one prognostic scenario. Tool performance is evaluated by strictly isolating the Test set, using it only at the end of the process to assess the real-world performance of models selected during Training and Validation.

## 2. Materials and Methods

### 2.1. Datasets and Study Design

Data derived from real-world clinical practice and well-established public databases were used, covering five domains of analysis (4 diagnostic-oriented and 1 prognostic):

1. **Breast Cancer (Diagnosis/Prognosis):** Features extracted from studies include texture, area, compactness, symmetry, fractal dimension, concavity, among others [3, 5].
2. **Hepatocellular Carcinoma (HCC):** Tumor markers (AFP, PIVKA-II, OPN, DKK-1), demographic variables, and liver function metrics (AGE, MELD, CTP) [2].
3. **Pulmonary Nodules:** Imaging variables (Nodule Diameter, CTR, CT value), serum biomarkers (CEA, CYFRA21-1, SCC, ProGRP, NSE), and age [1].
4. **Cervical Cancer:** Clinical, demographic, and lifestyle risk factors [4].

### 2.2. Usage Mode of TholdStormDX v0.0.1

The TholdStormDX v0.0.1 tool is designed to be very easy to use. Its graphical interface is highly intuitive, allowing even users without advanced technical knowledge to operate it.

**Figure 1.**
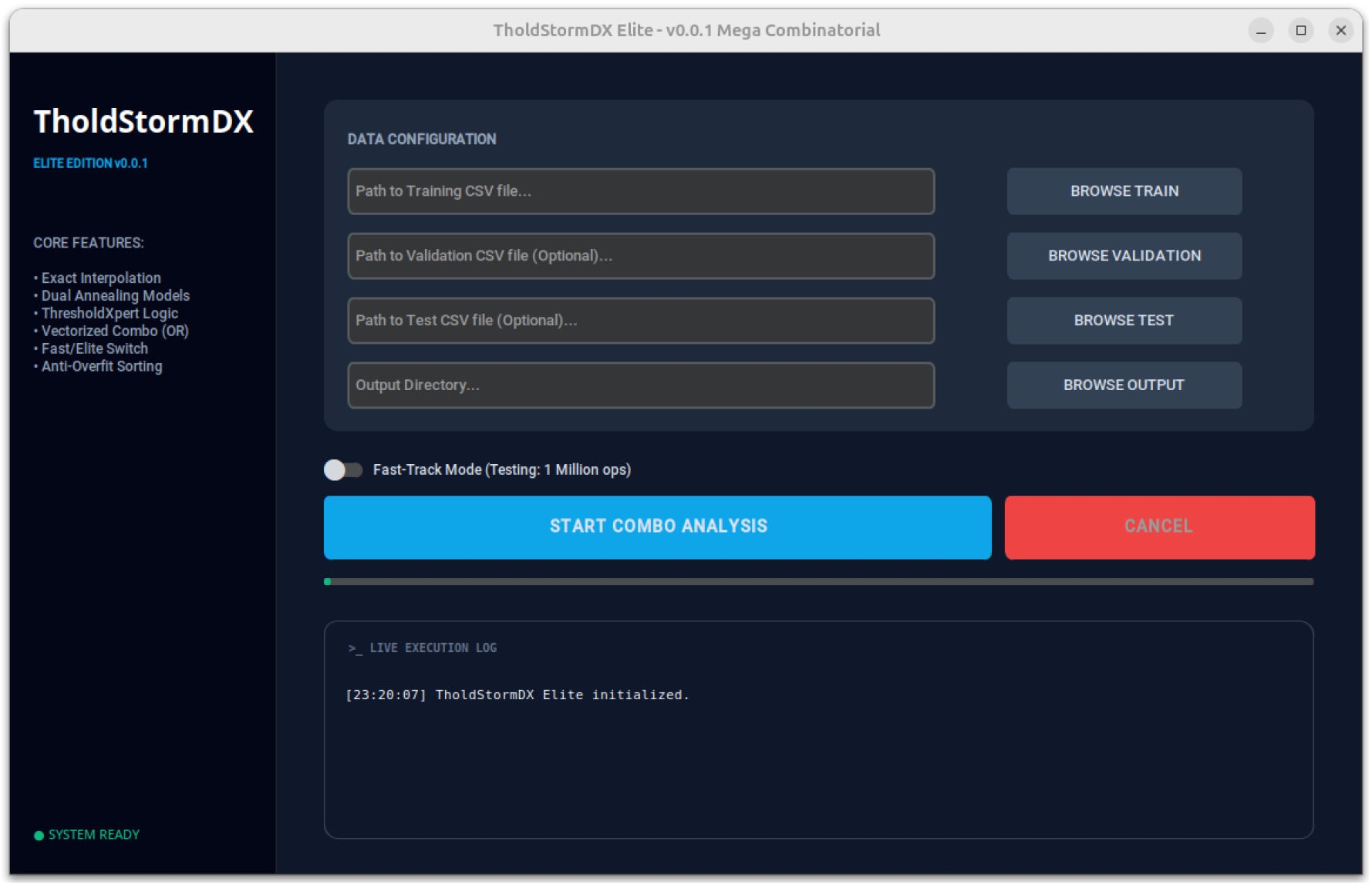
Graphical Interface of TholdStormDX v0.0.1

Ideally, the dataset should be divided into three parts: training, validation, and test sets. Although TholdStormDX v0.0.1 allows cut-off calculation using only training/validation, training/test, or training-only combinations, it is methodologically preferable to perform the split into all three sets mentioned.

In the presented examples, the training set accounted for approximately 60% of the data, while the validation and test sets each represented 20%; however, this proportion may vary depending on the initial dataset. The sets were kept approximately balanced (50% positive and 50% negative). In cases where an initial balance did not exist, the data were balanced, and the excess was incorporated into the test set.

After splitting the datasets, the next step is to load them into their corresponding sections of the interface (“Path to Training CSV File…”, “Path to Validation CSV File (Optional)…”, and “Path to Test CSV File (Optional)…”). It is essential that the input files are in CSV format, using a semicolon (;) as the delimiter and a dot (.) as the decimal separator. The target feature to be predicted must be located in the last column.

The generated files will be saved in a folder that must be specified in the “Output Directory…” section. If not manually defined, the default location will be the folder where the training dataset is stored.

By enabling the Fast-Track Mode option, results are produced more quickly at the cost of lower overall quality. Although this mode provides acceptable accuracy for individual cut-off calculations, it is recommended to keep it disabled to obtain significantly more precise results in the overall analysis.

Finally, click the “Start Combo Analysis” button and simply wait for the program to complete the calculations.

The results are provided in two files. The first is a PDF summarizing the most relevant information, including individual variable calculations and multivariable panels. The second is a CSV detailing all possible panels (unlike the PDF, which displays 200), along with their corresponding cut-off points and sensitivity and specificity metrics for the three datasets. This CSV allows filtering based on training and validation metrics to select the most balanced panel, and subsequently compare its performance against the test set metrics. In the PDF, the metrics corresponding to univariate analyses in the training set for logistic models are obtained through analytical calculation.

All generated files are available as supplementary material.

**Figure 2.**
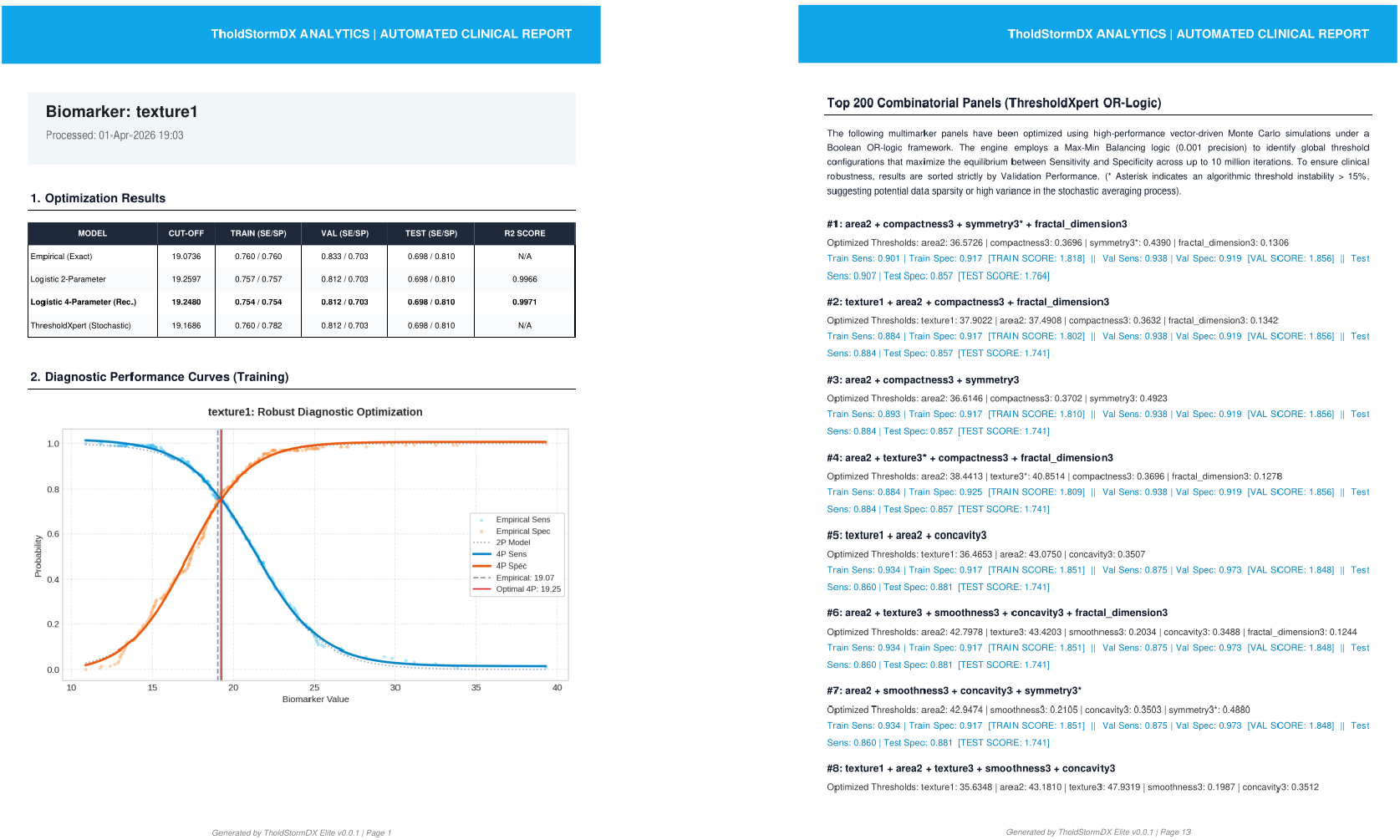
Example of the results displayed in the PDF file.

### 2.3. Architecture of the TholdStormDX v0.0.1 Engine

#### Stage 1: Individual Characterization (Logistic Modeling and Dual Annealing)

For each continuous variable, the software calculates the exact empirical intersection between the Sensitivity (SE) and Specificity (SP) curves. Subsequently, it independently fits the data to two models [7]:

- **2-Parameter Logistic Model (2P):** Assumes an ideal transition from 0 to 1.
- **4-Parameter Logistic Model (4P):** Introduces variable lower and upper asymptotes, allowing superior fitting in biomarkers with biological asymmetry or baseline noise.

Both models are optimized using the Dual Simulated Annealing algorithm [8], ensuring convergence toward the global optimum and avoiding entrapment in local minima.

Finally, as a non-parametric contrast to the logistic fittings, the tool subjects the individual variable to a fourth method: **Vectorized Stochastic Search (ThresholdXpert)** [6]. By replicating the core engine of Stage 2, this method evaluates millions of Monte Carlo iterations using Max-Min balancing logic to empirically identify the cut-off point that maximizes exact parity between Sensitivity and Specificity in the raw data.

#### Stage 2: Multimarker Combinatorial Optimization (OR Logic and Balancing)

The engine evaluates variable panels using **OR logic**. Given the risk of this logic collapsing specificity, TholdStormDX v0.0.1 employs **Max-Min balancing logic** (with 0.001 precision) through vectorized Monte Carlo simulations. The algorithm iteratively searches for the threshold vector that maximizes balance between SE and SP.

### 2.4. Strict Selection and Validation Criterion (Blind Opening)

To ensure clinical robustness and rigorously test the tool, a strict methodological criterion was imposed: the selection of the “Best Individual Variable” and the “Best Panel” was based on **maximizing both parity (balance) and joint magnitude** between Sensitivity and Specificity, evaluated exclusively on the Training (Train) and Validation (Val) cohorts. When two models showed similar scores, priority was given to the one with the smaller gap between SE and SP.

The Test set was kept completely blinded throughout this process and was only opened at the end to document the generalization performance of the model.

## 3. Results: Tool Validation

### 3.1. Effectiveness of Individual Modeling (Stage 1)

In the individual characterization of variables, the results confirmed that there is no universal mathematical model that perfectly fits the heterogeneity of all oncological biomarkers. The tool demonstrated that offering different calculation approaches is not redundancy, but a critical necessity to properly address complex biological variables.

### 3.2. Performance and Generalization in the Test Set

The following results are presented after applying the maximum parity criterion in Training and Validation, subsequently revealing performance in the Test set.

#### A. Cohort 1: Hepatocellular Carcinoma (HCC)

- **Best Individual Predictor Selected: *AFP*** (Cut-off: 10.0782 in the 4-parameter logistic model).
  - *Training Performance: Sens: 0*.*804 / Spe: 0*.*804*
  - *Validation Performance:* Sens: 0.634 / Spe: 0.821
  - *Test Performance:* **Sens: 0.610 / Spe: 0.692**
- **Best Optimized Panel (Selected: Panel #28):** AGE: 77.7087 | INV. MELD: 1.8855 | AFP: 11.9935 | OPN: 182.4645 | DKK-1: 22891.5806
  - *Training Performance: Sens: 0*.*762 / Spe: 0*.*774 (Score: 1*.*536)*
  - *Validation Performance:* Sens: 0.707 / Spe: 0.795 (Score: 1.502)
  - *Test Performance:* Sens: **0.707 / Spe: 0.718 (Score: 1.425)**

#### B. Cohort 2: Breast Cancer (Diagnosis)

- **Best Individual Predictor Selected: *perimeter3*** (Cut-off: 103.5714 in the 4-parameter logistic model).
  - *Training Performance: Sens: 0*.*917 / Spe: 0*.*917*
  - *Validation Performance:* Sens: 0.917 / Spe: 0.865
  - *Test Performance:* **Sens: 0.953 / Spe: 0.952**
- **Best Optimized Panel (Selected: Panel #48):** area2: 34.9496 | texture3: 46.6170 | smoothness3: 0.1762 | compactness3: 0.3759 | symmetry3: 0.4221 | fractal_dimension3: 0.1220
  - *Training Performance: Sens: 0*.*926 / Spe: 0*.*917 (Score: 1*.*843)*
  - *Validation Performance:* Sens: 0.917 / Spe: 0.919 (Score: 1.836)
  - *Test Performance:* **Sens: 0.930 / Spe: 0.857 (Score: 1.787)**

#### C.Cohort 3: Pulmonary Nodules (Lung Cancer)

- **Best Individual Predictor Selected:** *CT. Value* (Cut-off: −459.2655 in the 4-parameter logistic model).
  - *Training Performance: Sens: 0*.*824 / Spe: 0*.*824*
  - *Validation Performance:* Sens: 0.930 / Spe: 0.774
  - *Test Performance:* **Sens: 0.826 / Spe: 0.774**
- **Best Optimized Panel (Selected: Panel #387):** Nodule Diameter: 20.5678 | CT.value: - 404.1191
  - *Training Performance: Sens: 0.813 / Spe: 0.811 (Score: 1.624)*
  - *Validation Performance:* Sens: 0.895 / Spe: 0.806 (Score: 1.702)
  - *Test Performance:* **Sens: 0.744 / Spe: 0.817 (Score: 1.561)**

#### D. Cohort 4: Cervical Cancer (Clinical Risk Factors)

- **Best Individual Predictor Selected:** *Num of pregnancies* (Cut-off: 2.6705 in the 4-parameter logistic model).
  - *Training Performance: Sens: 0*.*510 / Spe: 0*.*510*
  - *Validation Performance:* Sens: 0.778 / Spe: 0.778
  - *Test Performance:* **Sens: 0.500 / Spe: 0.652**
- **Best Optimized Panel (Selected: Panel #6):** Age: 48.9232 | Number of sexual partners: 4.5042 | First sexual intercourse: 25.8064 | Smokes (packs/year): 1.1201 | Hormonal Contraceptives (years): 9.5310 | STDs (number): 0.3646
  - *Training Performance: Sens: 0*.*654 / Spe: 0*.*714 (Score: 1*.*368)*
  - *Validation Performance:* Sens: 0.778 / Spe: 0.778 (Score: 1.556)
  - *Test Performance:* **Sens: 0.300 / Spe: 0.727 (Score: 1.027)**

#### E.Cohort 5: Breast Cancer (Prognosis)

- **Best Individual Predictor Selected:** *lymph_node_status* (Cut-off: 2.6396 in the 4-parameter logistic model).
  - *Training Performance: Sens: 0*.*591 / Spe: 0*.*591*
  - *Validation Performance:* Sens: 0.778 / Spe: 0.700
  - *Test Performance:* **Sens: 0.778 / Spe: 0.623**
- **Best Optimized Panel (Selected: Panel #77):** texture1: 17.2479 | concave_points3: 0.6895 | lymph_node_status: 7.6368
  - *Training Performance: Sens: 0*.*724 / Spe: 0*.*815 (Score: 1*.*539)*
  - *Validation Performance:* Sens: 0.667 / Spe: 0.700 (Score: 1.367)
  - *Test Performance:* **Sens: 0.889 / Spe: 0.395 (Score: 1.284)**

## 4. Discussion

The rigorous analysis of the automated reports confirms that **TholdStormDX v0.0.1** is a highly reliable bioinformatics platform, ready for translational research in oncology. The methodology of not inspecting the Test set during the optimization phase has demonstrated that the generated thresholds are potentially real and generalizable to new populations, depending on the specific case.

### 4.1. Strengths of the Tool

1. **Immunity to Local Minima:** Unlike classical gradient descent, the integration of Dual Simulated Annealing [8] in Stage 1 ensures that the baseline cut-off point (Sens = Spe) corresponds to the true global mathematical optimum of the biomarker.
2. **Biological Flexibility:** The inclusion in Stage 1 of multiple approaches for calculating individual cut-off points proved to be critical. In clinical practice, many tumor markers exhibit baseline “noise” levels in healthy patients or fail to reach 100% sensitivity due to tumor heterogeneity. TholdStormDX v0.0.1 adapts to this noise, deriving more realistic thresholds.
3. **Vectorized Computational Power:** The engine’s ability to compute Monte Carlo simulations under OR logic at massive speeds allows it to find a needle in a haystack: the precise point where adding a variable increases sensitivity without collapsing specificity, regulated by the Max-Min algorithm.

### 4.2. Identified Limitations (Safety Mechanisms)

One of the most important features of TholdStormDX v0.0.1 is its self-auditing capability. Throughout the reports, certain variables within panels are marked with an **asterisk (*)** (e.g., CYFRA21-1* in lung cancer, or symmetry3* in breast cancer).

1. **Algorithmic Instability:** The asterisk alerts the clinician that the threshold of that variable fluctuates by more than 15% during the stochastic averaging process. This occurs when data are sparse (matrix scarcity) or when the mathematical contribution of the variable is marginal or redundant, creating a flat optimization landscape.
2. **Degradation Due to OR Logic:** Although OR logic is a classical strategy for highly sensitive screening tests, including purely noisy variables forces the algorithm to push the thresholds of other variables to suboptimal levels to compensate for the loss of specificity. This is typically observed in oversized panels.
3. **Evaluation of Non-specific or Diffuse Variables:** Variables of behavioral, demographic, or indirectly related nature tend to show limited performance in the Test set. While they may exhibit apparent associations in early phases (training or internal validation),these associations do not consistently hold when evaluated on independent data. This behavior is expected.
4. **Scalability and Combinatorial Complexity:** In the context of multimarker panels, the number of possible variable combinations grows rapidly with the size of the initial set. As a practical consequence, exhaustive search is not recommended with more than approximately 10–15 variables, as computation time can increase substantially.

## 5. Conclusion

**TholdStormDX v0.0.1** stands as a robust, transparent, and clinically safe algorithmic framework for the design of binary diagnostic and prognostic systems. By replacing subjective graphical estimates and “black-box” regression approximations with direct analytical calculations (Dual Annealing) and strict combinatorial optimization, the tool enables the discovery of high-performance parsimonious panels.

Independent cross-validation across four distinct oncological pathologies demonstrates that, as long as the algorithmic instability warning (asterisk) is respected and parsimony is prioritized, the thresholds calculated by TholdStormDX v0.0.1 can maintain their predictive integrity when applied to entirely new patient cohorts. This constitutes a fundamental step forward toward the implementation of critical medical decisions.

## Supporting information

Supplementary material. Outputs TholdStormDX for this study.

## Data Availability

The software is freely available and distributed under the GNU GPLv3 license. TholdStormDX v0.0.1 is written in Python, and its source code is available at the following GitHub repository:https://github.com/roberto117343/TholdStormDX. The datasets used for validation (Breast Cancer, Cervical Cancer, HCC, and Pulmonary Nodules) are from public repositories as described in the manuscript.

## 6. Availability

TholdStormDX v0.0.1 is open-source software programmed in Python and distributed under the terms of the GNU General Public License v3. It is available on GitHub at the following address: https://github.com/roberto117343/TholdStormDX.

## Appendix 1. Source code of TholdStormDX v0.0.1.

(https://github.com/roberto117343/TholdStormDX/blob/main/main.py)

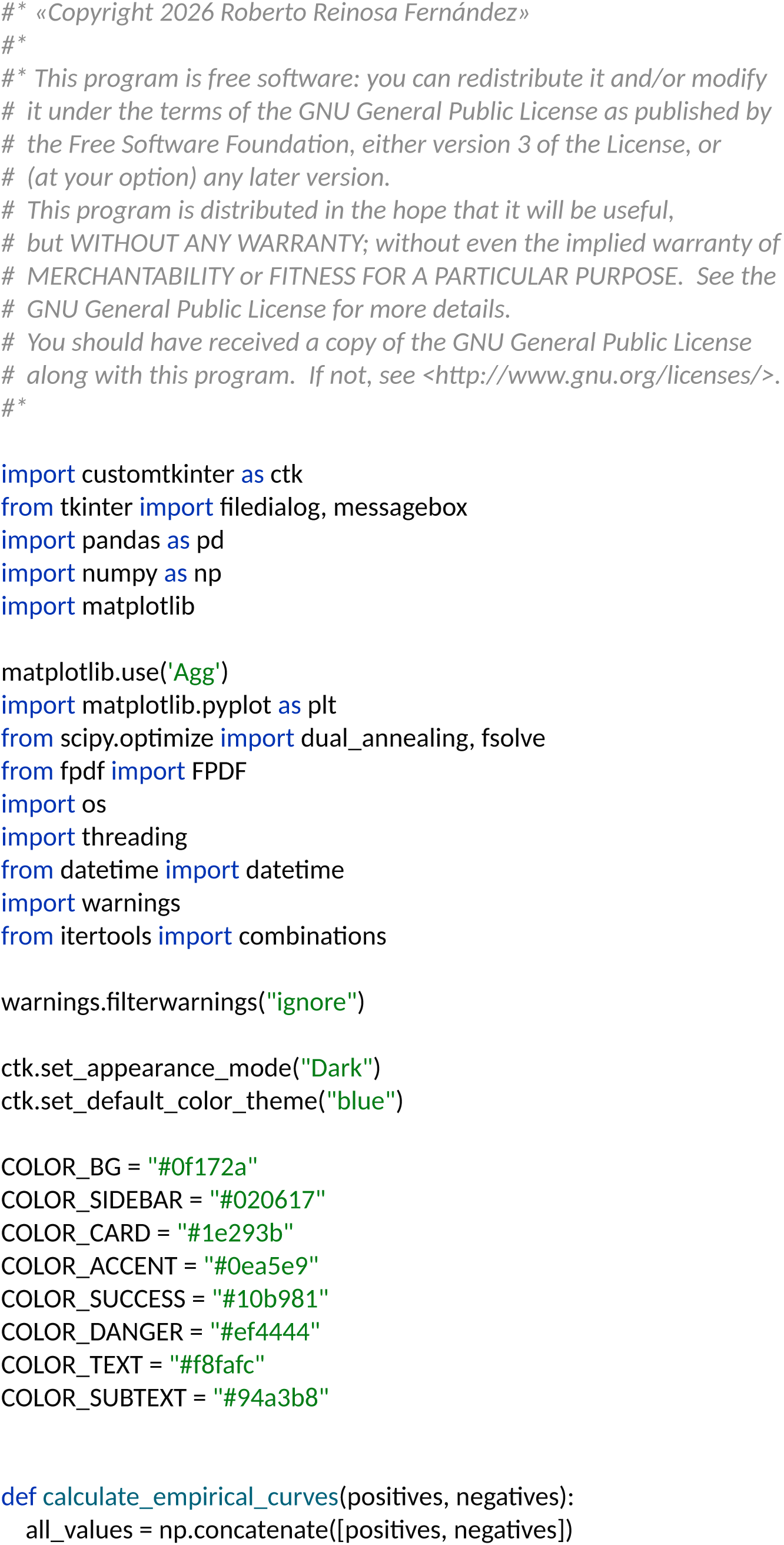
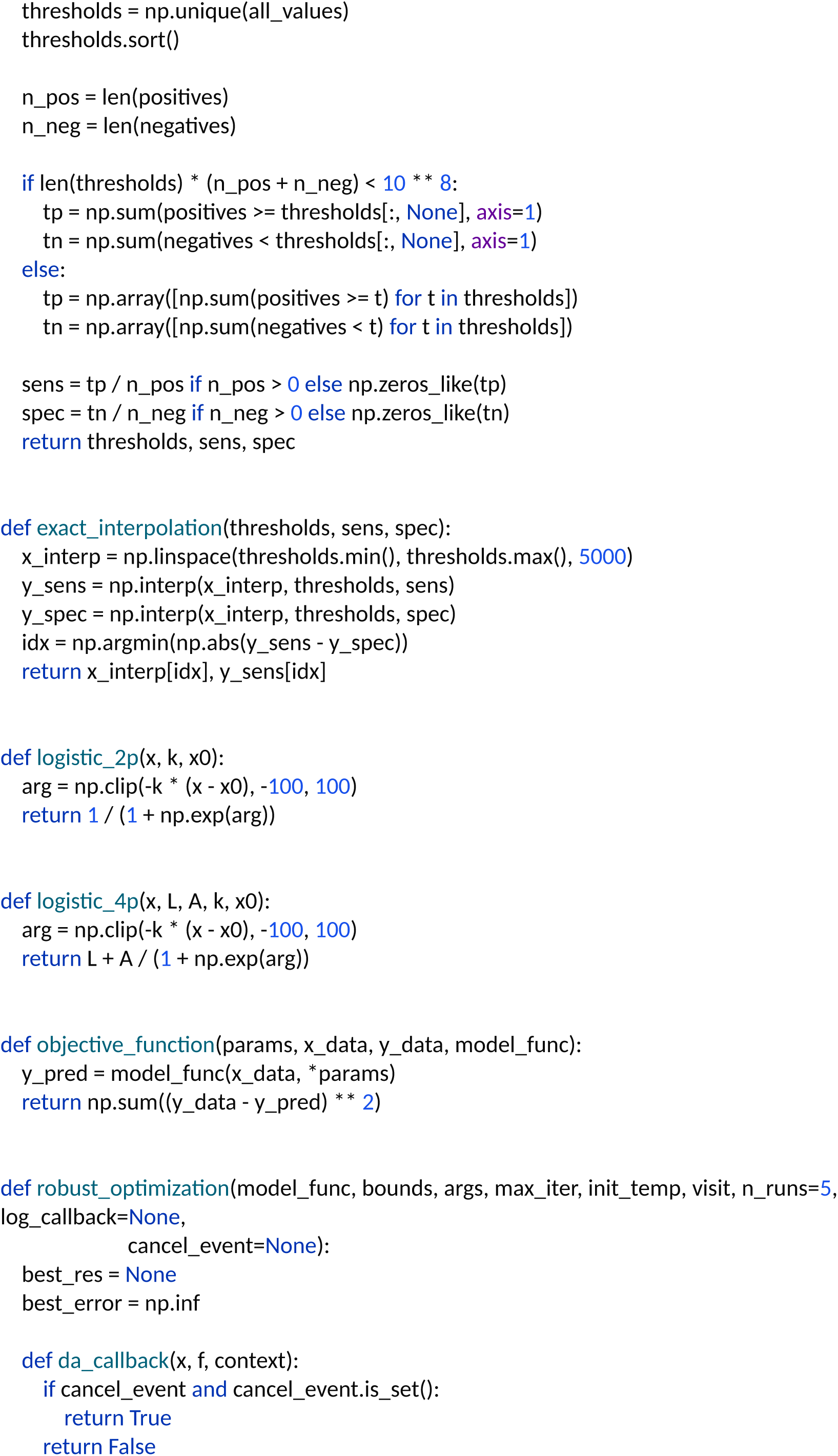
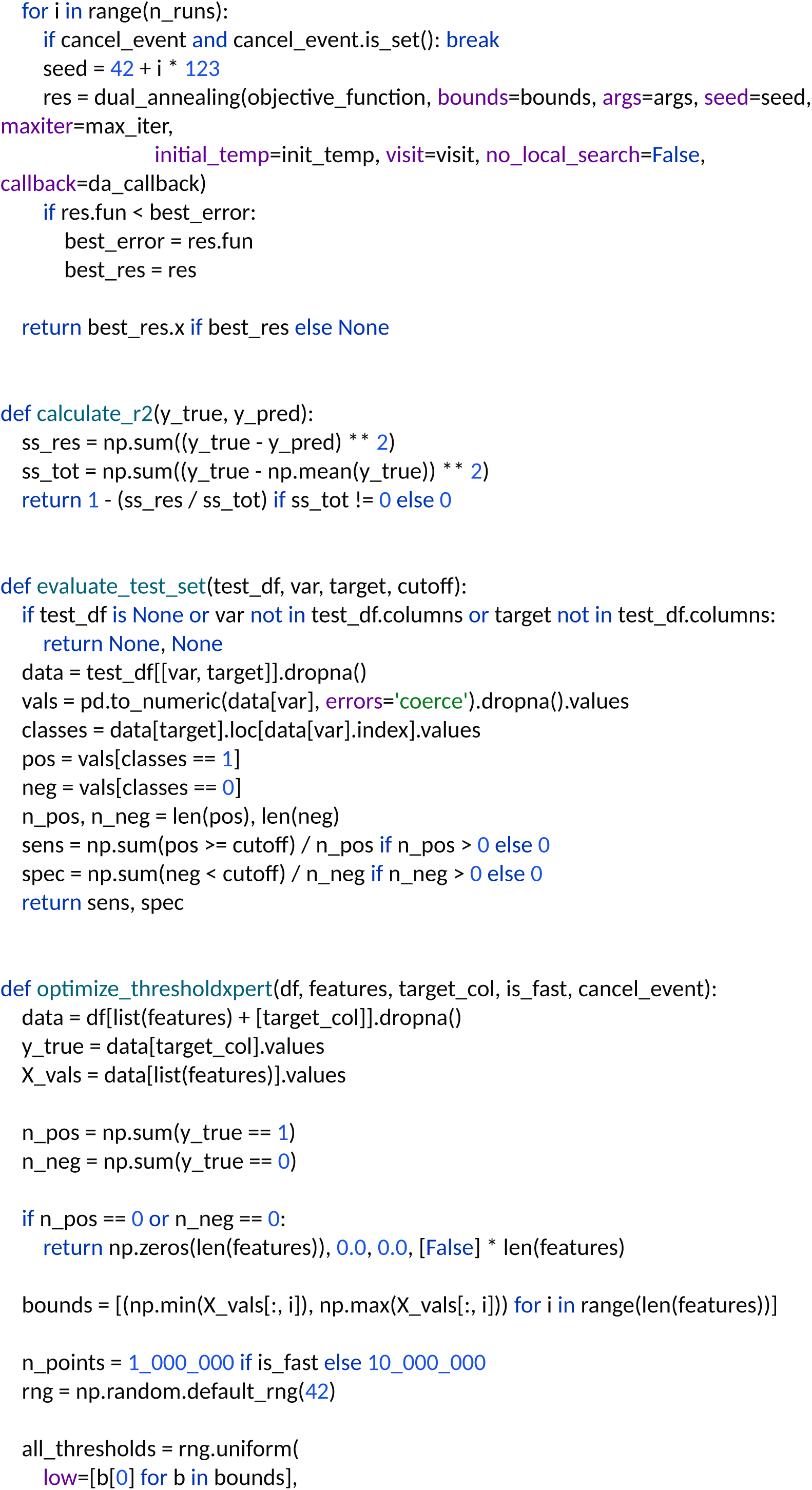
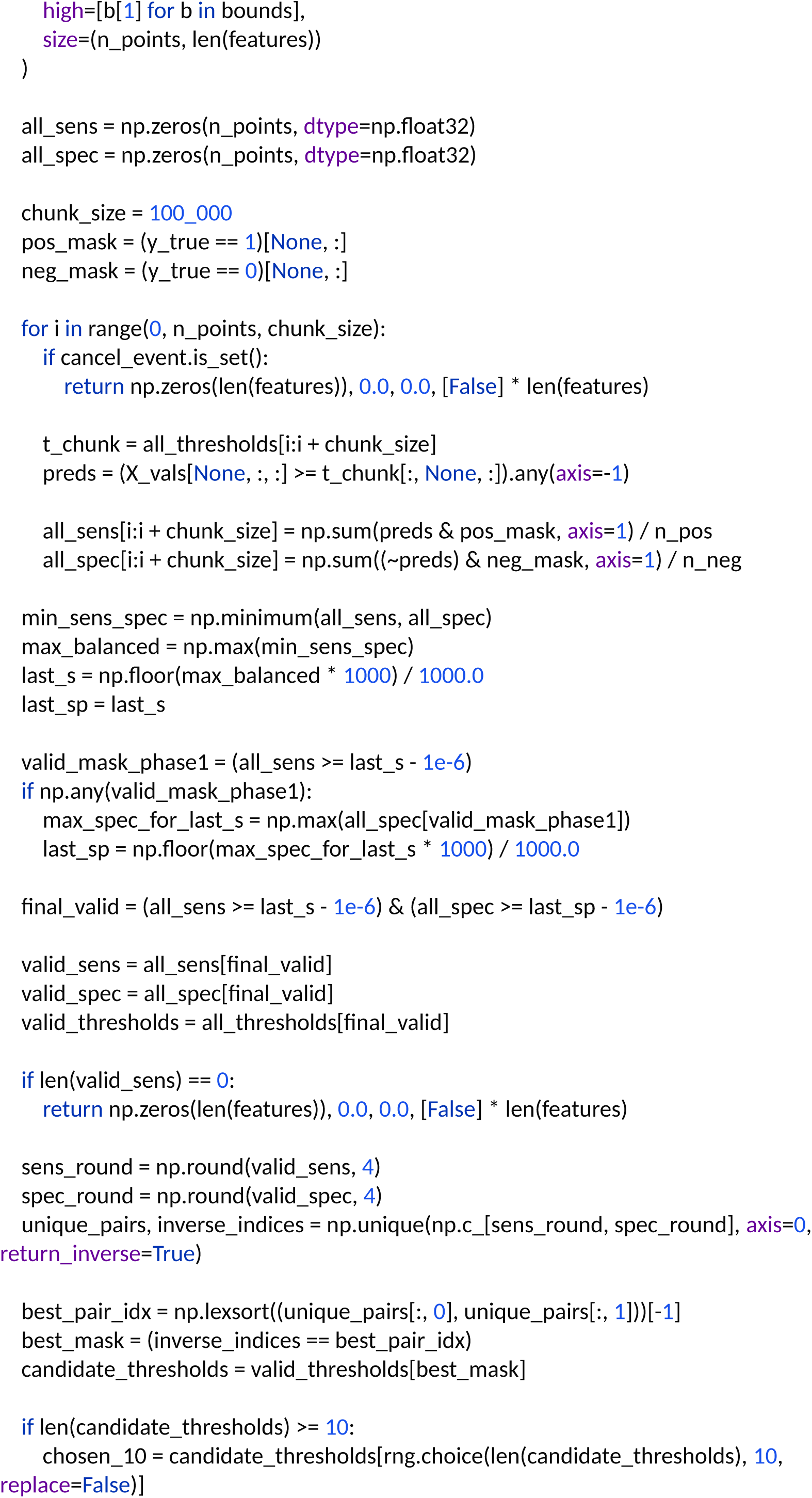
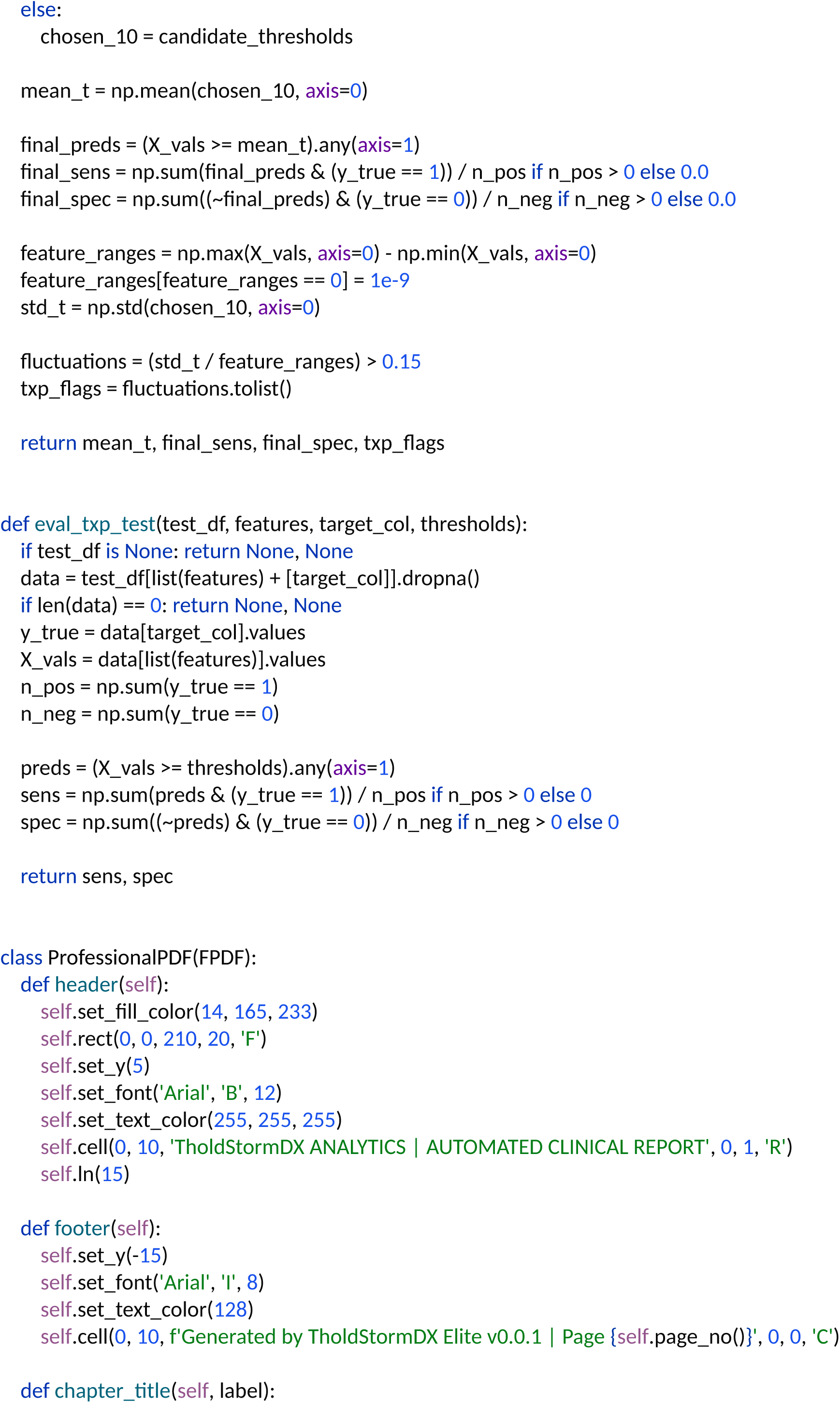
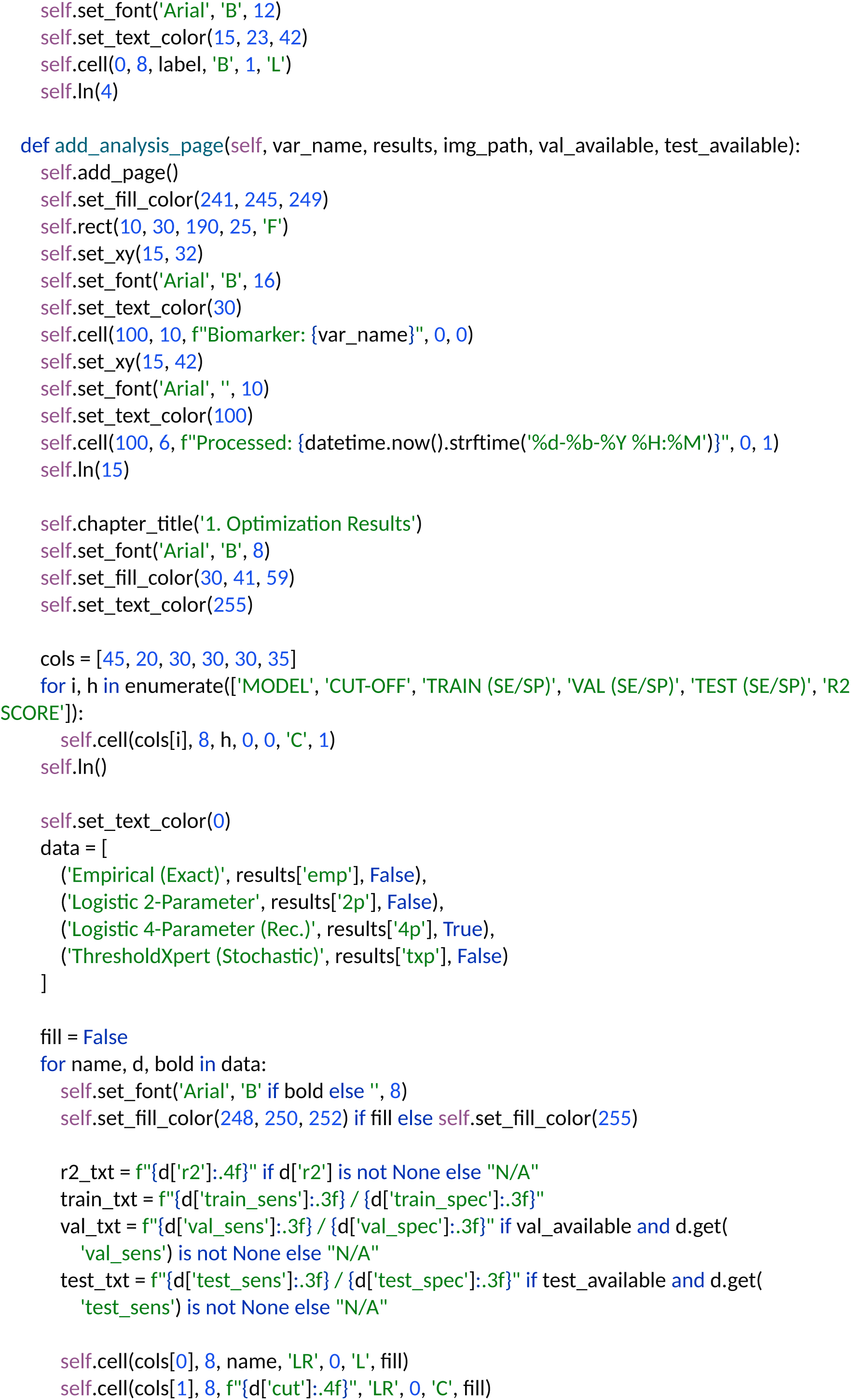
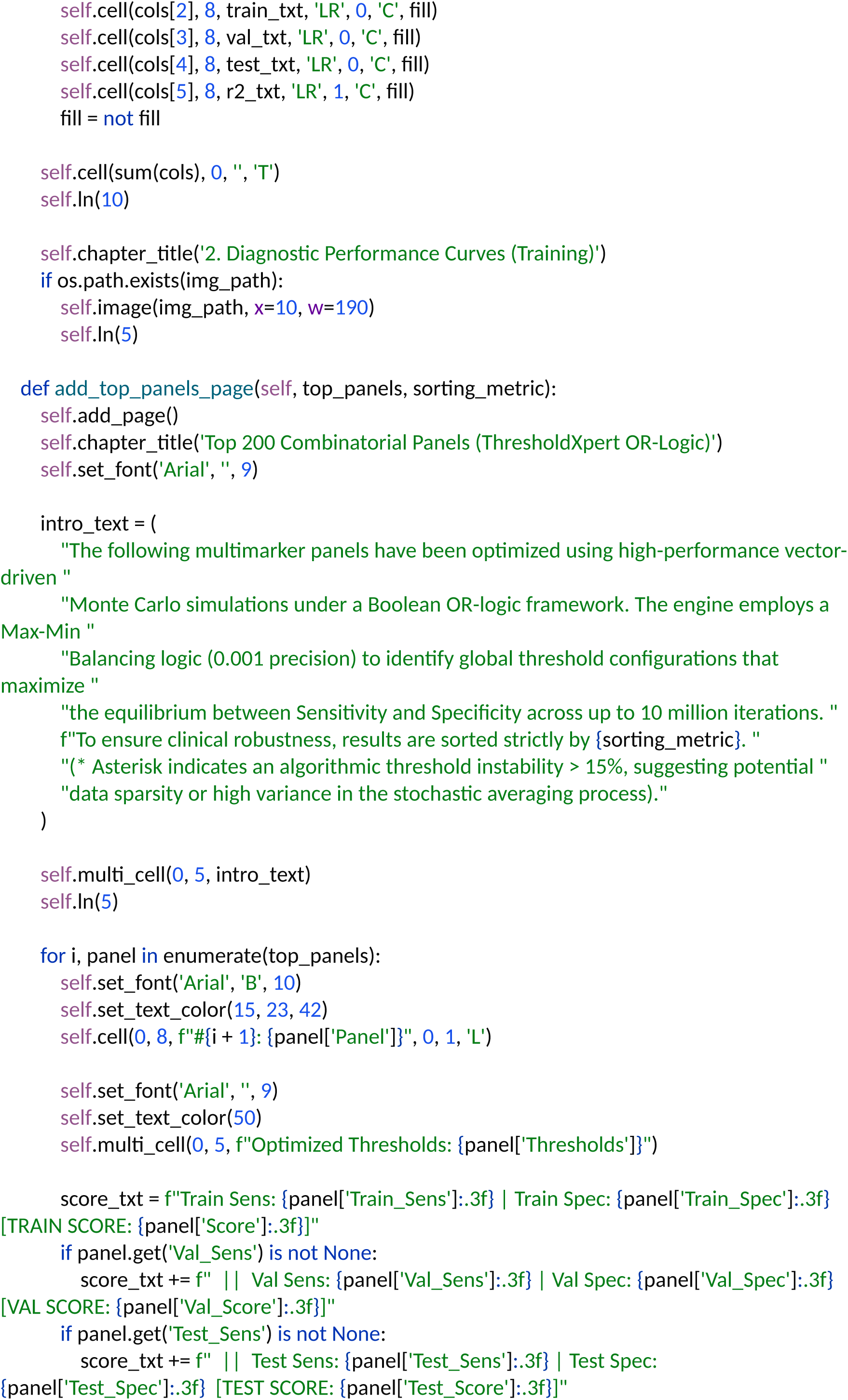
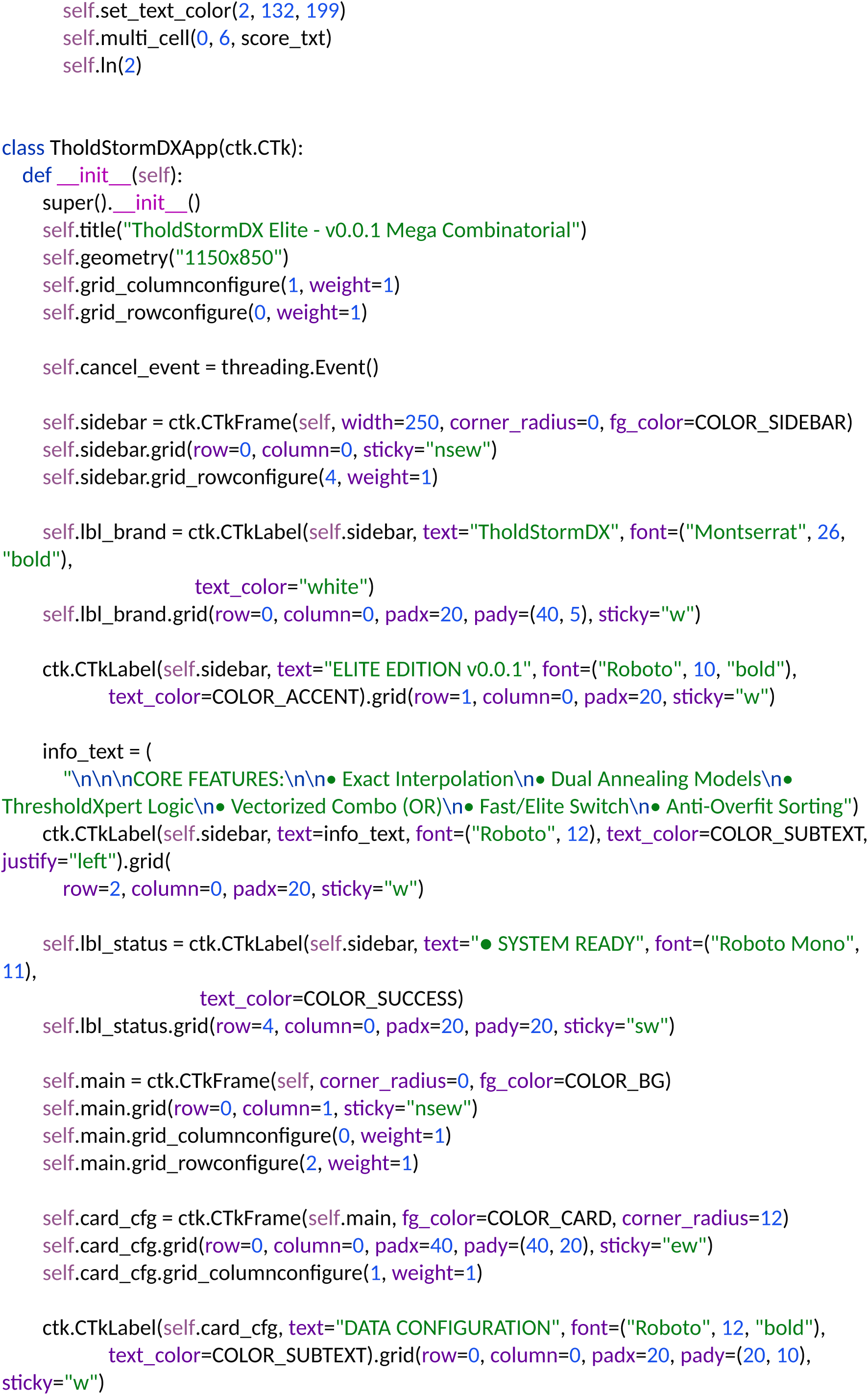
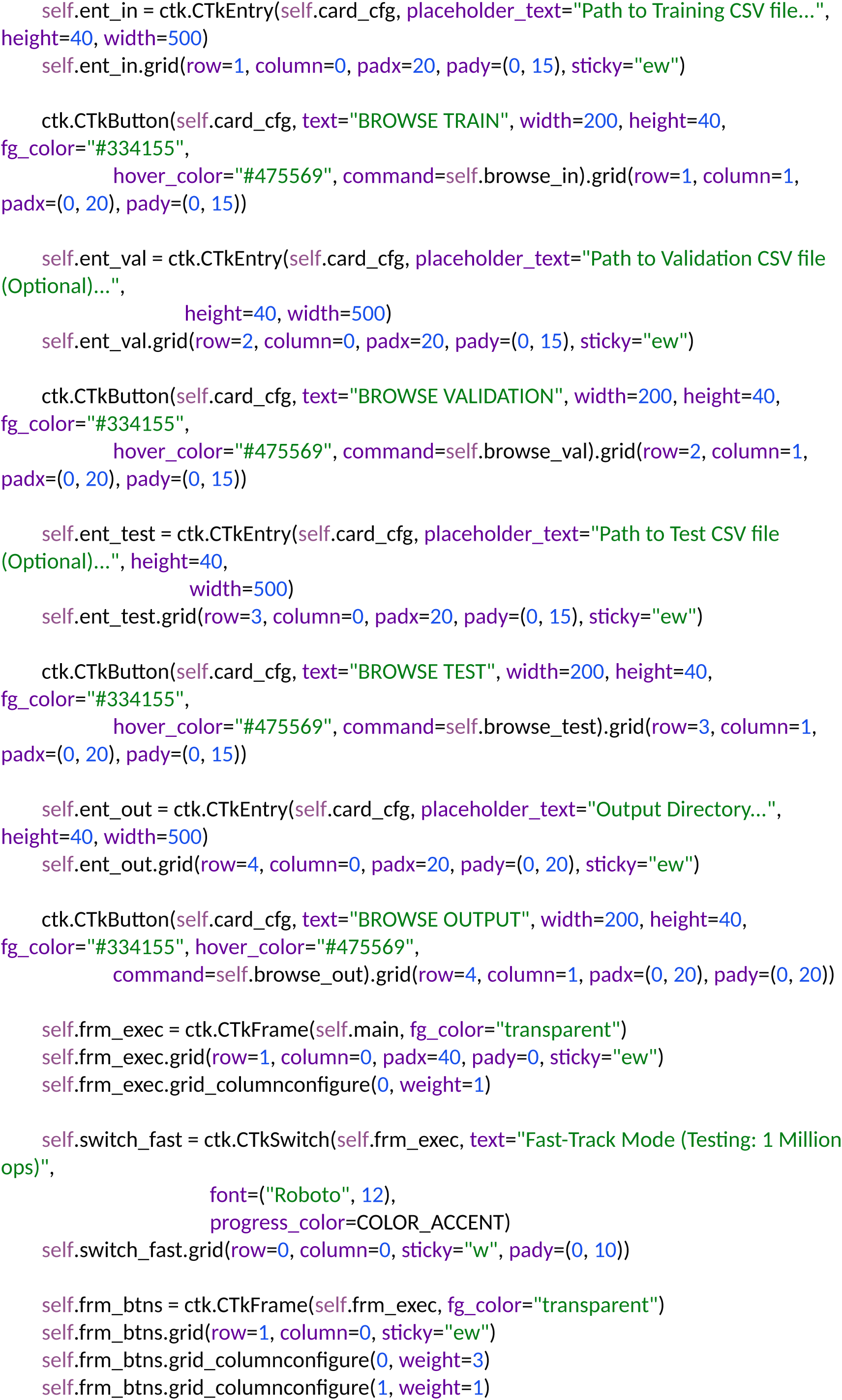
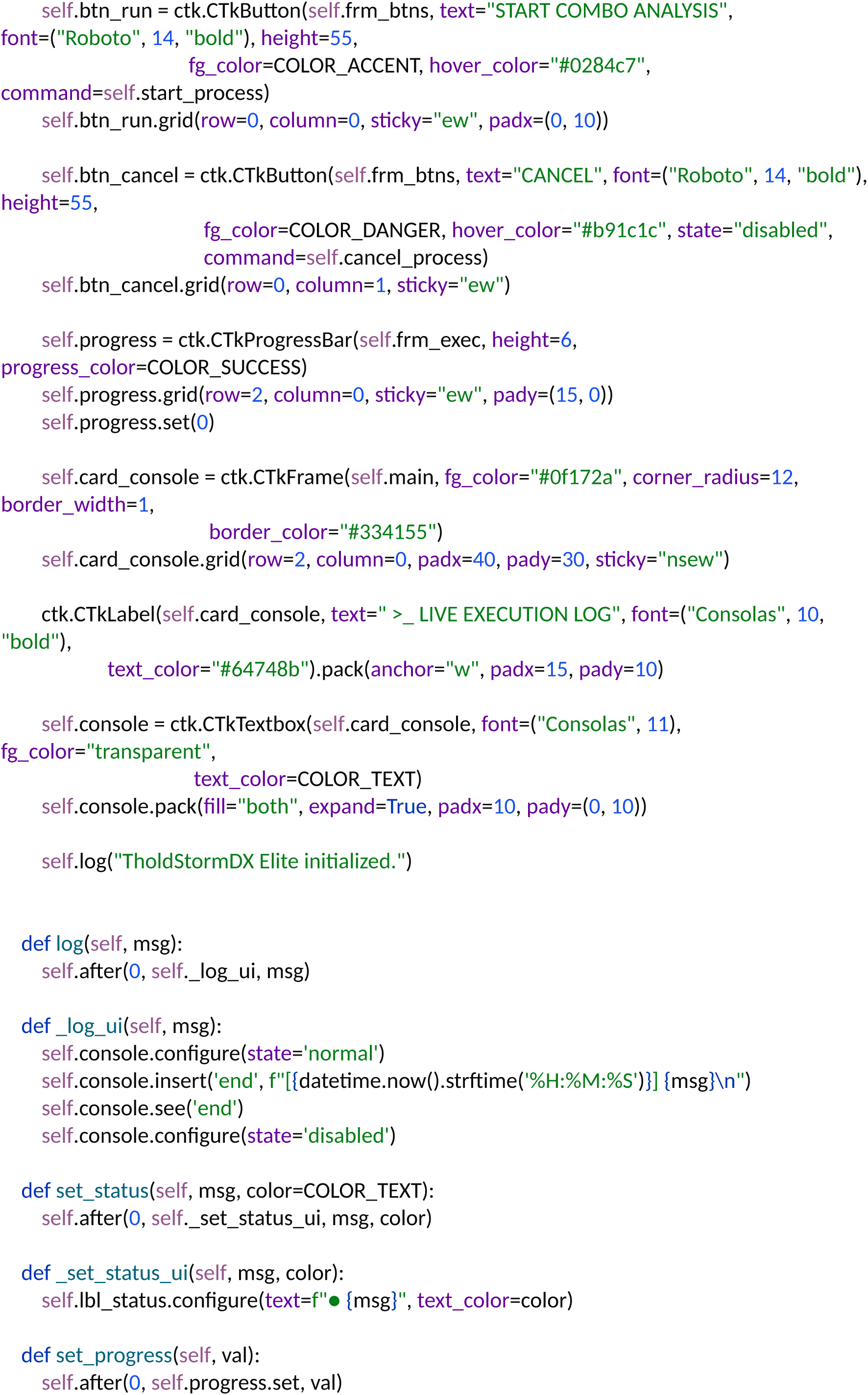
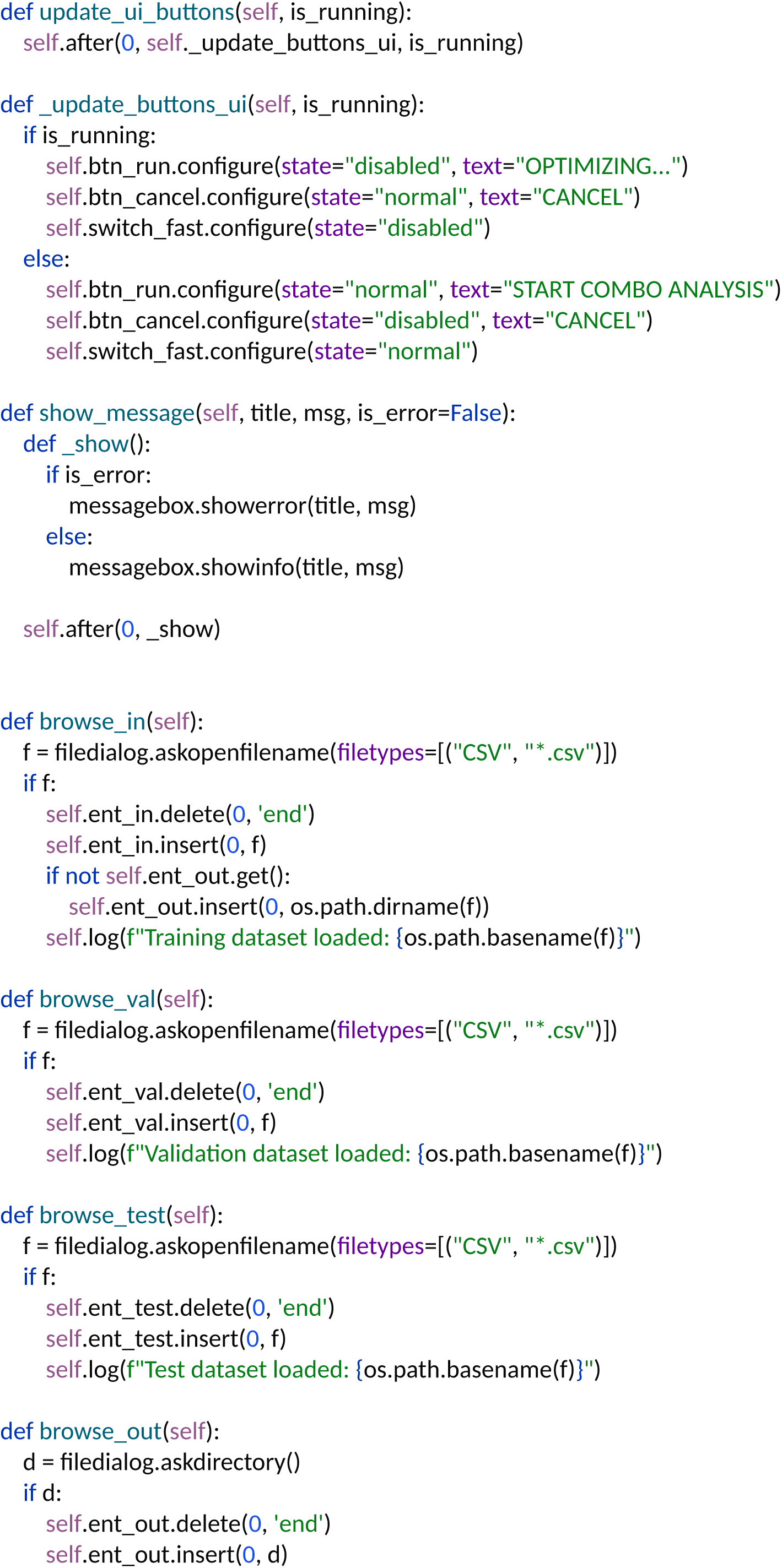
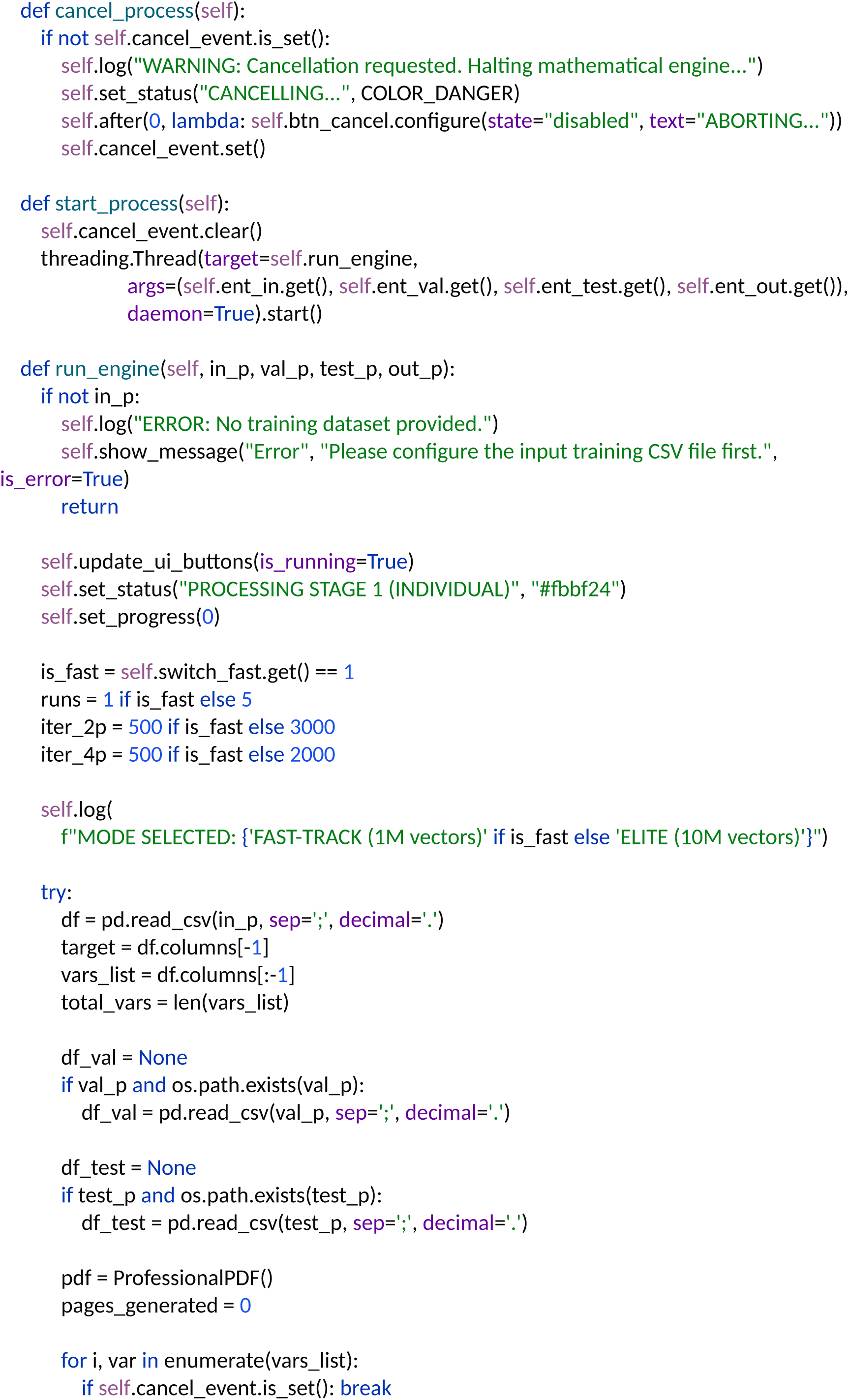
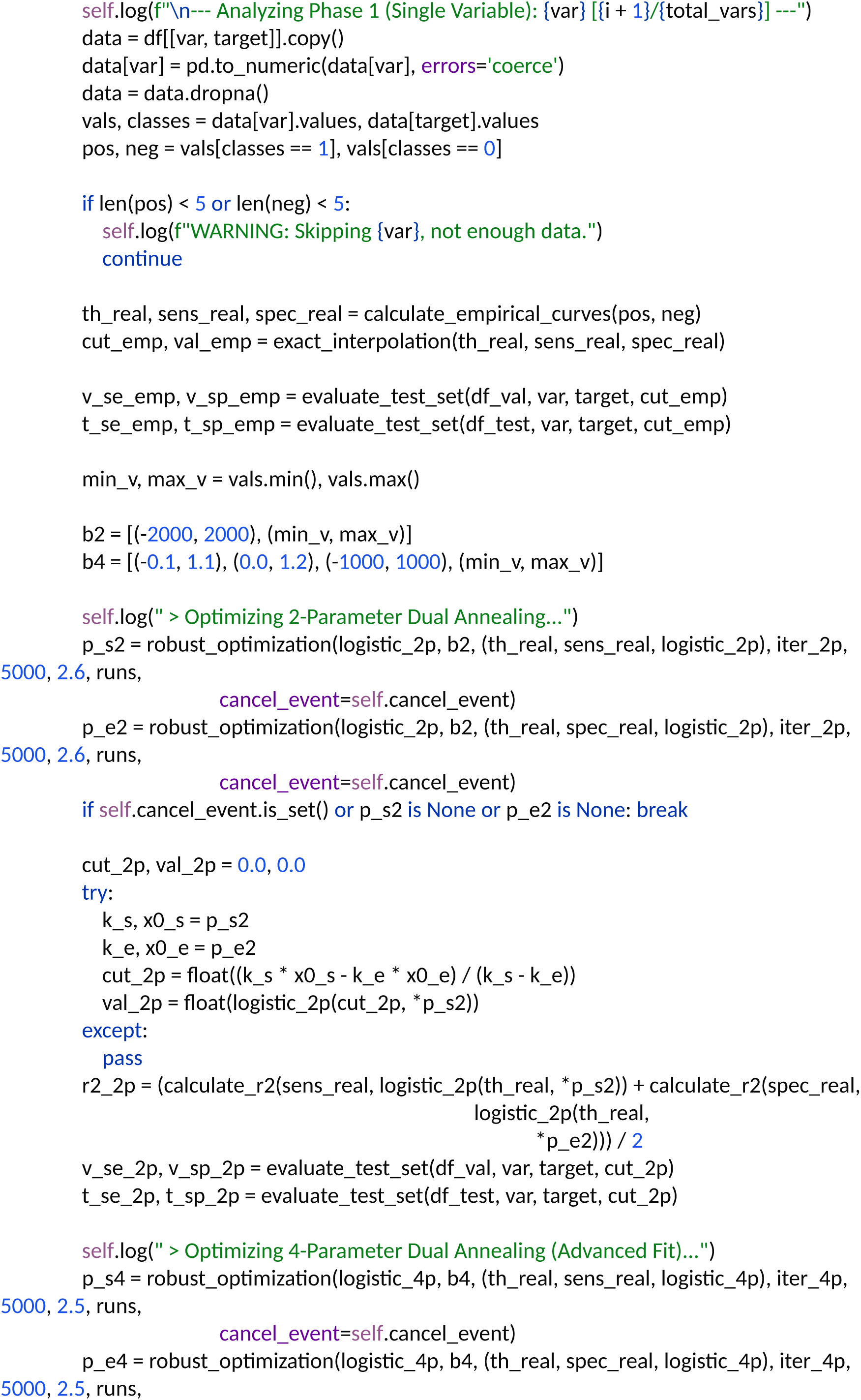
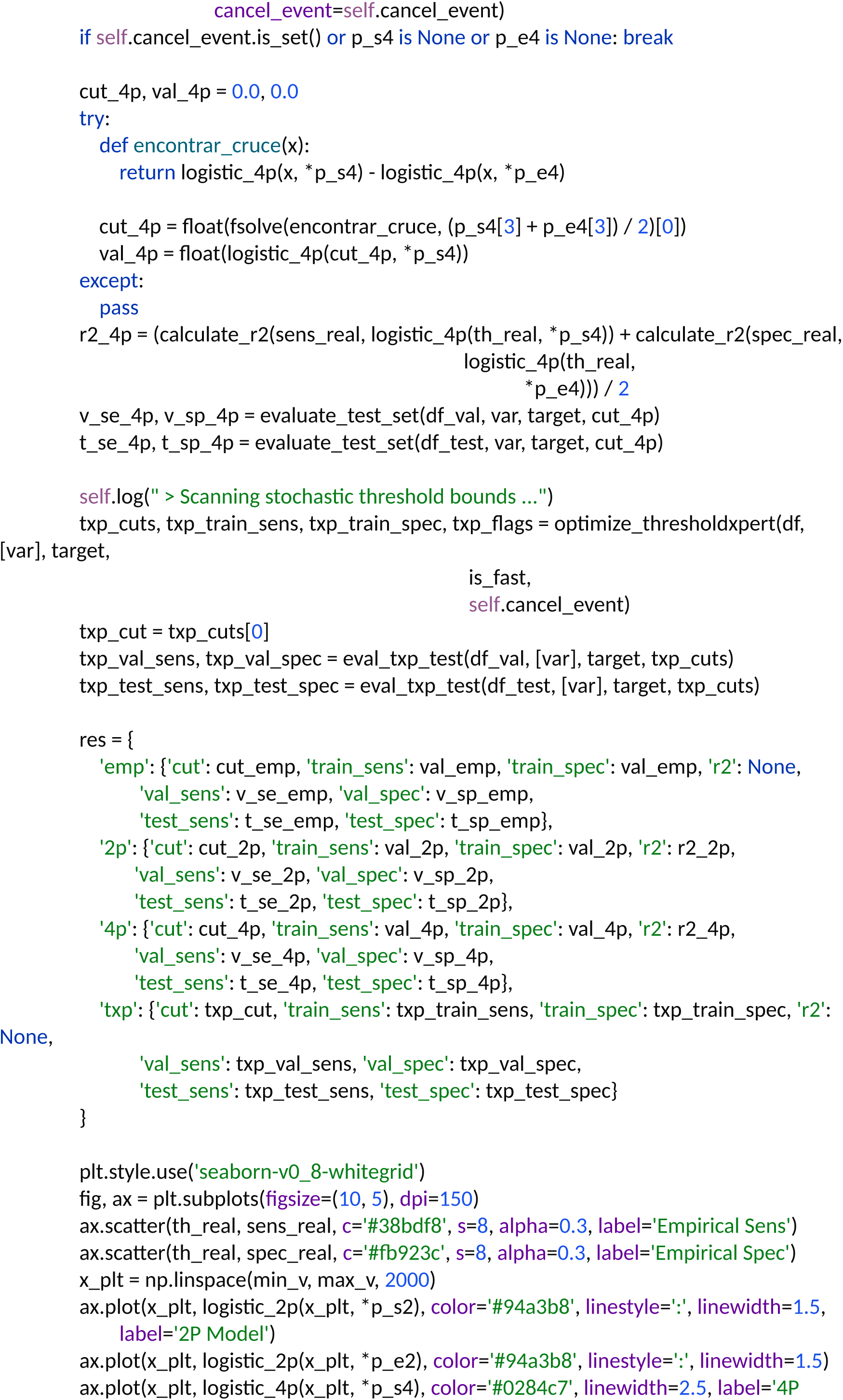
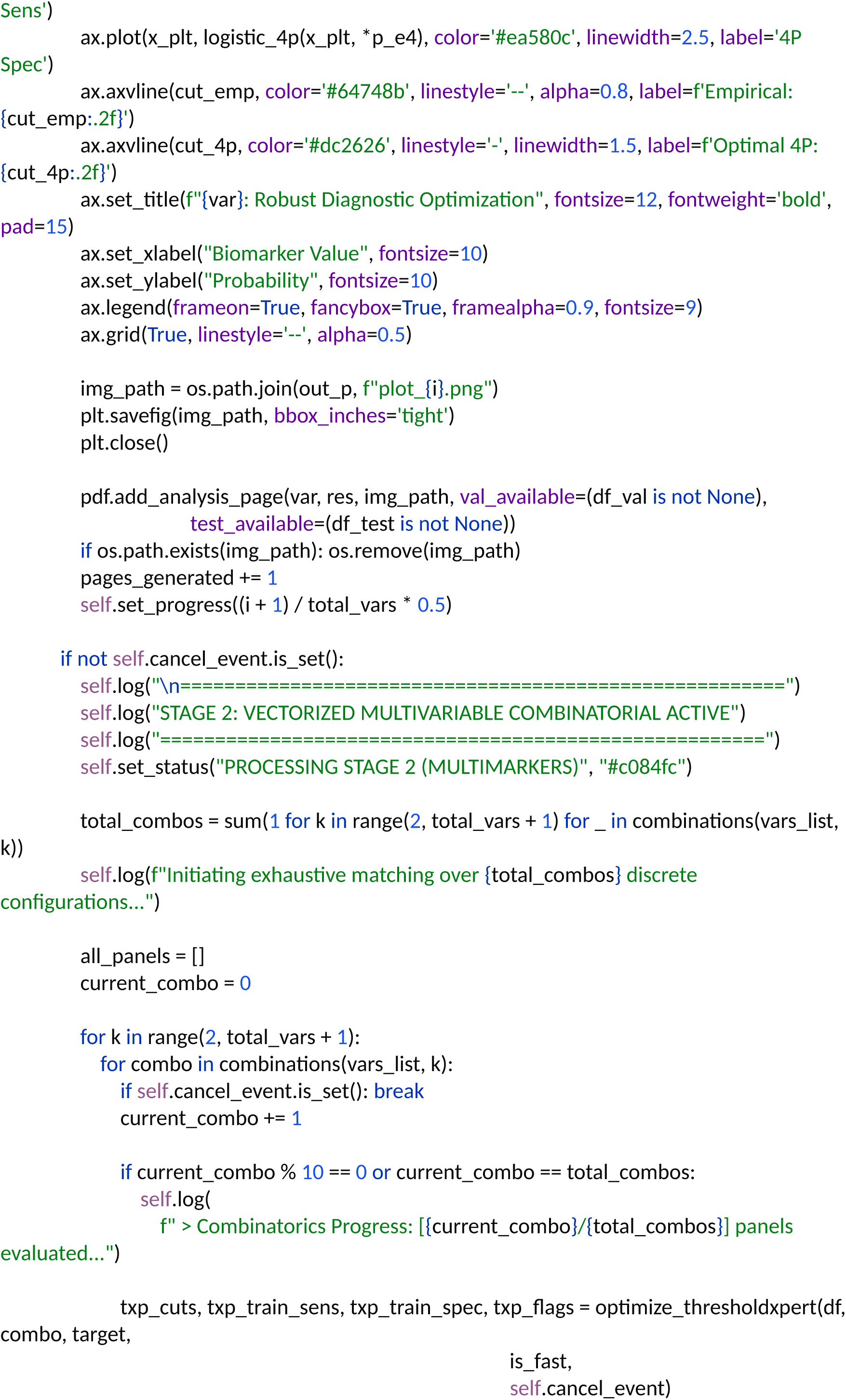
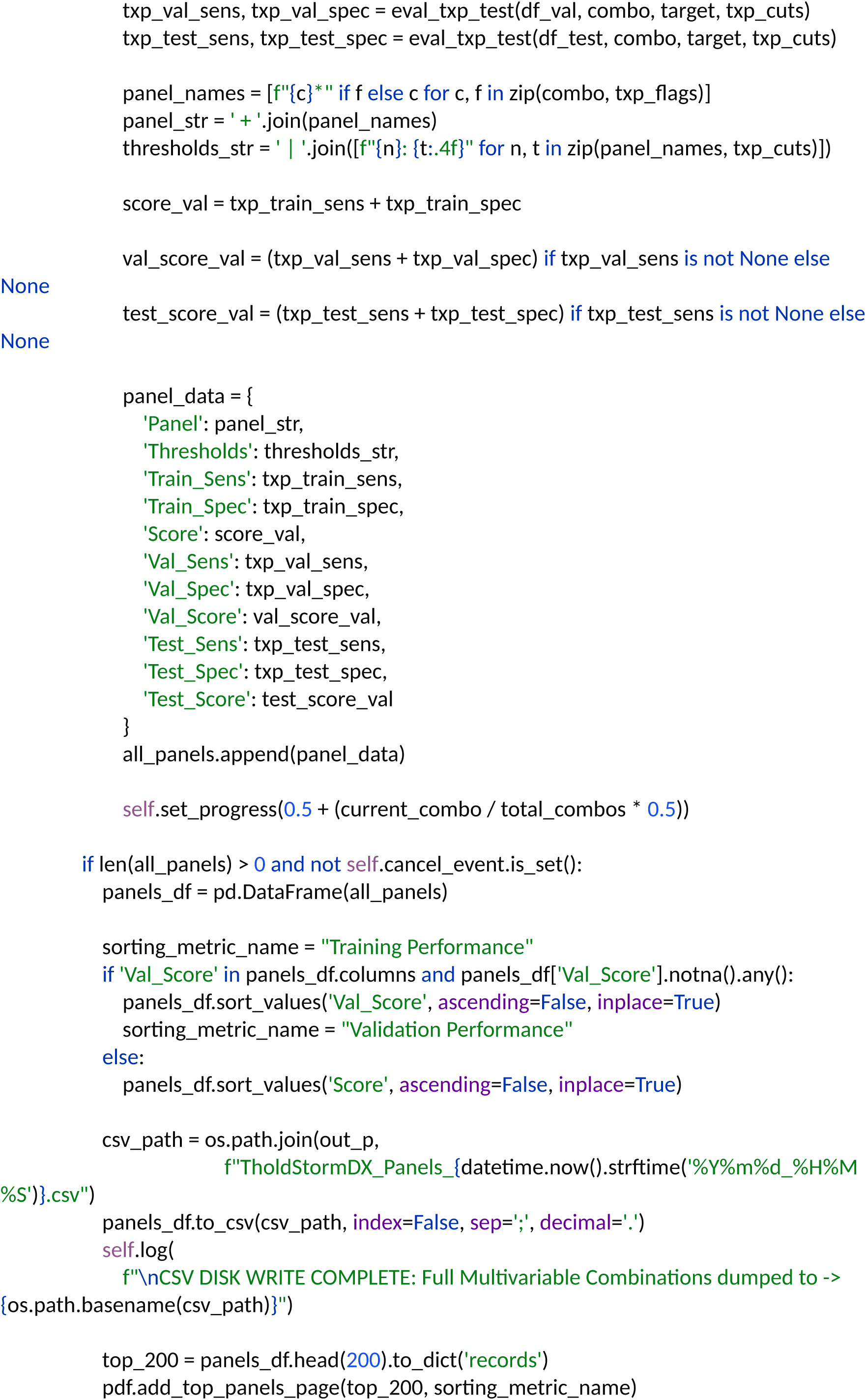
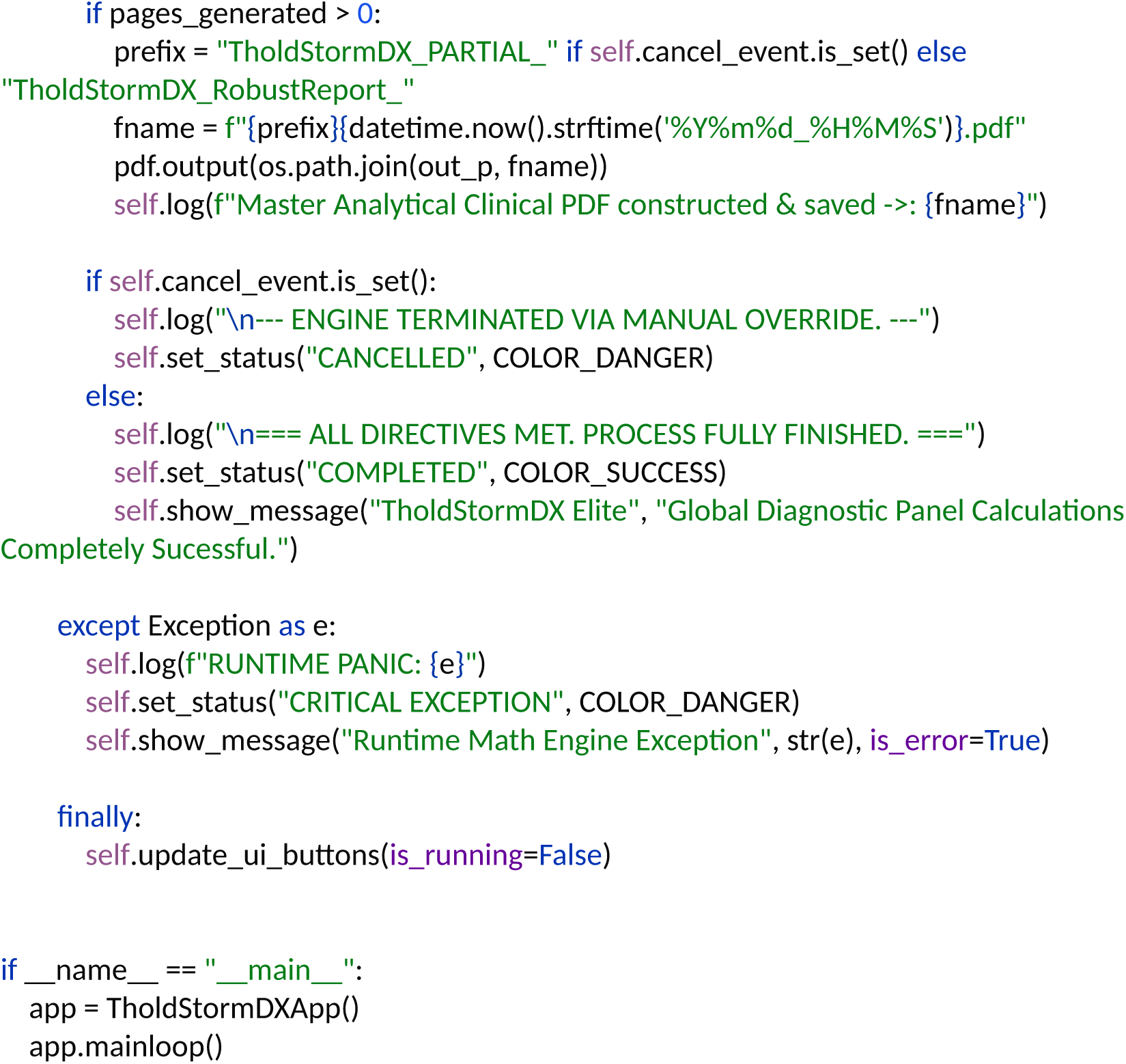

## References

1. Qiu J, Li R, Wang Y, Ma X, Qu C, Liu B, Yue W and Tian H (2023) A nomogram combining thoracic CT and tumor markers to predict the malignant grade of pulmonary nodules≤3 cm in diameter. Front. Oncol. 13:1196883. doi: 10.3389/fonc.2023.1196883

2. Jang ES, Jeong SH, Kim JW, Choi YS, Leissner P, et al. (2016) Diagnostic Performance of Alpha-Fetoprotein, Protein Induced by Vitamin K Absence, Osteopontin, Dickkopf-1 and Its Combinations for Hepatocellular Carcinoma. PLOS ONE 11(3): e0151069. 10.1371/journal.pone.0151069

3. Street, W.N., Wolberg, W.H. and Mangasarian, O.L. (1993) Nuclear Feature Extraction for Breast Tumor Diagnosis. International Symposium on Circuits and Systems, 5, 1945–1948. 10.1117/12.148698

4. Fernandes, K., Cardoso, J.S., Fernandes, J. (2017). Transfer Learning with Partial Observability Applied to Cervical Cancer Screening. In: Alexandre, L., Salvador Sánchez,J., Rodrigues, J. (eds) Pattern Recognition and Image Analysis. IbPRIA 2017. Lecture Notes in Computer Science(), vol 10255. Springer, Cham. 10.1007/978-3-319-58838-4_27

5. Olvi L. Mangasarian, W. Nick Street, William H. Wolberg, (1995) Breast Cancer Diagnosis and Prognosis Via Linear Programming. Operations Research 43(4):570–577. 10.1287/opre.43.4.570

6. Reinosa, R. (2025). Determination and Optimization of Diagnostic Cut-off Points for Multimarker Panels through a Two-Stage Approach and the Software Tool ThresholdXpert 1.0: Application to Risk Stratification of Pulmonary Nodules (Version 1). figshare. 10.6084/m9.figshare.30084427.v1

7. Reinosa Fernández, R. (2026). Integrated framework for the optimal determination of diagnostic cut-off points through empirical interpolation, logistic modeling optimized by dual annealing, and combinatorial optimization with ThresholdXpert: Application to hepatocellular carcinoma. medRxiv. 10.64898/2026.02.19.26346674

8. Xiang, Y., Gubian, S., Suomela, B., & Hoeng, J. (2013). Generalized simulated annealing for global optimization: The GenSA package. The R Journal, 5(1), 13–28. 10.32614/RJ-2013-002

