## Supplementary material. Outputs TholdStormDX for this study. for "Methodological and Clinical Validation of TholdStormDX v0.0.1: An Advanced Stochastic Engine for the Optimization of Thresholds and Multimarker Panels Applied to Oncology": Breast Diagnosis TholdStormDX_RobustReport_20260403_090535.pdf

### Biomarker: texture1

Processed: 01-Apr-2026 19:03

#### 1. Optimization Results

| MODEL | CUT-OFF | TRAIN (SE/SP) | VAL (SE/SP) | TEST (SE/SP) | R2 SCORE |
|---|---|---|---|---|---|
| Empirical (Exact) | 19.0736 | 0.760 / 0.760 | 0.833 / 0.703 | 0.698 / 0.810 | N/A |
| Logistic 2-Parameter | 19.2597 | 0.757 / 0.757 | 0.812 / 0.703 | 0.698 / 0.810 | 0.9966 |
| **Logistic 4-Parameter (Rec.)** | **19.2480** | **0.754 / 0.754** | **0.812 / 0.703** | **0.698 / 0.810** | **0.9971** |
| ThresholdXpert (Stochastic) | 19.1686 | 0.760 / 0.782 | 0.812 / 0.703 | 0.698 / 0.810 | N/A |

#### 2. Diagnostic Performance Curves (Training)

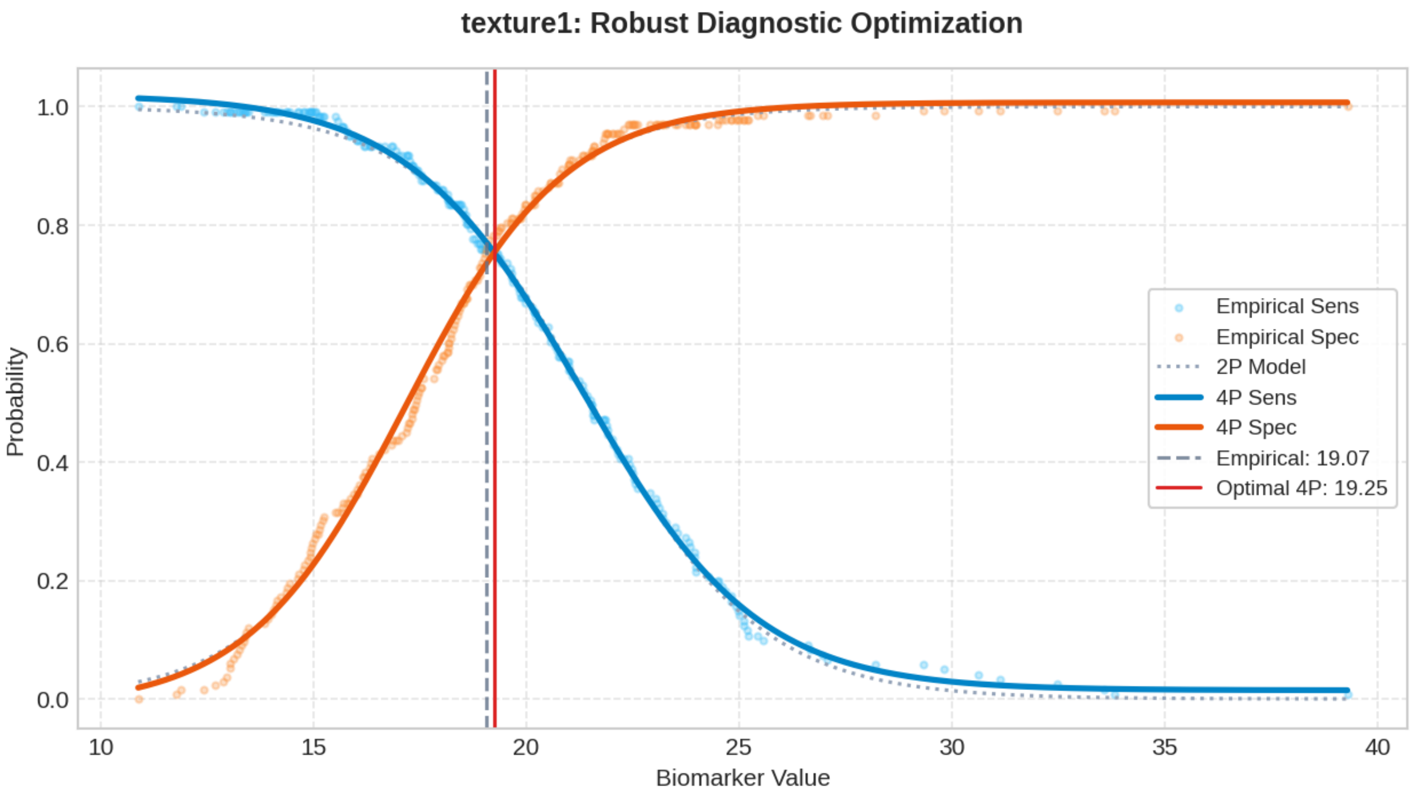

### Biomarker: area1

Processed: 01-Apr-2026 19:04

#### 1. Optimization Results

| MODEL | CUT-OFF | TRAIN (SE/SP) | VAL (SE/SP) | TEST (SE/SP) | R2 SCORE |
|---|---|---|---|---|---|
| Empirical (Exact) | 594.4329 | 0.884 / 0.884 | 0.771 / 0.811 | 0.930 / 0.881 | N/A |
| Logistic 2-Parameter | 595.3579 | 0.882 / 0.882 | 0.771 / 0.811 | 0.930 / 0.881 | 0.9926 |
| **Logistic 4-Parameter (Rec.)** | **596.1482** | **0.879 / 0.879** | **0.771 / 0.811** | **0.907 / 0.881** | **0.9970** |
| ThresholdXpert (Stochastic) | 593.2371 | 0.884 / 0.887 | 0.771 / 0.784 | 0.930 / 0.881 | N/A |

#### 2. Diagnostic Performance Curves (Training)

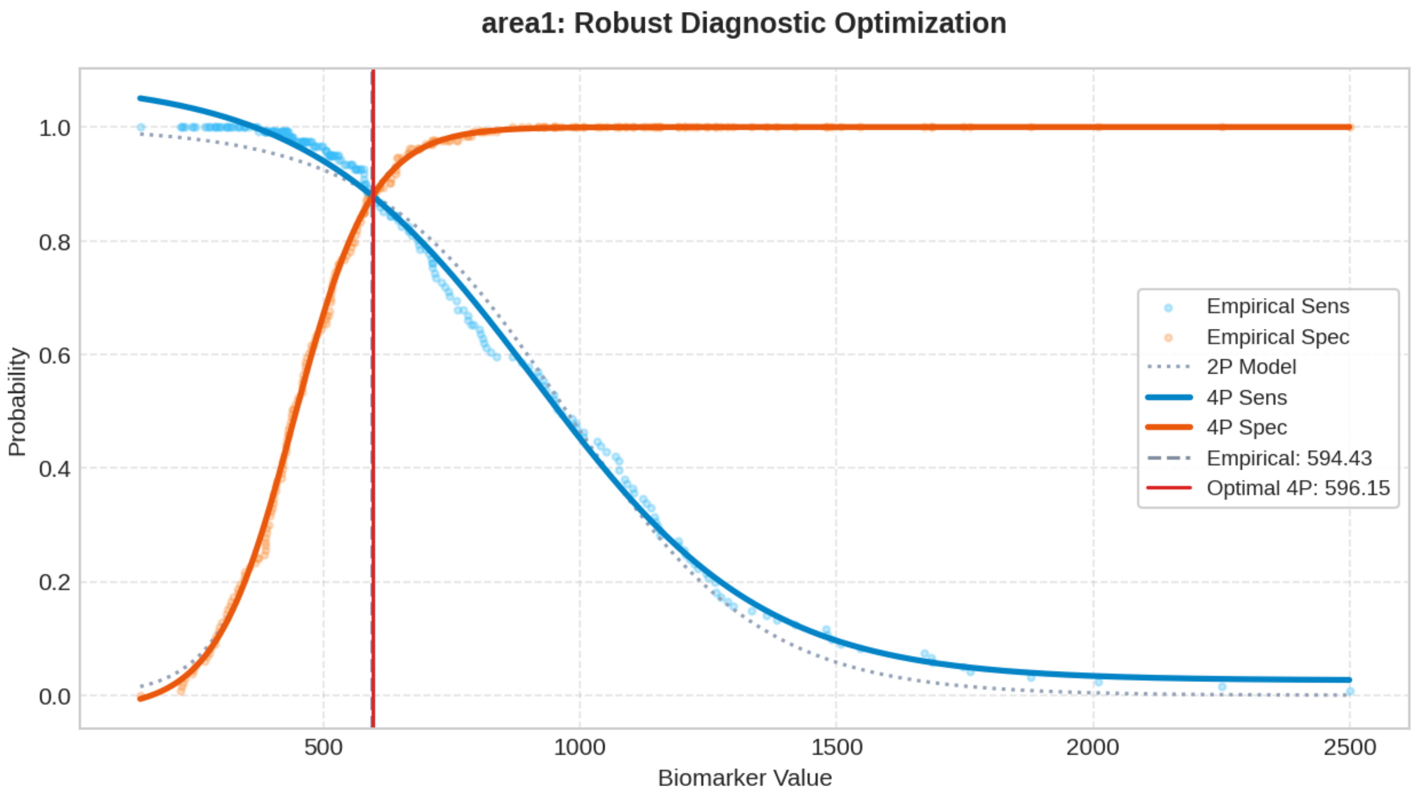

### Biomarker: concave_points1

Processed: 01-Apr-2026 19:07

#### 1. Optimization Results

| MODEL | CUT-OFF | TRAIN (SE/SP) | VAL (SE/SP) | TEST (SE/SP) | R2 SCORE |
|---|---|---|---|---|---|
| Empirical (Exact) | 0.0525 | 0.910 / 0.910 | 0.792 / 0.973 | 0.930 / 0.905 | N/A |
| Logistic 2-Parameter | 0.0444 | 0.919 / 0.919 | 0.875 / 0.946 | 0.977 / 0.881 | 0.9936 |
| **Logistic 4-Parameter (Rec.)** | **0.0464** | **0.918 / 0.918** | **0.854 / 0.973** | **0.977 / 0.881** | **0.9968** |
| ThresholdXpert (Stochastic) | 0.0498 | 0.926 / 0.910 | 0.833 / 0.973 | 0.953 / 0.881 | N/A |

#### 2. Diagnostic Performance Curves (Training)

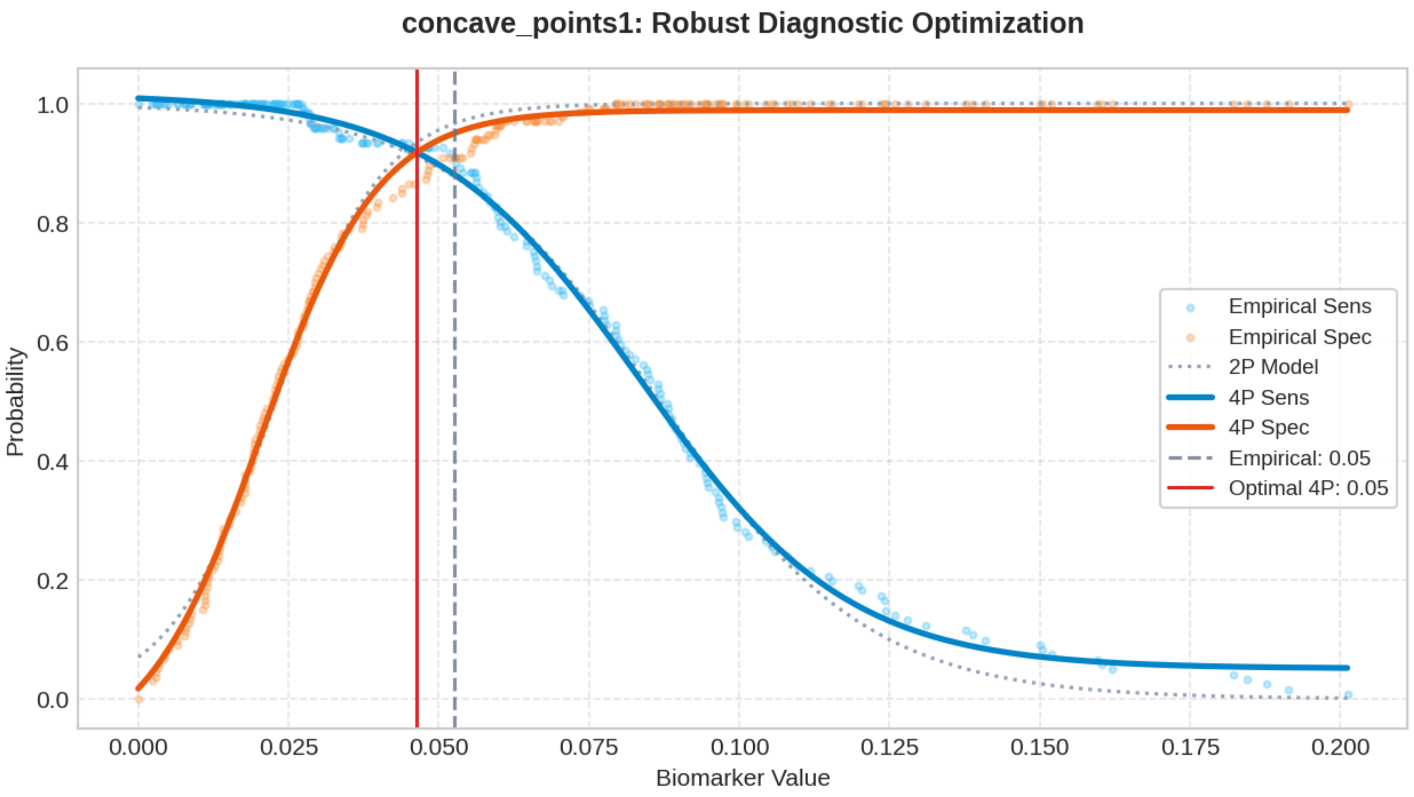

### Biomarker: area2

Processed: 01-Apr-2026 19:10

#### 1. Optimization Results

| MODEL | CUT-OFF | TRAIN (SE/SP) | VAL (SE/SP) | TEST (SE/SP) | R2 SCORE |
|---|---|---|---|---|---|
| Empirical (Exact) | 28.9003 | 0.868 / 0.868 | 0.792 / 0.811 | 0.860 / 0.857 | N/A |
| Logistic 2-Parameter | 28.0922 | 0.873 / 0.873 | 0.792 / 0.784 | 0.860 / 0.833 | 0.9816 |
| **Logistic 4-Parameter (Rec.)** | **28.5140** | **0.869 / 0.869** | **0.792 / 0.811** | **0.860 / 0.833** | **0.9974** |
| ThresholdXpert (Stochastic) | 28.9102 | 0.868 / 0.872 | 0.792 / 0.811 | 0.860 / 0.857 | N/A |

#### 2. Diagnostic Performance Curves (Training)

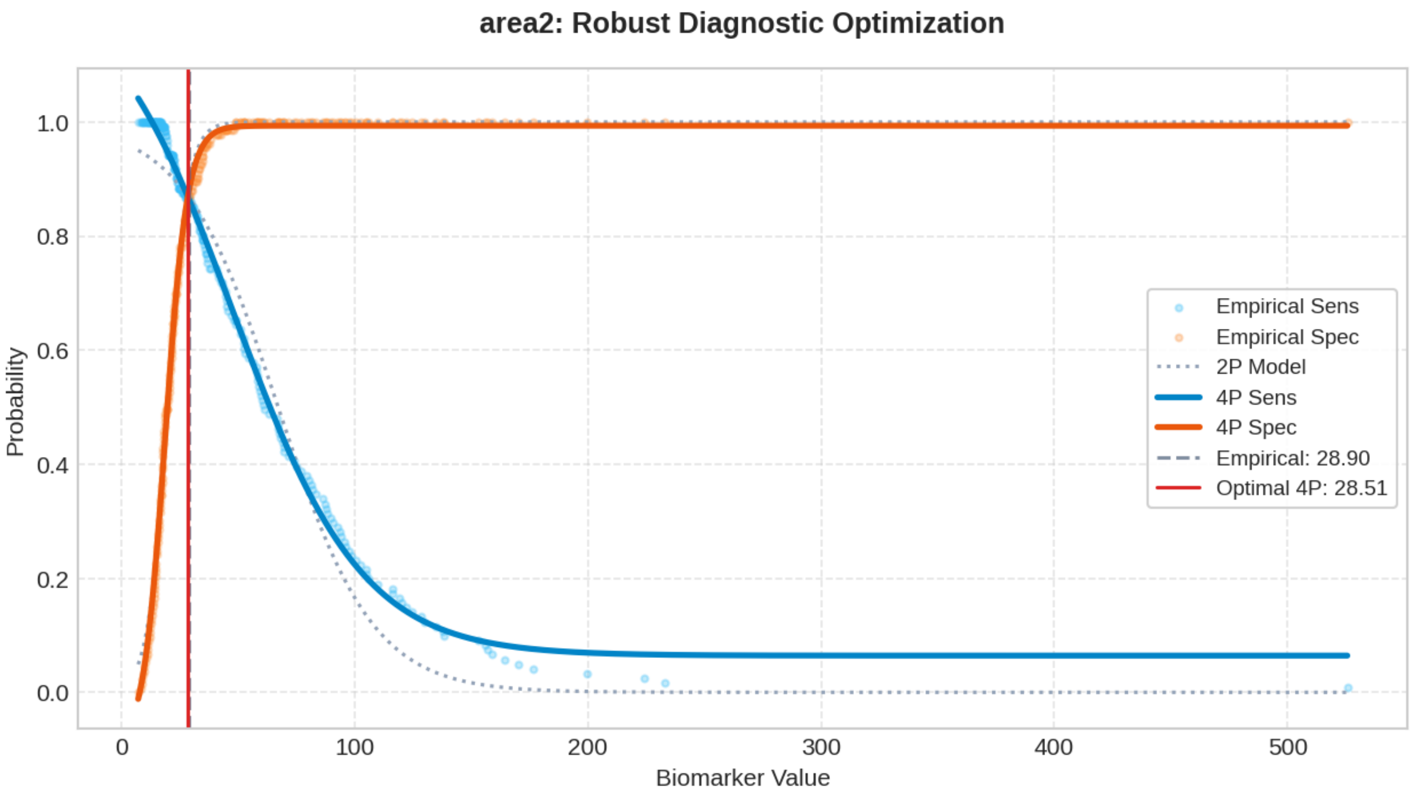

### Biomarker: texture3

Processed: 01-Apr-2026 19:13

#### 1. Optimization Results

| MODEL | CUT-OFF | TRAIN (SE/SP) | VAL (SE/SP) | TEST (SE/SP) | R2 SCORE |
|---|---|---|---|---|---|
| Empirical (Exact) | 25.5962 | 0.752 / 0.752 | 0.812 / 0.757 | 0.721 / 0.786 | N/A |
| Logistic 2-Parameter | 25.5860 | 0.759 / 0.759 | 0.812 / 0.757 | 0.721 / 0.786 | 0.9963 |
| **Logistic 4-Parameter (Rec.)** | **25.5509** | **0.757 / 0.757** | **0.812 / 0.730** | **0.721 / 0.786** | **0.9976** |
| ThresholdXpert (Stochastic) | 25.5005 | 0.760 / 0.752 | 0.812 / 0.730 | 0.721 / 0.786 | N/A |

#### 2. Diagnostic Performance Curves (Training)

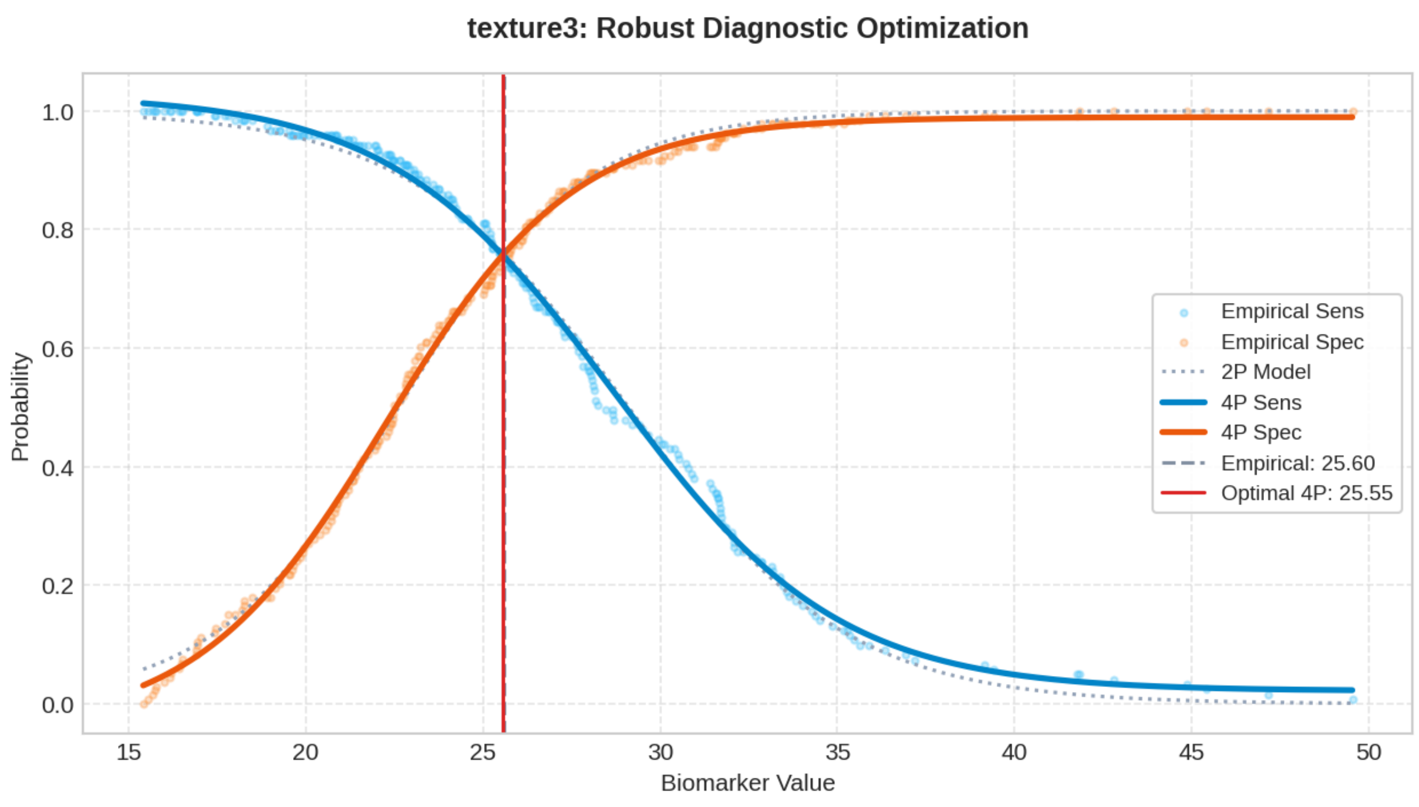

### Biomarker: perimeter3

Processed: 01-Apr-2026 19:17

#### 1. Optimization Results

| MODEL | CUT-OFF | TRAIN (SE/SP) | VAL (SE/SP) | TEST (SE/SP) | R2 SCORE |
|---|---|---|---|---|---|
| Empirical (Exact) | 105.8770 | 0.926 / 0.926 | 0.875 / 0.865 | 0.953 / 0.976 | N/A |
| Logistic 2-Parameter | 102.9635 | 0.914 / 0.914 | 0.917 / 0.865 | 0.953 / 0.952 | 0.9958 |
| **Logistic 4-Parameter (Rec.)** | **103.5714** | **0.917 / 0.917** | **0.917 / 0.865** | **0.953 / 0.952** | **0.9981** |
| ThresholdXpert (Stochastic) | 106.0561 | 0.926 / 0.932 | 0.875 / 0.865 | 0.930 / 1.000 | N/A |

#### 2. Diagnostic Performance Curves (Training)

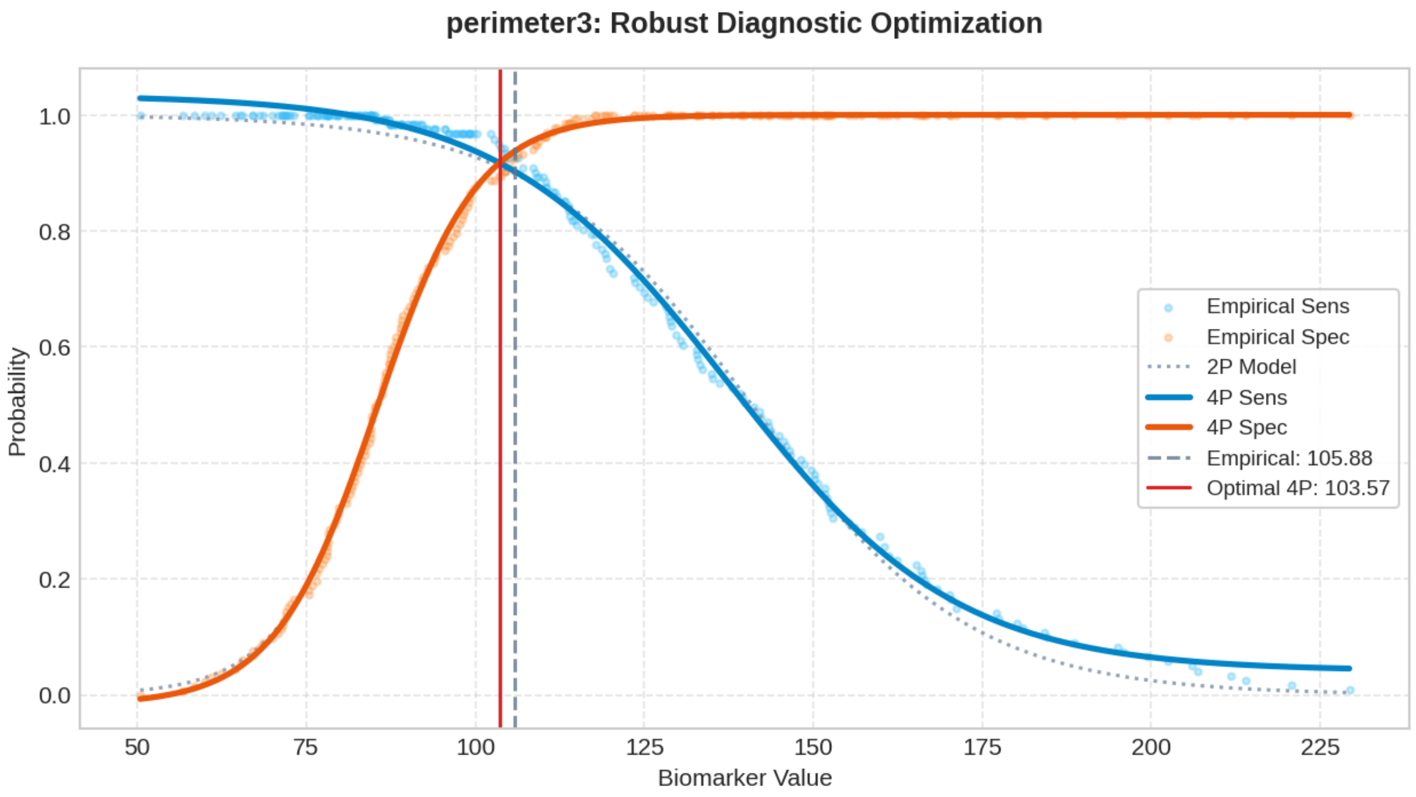

### Biomarker: smoothness3

Processed: 01-Apr-2026 19:20

#### 1. Optimization Results

| MODEL | CUT-OFF | TRAIN (SE/SP) | VAL (SE/SP) | TEST (SE/SP) | R2 SCORE |
|---|---|---|---|---|---|
| Empirical (Exact) | 0.1351 | 0.702 / 0.702 | 0.625 / 0.703 | 0.767 / 0.786 | N/A |
| Logistic 2-Parameter | 0.1346 | 0.696 / 0.696 | 0.646 / 0.703 | 0.767 / 0.762 | 0.9971 |
| **Logistic 4-Parameter (Rec.)** | **0.1346** | **0.692 / 0.692** | **0.646 / 0.703** | **0.767 / 0.762** | **0.9976** |
| ThresholdXpert (Stochastic) | 0.1351 | 0.702 / 0.707 | 0.625 / 0.703 | 0.767 / 0.786 | N/A |

#### 2. Diagnostic Performance Curves (Training)

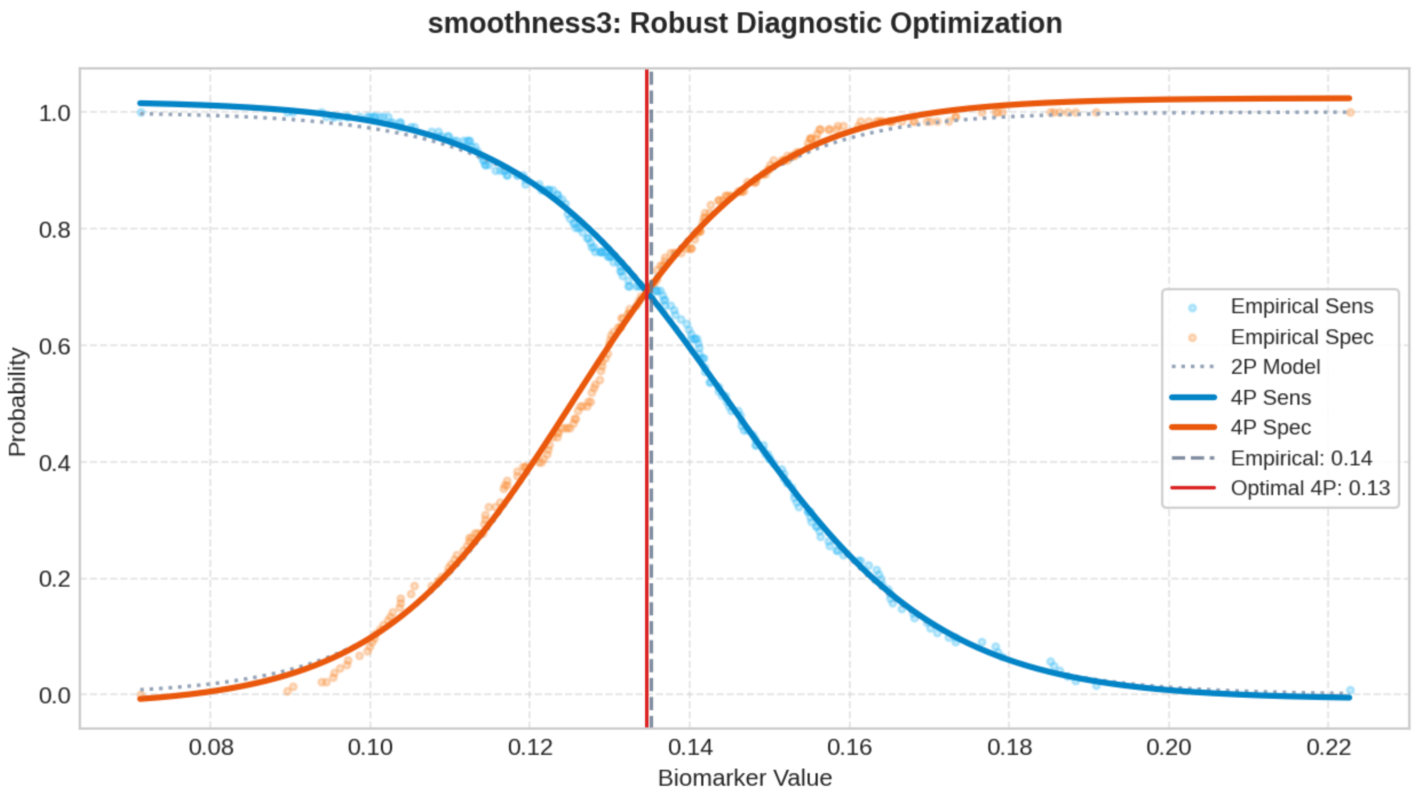

### Biomarker: compactness3

Processed: 01-Apr-2026 19:25

#### 1. Optimization Results

| MODEL | CUT-OFF | TRAIN (SE/SP) | VAL (SE/SP) | TEST (SE/SP) | R2 SCORE |
|---|---|---|---|---|---|
| Empirical (Exact) | 0.2298 | 0.776 / 0.776 | 0.812 / 0.865 | 0.837 / 0.667 | N/A |
| Logistic 2-Parameter | 0.2358 | 0.803 / 0.803 | 0.792 / 0.865 | 0.814 / 0.690 | 0.9914 |
| **Logistic 4-Parameter (Rec.)** | **0.2349** | **0.794 / 0.794** | **0.792 / 0.865** | **0.837 / 0.690** | **0.9965** |
| ThresholdXpert (Stochastic) | 0.2259 | 0.793 / 0.774 | 0.812 / 0.865 | 0.860 / 0.667 | N/A |

#### 2. Diagnostic Performance Curves (Training)

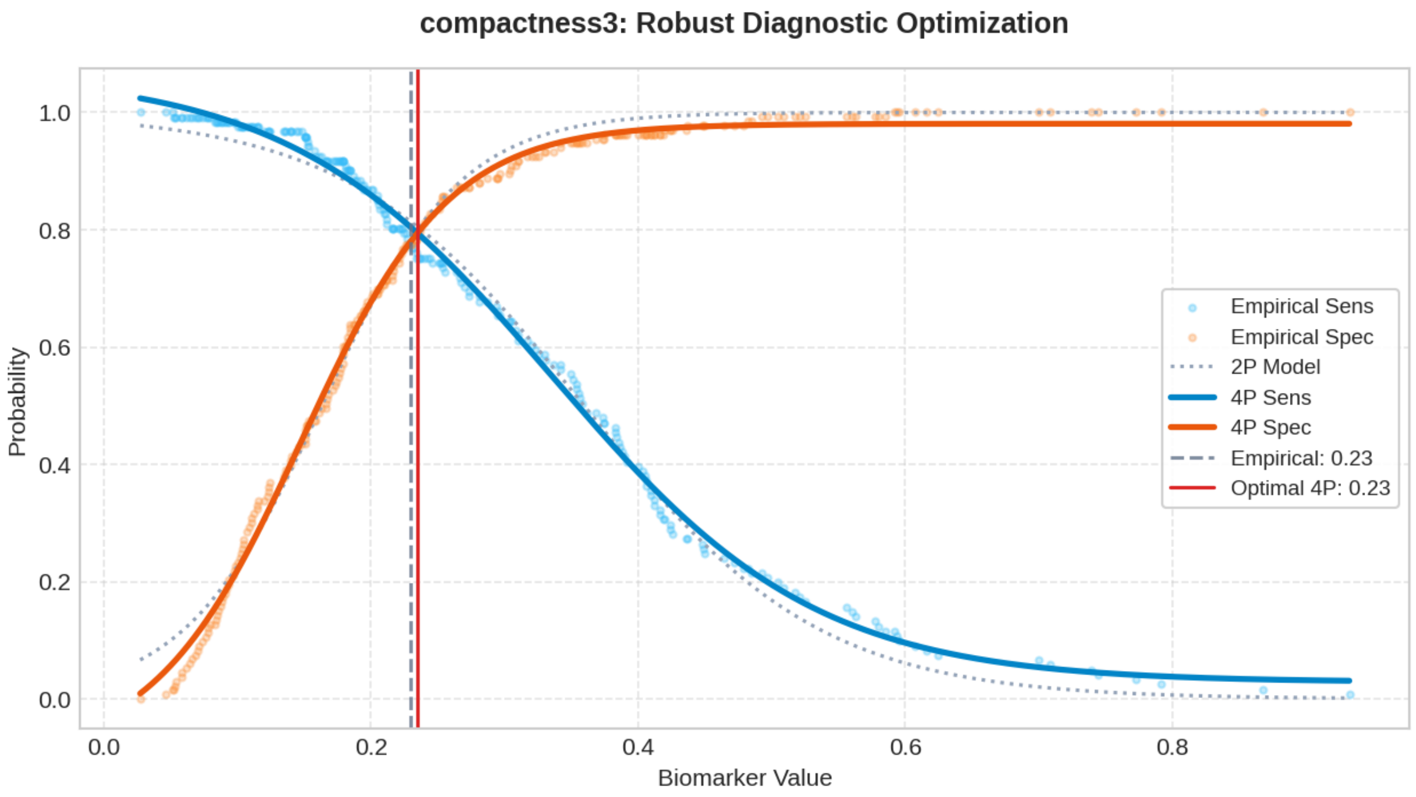

### Biomarker: concavity3

Processed: 01-Apr-2026 19:28

#### 1. Optimization Results

| MODEL | CUT-OFF | TRAIN (SE/SP) | VAL (SE/SP) | TEST (SE/SP) | R2 SCORE |
|---|---|---|---|---|---|
| Empirical (Exact) | 0.2657 | 0.876 / 0.876 | 0.875 / 0.892 | 0.884 / 0.833 | N/A |
| Logistic 2-Parameter | 0.2593 | 0.862 / 0.862 | 0.896 / 0.892 | 0.907 / 0.833 | 0.9909 |
| **Logistic 4-Parameter (Rec.)** | **0.2643** | **0.860 / 0.860** | **0.896 / 0.892** | **0.884 / 0.833** | **0.9973** |
| ThresholdXpert (Stochastic) | 0.2673 | 0.876 / 0.887 | 0.875 / 0.892 | 0.884 / 0.833 | N/A |

#### 2. Diagnostic Performance Curves (Training)

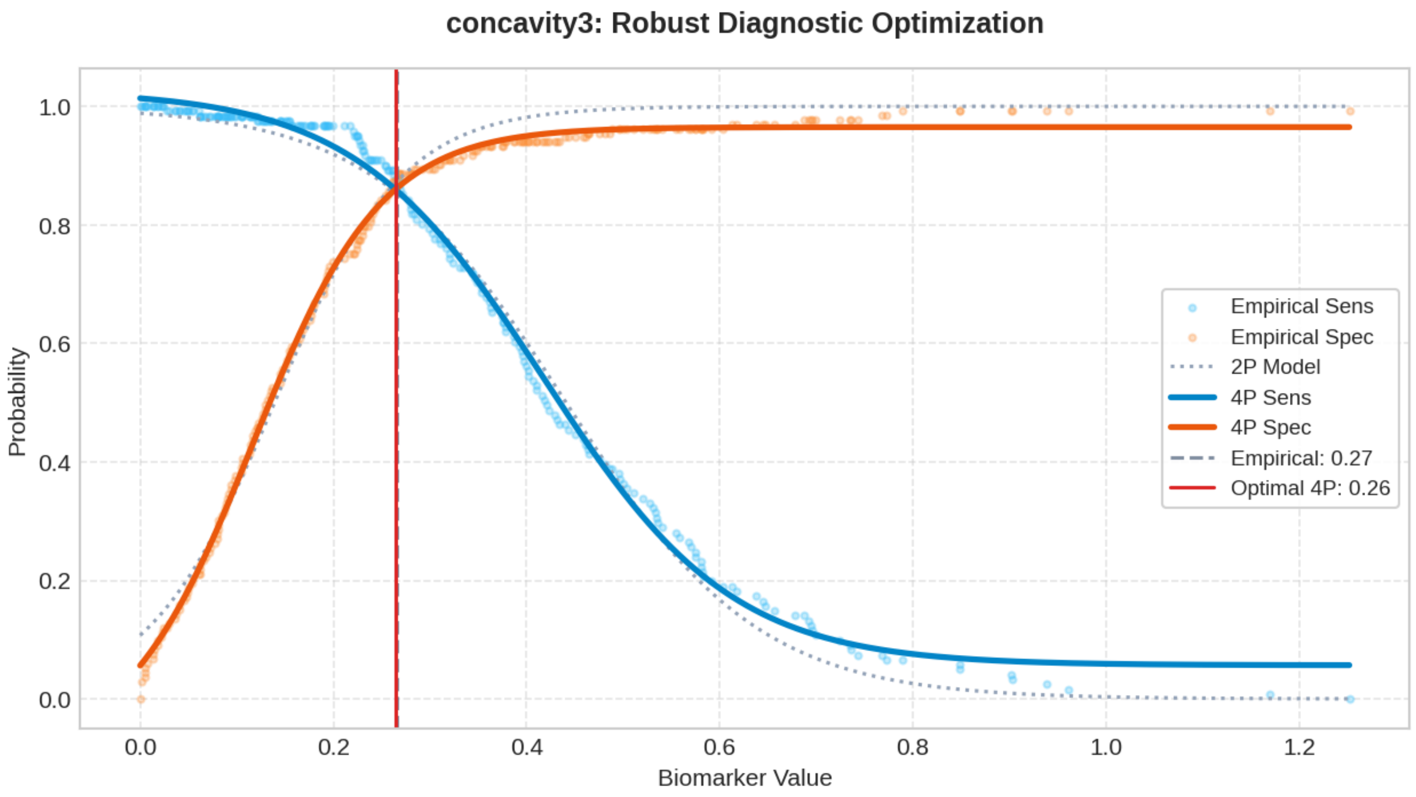

### Biomarker: concave_points3

Processed: 01-Apr-2026 19:32

#### 1. Optimization Results

| MODEL | CUT-OFF | TRAIN (SE/SP) | VAL (SE/SP) | TEST (SE/SP) | R2 SCORE |
|---|---|---|---|---|---|
| Empirical (Exact) | 0.1245 | 0.895 / 0.895 | 0.833 / 0.946 | 0.953 / 0.929 | N/A |
| Logistic 2-Parameter | 0.1212 | 0.904 / 0.904 | 0.854 / 0.946 | 0.953 / 0.905 | 0.9970 |
| **Logistic 4-Parameter (Rec.)** | **0.1228** | **0.902 / 0.902** | **0.833 / 0.946** | **0.953 / 0.929** | **0.9975** |
| ThresholdXpert (Stochastic) | 0.1222 | 0.901 / 0.895 | 0.854 / 0.946 | 0.953 / 0.929 | N/A |

#### 2. Diagnostic Performance Curves (Training)

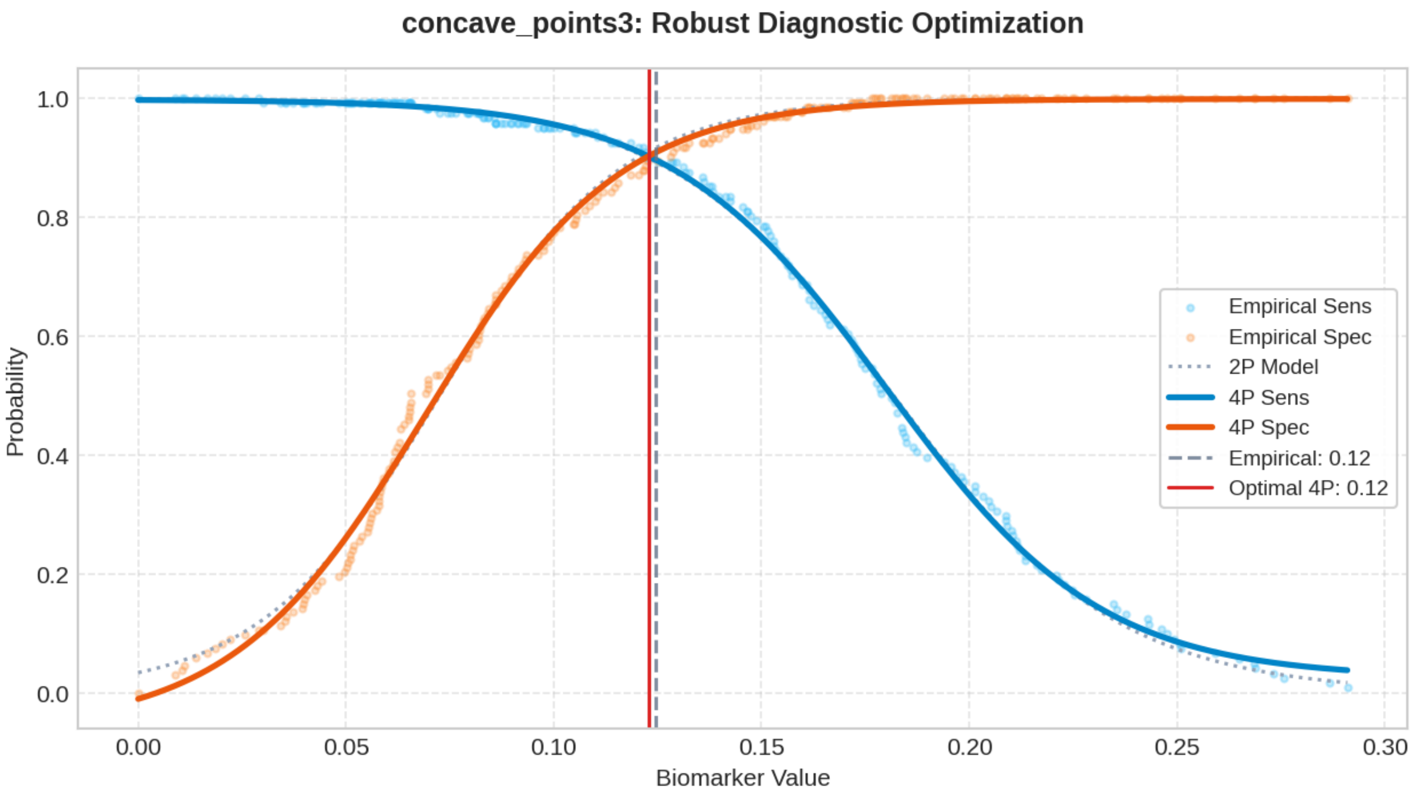

### Biomarker: symmetry3

Processed: 01-Apr-2026 19:36

#### 1. Optimization Results

| MODEL | CUT-OFF | TRAIN (SE/SP) | VAL (SE/SP) | TEST (SE/SP) | R2 SCORE |
|---|---|---|---|---|---|
| Empirical (Exact) | 0.2884 | 0.620 / 0.620 | 0.750 / 0.784 | 0.651 / 0.690 | N/A |
| Logistic 2-Parameter | 0.2900 | 0.650 / 0.650 | 0.729 / 0.784 | 0.628 / 0.690 | 0.9949 |
| **Logistic 4-Parameter (Rec.)** | **0.2897** | **0.646 / 0.646** | **0.729 / 0.784** | **0.628 / 0.690** | **0.9973** |
| ThresholdXpert (Stochastic) | 0.2915 | 0.620 / 0.654 | 0.708 / 0.784 | 0.605 / 0.690 | N/A |

#### 2. Diagnostic Performance Curves (Training)

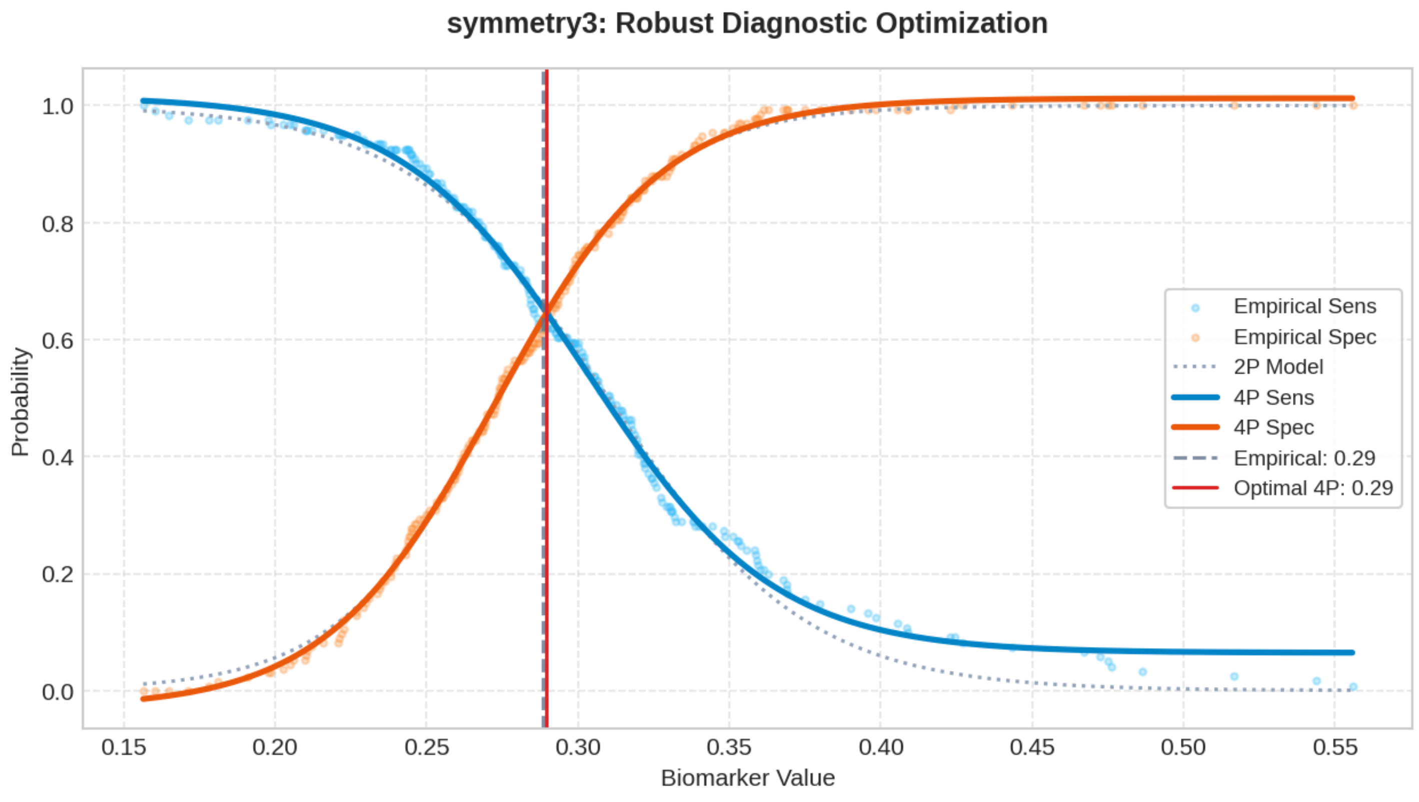

### Biomarker: fractal_dimension3

Processed: 01-Apr-2026 19:39

#### 1. Optimization Results

| MODEL | CUT-OFF | TRAIN (SE/SP) | VAL (SE/SP) | TEST (SE/SP) | R2 SCORE |
|---|---|---|---|---|---|
| Empirical (Exact) | 0.0807 | 0.632 / 0.632 | 0.646 / 0.649 | 0.698 / 0.643 | N/A |
| Logistic 2-Parameter | 0.0816 | 0.642 / 0.642 | 0.646 / 0.703 | 0.674 / 0.643 | 0.9870 |
| **Logistic 4-Parameter (Rec.)** | **0.0811** | **0.635 / 0.635** | **0.646 / 0.649** | **0.698 / 0.643** | **0.9954** |
| ThresholdXpert (Stochastic) | 0.0805 | 0.636 / 0.632 | 0.646 / 0.649 | 0.698 / 0.643 | N/A |

#### 2. Diagnostic Performance Curves (Training)

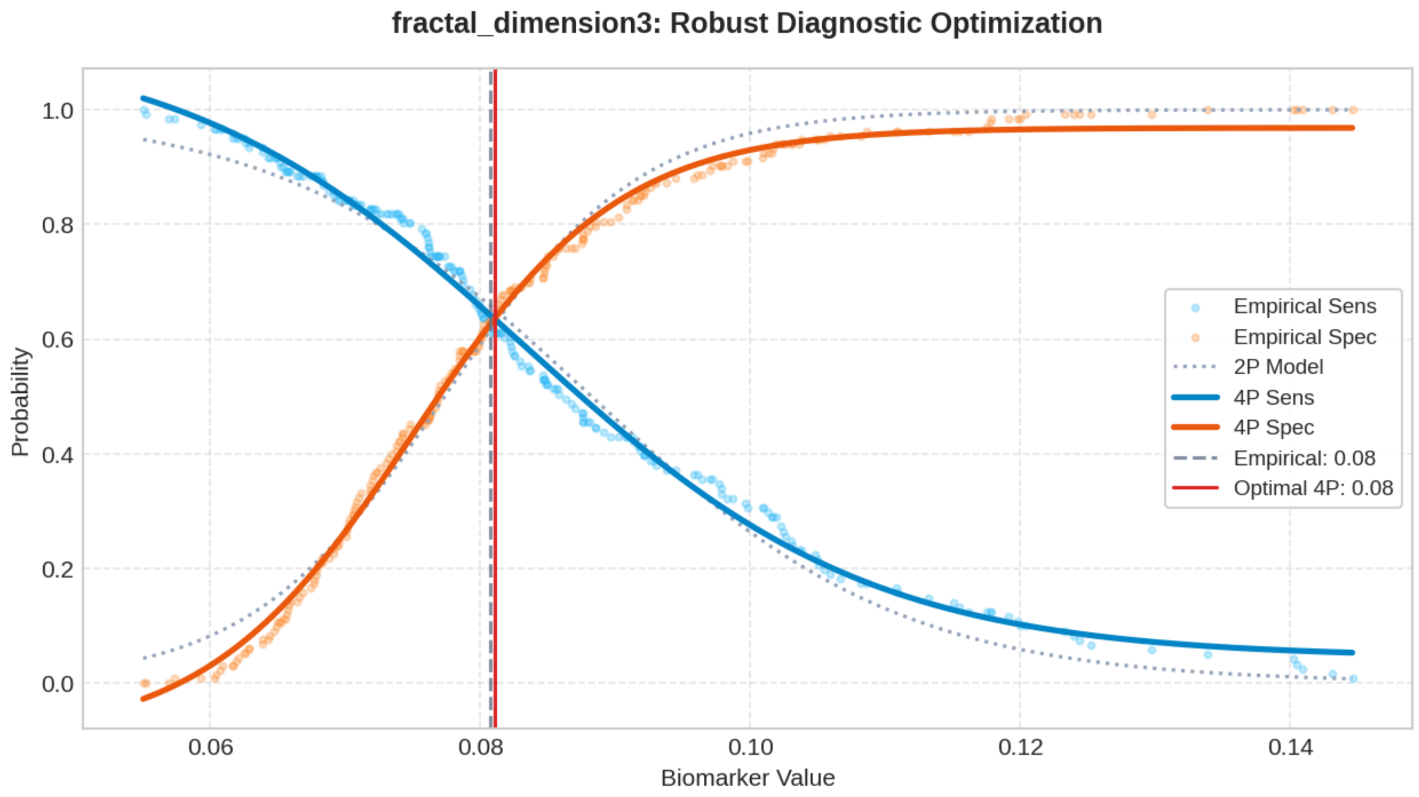

#### Top 200 Combinatorial Panels (ThresholdXpert OR-Logic)

The following multimarker panels have been optimized using high-performance vector-driven Monte Carlo simulations under a Boolean OR-logic framework. The engine employs a Max-Min Balancing logic (0.001 precision) to identify global threshold configurations that maximize the equilibrium between Sensitivity and Specificity across up to 10 million iterations. To ensure clinical robustness, results are sorted strictly by Validation Performance. (* Asterisk indicates an algorithmic threshold instability > 15%, suggesting potential data sparsity or high variance in the stochastic averaging process).

##### #1: area2 + compactness3 + symmetry3* + fractal_dimension3

Optimized Thresholds: area2: 36.5726 | compactness3: 0.3696 | symmetry3*: 0.4390 | fractal_dimension3: 0.1306

Train Sens: 0.901 | Train Spec: 0.917 [TRAIN SCORE: 1.818] || Val Sens: 0.938 | Val Spec: 0.919 [VAL SCORE: 1.856] || Test Sens: 0.907 | Test Spec: 0.857 [TEST SCORE: 1.764]

##### #2: texture1 + area2 + compactness3 + fractal_dimension3

Optimized Thresholds: texture1: 37.9022 | area2: 37.4908 | compactness3: 0.3632 | fractal_dimension3: 0.1342

Train Sens: 0.884 | Train Spec: 0.917 [TRAIN SCORE: 1.802] || Val Sens: 0.938 | Val Spec: 0.919 [VAL SCORE: 1.856] || Test Sens: 0.884 | Test Spec: 0.857 [TEST SCORE: 1.741]

##### #3: area2 + compactness3 + symmetry3

Optimized Thresholds: area2: 36.6146 | compactness3: 0.3702 | symmetry3: 0.4923

Train Sens: 0.893 | Train Spec: 0.917 [TRAIN SCORE: 1.810] || Val Sens: 0.938 | Val Spec: 0.919 [VAL SCORE: 1.856] || Test Sens: 0.884 | Test Spec: 0.857 [TEST SCORE: 1.741]

##### #4: area2 + texture3* + compactness3 + fractal_dimension3

Optimized Thresholds: area2: 38.4413 | texture3*: 40.8514 | compactness3: 0.3696 | fractal_dimension3: 0.1278

Train Sens: 0.884 | Train Spec: 0.925 [TRAIN SCORE: 1.809] || Val Sens: 0.938 | Val Spec: 0.919 [VAL SCORE: 1.856] || Test Sens: 0.884 | Test Spec: 0.857 [TEST SCORE: 1.741]

##### #5: texture1 + area2 + concavity3

Optimized Thresholds: texture1: 36.4653 | area2: 43.0750 | concavity3: 0.3507

Train Sens: 0.934 | Train Spec: 0.917 [TRAIN SCORE: 1.851] || Val Sens: 0.875 | Val Spec: 0.973 [VAL SCORE: 1.848] || Test Sens: 0.860 | Test Spec: 0.881 [TEST SCORE: 1.741]

##### #6: area2 + texture3 + smoothness3 + concavity3 + fractal_dimension3

Optimized Thresholds: area2: 42.7978 | texture3: 43.4203 | smoothness3: 0.2034 | concavity3: 0.3488 | fractal_dimension3: 0.1244

Train Sens: 0.934 | Train Spec: 0.917 [TRAIN SCORE: 1.851] || Val Sens: 0.875 | Val Spec: 0.973 [VAL SCORE: 1.848] || Test Sens: 0.860 | Test Spec: 0.881 [TEST SCORE: 1.741]

##### #7: area2 + smoothness3 + concavity3 + symmetry3*

Optimized Thresholds: area2: 42.9474 | smoothness3: 0.2105 | concavity3: 0.3503 | symmetry3*: 0.4880

Train Sens: 0.934 | Train Spec: 0.917 [TRAIN SCORE: 1.851] || Val Sens: 0.875 | Val Spec: 0.973 [VAL SCORE: 1.848] || Test Sens: 0.860 | Test Spec: 0.881 [TEST SCORE: 1.741]

##### #8: texture1 + area2 + texture3 + smoothness3 + concavity3

Optimized Thresholds: texture1: 35.6348 | area2: 43.1810 | texture3: 47.9319 | smoothness3: 0.1987 | concavity3: 0.3512

Train Sens: 0.917 | Train Spec: 0.917 [TRAIN SCORE: 1.835] || Val Sens: 0.875 | Val Spec: 0.973 [VAL SCORE: 1.848] || Test Sens: 0.860 | Test Spec: 0.881 [TEST SCORE: 1.741]

##### #9: area2 + texture3 + concavity3

Optimized Thresholds: area2: 42.9604 | texture3: 46.6856 | concavity3: 0.3497

Train Sens: 0.934 | Train Spec: 0.917 [TRAIN SCORE: 1.851] || Val Sens: 0.875 | Val Spec: 0.973 [VAL SCORE: 1.848] || Test Sens: 0.860 | Test Spec: 0.881 [TEST SCORE: 1.741]

##### #10: area2 + smoothness3 + concavity3

Optimized Thresholds: area2: 42.9662 | smoothness3: 0.2049 | concavity3: 0.3497

Train Sens: 0.934 | Train Spec: 0.917 [TRAIN SCORE: 1.851] || Val Sens: 0.875 | Val Spec: 0.973 [VAL SCORE: 1.848] || Test Sens: 0.860 | Test Spec: 0.881 [TEST SCORE: 1.741]

##### #11: area2 + smoothness3 + concavity3 + symmetry3 + fractal_dimension3

Optimized Thresholds: area2: 43.2743 | smoothness3: 0.1900 | concavity3: 0.3492 | symmetry3: 0.5317 | fractal_dimension3: 0.1286

Train Sens: 0.926 | Train Spec: 0.917 [TRAIN SCORE: 1.843] || Val Sens: 0.875 | Val Spec: 0.973 [VAL SCORE: 1.848] || Test Sens: 0.860 | Test Spec: 0.881 [TEST SCORE: 1.741]

##### #12: area2 + smoothness3 + concavity3 + fractal_dimension3

Optimized Thresholds: area2: 42.9484 | smoothness3: 0.2120 | concavity3: 0.3502 | fractal_dimension3: 0.1383

Train Sens: 0.934 | Train Spec: 0.917 [TRAIN SCORE: 1.851] || Val Sens: 0.875 | Val Spec: 0.973 [VAL SCORE: 1.848] || Test Sens: 0.860 | Test Spec: 0.881 [TEST SCORE: 1.741]

##### #13: area2 + concavity3

Optimized Thresholds: area2: 42.9625 | concavity3: 0.3497

Train Sens: 0.934 | Train Spec: 0.917 [TRAIN SCORE: 1.851] || Val Sens: 0.875 | Val Spec: 0.973 [VAL SCORE: 1.848] || Test Sens: 0.860 | Test Spec: 0.881 [TEST SCORE: 1.741]

##### #14: area2 + concavity3 + symmetry3

Optimized Thresholds: area2: 42.9566 | concavity3: 0.3496 | symmetry3: 0.4370

Train Sens: 0.934 | Train Spec: 0.917 [TRAIN SCORE: 1.851] || Val Sens: 0.875 | Val Spec: 0.973 [VAL SCORE: 1.848] || Test Sens: 0.884 | Test Spec: 0.881 [TEST SCORE: 1.765]

##### #15: area2* + texture3* + perimeter3 + compactness3 + concavity3 + concave_points3

Optimized Thresholds: area2*: 195.2117 | texture3*: 36.0632 | perimeter3: 114.9344 | compactness3: 0.7394 | concavity3: 0.9606 | concave_points3: 0.1668

Train Sens: 0.884 | Train Spec: 0.970 [TRAIN SCORE: 1.854] || Val Sens: 0.875 | Val Spec: 0.973 [VAL SCORE: 1.848] || Test Sens: 0.907 | Test Spec: 0.976 [TEST SCORE: 1.883]

##### #16: area2 + concavity3 + fractal_dimension3

Optimized Thresholds: area2: 42.9372 | concavity3: 0.3495 | fractal_dimension3: 0.1275

Train Sens: 0.934 | Train Spec: 0.917 [TRAIN SCORE: 1.851] || Val Sens: 0.875 | Val Spec: 0.973 [VAL SCORE: 1.848] || Test Sens: 0.860 | Test Spec: 0.881 [TEST SCORE: 1.741]

##### #17: area2 + texture3 + concavity3 + symmetry3 + fractal_dimension3

Optimized Thresholds: area2: 43.2743 | texture3: 42.1907 | concavity3: 0.3492 | symmetry3: 0.5317 | fractal_dimension3: 0.1286

Train Sens: 0.926 | Train Spec: 0.917 [TRAIN SCORE: 1.843] || Val Sens: 0.875 | Val Spec: 0.973 [VAL SCORE: 1.848] || Test Sens: 0.860 | Test Spec: 0.881 [TEST SCORE: 1.741]

###### #18: area2 + texture3 + smoothness3 + compactness3 + concavity3* + symmetry3

Optimized Thresholds: area2: 40.2771 | texture3: 45.3140 | smoothness3: 0.1831 | compactness3: 0.3665 | concavity3*: 0.8121 | symmetry3: 0.3974

Train Sens: 0.893 | Train Spec: 0.932 [TRAIN SCORE: 1.825] || Val Sens: 0.875 | Val Spec: 0.973 [VAL SCORE: 1.848] || Test Sens: 0.907 | Test Spec: 0.881 [TEST SCORE: 1.788]

###### #19: area2 + smoothness3 + compactness3 + symmetry3

Optimized Thresholds: area2: 40.2998 | smoothness3: 0.1827 | compactness3: 0.3691 | symmetry3: 0.4403

Train Sens: 0.893 | Train Spec: 0.932 [TRAIN SCORE: 1.825] || Val Sens: 0.875 | Val Spec: 0.973 [VAL SCORE: 1.848] || Test Sens: 0.907 | Test Spec: 0.881 [TEST SCORE: 1.788]

###### #20: area2 + texture3 + concavity3 + fractal_dimension3

Optimized Thresholds: area2: 42.9484 | texture3: 47.1405 | concavity3: 0.3502 | fractal_dimension3: 0.1383

Train Sens: 0.934 | Train Spec: 0.917 [TRAIN SCORE: 1.851] || Val Sens: 0.875 | Val Spec: 0.973 [VAL SCORE: 1.848] || Test Sens: 0.860 | Test Spec: 0.881 [TEST SCORE: 1.741]

###### #21: area1* + area2* + texture3 + perimeter3 + concavity3 + concave_points3 + fractal_dimension3

Optimized Thresholds: area1*: 1296.9868 | area2*: 193.2521 | texture3: 36.4042 | perimeter3: 114.4103 | concavity3: 1.0170 | concave_points3: 0.1630 | fractal_dimension3: 0.1393

Train Sens: 0.901 | Train Spec: 0.970 [TRAIN SCORE: 1.871] || Val Sens: 0.875 | Val Spec: 0.973 [VAL SCORE: 1.848] || Test Sens: 0.907 | Test Spec: 0.952 [TEST SCORE: 1.859]

###### #22: texture1 + area2 + texture3 + concavity3

Optimized Thresholds: texture1: 36.2558 | area2: 42.9794 | texture3: 44.4426 | concavity3: 0.3487

Train Sens: 0.934 | Train Spec: 0.917 [TRAIN SCORE: 1.851] || Val Sens: 0.875 | Val Spec: 0.973 [VAL SCORE: 1.848] || Test Sens: 0.860 | Test Spec: 0.881 [TEST SCORE: 1.741]

###### #23: texture1 + area2 + concavity3 + fractal_dimension3

Optimized Thresholds: texture1: 33.9234 | area2: 42.7791 | concavity3: 0.3500 | fractal_dimension3: 0.1283

Train Sens: 0.934 | Train Spec: 0.917 [TRAIN SCORE: 1.851] || Val Sens: 0.875 | Val Spec: 0.973 [VAL SCORE: 1.848] || Test Sens: 0.860 | Test Spec: 0.881 [TEST SCORE: 1.741]

###### #24: texture1 + area2 + texture3 + compactness3 + concavity3 + fractal_dimension3

Optimized Thresholds: texture1: 37.5311 | area2: 43.0989 | texture3: 46.8512 | compactness3: 0.3723 | concavity3: 0.3499 | fractal_dimension3: 0.1205

Train Sens: 0.934 | Train Spec: 0.917 [TRAIN SCORE: 1.851] || Val Sens: 0.875 | Val Spec: 0.973 [VAL SCORE: 1.848] || Test Sens: 0.884 | Test Spec: 0.857 [TEST SCORE: 1.741]

###### #25: texture1 + area2 + texture3 + smoothness3 + compactness3 + concavity3

Optimized Thresholds: texture1: 37.5255 | area2: 41.8686 | texture3: 44.1954 | smoothness3: 0.2077 | compactness3: 0.4136 | concavity3: 0.3643

Train Sens: 0.909 | Train Spec: 0.910 [TRAIN SCORE: 1.819] || Val Sens: 0.875 | Val Spec: 0.973 [VAL SCORE: 1.848] || Test Sens: 0.860 | Test Spec: 0.857 [TEST SCORE: 1.718]

**#26: area2 + texture3 + concavity3 + symmetry3***

Optimized Thresholds: area2: 42.9474 | texture3: 46.8108 | concavity3: 0.3503 | symmetry3*: 0.4880

Train Sens: 0.934 | Train Spec: 0.917 [TRAIN SCORE: 1.851] || Val Sens: 0.875 | Val Spec: 0.973 [VAL SCORE: 1.848] || Test Sens: 0.860 | Test Spec: 0.881 [TEST SCORE: 1.741]

**#27: area2 + concavity3 + symmetry3 + fractal_dimension3**

Optimized Thresholds: area2: 42.9495 | concavity3: 0.3498 | symmetry3: 0.4752 | fractal_dimension3: 0.1260

Train Sens: 0.934 | Train Spec: 0.917 [TRAIN SCORE: 1.851] || Val Sens: 0.875 | Val Spec: 0.973 [VAL SCORE: 1.848] || Test Sens: 0.860 | Test Spec: 0.881 [TEST SCORE: 1.741]

**#28: area1* + area2 + texture3* + perimeter3 + compactness3 + concavity3 + concave_points3**

Optimized Thresholds: area1*: 1486.4962 | area2: 56.0757 | texture3*: 41.4388 | perimeter3: 116.0215 | compactness3: 0.7273 | concavity3: 0.9904 | concave_points3: 0.1597

Train Sens: 0.901 | Train Spec: 0.962 [TRAIN SCORE: 1.863] || Val Sens: 0.875 | Val Spec: 0.973 [VAL SCORE: 1.848] || Test Sens: 0.907 | Test Spec: 0.976 [TEST SCORE: 1.883]

**#29: texture1 + area2 + smoothness3 + compactness3 + concavity3 + fractal_dimension3**

Optimized Thresholds: texture1: 37.5311 | area2: 43.0989 | smoothness3: 0.2107 | compactness3: 0.3723 | concavity3: 0.3499 | fractal_dimension3: 0.1205

Train Sens: 0.934 | Train Spec: 0.917 [TRAIN SCORE: 1.851] || Val Sens: 0.875 | Val Spec: 0.973 [VAL SCORE: 1.848] || Test Sens: 0.884 | Test Spec: 0.857 [TEST SCORE: 1.741]

**#30: area2 + texture3 + smoothness3 + concavity3 + symmetry3**

Optimized Thresholds: area2: 42.9678 | texture3: 46.4305 | smoothness3: 0.2109 | concavity3: 0.3489 | symmetry3: 0.4303

Train Sens: 0.934 | Train Spec: 0.917 [TRAIN SCORE: 1.851] || Val Sens: 0.875 | Val Spec: 0.973 [VAL SCORE: 1.848] || Test Sens: 0.884 | Test Spec: 0.881 [TEST SCORE: 1.765]

**#31: area2 + texture3 + smoothness3 + concavity3**

Optimized Thresholds: area2: 42.9983 | texture3: 46.4295 | smoothness3: 0.2026 | concavity3: 0.3494

Train Sens: 0.934 | Train Spec: 0.917 [TRAIN SCORE: 1.851] || Val Sens: 0.875 | Val Spec: 0.973 [VAL SCORE: 1.848] || Test Sens: 0.860 | Test Spec: 0.881 [TEST SCORE: 1.741]

**#32: texture1 + area2 + smoothness3 + concavity3**

Optimized Thresholds: texture1: 37.2234 | area2: 43.0204 | smoothness3: 0.1891 | concavity3: 0.3489

Train Sens: 0.934 | Train Spec: 0.917 [TRAIN SCORE: 1.851] || Val Sens: 0.875 | Val Spec: 0.973 [VAL SCORE: 1.848] || Test Sens: 0.860 | Test Spec: 0.881 [TEST SCORE: 1.741]

**#33: area1 + area2 + texture3 + perimeter3 + smoothness3 + concavity3 + concave_points3 + fractal_dimension3**

Optimized Thresholds: area1: 927.5709 | area2: 48.9277 | texture3: 33.0006 | perimeter3: 115.4127 | smoothness3: 0.1932 | concavity3: 1.0775 | concave_points3: 0.1790 | fractal_dimension3: 0.1373

Train Sens: 0.950 | Train Spec: 0.962 [TRAIN SCORE: 1.913] || Val Sens: 0.896 | Val Spec: 0.946 [VAL SCORE: 1.842] || Test Sens: 0.907 | Test Spec: 0.881 [TEST SCORE: 1.788]

##### #34: area1 + area2 + texture3 + perimeter3 + compactness3 + concavity3 + concave_points3 + fractal_dimension3

Optimized Thresholds: area1: 927.5709 | area2: 48.9277 | texture3: 33.0006 | perimeter3: 115.4127 | compactness3: 0.7569 | concavity3: 1.0775 | concave_points3: 0.1790 | fractal_dimension3: 0.1373

Train Sens: 0.950 | Train Spec: 0.962 [TRAIN SCORE: 1.913] || Val Sens: 0.896 | Val Spec: 0.946 [VAL SCORE: 1.842] || Test Sens: 0.907 | Test Spec: 0.881 [TEST SCORE: 1.788]

##### #35: area1 + area2 + texture3 + perimeter3 + concavity3 + concave_points3

Optimized Thresholds: area1: 1815.7478 | area2: 50.7333 | texture3: 32.5159 | perimeter3: 114.6056 | concavity3: 1.0931 | concave_points3: 0.1610

Train Sens: 0.959 | Train Spec: 0.947 [TRAIN SCORE: 1.906] || Val Sens: 0.896 | Val Spec: 0.946 [VAL SCORE: 1.842] || Test Sens: 0.930 | Test Spec: 0.929 [TEST SCORE: 1.859]

##### #36: area2 + texture3 + perimeter3 + concave_points3 + symmetry3 + fractal_dimension3

Optimized Thresholds: area2: 56.1275 | texture3: 33.5549 | perimeter3: 114.8468 | concave_points3: 0.1850 | symmetry3: 0.4167 | fractal_dimension3: 0.1333

Train Sens: 0.901 | Train Spec: 0.962 [TRAIN SCORE: 1.863] || Val Sens: 0.896 | Val Spec: 0.946 [VAL SCORE: 1.842] || Test Sens: 0.907 | Test Spec: 0.929 [TEST SCORE: 1.836]

##### #37: area1 + area2 + texture3 + perimeter3 + compactness3 + concavity3 + concave_points3 + symmetry3

Optimized Thresholds: area1: 927.5709 | area2: 48.9277 | texture3: 33.0006 | perimeter3: 115.4127 | compactness3: 0.7569 | concavity3: 1.0775 | concave_points3: 0.1790 | symmetry3: 0.5234

Train Sens: 0.950 | Train Spec: 0.962 [TRAIN SCORE: 1.913] || Val Sens: 0.896 | Val Spec: 0.946 [VAL SCORE: 1.842] || Test Sens: 0.907 | Test Spec: 0.905 [TEST SCORE: 1.812]

##### #38: area2 + texture3 + concave_points3 + symmetry3 + fractal_dimension3

Optimized Thresholds: area2: 42.9858 | texture3: 33.8003 | concave_points3: 0.1506 | symmetry3: 0.3695 | fractal_dimension3: 0.1411

Train Sens: 0.942 | Train Spec: 0.932 [TRAIN SCORE: 1.874] || Val Sens: 0.896 | Val Spec: 0.946 [VAL SCORE: 1.842] || Test Sens: 0.953 | Test Spec: 0.857 [TEST SCORE: 1.811]

##### #39: area2 + texture3 + smoothness3 + compactness3 + concavity3 + fractal_dimension3

Optimized Thresholds: area2: 39.9715 | texture3: 47.7861 | smoothness3: 0.2054 | compactness3: 0.3386 | concavity3: 0.3842 | fractal_dimension3: 0.1253

Train Sens: 0.934 | Train Spec: 0.917 [TRAIN SCORE: 1.851] || Val Sens: 0.896 | Val Spec: 0.946 [VAL SCORE: 1.842] || Test Sens: 0.884 | Test Spec: 0.857 [TEST SCORE: 1.741]

##### #40: area1 + area2 + texture3 + perimeter3 + smoothness3 + compactness3 + concave_points3 + fractal_dimension3

Optimized Thresholds: area1: 927.5709 | area2: 48.9277 | texture3: 33.0006 | perimeter3: 115.4127 | smoothness3: 0.1932 | compactness3: 0.8065 | concave_points3: 0.1790 | fractal_dimension3: 0.1373

Train Sens: 0.950 | Train Spec: 0.970 [TRAIN SCORE: 1.920] || Val Sens: 0.896 | Val Spec: 0.946 [VAL SCORE: 1.842] || Test Sens: 0.907 | Test Spec: 0.881 [TEST SCORE: 1.788]

##### #41: concave_points1 + area2 + texture3 + perimeter3 + concavity3 + concave_points3

Optimized Thresholds: concave_points1: 0.1428 | area2: 50.7333 | texture3: 32.5159 | perimeter3: 114.6056 | concavity3: 1.0931 | concave_points3: 0.1610

Train Sens: 0.959 | Train Spec: 0.947 [TRAIN SCORE: 1.906] || Val Sens: 0.896 | Val Spec: 0.946 [VAL SCORE: 1.842] || Test Sens: 0.930 | Test Spec: 0.929 [TEST SCORE: 1.859]

##### #42: area1 + area2 + texture3 + perimeter3 + smoothness3 + concavity3 + concave_points3 + symmetry3

Optimized Thresholds: area1: 927.5709 | area2: 48.9277 | texture3: 33.0006 | perimeter3: 115.4127 | smoothness3: 0.1932 | concavity3: 1.0775 | concave_points3: 0.1790 | symmetry3: 0.5234

Train Sens: 0.950 | Train Spec: 0.962 [TRAIN SCORE: 1.913] || Val Sens: 0.896 | Val Spec: 0.946 [VAL SCORE: 1.842] || Test Sens: 0.907 | Test Spec: 0.905 [TEST SCORE: 1.812]

##### #43: area1 + concave_points1* + area2 + texture3 + perimeter3 + compactness3 + concavity3 + concave_points3 + fract[illegible]

Optimized Thresholds: area1: 1930.4370 | concave_points1*: 0.1419 | area2: 69.5356 | texture3: 33.4430 | perimeter3: 115.9055 | compactness3: 0.8324 | concavity3: 1.0445 | concave_points3: 0.1627 | fractal_dimension3: 0.1405

Train Sens: 0.917 | Train Spec: 0.955 [TRAIN SCORE: 1.872] || Val Sens: 0.896 | Val Spec: 0.946 [VAL SCORE: 1.842] || Test Sens: 0.907 | Test Spec: 0.929 [TEST SCORE: 1.836]

##### #44: area1 + concave_points1 + area2 + texture3 + perimeter3 + concave_points3

Optimized Thresholds: area1: 1360.5648 | concave_points1: 0.0787 | area2: 49.5532 | texture3: 32.6158 | perimeter3: 115.0704 | concave_points3: 0.1776

Train Sens: 0.950 | Train Spec: 0.962 [TRAIN SCORE: 1.913] || Val Sens: 0.896 | Val Spec: 0.946 [VAL SCORE: 1.842] || Test Sens: 0.930 | Test Spec: 0.881 [TEST SCORE: 1.811]

##### #45: area1 + concave_points1* + area2* + texture3 + perimeter3 + concave_points3 + symmetry3*

Optimized Thresholds: area1: 1248.8565 | concave_points1*: 0.1288 | area2*: 268.4604 | texture3: 33.2269 | perimeter3: 114.6026 | concave_points3: 0.1763 | symmetry3*: 0.4451

Train Sens: 0.917 | Train Spec: 0.970 [TRAIN SCORE: 1.887] || Val Sens: 0.896 | Val Spec: 0.946 [VAL SCORE: 1.842] || Test Sens: 0.907 | Test Spec: 0.976 [TEST SCORE: 1.883]

##### #46: area1 + area2 + texture3 + perimeter3 + smoothness3 + compactness3 + concave_points3 + symmetry3

Optimized Thresholds: area1: 927.5709 | area2: 48.9277 | texture3: 33.0006 | perimeter3: 115.4127 | smoothness3: 0.1932 | compactness3: 0.8065 | concave_points3: 0.1790 | symmetry3: 0.5234

Train Sens: 0.950 | Train Spec: 0.970 [TRAIN SCORE: 1.920] || Val Sens: 0.896 | Val Spec: 0.946 [VAL SCORE: 1.842] || Test Sens: 0.907 | Test Spec: 0.905 [TEST SCORE: 1.812]

##### #47: area2 + texture3* + compactness3

Optimized Thresholds: area2: 37.1246 | texture3*: 42.3385 | compactness3: 0.3756

Train Sens: 0.884 | Train Spec: 0.925 [TRAIN SCORE: 1.809] || Val Sens: 0.917 | Val Spec: 0.919 [VAL SCORE: 1.836] || Test Sens: 0.884 | Test Spec: 0.857 [TEST SCORE: 1.741]

##### #48: area2 + texture3 + smoothness3 + compactness3 + symmetry3 + fractal_dimension3

Optimized Thresholds: area2: 34.9496 | texture3: 46.6170 | smoothness3: 0.1762 | compactness3: 0.3759 | symmetry3: 0.4221 | fractal_dimension3: 0.1220

Train Sens: 0.926 | Train Spec: 0.917 [TRAIN SCORE: 1.843] || Val Sens: 0.917 | Val Spec: 0.919 [VAL SCORE: 1.836] || Test Sens: 0.930 | Test Spec: 0.857 [TEST SCORE: 1.787]

##### #49: area2 + texture3* + compactness3 + symmetry3* + fractal_dimension3

Optimized Thresholds: area2: 36.8945 | texture3*: 43.3642 | compactness3: 0.3816 | symmetry3*: 0.4643 | fractal_dimension3:

0.1271

Train Sens: 0.884 | Train Spec: 0.925 [TRAIN SCORE: 1.809] || Val Sens: 0.917 | Val Spec: 0.919 [VAL SCORE: 1.836] || Test Sens: 0.907 | Test Spec: 0.857 [TEST SCORE: 1.764]

##### #50: texture1 + area2 + texture3 + smoothness3 + compactness3 + concavity3 + fractal_dimension3

Optimized Thresholds: texture1: 35.3205 | area2: 35.0158 | texture3: 45.7119 | smoothness3: 0.1824 | compactness3: 0.3755 | concavity3: 1.0165 | fractal_dimension3: 0.1429

Train Sens: 0.926 | Train Spec: 0.917 [TRAIN SCORE: 1.843] || Val Sens: 0.917 | Val Spec: 0.919 [VAL SCORE: 1.836] || Test Sens: 0.907 | Test Spec: 0.857 [TEST SCORE: 1.764]

##### #51: texture1* + area2 + compactness3

Optimized Thresholds: texture1*: 33.4513 | area2: 37.5473 | compactness3: 0.3762

Train Sens: 0.884 | Train Spec: 0.917 [TRAIN SCORE: 1.802] || Val Sens: 0.917 | Val Spec: 0.919 [VAL SCORE: 1.836] || Test Sens: 0.884 | Test Spec: 0.857 [TEST SCORE: 1.741]

##### #52: area2 + texture3* + compactness3 + symmetry3

Optimized Thresholds: area2: 37.1004 | texture3*: 42.0963 | compactness3: 0.3764 | symmetry3: 0.4455

Train Sens: 0.884 | Train Spec: 0.925 [TRAIN SCORE: 1.809] || Val Sens: 0.917 | Val Spec: 0.919 [VAL SCORE: 1.836] || Test Sens: 0.907 | Test Spec: 0.857 [TEST SCORE: 1.764]

##### #53: texture1* + area2 + smoothness3 + compactness3 + fractal_dimension3

Optimized Thresholds: texture1*: 35.7465 | area2: 38.0254 | smoothness3: 0.1875 | compactness3: 0.3813 | fractal_dimension3: 0.1288

Train Sens: 0.893 | Train Spec: 0.932 [TRAIN SCORE: 1.825] || Val Sens: 0.917 | Val Spec: 0.919 [VAL SCORE: 1.836] || Test Sens: 0.884 | Test Spec: 0.857 [TEST SCORE: 1.741]

##### #54: texture1 + area2 + texture3* + compactness3 + fractal_dimension3

Optimized Thresholds: texture1: 35.5318 | area2: 37.1649 | texture3*: 39.4373 | compactness3: 0.3800 | fractal_dimension3: 0.1306

Train Sens: 0.884 | Train Spec: 0.917 [TRAIN SCORE: 1.802] || Val Sens: 0.917 | Val Spec: 0.919 [VAL SCORE: 1.836] || Test Sens: 0.884 | Test Spec: 0.833 [TEST SCORE: 1.717]

##### #55: area1 + concave_points1 + texture3 + perimeter3 + smoothness3 + compactness3 + concavity3 + concave_points3 [illegible]

Optimized Thresholds: area1: 2165.3246 | concave_points1: 0.1398 | texture3: 45.8236 | perimeter3: 109.6557 | smoothness3: 0.1755 | compactness3: 0.6575 | concavity3: 1.1478 | concave_points3: 0.1799 | fractal_dimension3: 0.1375

Train Sens: 0.942 | Train Spec: 0.955 [TRAIN SCORE: 1.897] || Val Sens: 0.938 | Val Spec: 0.892 [VAL SCORE: 1.829] || Test Sens: 0.930 | Test Spec: 0.976 [TEST SCORE: 1.906]

##### #56: texture3 + perimeter3 + compactness3 + concavity3 + concave_points3 + fractal_dimension3

Optimized Thresholds: texture3: 38.4596 | perimeter3: 111.7221 | compactness3: 0.6954 | concavity3: 0.9749 | concave_points3: 0.1627 | fractal_dimension3: 0.1364

Train Sens: 0.917 | Train Spec: 0.955 [TRAIN SCORE: 1.872] || Val Sens: 0.938 | Val Spec: 0.892 [VAL SCORE: 1.829] || Test Sens: 0.907 | Test Spec: 0.952 [TEST SCORE: 1.859]

##### #57: area2 + texture3 + perimeter3 + compactness3 + concavity3 + concave_points3 + fractal_dimension3

Optimized Thresholds: area2: 53.9321 | texture3: 46.4145 | perimeter3: 109.7447 | compactness3: 0.7138 | concavity3: 1.0396 | concave_points3: 0.1603 | fractal_dimension3: 0.1403

Train Sens: 0.950 | Train Spec: 0.947 [TRAIN SCORE: 1.898] || Val Sens: 0.938 | Val Spec: 0.892 [VAL SCORE: 1.829] || Test Sens: 0.907 | Test Spec: 0.952 [TEST SCORE: 1.859]

**#58: texture1 + area1 + concave_points1 + area2 + perimeter3 + smoothness3 + concave_points3 + symmetry3 + fractal**

Optimized Thresholds: texture1: 37.2493 | area1: 1589.5371 | concave_points1: 0.1551 | area2: 64.1871 | perimeter3: 109.8031 | smoothness3: 0.1740 | concave_points3: 0.2142 | symmetry3: 0.4527 | fractal_dimension3: 0.1356

Train Sens: 0.950 | Train Spec: 0.962 [TRAIN SCORE: 1.913] || Val Sens: 0.938 | Val Spec: 0.892 [VAL SCORE: 1.829] || Test Sens: 0.907 | Test Spec: 0.952 [TEST SCORE: 1.859]

**#59: texture1 + area2 + perimeter3 + smoothness3 + concavity3 + concave_points3 + symmetry3 + fractal_dimension3**

Optimized Thresholds: texture1: 37.0747 | area2: 97.8350 | perimeter3: 110.0622 | smoothness3: 0.1761 | concavity3: 1.0082 | concave_points3: 0.2251 | symmetry3: 0.4958 | fractal_dimension3: 0.1368

Train Sens: 0.950 | Train Spec: 0.955 [TRAIN SCORE: 1.905] || Val Sens: 0.938 | Val Spec: 0.892 [VAL SCORE: 1.829] || Test Sens: 0.907 | Test Spec: 0.976 [TEST SCORE: 1.883]

**#60: texture1 + area1 + concave_points1 + area2 + perimeter3 + smoothness3 + concave_points3 + fractal_dimension3**

Optimized Thresholds: texture1: 38.3989 | area1: 751.2675 | concave_points1: 0.1856 | area2: 85.8681 | perimeter3: 109.5069 | smoothness3: 0.1763 | concave_points3: 0.1776 | fractal_dimension3: 0.1334

Train Sens: 0.950 | Train Spec: 0.962 [TRAIN SCORE: 1.913] || Val Sens: 0.938 | Val Spec: 0.892 [VAL SCORE: 1.829] || Test Sens: 0.930 | Test Spec: 0.976 [TEST SCORE: 1.906]

**#61: area1* + texture3* + perimeter3 + smoothness3 + compactness3 + concavity3 + concave_points3 + symmetry3 + fr**

Optimized Thresholds: area1*: 1507.4262 | texture3*: 39.0670 | perimeter3: 111.5332 | smoothness3: 0.1770 | compactness3: 0.6965 | concavity3: 1.1617 | concave_points3: 0.2212 | symmetry3: 0.5323 | fractal_dimension3: 0.1348

Train Sens: 0.917 | Train Spec: 0.962 [TRAIN SCORE: 1.880] || Val Sens: 0.938 | Val Spec: 0.892 [VAL SCORE: 1.829] || Test Sens: 0.907 | Test Spec: 0.952 [TEST SCORE: 1.859]

**#62: area1* + area2 + texture3 + perimeter3 + smoothness3 + concave_points3 + symmetry3**

Optimized Thresholds: area1*: 1554.2994 | area2: 97.7352 | texture3: 45.9524 | perimeter3: 109.6929 | smoothness3: 0.1747 | concave_points3: 0.2023 | symmetry3: 0.4661

Train Sens: 0.950 | Train Spec: 0.962 [TRAIN SCORE: 1.913] || Val Sens: 0.938 | Val Spec: 0.892 [VAL SCORE: 1.829] || Test Sens: 0.930 | Test Spec: 1.000 [TEST SCORE: 1.930]

**#63: area2 + texture3* + perimeter3 + compactness3 + concavity3 + concave_points3 + symmetry3***

Optimized Thresholds: area2: 108.3729 | texture3*: 39.2576 | perimeter3: 111.6134 | compactness3: 0.7553 | concavity3: 0.9694 | concave_points3: 0.1665 | symmetry3*: 0.4279

Train Sens: 0.909 | Train Spec: 0.955 [TRAIN SCORE: 1.864] || Val Sens: 0.938 | Val Spec: 0.892 [VAL SCORE: 1.829] || Test Sens: 0.907 | Test Spec: 0.976 [TEST SCORE: 1.883]

**#64: area1 + concave_points1 + area2 + texture3 + perimeter3 + smoothness3 + concavity3 + concave_points3**

Optimized Thresholds: area1: 1981.8632 | concave_points1: 0.1212 | area2: 79.9502 | texture3: 48.2566 | perimeter3: 109.9155 | smoothness3: 0.1783 | concavity3: 0.8391 | concave_points3: 0.1740

Train Sens: 0.950 | Train Spec: 0.955 [TRAIN SCORE: 1.905] || Val Sens: 0.938 | Val Spec: 0.892 [VAL SCORE: 1.829] || Test Sens: 0.930 | Test Spec: 1.000 [TEST SCORE: 1.930]

**#65: area1 + area2 + perimeter3 + smoothness3 + concave_points3**

Optimized Thresholds: area1: 1765.9210 | area2: 59.9000 | perimeter3: 110.3588 | smoothness3: 0.1763 | concave_points3: 0.1764

Train Sens: 0.934 | Train Spec: 0.962 [TRAIN SCORE: 1.896] || Val Sens: 0.938 | Val Spec: 0.892 [VAL SCORE: 1.829] || Test Sens: 0.930 | Test Spec: 0.976 [TEST SCORE: 1.906]

##### #66: concave_points1 + area2 + texture3 + perimeter3 + smoothness3 + concavity3 + concave_points3 + symmetry3 + f

Optimized Thresholds: concave_points1: 0.1050 | area2: 62.0077 | texture3: 46.4319 | perimeter3: 109.3554 | smoothness3: 0.1763 | concavity3: 0.8383 | concave_points3: 0.2073 | symmetry3: 0.3850 | fractal_dimension3: 0.1333

Train Sens: 0.950 | Train Spec: 0.947 [TRAIN SCORE: 1.898] || Val Sens: 0.938 | Val Spec: 0.892 [VAL SCORE: 1.829] || Test Sens: 0.953 | Test Spec: 0.929 [TEST SCORE: 1.882]

##### #67: concave_points1* + texture3* + perimeter3 + concavity3 + concave_points3 + fractal_dimension3

Optimized Thresholds: concave_points1*: 0.1373 | texture3*: 38.7315 | perimeter3: 111.1247 | concavity3: 1.0868 | concave_points3: 0.1719 | fractal_dimension3: 0.1386

Train Sens: 0.909 | Train Spec: 0.955 [TRAIN SCORE: 1.864] || Val Sens: 0.938 | Val Spec: 0.892 [VAL SCORE: 1.829] || Test Sens: 0.907 | Test Spec: 0.952 [TEST SCORE: 1.859]

##### #68: concave_points1 + area2 + texture3 + perimeter3 + smoothness3 + concave_points3 + symmetry3 + fractal_dimen

Optimized Thresholds: concave_points1: 0.1582 | area2: 65.9362 | texture3: 47.1454 | perimeter3: 109.7138 | smoothness3: 0.1896 | concave_points3: 0.1977 | symmetry3: 0.4766 | fractal_dimension3: 0.1202

Train Sens: 0.950 | Train Spec: 0.955 [TRAIN SCORE: 1.905] || Val Sens: 0.938 | Val Spec: 0.892 [VAL SCORE: 1.829] || Test Sens: 0.907 | Test Spec: 0.929 [TEST SCORE: 1.836]

##### #69: concave_points1* + perimeter3 + smoothness3 + concave_points3

Optimized Thresholds: concave_points1*: 0.1291 | perimeter3: 109.7365 | smoothness3: 0.1781 | concave_points3: 0.1633

Train Sens: 0.950 | Train Spec: 0.947 [TRAIN SCORE: 1.898] || Val Sens: 0.938 | Val Spec: 0.892 [VAL SCORE: 1.829] || Test Sens: 0.930 | Test Spec: 1.000 [TEST SCORE: 1.930]

##### #70: texture3* + perimeter3 + smoothness3 + concave_points3 + symmetry3 + fractal_dimension3

Optimized Thresholds: texture3*: 37.3141 | perimeter3: 111.3256 | smoothness3: 0.1855 | concave_points3: 0.1947 | symmetry3: 0.4714 | fractal_dimension3: 0.1352

Train Sens: 0.909 | Train Spec: 0.962 [TRAIN SCORE: 1.871] || Val Sens: 0.938 | Val Spec: 0.892 [VAL SCORE: 1.829] || Test Sens: 0.907 | Test Spec: 0.952 [TEST SCORE: 1.859]

##### #71: perimeter3 + compactness3 + concavity3

Optimized Thresholds: perimeter3: 110.6666 | compactness3: 0.3984 | concavity3: 0.9857

Train Sens: 0.917 | Train Spec: 0.932 [TRAIN SCORE: 1.850] || Val Sens: 0.938 | Val Spec: 0.892 [VAL SCORE: 1.829] || Test Sens: 0.907 | Test Spec: 0.952 [TEST SCORE: 1.859]

##### #72: area1 + concave_points1 + area2 + texture3 + perimeter3 + smoothness3 + concave_points3 + symmetry3 + fractal

Optimized Thresholds: area1: 2131.2570 | concave_points1: 0.1099 | area2: 72.8558 | texture3: 45.9661 | perimeter3: 109.8557 | smoothness3: 0.1743 | concave_points3: 0.2080 | symmetry3: 0.5450 | fractal_dimension3: 0.1416

Train Sens: 0.950 | Train Spec: 0.962 [TRAIN SCORE: 1.913] || Val Sens: 0.938 | Val Spec: 0.892 [VAL SCORE: 1.829] || Test Sens: 0.907 | Test Spec: 0.952 [TEST SCORE: 1.859]

##### #73: texture1 + area1 + area2 + perimeter3 + smoothness3 + concavity3 + concave_points3 + symmetry3 + fractal_dime

Optimized Thresholds: texture1: 37.0182 | area1: 1011.2080 | area2: 58.3681 | perimeter3: 109.7858 | smoothness3: 0.1784 |

concavity3: 1.0471 | concave_points3: 0.1639 | symmetry3: 0.4378 | fractal_dimension3: 0.1356

Train Sens: 0.959 | Train Spec: 0.947 [TRAIN SCORE: 1.906] || Val Sens: 0.938 | Val Spec: 0.892 [VAL SCORE: 1.829] || Test Sens: 0.930 | Test Spec: 0.952 [TEST SCORE: 1.883]

##### #74: area2 + texture3 + perimeter3 + concavity3 + concave_points3 + fractal_dimension3

Optimized Thresholds: area2: 53.2799 | texture3: 33.7876 | perimeter3: 113.0969 | concavity3: 0.8908 | concave_points3: 0.1824 | fractal_dimension3: 0.1364

Train Sens: 0.926 | Train Spec: 0.947 [TRAIN SCORE: 1.873] || Val Sens: 0.938 | Val Spec: 0.892 [VAL SCORE: 1.829] || Test Sens: 0.907 | Test Spec: 0.905 [TEST SCORE: 1.812]

##### #75: area1* + concave_points1 + area2 + perimeter3 + smoothness3 + concave_points3 + symmetry3 + fractal_dimensi[illegible]

Optimized Thresholds: area1*: 1424.6228 | concave_points1: 0.1043 | area2: 60.5049 | perimeter3: 109.7577 | smoothness3: 0.1776 | concave_points3: 0.1873 | symmetry3: 0.4846 | fractal_dimension3: 0.1381

Train Sens: 0.950 | Train Spec: 0.962 [TRAIN SCORE: 1.913] || Val Sens: 0.938 | Val Spec: 0.892 [VAL SCORE: 1.829] || Test Sens: 0.930 | Test Spec: 0.952 [TEST SCORE: 1.883]

##### #76: area1 + concave_points1 + area2 + perimeter3 + smoothness3 + concave_points3

Optimized Thresholds: area1: 1343.2136 | concave_points1: 0.1302 | area2: 79.8334 | perimeter3: 109.6926 | smoothness3: 0.1762 | concave_points3: 0.1768

Train Sens: 0.950 | Train Spec: 0.962 [TRAIN SCORE: 1.913] || Val Sens: 0.938 | Val Spec: 0.892 [VAL SCORE: 1.829] || Test Sens: 0.930 | Test Spec: 1.000 [TEST SCORE: 1.930]

##### #77: area1 + perimeter3 + compactness3 + concavity3

Optimized Thresholds: area1: 696.9721 | perimeter3: 112.0688 | compactness3: 0.3852 | concavity3: 1.0110

Train Sens: 0.934 | Train Spec: 0.932 [TRAIN SCORE: 1.866] || Val Sens: 0.938 | Val Spec: 0.892 [VAL SCORE: 1.829] || Test Sens: 0.953 | Test Spec: 0.952 [TEST SCORE: 1.906]

##### #78: texture1 + concave_points1 + area2 + texture3 + perimeter3 + smoothness3 + concave_points3 + symmetry3 + frac[illegible]

Optimized Thresholds: texture1: 34.8477 | concave_points1: 0.1099 | area2: 72.8558 | texture3: 45.9661 | perimeter3: 109.8557 | smoothness3: 0.1743 | concave_points3: 0.2080 | symmetry3: 0.5450 | fractal_dimension3: 0.1416

Train Sens: 0.950 | Train Spec: 0.962 [TRAIN SCORE: 1.913] || Val Sens: 0.938 | Val Spec: 0.892 [VAL SCORE: 1.829] || Test Sens: 0.907 | Test Spec: 0.952 [TEST SCORE: 1.859]

##### #79: area2 + perimeter3 + smoothness3 + concave_points3 + symmetry3

Optimized Thresholds: area2: 73.6228 | perimeter3: 109.7867 | smoothness3: 0.1757 | concave_points3: 0.1898 | symmetry3: 0.4492

Train Sens: 0.950 | Train Spec: 0.962 [TRAIN SCORE: 1.913] || Val Sens: 0.938 | Val Spec: 0.892 [VAL SCORE: 1.829] || Test Sens: 0.930 | Test Spec: 0.976 [TEST SCORE: 1.906]

##### #80: area2 + compactness3 + fractal_dimension3

Optimized Thresholds: area2: 38.1989 | compactness3: 0.3592 | fractal_dimension3: 0.1341

Train Sens: 0.884 | Train Spec: 0.917 [TRAIN SCORE: 1.802] || Val Sens: 0.938 | Val Spec: 0.892 [VAL SCORE: 1.829] || Test Sens: 0.884 | Test Spec: 0.857 [TEST SCORE: 1.741]

##### #81: concave_points1* + area2 + texture3 + perimeter3 + smoothness3 + concave_points3 + symmetry3

Optimized Thresholds: concave_points1*: 0.1205 | area2: 97.7352 | texture3: 45.9524 | perimeter3: 109.6929 | smoothness3: 0.1747 | concave_points3: 0.2023 | symmetry3: 0.4661

Train Sens: 0.950 | Train Spec: 0.962 [TRAIN SCORE: 1.913] || Val Sens: 0.938 | Val Spec: 0.892 [VAL SCORE: 1.829] || Test Sens: 0.930 | Test Spec: 1.000 [TEST SCORE: 1.930]

**#82: concave_points1* + texture3* + perimeter3 + smoothness3 + compactness3 + concave_points3 + symmetry3**

Optimized Thresholds: concave_points1*: 0.1360 | texture3*: 38.0489 | perimeter3: 110.8592 | smoothness3: 0.1841 | compactness3: 0.7432 | concave_points3: 0.1910 | symmetry3: 0.4546

Train Sens: 0.909 | Train Spec: 0.962 [TRAIN SCORE: 1.871] || Val Sens: 0.938 | Val Spec: 0.892 [VAL SCORE: 1.829] || Test Sens: 0.907 | Test Spec: 0.976 [TEST SCORE: 1.883]

**#83: texture1 + area1 + area2 + texture3 + perimeter3 + smoothness3 + concave_points3 + symmetry3 + fractal_dimens[illegible]**

Optimized Thresholds: texture1: 34.8477 | area1: 1429.6346 | area2: 72.8558 | texture3: 45.9661 | perimeter3: 109.8557 | smoothness3: 0.1743 | concave_points3: 0.2080 | symmetry3: 0.5450 | fractal_dimension3: 0.1416

Train Sens: 0.950 | Train Spec: 0.962 [TRAIN SCORE: 1.913] || Val Sens: 0.938 | Val Spec: 0.892 [VAL SCORE: 1.829] || Test Sens: 0.907 | Test Spec: 0.952 [TEST SCORE: 1.859]

**#84: concave_points1 + area2 + perimeter3 + smoothness3 + concave_points3 + symmetry3**

Optimized Thresholds: concave_points1: 0.1811 | area2: 43.9711 | perimeter3: 109.9201 | smoothness3: 0.1768 | concave_points3: 0.1968 | symmetry3: 0.4900

Train Sens: 0.959 | Train Spec: 0.955 [TRAIN SCORE: 1.914] || Val Sens: 0.938 | Val Spec: 0.892 [VAL SCORE: 1.829] || Test Sens: 0.930 | Test Spec: 0.929 [TEST SCORE: 1.859]

**#85: area1* + texture3* + perimeter3 + smoothness3 + compactness3 + concavity3 + concave_points3**

Optimized Thresholds: area1*: 1598.6598 | texture3*: 39.1911 | perimeter3: 110.6734 | smoothness3: 0.2040 | compactness3: 0.7365 | concavity3: 1.0511 | concave_points3: 0.1681

Train Sens: 0.909 | Train Spec: 0.947 [TRAIN SCORE: 1.856] || Val Sens: 0.938 | Val Spec: 0.892 [VAL SCORE: 1.829] || Test Sens: 0.907 | Test Spec: 0.976 [TEST SCORE: 1.883]

**#86: area2 + texture3 + perimeter3 + smoothness3 + concave_points3 + symmetry3**

Optimized Thresholds: area2: 73.3764 | texture3: 47.3313 | perimeter3: 109.8893 | smoothness3: 0.1767 | concave_points3: 0.2016 | symmetry3: 0.4545

Train Sens: 0.950 | Train Spec: 0.962 [TRAIN SCORE: 1.913] || Val Sens: 0.938 | Val Spec: 0.892 [VAL SCORE: 1.829] || Test Sens: 0.930 | Test Spec: 0.976 [TEST SCORE: 1.906]

**#87: texture3 + perimeter3 + smoothness3 + compactness3 + concavity3 + concave_points3**

Optimized Thresholds: texture3: 41.9318 | perimeter3: 110.0186 | smoothness3: 0.1839 | compactness3: 0.7133 | concavity3: 0.8263 | concave_points3: 0.1602

Train Sens: 0.950 | Train Spec: 0.947 [TRAIN SCORE: 1.898] || Val Sens: 0.938 | Val Spec: 0.892 [VAL SCORE: 1.829] || Test Sens: 0.907 | Test Spec: 1.000 [TEST SCORE: 1.907]

**#88: texture1 + area2 + perimeter3 + smoothness3 + compactness3 + concave_points3 + symmetry3 + fractal_dimensic[illegible]**

Optimized Thresholds: texture1: 37.0747 | area2: 97.8350 | perimeter3: 110.0622 | smoothness3: 0.1761 | compactness3: 0.7564 | concave_points3: 0.2251 | symmetry3: 0.4958 | fractal_dimension3: 0.1368

Train Sens: 0.950 | Train Spec: 0.962 [TRAIN SCORE: 1.913] || Val Sens: 0.938 | Val Spec: 0.892 [VAL SCORE: 1.829] || Test Sens: 0.907 | Test Spec: 0.976 [TEST SCORE: 1.883]

**#89: area1 + texture3 + perimeter3 + smoothness3 + compactness3 + concave_points3 + symmetry3**

Optimized Thresholds: area1: 694.9432 | texture3: 41.8257 | perimeter3: 109.7751 | smoothness3: 0.1764 | compactness3: 0.7763 | concave_points3: 0.2094 | symmetry3: 0.5261

Train Sens: 0.950 | Train Spec: 0.955 [TRAIN SCORE: 1.905] || Val Sens: 0.938 | Val Spec: 0.892 [VAL SCORE: 1.829] || Test Sens: 0.930 | Test Spec: 1.000 [TEST SCORE: 1.930]

**#90: area1 + concave_points1 + perimeter3 + smoothness3 + compactness3 + concave_points3 + symmetry3**

Optimized Thresholds: area1: 694.9432 | concave_points1: 0.1557 | perimeter3: 109.7751 | smoothness3: 0.1764 | compactness3: 0.7763 | concave_points3: 0.2094 | symmetry3: 0.5261

Train Sens: 0.950 | Train Spec: 0.955 [TRAIN SCORE: 1.905] || Val Sens: 0.938 | Val Spec: 0.892 [VAL SCORE: 1.829] || Test Sens: 0.930 | Test Spec: 1.000 [TEST SCORE: 1.930]

**#91: concave_points1 + area2 + perimeter3 + smoothness3 + concave_points3**

Optimized Thresholds: concave_points1: 0.1386 | area2: 59.9000 | perimeter3: 110.3588 | smoothness3: 0.1763 | concave_points3: 0.1764

Train Sens: 0.934 | Train Spec: 0.962 [TRAIN SCORE: 1.896] || Val Sens: 0.938 | Val Spec: 0.892 [VAL SCORE: 1.829] || Test Sens: 0.930 | Test Spec: 0.976 [TEST SCORE: 1.906]

**#92: area1* + area2 + perimeter3 + smoothness3 + concave_points3 + symmetry3**

Optimized Thresholds: area1*: 1777.7068 | area2: 44.7439 | perimeter3: 109.9672 | smoothness3: 0.1756 | concave_points3: 0.1966 | symmetry3: 0.4756

Train Sens: 0.959 | Train Spec: 0.955 [TRAIN SCORE: 1.914] || Val Sens: 0.938 | Val Spec: 0.892 [VAL SCORE: 1.829] || Test Sens: 0.930 | Test Spec: 0.929 [TEST SCORE: 1.859]

**#93: area1 + concave_points1* + texture3 + perimeter3 + smoothness3 + concavity3 + concave_points3**

Optimized Thresholds: area1: 1481.5231 | concave_points1*: 0.1415 | texture3: 38.5800 | perimeter3: 111.3112 | smoothness3: 0.1803 | concavity3: 0.9050 | concave_points3: 0.1774

Train Sens: 0.901 | Train Spec: 0.955 [TRAIN SCORE: 1.856] || Val Sens: 0.938 | Val Spec: 0.892 [VAL SCORE: 1.829] || Test Sens: 0.907 | Test Spec: 0.976 [TEST SCORE: 1.883]

**#94: area2 + perimeter3 + smoothness3 + concave_points3**

Optimized Thresholds: area2: 68.7367 | perimeter3: 110.5604 | smoothness3: 0.1759 | concave_points3: 0.1769

Train Sens: 0.934 | Train Spec: 0.970 [TRAIN SCORE: 1.904] || Val Sens: 0.938 | Val Spec: 0.892 [VAL SCORE: 1.829] || Test Sens: 0.930 | Test Spec: 0.976 [TEST SCORE: 1.906]

**#95: texture1 + area1 + area2 + texture3 + perimeter3 + smoothness3 + concavity3 + concave_points3 + symmetry3 + fr**

Optimized Thresholds: texture1: 31.7324 | area1: 958.3723 | area2: 74.0776 | texture3: 45.5718 | perimeter3: 109.9813 | smoothness3: 0.1744 | concavity3: 0.8401 | concave_points3: 0.2019 | symmetry3: 0.4869 | fractal_dimension3: 0.1303

Train Sens: 0.950 | Train Spec: 0.947 [TRAIN SCORE: 1.898] || Val Sens: 0.938 | Val Spec: 0.892 [VAL SCORE: 1.829] || Test Sens: 0.930 | Test Spec: 0.952 [TEST SCORE: 1.883]

**#96: texture1 + area1 + area2 + texture3 + perimeter3 + smoothness3 + compactness3 + concave_points3 + symmetry3**

Optimized Thresholds: texture1: 31.7324 | area1: 958.3723 | area2: 74.0776 | texture3: 45.5718 | perimeter3: 109.9813 | smoothness3: 0.1744 | compactness3: 0.6348 | concave_points3: 0.2019 | symmetry3: 0.4869 | fractal_dimension3: 0.1303

Train Sens: 0.950 | Train Spec: 0.955 [TRAIN SCORE: 1.905] || Val Sens: 0.938 | Val Spec: 0.892 [VAL SCORE: 1.829] || Test Sens: 0.930 | Test Spec: 0.952 [TEST SCORE: 1.883]

##### #97: texture1 + area1 + concave_points1* + area2 + smoothness3 + compactness3 + concavity3

Optimized Thresholds: texture1: 37.4618 | area1: 962.0549 | concave_points1*: 0.1057 | area2: 40.3507 | smoothness3: 0.1749 | compactness3: 0.3918 | concavity3: 1.0061

Train Sens: 0.909 | Train Spec: 0.940 [TRAIN SCORE: 1.849] || Val Sens: 0.854 | Val Spec: 0.973 [VAL SCORE: 1.827] || Test Sens: 0.884 | Test Spec: 0.881 [TEST SCORE: 1.765]

##### #98: area2 + texture3 + smoothness3 + concavity3 + symmetry3 + fractal_dimension3

Optimized Thresholds: area2: 43.6156 | texture3: 43.9649 | smoothness3: 0.1785 | concavity3: 0.3494 | symmetry3: 0.5113 | fractal_dimension3: 0.1337

Train Sens: 0.926 | Train Spec: 0.917 [TRAIN SCORE: 1.843] || Val Sens: 0.854 | Val Spec: 0.973 [VAL SCORE: 1.827] || Test Sens: 0.884 | Test Spec: 0.881 [TEST SCORE: 1.765]

##### #99: area2 + texture3 + compactness3* + concavity3 + fractal_dimension3

Optimized Thresholds: area2: 41.6739 | texture3: 46.4745 | compactness3*: 0.5094 | concavity3: 0.3664 | fractal_dimension3: 0.1235

Train Sens: 0.909 | Train Spec: 0.910 [TRAIN SCORE: 1.819] || Val Sens: 0.854 | Val Spec: 0.973 [VAL SCORE: 1.827] || Test Sens: 0.860 | Test Spec: 0.881 [TEST SCORE: 1.741]

##### #100: concave_points1* + area2 + texture3* + perimeter3 + compactness3 + concavity3 + concave_points3

Optimized Thresholds: concave_points1*: 0.1240 | area2: 46.7176 | texture3*: 42.9497 | perimeter3: 116.1344 | compactness3: 0.6576 | concavity3: 0.9520 | concave_points3: 0.1682

Train Sens: 0.901 | Train Spec: 0.962 [TRAIN SCORE: 1.863] || Val Sens: 0.854 | Val Spec: 0.973 [VAL SCORE: 1.827] || Test Sens: 0.907 | Test Spec: 0.929 [TEST SCORE: 1.836]

##### #101: texture1 + area2 + smoothness3 + concavity3 + fractal_dimension3

Optimized Thresholds: texture1: 39.2254 | area2: 43.5430 | smoothness3: 0.1804 | concavity3: 0.3488 | fractal_dimension3: 0.1281

Train Sens: 0.926 | Train Spec: 0.917 [TRAIN SCORE: 1.843] || Val Sens: 0.854 | Val Spec: 0.973 [VAL SCORE: 1.827] || Test Sens: 0.860 | Test Spec: 0.881 [TEST SCORE: 1.741]

##### #102: area2 + smoothness3 + compactness3 + symmetry3 + fractal_dimension3

Optimized Thresholds: area2: 40.9333 | smoothness3: 0.1791 | compactness3: 0.3634 | symmetry3: 0.4696 | fractal_dimension3: 0.1262

Train Sens: 0.884 | Train Spec: 0.932 [TRAIN SCORE: 1.817] || Val Sens: 0.854 | Val Spec: 0.973 [VAL SCORE: 1.827] || Test Sens: 0.884 | Test Spec: 0.881 [TEST SCORE: 1.765]

##### #103: area2* + perimeter3 + smoothness3 + concavity3 + concave_points3 + symmetry3 + fractal_dimension3

Optimized Thresholds: area2*: 280.9770 | perimeter3: 109.5216 | smoothness3: 0.1776 | concavity3: 0.9608 | concave_points3: 0.2127 | symmetry3: 0.3645 | fractal_dimension3: 0.1312

Train Sens: 0.942 | Train Spec: 0.955 [TRAIN SCORE: 1.897] || Val Sens: 0.958 | Val Spec: 0.865 [VAL SCORE: 1.823] || Test Sens: 0.930 | Test Spec: 0.952 [TEST SCORE: 1.883]

##### #104: concave_points1 + perimeter3 + smoothness3 + compactness3 + concave_points3 + symmetry3 + fractal_dimens[illegible]

Optimized Thresholds: concave_points1: 0.1760 | perimeter3: 109.6221 | smoothness3: 0.1738 | compactness3: 0.6569 | concave_points3: 0.1793 | symmetry3: 0.3605 | fractal_dimension3: 0.1324

Train Sens: 0.950 | Train Spec: 0.955 [TRAIN SCORE: 1.905] || Val Sens: 0.958 | Val Spec: 0.865 [VAL SCORE: 1.823] || Test Sens: 0.930 | Test Spec: 0.952 [TEST SCORE: 1.883]

##### #105: area1 + concave_points1 + texture3 + perimeter3 + smoothness3 + concave_points3 + symmetry3 + fractal_dime[illegible]

Optimized Thresholds: area1: 719.4099 | concave_points1: 0.1689 | texture3: 43.2607 | perimeter3: 110.0273 | smoothness3: 0.1740 | concave_points3: 0.2180 | symmetry3: 0.3625 | fractal_dimension3: 0.1394

Train Sens: 0.950 | Train Spec: 0.955 [TRAIN SCORE: 1.905] || Val Sens: 0.958 | Val Spec: 0.865 [VAL SCORE: 1.823] || Test Sens: 0.953 | Test Spec: 0.952 [TEST SCORE: 1.906]

##### #106: concave_points1 + perimeter3 + smoothness3 + concavity3 + concave_points3 + symmetry3 + fractal_dimension[illegible]

Optimized Thresholds: concave_points1: 0.1760 | perimeter3: 109.6221 | smoothness3: 0.1738 | concavity3: 0.8707 | concave_points3: 0.1793 | symmetry3: 0.3605 | fractal_dimension3: 0.1324

Train Sens: 0.950 | Train Spec: 0.955 [TRAIN SCORE: 1.905] || Val Sens: 0.958 | Val Spec: 0.865 [VAL SCORE: 1.823] || Test Sens: 0.930 | Test Spec: 0.952 [TEST SCORE: 1.883]

##### #107: concave_points1* + perimeter3 + smoothness3 + compactness3 + concave_points3 + symmetry3

Optimized Thresholds: concave_points1*: 0.1493 | perimeter3: 109.7339 | smoothness3: 0.1747 | compactness3: 0.6716 | concave_points3: 0.2050 | symmetry3: 0.3618

Train Sens: 0.950 | Train Spec: 0.955 [TRAIN SCORE: 1.905] || Val Sens: 0.958 | Val Spec: 0.865 [VAL SCORE: 1.823] || Test Sens: 0.930 | Test Spec: 0.976 [TEST SCORE: 1.906]

##### #108: texture3 + perimeter3 + smoothness3 + concave_points3 + symmetry3

Optimized Thresholds: texture3: 46.0230 | perimeter3: 109.8933 | smoothness3: 0.1784 | concave_points3: 0.1928 | symmetry3: 0.3619

Train Sens: 0.950 | Train Spec: 0.955 [TRAIN SCORE: 1.905] || Val Sens: 0.958 | Val Spec: 0.865 [VAL SCORE: 1.823] || Test Sens: 0.930 | Test Spec: 0.976 [TEST SCORE: 1.906]

##### #109: texture3 + perimeter3 + smoothness3 + concavity3 + concave_points3 + symmetry3

Optimized Thresholds: texture3: 48.8364 | perimeter3: 109.9615 | smoothness3: 0.1754 | concavity3: 0.9219 | concave_points3: 0.2200 | symmetry3: 0.3625

Train Sens: 0.950 | Train Spec: 0.955 [TRAIN SCORE: 1.905] || Val Sens: 0.958 | Val Spec: 0.865 [VAL SCORE: 1.823] || Test Sens: 0.930 | Test Spec: 0.976 [TEST SCORE: 1.906]

##### #110: texture3 + perimeter3 + smoothness3 + concavity3 + concave_points3 + symmetry3 + fractal_dimension3

Optimized Thresholds: texture3: 45.2602 | perimeter3: 109.6221 | smoothness3: 0.1738 | concavity3: 0.8707 | concave_points3: 0.1793 | symmetry3: 0.3605 | fractal_dimension3: 0.1324

Train Sens: 0.950 | Train Spec: 0.955 [TRAIN SCORE: 1.905] || Val Sens: 0.958 | Val Spec: 0.865 [VAL SCORE: 1.823] || Test Sens: 0.930 | Test Spec: 0.952 [TEST SCORE: 1.883]

##### #111: area1 + perimeter3 + smoothness3 + concavity3 + concave_points3 + symmetry3 + fractal_dimension3

Optimized Thresholds: area1: 2203.7137 | perimeter3: 109.6221 | smoothness3: 0.1738 | concavity3: 0.8707 | concave_points3: 0.1793 | symmetry3: 0.3605 | fractal_dimension3: 0.1324

Train Sens: 0.950 | Train Spec: 0.955 [TRAIN SCORE: 1.905] || Val Sens: 0.958 | Val Spec: 0.865 [VAL SCORE: 1.823] || Test Sens: 0.930 | Test Spec: 0.952 [TEST SCORE: 1.883]

##### #112: area1 + perimeter3 + smoothness3 + compactness3 + concave_points3 + symmetry3 + fractal_dimension3

Optimized Thresholds: area1: 2203.7137 | perimeter3: 109.6221 | smoothness3: 0.1738 | compactness3: 0.6569 | concave_points3: 0.1793 | symmetry3: 0.3605 | fractal_dimension3: 0.1324

Train Sens: 0.950 | Train Spec: 0.955 [TRAIN SCORE: 1.905] || Val Sens: 0.958 | Val Spec: 0.865 [VAL SCORE: 1.823] || Test Sens: 0.930 | Test Spec: 0.952 [TEST SCORE: 1.883]

**#113: texture3 + perimeter3 + smoothness3 + compactness3 + concave_points3 + symmetry3 + fractal_dimension3**

Optimized Thresholds: texture3: 45.2602 | perimeter3: 109.6221 | smoothness3: 0.1738 | compactness3: 0.6569 | concave_points3: 0.1793 | symmetry3: 0.3605 | fractal_dimension3: 0.1324

Train Sens: 0.950 | Train Spec: 0.955 [TRAIN SCORE: 1.905] || Val Sens: 0.958 | Val Spec: 0.865 [VAL SCORE: 1.823] || Test Sens: 0.930 | Test Spec: 0.952 [TEST SCORE: 1.883]

**#114: texture3 + perimeter3 + smoothness3 + compactness3 + concave_points3 + symmetry3**

Optimized Thresholds: texture3: 48.8364 | perimeter3: 109.9615 | smoothness3: 0.1754 | compactness3: 0.6940 | concave_points3: 0.2200 | symmetry3: 0.3625

Train Sens: 0.950 | Train Spec: 0.955 [TRAIN SCORE: 1.905] || Val Sens: 0.958 | Val Spec: 0.865 [VAL SCORE: 1.823] || Test Sens: 0.930 | Test Spec: 0.976 [TEST SCORE: 1.906]

**#115: texture1 + perimeter3 + smoothness3 + compactness3 + concave_points3 + symmetry3 + fractal_dimension3**

Optimized Thresholds: texture1: 35.7210 | perimeter3: 109.6221 | smoothness3: 0.1738 | compactness3: 0.6569 | concave_points3: 0.1793 | symmetry3: 0.3605 | fractal_dimension3: 0.1324

Train Sens: 0.950 | Train Spec: 0.955 [TRAIN SCORE: 1.905] || Val Sens: 0.958 | Val Spec: 0.865 [VAL SCORE: 1.823] || Test Sens: 0.930 | Test Spec: 0.952 [TEST SCORE: 1.883]

**#116: area1* + perimeter3 + smoothness3 + concavity3 + concave_points3 + symmetry3**

Optimized Thresholds: area1*: 1891.7674 | perimeter3: 109.7339 | smoothness3: 0.1747 | concavity3: 0.8910 | concave_points3: 0.2050 | symmetry3: 0.3618

Train Sens: 0.950 | Train Spec: 0.955 [TRAIN SCORE: 1.905] || Val Sens: 0.958 | Val Spec: 0.865 [VAL SCORE: 1.823] || Test Sens: 0.930 | Test Spec: 0.976 [TEST SCORE: 1.906]

**#117: texture1 + perimeter3 + smoothness3 + concavity3 + concave_points3 + symmetry3 + fractal_dimension3**

Optimized Thresholds: texture1: 35.7210 | perimeter3: 109.6221 | smoothness3: 0.1738 | concavity3: 0.8707 | concave_points3: 0.1793 | symmetry3: 0.3605 | fractal_dimension3: 0.1324

Train Sens: 0.950 | Train Spec: 0.955 [TRAIN SCORE: 1.905] || Val Sens: 0.958 | Val Spec: 0.865 [VAL SCORE: 1.823] || Test Sens: 0.930 | Test Spec: 0.952 [TEST SCORE: 1.883]

**#118: concave_points1* + perimeter3 + smoothness3 + concavity3 + concave_points3 + symmetry3**

Optimized Thresholds: concave_points1*: 0.1493 | perimeter3: 109.7339 | smoothness3: 0.1747 | concavity3: 0.8910 | concave_points3: 0.2050 | symmetry3: 0.3618

Train Sens: 0.950 | Train Spec: 0.955 [TRAIN SCORE: 1.905] || Val Sens: 0.958 | Val Spec: 0.865 [VAL SCORE: 1.823] || Test Sens: 0.930 | Test Spec: 0.976 [TEST SCORE: 1.906]

**#119: area1* + perimeter3 + smoothness3 + compactness3 + concave_points3 + symmetry3**

Optimized Thresholds: area1*: 1891.7674 | perimeter3: 109.7339 | smoothness3: 0.1747 | compactness3: 0.6716 | concave_points3: 0.2050 | symmetry3: 0.3618

Train Sens: 0.950 | Train Spec: 0.955 [TRAIN SCORE: 1.905] || Val Sens: 0.958 | Val Spec: 0.865 [VAL SCORE: 1.823] || Test Sens: 0.930 | Test Spec: 0.976 [TEST SCORE: 1.906]

**#120: concave_points1 + area2 + texture3 + perimeter3 + compactness3 + concavity3 + concave_points3 + symmetry3**

Optimized Thresholds: concave_points1: 0.1039 | area2: 93.0484 | texture3: 35.8450 | perimeter3: 109.5293 | compactness3: 0.7577 | concavity3: 0.7821 | concave_points3: 0.1938 | symmetry3: 0.3621

Train Sens: 0.950 | Train Spec: 0.947 [TRAIN SCORE: 1.898] || Val Sens: 0.958 | Val Spec: 0.865 [VAL SCORE: 1.823] || Test Sens: 0.930 | Test Spec: 0.929 [TEST SCORE: 1.859]

**#121: area2 + texture3 + compactness3 + concavity3 + symmetry3 + fractal_dimension3**

Optimized Thresholds: area2: 41.2489 | texture3: 43.7415 | compactness3: 0.3383 | concavity3: 0.3811 | symmetry3: 0.4495 | fractal_dimension3: 0.1208

Train Sens: 0.934 | Train Spec: 0.917 [TRAIN SCORE: 1.851] || Val Sens: 0.875 | Val Spec: 0.946 [VAL SCORE: 1.821] || Test Sens: 0.907 | Test Spec: 0.857 [TEST SCORE: 1.764]

**#122: area2 + texture3 + compactness3 + concave_points3 + symmetry3**

Optimized Thresholds: area2: 49.0094 | texture3: 33.2849 | compactness3: 0.8317 | concave_points3: 0.1501 | symmetry3: 0.3679

Train Sens: 0.934 | Train Spec: 0.940 [TRAIN SCORE: 1.874] || Val Sens: 0.875 | Val Spec: 0.946 [VAL SCORE: 1.821] || Test Sens: 0.953 | Test Spec: 0.881 [TEST SCORE: 1.834]

**#123: area1 + area2 + smoothness3 + concavity3 + concave_points3 + fractal_dimension3**

Optimized Thresholds: area1: 691.5235 | area2: 288.3208 | smoothness3: 0.1747 | concavity3: 1.0525 | concave_points3: 0.1602 | fractal_dimension3: 0.1301

Train Sens: 0.942 | Train Spec: 0.955 [TRAIN SCORE: 1.897] || Val Sens: 0.875 | Val Spec: 0.946 [VAL SCORE: 1.821] || Test Sens: 0.953 | Test Spec: 0.976 [TEST SCORE: 1.930]

**#124: area2 + texture3 + compactness3 + concavity3 + symmetry3**

Optimized Thresholds: area2: 42.8613 | texture3: 49.4692 | compactness3: 0.3539 | concavity3: 0.3981 | symmetry3: 0.3992

Train Sens: 0.917 | Train Spec: 0.925 [TRAIN SCORE: 1.842] || Val Sens: 0.875 | Val Spec: 0.946 [VAL SCORE: 1.821] || Test Sens: 0.907 | Test Spec: 0.881 [TEST SCORE: 1.788]

**#125: texture1* + area1 + texture3 + compactness3 + concavity3**

Optimized Thresholds: texture1*: 33.0784 | area1: 695.2762 | texture3: 44.0490 | compactness3: 0.3811 | concavity3: 0.8969

Train Sens: 0.909 | Train Spec: 0.925 [TRAIN SCORE: 1.834] || Val Sens: 0.875 | Val Spec: 0.946 [VAL SCORE: 1.821] || Test Sens: 0.930 | Test Spec: 0.952 [TEST SCORE: 1.883]

**#126: area1 + texture3 + smoothness3 + concavity3 + concave_points3**

Optimized Thresholds: area1: 694.7156 | texture3: 45.9178 | smoothness3: 0.1776 | concavity3: 1.0098 | concave_points3: 0.1540

Train Sens: 0.950 | Train Spec: 0.940 [TRAIN SCORE: 1.890] || Val Sens: 0.875 | Val Spec: 0.946 [VAL SCORE: 1.821] || Test Sens: 0.977 | Test Spec: 0.976 [TEST SCORE: 1.953]

**#127: smoothness3 + compactness3 + concave_points3 + symmetry3**

Optimized Thresholds: smoothness3: 0.2034 | compactness3: 0.7003 | concave_points3: 0.1224 | symmetry3: 0.4723

Train Sens: 0.901 | Train Spec: 0.895 [TRAIN SCORE: 1.796] || Val Sens: 0.875 | Val Spec: 0.946 [VAL SCORE: 1.821] || Test Sens: 0.953 | Test Spec: 0.929 [TEST SCORE: 1.882]

**#128: area1 + area2* + smoothness3 + concave_points3 + symmetry3 + fractal_dimension3**

Optimized Thresholds: area1: 694.1693 | area2*: 331.1761 | smoothness3: 0.1771 | concave_points3: 0.1605 | symmetry3: 0.3884 | fractal_dimension3: 0.1333

Train Sens: 0.942 | Train Spec: 0.955 [TRAIN SCORE: 1.897] || Val Sens: 0.875 | Val Spec: 0.946 [VAL SCORE: 1.821] || Test Sens: 0.953 | Test Spec: 0.952 [TEST SCORE: 1.906]

**#129: texture1 + area2 + compactness3 + concavity3**

Optimized Thresholds: texture1: 35.3658 | area2: 43.0879 | compactness3: 0.3373 | concavity3: 0.3898

Train Sens: 0.934 | Train Spec: 0.925 [TRAIN SCORE: 1.859] || Val Sens: 0.875 | Val Spec: 0.946 [VAL SCORE: 1.821] || Test Sens: 0.884 | Test Spec: 0.857 [TEST SCORE: 1.741]

**#130: area2 + texture3 + concavity3 + concave_points3 + symmetry3**

Optimized Thresholds: area2: 49.0094 | texture3: 33.2849 | concavity3: 1.1124 | concave_points3: 0.1501 | symmetry3: 0.3679

Train Sens: 0.934 | Train Spec: 0.940 [TRAIN SCORE: 1.874] || Val Sens: 0.875 | Val Spec: 0.946 [VAL SCORE: 1.821] || Test Sens: 0.953 | Test Spec: 0.881 [TEST SCORE: 1.834]

**#131: area2 + texture3 + compactness3 + concavity3**

Optimized Thresholds: area2: 43.0942 | texture3: 46.8446 | compactness3: 0.3452 | concavity3: 0.3994

Train Sens: 0.917 | Train Spec: 0.925 [TRAIN SCORE: 1.842] || Val Sens: 0.875 | Val Spec: 0.946 [VAL SCORE: 1.821] || Test Sens: 0.884 | Test Spec: 0.857 [TEST SCORE: 1.741]

**#132: texture1 + area2 + texture3 + compactness3 + concavity3 + symmetry3**

Optimized Thresholds: texture1: 36.6200 | area2: 42.8613 | texture3: 49.4692 | compactness3: 0.3539 | concavity3: 0.3981 | symmetry3: 0.3992

Train Sens: 0.917 | Train Spec: 0.925 [TRAIN SCORE: 1.842] || Val Sens: 0.875 | Val Spec: 0.946 [VAL SCORE: 1.821] || Test Sens: 0.907 | Test Spec: 0.881 [TEST SCORE: 1.788]

**#133: area2 + compactness3 + concavity3**

Optimized Thresholds: area2: 43.0879 | compactness3: 0.3373 | concavity3: 0.3898

Train Sens: 0.934 | Train Spec: 0.925 [TRAIN SCORE: 1.859] || Val Sens: 0.875 | Val Spec: 0.946 [VAL SCORE: 1.821] || Test Sens: 0.884 | Test Spec: 0.857 [TEST SCORE: 1.741]

**#134: texture1 + area1 + concave_points1 + area2 + compactness3 + concavity3 + fractal_dimension3**

Optimized Thresholds: texture1: 36.1944 | area1: 792.3991 | concave_points1: 0.0613 | area2: 40.5356 | compactness3: 0.3966 | concavity3: 1.1940 | fractal_dimension3: 0.1215

Train Sens: 0.934 | Train Spec: 0.932 [TRAIN SCORE: 1.866] || Val Sens: 0.875 | Val Spec: 0.946 [VAL SCORE: 1.821] || Test Sens: 0.953 | Test Spec: 0.857 [TEST SCORE: 1.811]

**#135: concave_points3 + symmetry3***

Optimized Thresholds: concave_points3: 0.1224 | symmetry3*: 0.4491

Train Sens: 0.901 | Train Spec: 0.895 [TRAIN SCORE: 1.796] || Val Sens: 0.875 | Val Spec: 0.946 [VAL SCORE: 1.821] || Test Sens: 0.977 | Test Spec: 0.929 [TEST SCORE: 1.905]

**#136: area2 + smoothness3 + compactness3 + concavity3 + symmetry3 + fractal_dimension3**

Optimized Thresholds: area2: 41.2489 | smoothness3: 0.1969 | compactness3: 0.3383 | concavity3: 0.3811 | symmetry3: 0.4495 | fractal_dimension3: 0.1208

Train Sens: 0.934 | Train Spec: 0.917 [TRAIN SCORE: 1.851] || Val Sens: 0.875 | Val Spec: 0.946 [VAL SCORE: 1.821] || Test Sens: 0.907 | Test Spec: 0.857 [TEST SCORE: 1.764]

##### #137: area1 + concave_points1 + texture3 + smoothness3 + concave_points3 + fractal_dimension3

Optimized Thresholds: area1: 693.7209 | concave_points1: 0.1010 | texture3: 45.4632 | smoothness3: 0.1762 | concave_points3: 0.1607 | fractal_dimension3: 0.1348

Train Sens: 0.942 | Train Spec: 0.955 [TRAIN SCORE: 1.897] || Val Sens: 0.875 | Val Spec: 0.946 [VAL SCORE: 1.821] || Test Sens: 0.953 | Test Spec: 0.976 [TEST SCORE: 1.930]

##### #138: texture1 + area1 + concave_points1 + smoothness3 + concave_points3 + symmetry3

Optimized Thresholds: texture1: 36.9965 | area1: 696.8947 | concave_points1: 0.1096 | smoothness3: 0.1780 | concave_points3: 0.1602 | symmetry3: 0.4761

Train Sens: 0.942 | Train Spec: 0.955 [TRAIN SCORE: 1.897] || Val Sens: 0.875 | Val Spec: 0.946 [VAL SCORE: 1.821] || Test Sens: 0.953 | Test Spec: 1.000 [TEST SCORE: 1.953]

##### #139: texture1 + area1 + area2 + compactness3 + concavity3

Optimized Thresholds: texture1: 34.1123 | area1: 919.7682 | area2: 42.7705 | compactness3: 0.3382 | concavity3: 0.8461

Train Sens: 0.934 | Train Spec: 0.925 [TRAIN SCORE: 1.859] || Val Sens: 0.875 | Val Spec: 0.946 [VAL SCORE: 1.821] || Test Sens: 0.907 | Test Spec: 0.857 [TEST SCORE: 1.764]

##### #140: texture1 + area1 + texture3 + smoothness3 + concavity3 + concave_points3

Optimized Thresholds: texture1: 34.4565 | area1: 691.5012 | texture3: 47.7034 | smoothness3: 0.1739 | concavity3: 0.8546 | concave_points3: 0.1505

Train Sens: 0.950 | Train Spec: 0.940 [TRAIN SCORE: 1.890] || Val Sens: 0.875 | Val Spec: 0.946 [VAL SCORE: 1.821] || Test Sens: 0.977 | Test Spec: 0.976 [TEST SCORE: 1.953]

##### #141: concavity3 + concave_points3 + symmetry3

Optimized Thresholds: concavity3: 0.9332 | concave_points3: 0.1223 | symmetry3: 0.4489

Train Sens: 0.901 | Train Spec: 0.895 [TRAIN SCORE: 1.796] || Val Sens: 0.875 | Val Spec: 0.946 [VAL SCORE: 1.821] || Test Sens: 0.977 | Test Spec: 0.929 [TEST SCORE: 1.905]

##### #142: smoothness3 + concave_points3 + symmetry3*

Optimized Thresholds: smoothness3: 0.2017 | concave_points3: 0.1224 | symmetry3*: 0.4594

Train Sens: 0.901 | Train Spec: 0.895 [TRAIN SCORE: 1.796] || Val Sens: 0.875 | Val Spec: 0.946 [VAL SCORE: 1.821] || Test Sens: 0.977 | Test Spec: 0.929 [TEST SCORE: 1.905]

##### #143: compactness3 + concave_points3 + symmetry3

Optimized Thresholds: compactness3: 0.7183 | concave_points3: 0.1223 | symmetry3: 0.4630

Train Sens: 0.901 | Train Spec: 0.895 [TRAIN SCORE: 1.796] || Val Sens: 0.875 | Val Spec: 0.946 [VAL SCORE: 1.821] || Test Sens: 0.977 | Test Spec: 0.929 [TEST SCORE: 1.905]

##### #144: texture1 + area1 + concave_points1 + area2 + texture3 + compactness3 + concavity3 + fractal_dimension3

Optimized Thresholds: texture1: 34.7360 | area1: 863.7121 | concave_points1: 0.1619 | area2: 43.0588 | texture3: 45.5117 | compactness3: 0.3455 | concavity3: 0.7241 | fractal_dimension3: 0.1242

Train Sens: 0.926 | Train Spec: 0.932 [TRAIN SCORE: 1.858] || Val Sens: 0.875 | Val Spec: 0.946 [VAL SCORE: 1.821] || Test Sens: 0.907 | Test Spec: 0.881 [TEST SCORE: 1.788]

##### #145: area1 + area2 + compactness3 + concavity3

Optimized Thresholds: area1: 903.3985 | area2: 43.0811 | compactness3: 0.3495 | concavity3: 1.1803

Train Sens: 0.926 | Train Spec: 0.932 [TRAIN SCORE: 1.858] || Val Sens: 0.875 | Val Spec: 0.946 [VAL SCORE: 1.821] || Test Sens: 0.907 | Test Spec: 0.881 [TEST SCORE: 1.788]

**#146: area1 + area2 + compactness3 + fractal_dimension3**

Optimized Thresholds: area1: 903.3985 | area2: 43.0811 | compactness3: 0.3495 | fractal_dimension3: 0.1395

Train Sens: 0.926 | Train Spec: 0.932 [TRAIN SCORE: 1.858] || Val Sens: 0.875 | Val Spec: 0.946 [VAL SCORE: 1.821] || Test Sens: 0.907 | Test Spec: 0.881 [TEST SCORE: 1.788]

**#147: texture1* + area2* + perimeter3 + concavity3 + concave_points3 + fractal_dimension3**

Optimized Thresholds: texture1*: 24.8775 | area2*: 140.4740 | perimeter3: 115.3840 | concavity3: 0.9318 | concave_points3: 0.1666 | fractal_dimension3: 0.1377

Train Sens: 0.901 | Train Spec: 0.955 [TRAIN SCORE: 1.856] || Val Sens: 0.875 | Val Spec: 0.946 [VAL SCORE: 1.821] || Test Sens: 0.907 | Test Spec: 0.952 [TEST SCORE: 1.859]

**#148: texture1* + area2 + perimeter3 + concavity3 + concave_points3 + symmetry3**

Optimized Thresholds: texture1*: 26.2606 | area2: 107.6638 | perimeter3: 115.2040 | concavity3: 0.9086 | concave_points3: 0.1671 | symmetry3: 0.4608

Train Sens: 0.884 | Train Spec: 0.962 [TRAIN SCORE: 1.847] || Val Sens: 0.875 | Val Spec: 0.946 [VAL SCORE: 1.821] || Test Sens: 0.907 | Test Spec: 0.976 [TEST SCORE: 1.883]

**#149: smoothness3 + compactness3* + concavity3 + concave_points3 + symmetry3**

Optimized Thresholds: smoothness3: 0.2016 | compactness3*: 0.6866 | concavity3: 0.9766 | concave_points3: 0.1223 | symmetry3: 0.4244

Train Sens: 0.901 | Train Spec: 0.895 [TRAIN SCORE: 1.796] || Val Sens: 0.875 | Val Spec: 0.946 [VAL SCORE: 1.821] || Test Sens: 0.977 | Test Spec: 0.929 [TEST SCORE: 1.905]

**#150: smoothness3 + compactness3 + concave_points3 + symmetry3 + fractal_dimension3**

Optimized Thresholds: smoothness3: 0.1914 | compactness3: 0.7456 | concave_points3: 0.1222 | symmetry3: 0.4509 | fractal_dimension3: 0.1400

Train Sens: 0.901 | Train Spec: 0.895 [TRAIN SCORE: 1.796] || Val Sens: 0.875 | Val Spec: 0.946 [VAL SCORE: 1.821] || Test Sens: 0.977 | Test Spec: 0.905 [TEST SCORE: 1.882]

**#151: texture1 + area1 + concave_points1 + smoothness3 + compactness3 + concavity3 + concave_points3**

Optimized Thresholds: texture1: 39.2588 | area1: 691.1969 | concave_points1: 0.0844 | smoothness3: 0.1781 | compactness3: 0.5752 | concavity3: 1.0644 | concave_points3: 0.1549

Train Sens: 0.950 | Train Spec: 0.940 [TRAIN SCORE: 1.890] || Val Sens: 0.875 | Val Spec: 0.946 [VAL SCORE: 1.821] || Test Sens: 0.977 | Test Spec: 0.952 [TEST SCORE: 1.929]

**#152: texture1 + area2* + texture3* + perimeter3 + smoothness3 + concavity3 + concave_points3**

Optimized Thresholds: texture1: 24.0559 | area2*: 245.5287 | texture3*: 39.7650 | perimeter3: 114.8002 | smoothness3: 0.1794 | concavity3: 0.9410 | concave_points3: 0.1790

Train Sens: 0.893 | Train Spec: 0.955 [TRAIN SCORE: 1.847] || Val Sens: 0.875 | Val Spec: 0.946 [VAL SCORE: 1.821] || Test Sens: 0.930 | Test Spec: 0.976 [TEST SCORE: 1.906]

**#153: texture1* + area1 + compactness3 + concavity3**

Optimized Thresholds: texture1*: 32.9433 | area1: 692.9038 | compactness3: 0.3813 | concavity3: 0.8515

Train Sens: 0.909 | Train Spec: 0.925 [TRAIN SCORE: 1.834] || Val Sens: 0.875 | Val Spec: 0.946 [VAL SCORE: 1.821] || Test Sens: 0.930 | Test Spec: 0.952 [TEST SCORE: 1.883]

##### #154: texture1 + area2 + texture3 + compactness3 + concavity3

Optimized Thresholds: texture1: 38.2257 | area2: 40.6933 | texture3: 46.0207 | compactness3: 0.3364 | concavity3: 0.3827

Train Sens: 0.934 | Train Spec: 0.910 [TRAIN SCORE: 1.844] || Val Sens: 0.875 | Val Spec: 0.946 [VAL SCORE: 1.821] || Test Sens: 0.884 | Test Spec: 0.857 [TEST SCORE: 1.741]

##### #155: compactness3 + concavity3 + concave_points3 + symmetry3*

Optimized Thresholds: compactness3: 0.6986 | concavity3: 1.0296 | concave_points3: 0.1223 | symmetry3*: 0.4711

Train Sens: 0.901 | Train Spec: 0.895 [TRAIN SCORE: 1.796] || Val Sens: 0.875 | Val Spec: 0.946 [VAL SCORE: 1.821] || Test Sens: 0.953 | Test Spec: 0.929 [TEST SCORE: 1.882]

##### #156: texture1* + area1 + concave_points1* + texture3 + perimeter3 + concavity3 + concave_points3 + symmetry3

Optimized Thresholds: texture1*: 25.7534 | area1: 1579.2927 | concave_points1*: 0.1185 | texture3: 34.8258 | perimeter3: 114.9789 | concavity3: 1.0456 | concave_points3: 0.1772 | symmetry3: 0.4397

Train Sens: 0.884 | Train Spec: 0.962 [TRAIN SCORE: 1.847] || Val Sens: 0.875 | Val Spec: 0.946 [VAL SCORE: 1.821] || Test Sens: 0.907 | Test Spec: 0.976 [TEST SCORE: 1.883]

##### #157: smoothness3 + concave_points3 + symmetry3 + fractal_dimension3

Optimized Thresholds: smoothness3: 0.1896 | concave_points3: 0.1223 | symmetry3: 0.4515 | fractal_dimension3: 0.1374

Train Sens: 0.901 | Train Spec: 0.895 [TRAIN SCORE: 1.796] || Val Sens: 0.875 | Val Spec: 0.946 [VAL SCORE: 1.821] || Test Sens: 0.977 | Test Spec: 0.905 [TEST SCORE: 1.882]

##### #158: area1 + area2 + smoothness3 + compactness3 + concave_points3 + fractal_dimension3

Optimized Thresholds: area1: 691.5235 | area2: 288.3208 | smoothness3: 0.1747 | compactness3: 0.7884 | concave_points3: 0.1602 | fractal_dimension3: 0.1301

Train Sens: 0.942 | Train Spec: 0.955 [TRAIN SCORE: 1.897] || Val Sens: 0.875 | Val Spec: 0.946 [VAL SCORE: 1.821] || Test Sens: 0.953 | Test Spec: 0.976 [TEST SCORE: 1.930]

##### #159: texture1 + area2 + compactness3 + concavity3 + fractal_dimension3

Optimized Thresholds: texture1: 39.1486 | area2: 42.9197 | compactness3: 0.3536 | concavity3: 0.3971 | fractal_dimension3: 0.1202

Train Sens: 0.917 | Train Spec: 0.925 [TRAIN SCORE: 1.842] || Val Sens: 0.875 | Val Spec: 0.946 [VAL SCORE: 1.821] || Test Sens: 0.884 | Test Spec: 0.881 [TEST SCORE: 1.765]

##### #160: smoothness3 + concavity3 + concave_points3 + symmetry3*

Optimized Thresholds: smoothness3: 0.2022 | concavity3: 1.0240 | concave_points3: 0.1224 | symmetry3*: 0.4589

Train Sens: 0.901 | Train Spec: 0.895 [TRAIN SCORE: 1.796] || Val Sens: 0.875 | Val Spec: 0.946 [VAL SCORE: 1.821] || Test Sens: 0.977 | Test Spec: 0.929 [TEST SCORE: 1.905]

##### #161: concave_points3 + symmetry3* + fractal_dimension3

Optimized Thresholds: concave_points3: 0.1224 | symmetry3*: 0.4585 | fractal_dimension3: 0.1382

Train Sens: 0.901 | Train Spec: 0.895 [TRAIN SCORE: 1.796] || Val Sens: 0.875 | Val Spec: 0.946 [VAL SCORE: 1.821] || Test Sens: 0.977 | Test Spec: 0.905 [TEST SCORE: 1.882]

##### #162: area1 + area2 + texture3 + compactness3

Optimized Thresholds: area1: 924.6689 | area2: 42.8103 | texture3: 42.5927 | compactness3: 0.3492

Train Sens: 0.926 | Train Spec: 0.932 [TRAIN SCORE: 1.858] || Val Sens: 0.875 | Val Spec: 0.946 [VAL SCORE: 1.821] || Test Sens: 0.907 | Test Spec: 0.881 [TEST SCORE: 1.788]

##### #163: concavity3 + concave_points3 + symmetry3 + fractal_dimension3

Optimized Thresholds: concavity3: 1.0040 | concave_points3: 0.1223 | symmetry3: 0.4375 | fractal_dimension3: 0.1395

Train Sens: 0.901 | Train Spec: 0.895 [TRAIN SCORE: 1.796] || Val Sens: 0.875 | Val Spec: 0.946 [VAL SCORE: 1.821] || Test Sens: 0.977 | Test Spec: 0.905 [TEST SCORE: 1.882]

##### #164: area1 + area2 + smoothness3 + compactness3 + concavity3

Optimized Thresholds: area1: 888.6843 | area2: 42.7636 | smoothness3: 0.1883 | compactness3: 0.3470 | concavity3: 0.7709

Train Sens: 0.934 | Train Spec: 0.932 [TRAIN SCORE: 1.866] || Val Sens: 0.875 | Val Spec: 0.946 [VAL SCORE: 1.821] || Test Sens: 0.907 | Test Spec: 0.881 [TEST SCORE: 1.788]

##### #165: area1 + area2 + compactness3 + concavity3 + fractal_dimension3

Optimized Thresholds: area1: 929.4392 | area2: 43.0126 | compactness3: 0.3482 | concavity3: 0.5231 | fractal_dimension3: 0.1414

Train Sens: 0.926 | Train Spec: 0.932 [TRAIN SCORE: 1.858] || Val Sens: 0.875 | Val Spec: 0.946 [VAL SCORE: 1.821] || Test Sens: 0.884 | Test Spec: 0.881 [TEST SCORE: 1.765]

##### #166: area1 + concave_points1 + smoothness3 + compactness3 + concave_points3 + fractal_dimension3

Optimized Thresholds: area1: 691.5235 | concave_points1: 0.1091 | smoothness3: 0.1747 | compactness3: 0.7884 | concave_points3: 0.1602 | fractal_dimension3: 0.1301

Train Sens: 0.942 | Train Spec: 0.955 [TRAIN SCORE: 1.897] || Val Sens: 0.875 | Val Spec: 0.946 [VAL SCORE: 1.821] || Test Sens: 0.953 | Test Spec: 0.976 [TEST SCORE: 1.930]

##### #167: area1 + area2 + texture3 + smoothness3 + concave_points3 + fractal_dimension3

Optimized Thresholds: area1: 693.7209 | area2: 267.5245 | texture3: 45.4632 | smoothness3: 0.1762 | concave_points3: 0.1607 | fractal_dimension3: 0.1348

Train Sens: 0.942 | Train Spec: 0.955 [TRAIN SCORE: 1.897] || Val Sens: 0.875 | Val Spec: 0.946 [VAL SCORE: 1.821] || Test Sens: 0.953 | Test Spec: 0.976 [TEST SCORE: 1.930]

##### #168: area1 + area2 + smoothness3 + concave_points3 + fractal_dimension3

Optimized Thresholds: area1: 693.5656 | area2: 49.8940 | smoothness3: 0.1747 | concave_points3: 0.1591 | fractal_dimension3: 0.1397

Train Sens: 0.950 | Train Spec: 0.947 [TRAIN SCORE: 1.898] || Val Sens: 0.875 | Val Spec: 0.946 [VAL SCORE: 1.821] || Test Sens: 0.953 | Test Spec: 0.929 [TEST SCORE: 1.882]

##### #169: area1 + area2 + texture3 + compactness3 + concavity3

Optimized Thresholds: area1: 888.6843 | area2: 42.7636 | texture3: 41.8168 | compactness3: 0.3470 | concavity3: 0.7709

Train Sens: 0.926 | Train Spec: 0.932 [TRAIN SCORE: 1.858] || Val Sens: 0.875 | Val Spec: 0.946 [VAL SCORE: 1.821] || Test Sens: 0.907 | Test Spec: 0.881 [TEST SCORE: 1.788]

##### #170: area1 + concave_points1* + smoothness3 + concave_points3 + symmetry3 + fractal_dimension3

Optimized Thresholds: area1: 694.1693 | concave_points1*: 0.1257 | smoothness3: 0.1771 | concave_points3: 0.1605 | symmetry3:

0.3884 | fractal_dimension3: 0.1333

Train Sens: 0.942 | Train Spec: 0.955 [TRAIN SCORE: 1.897] || Val Sens: 0.875 | Val Spec: 0.946 [VAL SCORE: 1.821] || Test Sens: 0.953 | Test Spec: 0.952 [TEST SCORE: 1.906]

**#171: area1 + concave_points1 + smoothness3 + concavity3 + concave_points3 + fractal_dimension3**

Optimized Thresholds: area1: 691.5235 | concave_points1: 0.1091 | smoothness3: 0.1747 | concavity3: 1.0525 | concave_points3: 0.1602 | fractal_dimension3: 0.1301

Train Sens: 0.942 | Train Spec: 0.955 [TRAIN SCORE: 1.897] || Val Sens: 0.875 | Val Spec: 0.946 [VAL SCORE: 1.821] || Test Sens: 0.953 | Test Spec: 0.976 [TEST SCORE: 1.930]

**#172: texture1 + area1 + concave_points1 + area2 + compactness3 + fractal_dimension3**

Optimized Thresholds: texture1: 33.9657 | area1: 859.4607 | concave_points1: 0.0602 | area2: 39.6811 | compactness3: 0.3868 | fractal_dimension3: 0.1298

Train Sens: 0.934 | Train Spec: 0.940 [TRAIN SCORE: 1.874] || Val Sens: 0.896 | Val Spec: 0.919 [VAL SCORE: 1.815] || Test Sens: 0.930 | Test Spec: 0.857 [TEST SCORE: 1.787]

**#173: texture1 + area1 + concave_points1 + area2 + compactness3 + concavity3**

Optimized Thresholds: texture1: 33.9657 | area1: 859.4607 | concave_points1: 0.0602 | area2: 39.6811 | compactness3: 0.3868 | concavity3: 1.0450

Train Sens: 0.934 | Train Spec: 0.940 [TRAIN SCORE: 1.874] || Val Sens: 0.896 | Val Spec: 0.919 [VAL SCORE: 1.815] || Test Sens: 0.930 | Test Spec: 0.857 [TEST SCORE: 1.787]

**#174: area1 + concave_points1* + area2* + texture3 + perimeter3 + compactness3 + concavity3 + concave_points3 + sy**

Optimized Thresholds: area1: 1791.0362 | concave_points1*: 0.1157 | area2*: 132.2703 | texture3: 35.4950 | perimeter3: 113.7700 | compactness3: 0.7804 | concavity3: 1.0837 | concave_points3: 0.1795 | symmetry3*: 0.4659

Train Sens: 0.884 | Train Spec: 0.962 [TRAIN SCORE: 1.847] || Val Sens: 0.896 | Val Spec: 0.919 [VAL SCORE: 1.815] || Test Sens: 0.907 | Test Spec: 0.976 [TEST SCORE: 1.883]

**#175: area1 + concave_points1 + area2 + texture3 + concavity3 + concave_points3 + symmetry3**

Optimized Thresholds: area1: 926.8509 | concave_points1: 0.1854 | area2: 40.1977 | texture3: 33.2968 | concavity3: 0.8705 | concave_points3: 0.1647 | symmetry3: 0.3655

Train Sens: 0.950 | Train Spec: 0.940 [TRAIN SCORE: 1.890] || Val Sens: 0.896 | Val Spec: 0.919 [VAL SCORE: 1.815] || Test Sens: 0.907 | Test Spec: 0.881 [TEST SCORE: 1.788]

**#176: texture1 + concave_points1 + area2 + perimeter3 + smoothness3 + concavity3 + symmetry3**

Optimized Thresholds: texture1: 37.2718 | concave_points1: 0.0785 | area2: 76.1985 | perimeter3: 110.0196 | smoothness3: 0.1739 | concavity3: 1.0289 | symmetry3: 0.4851

Train Sens: 0.950 | Train Spec: 0.955 [TRAIN SCORE: 1.905] || Val Sens: 0.917 | Val Spec: 0.892 [VAL SCORE: 1.809] || Test Sens: 0.930 | Test Spec: 0.929 [TEST SCORE: 1.859]

**#177: area1* + area2 + perimeter3 + compactness3 + concave_points3 + fractal_dimension3**

Optimized Thresholds: area1*: 1606.6574 | area2: 48.0482 | perimeter3: 110.3609 | compactness3: 0.6735 | concave_points3: 0.1812 | fractal_dimension3: 0.1349

Train Sens: 0.926 | Train Spec: 0.955 [TRAIN SCORE: 1.881] || Val Sens: 0.917 | Val Spec: 0.892 [VAL SCORE: 1.809] || Test Sens: 0.907 | Test Spec: 0.905 [TEST SCORE: 1.812]

###### #178: texture1 + area1 + concave_points1 + area2 + perimeter3 + smoothness3 + concavity3 + symmetry3 + fractal_dim

Optimized Thresholds: texture1: 37.2493 | area1: 1589.5371 | concave_points1: 0.1551 | area2: 64.1871 | perimeter3: 109.8031 | smoothness3: 0.1740 | concavity3: 0.9218 | symmetry3: 0.4527 | fractal_dimension3: 0.1356

Train Sens: 0.950 | Train Spec: 0.955 [TRAIN SCORE: 1.905] || Val Sens: 0.917 | Val Spec: 0.892 [VAL SCORE: 1.809] || Test Sens: 0.907 | Test Spec: 0.952 [TEST SCORE: 1.859]

###### #179: texture1 + area1 + concave_points1 + area2 + perimeter3 + smoothness3 + concavity3 + concave_points3 + fracta

Optimized Thresholds: texture1: 37.2493 | area1: 1589.5371 | concave_points1: 0.1551 | area2: 64.1871 | perimeter3: 109.8031 | smoothness3: 0.1740 | concavity3: 0.9218 | concave_points3: 0.2159 | fractal_dimension3: 0.1356

Train Sens: 0.950 | Train Spec: 0.955 [TRAIN SCORE: 1.905] || Val Sens: 0.917 | Val Spec: 0.892 [VAL SCORE: 1.809] || Test Sens: 0.907 | Test Spec: 0.952 [TEST SCORE: 1.859]

###### #180: area2 + perimeter3 + smoothness3 + compactness3 + concavity3 + concave_points3 + fractal_dimension3

Optimized Thresholds: area2: 78.8811 | perimeter3: 109.7266 | smoothness3: 0.1802 | compactness3: 0.8380 | concavity3: 0.9773 | concave_points3: 0.1712 | fractal_dimension3: 0.1383

Train Sens: 0.950 | Train Spec: 0.955 [TRAIN SCORE: 1.905] || Val Sens: 0.917 | Val Spec: 0.892 [VAL SCORE: 1.809] || Test Sens: 0.907 | Test Spec: 0.976 [TEST SCORE: 1.883]

###### #181: concave_points1 + area2 + perimeter3 + concave_points3 + symmetry3 + fractal_dimension3

Optimized Thresholds: concave_points1: 0.0794 | area2: 46.8611 | perimeter3: 109.7700 | concave_points3: 0.1824 | symmetry3: 0.4939 | fractal_dimension3: 0.1327

Train Sens: 0.950 | Train Spec: 0.955 [TRAIN SCORE: 1.905] || Val Sens: 0.917 | Val Spec: 0.892 [VAL SCORE: 1.809] || Test Sens: 0.907 | Test Spec: 0.881 [TEST SCORE: 1.788]

###### #182: texture1 + area1 + concave_points1 + area2 + perimeter3 + smoothness3 + compactness3 + concave_points3 + fr

Optimized Thresholds: texture1: 37.2493 | area1: 1589.5371 | concave_points1: 0.1551 | area2: 64.1871 | perimeter3: 109.8031 | smoothness3: 0.1740 | compactness3: 0.6939 | concave_points3: 0.2159 | fractal_dimension3: 0.1356

Train Sens: 0.950 | Train Spec: 0.962 [TRAIN SCORE: 1.913] || Val Sens: 0.917 | Val Spec: 0.892 [VAL SCORE: 1.809] || Test Sens: 0.907 | Test Spec: 0.952 [TEST SCORE: 1.859]

###### #183: concave_points1* + area2 + perimeter3 + concavity3 + concave_points3 + fractal_dimension3

Optimized Thresholds: concave_points1*: 0.1309 | area2: 53.8254 | perimeter3: 110.2868 | concavity3: 0.9349 | concave_points3: 0.1718 | fractal_dimension3: 0.1386

Train Sens: 0.934 | Train Spec: 0.955 [TRAIN SCORE: 1.889] || Val Sens: 0.917 | Val Spec: 0.892 [VAL SCORE: 1.809] || Test Sens: 0.907 | Test Spec: 0.952 [TEST SCORE: 1.859]

###### #184: texture1 + area1 + concave_points1 + area2 + perimeter3 + smoothness3 + compactness3 + symmetry3 + fractal_

Optimized Thresholds: texture1: 37.2493 | area1: 1589.5371 | concave_points1: 0.1551 | area2: 64.1871 | perimeter3: 109.8031 | smoothness3: 0.1740 | compactness3: 0.6939 | symmetry3: 0.4527 | fractal_dimension3: 0.1356

Train Sens: 0.950 | Train Spec: 0.962 [TRAIN SCORE: 1.913] || Val Sens: 0.917 | Val Spec: 0.892 [VAL SCORE: 1.809] || Test Sens: 0.907 | Test Spec: 0.952 [TEST SCORE: 1.859]

###### #185: area1* + concave_points1 + perimeter3 + smoothness3 + compactness3 + concavity3 + concave_points3 + symm

Optimized Thresholds: area1*: 2036.0026 | concave_points1: 0.1061 | perimeter3: 109.6912 | smoothness3: 0.1838 | compactness3: 0.7369 | concavity3: 1.1345 | concave_points3: 0.1908 | symmetry3: 0.4094

Train Sens: 0.934 | Train Spec: 0.955 [TRAIN SCORE: 1.889] || Val Sens: 0.917 | Val Spec: 0.892 [VAL SCORE: 1.809] || Test Sens: 0.907 | Test Spec: 0.976 [TEST SCORE: 1.883]

**#186: perimeter3 + smoothness3 + compactness3 + concave_points3 + symmetry3 + fractal_dimension3**

Optimized Thresholds: perimeter3: 109.5769 | smoothness3: 0.1829 | compactness3: 0.7173 | concave_points3: 0.1923 | symmetry3: 0.4117 | fractal_dimension3: 0.1355

Train Sens: 0.934 | Train Spec: 0.955 [TRAIN SCORE: 1.889] || Val Sens: 0.917 | Val Spec: 0.892 [VAL SCORE: 1.809] || Test Sens: 0.907 | Test Spec: 0.952 [TEST SCORE: 1.859]

**#187: texture1 + concave_points1* + area2 + perimeter3 + smoothness3 + compactness3 + symmetry3**

Optimized Thresholds: texture1: 38.2172 | concave_points1*: 0.1168 | area2: 73.0588 | perimeter3: 109.9426 | smoothness3: 0.1747 | compactness3: 0.7284 | symmetry3: 0.4671

Train Sens: 0.950 | Train Spec: 0.962 [TRAIN SCORE: 1.913] || Val Sens: 0.917 | Val Spec: 0.892 [VAL SCORE: 1.809] || Test Sens: 0.907 | Test Spec: 0.976 [TEST SCORE: 1.883]

**#188: concave_points1* + area2 + perimeter3 + concavity3 + concave_points3 + symmetry3**

Optimized Thresholds: concave_points1*: 0.1258 | area2: 62.0940 | perimeter3: 109.6676 | concavity3: 0.9974 | concave_points3: 0.1656 | symmetry3: 0.4721

Train Sens: 0.942 | Train Spec: 0.947 [TRAIN SCORE: 1.890] || Val Sens: 0.917 | Val Spec: 0.892 [VAL SCORE: 1.809] || Test Sens: 0.907 | Test Spec: 0.976 [TEST SCORE: 1.883]

**#189: texture1* + concave_points1* + area2* + perimeter3 + smoothness3 + symmetry3 + fractal_dimension3**

Optimized Thresholds: texture1*: 35.2775 | concave_points1*: 0.1219 | area2*: 109.6497 | perimeter3: 110.7954 | smoothness3: 0.1768 | symmetry3: 0.4538 | fractal_dimension3: 0.1312

Train Sens: 0.917 | Train Spec: 0.970 [TRAIN SCORE: 1.887] || Val Sens: 0.917 | Val Spec: 0.892 [VAL SCORE: 1.809] || Test Sens: 0.907 | Test Spec: 0.976 [TEST SCORE: 1.883]

**#190: area1* + concave_points1 + area2* + texture3* + perimeter3 + compactness3 + concavity3 + concave_points3 + sy**

Optimized Thresholds: area1*: 1635.2604 | concave_points1: 0.1275 | area2*: 157.2029 | texture3*: 39.2909 | perimeter3: 112.1436 | compactness3: 0.7284 | concavity3: 0.9129 | concave_points3: 0.1801 | symmetry3: 0.4159 | fractal_dimension3: 0.1354

Train Sens: 0.884 | Train Spec: 0.962 [TRAIN SCORE: 1.847] || Val Sens: 0.917 | Val Spec: 0.892 [VAL SCORE: 1.809] || Test Sens: 0.907 | Test Spec: 0.952 [TEST SCORE: 1.859]

**#191: area1* + perimeter3 + compactness3 + concavity3 + concave_points3**

Optimized Thresholds: area1*: 1408.2739 | perimeter3: 109.8708 | compactness3: 0.7857 | concavity3: 1.0515 | concave_points3: 0.1628

Train Sens: 0.942 | Train Spec: 0.947 [TRAIN SCORE: 1.890] || Val Sens: 0.917 | Val Spec: 0.892 [VAL SCORE: 1.809] || Test Sens: 0.907 | Test Spec: 1.000 [TEST SCORE: 1.907]

**#192: area1* + perimeter3 + compactness3 + concave_points3 + fractal_dimension3**

Optimized Thresholds: area1*: 1603.5470 | perimeter3: 109.6882 | compactness3: 0.7308 | concave_points3: 0.1616 | fractal_dimension3: 0.1368

Train Sens: 0.942 | Train Spec: 0.947 [TRAIN SCORE: 1.890] || Val Sens: 0.917 | Val Spec: 0.892 [VAL SCORE: 1.809] || Test Sens: 0.907 | Test Spec: 0.976 [TEST SCORE: 1.883]

**#193: area1 + concave_points1 + perimeter3 + smoothness3 + concavity3 + concave_points3 + symmetry3* + fractal_di**

Optimized Thresholds: area1: 1020.1222 | concave_points1: 0.0991 | perimeter3: 109.6111 | smoothness3: 0.1953 | concavity3: 0.9071 | concave_points3: 0.1974 | symmetry3*: 0.4150 | fractal_dimension3: 0.1412

Train Sens: 0.926 | Train Spec: 0.955 [TRAIN SCORE: 1.881] || Val Sens: 0.917 | Val Spec: 0.892 [VAL SCORE: 1.809] || Test Sens: 0.907 | Test Spec: 0.976 [TEST SCORE: 1.883]

**#194: area1 + area2 + texture3 + perimeter3 + smoothness3 + compactness3 + concavity3 + symmetry3 + fractal_dimen[illegible]**

Optimized Thresholds: area1: 743.7372 | area2: 93.5469 | texture3: 46.6155 | perimeter3: 109.7466 | smoothness3: 0.1745 | compactness3: 0.7211 | concavity3: 1.1638 | symmetry3: 0.5314 | fractal_dimension3: 0.1403

Train Sens: 0.950 | Train Spec: 0.955 [TRAIN SCORE: 1.905] || Val Sens: 0.917 | Val Spec: 0.892 [VAL SCORE: 1.809] || Test Sens: 0.907 | Test Spec: 0.976 [TEST SCORE: 1.883]

**#195: perimeter3 + smoothness3 + compactness3 + concavity3 + concave_points3 + symmetry3 + fractal_dimension3**

Optimized Thresholds: perimeter3: 109.8385 | smoothness3: 0.1848 | compactness3: 0.6487 | concavity3: 1.0718 | concave_points3: 0.1632 | symmetry3: 0.4588 | fractal_dimension3: 0.1401

Train Sens: 0.950 | Train Spec: 0.947 [TRAIN SCORE: 1.898] || Val Sens: 0.917 | Val Spec: 0.892 [VAL SCORE: 1.809] || Test Sens: 0.907 | Test Spec: 0.976 [TEST SCORE: 1.883]

**#196: concave_points1 + texture3* + perimeter3 + smoothness3 + concavity3 + concave_points3**

Optimized Thresholds: concave_points1: 0.1359 | texture3*: 40.5082 | perimeter3: 111.6910 | smoothness3: 0.1957 | concavity3: 0.9648 | concave_points3: 0.1640

Train Sens: 0.917 | Train Spec: 0.955 [TRAIN SCORE: 1.872] || Val Sens: 0.917 | Val Spec: 0.892 [VAL SCORE: 1.809] || Test Sens: 0.907 | Test Spec: 0.976 [TEST SCORE: 1.883]

**#197: area1 + concave_points1 + perimeter3 + smoothness3 + compactness3 + concave_points3 + symmetry3* + fracta[illegible]**

Optimized Thresholds: area1: 1020.1222 | concave_points1: 0.0991 | perimeter3: 109.6111 | smoothness3: 0.1953 | compactness3: 0.6833 | concave_points3: 0.1974 | symmetry3*: 0.4150 | fractal_dimension3: 0.1412

Train Sens: 0.926 | Train Spec: 0.955 [TRAIN SCORE: 1.881] || Val Sens: 0.917 | Val Spec: 0.892 [VAL SCORE: 1.809] || Test Sens: 0.907 | Test Spec: 0.976 [TEST SCORE: 1.883]

**#198: texture1 + concave_points1 + perimeter3 + smoothness3 + compactness3 + symmetry3 + fractal_dimension3**

Optimized Thresholds: texture1: 36.1665 | concave_points1: 0.1879 | perimeter3: 109.6634 | smoothness3: 0.1759 | compactness3: 0.7360 | symmetry3: 0.4891 | fractal_dimension3: 0.1397

Train Sens: 0.942 | Train Spec: 0.962 [TRAIN SCORE: 1.905] || Val Sens: 0.917 | Val Spec: 0.892 [VAL SCORE: 1.809] || Test Sens: 0.907 | Test Spec: 0.976 [TEST SCORE: 1.883]

**#199: area2 + perimeter3 + smoothness3 + compactness3 + concave_points3 + symmetry3 + fractal_dimension3**

Optimized Thresholds: area2: 42.8503 | perimeter3: 109.4448 | smoothness3: 0.1768 | compactness3: 0.6899 | concave_points3: 0.2800 | symmetry3: 0.4306 | fractal_dimension3: 0.1370

Train Sens: 0.967 | Train Spec: 0.955 [TRAIN SCORE: 1.922] || Val Sens: 0.917 | Val Spec: 0.892 [VAL SCORE: 1.809] || Test Sens: 0.953 | Test Spec: 0.905 [TEST SCORE: 1.858]

**#200: perimeter3 + smoothness3 + concavity3 + concave_points3 + symmetry3 + fractal_dimension3**

Optimized Thresholds: perimeter3: 109.5769 | smoothness3: 0.1829 | concavity3: 0.9542 | concave_points3: 0.1923 | symmetry3: 0.4117 | fractal_dimension3: 0.1355

Train Sens: 0.934 | Train Spec: 0.955 [TRAIN SCORE: 1.889] || Val Sens: 0.917 | Val Spec: 0.892 [VAL SCORE: 1.809] || Test Sens: 0.907 | Test Spec: 0.952 [TEST SCORE: 1.859]
