## Supplementary material. Outputs TholdStormDX for this study. for "Methodological and Clinical Validation of TholdStormDX v0.0.1: An Advanced Stochastic Engine for the Optimization of Thresholds and Multimarker Panels Applied to Oncology": Breast Prognosis TholdStormDX_RobustReport_20260404_200738.pdf

### Biomarker: texture1

Processed: 04-Apr-2026 18:28

#### 1. Optimization Results

| MODEL | CUT-OFF | TRAIN (SE/SP) | VAL (SE/SP) | TEST (SE/SP) | R2 SCORE |
|---|---|---|---|---|---|
| Empirical (Exact) | 16.7272 | 0.655 / 0.655 | 0.667 / 0.700 | 0.889 / 0.430 | N/A |
| Logistic 2-Parameter | 16.5527 | 0.685 / 0.685 | 0.667 / 0.600 | 0.889 / 0.412 | 0.9879 |
| **Logistic 4-Parameter (Rec.)** | **16.5190** | **0.675 / 0.675** | **0.667 / 0.600** | **0.889 / 0.412** | **0.9899** |
| ThresholdXpert (Stochastic) | 17.1796 | 0.655 / 0.815 | 0.667 / 0.700 | 0.889 / 0.456 | N/A |

#### 2. Diagnostic Performance Curves (Training)

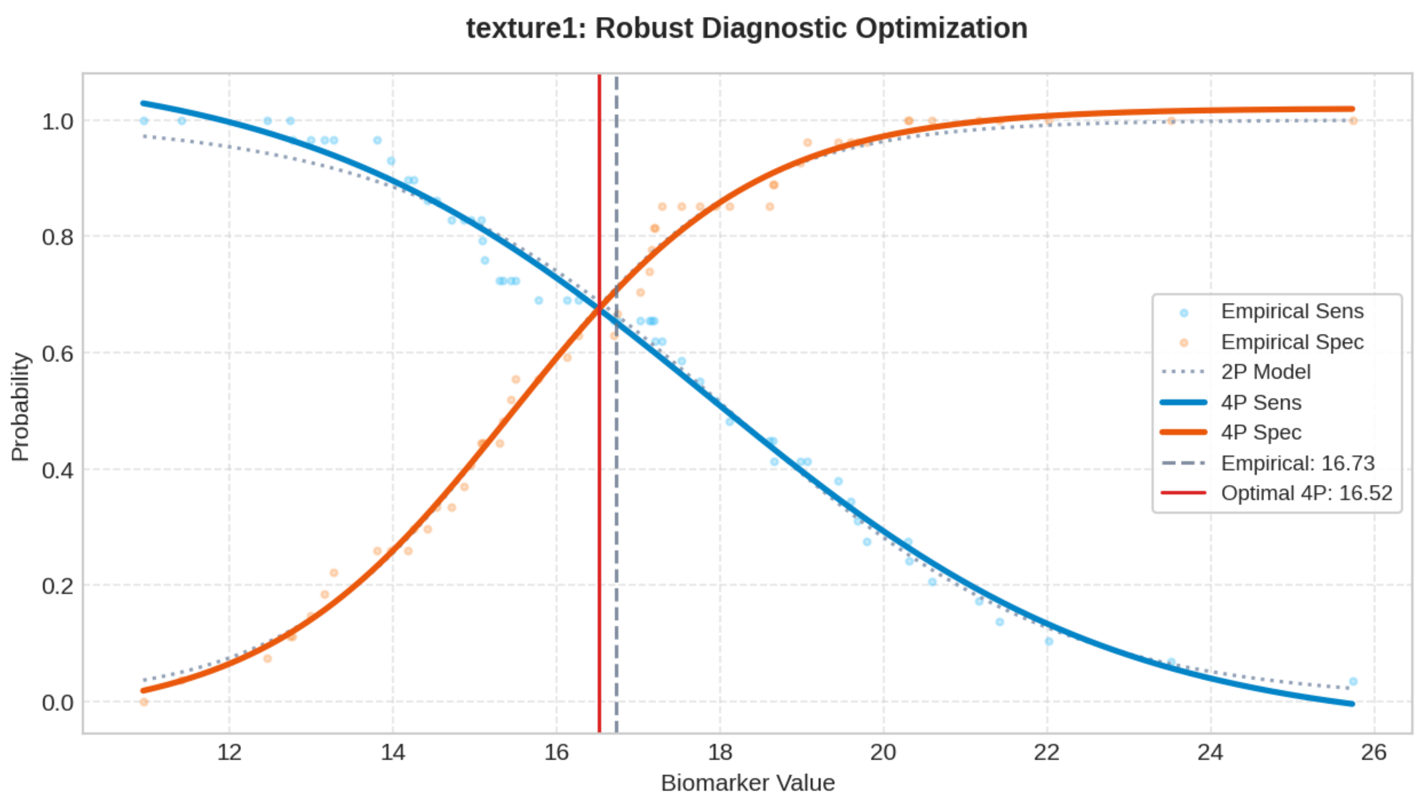

### Biomarker: area3

Processed: 04-Apr-2026 18:31

#### 1. Optimization Results

| MODEL | CUT-OFF | TRAIN (SE/SP) | VAL (SE/SP) | TEST (SE/SP) | R2 SCORE |
|---|---|---|---|---|---|
| Empirical (Exact) | 133.3200 | 0.621 / 0.621 | 0.667 / 0.600 | 0.889 / 0.474 | N/A |
| Logistic 2-Parameter | 134.6159 | 0.658 / 0.658 | 0.667 / 0.600 | 0.889 / 0.482 | 0.9782 |
| **Logistic 4-Parameter (Rec.)** | **134.1363** | **0.642 / 0.642** | **0.667 / 0.600** | **0.889 / 0.482** | **0.9888** |
| ThresholdXpert (Stochastic) | 137.4416 | 0.621 / 0.741 | 0.556 / 0.600 | 0.889 / 0.518 | N/A |

#### 2. Diagnostic Performance Curves (Training)

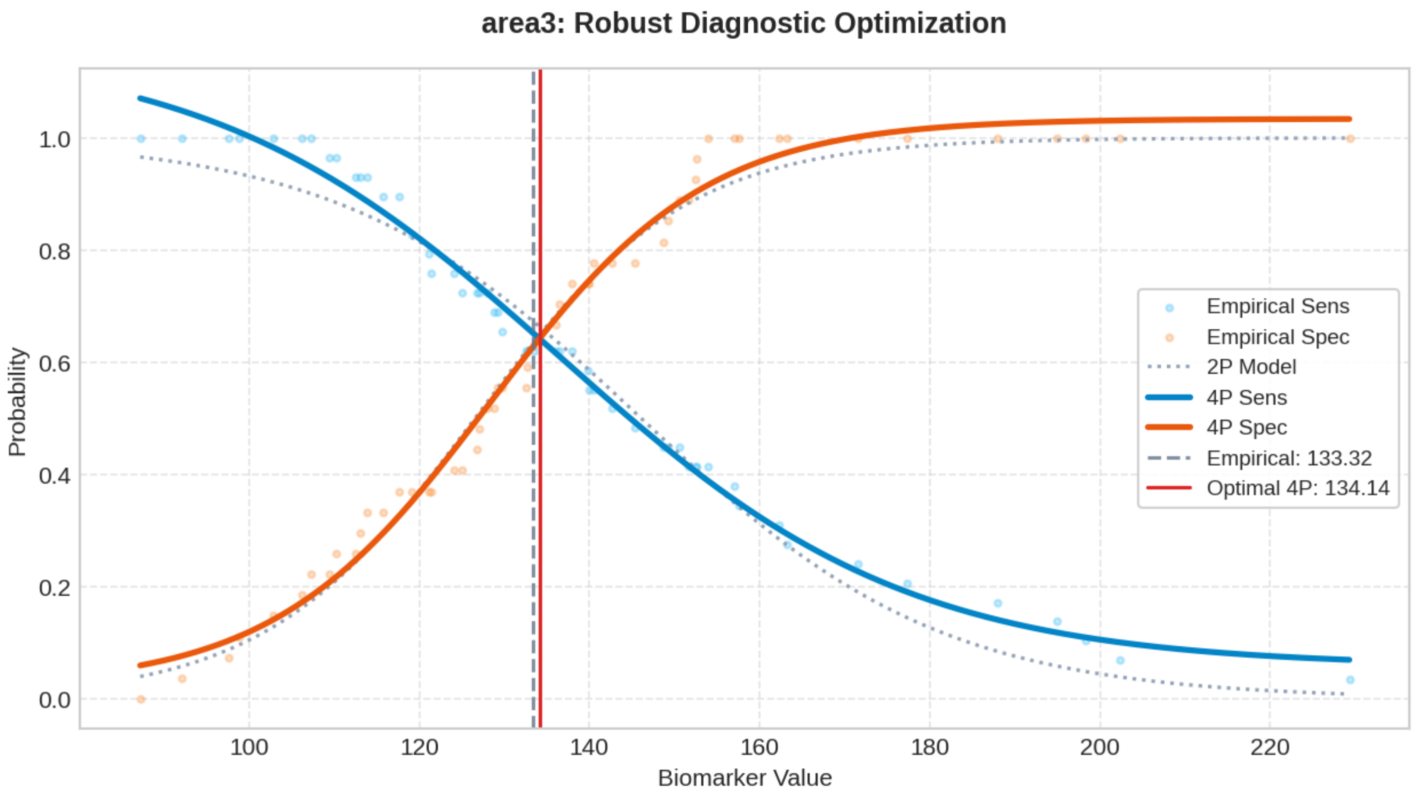

### Biomarker: smoothness3

Processed: 04-Apr-2026 18:32

#### 1. Optimization Results

| MODEL | CUT-OFF | TRAIN (SE/SP) | VAL (SE/SP) | TEST (SE/SP) | R2 SCORE |
|---|---|---|---|---|---|
| Empirical (Exact) | 1268.1348 | 0.624 / 0.624 | 0.778 / 0.600 | 0.889 / 0.482 | N/A |
| Logistic 2-Parameter | 1264.6632 | 0.668 / 0.668 | 0.778 / 0.600 | 0.889 / 0.482 | 0.9797 |
| **Logistic 4-Parameter (Rec.)** | **1250.5308** | **0.653 / 0.653** | **0.778 / 0.600** | **0.889 / 0.465** | **0.9897** |
| ThresholdXpert (Stochastic) | 1283.6028 | 0.621 / 0.667 | 0.778 / 0.600 | 0.889 / 0.500 | N/A |

#### 2. Diagnostic Performance Curves (Training)

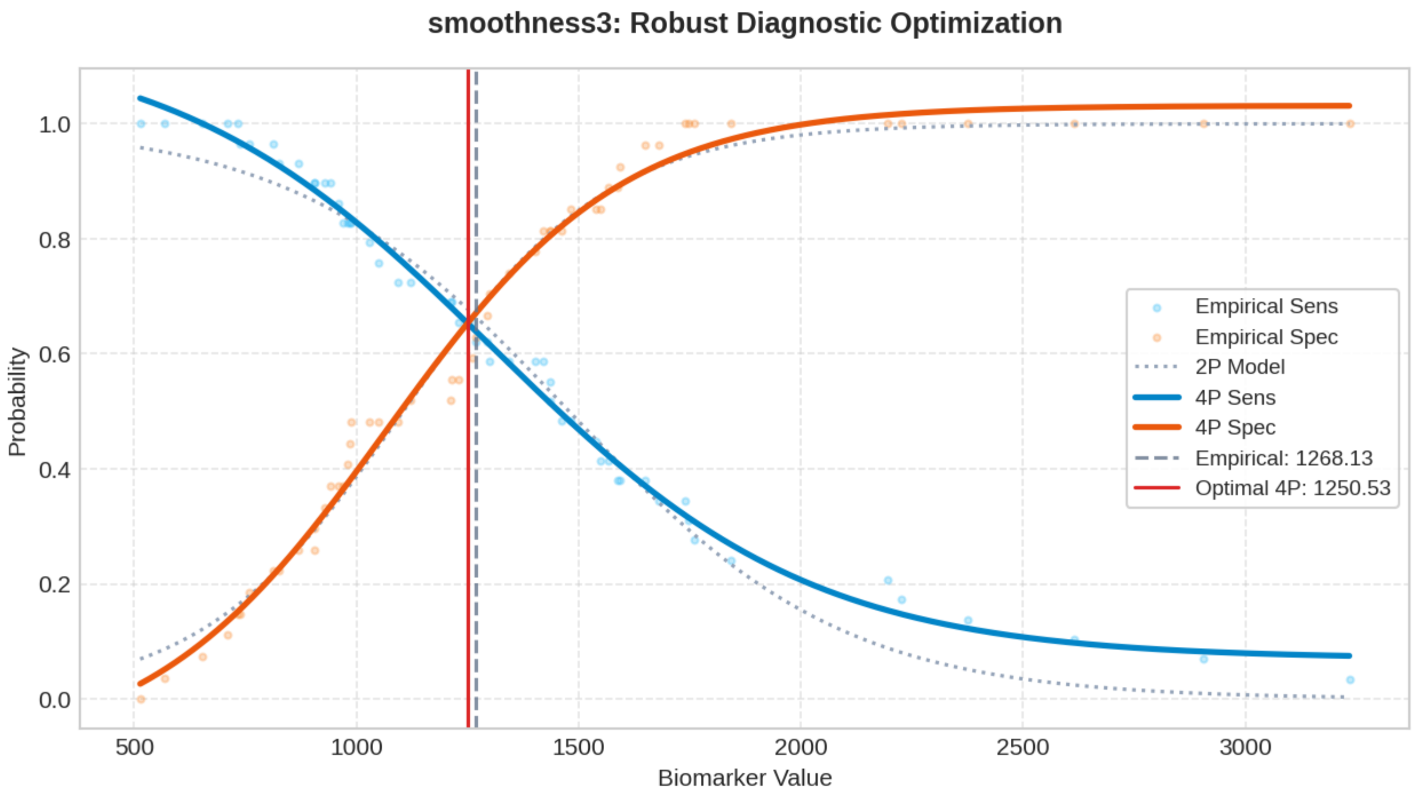

### Biomarker: concavity3

Processed: 04-Apr-2026 18:35

#### 1. Optimization Results

| MODEL | CUT-OFF | TRAIN (SE/SP) | VAL (SE/SP) | TEST (SE/SP) | R2 SCORE |
|---|---|---|---|---|---|
| Empirical (Exact) | 0.3620 | 0.408 / 0.408 | 0.556 / 0.600 | 0.556 / 0.579 | N/A |
| Logistic 2-Parameter | 0.3675 | 0.412 / 0.412 | 0.556 / 0.600 | 0.556 / 0.579 | 0.9749 |
| **Logistic 4-Parameter (Rec.)** | **0.3617** | **0.418 / 0.418** | **0.556 / 0.600** | **0.556 / 0.579** | **0.9877** |
| ThresholdXpert (Stochastic) | 0.3564 | 0.448 / 0.407 | 0.556 / 0.500 | 0.556 / 0.561 | N/A |

#### 2. Diagnostic Performance Curves (Training)

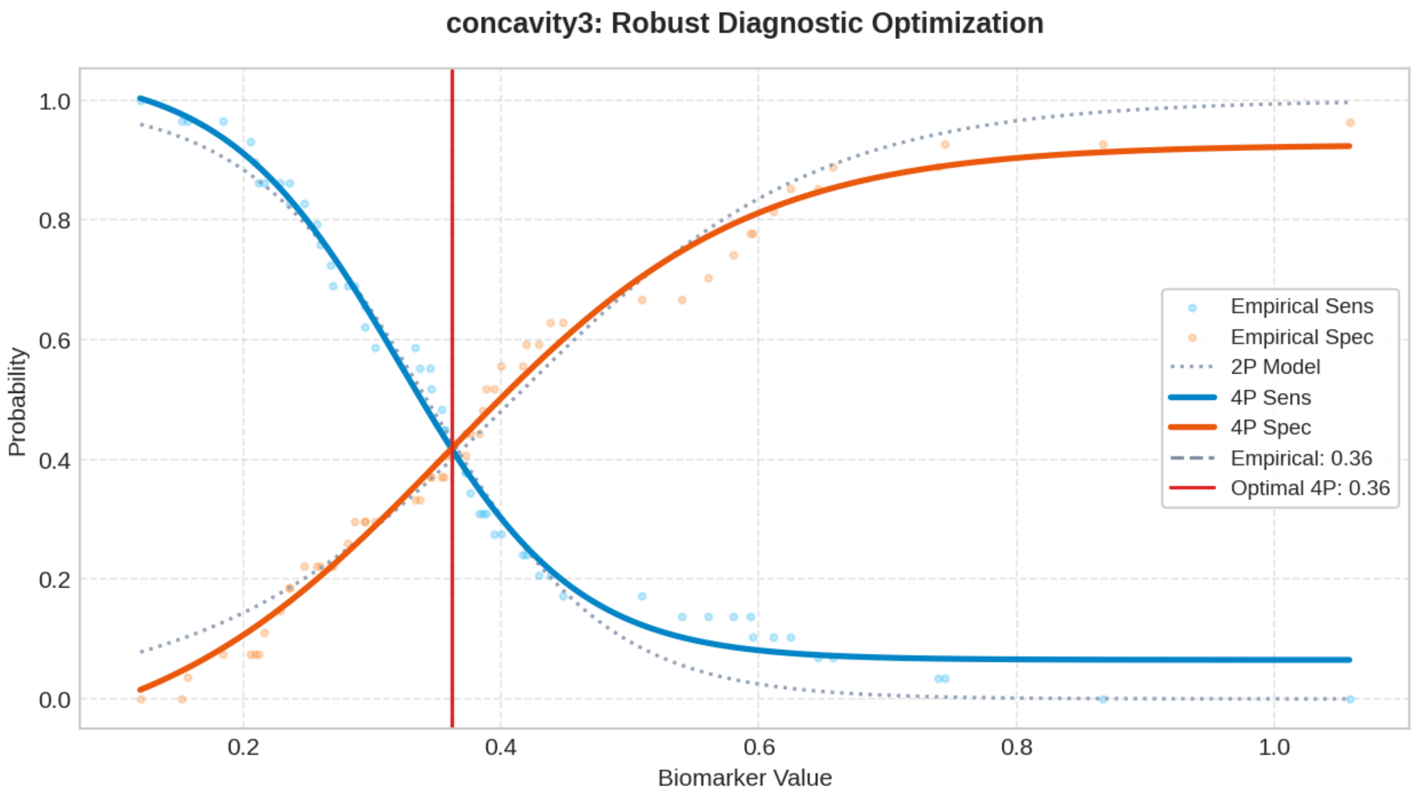

### Biomarker: concave_points3

Processed: 04-Apr-2026 18:37

#### 1. Optimization Results

| MODEL | CUT-OFF | TRAIN (SE/SP) | VAL (SE/SP) | TEST (SE/SP) | R2 SCORE |
|---|---|---|---|---|---|
| Empirical (Exact) | 0.4207 | 0.518 / 0.518 | 0.556 / 0.500 | 0.333 / 0.561 | N/A |
| Logistic 2-Parameter | 0.4517 | 0.505 / 0.505 | 0.444 / 0.500 | 0.222 / 0.614 | 0.9704 |
| **Logistic 4-Parameter (Rec.)** | **0.4440** | **0.501 / 0.501** | **0.444 / 0.500** | **0.222 / 0.596** | **0.9800** |
| ThresholdXpert (Stochastic) | 0.4026 | 0.586 / 0.519 | 0.556 / 0.500 | 0.333 / 0.518 | N/A |

#### 2. Diagnostic Performance Curves (Training)

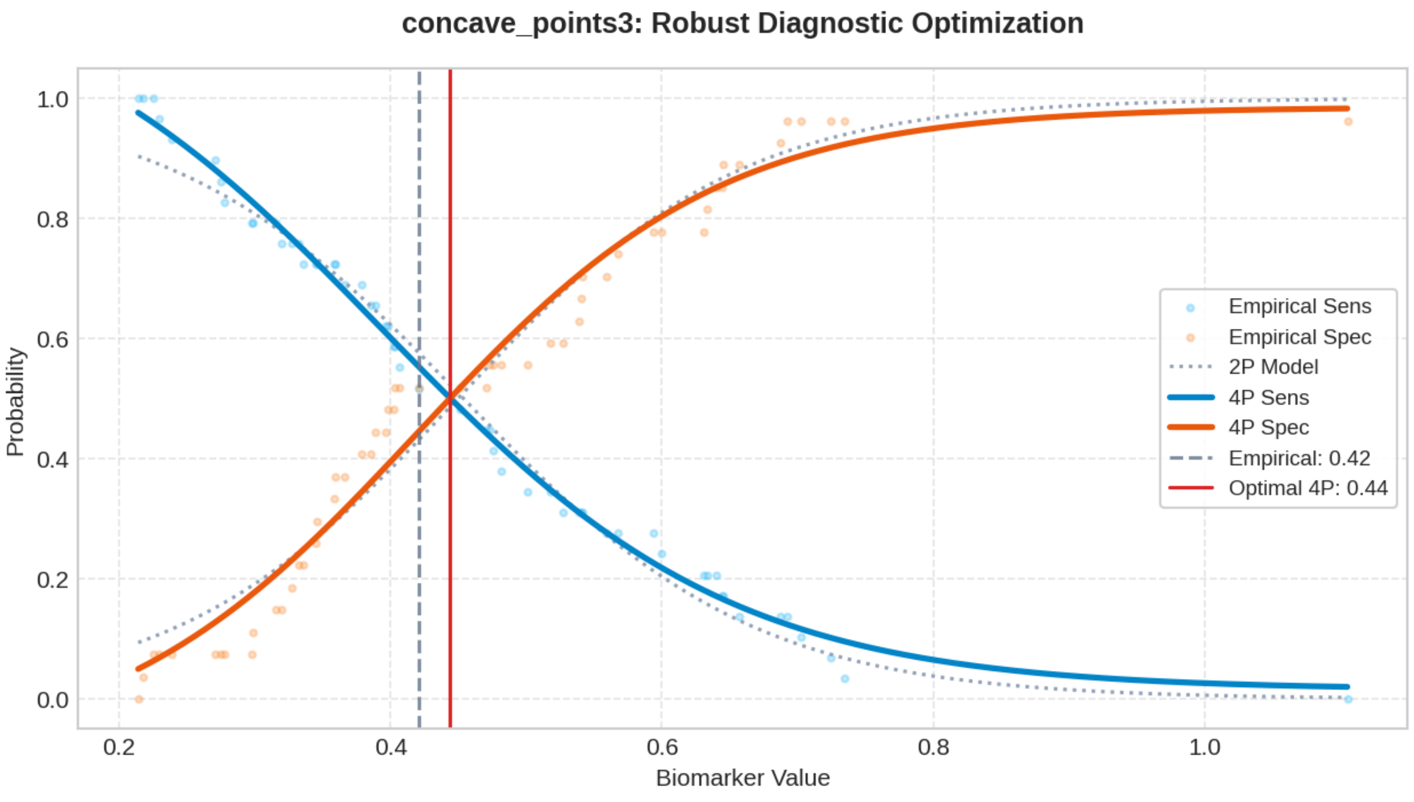

### Biomarker: symmetry3

Processed: 04-Apr-2026 18:40

#### 1. Optimization Results

| MODEL | CUT-OFF | TRAIN (SE/SP) | VAL (SE/SP) | TEST (SE/SP) | R2 SCORE |
|---|---|---|---|---|---|
| Empirical (Exact) | 0.1826 | 0.517 / 0.517 | 0.444 / 0.500 | 0.667 / 0.553 | N/A |
| Logistic 2-Parameter | 0.1827 | 0.525 / 0.525 | 0.444 / 0.500 | 0.667 / 0.570 | 0.9880 |
| **Logistic 4-Parameter (Rec.)** | **0.1826** | **0.520 / 0.520** | **0.444 / 0.500** | **0.667 / 0.553** | **0.9906** |
| ThresholdXpert (Stochastic) | 0.1819 | 0.517 / 0.519 | 0.444 / 0.500 | 0.667 / 0.535 | N/A |

#### 2. Diagnostic Performance Curves (Training)

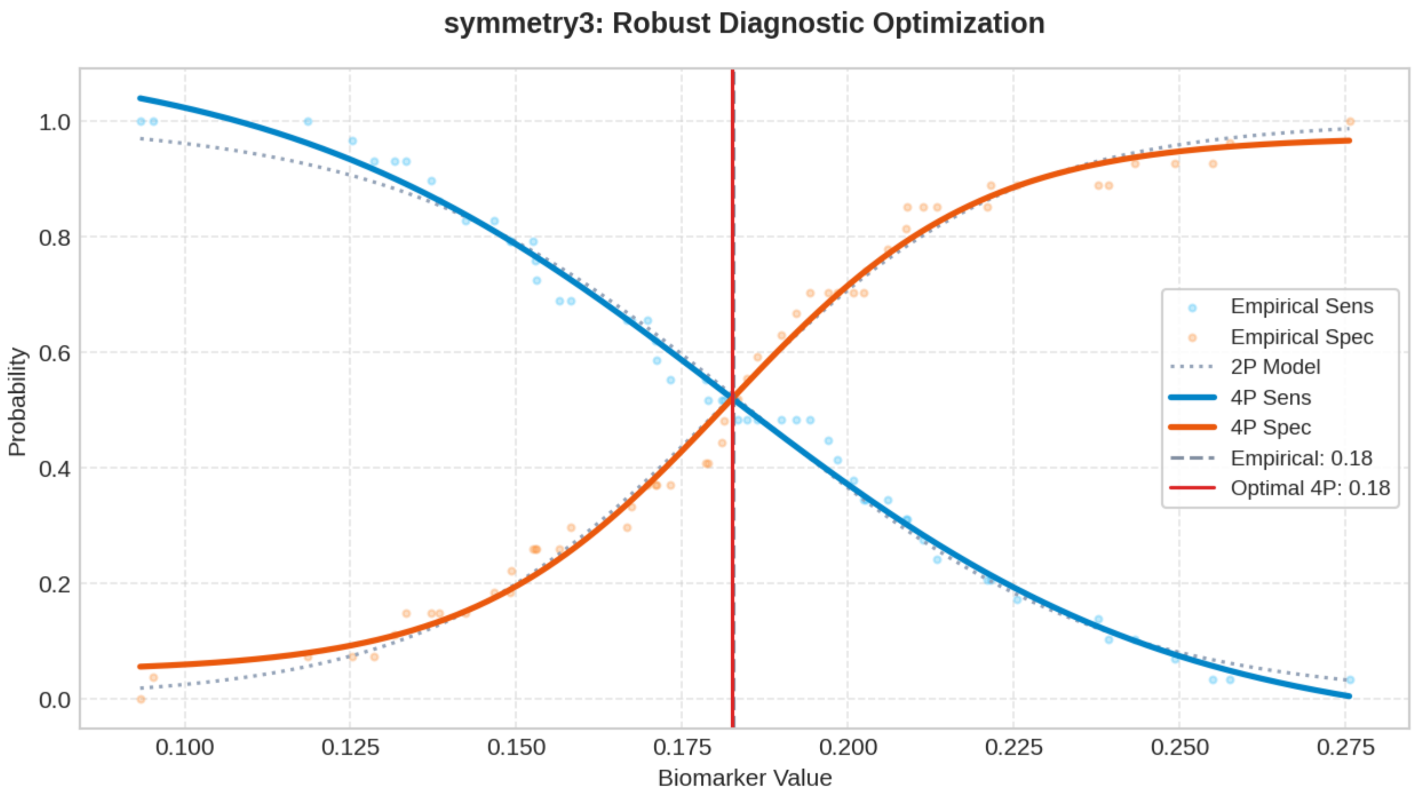

### Biomarker: fractal_dimension3

Processed: 04-Apr-2026 18:43

#### 1. Optimization Results

| MODEL | CUT-OFF | TRAIN (SE/SP) | VAL (SE/SP) | TEST (SE/SP) | R2 SCORE |
|---|---|---|---|---|---|
| Empirical (Exact) | 0.3279 | 0.334 / 0.334 | 0.333 / 0.500 | 0.222 / 0.675 | N/A |
| Logistic 2-Parameter | 0.3320 | 0.374 / 0.374 | 0.333 / 0.500 | 0.222 / 0.702 | 0.9736 |
| **Logistic 4-Parameter (Rec.)** | **0.3279** | **0.370 / 0.370** | **0.333 / 0.500** | **0.222 / 0.675** | **0.9898** |
| ThresholdXpert (Stochastic) | 0.3050 | 0.517 / 0.333 | 0.556 / 0.400 | 0.333 / 0.474 | N/A |

#### 2. Diagnostic Performance Curves (Training)

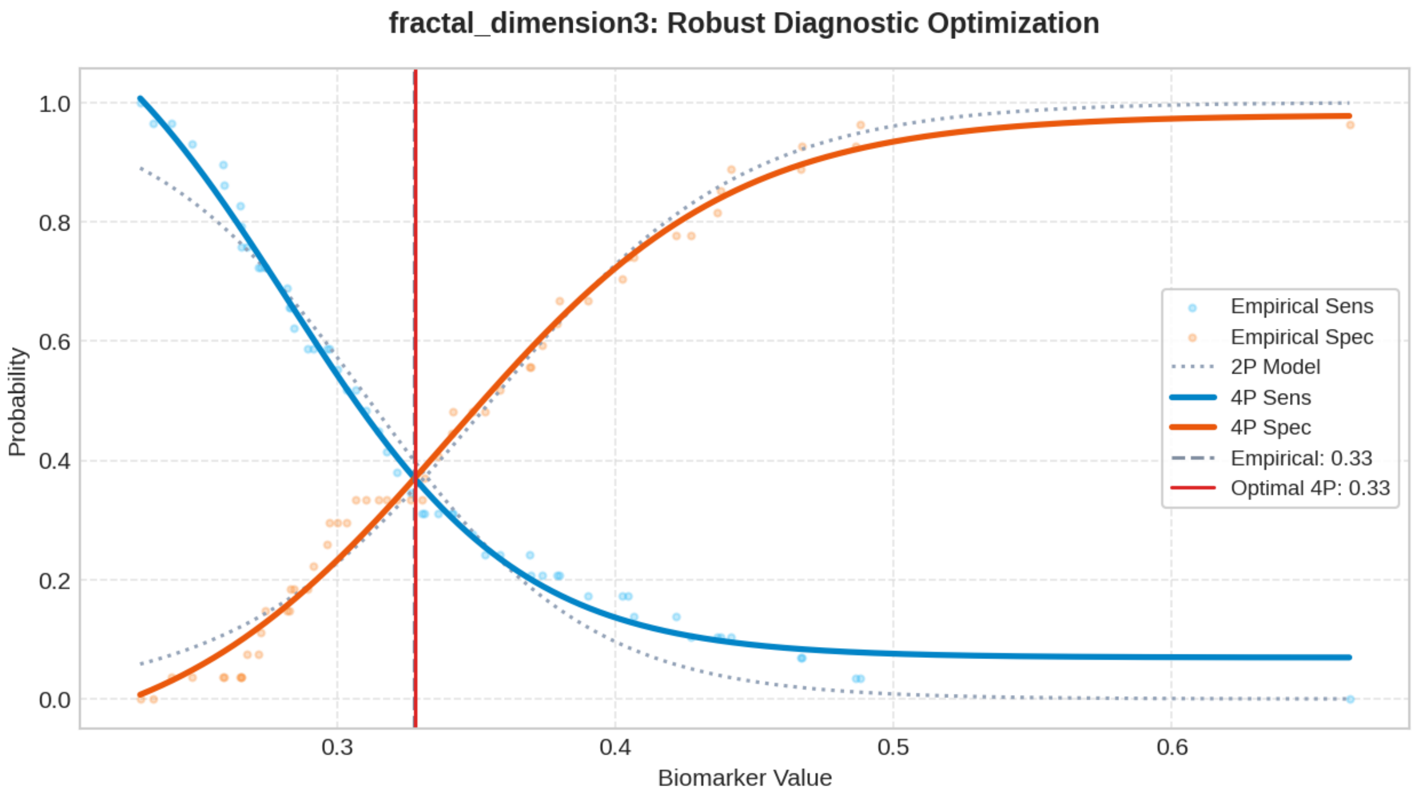

### Biomarker: tumor_size

Processed: 04-Apr-2026 18:45

#### 1. Optimization Results

| MODEL | CUT-OFF | TRAIN (SE/SP) | VAL (SE/SP) | TEST (SE/SP) | R2 SCORE |
|---|---|---|---|---|---|
| Empirical (Exact) | 0.0910 | 0.370 / 0.370 | 0.444 / 0.600 | 0.444 / 0.588 | N/A |
| Logistic 2-Parameter | 0.0923 | 0.373 / 0.373 | 0.333 / 0.600 | 0.333 / 0.623 | 0.9802 |
| **Logistic 4-Parameter (Rec.)** | **0.0919** | **0.375 / 0.375** | **0.444 / 0.600** | **0.444 / 0.605** | **0.9855** |
| ThresholdXpert (Stochastic) | 0.0872 | 0.517 / 0.370 | 0.444 / 0.400 | 0.556 / 0.553 | N/A |

#### 2. Diagnostic Performance Curves (Training)

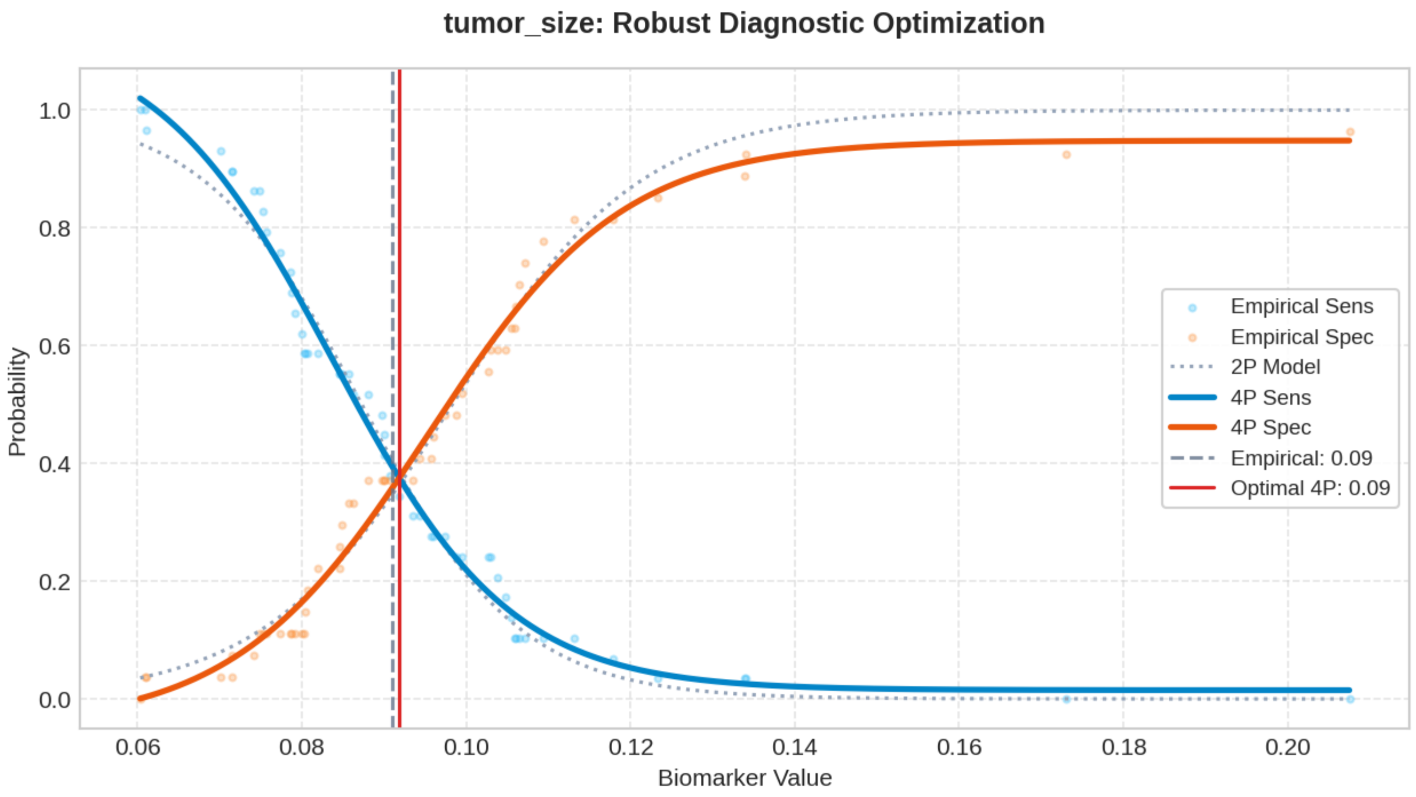

### Biomarker: lymph_node_status

Processed: 04-Apr-2026 18:46

#### 1. Optimization Results

| MODEL | CUT-OFF | TRAIN (SE/SP) | VAL (SE/SP) | TEST (SE/SP) | R2 SCORE |
|---|---|---|---|---|---|
| Empirical (Exact) | 2.5758 | 0.585 / 0.585 | 0.778 / 0.700 | 0.778 / 0.614 | N/A |
| Logistic 2-Parameter | 2.7671 | 0.588 / 0.588 | 0.778 / 0.700 | 0.778 / 0.632 | 0.9568 |
| **Logistic 4-Parameter (Rec.)** | **2.6396** | **0.591 / 0.591** | **0.778 / 0.700** | **0.778 / 0.623** | **0.9825** |
| ThresholdXpert (Stochastic) | 2.1773 | 0.724 / 0.556 | 0.778 / 0.500 | 0.889 / 0.518 | N/A |

#### 2. Diagnostic Performance Curves (Training)

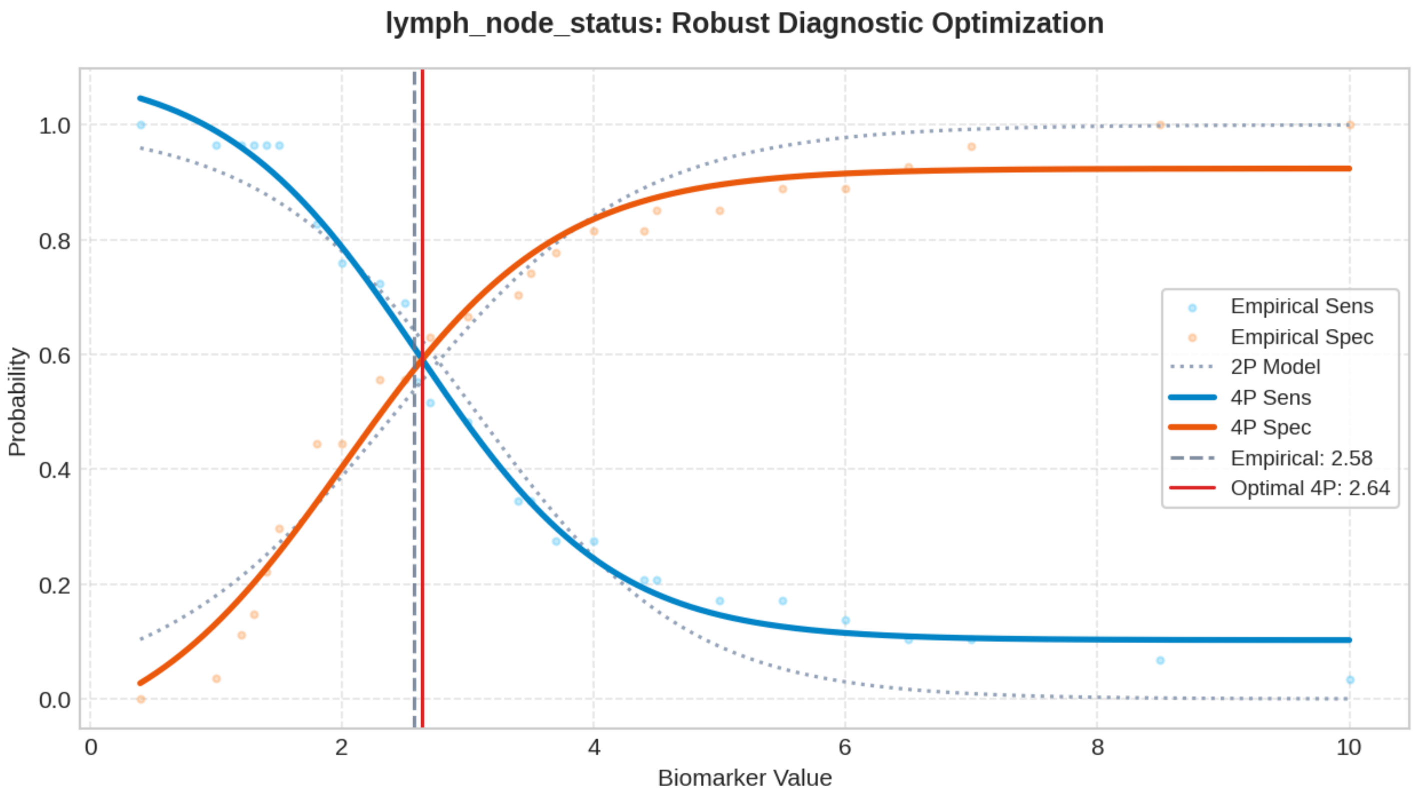

#### Top 200 Combinatorial Panels (ThresholdXpert OR-Logic)

The following multimarker panels have been optimized using high-performance vector-driven Monte Carlo simulations under a Boolean OR-logic framework. The engine employs a Max-Min Balancing logic (0.001 precision) to identify global threshold configurations that maximize the equilibrium between Sensitivity and Specificity across up to 10 million iterations. To ensure clinical robustness, results are sorted strictly by Validation Performance. (* Asterisk indicates an algorithmic threshold instability > 15%, suggesting potential data sparsity or high variance in the stochastic averaging process).

##### #1: concavity3 + fractal_dimension3 + lymph_node_status

Optimized Thresholds: concavity3: 0.6967 | fractal_dimension3: 0.5573 | lymph_node_status: 2.5587

Train Sens: 0.586 | Train Spec: 0.556  [TRAIN SCORE: 1.142]  ||  Val Sens: 0.778 | Val Spec: 0.700  [VAL SCORE: 1.478]  ||  Test Sens: 0.778 | Test Spec: 0.579  [TEST SCORE: 1.357]

##### #2: concavity3* + concave_points3 + fractal_dimension3 + lymph_node_status

Optimized Thresholds: concavity3*: 0.8765 | concave_points3: 0.6641 | fractal_dimension3: 0.5834 | lymph_node_status: 2.6845

Train Sens: 0.621 | Train Spec: 0.593  [TRAIN SCORE: 1.213]  ||  Val Sens: 0.778 | Val Spec: 0.700  [VAL SCORE: 1.478]  ||  Test Sens: 0.778 | Test Spec: 0.570  [TEST SCORE: 1.348]

##### #3: concavity3 + concave_points3 + lymph_node_status

Optimized Thresholds: concavity3: 0.9684 | concave_points3: 0.6894 | lymph_node_status: 2.6431

Train Sens: 0.621 | Train Spec: 0.630  [TRAIN SCORE: 1.250]  ||  Val Sens: 0.778 | Val Spec: 0.700  [VAL SCORE: 1.478]  ||  Test Sens: 0.778 | Test Spec: 0.588  [TEST SCORE: 1.365]

##### #4: concavity3 + concave_points3 + fractal_dimension3 + tumor_size* + lymph_node_status

Optimized Thresholds: concavity3: 0.8816 | concave_points3: 0.6765 | fractal_dimension3: 0.5738 | tumor_size*: 0.1785 | lymph_node_status: 2.6494

Train Sens: 0.621 | Train Spec: 0.593  [TRAIN SCORE: 1.213]  ||  Val Sens: 0.778 | Val Spec: 0.700  [VAL SCORE: 1.478]  ||  Test Sens: 0.778 | Test Spec: 0.561  [TEST SCORE: 1.339]

##### #5: concave_points3 + fractal_dimension3 + tumor_size* + lymph_node_status

Optimized Thresholds: concave_points3: 0.6594 | fractal_dimension3: 0.5836 | tumor_size*: 0.1636 | lymph_node_status: 2.6949

Train Sens: 0.621 | Train Spec: 0.593  [TRAIN SCORE: 1.213]  ||  Val Sens: 0.778 | Val Spec: 0.700  [VAL SCORE: 1.478]  ||  Test Sens: 0.778 | Test Spec: 0.561  [TEST SCORE: 1.339]

##### #6: fractal_dimension3 + lymph_node_status

Optimized Thresholds: fractal_dimension3: 0.5424 | lymph_node_status: 2.5756

Train Sens: 0.552 | Train Spec: 0.556  [TRAIN SCORE: 1.107]  ||  Val Sens: 0.778 | Val Spec: 0.700  [VAL SCORE: 1.478]  ||  Test Sens: 0.778 | Test Spec: 0.596  [TEST SCORE: 1.374]

##### #7: fractal_dimension3 + tumor_size* + lymph_node_status

Optimized Thresholds: fractal_dimension3: 0.5656 | tumor_size*: 0.1719 | lymph_node_status: 2.5800

Train Sens: 0.552 | Train Spec: 0.556  [TRAIN SCORE: 1.107]  ||  Val Sens: 0.778 | Val Spec: 0.700  [VAL SCORE: 1.478]  ||  Test Sens: 0.778 | Test Spec: 0.605  [TEST SCORE: 1.383]

##### #8: concavity3 + fractal_dimension3* + tumor_size + lymph_node_status

Optimized Thresholds: concavity3: 0.6819 | fractal_dimension3*: 0.5816 | tumor_size: 0.1639 | lymph_node_status: 2.5394

Train Sens: 0.586 | Train Spec: 0.556 [TRAIN SCORE: 1.142] || Val Sens: 0.778 | Val Spec: 0.700 [VAL SCORE: 1.478] || Test Sens: 0.778 | Test Spec: 0.588 [TEST SCORE: 1.365]

###### #9: concave_points3 + lymph_node_status

Optimized Thresholds: concave_points3: 0.6899 | lymph_node_status: 2.6494

Train Sens: 0.621 | Train Spec: 0.630 [TRAIN SCORE: 1.250] || Val Sens: 0.778 | Val Spec: 0.700 [VAL SCORE: 1.478] || Test Sens: 0.778 | Test Spec: 0.588 [TEST SCORE: 1.365]

###### #10: concave_points3 + tumor_size + lymph_node_status

Optimized Thresholds: concave_points3: 0.6897 | tumor_size: 0.1885 | lymph_node_status: 2.6540

Train Sens: 0.621 | Train Spec: 0.630 [TRAIN SCORE: 1.250] || Val Sens: 0.778 | Val Spec: 0.700 [VAL SCORE: 1.478] || Test Sens: 0.778 | Test Spec: 0.588 [TEST SCORE: 1.365]

###### #11: concavity3 + concave_points3 + tumor_size + lymph_node_status

Optimized Thresholds: concavity3: 0.9366 | concave_points3: 0.6901 | tumor_size: 0.1916 | lymph_node_status: 2.6472

Train Sens: 0.621 | Train Spec: 0.630 [TRAIN SCORE: 1.250] || Val Sens: 0.778 | Val Spec: 0.700 [VAL SCORE: 1.478] || Test Sens: 0.778 | Test Spec: 0.588 [TEST SCORE: 1.365]

###### #12: concave_points3 + fractal_dimension3 + lymph_node_status

Optimized Thresholds: concave_points3: 0.6605 | fractal_dimension3: 0.5884 | lymph_node_status: 2.6831

Train Sens: 0.621 | Train Spec: 0.593 [TRAIN SCORE: 1.213] || Val Sens: 0.778 | Val Spec: 0.700 [VAL SCORE: 1.478] || Test Sens: 0.778 | Test Spec: 0.570 [TEST SCORE: 1.348]

###### #13: smoothness3 + concavity3 + symmetry3 + fractal_dimension3 + tumor_size + lymph_node_status

Optimized Thresholds: smoothness3: 1412.6295 | concavity3: 0.8208 | symmetry3: 0.2566 | fractal_dimension3: 0.6003 | tumor_size: 0.1110 | lymph_node_status: 8.4730

Train Sens: 0.690 | Train Spec: 0.667 [TRAIN SCORE: 1.356] || Val Sens: 0.889 | Val Spec: 0.500 [VAL SCORE: 1.389] || Test Sens: 0.889 | Test Spec: 0.465 [TEST SCORE: 1.354]

###### #14: smoothness3 + symmetry3 + tumor_size + lymph_node_status

Optimized Thresholds: smoothness3: 1412.1093 | symmetry3: 0.2610 | tumor_size: 0.1114 | lymph_node_status: 8.7510

Train Sens: 0.690 | Train Spec: 0.667 [TRAIN SCORE: 1.356] || Val Sens: 0.889 | Val Spec: 0.500 [VAL SCORE: 1.389] || Test Sens: 0.889 | Test Spec: 0.474 [TEST SCORE: 1.363]

###### #15: smoothness3 + symmetry3 + fractal_dimension3

Optimized Thresholds: smoothness3: 1283.1033 | symmetry3: 0.2547 | fractal_dimension3: 0.4871

Train Sens: 0.655 | Train Spec: 0.630 [TRAIN SCORE: 1.285] || Val Sens: 0.778 | Val Spec: 0.600 [VAL SCORE: 1.378] || Test Sens: 0.889 | Test Spec: 0.465 [TEST SCORE: 1.354]

###### #16: smoothness3 + symmetry3

Optimized Thresholds: smoothness3: 1283.4400 | symmetry3: 0.2650

Train Sens: 0.621 | Train Spec: 0.667 [TRAIN SCORE: 1.287] || Val Sens: 0.778 | Val Spec: 0.600 [VAL SCORE: 1.378] || Test Sens: 0.889 | Test Spec: 0.491 [TEST SCORE: 1.380]

###### #17: smoothness3 + fractal_dimension3

Optimized Thresholds: smoothness3: 1283.5067 | fractal_dimension3: 0.4874

Train Sens: 0.655 | Train Spec: 0.630 [TRAIN SCORE: 1.285] || Val Sens: 0.778 | Val Spec: 0.600 [VAL SCORE: 1.378] || Test Sens: 0.889 | Test Spec: 0.482 [TEST SCORE: 1.371]

###### #18: area3 + concavity3 + concave_points3 + symmetry3 + tumor_size + lymph_node_status

Optimized Thresholds: area3: 154.8575 | concavity3: 0.9627 | concave_points3: 0.6894 | symmetry3: 0.2647 | tumor_size: 0.1799 | lymph_node_status: 2.7952

Train Sens: 0.724 | Train Spec: 0.667 [TRAIN SCORE: 1.391] || Val Sens: 0.778 | Val Spec: 0.600 [VAL SCORE: 1.378] || Test Sens: 1.000 | Test Spec: 0.439 [TEST SCORE: 1.439]

###### #19: area3 + concavity3 + symmetry3 + fractal_dimension3 + tumor_size + lymph_node_status

Optimized Thresholds: area3: 136.5019 | concavity3: 0.6488 | symmetry3: 0.2610 | fractal_dimension3: 0.4875 | tumor_size: 0.1920 | lymph_node_status: 7.0142

Train Sens: 0.690 | Train Spec: 0.667 [TRAIN SCORE: 1.356] || Val Sens: 0.778 | Val Spec: 0.600 [VAL SCORE: 1.378] || Test Sens: 0.889 | Test Spec: 0.447 [TEST SCORE: 1.336]

###### #20: texture1 + concavity3 + concave_points3 + symmetry3 + tumor_size*

Optimized Thresholds: texture1: 17.1796 | concavity3: 0.8060 | concave_points3: 0.6641 | symmetry3: 0.2629 | tumor_size*: 0.1763

Train Sens: 0.724 | Train Spec: 0.741 [TRAIN SCORE: 1.465] || Val Sens: 0.667 | Val Spec: 0.700 [VAL SCORE: 1.367] || Test Sens: 0.889 | Test Spec: 0.395 [TEST SCORE: 1.284]

###### #21: texture1 + concavity3 + concave_points3 + fractal_dimension3 + tumor_size*

Optimized Thresholds: texture1: 17.1813 | concavity3: 0.8913 | concave_points3: 0.6718 | fractal_dimension3: 0.5468 | tumor_size*: 0.1506

Train Sens: 0.724 | Train Spec: 0.741 [TRAIN SCORE: 1.465] || Val Sens: 0.667 | Val Spec: 0.700 [VAL SCORE: 1.367] || Test Sens: 0.889 | Test Spec: 0.395 [TEST SCORE: 1.284]

###### #22: texture1 + concavity3 + concave_points3 + fractal_dimension3 + lymph_node_status

Optimized Thresholds: texture1: 17.2474 | concavity3: 0.8738 | concave_points3: 0.6703 | fractal_dimension3: 0.5477 | lymph_node_status: 7.6367

Train Sens: 0.724 | Train Spec: 0.778 [TRAIN SCORE: 1.502] || Val Sens: 0.667 | Val Spec: 0.700 [VAL SCORE: 1.367] || Test Sens: 0.889 | Test Spec: 0.377 [TEST SCORE: 1.266]

###### #23: texture1 + concavity3 + concave_points3 + symmetry3 + lymph_node_status

Optimized Thresholds: texture1: 17.2484 | concavity3: 0.9143 | concave_points3: 0.6745 | symmetry3: 0.2637 | lymph_node_status: 7.3793

Train Sens: 0.724 | Train Spec: 0.778 [TRAIN SCORE: 1.502] || Val Sens: 0.667 | Val Spec: 0.700 [VAL SCORE: 1.367] || Test Sens: 0.889 | Test Spec: 0.377 [TEST SCORE: 1.266]

###### #24: texture1 + concavity3 + concave_points3 + symmetry3 + fractal_dimension3 + lymph_node_status

Optimized Thresholds: texture1: 17.2422 | concavity3: 0.9106 | concave_points3: 0.6735 | symmetry3: 0.2622 | fractal_dimension3: 0.5617 | lymph_node_status: 7.2787

Train Sens: 0.724 | Train Spec: 0.778 [TRAIN SCORE: 1.502] || Val Sens: 0.667 | Val Spec: 0.700 [VAL SCORE: 1.367] || Test Sens: 0.889 | Test Spec: 0.368 [TEST SCORE: 1.257]

**#25: texture1 + concavity3 + concave_points3 + symmetry3 + fractal_dimension3**

Optimized Thresholds: texture1: 17.1814 | concavity3: 0.8366 | concave_points3: 0.6660 | symmetry3: 0.2621 | fractal_dimension3: 0.6169

Train Sens: 0.724 | Train Spec: 0.741 [TRAIN SCORE: 1.465] || Val Sens: 0.667 | Val Spec: 0.700 [VAL SCORE: 1.367] || Test Sens: 0.889 | Test Spec: 0.395 [TEST SCORE: 1.284]

**#26: texture1 + area3* + concave_points3 + symmetry3 + fractal_dimension3 + lymph_node_status**

Optimized Thresholds: texture1: 17.3024 | area3*: 187.5069 | concave_points3: 0.6747 | symmetry3: 0.2622 | fractal_dimension3: 0.5525 | lymph_node_status: 7.3889

Train Sens: 0.690 | Train Spec: 0.778 [TRAIN SCORE: 1.467] || Val Sens: 0.667 | Val Spec: 0.700 [VAL SCORE: 1.367] || Test Sens: 0.889 | Test Spec: 0.386 [TEST SCORE: 1.275]

**#27: texture1 + concavity3 + symmetry3 + fractal_dimension3 + tumor_size**

Optimized Thresholds: texture1: 17.1787 | concavity3: 0.6189 | symmetry3: 0.2642 | fractal_dimension3: 0.5640 | tumor_size: 0.1564

Train Sens: 0.724 | Train Spec: 0.704 [TRAIN SCORE: 1.428] || Val Sens: 0.667 | Val Spec: 0.700 [VAL SCORE: 1.367] || Test Sens: 0.889 | Test Spec: 0.412 [TEST SCORE: 1.301]

**#28: area3 + tumor_size* + lymph_node_status***

Optimized Thresholds: area3: 140.4909 | tumor_size*: 0.1756 | lymph_node_status*: 5.1528

Train Sens: 0.586 | Train Spec: 0.704 [TRAIN SCORE: 1.290] || Val Sens: 0.667 | Val Spec: 0.700 [VAL SCORE: 1.367] || Test Sens: 0.889 | Test Spec: 0.500 [TEST SCORE: 1.389]

**#29: area3 + symmetry3 + tumor_size* + lymph_node_status***

Optimized Thresholds: area3: 141.2093 | symmetry3: 0.2624 | tumor_size*: 0.1598 | lymph_node_status*: 5.7929

Train Sens: 0.586 | Train Spec: 0.667 [TRAIN SCORE: 1.253] || Val Sens: 0.667 | Val Spec: 0.700 [VAL SCORE: 1.367] || Test Sens: 0.889 | Test Spec: 0.518 [TEST SCORE: 1.406]

**#30: texture1 + concavity3 + concave_points3 + symmetry3 + fractal_dimension3 + tumor_size***

Optimized Thresholds: texture1: 17.1837 | concavity3: 0.8727 | concave_points3: 0.6712 | symmetry3: 0.2671 | fractal_dimension3: 0.5614 | tumor_size*: 0.1543

Train Sens: 0.724 | Train Spec: 0.741 [TRAIN SCORE: 1.465] || Val Sens: 0.667 | Val Spec: 0.700 [VAL SCORE: 1.367] || Test Sens: 0.889 | Test Spec: 0.386 [TEST SCORE: 1.275]

**#31: texture1 + concave_points3 + fractal_dimension3 + tumor_size + lymph_node_status**

Optimized Thresholds: texture1: 17.2484 | concave_points3: 0.6738 | fractal_dimension3: 0.5807 | tumor_size: 0.1723 | lymph_node_status: 7.4501

Train Sens: 0.724 | Train Spec: 0.778 [TRAIN SCORE: 1.502] || Val Sens: 0.667 | Val Spec: 0.700 [VAL SCORE: 1.367] || Test Sens: 0.889 | Test Spec: 0.386 [TEST SCORE: 1.275]

**#32: texture1 + concavity3 + concave_points3 + symmetry3 + tumor_size + lymph_node_status**

Optimized Thresholds: texture1: 17.2398 | concavity3: 0.8990 | concave_points3: 0.6743 | symmetry3: 0.2620 | tumor_size: 0.1697 | lymph_node_status: 7.3349

Train Sens: 0.724 | Train Spec: 0.778 [TRAIN SCORE: 1.502] || Val Sens: 0.667 | Val Spec: 0.700 [VAL SCORE: 1.367] || Test Sens: 0.889 | Test Spec: 0.377 [TEST SCORE: 1.266]

###### #33: texture1 + concavity3 + concave_points3 + fractal_dimension3 + tumor_size + lymph_node_status

Optimized Thresholds: texture1: 17.2322 | concavity3: 0.9095 | concave_points3: 0.6751 | fractal_dimension3: 0.5733 | tumor_size: 0.1790 | lymph_node_status: 7.5369

Train Sens: 0.724 | Train Spec: 0.778 [TRAIN SCORE: 1.502] || Val Sens: 0.667 | Val Spec: 0.700 [VAL SCORE: 1.367] || Test Sens: 0.889 | Test Spec: 0.377 [TEST SCORE: 1.266]

###### #34: texture1 + concavity3 + symmetry3 + fractal_dimension3 + tumor_size* + lymph_node_status

Optimized Thresholds: texture1: 17.2284 | concavity3: 0.6162 | symmetry3: 0.2672 | fractal_dimension3: 0.5602 | tumor_size*: 0.1719 | lymph_node_status: 7.7056

Train Sens: 0.724 | Train Spec: 0.704 [TRAIN SCORE: 1.428] || Val Sens: 0.667 | Val Spec: 0.700 [VAL SCORE: 1.367] || Test Sens: 0.889 | Test Spec: 0.395 [TEST SCORE: 1.284]

###### #35: texture1 + concave_points3 + symmetry3 + fractal_dimension3 + tumor_size + lymph_node_status

Optimized Thresholds: texture1: 17.2503 | concave_points3: 0.6770 | symmetry3: 0.2670 | fractal_dimension3: 0.5769 | tumor_size: 0.1725 | lymph_node_status: 7.2562

Train Sens: 0.724 | Train Spec: 0.778 [TRAIN SCORE: 1.502] || Val Sens: 0.667 | Val Spec: 0.700 [VAL SCORE: 1.367] || Test Sens: 0.889 | Test Spec: 0.368 [TEST SCORE: 1.257]

###### #36: texture1 + concavity3 + concave_points3 + tumor_size + lymph_node_status

Optimized Thresholds: texture1: 17.2569 | concavity3: 0.9483 | concave_points3: 0.6877 | tumor_size: 0.1827 | lymph_node_status: 8.1834

Train Sens: 0.724 | Train Spec: 0.815 [TRAIN SCORE: 1.539] || Val Sens: 0.667 | Val Spec: 0.700 [VAL SCORE: 1.367] || Test Sens: 0.889 | Test Spec: 0.404 [TEST SCORE: 1.292]

###### #37: texture1 + concave_points3 + symmetry3 + fractal_dimension3 + tumor_size*

Optimized Thresholds: texture1: 17.1824 | concave_points3: 0.6755 | symmetry3: 0.2631 | fractal_dimension3: 0.6047 | tumor_size*: 0.1745

Train Sens: 0.724 | Train Spec: 0.741 [TRAIN SCORE: 1.465] || Val Sens: 0.667 | Val Spec: 0.700 [VAL SCORE: 1.367] || Test Sens: 0.889 | Test Spec: 0.395 [TEST SCORE: 1.284]

###### #38: texture1 + concavity3 + symmetry3 + fractal_dimension3 + lymph_node_status

Optimized Thresholds: texture1: 17.2390 | concavity3: 0.6184 | symmetry3: 0.2624 | fractal_dimension3: 0.5849 | lymph_node_status: 7.4099

Train Sens: 0.724 | Train Spec: 0.704 [TRAIN SCORE: 1.428] || Val Sens: 0.667 | Val Spec: 0.700 [VAL SCORE: 1.367] || Test Sens: 0.889 | Test Spec: 0.404 [TEST SCORE: 1.292]

###### #39: texture1 + concavity3 + tumor_size* + lymph_node_status

Optimized Thresholds: texture1: 17.2378 | concavity3: 0.6178 | tumor_size*: 0.1653 | lymph_node_status: 7.5187

Train Sens: 0.724 | Train Spec: 0.704 [TRAIN SCORE: 1.428] || Val Sens: 0.667 | Val Spec: 0.700 [VAL SCORE: 1.367] || Test Sens: 0.889 | Test Spec: 0.404 [TEST SCORE: 1.292]

###### #40: texture1 + concavity3 + concave_points3 + symmetry3

Optimized Thresholds: texture1: 17.1780 | concavity3: 0.9934 | concave_points3: 0.6918 | symmetry3: 0.2725

Train Sens: 0.724 | Train Spec: 0.778 [TRAIN SCORE: 1.502] || Val Sens: 0.667 | Val Spec: 0.700 [VAL SCORE: 1.367] || Test Sens: 0.889 | Test Spec: 0.412 [TEST SCORE: 1.301]

##### #41: texture1 + concavity3 + concave_points3 + fractal_dimension3

Optimized Thresholds: texture1: 17.1814 | concavity3: 0.8623 | concave_points3: 0.6713 | fractal_dimension3: 0.5459

Train Sens: 0.724 | Train Spec: 0.741 [TRAIN SCORE: 1.465] || Val Sens: 0.667 | Val Spec: 0.700 [VAL SCORE: 1.367] || Test Sens: 0.889 | Test Spec: 0.395 [TEST SCORE: 1.284]

##### #42: texture1 + concavity3 + concave_points3 + tumor_size

Optimized Thresholds: texture1: 17.1823 | concavity3: 0.9952 | concave_points3: 0.6897 | tumor_size: 0.1870

Train Sens: 0.724 | Train Spec: 0.778 [TRAIN SCORE: 1.502] || Val Sens: 0.667 | Val Spec: 0.700 [VAL SCORE: 1.367] || Test Sens: 0.889 | Test Spec: 0.412 [TEST SCORE: 1.301]

##### #43: texture1 + concavity3 + concave_points3 + lymph_node_status

Optimized Thresholds: texture1: 17.2475 | concavity3: 0.9516 | concave_points3: 0.6893 | lymph_node_status: 7.6152

Train Sens: 0.724 | Train Spec: 0.815 [TRAIN SCORE: 1.539] || Val Sens: 0.667 | Val Spec: 0.700 [VAL SCORE: 1.367] || Test Sens: 0.889 | Test Spec: 0.395 [TEST SCORE: 1.284]

##### #44: texture1 + concavity3 + symmetry3 + fractal_dimension3

Optimized Thresholds: texture1: 17.1791 | concavity3: 0.6187 | symmetry3: 0.2665 | fractal_dimension3: 0.5628

Train Sens: 0.724 | Train Spec: 0.704 [TRAIN SCORE: 1.428] || Val Sens: 0.667 | Val Spec: 0.700 [VAL SCORE: 1.367] || Test Sens: 0.889 | Test Spec: 0.412 [TEST SCORE: 1.301]

##### #45: texture1 + concavity3 + symmetry3 + tumor_size*

Optimized Thresholds: texture1: 17.1799 | concavity3: 0.6186 | symmetry3: 0.2667 | tumor_size*: 0.1604

Train Sens: 0.724 | Train Spec: 0.704 [TRAIN SCORE: 1.428] || Val Sens: 0.667 | Val Spec: 0.700 [VAL SCORE: 1.367] || Test Sens: 0.889 | Test Spec: 0.421 [TEST SCORE: 1.310]

##### #46: texture1 + concavity3 + symmetry3 + lymph_node_status

Optimized Thresholds: texture1: 17.2374 | concavity3: 0.6171 | symmetry3: 0.2661 | lymph_node_status: 7.5375

Train Sens: 0.724 | Train Spec: 0.704 [TRAIN SCORE: 1.428] || Val Sens: 0.667 | Val Spec: 0.700 [VAL SCORE: 1.367] || Test Sens: 0.889 | Test Spec: 0.404 [TEST SCORE: 1.292]

##### #47: area3 + concavity3 + symmetry3 + tumor_size* + lymph_node_status*

Optimized Thresholds: area3: 140.8536 | concavity3: 0.8502 | symmetry3: 0.2610 | tumor_size*: 0.1574 | lymph_node_status*: 6.0309

Train Sens: 0.552 | Train Spec: 0.667 [TRAIN SCORE: 1.218] || Val Sens: 0.667 | Val Spec: 0.700 [VAL SCORE: 1.367] || Test Sens: 0.889 | Test Spec: 0.482 [TEST SCORE: 1.371]

##### #48: texture1 + concavity3 + fractal_dimension3 + tumor_size

Optimized Thresholds: texture1: 17.1834 | concavity3: 0.6171 | fractal_dimension3: 0.5523 | tumor_size: 0.1719

Train Sens: 0.724 | Train Spec: 0.704 [TRAIN SCORE: 1.428] || Val Sens: 0.667 | Val Spec: 0.700 [VAL SCORE: 1.367] || Test Sens: 0.889 | Test Spec: 0.412 [TEST SCORE: 1.301]

##### #49: texture1 + concavity3 + fractal_dimension3 + lymph_node_status

Optimized Thresholds: texture1: 17.2205 | concavity3: 0.6187 | fractal_dimension3: 0.5806 | lymph_node_status: 7.4400

Train Sens: 0.724 | Train Spec: 0.704 [TRAIN SCORE: 1.428] || Val Sens: 0.667 | Val Spec: 0.700 [VAL SCORE: 1.367] || Test Sens: 0.889 | Test Spec: 0.404 [TEST SCORE: 1.292]

##### #50: texture1 + concave_points3 + symmetry3 + fractal_dimension3

Optimized Thresholds: texture1: 17.1780 | concave_points3: 0.6672 | symmetry3: 0.2650 | fractal_dimension3: 0.5829

Train Sens: 0.724 | Train Spec: 0.741 [TRAIN SCORE: 1.465] || Val Sens: 0.667 | Val Spec: 0.700 [VAL SCORE: 1.367] || Test Sens: 0.889 | Test Spec: 0.395 [TEST SCORE: 1.284]

##### #51: texture1 + concavity3 + symmetry3 + tumor_size* + lymph_node_status

Optimized Thresholds: texture1: 17.2342 | concavity3: 0.6195 | symmetry3: 0.2644 | tumor_size*: 0.1692 | lymph_node_status: 7.7219

Train Sens: 0.724 | Train Spec: 0.704 [TRAIN SCORE: 1.428] || Val Sens: 0.667 | Val Spec: 0.700 [VAL SCORE: 1.367] || Test Sens: 0.889 | Test Spec: 0.404 [TEST SCORE: 1.292]

##### #52: texture1 + concave_points3 + symmetry3 + tumor_size*

Optimized Thresholds: texture1: 17.1782 | concave_points3: 0.6627 | symmetry3: 0.2665 | tumor_size*: 0.1756

Train Sens: 0.724 | Train Spec: 0.741 [TRAIN SCORE: 1.465] || Val Sens: 0.667 | Val Spec: 0.700 [VAL SCORE: 1.367] || Test Sens: 0.889 | Test Spec: 0.395 [TEST SCORE: 1.284]

##### #53: texture1 + concave_points3 + symmetry3 + lymph_node_status

Optimized Thresholds: texture1: 17.2543 | concave_points3: 0.6898 | symmetry3: 0.2673 | lymph_node_status: 7.6539

Train Sens: 0.724 | Train Spec: 0.815 [TRAIN SCORE: 1.539] || Val Sens: 0.667 | Val Spec: 0.700 [VAL SCORE: 1.367] || Test Sens: 0.889 | Test Spec: 0.386 [TEST SCORE: 1.275]

##### #54: texture1 + concave_points3 + fractal_dimension3 + tumor_size*

Optimized Thresholds: texture1: 17.1810 | concave_points3: 0.6686 | fractal_dimension3: 0.5976 | tumor_size*: 0.1736

Train Sens: 0.724 | Train Spec: 0.741 [TRAIN SCORE: 1.465] || Val Sens: 0.667 | Val Spec: 0.700 [VAL SCORE: 1.367] || Test Sens: 0.889 | Test Spec: 0.404 [TEST SCORE: 1.292]

##### #55: texture1 + concave_points3 + fractal_dimension3 + lymph_node_status

Optimized Thresholds: texture1: 17.2558 | concave_points3: 0.6724 | fractal_dimension3: 0.5730 | lymph_node_status: 7.9344

Train Sens: 0.724 | Train Spec: 0.778 [TRAIN SCORE: 1.502] || Val Sens: 0.667 | Val Spec: 0.700 [VAL SCORE: 1.367] || Test Sens: 0.889 | Test Spec: 0.377 [TEST SCORE: 1.266]

##### #56: texture1 + concave_points3 + tumor_size + lymph_node_status

Optimized Thresholds: texture1: 17.2437 | concave_points3: 0.6897 | tumor_size: 0.1906 | lymph_node_status: 7.4064

Train Sens: 0.724 | Train Spec: 0.815 [TRAIN SCORE: 1.539] || Val Sens: 0.667 | Val Spec: 0.700 [VAL SCORE: 1.367] || Test Sens: 0.889 | Test Spec: 0.395 [TEST SCORE: 1.284]

##### #57: texture1 + concave_points3 + symmetry3 + fractal_dimension3 + lymph_node_status

Optimized Thresholds: texture1: 17.2405 | concave_points3: 0.6719 | symmetry3: 0.2648 | fractal_dimension3: 0.5568 | lymph_node_status: 7.3476

Train Sens: 0.724 | Train Spec: 0.778 [TRAIN SCORE: 1.502] || Val Sens: 0.667 | Val Spec: 0.700 [VAL SCORE: 1.367] || Test Sens: 0.889 | Test Spec: 0.368 [TEST SCORE: 1.257]

##### #58: texture1 + symmetry3 + fractal_dimension3 + lymph_node_status

Optimized Thresholds: texture1: 17.2603 | symmetry3: 0.2633 | fractal_dimension3: 0.5850 | lymph_node_status: 5.4796

Train Sens: 0.690 | Train Spec: 0.741 [TRAIN SCORE: 1.430] || Val Sens: 0.667 | Val Spec: 0.700 [VAL SCORE: 1.367] || Test

Sens: 0.889 | Test Spec: 0.430 [TEST SCORE: 1.319]

###### #59: texture1 + symmetry3 + tumor_size + lymph_node_status

Optimized Thresholds: texture1: 17.2441 | symmetry3: 0.2668 | tumor_size: 0.1889 | lymph_node_status: 5.5565

Train Sens: 0.690 | Train Spec: 0.778 [TRAIN SCORE: 1.467] || Val Sens: 0.667 | Val Spec: 0.700 [VAL SCORE: 1.367] || Test Sens: 0.889 | Test Spec: 0.430 [TEST SCORE: 1.319]

###### #60: texture1 + fractal_dimension3 + lymph_node_status

Optimized Thresholds: texture1: 17.2357 | fractal_dimension3: 0.5460 | lymph_node_status: 5.4447

Train Sens: 0.690 | Train Spec: 0.741 [TRAIN SCORE: 1.430] || Val Sens: 0.667 | Val Spec: 0.700 [VAL SCORE: 1.367] || Test Sens: 0.889 | Test Spec: 0.430 [TEST SCORE: 1.319]

###### #61: texture1 + concavity3 + fractal_dimension3 + tumor_size + lymph_node_status

Optimized Thresholds: texture1: 17.2258 | concavity3: 0.6193 | fractal_dimension3: 0.5775 | tumor_size: 0.1627 | lymph_node_status: 7.6292

Train Sens: 0.724 | Train Spec: 0.704 [TRAIN SCORE: 1.428] || Val Sens: 0.667 | Val Spec: 0.700 [VAL SCORE: 1.367] || Test Sens: 0.889 | Test Spec: 0.404 [TEST SCORE: 1.292]

###### #62: texture1 + tumor_size + lymph_node_status

Optimized Thresholds: texture1: 17.2307 | tumor_size: 0.1934 | lymph_node_status: 5.4497

Train Sens: 0.690 | Train Spec: 0.778 [TRAIN SCORE: 1.467] || Val Sens: 0.667 | Val Spec: 0.700 [VAL SCORE: 1.367] || Test Sens: 0.889 | Test Spec: 0.439 [TEST SCORE: 1.327]

###### #63: texture1 + concave_points3 + symmetry3 + tumor_size + lymph_node_status

Optimized Thresholds: texture1: 17.2628 | concave_points3: 0.6890 | symmetry3: 0.2681 | tumor_size: 0.1954 | lymph_node_status: 7.5213

Train Sens: 0.724 | Train Spec: 0.815 [TRAIN SCORE: 1.539] || Val Sens: 0.667 | Val Spec: 0.700 [VAL SCORE: 1.367] || Test Sens: 0.889 | Test Spec: 0.386 [TEST SCORE: 1.275]

###### #64: texture1 + concavity3 + concave_points3 + symmetry3 + fractal_dimension3 + tumor_size + lymph_node_status

Optimized Thresholds: texture1: 17.2400 | concavity3: 0.8581 | concave_points3: 0.6779 | symmetry3: 0.2641 | fractal_dimension3: 0.5702 | tumor_size: 0.1905 | lymph_node_status: 7.6229

Train Sens: 0.724 | Train Spec: 0.778 [TRAIN SCORE: 1.502] || Val Sens: 0.667 | Val Spec: 0.700 [VAL SCORE: 1.367] || Test Sens: 0.889 | Test Spec: 0.368 [TEST SCORE: 1.257]

###### #65: texture1 + concave_points3 + fractal_dimension3

Optimized Thresholds: texture1: 17.1820 | concave_points3: 0.6651 | fractal_dimension3: 0.6009

Train Sens: 0.724 | Train Spec: 0.741 [TRAIN SCORE: 1.465] || Val Sens: 0.667 | Val Spec: 0.700 [VAL SCORE: 1.367] || Test Sens: 0.889 | Test Spec: 0.404 [TEST SCORE: 1.292]

###### #66: texture1 + symmetry3 + lymph_node_status

Optimized Thresholds: texture1: 17.2514 | symmetry3: 0.2640 | lymph_node_status: 5.5558

Train Sens: 0.690 | Train Spec: 0.778 [TRAIN SCORE: 1.467] || Val Sens: 0.667 | Val Spec: 0.700 [VAL SCORE: 1.367] || Test Sens: 0.889 | Test Spec: 0.430 [TEST SCORE: 1.319]

##### #67: texture1 + concavity3 + symmetry3

Optimized Thresholds: texture1: 17.1782 | concavity3: 0.6188 | symmetry3: 0.2666

Train Sens: 0.724 | Train Spec: 0.704 [TRAIN SCORE: 1.428] || Val Sens: 0.667 | Val Spec: 0.700 [VAL SCORE: 1.367] || Test Sens: 0.889 | Test Spec: 0.421 [TEST SCORE: 1.310]

##### #68: texture1 + lymph_node_status

Optimized Thresholds: texture1: 17.2517 | lymph_node_status: 5.5751

Train Sens: 0.690 | Train Spec: 0.778 [TRAIN SCORE: 1.467] || Val Sens: 0.667 | Val Spec: 0.700 [VAL SCORE: 1.367] || Test Sens: 0.889 | Test Spec: 0.439 [TEST SCORE: 1.327]

##### #69: texture1 + fractal_dimension3

Optimized Thresholds: texture1: 17.1812 | fractal_dimension3: 0.5845

Train Sens: 0.655 | Train Spec: 0.778 [TRAIN SCORE: 1.433] || Val Sens: 0.667 | Val Spec: 0.700 [VAL SCORE: 1.367] || Test Sens: 0.889 | Test Spec: 0.456 [TEST SCORE: 1.345]

##### #70: texture1 + concavity3 + fractal_dimension3

Optimized Thresholds: texture1: 17.1791 | concavity3: 0.6185 | fractal_dimension3: 0.6104

Train Sens: 0.724 | Train Spec: 0.704 [TRAIN SCORE: 1.428] || Val Sens: 0.667 | Val Spec: 0.700 [VAL SCORE: 1.367] || Test Sens: 0.889 | Test Spec: 0.421 [TEST SCORE: 1.310]

##### #71: texture1 + concavity3 + tumor_size*

Optimized Thresholds: texture1: 17.1793 | concavity3: 0.6175 | tumor_size*: 0.1608

Train Sens: 0.724 | Train Spec: 0.704 [TRAIN SCORE: 1.428] || Val Sens: 0.667 | Val Spec: 0.700 [VAL SCORE: 1.367] || Test Sens: 0.889 | Test Spec: 0.421 [TEST SCORE: 1.310]

##### #72: texture1 + concavity3 + lymph_node_status

Optimized Thresholds: texture1: 17.2316 | concavity3: 0.6185 | lymph_node_status: 7.9646

Train Sens: 0.724 | Train Spec: 0.704 [TRAIN SCORE: 1.428] || Val Sens: 0.667 | Val Spec: 0.700 [VAL SCORE: 1.367] || Test Sens: 0.889 | Test Spec: 0.404 [TEST SCORE: 1.292]

##### #73: texture1 + concave_points3 + symmetry3

Optimized Thresholds: texture1: 17.1761 | concave_points3: 0.6897 | symmetry3: 0.2663

Train Sens: 0.724 | Train Spec: 0.778 [TRAIN SCORE: 1.502] || Val Sens: 0.667 | Val Spec: 0.700 [VAL SCORE: 1.367] || Test Sens: 0.889 | Test Spec: 0.404 [TEST SCORE: 1.292]

##### #74: texture1 + concave_points3

Optimized Thresholds: texture1: 17.1833 | concave_points3: 0.6896

Train Sens: 0.724 | Train Spec: 0.778 [TRAIN SCORE: 1.502] || Val Sens: 0.667 | Val Spec: 0.700 [VAL SCORE: 1.367] || Test Sens: 0.889 | Test Spec: 0.412 [TEST SCORE: 1.301]

##### #75: texture1 + concave_points3 + tumor_size

Optimized Thresholds: texture1: 17.1811 | concave_points3: 0.6892 | tumor_size: 0.1853

Train Sens: 0.724 | Train Spec: 0.778 [TRAIN SCORE: 1.502] || Val Sens: 0.667 | Val Spec: 0.700 [VAL SCORE: 1.367] || Test Sens: 0.889 | Test Spec: 0.412 [TEST SCORE: 1.301]

##### #76: texture1 + area3* + smoothness3* + concave_points3 + symmetry3 + fractal_dimension3 + lymph_node_status

Optimized Thresholds: texture1: 17.2479 | area3*: 187.3801 | smoothness3*: 2335.6665 | concave_points3: 0.6859 | symmetry3: 0.2648 | fractal_dimension3: 0.5607 | lymph_node_status: 7.5945

Train Sens: 0.724 | Train Spec: 0.778 [TRAIN SCORE: 1.502] || Val Sens: 0.667 | Val Spec: 0.700 [VAL SCORE: 1.367] || Test Sens: 0.889 | Test Spec: 0.368 [TEST SCORE: 1.257]

##### #77: texture1 + concave_points3 + lymph_node_status

Optimized Thresholds: texture1: 17.2479 | concave_points3: 0.6895 | lymph_node_status: 7.6368

Train Sens: 0.724 | Train Spec: 0.815 [TRAIN SCORE: 1.539] || Val Sens: 0.667 | Val Spec: 0.700 [VAL SCORE: 1.367] || Test Sens: 0.889 | Test Spec: 0.395 [TEST SCORE: 1.284]

##### #78: texture1 + area3* + smoothness3* + concave_points3 + symmetry3 + tumor_size + lymph_node_status

Optimized Thresholds: texture1: 17.2479 | area3*: 187.3801 | smoothness3*: 2335.6665 | concave_points3: 0.6859 | symmetry3: 0.2648 | tumor_size: 0.1726 | lymph_node_status: 7.5945

Train Sens: 0.724 | Train Spec: 0.778 [TRAIN SCORE: 1.502] || Val Sens: 0.667 | Val Spec: 0.700 [VAL SCORE: 1.367] || Test Sens: 0.889 | Test Spec: 0.377 [TEST SCORE: 1.266]

##### #79: texture1 + concavity3 + concave_points3

Optimized Thresholds: texture1: 17.1793 | concavity3: 0.9440 | concave_points3: 0.6905

Train Sens: 0.724 | Train Spec: 0.778 [TRAIN SCORE: 1.502] || Val Sens: 0.667 | Val Spec: 0.700 [VAL SCORE: 1.367] || Test Sens: 0.889 | Test Spec: 0.412 [TEST SCORE: 1.301]

##### #80: texture1 + area3* + smoothness3* + concavity3 + symmetry3 + tumor_size + lymph_node_status

Optimized Thresholds: texture1: 17.2358 | area3*: 185.4646 | smoothness3*: 2327.7646 | concavity3: 0.9526 | symmetry3: 0.2654 | tumor_size: 0.1807 | lymph_node_status: 5.4107

Train Sens: 0.690 | Train Spec: 0.778 [TRAIN SCORE: 1.467] || Val Sens: 0.667 | Val Spec: 0.700 [VAL SCORE: 1.367] || Test Sens: 0.889 | Test Spec: 0.430 [TEST SCORE: 1.319]

##### #81: texture1 + concavity3

Optimized Thresholds: texture1: 17.1826 | concavity3: 0.6179

Train Sens: 0.724 | Train Spec: 0.704 [TRAIN SCORE: 1.428] || Val Sens: 0.667 | Val Spec: 0.700 [VAL SCORE: 1.367] || Test Sens: 0.889 | Test Spec: 0.421 [TEST SCORE: 1.310]

##### #82: concavity3 + symmetry3 + lymph_node_status

Optimized Thresholds: concavity3: 0.7978 | symmetry3: 0.2106 | lymph_node_status: 2.8439

Train Sens: 0.621 | Train Spec: 0.593 [TRAIN SCORE: 1.213] || Val Sens: 0.778 | Val Spec: 0.500 [VAL SCORE: 1.278] || Test Sens: 0.778 | Test Spec: 0.474 [TEST SCORE: 1.251]

##### #83: smoothness3 + tumor_size

Optimized Thresholds: smoothness3: 1412.4362 | tumor_size: 0.1112

Train Sens: 0.690 | Train Spec: 0.667 [TRAIN SCORE: 1.356] || Val Sens: 0.778 | Val Spec: 0.500 [VAL SCORE: 1.278] || Test Sens: 0.889 | Test Spec: 0.491 [TEST SCORE: 1.380]

##### #84: smoothness3 + fractal_dimension3 + tumor_size

Optimized Thresholds: smoothness3: 1410.0060 | fractal_dimension3: 0.4871 | tumor_size: 0.1115

Train Sens: 0.724 | Train Spec: 0.667 [TRAIN SCORE: 1.391] || Val Sens: 0.778 | Val Spec: 0.500 [VAL SCORE: 1.278] || Test Sens: 0.889 | Test Spec: 0.482 [TEST SCORE: 1.371]

###### #85: smoothness3 + concavity3 + symmetry3 + tumor_size* + lymph_node_status*

Optimized Thresholds: smoothness3: 1411.7110 | concavity3: 0.8717 | symmetry3: 0.2604 | tumor_size*: 0.1199 | lymph_node_status*: 8.2220

Train Sens: 0.621 | Train Spec: 0.704 [TRAIN SCORE: 1.324] || Val Sens: 0.778 | Val Spec: 0.500 [VAL SCORE: 1.278] || Test Sens: 0.778 | Test Spec: 0.500 [TEST SCORE: 1.278]

###### #86: concavity3 + lymph_node_status

Optimized Thresholds: concavity3: 0.9581 | lymph_node_status: 2.2212

Train Sens: 0.724 | Train Spec: 0.556 [TRAIN SCORE: 1.280] || Val Sens: 0.778 | Val Spec: 0.500 [VAL SCORE: 1.278] || Test Sens: 0.889 | Test Spec: 0.518 [TEST SCORE: 1.406]

###### #87: area3* + smoothness3 + tumor_size* + lymph_node_status

Optimized Thresholds: area3*: 184.8793 | smoothness3: 1411.8219 | tumor_size*: 0.1197 | lymph_node_status: 7.2051

Train Sens: 0.621 | Train Spec: 0.704 [TRAIN SCORE: 1.324] || Val Sens: 0.778 | Val Spec: 0.500 [VAL SCORE: 1.278] || Test Sens: 0.778 | Test Spec: 0.491 [TEST SCORE: 1.269]

###### #88: smoothness3 + concavity3* + tumor_size + lymph_node_status*

Optimized Thresholds: smoothness3: 1412.3679 | concavity3*: 0.8245 | tumor_size: 0.1146 | lymph_node_status*: 7.7103

Train Sens: 0.655 | Train Spec: 0.667 [TRAIN SCORE: 1.322] || Val Sens: 0.778 | Val Spec: 0.500 [VAL SCORE: 1.278] || Test Sens: 0.889 | Test Spec: 0.474 [TEST SCORE: 1.363]

###### #89: smoothness3 + concave_points3 + symmetry3 + tumor_size

Optimized Thresholds: smoothness3: 1411.7716 | concave_points3: 0.8111 | symmetry3: 0.2569 | tumor_size: 0.1123

Train Sens: 0.690 | Train Spec: 0.667 [TRAIN SCORE: 1.356] || Val Sens: 0.778 | Val Spec: 0.500 [VAL SCORE: 1.278] || Test Sens: 0.889 | Test Spec: 0.482 [TEST SCORE: 1.371]

###### #90: smoothness3 + symmetry3 + tumor_size

Optimized Thresholds: smoothness3: 1410.0067 | symmetry3: 0.2561 | tumor_size: 0.1117

Train Sens: 0.690 | Train Spec: 0.667 [TRAIN SCORE: 1.356] || Val Sens: 0.778 | Val Spec: 0.500 [VAL SCORE: 1.278] || Test Sens: 0.889 | Test Spec: 0.482 [TEST SCORE: 1.371]

###### #91: area3* + smoothness3 + fractal_dimension3 + tumor_size

Optimized Thresholds: area3*: 192.8023 | smoothness3: 1410.5736 | fractal_dimension3: 0.4872 | tumor_size: 0.1111

Train Sens: 0.724 | Train Spec: 0.667 [TRAIN SCORE: 1.391] || Val Sens: 0.778 | Val Spec: 0.500 [VAL SCORE: 1.278] || Test Sens: 0.889 | Test Spec: 0.482 [TEST SCORE: 1.371]

###### #92: smoothness3 + symmetry3 + fractal_dimension3 + tumor_size

Optimized Thresholds: smoothness3: 1417.2495 | symmetry3: 0.2705 | fractal_dimension3: 0.4865 | tumor_size: 0.1123

Train Sens: 0.724 | Train Spec: 0.667 [TRAIN SCORE: 1.391] || Val Sens: 0.778 | Val Spec: 0.500 [VAL SCORE: 1.278] || Test Sens: 0.889 | Test Spec: 0.491 [TEST SCORE: 1.380]

###### #93: symmetry3 + lymph_node_status

Optimized Thresholds: symmetry3: 0.2114 | lymph_node_status: 2.9124

Train Sens: 0.621 | Train Spec: 0.593 [TRAIN SCORE: 1.213] || Val Sens: 0.778 | Val Spec: 0.500 [VAL SCORE: 1.278] || Test Sens: 0.778 | Test Spec: 0.482 [TEST SCORE: 1.260]

###### #94: area3* + smoothness3 + symmetry3 + tumor_size

Optimized Thresholds: area3*: 180.1252 | smoothness3: 1410.7448 | symmetry3: 0.2562 | tumor_size: 0.1114

Train Sens: 0.690 | Train Spec: 0.667 [TRAIN SCORE: 1.356] || Val Sens: 0.778 | Val Spec: 0.500 [VAL SCORE: 1.278] || Test Sens: 0.889 | Test Spec: 0.482 [TEST SCORE: 1.371]

###### #95: concavity3 + tumor_size + lymph_node_status

Optimized Thresholds: concavity3: 0.9794 | tumor_size: 0.1947 | lymph_node_status: 2.0982

Train Sens: 0.724 | Train Spec: 0.556 [TRAIN SCORE: 1.280] || Val Sens: 0.778 | Val Spec: 0.500 [VAL SCORE: 1.278] || Test Sens: 0.889 | Test Spec: 0.509 [TEST SCORE: 1.398]

###### #96: smoothness3 + concavity3 + symmetry3 + tumor_size

Optimized Thresholds: smoothness3: 1411.3902 | concavity3: 0.8945 | symmetry3: 0.2603 | tumor_size: 0.1118

Train Sens: 0.690 | Train Spec: 0.667 [TRAIN SCORE: 1.356] || Val Sens: 0.778 | Val Spec: 0.500 [VAL SCORE: 1.278] || Test Sens: 0.889 | Test Spec: 0.482 [TEST SCORE: 1.371]

###### #97: tumor_size + lymph_node_status

Optimized Thresholds: tumor_size: 0.1938 | lymph_node_status: 2.1747

Train Sens: 0.724 | Train Spec: 0.556 [TRAIN SCORE: 1.280] || Val Sens: 0.778 | Val Spec: 0.500 [VAL SCORE: 1.278] || Test Sens: 0.889 | Test Spec: 0.518 [TEST SCORE: 1.406]

###### #98: smoothness3 + concavity3 + tumor_size

Optimized Thresholds: smoothness3: 1413.3941 | concavity3: 0.8800 | tumor_size: 0.1118

Train Sens: 0.690 | Train Spec: 0.667 [TRAIN SCORE: 1.356] || Val Sens: 0.778 | Val Spec: 0.500 [VAL SCORE: 1.278] || Test Sens: 0.889 | Test Spec: 0.491 [TEST SCORE: 1.380]

###### #99: smoothness3 + concavity3 + concave_points3* + tumor_size

Optimized Thresholds: smoothness3: 1413.2059 | concavity3: 0.8640 | concave_points3*: 0.8526 | tumor_size: 0.1120

Train Sens: 0.690 | Train Spec: 0.667 [TRAIN SCORE: 1.356] || Val Sens: 0.778 | Val Spec: 0.500 [VAL SCORE: 1.278] || Test Sens: 0.889 | Test Spec: 0.482 [TEST SCORE: 1.371]

###### #100: area3 + smoothness3 + symmetry3 + fractal_dimension3 + tumor_size + lymph_node_status

Optimized Thresholds: area3: 217.5771 | smoothness3: 1405.5352 | symmetry3: 0.2700 | fractal_dimension3: 0.4872 | tumor_size: 0.1119 | lymph_node_status: 9.9453

Train Sens: 0.724 | Train Spec: 0.667 [TRAIN SCORE: 1.391] || Val Sens: 0.778 | Val Spec: 0.500 [VAL SCORE: 1.278] || Test Sens: 0.889 | Test Spec: 0.456 [TEST SCORE: 1.345]

###### #101: area3 + smoothness3 + concavity3 + fractal_dimension3 + tumor_size + lymph_node_status

Optimized Thresholds: area3: 217.5771 | smoothness3: 1405.5352 | concavity3: 1.0294 | fractal_dimension3: 0.4872 | tumor_size: 0.1119 | lymph_node_status: 9.9453

Train Sens: 0.724 | Train Spec: 0.667 [TRAIN SCORE: 1.391] || Val Sens: 0.778 | Val Spec: 0.500 [VAL SCORE: 1.278] || Test Sens: 0.889 | Test Spec: 0.456 [TEST SCORE: 1.345]

**#102: area3* + smoothness3 + tumor_size**

Optimized Thresholds: area3*: 186.4783 | smoothness3: 1415.4283 | tumor_size: 0.1109

Train Sens: 0.690 | Train Spec: 0.667 [TRAIN SCORE: 1.356] || Val Sens: 0.778 | Val Spec: 0.500 [VAL SCORE: 1.278] || Test Sens: 0.889 | Test Spec: 0.491 [TEST SCORE: 1.380]

**#103: area3 + smoothness3 + concavity3 + symmetry3 + tumor_size + lymph_node_status***

Optimized Thresholds: area3: 200.7007 | smoothness3: 1412.1500 | concavity3: 0.9548 | symmetry3: 0.2574 | tumor_size: 0.1176 | lymph_node_status*: 7.8273

Train Sens: 0.655 | Train Spec: 0.667 [TRAIN SCORE: 1.322] || Val Sens: 0.778 | Val Spec: 0.500 [VAL SCORE: 1.278] || Test Sens: 0.778 | Test Spec: 0.465 [TEST SCORE: 1.243]

**#104: area3 + smoothness3 + concavity3 + symmetry3 + tumor_size**

Optimized Thresholds: area3: 201.4076 | smoothness3: 1410.8963 | concavity3: 0.9106 | symmetry3: 0.2618 | tumor_size: 0.1112

Train Sens: 0.690 | Train Spec: 0.667 [TRAIN SCORE: 1.356] || Val Sens: 0.778 | Val Spec: 0.500 [VAL SCORE: 1.278] || Test Sens: 0.889 | Test Spec: 0.491 [TEST SCORE: 1.380]

**#105: area3* + smoothness3 + concavity3 + tumor_size + lymph_node_status***

Optimized Thresholds: area3*: 203.3607 | smoothness3: 1410.6590 | concavity3: 0.8546 | tumor_size: 0.1186 | lymph_node_status*: 7.8348

Train Sens: 0.621 | Train Spec: 0.704 [TRAIN SCORE: 1.324] || Val Sens: 0.778 | Val Spec: 0.500 [VAL SCORE: 1.278] || Test Sens: 0.778 | Test Spec: 0.482 [TEST SCORE: 1.260]

**#106: smoothness3 + concave_points3 + tumor_size**

Optimized Thresholds: smoothness3: 1408.5233 | concave_points3: 0.8542 | tumor_size: 0.1119

Train Sens: 0.690 | Train Spec: 0.667 [TRAIN SCORE: 1.356] || Val Sens: 0.778 | Val Spec: 0.500 [VAL SCORE: 1.278] || Test Sens: 0.889 | Test Spec: 0.474 [TEST SCORE: 1.363]

**#107: area3* + smoothness3 + concavity3 + tumor_size**

Optimized Thresholds: area3*: 192.4432 | smoothness3: 1411.3764 | concavity3: 0.8917 | tumor_size: 0.1113

Train Sens: 0.690 | Train Spec: 0.667 [TRAIN SCORE: 1.356] || Val Sens: 0.778 | Val Spec: 0.500 [VAL SCORE: 1.278] || Test Sens: 0.889 | Test Spec: 0.491 [TEST SCORE: 1.380]

**#108: smoothness3 + concavity3 + concave_points3 + symmetry3 + tumor_size**

Optimized Thresholds: smoothness3: 1411.3179 | concavity3: 0.8870 | concave_points3: 0.7772 | symmetry3: 0.2600 | tumor_size: 0.1117

Train Sens: 0.690 | Train Spec: 0.667 [TRAIN SCORE: 1.356] || Val Sens: 0.778 | Val Spec: 0.500 [VAL SCORE: 1.278] || Test Sens: 0.889 | Test Spec: 0.482 [TEST SCORE: 1.371]

**#109: texture1 + area3 + concavity3 + concave_points3 + tumor_size**

Optimized Thresholds: texture1: 17.1806 | area3: 153.1193 | concavity3: 0.8859 | concave_points3: 0.6687 | tumor_size: 0.1516

Train Sens: 0.759 | Train Spec: 0.741 [TRAIN SCORE: 1.499] || Val Sens: 0.667 | Val Spec: 0.600 [VAL SCORE: 1.267] || Test Sens: 0.889 | Test Spec: 0.404 [TEST SCORE: 1.292]

**#110: texture1 + area3 + smoothness3 + concave_points3 + tumor_size**

Optimized Thresholds: texture1: 17.1778 | area3: 216.3871 | smoothness3: 1682.8785 | concave_points3: 0.6902 | tumor_size:

0.1972
Train Sens: 0.759 | Train Spec: 0.778 [TRAIN SCORE: 1.536] || Val Sens: 0.667 | Val Spec: 0.600 [VAL SCORE: 1.267] || Test Sens: 0.889 | Test Spec: 0.412 [TEST SCORE: 1.301]

**#111: texture1 + area3 + smoothness3 + concave_points3 + lymph_node_status**

Optimized Thresholds: texture1: 17.2123 | area3: 159.2328 | smoothness3: 1686.4514 | concave_points3: 0.6874 | lymph_node_status: 8.2599
Train Sens: 0.759 | Train Spec: 0.815 [TRAIN SCORE: 1.573] || Val Sens: 0.667 | Val Spec: 0.600 [VAL SCORE: 1.267] || Test Sens: 0.889 | Test Spec: 0.404 [TEST SCORE: 1.292]

**#112: texture1 + smoothness3**

Optimized Thresholds: texture1: 17.1795 | smoothness3: 1561.8471
Train Sens: 0.690 | Train Spec: 0.815 [TRAIN SCORE: 1.504] || Val Sens: 0.667 | Val Spec: 0.600 [VAL SCORE: 1.267] || Test Sens: 0.889 | Test Spec: 0.456 [TEST SCORE: 1.345]

**#113: texture1 + area3 + smoothness3* + concavity3 + lymph_node_status**

Optimized Thresholds: texture1: 17.2481 | area3: 169.0676 | smoothness3*: 2256.1292 | concavity3: 0.9404 | lymph_node_status: 5.5671
Train Sens: 0.690 | Train Spec: 0.778 [TRAIN SCORE: 1.467] || Val Sens: 0.667 | Val Spec: 0.600 [VAL SCORE: 1.267] || Test Sens: 0.889 | Test Spec: 0.439 [TEST SCORE: 1.327]

**#114: texture1 + area3 + concavity3 + concave_points3 + lymph_node_status**

Optimized Thresholds: texture1: 17.2715 | area3: 154.8885 | concavity3: 0.9083 | concave_points3: 0.6707 | lymph_node_status: 6.7803
Train Sens: 0.759 | Train Spec: 0.778 [TRAIN SCORE: 1.536] || Val Sens: 0.667 | Val Spec: 0.600 [VAL SCORE: 1.267] || Test Sens: 0.889 | Test Spec: 0.386 [TEST SCORE: 1.275]

**#115: texture1 + area3 + concavity3 + symmetry3 + fractal_dimension3**

Optimized Thresholds: texture1: 17.1835 | area3: 154.8260 | concavity3: 0.6194 | symmetry3: 0.2608 | fractal_dimension3: 0.5733
Train Sens: 0.759 | Train Spec: 0.704 [TRAIN SCORE: 1.462] || Val Sens: 0.667 | Val Spec: 0.600 [VAL SCORE: 1.267] || Test Sens: 0.889 | Test Spec: 0.412 [TEST SCORE: 1.301]

**#116: texture1 + area3 + concavity3 + concave_points3 + fractal_dimension3**

Optimized Thresholds: texture1: 17.1841 | area3: 152.7445 | concavity3: 0.8919 | concave_points3: 0.6798 | fractal_dimension3: 0.5348
Train Sens: 0.759 | Train Spec: 0.741 [TRAIN SCORE: 1.499] || Val Sens: 0.667 | Val Spec: 0.600 [VAL SCORE: 1.267] || Test Sens: 0.889 | Test Spec: 0.395 [TEST SCORE: 1.284]

**#117: texture1 + area3 + concavity3 + concave_points3 + symmetry3**

Optimized Thresholds: texture1: 17.2150 | area3: 154.3257 | concavity3: 0.9215 | concave_points3: 0.6887 | symmetry3: 0.2631
Train Sens: 0.724 | Train Spec: 0.815 [TRAIN SCORE: 1.539] || Val Sens: 0.667 | Val Spec: 0.600 [VAL SCORE: 1.267] || Test Sens: 0.889 | Test Spec: 0.404 [TEST SCORE: 1.292]

**#118: texture1 + area3 + smoothness3* + tumor_size + lymph_node_status**

Optimized Thresholds: texture1: 17.2287 | area3: 161.6018 | smoothness3*: 2175.6121 | tumor_size: 0.1889 | lymph_node_status: 5.5476

Train Sens: 0.690 | Train Spec: 0.778 [TRAIN SCORE: 1.467] || Val Sens: 0.667 | Val Spec: 0.600 [VAL SCORE: 1.267] || Test Sens: 0.889 | Test Spec: 0.439 [TEST SCORE: 1.327]

**#119: texture1 + area3 + smoothness3* + fractal_dimension3 + lymph_node_status**

Optimized Thresholds: texture1: 17.2491 | area3: 165.9573 | smoothness3*: 2232.8017 | fractal_dimension3: 0.5845 | lymph_node_status: 5.5060

Train Sens: 0.690 | Train Spec: 0.741 [TRAIN SCORE: 1.430] || Val Sens: 0.667 | Val Spec: 0.600 [VAL SCORE: 1.267] || Test Sens: 0.889 | Test Spec: 0.439 [TEST SCORE: 1.327]

**#120: texture1 + area3* + smoothness3* + symmetry3 + fractal_dimension3**

Optimized Thresholds: texture1: 17.1774 | area3*: 191.1834 | smoothness3*: 1814.9364 | symmetry3: 0.2628 | fractal_dimension3: 0.5961

Train Sens: 0.655 | Train Spec: 0.778 [TRAIN SCORE: 1.433] || Val Sens: 0.667 | Val Spec: 0.600 [VAL SCORE: 1.267] || Test Sens: 0.889 | Test Spec: 0.447 [TEST SCORE: 1.336]

**#121: texture1 + area3 + smoothness3* + symmetry3 + lymph_node_status**

Optimized Thresholds: texture1: 17.2464 | area3: 168.6754 | smoothness3*: 2113.0779 | symmetry3: 0.2691 | lymph_node_status: 5.6102

Train Sens: 0.690 | Train Spec: 0.778 [TRAIN SCORE: 1.467] || Val Sens: 0.667 | Val Spec: 0.600 [VAL SCORE: 1.267] || Test Sens: 0.889 | Test Spec: 0.430 [TEST SCORE: 1.319]

**#122: texture1 + area3 + concavity3 + symmetry3 + tumor_size**

Optimized Thresholds: texture1: 17.1835 | area3: 154.8260 | concavity3: 0.6194 | symmetry3: 0.2608 | tumor_size: 0.1768

Train Sens: 0.759 | Train Spec: 0.704 [TRAIN SCORE: 1.462] || Val Sens: 0.667 | Val Spec: 0.600 [VAL SCORE: 1.267] || Test Sens: 0.889 | Test Spec: 0.421 [TEST SCORE: 1.310]

**#123: texture1 + area3* + smoothness3 + concave_points3 + fractal_dimension3**

Optimized Thresholds: texture1: 17.1817 | area3*: 186.6784 | smoothness3: 1744.9939 | concave_points3: 0.6696 | fractal_dimension3: 0.5746

Train Sens: 0.759 | Train Spec: 0.741 [TRAIN SCORE: 1.499] || Val Sens: 0.667 | Val Spec: 0.600 [VAL SCORE: 1.267] || Test Sens: 0.889 | Test Spec: 0.395 [TEST SCORE: 1.284]

**#124: texture1 + area3 + concave_points3 + fractal_dimension3 + lymph_node_status**

Optimized Thresholds: texture1: 17.2381 | area3: 153.3805 | concave_points3: 0.6756 | fractal_dimension3: 0.5872 | lymph_node_status: 7.5173

Train Sens: 0.759 | Train Spec: 0.778 [TRAIN SCORE: 1.536] || Val Sens: 0.667 | Val Spec: 0.600 [VAL SCORE: 1.267] || Test Sens: 0.889 | Test Spec: 0.386 [TEST SCORE: 1.275]

**#125: texture1 + area3 + concavity3 + symmetry3 + lymph_node_status**

Optimized Thresholds: texture1: 17.2180 | area3: 156.3258 | concavity3: 1.0061 | symmetry3: 0.2707 | lymph_node_status: 5.6698

Train Sens: 0.724 | Train Spec: 0.778 [TRAIN SCORE: 1.502] || Val Sens: 0.667 | Val Spec: 0.600 [VAL SCORE: 1.267] || Test Sens: 0.889 | Test Spec: 0.439 [TEST SCORE: 1.327]

**#126: texture1 + smoothness3 + concave_points3 + symmetry3 + fractal_dimension3**

Optimized Thresholds: texture1: 17.1787 | smoothness3: 1603.7684 | concave_points3: 0.6735 | symmetry3: 0.2668 | fractal_dimension3: 0.6112

Train Sens: 0.759 | Train Spec: 0.741 [TRAIN SCORE: 1.499] || Val Sens: 0.667 | Val Spec: 0.600 [VAL SCORE: 1.267] || Test Sens: 0.889 | Test Spec: 0.395 [TEST SCORE: 1.284]

###### #127: texture1 + area3 + concave_points3 + tumor_size + lymph_node_status

Optimized Thresholds: texture1: 17.2264 | area3: 152.4336 | concave_points3: 0.6871 | tumor_size: 0.1748 | lymph_node_status: 7.7550

Train Sens: 0.759 | Train Spec: 0.815 [TRAIN SCORE: 1.573] || Val Sens: 0.667 | Val Spec: 0.600 [VAL SCORE: 1.267] || Test Sens: 0.889 | Test Spec: 0.386 [TEST SCORE: 1.275]

###### #128: texture1 + area3 + symmetry3 + fractal_dimension3 + lymph_node_status

Optimized Thresholds: texture1: 17.2453 | area3: 154.9147 | symmetry3: 0.2666 | fractal_dimension3: 0.5928 | lymph_node_status: 5.5779

Train Sens: 0.724 | Train Spec: 0.741 [TRAIN SCORE: 1.465] || Val Sens: 0.667 | Val Spec: 0.600 [VAL SCORE: 1.267] || Test Sens: 0.889 | Test Spec: 0.430 [TEST SCORE: 1.319]

###### #129: texture1 + area3 + symmetry3 + tumor_size + lymph_node_status

Optimized Thresholds: texture1: 17.2402 | area3: 154.9329 | symmetry3: 0.2692 | tumor_size: 0.1921 | lymph_node_status: 5.5795

Train Sens: 0.724 | Train Spec: 0.778 [TRAIN SCORE: 1.502] || Val Sens: 0.667 | Val Spec: 0.600 [VAL SCORE: 1.267] || Test Sens: 0.889 | Test Spec: 0.430 [TEST SCORE: 1.319]

###### #130: texture1 + area3 + fractal_dimension3 + tumor_size + lymph_node_status

Optimized Thresholds: texture1: 17.2615 | area3: 155.0258 | fractal_dimension3: 0.5900 | tumor_size: 0.1421 | lymph_node_status: 6.0277

Train Sens: 0.690 | Train Spec: 0.741 [TRAIN SCORE: 1.430] || Val Sens: 0.667 | Val Spec: 0.600 [VAL SCORE: 1.267] || Test Sens: 0.889 | Test Spec: 0.430 [TEST SCORE: 1.319]

###### #131: texture1 + smoothness3 + concavity3 + concave_points3 + symmetry3

Optimized Thresholds: texture1: 17.1904 | smoothness3: 1543.1136 | concavity3: 0.9031 | concave_points3: 0.6852 | symmetry3: 0.2647

Train Sens: 0.724 | Train Spec: 0.741 [TRAIN SCORE: 1.465] || Val Sens: 0.667 | Val Spec: 0.600 [VAL SCORE: 1.267] || Test Sens: 0.889 | Test Spec: 0.395 [TEST SCORE: 1.284]

###### #132: texture1 + smoothness3 + concavity3 + concave_points3 + fractal_dimension3

Optimized Thresholds: texture1: 17.1801 | smoothness3: 1605.8305 | concavity3: 0.8646 | concave_points3: 0.6747 | fractal_dimension3: 0.5841

Train Sens: 0.759 | Train Spec: 0.741 [TRAIN SCORE: 1.499] || Val Sens: 0.667 | Val Spec: 0.600 [VAL SCORE: 1.267] || Test Sens: 0.889 | Test Spec: 0.404 [TEST SCORE: 1.292]

###### #133: texture1 + smoothness3 + concavity3 + concave_points3 + tumor_size*

Optimized Thresholds: texture1: 17.1776 | smoothness3: 1578.4050 | concavity3: 0.8540 | concave_points3: 0.6699 | tumor_size*: 0.1661

Train Sens: 0.759 | Train Spec: 0.741 [TRAIN SCORE: 1.499] || Val Sens: 0.667 | Val Spec: 0.600 [VAL SCORE: 1.267] || Test Sens: 0.889 | Test Spec: 0.404 [TEST SCORE: 1.292]

###### #134: texture1 + smoothness3 + concavity3 + concave_points3 + lymph_node_status

Optimized Thresholds: texture1: 17.2388 | smoothness3: 1723.3648 | concavity3: 1.0280 | concave_points3: 0.6873 |

lymph_node_status: 7.6158

Train Sens: 0.759 | Train Spec: 0.815 [TRAIN SCORE: 1.573] || Val Sens: 0.667 | Val Spec: 0.600 [VAL SCORE: 1.267] || Test Sens: 0.889 | Test Spec: 0.395 [TEST SCORE: 1.284]

**#135: texture1 + smoothness3 + concavity3 + symmetry3 + fractal_dimension3**

Optimized Thresholds: texture1: 17.1792 | smoothness3: 1635.1134 | concavity3: 0.6224 | symmetry3: 0.2661 | fractal_dimension3: 0.5743

Train Sens: 0.759 | Train Spec: 0.704 [TRAIN SCORE: 1.462] || Val Sens: 0.667 | Val Spec: 0.600 [VAL SCORE: 1.267] || Test Sens: 0.889 | Test Spec: 0.412 [TEST SCORE: 1.301]

**#136: texture1 + smoothness3 + concavity3 + symmetry3 + lymph_node_status**

Optimized Thresholds: texture1: 17.2345 | smoothness3: 1687.8584 | concavity3: 0.9451 | symmetry3: 0.2652 | lymph_node_status: 5.6770

Train Sens: 0.724 | Train Spec: 0.778 [TRAIN SCORE: 1.502] || Val Sens: 0.667 | Val Spec: 0.600 [VAL SCORE: 1.267] || Test Sens: 0.889 | Test Spec: 0.430 [TEST SCORE: 1.319]

**#137: texture1 + smoothness3 + concavity3 + fractal_dimension3 + lymph_node_status**

Optimized Thresholds: texture1: 17.2457 | smoothness3: 1713.4769 | concavity3: 0.8297 | fractal_dimension3: 0.5988 | lymph_node_status: 5.9253

Train Sens: 0.724 | Train Spec: 0.741 [TRAIN SCORE: 1.465] || Val Sens: 0.667 | Val Spec: 0.600 [VAL SCORE: 1.267] || Test Sens: 0.889 | Test Spec: 0.421 [TEST SCORE: 1.310]

**#138: texture1 + smoothness3 + concavity3 + tumor_size + lymph_node_status**

Optimized Thresholds: texture1: 17.2323 | smoothness3: 1714.5678 | concavity3: 0.9425 | tumor_size: 0.1873 | lymph_node_status: 5.6507

Train Sens: 0.724 | Train Spec: 0.778 [TRAIN SCORE: 1.502] || Val Sens: 0.667 | Val Spec: 0.600 [VAL SCORE: 1.267] || Test Sens: 0.889 | Test Spec: 0.439 [TEST SCORE: 1.327]

**#139: texture1 + smoothness3 + concave_points3 + symmetry3 + tumor_size**

Optimized Thresholds: texture1: 17.1800 | smoothness3: 1616.3503 | concave_points3: 0.6755 | symmetry3: 0.2679 | tumor_size: 0.1787

Train Sens: 0.759 | Train Spec: 0.741 [TRAIN SCORE: 1.499] || Val Sens: 0.667 | Val Spec: 0.600 [VAL SCORE: 1.267] || Test Sens: 0.889 | Test Spec: 0.395 [TEST SCORE: 1.284]

**#140: texture1 + area3 + concavity3 + fractal_dimension3 + lymph_node_status**

Optimized Thresholds: texture1: 17.2400 | area3: 154.3764 | concavity3: 0.8362 | fractal_dimension3: 0.5693 | lymph_node_status: 5.8433

Train Sens: 0.724 | Train Spec: 0.741 [TRAIN SCORE: 1.465] || Val Sens: 0.667 | Val Spec: 0.600 [VAL SCORE: 1.267] || Test Sens: 0.889 | Test Spec: 0.412 [TEST SCORE: 1.301]

**#141: texture1 + area3 + concave_points3 + fractal_dimension3 + tumor_size**

Optimized Thresholds: texture1: 17.1802 | area3: 154.7271 | concave_points3: 0.6741 | fractal_dimension3: 0.5804 | tumor_size: 0.1683

Train Sens: 0.759 | Train Spec: 0.741 [TRAIN SCORE: 1.499] || Val Sens: 0.667 | Val Spec: 0.600 [VAL SCORE: 1.267] || Test Sens: 0.889 | Test Spec: 0.404 [TEST SCORE: 1.292]

**#142: texture1 + smoothness3 + concave_points3 + symmetry3 + lymph_node_status**

Optimized Thresholds: texture1: 17.2371 | smoothness3: 1699.6041 | concave_points3: 0.6783 | symmetry3: 0.2644 | lymph_node_status: 7.3410

Train Sens: 0.759 | Train Spec: 0.778 [TRAIN SCORE: 1.536] || Val Sens: 0.667 | Val Spec: 0.600 [VAL SCORE: 1.267] || Test Sens: 0.889 | Test Spec: 0.377 [TEST SCORE: 1.266]

**#143: texture1 + smoothness3 + concave_points3 + fractal_dimension3 + tumor_size***

Optimized Thresholds: texture1: 17.1782 | smoothness3: 1654.6411 | concave_points3: 0.6670 | fractal_dimension3: 0.5663 | tumor_size*: 0.1668

Train Sens: 0.759 | Train Spec: 0.741 [TRAIN SCORE: 1.499] || Val Sens: 0.667 | Val Spec: 0.600 [VAL SCORE: 1.267] || Test Sens: 0.889 | Test Spec: 0.395 [TEST SCORE: 1.284]

**#144: texture1 + smoothness3 + concave_points3 + fractal_dimension3 + lymph_node_status**

Optimized Thresholds: texture1: 17.2451 | smoothness3: 1727.4790 | concave_points3: 0.6682 | fractal_dimension3: 0.5683 | lymph_node_status: 7.7017

Train Sens: 0.759 | Train Spec: 0.778 [TRAIN SCORE: 1.536] || Val Sens: 0.667 | Val Spec: 0.600 [VAL SCORE: 1.267] || Test Sens: 0.889 | Test Spec: 0.377 [TEST SCORE: 1.266]

**#145: texture1 + smoothness3 + concave_points3 + tumor_size + lymph_node_status**

Optimized Thresholds: texture1: 17.2279 | smoothness3: 1717.5452 | concave_points3: 0.6762 | tumor_size: 0.1820 | lymph_node_status: 7.5800

Train Sens: 0.759 | Train Spec: 0.778 [TRAIN SCORE: 1.536] || Val Sens: 0.667 | Val Spec: 0.600 [VAL SCORE: 1.267] || Test Sens: 0.889 | Test Spec: 0.386 [TEST SCORE: 1.275]

**#146: texture1 + smoothness3 + symmetry3 + fractal_dimension3 + lymph_node_status**

Optimized Thresholds: texture1: 17.2323 | smoothness3: 1725.9264 | symmetry3: 0.2661 | fractal_dimension3: 0.5465 | lymph_node_status: 5.6710

Train Sens: 0.724 | Train Spec: 0.741 [TRAIN SCORE: 1.465] || Val Sens: 0.667 | Val Spec: 0.600 [VAL SCORE: 1.267] || Test Sens: 0.889 | Test Spec: 0.421 [TEST SCORE: 1.310]

**#147: texture1 + area3 + concave_points3 + symmetry3 + lymph_node_status**

Optimized Thresholds: texture1: 17.2149 | area3: 154.8875 | concave_points3: 0.6805 | symmetry3: 0.2646 | lymph_node_status: 7.7397

Train Sens: 0.759 | Train Spec: 0.778 [TRAIN SCORE: 1.536] || Val Sens: 0.667 | Val Spec: 0.600 [VAL SCORE: 1.267] || Test Sens: 0.889 | Test Spec: 0.377 [TEST SCORE: 1.266]

**#148: texture1 + smoothness3 + symmetry3 + tumor_size + lymph_node_status**

Optimized Thresholds: texture1: 17.2262 | smoothness3: 1720.1629 | symmetry3: 0.2671 | tumor_size: 0.1878 | lymph_node_status: 5.8191

Train Sens: 0.724 | Train Spec: 0.778 [TRAIN SCORE: 1.502] || Val Sens: 0.667 | Val Spec: 0.600 [VAL SCORE: 1.267] || Test Sens: 0.889 | Test Spec: 0.430 [TEST SCORE: 1.319]

**#149: texture1 + smoothness3 + fractal_dimension3 + tumor_size* + lymph_node_status**

Optimized Thresholds: texture1: 17.2384 | smoothness3: 1633.4365 | fractal_dimension3: 0.5579 | tumor_size*: 0.1422 | lymph_node_status: 7.2008

Train Sens: 0.690 | Train Spec: 0.741 [TRAIN SCORE: 1.430] || Val Sens: 0.667 | Val Spec: 0.600 [VAL SCORE: 1.267] || Test Sens: 0.889 | Test Spec: 0.421 [TEST SCORE: 1.310]

###### #150: texture1 + area3 + concave_points3 + symmetry3 + tumor_size

Optimized Thresholds: texture1: 17.1791 | area3: 154.3426 | concave_points3: 0.6874 | symmetry3: 0.2577 | tumor_size: 0.1874

Train Sens: 0.759 | Train Spec: 0.778 [TRAIN SCORE: 1.536] || Val Sens: 0.667 | Val Spec: 0.600 [VAL SCORE: 1.267] || Test Sens: 0.889 | Test Spec: 0.404 [TEST SCORE: 1.292]

###### #151: texture1 + area3 + concave_points3 + symmetry3 + fractal_dimension3

Optimized Thresholds: texture1: 17.1811 | area3: 155.2477 | concave_points3: 0.6789 | symmetry3: 0.2654 | fractal_dimension3: 0.5706

Train Sens: 0.759 | Train Spec: 0.741 [TRAIN SCORE: 1.499] || Val Sens: 0.667 | Val Spec: 0.600 [VAL SCORE: 1.267] || Test Sens: 0.889 | Test Spec: 0.386 [TEST SCORE: 1.275]

###### #152: texture1 + area3 + concavity3 + tumor_size + lymph_node_status

Optimized Thresholds: texture1: 17.2366 | area3: 154.9507 | concavity3: 1.0062 | tumor_size: 0.1904 | lymph_node_status: 5.5261

Train Sens: 0.724 | Train Spec: 0.778 [TRAIN SCORE: 1.502] || Val Sens: 0.667 | Val Spec: 0.600 [VAL SCORE: 1.267] || Test Sens: 0.889 | Test Spec: 0.439 [TEST SCORE: 1.327]

###### #153: texture1 + area3

Optimized Thresholds: texture1: 17.1786 | area3: 154.9662

Train Sens: 0.690 | Train Spec: 0.815 [TRAIN SCORE: 1.504] || Val Sens: 0.667 | Val Spec: 0.600 [VAL SCORE: 1.267] || Test Sens: 0.889 | Test Spec: 0.456 [TEST SCORE: 1.345]

###### #154: texture1 + area3* + smoothness3* + concavity3 + fractal_dimension3 + lymph_node_status

Optimized Thresholds: texture1: 17.2504 | area3*: 172.4409 | smoothness3*: 2065.9788 | concavity3: 0.8254 | fractal_dimension3: 0.5751 | lymph_node_status: 5.9498

Train Sens: 0.690 | Train Spec: 0.741 [TRAIN SCORE: 1.430] || Val Sens: 0.667 | Val Spec: 0.600 [VAL SCORE: 1.267] || Test Sens: 0.889 | Test Spec: 0.412 [TEST SCORE: 1.301]

###### #155: texture1 + symmetry3 + fractal_dimension3 + tumor_size + lymph_node_status

Optimized Thresholds: texture1: 17.2044 | symmetry3: 0.2612 | fractal_dimension3: 0.6569 | tumor_size: 0.1341 | lymph_node_status: 7.2568

Train Sens: 0.690 | Train Spec: 0.778 [TRAIN SCORE: 1.467] || Val Sens: 0.667 | Val Spec: 0.600 [VAL SCORE: 1.267] || Test Sens: 0.889 | Test Spec: 0.395 [TEST SCORE: 1.284]

###### #156: smoothness3 + concavity3 + concave_points3 + symmetry3 + tumor_size + lymph_node_status

Optimized Thresholds: smoothness3: 1416.4095 | concavity3: 0.8149 | concave_points3: 0.6758 | symmetry3: 0.2625 | tumor_size: 0.1477 | lymph_node_status: 3.7489

Train Sens: 0.690 | Train Spec: 0.704 [TRAIN SCORE: 1.393] || Val Sens: 0.667 | Val Spec: 0.600 [VAL SCORE: 1.267] || Test Sens: 0.889 | Test Spec: 0.447 [TEST SCORE: 1.336]

###### #157: texture1 + area3* + smoothness3* + concave_points3 + symmetry3 + fractal_dimension3 + tumor_size

Optimized Thresholds: texture1: 17.2395 | area3*: 177.3456 | smoothness3*: 2130.5997 | concave_points3: 0.6762 | symmetry3: 0.2677 | fractal_dimension3: 0.5392 | tumor_size: 0.1828

Train Sens: 0.690 | Train Spec: 0.778 [TRAIN SCORE: 1.467] || Val Sens: 0.667 | Val Spec: 0.600 [VAL SCORE: 1.267] || Test Sens: 0.889 | Test Spec: 0.386 [TEST SCORE: 1.275]

**#158: texture1 + area3* + smoothness3 + concavity3* + fractal_dimension3 + tumor_size* + lymph_node_status**

Optimized Thresholds: texture1: 17.2199 | area3*: 188.5782 | smoothness3: 1715.5146 | concavity3*: 0.8236 | fractal_dimension3: 0.5848 | tumor_size*: 0.1414 | lymph_node_status: 7.2827

Train Sens: 0.690 | Train Spec: 0.778 [TRAIN SCORE: 1.467] || Val Sens: 0.667 | Val Spec: 0.600 [VAL SCORE: 1.267] || Test Sens: 0.889 | Test Spec: 0.421 [TEST SCORE: 1.310]

**#159: texture1 + area3* + smoothness3* + concavity3 + symmetry3 + fractal_dimension3 + lymph_node_status**

Optimized Thresholds: texture1: 17.2345 | area3*: 184.2448 | smoothness3*: 2190.0975 | concavity3: 0.9016 | symmetry3: 0.2652 | fractal_dimension3: 0.5788 | lymph_node_status: 5.9105

Train Sens: 0.690 | Train Spec: 0.741 [TRAIN SCORE: 1.430] || Val Sens: 0.667 | Val Spec: 0.600 [VAL SCORE: 1.267] || Test Sens: 0.889 | Test Spec: 0.412 [TEST SCORE: 1.301]

**#160: texture1 + area3 + smoothness3* + concavity3 + concave_points3 + tumor_size + lymph_node_status**

Optimized Thresholds: texture1: 17.2471 | area3: 159.8928 | smoothness3*: 2200.2131 | concavity3: 0.9950 | concave_points3: 0.6822 | tumor_size: 0.1830 | lymph_node_status: 6.9991

Train Sens: 0.724 | Train Spec: 0.778 [TRAIN SCORE: 1.502] || Val Sens: 0.667 | Val Spec: 0.600 [VAL SCORE: 1.267] || Test Sens: 0.889 | Test Spec: 0.386 [TEST SCORE: 1.275]

**#161: texture1 + area3 + smoothness3* + concavity3 + concave_points3 + fractal_dimension3 + lymph_node_status**

Optimized Thresholds: texture1: 17.2620 | area3: 159.0457 | smoothness3*: 2453.5202 | concavity3: 0.9722 | concave_points3: 0.6772 | fractal_dimension3: 0.5568 | lymph_node_status: 7.4661

Train Sens: 0.724 | Train Spec: 0.778 [TRAIN SCORE: 1.502] || Val Sens: 0.667 | Val Spec: 0.600 [VAL SCORE: 1.267] || Test Sens: 0.889 | Test Spec: 0.377 [TEST SCORE: 1.266]

**#162: texture1 + area3* + smoothness3 + concavity3 + concave_points3 + fractal_dimension3 + tumor_size**

Optimized Thresholds: texture1: 17.1804 | area3*: 181.5210 | smoothness3: 1606.2960 | concavity3: 0.7862 | concave_points3: 0.6693 | fractal_dimension3: 0.5937 | tumor_size: 0.1524

Train Sens: 0.759 | Train Spec: 0.741 [TRAIN SCORE: 1.499] || Val Sens: 0.667 | Val Spec: 0.600 [VAL SCORE: 1.267] || Test Sens: 0.889 | Test Spec: 0.404 [TEST SCORE: 1.292]

**#163: texture1 + area3 + smoothness3 + concavity3 + concave_points3 + symmetry3 + lymph_node_status**

Optimized Thresholds: texture1: 17.2173 | area3: 161.5871 | smoothness3: 1693.5991 | concavity3: 1.0407 | concave_points3: 0.6921 | symmetry3: 0.2743 | lymph_node_status: 6.0650

Train Sens: 0.759 | Train Spec: 0.778 [TRAIN SCORE: 1.536] || Val Sens: 0.667 | Val Spec: 0.600 [VAL SCORE: 1.267] || Test Sens: 0.889 | Test Spec: 0.395 [TEST SCORE: 1.284]

**#164: texture1 + area3 + smoothness3 + concavity3 + concave_points3 + symmetry3 + fractal_dimension3**

Optimized Thresholds: texture1: 17.1848 | area3: 152.6532 | smoothness3: 1731.2902 | concavity3: 0.8500 | concave_points3: 0.6777 | symmetry3: 0.2677 | fractal_dimension3: 0.5139

Train Sens: 0.759 | Train Spec: 0.741 [TRAIN SCORE: 1.499] || Val Sens: 0.667 | Val Spec: 0.600 [VAL SCORE: 1.267] || Test Sens: 0.889 | Test Spec: 0.386 [TEST SCORE: 1.275]

**#165: smoothness3 + concavity3 + concave_points3 + fractal_dimension3 + tumor_size + lymph_node_status**

Optimized Thresholds: smoothness3: 1415.1114 | concavity3: 0.6612 | concave_points3: 0.6909 | fractal_dimension3: 0.4870 | tumor_size: 0.1610 | lymph_node_status: 3.9040

Train Sens: 0.724 | Train Spec: 0.704 [TRAIN SCORE: 1.428] || Val Sens: 0.667 | Val Spec: 0.600 [VAL SCORE: 1.267] || Test Sens: 0.889 | Test Spec: 0.447 [TEST SCORE: 1.336]

**#166: area3 + smoothness3 + concavity3 + concave_points3 + tumor_size + lymph_node_status**

Optimized Thresholds: area3: 213.7848 | smoothness3: 1411.3247 | concavity3: 0.9088 | concave_points3: 0.6884 | tumor_size: 0.1931 | lymph_node_status: 3.8434

Train Sens: 0.690 | Train Spec: 0.741 [TRAIN SCORE: 1.430] || Val Sens: 0.667 | Val Spec: 0.600 [VAL SCORE: 1.267] || Test Sens: 0.889 | Test Spec: 0.465 [TEST SCORE: 1.354]

**#167: area3 + smoothness3 + concavity3 + concave_points3 + lymph_node_status**

Optimized Thresholds: area3: 212.6865 | smoothness3: 1414.9187 | concavity3: 1.0370 | concave_points3: 0.6921 | lymph_node_status: 3.7288

Train Sens: 0.690 | Train Spec: 0.741 [TRAIN SCORE: 1.430] || Val Sens: 0.667 | Val Spec: 0.600 [VAL SCORE: 1.267] || Test Sens: 0.889 | Test Spec: 0.465 [TEST SCORE: 1.354]

**#168: area3 + smoothness3 + concavity3 + concave_points3 + symmetry3 + lymph_node_status**

Optimized Thresholds: area3: 213.7848 | smoothness3: 1411.3247 | concavity3: 0.9088 | concave_points3: 0.6884 | symmetry3: 0.2578 | lymph_node_status: 3.8434

Train Sens: 0.690 | Train Spec: 0.741 [TRAIN SCORE: 1.430] || Val Sens: 0.667 | Val Spec: 0.600 [VAL SCORE: 1.267] || Test Sens: 0.889 | Test Spec: 0.456 [TEST SCORE: 1.345]

**#169: texture1 + smoothness3 + concave_points3 + fractal_dimension3 + tumor_size + lymph_node_status**

Optimized Thresholds: texture1: 17.2485 | smoothness3: 1729.0770 | concave_points3: 0.6743 | fractal_dimension3: 0.5254 | tumor_size: 0.1470 | lymph_node_status: 8.0802

Train Sens: 0.759 | Train Spec: 0.778 [TRAIN SCORE: 1.536] || Val Sens: 0.667 | Val Spec: 0.600 [VAL SCORE: 1.267] || Test Sens: 0.889 | Test Spec: 0.386 [TEST SCORE: 1.275]

**#170: texture1 + smoothness3 + concave_points3 + symmetry3 + tumor_size + lymph_node_status**

Optimized Thresholds: texture1: 17.2338 | smoothness3: 1570.4690 | concave_points3: 0.6628 | symmetry3: 0.2658 | tumor_size: 0.1684 | lymph_node_status: 7.6768

Train Sens: 0.759 | Train Spec: 0.741 [TRAIN SCORE: 1.499] || Val Sens: 0.667 | Val Spec: 0.600 [VAL SCORE: 1.267] || Test Sens: 0.889 | Test Spec: 0.377 [TEST SCORE: 1.266]

**#171: texture1 + smoothness3 + concave_points3 + symmetry3 + fractal_dimension3 + lymph_node_status**

Optimized Thresholds: texture1: 17.2338 | smoothness3: 1570.4690 | concave_points3: 0.6628 | symmetry3: 0.2658 | fractal_dimension3: 0.5483 | lymph_node_status: 7.6768

Train Sens: 0.759 | Train Spec: 0.741 [TRAIN SCORE: 1.499] || Val Sens: 0.667 | Val Spec: 0.600 [VAL SCORE: 1.267] || Test Sens: 0.889 | Test Spec: 0.368 [TEST SCORE: 1.257]

**#172: texture1 + smoothness3 + concavity3 + fractal_dimension3 + tumor_size* + lymph_node_status**

Optimized Thresholds: texture1: 17.2226 | smoothness3: 1620.1902 | concavity3: 0.8200 | fractal_dimension3: 0.5861 | tumor_size*: 0.1426 | lymph_node_status: 8.0719

Train Sens: 0.690 | Train Spec: 0.741 [TRAIN SCORE: 1.430] || Val Sens: 0.667 | Val Spec: 0.600 [VAL SCORE: 1.267] || Test Sens: 0.889 | Test Spec: 0.430 [TEST SCORE: 1.319]

###### #173: texture1 + smoothness3 + concavity3 + symmetry3 + tumor_size* + lymph_node_status*

Optimized Thresholds: texture1: 17.2440 | smoothness3: 1571.3455 | concavity3: 0.8198 | symmetry3: 0.2619 | tumor_size*: 0.1618 | lymph_node_status*: 7.0094

Train Sens: 0.690 | Train Spec: 0.741 [TRAIN SCORE: 1.430] || Val Sens: 0.667 | Val Spec: 0.600 [VAL SCORE: 1.267] || Test Sens: 0.889 | Test Spec: 0.412 [TEST SCORE: 1.301]

###### #174: texture1 + smoothness3 + concavity3 + symmetry3 + fractal_dimension3 + lymph_node_status

Optimized Thresholds: texture1: 17.2401 | smoothness3: 1584.3773 | concavity3: 0.6935 | symmetry3: 0.2622 | fractal_dimension3: 0.5598 | lymph_node_status: 7.1796

Train Sens: 0.690 | Train Spec: 0.741 [TRAIN SCORE: 1.430] || Val Sens: 0.667 | Val Spec: 0.600 [VAL SCORE: 1.267] || Test Sens: 0.889 | Test Spec: 0.395 [TEST SCORE: 1.284]

###### #175: texture1 + smoothness3 + concavity3 + concave_points3 + tumor_size* + lymph_node_status

Optimized Thresholds: texture1: 17.2172 | smoothness3: 1525.9966 | concavity3: 0.8523 | concave_points3: 0.6727 | tumor_size*: 0.1659 | lymph_node_status: 7.2890

Train Sens: 0.759 | Train Spec: 0.741 [TRAIN SCORE: 1.499] || Val Sens: 0.667 | Val Spec: 0.600 [VAL SCORE: 1.267] || Test Sens: 0.889 | Test Spec: 0.386 [TEST SCORE: 1.275]

###### #176: texture1 + smoothness3 + concavity3 + concave_points3 + fractal_dimension3 + lymph_node_status

Optimized Thresholds: texture1: 17.2239 | smoothness3: 1575.7663 | concavity3: 0.8430 | concave_points3: 0.6672 | fractal_dimension3: 0.5709 | lymph_node_status: 7.0356

Train Sens: 0.759 | Train Spec: 0.741 [TRAIN SCORE: 1.499] || Val Sens: 0.667 | Val Spec: 0.600 [VAL SCORE: 1.267] || Test Sens: 0.889 | Test Spec: 0.377 [TEST SCORE: 1.266]

###### #177: texture1 + area3* + smoothness3* + concave_points3 + fractal_dimension3 + tumor_size + lymph_node_status

Optimized Thresholds: texture1: 17.2463 | area3*: 176.6868 | smoothness3*: 2529.9697 | concave_points3: 0.6877 | fractal_dimension3: 0.6236 | tumor_size: 0.1774 | lymph_node_status: 7.2678

Train Sens: 0.724 | Train Spec: 0.778 [TRAIN SCORE: 1.502] || Val Sens: 0.667 | Val Spec: 0.600 [VAL SCORE: 1.267] || Test Sens: 0.889 | Test Spec: 0.395 [TEST SCORE: 1.284]

###### #178: texture1 + area3* + smoothness3* + symmetry3 + fractal_dimension3 + tumor_size* + lymph_node_status

Optimized Thresholds: texture1: 17.2192 | area3*: 178.9905 | smoothness3*: 1953.4585 | symmetry3: 0.2634 | fractal_dimension3: 0.5894 | tumor_size*: 0.1525 | lymph_node_status: 6.8160

Train Sens: 0.655 | Train Spec: 0.778 [TRAIN SCORE: 1.433] || Val Sens: 0.667 | Val Spec: 0.600 [VAL SCORE: 1.267] || Test Sens: 0.889 | Test Spec: 0.430 [TEST SCORE: 1.319]

###### #179: texture1 + area3 + concavity3 + concave_points3 + symmetry3 + fractal_dimension3 + tumor_size

Optimized Thresholds: texture1: 17.2453 | area3: 154.9357 | concavity3: 0.9560 | concave_points3: 0.6870 | symmetry3: 0.2668 | fractal_dimension3: 0.5431 | tumor_size: 0.1600

Train Sens: 0.724 | Train Spec: 0.778 [TRAIN SCORE: 1.502] || Val Sens: 0.667 | Val Spec: 0.600 [VAL SCORE: 1.267] || Test Sens: 0.889 | Test Spec: 0.386 [TEST SCORE: 1.275]

###### #180: texture1 + area3 + concavity3 + concave_points3 + symmetry3 + fractal_dimension3 + lymph_node_status

Optimized Thresholds: texture1: 17.2453 | area3: 154.9357 | concavity3: 0.9560 | concave_points3: 0.6870 | symmetry3: 0.2668 | fractal_dimension3: 0.5431 | lymph_node_status: 6.9003

Train Sens: 0.759 | Train Spec: 0.778 [TRAIN SCORE: 1.536] || Val Sens: 0.667 | Val Spec: 0.600 [VAL SCORE: 1.267] || Test Sens: 0.889 | Test Spec: 0.368 [TEST SCORE: 1.257]

**#181: texture1 + area3 + concavity3 + concave_points3 + symmetry3 + fractal_dimension3 + tumor_size + lymph_node_**

Optimized Thresholds: texture1: 17.2204 | area3: 152.2456 | concavity3: 0.7755 | concave_points3: 0.6596 | symmetry3: 0.2399 | fractal_dimension3: 0.6089 | tumor_size: 0.1908 | lymph_node_status: 6.9731

Train Sens: 0.759 | Train Spec: 0.741 [TRAIN SCORE: 1.499] || Val Sens: 0.667 | Val Spec: 0.600 [VAL SCORE: 1.267] || Test Sens: 0.889 | Test Spec: 0.360 [TEST SCORE: 1.249]

**#182: texture1 + area3 + smoothness3 + concave_points3 + symmetry3 + fractal_dimension3 + tumor_size + lymph_no**

Optimized Thresholds: texture1: 17.2204 | area3: 152.2456 | smoothness3: 2414.6303 | concave_points3: 0.6596 | symmetry3: 0.2399 | fractal_dimension3: 0.6089 | tumor_size: 0.1908 | lymph_node_status: 6.9731

Train Sens: 0.759 | Train Spec: 0.741 [TRAIN SCORE: 1.499] || Val Sens: 0.667 | Val Spec: 0.600 [VAL SCORE: 1.267] || Test Sens: 0.889 | Test Spec: 0.360 [TEST SCORE: 1.249]

**#183: texture1 + area3 + smoothness3* + concavity3 + symmetry3 + fractal_dimension3 + tumor_size + lymph_node_st**

Optimized Thresholds: texture1: 17.2354 | area3: 169.1384 | smoothness3*: 2151.8512 | concavity3: 0.8693 | symmetry3: 0.2654 | fractal_dimension3: 0.5333 | tumor_size: 0.1427 | lymph_node_status: 6.3214

Train Sens: 0.655 | Train Spec: 0.741 [TRAIN SCORE: 1.396] || Val Sens: 0.667 | Val Spec: 0.600 [VAL SCORE: 1.267] || Test Sens: 0.889 | Test Spec: 0.404 [TEST SCORE: 1.292]

**#184: texture1 + area3 + smoothness3 + concavity3 + concave_points3 + fractal_dimension3 + tumor_size + lymph_no**

Optimized Thresholds: texture1: 17.2461 | area3: 174.2563 | smoothness3: 1865.2300 | concavity3: 0.8563 | concave_points3: 0.6676 | fractal_dimension3: 0.5695 | tumor_size: 0.1733 | lymph_node_status: 7.1259

Train Sens: 0.724 | Train Spec: 0.778 [TRAIN SCORE: 1.502] || Val Sens: 0.667 | Val Spec: 0.600 [VAL SCORE: 1.267] || Test Sens: 0.889 | Test Spec: 0.377 [TEST SCORE: 1.266]

**#185: texture1 + area3* + smoothness3 + concavity3 + concave_points3 + symmetry3 + tumor_size + lymph_node_statu**

Optimized Thresholds: texture1: 17.2204 | area3*: 187.2350 | smoothness3: 1850.9876 | concavity3: 0.8080 | concave_points3: 0.6624 | symmetry3: 0.2676 | tumor_size: 0.1842 | lymph_node_status: 7.2741

Train Sens: 0.724 | Train Spec: 0.778 [TRAIN SCORE: 1.502] || Val Sens: 0.667 | Val Spec: 0.600 [VAL SCORE: 1.267] || Test Sens: 0.889 | Test Spec: 0.377 [TEST SCORE: 1.266]

**#186: texture1 + area3* + smoothness3 + concavity3 + concave_points3 + symmetry3 + fractal_dimension3 + lymph_no**

Optimized Thresholds: texture1: 17.2204 | area3*: 187.2350 | smoothness3: 1850.9876 | concavity3: 0.8080 | concave_points3: 0.6624 | symmetry3: 0.2676 | fractal_dimension3: 0.5950 | lymph_node_status: 7.2741

Train Sens: 0.724 | Train Spec: 0.778 [TRAIN SCORE: 1.502] || Val Sens: 0.667 | Val Spec: 0.600 [VAL SCORE: 1.267] || Test Sens: 0.889 | Test Spec: 0.377 [TEST SCORE: 1.266]

**#187: texture1 + area3 + smoothness3 + concavity3 + concave_points3 + symmetry3 + fractal_dimension3 + tumor_size**

Optimized Thresholds: texture1: 17.1751 | area3: 154.6877 | smoothness3: 2343.8434 | concavity3: 0.8082 | concave_points3: 0.6805 | symmetry3: 0.2732 | fractal_dimension3: 0.5306 | tumor_size: 0.1811

Train Sens: 0.759 | Train Spec: 0.741 [TRAIN SCORE: 1.499] || Val Sens: 0.667 | Val Spec: 0.600 [VAL SCORE: 1.267] || Test Sens: 0.889 | Test Spec: 0.395 [TEST SCORE: 1.284]

**#188: area3 + smoothness3 + concavity3* + concave_points3 + fractal_dimension3 + tumor_size + lymph_node_status**

Optimized Thresholds: area3: 169.5026 | smoothness3: 1415.8527 | concavity3*: 0.8288 | concave_points3: 0.6614 | fractal_dimension3: 0.4878 | tumor_size: 0.1769 | lymph_node_status: 3.9254

Train Sens: 0.724 | Train Spec: 0.704 [TRAIN SCORE: 1.428] || Val Sens: 0.667 | Val Spec: 0.600 [VAL SCORE: 1.267] || Test Sens: 0.889 | Test Spec: 0.447 [TEST SCORE: 1.336]

##### #189: area3 + smoothness3 + concavity3 + concave_points3 + symmetry3 + tumor_size + lymph_node_status

Optimized Thresholds: area3: 191.5074 | smoothness3: 1411.5966 | concavity3: 0.7735 | concave_points3: 0.6508 | symmetry3: 0.2543 | tumor_size: 0.1838 | lymph_node_status: 3.8122

Train Sens: 0.690 | Train Spec: 0.704 [TRAIN SCORE: 1.393] || Val Sens: 0.667 | Val Spec: 0.600 [VAL SCORE: 1.267] || Test Sens: 0.889 | Test Spec: 0.447 [TEST SCORE: 1.336]

##### #190: texture1 + smoothness3 + concave_points3 + symmetry3 + fractal_dimension3 + tumor_size + lymph_node_statu

Optimized Thresholds: texture1: 17.2281 | smoothness3: 1701.9053 | concave_points3: 0.6622 | symmetry3: 0.2746 | fractal_dimension3: 0.5018 | tumor_size: 0.1577 | lymph_node_status: 7.6373

Train Sens: 0.759 | Train Spec: 0.778 [TRAIN SCORE: 1.536] || Val Sens: 0.667 | Val Spec: 0.600 [VAL SCORE: 1.267] || Test Sens: 0.889 | Test Spec: 0.377 [TEST SCORE: 1.266]

##### #191: texture1 + smoothness3 + concavity3 + symmetry3 + fractal_dimension3 + tumor_size + lymph_node_status

Optimized Thresholds: texture1: 17.2410 | smoothness3: 1710.5366 | concavity3: 0.8817 | symmetry3: 0.2637 | fractal_dimension3: 0.5786 | tumor_size: 0.1515 | lymph_node_status: 5.9107

Train Sens: 0.724 | Train Spec: 0.741 [TRAIN SCORE: 1.465] || Val Sens: 0.667 | Val Spec: 0.600 [VAL SCORE: 1.267] || Test Sens: 0.889 | Test Spec: 0.412 [TEST SCORE: 1.301]

##### #192: texture1 + smoothness3 + concavity3 + concave_points3 + fractal_dimension3 + tumor_size + lymph_node_statu

Optimized Thresholds: texture1: 17.2599 | smoothness3: 1717.8079 | concavity3: 0.8157 | concave_points3: 0.6769 | fractal_dimension3: 0.5807 | tumor_size: 0.1612 | lymph_node_status: 7.6130

Train Sens: 0.759 | Train Spec: 0.778 [TRAIN SCORE: 1.536] || Val Sens: 0.667 | Val Spec: 0.600 [VAL SCORE: 1.267] || Test Sens: 0.889 | Test Spec: 0.386 [TEST SCORE: 1.275]

##### #193: texture1 + smoothness3 + concavity3 + concave_points3 + symmetry3 + tumor_size + lymph_node_status

Optimized Thresholds: texture1: 17.2526 | smoothness3: 1711.7113 | concavity3: 0.8506 | concave_points3: 0.6790 | symmetry3: 0.2667 | tumor_size: 0.1649 | lymph_node_status: 7.7884

Train Sens: 0.759 | Train Spec: 0.778 [TRAIN SCORE: 1.536] || Val Sens: 0.667 | Val Spec: 0.600 [VAL SCORE: 1.267] || Test Sens: 0.889 | Test Spec: 0.377 [TEST SCORE: 1.266]

##### #194: texture1 + smoothness3 + concavity3 + concave_points3 + symmetry3 + fractal_dimension3 + lymph_node_statu

Optimized Thresholds: texture1: 17.2526 | smoothness3: 1711.7113 | concavity3: 0.8506 | concave_points3: 0.6790 | symmetry3: 0.2667 | fractal_dimension3: 0.5379 | lymph_node_status: 7.7884

Train Sens: 0.759 | Train Spec: 0.778 [TRAIN SCORE: 1.536] || Val Sens: 0.667 | Val Spec: 0.600 [VAL SCORE: 1.267] || Test Sens: 0.889 | Test Spec: 0.368 [TEST SCORE: 1.257]

##### #195: texture1 + smoothness3 + concavity3 + concave_points3 + symmetry3 + fractal_dimension3 + tumor_size

Optimized Thresholds: texture1: 17.2526 | smoothness3: 1711.7113 | concavity3: 0.8506 | concave_points3: 0.6790 | symmetry3: 0.2667 | fractal_dimension3: 0.5379 | tumor_size: 0.1736

Train Sens: 0.724 | Train Spec: 0.778 [TRAIN SCORE: 1.502] || Val Sens: 0.667 | Val Spec: 0.600 [VAL SCORE: 1.267] || Test Sens: 0.889 | Test Spec: 0.386 [TEST SCORE: 1.275]

**#196: texture1 + area3 + concave_points3 + symmetry3 + fractal_dimension3 + tumor_size* + lymph_node_status**

Optimized Thresholds: texture1: 17.1842 | area3: 154.5597 | concave_points3: 0.6669 | symmetry3: 0.2665 | fractal_dimension3: 0.5685 | tumor_size*: 0.1474 | lymph_node_status: 8.6095

Train Sens: 0.759 | Train Spec: 0.741 [TRAIN SCORE: 1.499] || Val Sens: 0.667 | Val Spec: 0.600 [VAL SCORE: 1.267] || Test Sens: 0.889 | Test Spec: 0.377 [TEST SCORE: 1.266]

**#197: texture1 + area3 + concavity3 + symmetry3 + fractal_dimension3 + tumor_size* + lymph_node_status**

Optimized Thresholds: texture1: 17.2208 | area3: 154.3230 | concavity3: 0.8566 | symmetry3: 0.2614 | fractal_dimension3: 0.6074 | tumor_size*: 0.1556 | lymph_node_status: 6.2164

Train Sens: 0.690 | Train Spec: 0.741 [TRAIN SCORE: 1.430] || Val Sens: 0.667 | Val Spec: 0.600 [VAL SCORE: 1.267] || Test Sens: 0.889 | Test Spec: 0.412 [TEST SCORE: 1.301]

**#198: texture1 + area3 + concavity3 + concave_points3 + fractal_dimension3 + tumor_size + lymph_node_status**

Optimized Thresholds: texture1: 17.2442 | area3: 155.1180 | concavity3: 0.9526 | concave_points3: 0.6891 | fractal_dimension3: 0.6189 | tumor_size: 0.1769 | lymph_node_status: 6.7573

Train Sens: 0.759 | Train Spec: 0.778 [TRAIN SCORE: 1.536] || Val Sens: 0.667 | Val Spec: 0.600 [VAL SCORE: 1.267] || Test Sens: 0.889 | Test Spec: 0.395 [TEST SCORE: 1.284]

**#199: texture1 + area3 + concavity3 + concave_points3 + symmetry3 + tumor_size + lymph_node_status**

Optimized Thresholds: texture1: 17.2453 | area3: 154.9357 | concavity3: 0.9560 | concave_points3: 0.6870 | symmetry3: 0.2668 | tumor_size: 0.1666 | lymph_node_status: 6.9003

Train Sens: 0.759 | Train Spec: 0.778 [TRAIN SCORE: 1.536] || Val Sens: 0.667 | Val Spec: 0.600 [VAL SCORE: 1.267] || Test Sens: 0.889 | Test Spec: 0.377 [TEST SCORE: 1.266]

**#200: texture1 + smoothness3 + concavity3 + concave_points3 + fractal_dimension3 + tumor_size**

Optimized Thresholds: texture1: 17.1796 | smoothness3: 1634.8092 | concavity3: 0.9601 | concave_points3: 0.6625 | fractal_dimension3: 0.5592 | tumor_size: 0.1508

Train Sens: 0.759 | Train Spec: 0.741 [TRAIN SCORE: 1.499] || Val Sens: 0.667 | Val Spec: 0.600 [VAL SCORE: 1.267] || Test Sens: 0.889 | Test Spec: 0.395 [TEST SCORE: 1.284]
