## Supplementary material. Outputs TholdStormDX for this study. for "Methodological and Clinical Validation of TholdStormDX v0.0.1: An Advanced Stochastic Engine for the Optimization of Thresholds and Multimarker Panels Applied to Oncology": Cervical TholdStormDX_RobustReport_20260331_224237.pdf

### Biomarker: Age

Processed: 31-Mar-2026 21:41

#### 1. Optimization Results

| MODEL | CUT-OFF | TRAIN (SE/SP) | VAL (SE/SP) | TEST (SE/SP) | R2 SCORE |
|---|---|---|---|---|---|
| Empirical (Exact) | 28.3083 | 0.464 / 0.464 | 0.444 / 0.333 | 0.500 / 0.626 | N/A |
| Logistic 2-Parameter | 28.6998 | 0.475 / 0.475 | 0.444 / 0.333 | 0.500 / 0.626 | 0.9605 |
| **Logistic 4-Parameter (Rec.)** | **28.2029** | **0.457 / 0.457** | **0.444 / 0.333** | **0.500 / 0.626** | **0.9765** |
| ThresholdXpert (Stochastic) | 25.6245 | 0.538 / 0.464 | 0.556 / 0.222 | 0.600 / 0.491 | N/A |

#### 2. Diagnostic Performance Curves (Training)

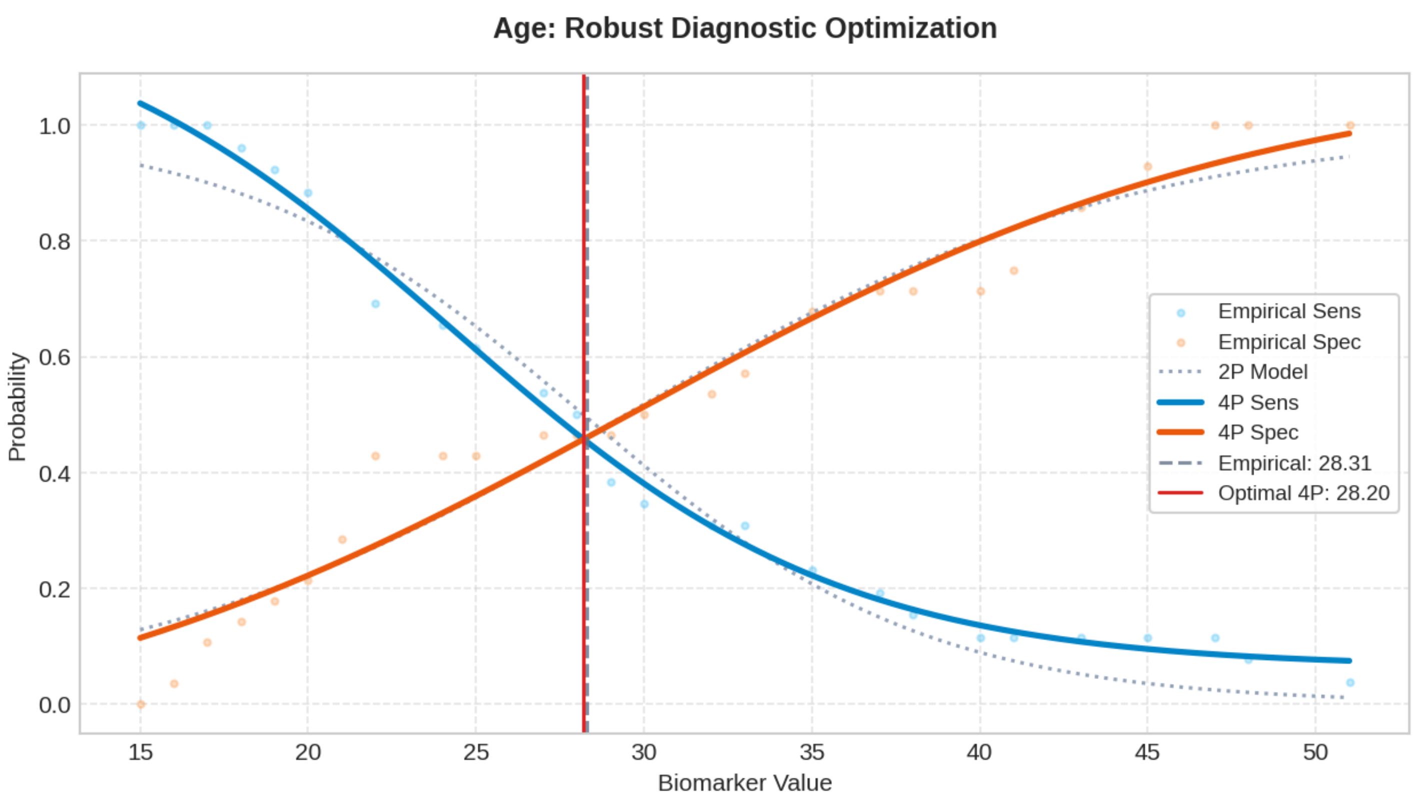

### Biomarker: Number of sexual partners

Processed: 31-Mar-2026 21:43

#### 1. Optimization Results

| MODEL | CUT-OFF | TRAIN (SE/SP) | VAL (SE/SP) | TEST (SE/SP) | R2 SCORE |
|---|---|---|---|---|---|
| Empirical (Exact) | 2.7914 | 0.563 / 0.563 | 0.333 / 0.667 | 0.300 / 0.561 | N/A |
| Logistic 2-Parameter | 2.7960 | 0.557 / 0.557 | 0.333 / 0.667 | 0.300 / 0.561 | 0.9680 |
| **Logistic 4-Parameter (Rec.)** | **2.7222** | **0.546 / 0.546** | **0.333 / 0.667** | **0.300 / 0.561** | **0.9798** |
| ThresholdXpert (Stochastic) | 2.4916 | 0.538 / 0.607 | 0.333 / 0.667 | 0.300 / 0.561 | N/A |

#### 2. Diagnostic Performance Curves (Training)

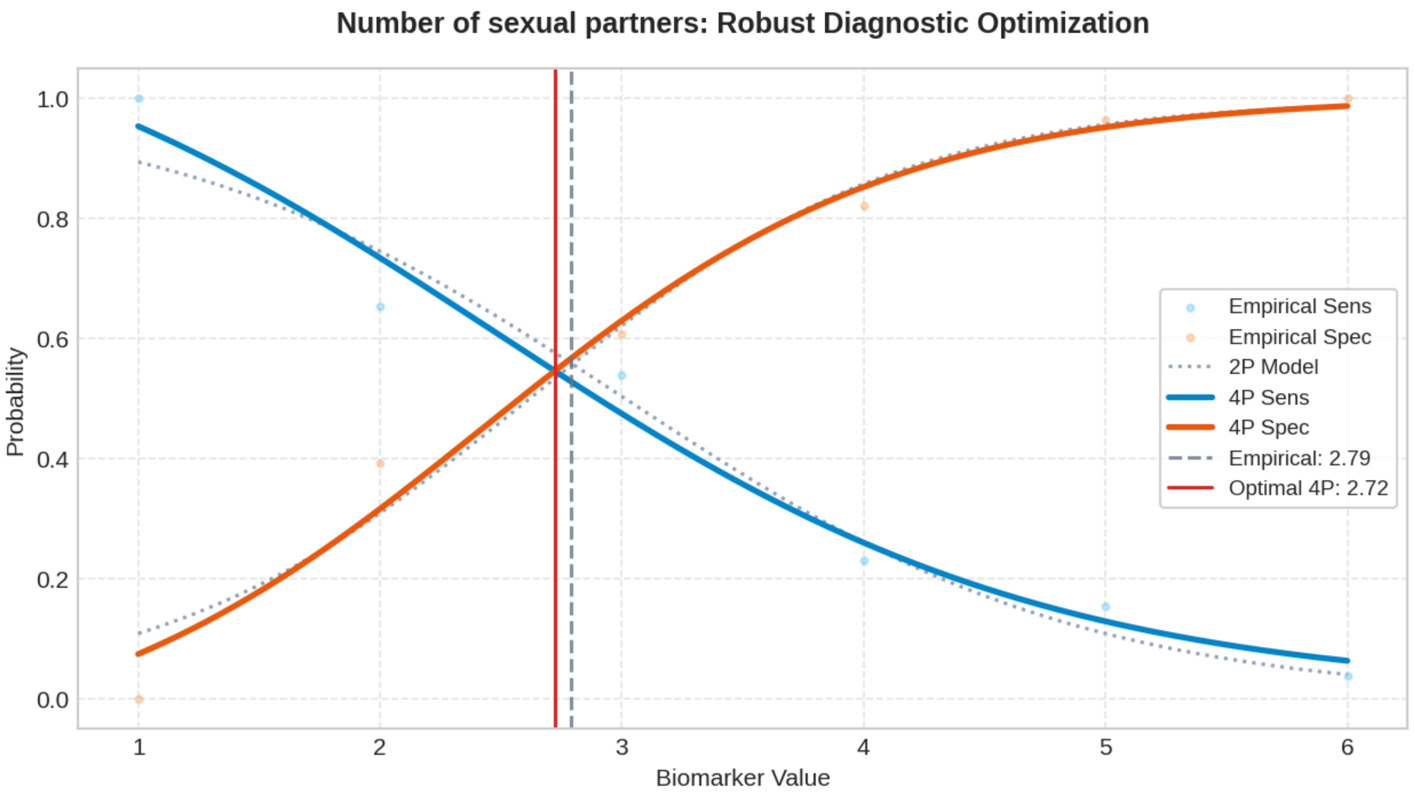

### Biomarker: First sexual intercourse

Processed: 31-Mar-2026 21:45

#### 1. Optimization Results

| MODEL | CUT-OFF | TRAIN (SE/SP) | VAL (SE/SP) | TEST (SE/SP) | R2 SCORE |
|---|---|---|---|---|---|
| Empirical (Exact) | 17.5047 | 0.537 / 0.537 | 0.667 / 0.667 | 0.400 / 0.628 | N/A |
| Logistic 2-Parameter | 17.5827 | 0.523 / 0.523 | 0.667 / 0.667 | 0.400 / 0.628 | 0.9871 |
| **Logistic 4-Parameter (Rec.)** | **17.5197** | **0.524 / 0.524** | **0.667 / 0.667** | **0.400 / 0.628** | **0.9930** |
| ThresholdXpert (Stochastic) | 16.3017 | 0.692 / 0.429 | 0.778 / 0.444 | 0.700 / 0.461 | N/A |

#### 2. Diagnostic Performance Curves (Training)

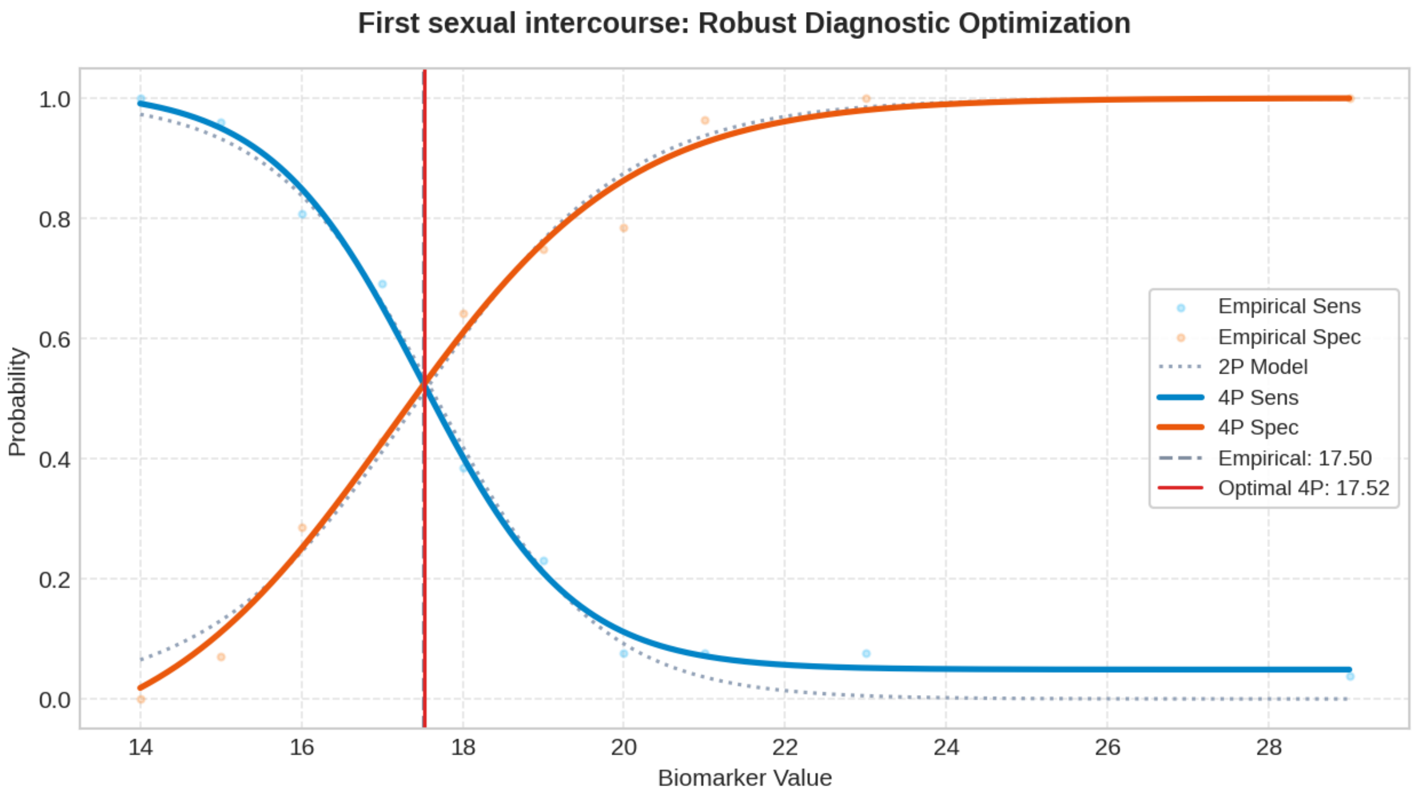

### Biomarker: Num of pregnancies

Processed: 31-Mar-2026 21:47

#### 1. Optimization Results

| MODEL | CUT-OFF | TRAIN (SE/SP) | VAL (SE/SP) | TEST (SE/SP) | R2 SCORE |
|---|---|---|---|---|---|
| Empirical (Exact) | 2.6437 | 0.505 / 0.505 | 0.778 / 0.778 | 0.500 / 0.652 | N/A |
| Logistic 2-Parameter | 2.7533 | 0.502 / 0.502 | 0.778 / 0.778 | 0.500 / 0.652 | 0.9861 |
| **Logistic 4-Parameter (Rec.)** | **2.6705** | **0.510 / 0.510** | **0.778 / 0.778** | **0.500 / 0.652** | **0.9944** |
| ThresholdXpert (Stochastic) | 2.4454 | 0.423 / 0.607 | 0.778 / 0.778 | 0.500 / 0.652 | N/A |

#### 2. Diagnostic Performance Curves (Training)

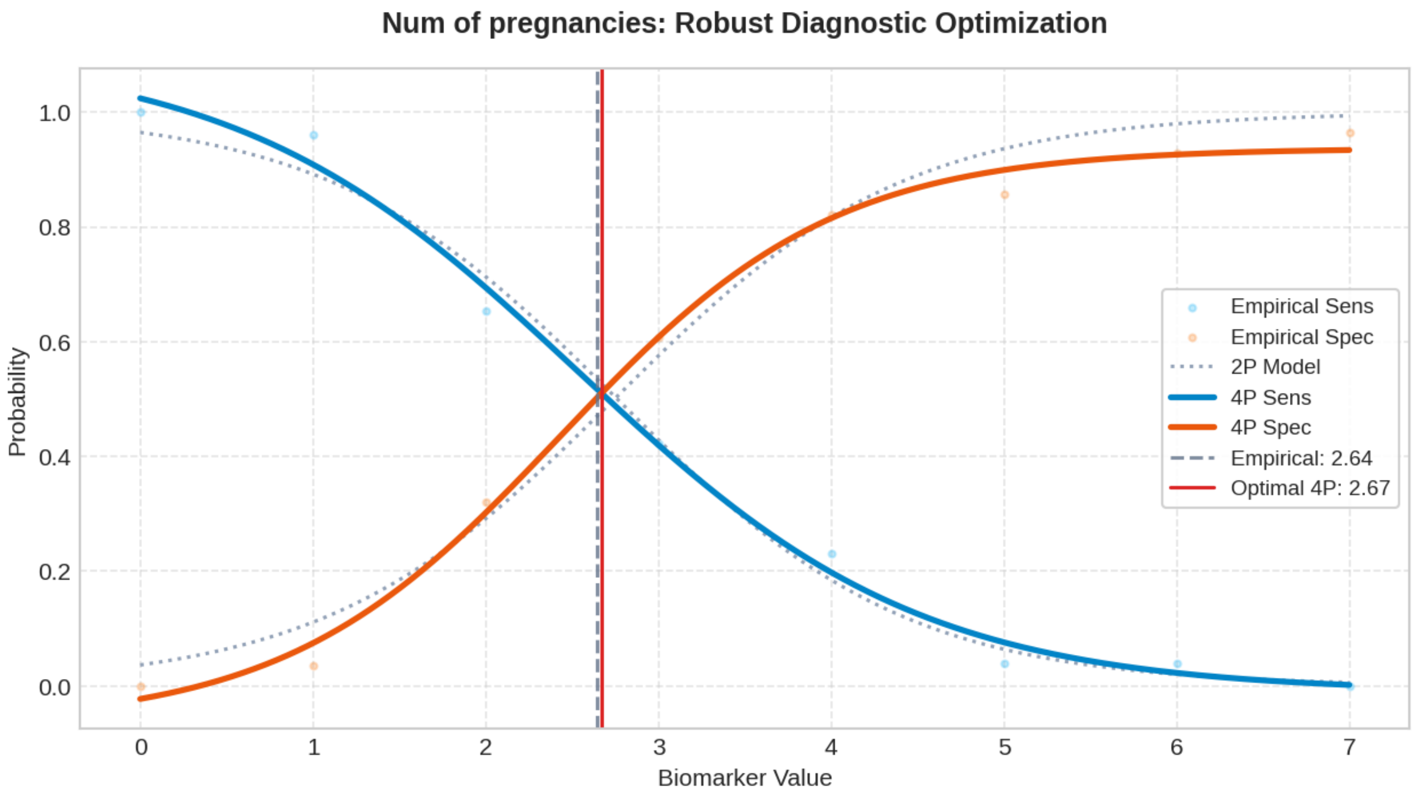

### Biomarker: Smokes (packs/year)

Processed: 31-Mar-2026 21:50

#### 1. Optimization Results

| MODEL | CUT-OFF | TRAIN (SE/SP) | VAL (SE/SP) | TEST (SE/SP) | R2 SCORE |
|---|---|---|---|---|---|
| Empirical (Exact) | 0.0294 | 0.525 / 0.525 | 0.222 / 0.778 | 0.100 / 0.862 | N/A |
| Logistic 2-Parameter | 0.0480 | 0.835 / 0.835 | 0.222 / 0.778 | 0.100 / 0.863 | 0.9540 |
| **Logistic 4-Parameter (Rec.)** | **0.0304** | **0.619 / 0.619** | **0.222 / 0.778** | **0.100 / 0.862** | **0.9889** |
| ThresholdXpert (Stochastic) | 0.0295 | 0.192 / 0.929 | 0.222 / 0.778 | 0.100 / 0.862 | N/A |

#### 2. Diagnostic Performance Curves (Training)

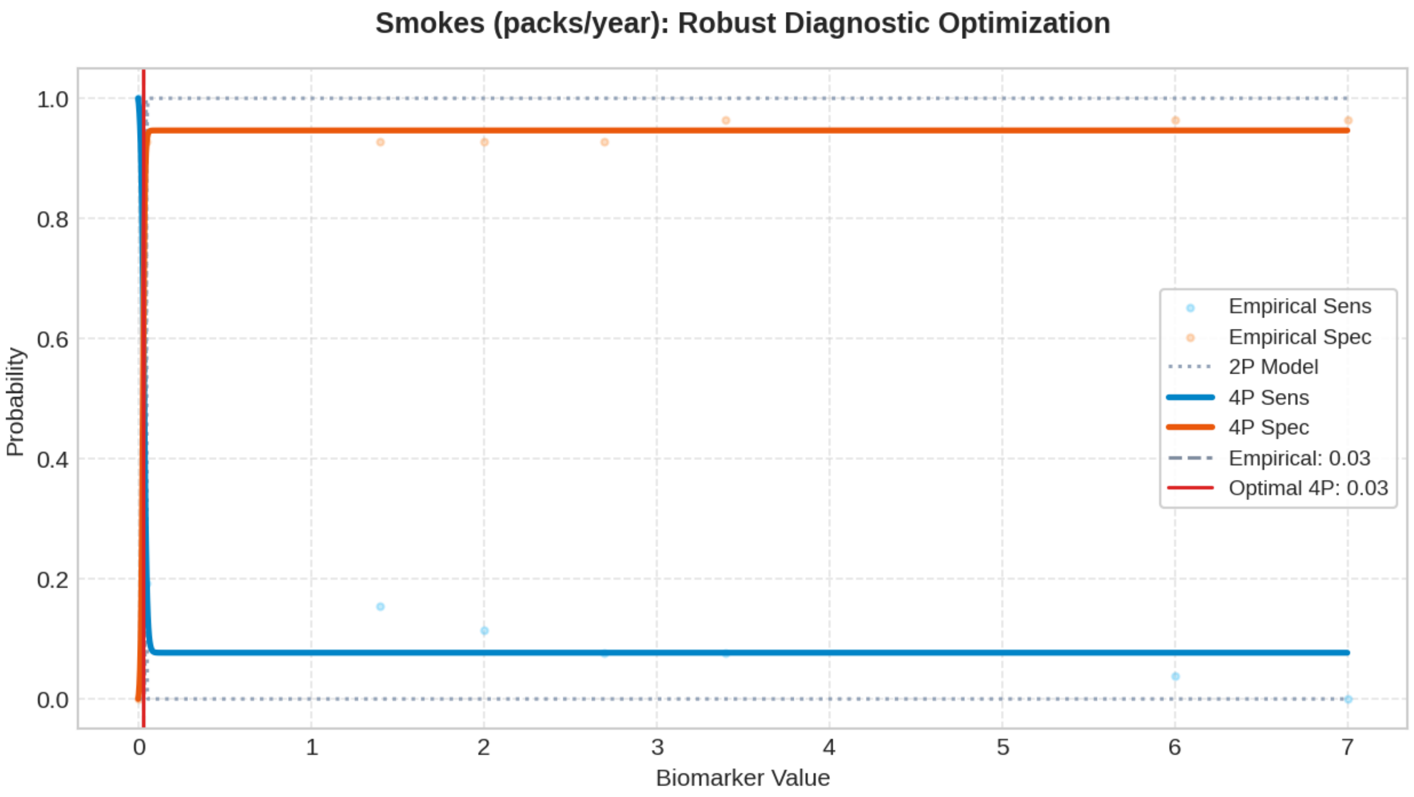

### Biomarker: Hormonal Contraceptives (years)

Processed: 31-Mar-2026 21:53

#### 1. Optimization Results

| MODEL | CUT-OFF | TRAIN (SE/SP) | VAL (SE/SP) | TEST (SE/SP) | R2 SCORE |
|---|---|---|---|---|---|
| Empirical (Exact) | 0.5897 | 0.572 / 0.572 | 0.222 / 0.667 | 0.400 / 0.524 | N/A |
| Logistic 2-Parameter | 1.7183 | 0.534 / 0.534 | 0.222 / 0.667 | 0.300 / 0.659 | 0.8007 |
| **Logistic 4-Parameter (Rec.)** | **0.6812** | **0.625 / 0.625** | **0.222 / 0.667** | **0.400 / 0.534** | **0.8438** |
| ThresholdXpert (Stochastic) | 0.3611 | 0.654 / 0.571 | 0.333 / 0.556 | 0.500 / 0.476 | N/A |

#### 2. Diagnostic Performance Curves (Training)

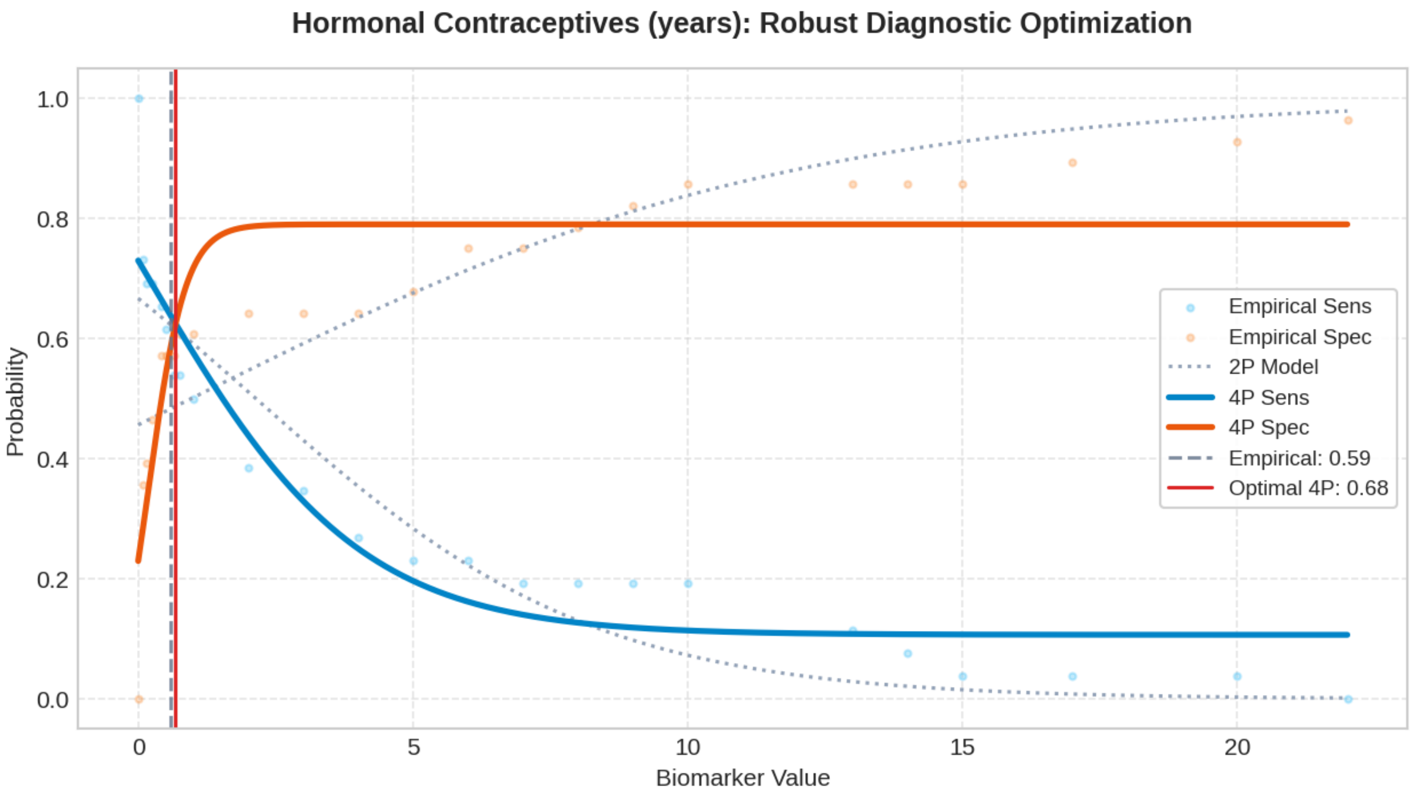

### Biomarker: IUD (years)

Processed: 31-Mar-2026 21:55

#### 1. Optimization Results

| MODEL | CUT-OFF | TRAIN (SE/SP) | VAL (SE/SP) | TEST (SE/SP) | R2 SCORE |
|---|---|---|---|---|---|
| Empirical (Exact) | 0.3403 | 0.504 / 0.504 | 0.444 / 0.778 | 0.100 / 0.908 | N/A |
| Logistic 2-Parameter | 0.4108 | 0.494 / 0.494 | 0.444 / 0.778 | 0.100 / 0.908 | 0.9555 |
| **Logistic 4-Parameter (Rec.)** | **0.3809** | **0.611 / 0.611** | **0.444 / 0.778** | **0.100 / 0.908** | **0.9904** |
| ThresholdXpert (Stochastic) | 0.7782 | 0.154 / 0.893 | 0.444 / 0.778 | 0.100 / 0.911 | N/A |

#### 2. Diagnostic Performance Curves (Training)

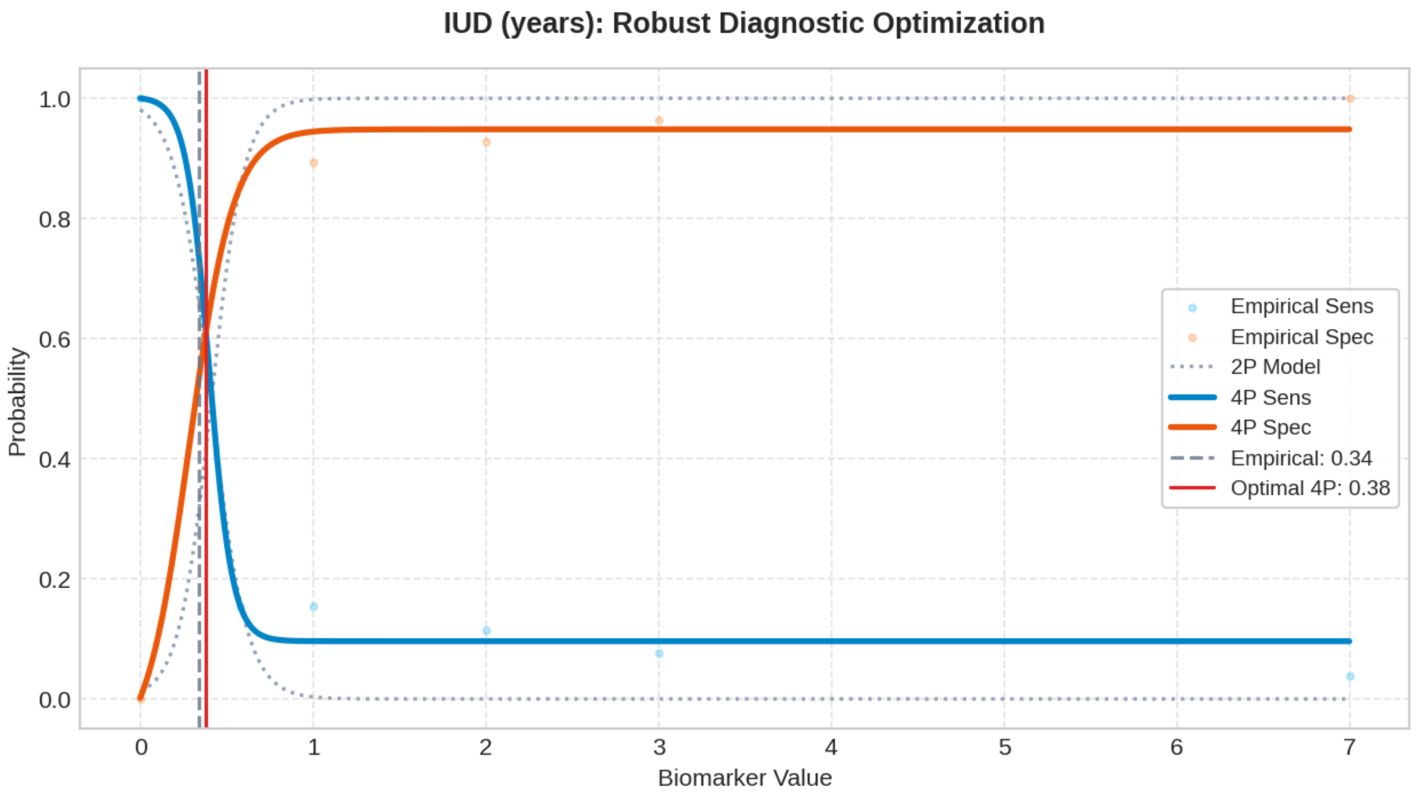

### Biomarker: STDs (number)

Processed: 31-Mar-2026 21:57

#### 1. Optimization Results

| MODEL | CUT-OFF | TRAIN (SE/SP) | VAL (SE/SP) | TEST (SE/SP) | R2 SCORE |
|---|---|---|---|---|---|
| Empirical (Exact) | 0.5887 | 0.547 / 0.547 | 0.333 / 0.889 | 0.100 / 0.911 | N/A |
| Logistic 2-Parameter | 0.7614 | 0.525 / 0.525 | 0.333 / 0.889 | 0.100 / 0.911 | 0.9745 |
| **Logistic 4-Parameter (Rec.)** | **0.9080** | **0.516 / 0.516** | **0.333 / 0.889** | **0.100 / 0.911** | **0.9923** |
| ThresholdXpert (Stochastic) | 0.5024 | 0.231 / 0.929 | 0.333 / 0.889 | 0.100 / 0.911 | N/A |

#### 2. Diagnostic Performance Curves (Training)

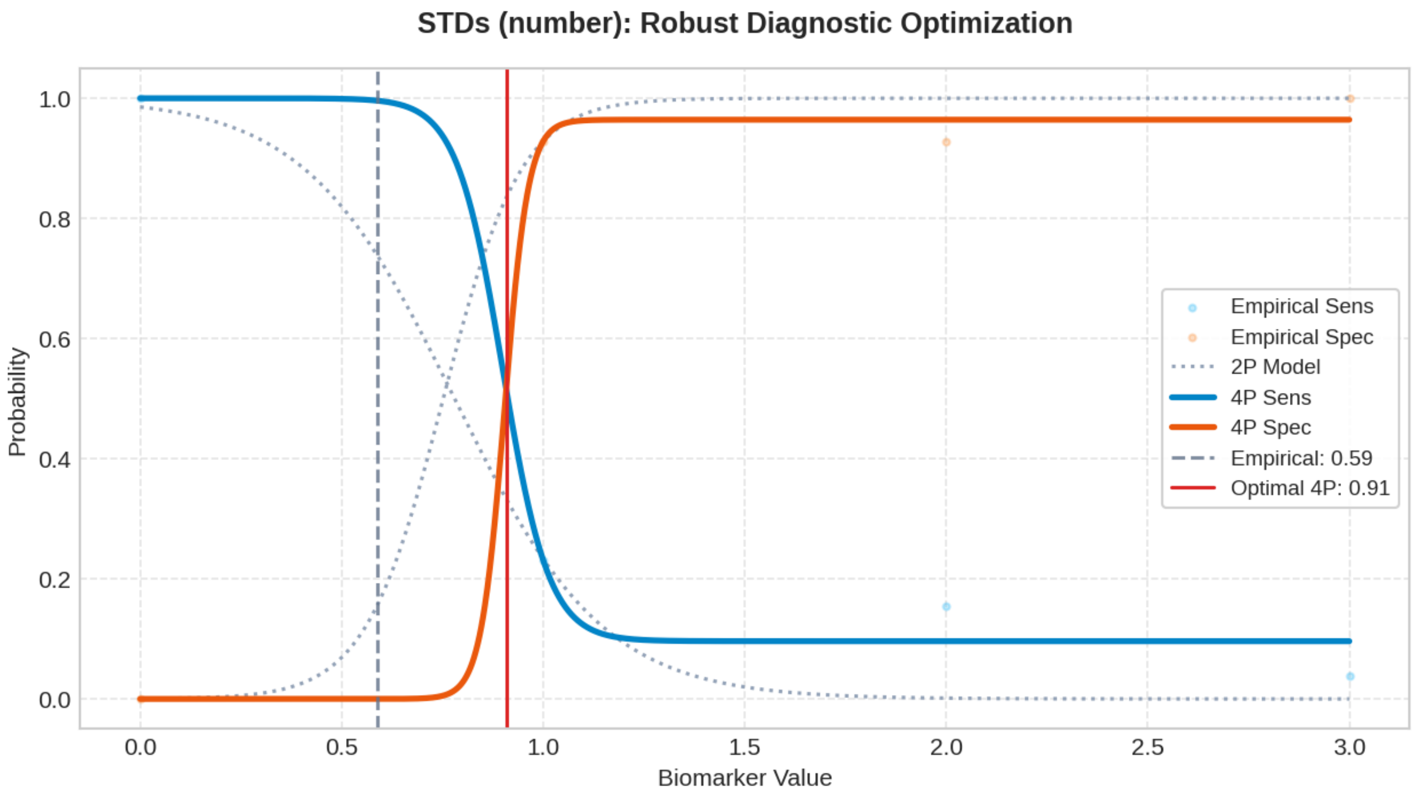

#### Top 200 Combinatorial Panels (ThresholdXpert OR-Logic)

The following multimarker panels have been optimized using high-performance vector-driven Monte Carlo simulations under a Boolean OR-logic framework. The engine employs a Max-Min Balancing logic (0.001 precision) to identify global threshold configurations that maximize the equilibrium between Sensitivity and Specificity across up to 10 million iterations. To ensure clinical robustness, results are sorted strictly by Validation Performance. (* Asterisk indicates an algorithmic threshold instability > 15%, suggesting potential data sparsity or high variance in the stochastic averaging process).

##### #1: Num of pregnancies + IUD (years)* + STDs (number)

Optimized Thresholds: Num of pregnancies: 2.4778 | IUD (years)*: 3.0856 | STDs (number): 0.6346

Train Sens: 0.615 | Train Spec: 0.536 [TRAIN SCORE: 1.151] || Val Sens: 1.000 | Val Spec: 0.667 [VAL SCORE: 1.667] || Test Sens: 0.600 | Test Spec: 0.578 [TEST SCORE: 1.178]

##### #2: Num of pregnancies + STDs (number)

Optimized Thresholds: Num of pregnancies: 2.5698 | STDs (number): 0.5168

Train Sens: 0.615 | Train Spec: 0.536 [TRAIN SCORE: 1.151] || Val Sens: 1.000 | Val Spec: 0.667 [VAL SCORE: 1.667] || Test Sens: 0.600 | Test Spec: 0.601 [TEST SCORE: 1.201]

##### #3: First sexual intercourse + Num of pregnancies + Hormonal Contraceptives (years) + STDs (number)

Optimized Thresholds: First sexual intercourse: 25.5591 | Num of pregnancies: 3.4670 | Hormonal Contraceptives (years): 10.8063 | STDs (number): 0.4210

Train Sens: 0.577 | Train Spec: 0.679 [TRAIN SCORE: 1.255] || Val Sens: 0.778 | Val Spec: 0.889 [VAL SCORE: 1.667] || Test Sens: 0.400 | Test Spec: 0.715 [TEST SCORE: 1.115]

##### #4: Num of pregnancies + Smokes (packs/year)* + STDs (number)

Optimized Thresholds: Num of pregnancies: 2.4778 | Smokes (packs/year)*: 3.0856 | STDs (number): 0.6346

Train Sens: 0.615 | Train Spec: 0.536 [TRAIN SCORE: 1.151] || Val Sens: 1.000 | Val Spec: 0.556 [VAL SCORE: 1.556] || Test Sens: 0.600 | Test Spec: 0.587 [TEST SCORE: 1.187]

##### #5: Number of sexual partners* + Num of pregnancies* + Hormonal Contraceptives (years)* + IUD (years)

Optimized Thresholds: Number of sexual partners*: 4.0701 | Num of pregnancies*: 3.3718 | Hormonal Contraceptives (years)*: 13.7431 | IUD (years): 1.0214

Train Sens: 0.423 | Train Spec: 0.679 [TRAIN SCORE: 1.102] || Val Sens: 0.778 | Val Spec: 0.778 [VAL SCORE: 1.556] || Test Sens: 0.400 | Test Spec: 0.701 [TEST SCORE: 1.101]

##### #6: Age + Number of sexual partners + First sexual intercourse + Smokes (packs/year) + Hormonal Contraceptives (yea

Optimized Thresholds: Age: 48.9232 | Number of sexual partners: 4.5042 | First sexual intercourse: 25.8064 | Smokes (packs/year): 1.1201 | Hormonal Contraceptives (years): 9.5310 | STDs (number): 0.3646

Train Sens: 0.654 | Train Spec: 0.714 [TRAIN SCORE: 1.368] || Val Sens: 0.778 | Val Spec: 0.778 [VAL SCORE: 1.556] || Test Sens: 0.300 | Test Spec: 0.727 [TEST SCORE: 1.027]

##### #7: Age + Smokes (packs/year) + STDs (number)

Optimized Thresholds: Age: 34.0554 | Smokes (packs/year): 0.9179 | STDs (number): 0.5015

Train Sens: 0.500 | Train Spec: 0.607 [TRAIN SCORE: 1.107] || Val Sens: 0.778 | Val Spec: 0.778 [VAL SCORE: 1.556] || Test Sens: 0.400 | Test Spec: 0.701 [TEST SCORE: 1.101]

##### #8: Age + Number of sexual partners + Num of pregnancies + Smokes (packs/year) + Hormonal Contraceptives (years)

Optimized Thresholds: Age: 48.2880 | Number of sexual partners: 3.5143 | Num of pregnancies: 6.2501 | Smokes (packs/year): 0.7505 | Hormonal Contraceptives (years): 9.5359 | STDs (number): 0.4261

Train Sens: 0.654 | Train Spec: 0.643 [TRAIN SCORE: 1.297] || Val Sens: 0.778 | Val Spec: 0.778 [VAL SCORE: 1.556] || Test Sens: 0.500 | Test Spec: 0.679 [TEST SCORE: 1.179]

##### #9: Age + Num of pregnancies + Smokes (packs/year) + Hormonal Contraceptives (years) + STDs (number)

Optimized Thresholds: Age: 47.1286 | Num of pregnancies: 3.5790 | Smokes (packs/year): 0.8309 | Hormonal Contraceptives (years): 11.3924 | STDs (number): 0.4747

Train Sens: 0.577 | Train Spec: 0.643 [TRAIN SCORE: 1.220] || Val Sens: 0.778 | Val Spec: 0.778 [VAL SCORE: 1.556] || Test Sens: 0.500 | Test Spec: 0.698 [TEST SCORE: 1.198]

##### #10: Age + First sexual intercourse* + Num of pregnancies + Smokes (packs/year) + Hormonal Contraceptives (years) +

Optimized Thresholds: Age: 47.2834 | First sexual intercourse*: 25.4778 | Num of pregnancies: 3.5473 | Smokes (packs/year): 1.2712 | Hormonal Contraceptives (years): 12.1420 | STDs (number): 0.6011

Train Sens: 0.615 | Train Spec: 0.643 [TRAIN SCORE: 1.258] || Val Sens: 0.778 | Val Spec: 0.778 [VAL SCORE: 1.556] || Test Sens: 0.400 | Test Spec: 0.683 [TEST SCORE: 1.083]

##### #11: Age + Number of sexual partners + Smokes (packs/year) + Hormonal Contraceptives (years) + STDs (number)

Optimized Thresholds: Age: 47.6517 | Number of sexual partners: 3.5161 | Smokes (packs/year): 0.8969 | Hormonal Contraceptives (years): 9.6799 | STDs (number): 0.4842

Train Sens: 0.654 | Train Spec: 0.643 [TRAIN SCORE: 1.297] || Val Sens: 0.778 | Val Spec: 0.778 [VAL SCORE: 1.556] || Test Sens: 0.500 | Test Spec: 0.688 [TEST SCORE: 1.188]

##### #12: Age + Number of sexual partners + Num of pregnancies + Smokes (packs/year) + STDs (number)*

Optimized Thresholds: Age: 44.6280 | Number of sexual partners: 3.5166 | Num of pregnancies: 3.7797 | Smokes (packs/year): 0.7339 | STDs (number)*: 0.6448

Train Sens: 0.615 | Train Spec: 0.607 [TRAIN SCORE: 1.223] || Val Sens: 0.778 | Val Spec: 0.778 [VAL SCORE: 1.556] || Test Sens: 0.500 | Test Spec: 0.597 [TEST SCORE: 1.097]

##### #13: Num of pregnancies + IUD (years)

Optimized Thresholds: Num of pregnancies: 2.5772 | IUD (years): 1.1189

Train Sens: 0.462 | Train Spec: 0.607 [TRAIN SCORE: 1.069] || Val Sens: 0.778 | Val Spec: 0.778 [VAL SCORE: 1.556] || Test Sens: 0.500 | Test Spec: 0.619 [TEST SCORE: 1.119]

##### #14: Number of sexual partners + Num of pregnancies + Smokes (packs/year) + STDs (number)

Optimized Thresholds: Number of sexual partners: 3.3837 | Num of pregnancies: 3.4343 | Smokes (packs/year): 0.9991 | STDs (number): 0.5639

Train Sens: 0.577 | Train Spec: 0.607 [TRAIN SCORE: 1.184] || Val Sens: 0.778 | Val Spec: 0.778 [VAL SCORE: 1.556] || Test Sens: 0.500 | Test Spec: 0.611 [TEST SCORE: 1.111]

##### #15: First sexual intercourse* + Num of pregnancies + Smokes (packs/year) + Hormonal Contraceptives (years) + STDs

Optimized Thresholds: First sexual intercourse*: 25.2397 | Num of pregnancies: 3.5135 | Smokes (packs/year): 1.0603 | Hormonal Contraceptives (years): 11.1260 | STDs (number): 0.5242

Train Sens: 0.615 | Train Spec: 0.643 [TRAIN SCORE: 1.258] || Val Sens: 0.778 | Val Spec: 0.778 [VAL SCORE: 1.556] || Test

Sens: 0.500 | Test Spec: 0.679 [TEST SCORE: 1.179]

##### #16: Age + Number of sexual partners + First sexual intercourse + Num of pregnancies + Smokes (packs/year) + Hormo

Optimized Thresholds: Age: 48.2090 | Number of sexual partners: 4.5342 | First sexual intercourse: 24.5757 | Num of pregnancies: 5.7641 | Smokes (packs/year): 0.9806 | Hormonal Contraceptives (years): 9.3242 | STDs (number): 0.3403

Train Sens: 0.654 | Train Spec: 0.714 [TRAIN SCORE: 1.368] || Val Sens: 0.778 | Val Spec: 0.778 [VAL SCORE: 1.556] || Test Sens: 0.300 | Test Spec: 0.701 [TEST SCORE: 1.001]

##### #17: Age + Number of sexual partners + Num of pregnancies + Smokes (packs/year) + Hormonal Contraceptives (years

Optimized Thresholds: Age: 48.1366 | Number of sexual partners: 3.4582 | Num of pregnancies: 5.8743 | Smokes (packs/year): 0.9949 | Hormonal Contraceptives (years): 10.0426 | IUD (years)*: 2.7632 | STDs (number): 0.5508

Train Sens: 0.615 | Train Spec: 0.643 [TRAIN SCORE: 1.258] || Val Sens: 1.000 | Val Spec: 0.556 [VAL SCORE: 1.556] || Test Sens: 0.600 | Test Spec: 0.637 [TEST SCORE: 1.237]

##### #18: Num of pregnancies + Smokes (packs/year) + Hormonal Contraceptives (years) + STDs (number)

Optimized Thresholds: Num of pregnancies: 3.5769 | Smokes (packs/year): 1.0727 | Hormonal Contraceptives (years): 11.5617 | STDs (number): 0.4534

Train Sens: 0.577 | Train Spec: 0.643 [TRAIN SCORE: 1.220] || Val Sens: 0.778 | Val Spec: 0.778 [VAL SCORE: 1.556] || Test Sens: 0.500 | Test Spec: 0.706 [TEST SCORE: 1.206]

##### #19: Age + Number of sexual partners + First sexual intercourse + Num of pregnancies + Hormonal Contraceptives (yea

Optimized Thresholds: Age: 48.8821 | Number of sexual partners: 4.0756 | First sexual intercourse: 25.8413 | Num of pregnancies: 5.9295 | Hormonal Contraceptives (years): 7.9538 | STDs (number): 0.6015

Train Sens: 0.615 | Train Spec: 0.679 [TRAIN SCORE: 1.294] || Val Sens: 0.667 | Val Spec: 0.889 [VAL SCORE: 1.556] || Test Sens: 0.300 | Test Spec: 0.715 [TEST SCORE: 1.015]

##### #20: Age + Number of sexual partners + First sexual intercourse* + Hormonal Contraceptives (years) + STDs (number)

Optimized Thresholds: Age: 46.9926 | Number of sexual partners: 4.1458 | First sexual intercourse*: 24.6275 | Hormonal Contraceptives (years): 7.3809 | STDs (number): 0.4563

Train Sens: 0.615 | Train Spec: 0.679 [TRAIN SCORE: 1.294] || Val Sens: 0.667 | Val Spec: 0.889 [VAL SCORE: 1.556] || Test Sens: 0.300 | Test Spec: 0.730 [TEST SCORE: 1.030]

##### #21: Age + Number of sexual partners + Hormonal Contraceptives (years) + STDs (number)

Optimized Thresholds: Age: 48.5184 | Number of sexual partners: 4.2169 | Hormonal Contraceptives (years): 7.0578 | STDs (number): 0.4937

Train Sens: 0.577 | Train Spec: 0.679 [TRAIN SCORE: 1.255] || Val Sens: 0.667 | Val Spec: 0.889 [VAL SCORE: 1.556] || Test Sens: 0.300 | Test Spec: 0.759 [TEST SCORE: 1.059]

##### #22: Number of sexual partners + First sexual intercourse + Smokes (packs/year) + Hormonal Contraceptives (years) +

Optimized Thresholds: Number of sexual partners: 4.4208 | First sexual intercourse: 23.3938 | Smokes (packs/year): 0.8124 | Hormonal Contraceptives (years): 9.4967 | STDs (number): 0.3654

Train Sens: 0.654 | Train Spec: 0.714 [TRAIN SCORE: 1.368] || Val Sens: 0.667 | Val Spec: 0.778 [VAL SCORE: 1.444] || Test Sens: 0.300 | Test Spec: 0.717 [TEST SCORE: 1.017]

##### #23: Age + Number of sexual partners + First sexual intercourse* + Smokes (packs/year) + IUD (years) + STDs (number

Optimized Thresholds: Age: 45.9648 | Number of sexual partners: 3.6301 | First sexual intercourse*: 24.3768 | Smokes (packs/year):

1.0380 | IUD (years): 1.4830 | STDs (number): 0.2632

Train Sens: 0.654 | Train Spec: 0.714 [TRAIN SCORE: 1.368] || Val Sens: 0.889 | Val Spec: 0.556 [VAL SCORE: 1.444] || Test Sens: 0.500 | Test Spec: 0.635 [TEST SCORE: 1.135]

###### #24: Smokes (packs/year) + IUD (years) + STDs (number)

Optimized Thresholds: Smokes (packs/year): 1.0338 | IUD (years): 0.8522 | STDs (number): 0.6397

Train Sens: 0.423 | Train Spec: 0.786 [TRAIN SCORE: 1.209] || Val Sens: 0.889 | Val Spec: 0.556 [VAL SCORE: 1.444] || Test Sens: 0.300 | Test Spec: 0.782 [TEST SCORE: 1.082]

###### #25: Number of sexual partners + First sexual intercourse + Num of pregnancies + Hormonal Contraceptives (years) + S

Optimized Thresholds: Number of sexual partners: 4.0631 | First sexual intercourse: 24.8726 | Num of pregnancies: 5.4500 | Hormonal Contraceptives (years): 7.4622 | STDs (number): 0.5095

Train Sens: 0.615 | Train Spec: 0.679 [TRAIN SCORE: 1.294] || Val Sens: 0.556 | Val Spec: 0.889 [VAL SCORE: 1.444] || Test Sens: 0.300 | Test Spec: 0.715 [TEST SCORE: 1.015]

###### #26: Age + First sexual intercourse + Num of pregnancies + IUD (years)

Optimized Thresholds: Age: 34.2649 | First sexual intercourse: 22.2331 | Num of pregnancies: 2.4913 | IUD (years): 1.3249

Train Sens: 0.577 | Train Spec: 0.607 [TRAIN SCORE: 1.184] || Val Sens: 0.778 | Val Spec: 0.667 [VAL SCORE: 1.444] || Test Sens: 0.700 | Test Spec: 0.551 [TEST SCORE: 1.251]

###### #27: Num of pregnancies + Smokes (packs/year) + IUD (years) + STDs (number)

Optimized Thresholds: Num of pregnancies: 2.4314 | Smokes (packs/year): 0.0189 | IUD (years): 0.6149 | STDs (number): 2.3253

Train Sens: 0.538 | Train Spec: 0.607 [TRAIN SCORE: 1.146] || Val Sens: 0.889 | Val Spec: 0.556 [VAL SCORE: 1.444] || Test Sens: 0.600 | Test Spec: 0.527 [TEST SCORE: 1.127]

###### #28: Age + Number of sexual partners + Num of pregnancies + Smokes (packs/year) + IUD (years) + STDs (number)

Optimized Thresholds: Age: 45.9626 | Number of sexual partners: 3.5617 | Num of pregnancies: 6.4947 | Smokes (packs/year): 0.8518 | IUD (years): 1.4755 | STDs (number): 0.5096

Train Sens: 0.615 | Train Spec: 0.714 [TRAIN SCORE: 1.330] || Val Sens: 0.889 | Val Spec: 0.556 [VAL SCORE: 1.444] || Test Sens: 0.500 | Test Spec: 0.648 [TEST SCORE: 1.148]

###### #29: Num of pregnancies + Smokes (packs/year) + IUD (years)

Optimized Thresholds: Num of pregnancies: 2.3511 | Smokes (packs/year): 0.0159 | IUD (years): 1.5386

Train Sens: 0.500 | Train Spec: 0.607 [TRAIN SCORE: 1.107] || Val Sens: 0.889 | Val Spec: 0.556 [VAL SCORE: 1.444] || Test Sens: 0.600 | Test Spec: 0.539 [TEST SCORE: 1.139]

###### #30: Age + Num of pregnancies + Hormonal Contraceptives (years) + IUD (years) + STDs (number)

Optimized Thresholds: Age: 34.2164 | Num of pregnancies: 2.4840 | Hormonal Contraceptives (years): 11.6032 | IUD (years): 1.8373 | STDs (number): 2.4657

Train Sens: 0.577 | Train Spec: 0.607 [TRAIN SCORE: 1.184] || Val Sens: 0.778 | Val Spec: 0.667 [VAL SCORE: 1.444] || Test Sens: 0.700 | Test Spec: 0.560 [TEST SCORE: 1.260]

###### #31: Age + Number of sexual partners + Num of pregnancies + Hormonal Contraceptives (years) + IUD (years)* + STDs

Optimized Thresholds: Age: 47.8587 | Number of sexual partners: 3.9465 | Num of pregnancies: 6.2015 | Hormonal Contraceptives (years): 8.3759 | IUD (years)*: 2.8838 | STDs (number): 0.6034

Train Sens: 0.615 | Train Spec: 0.607 [TRAIN SCORE: 1.223] || Val Sens: 0.889 | Val Spec: 0.556 [VAL SCORE: 1.444] || Test Sens: 0.500 | Test Spec: 0.660 [TEST SCORE: 1.160]

###### #32: Age + Number of sexual partners + First sexual intercourse + Smokes (packs/year) + STDs (number)

Optimized Thresholds: Age: 46.0788 | Number of sexual partners: 3.6055 | First sexual intercourse: 23.9294 | Smokes (packs/year): 0.8106 | STDs (number): 0.5703

Train Sens: 0.615 | Train Spec: 0.714 [TRAIN SCORE: 1.330] || Val Sens: 0.667 | Val Spec: 0.778 [VAL SCORE: 1.444] || Test Sens: 0.400 | Test Spec: 0.676 [TEST SCORE: 1.076]

###### #33: Number of sexual partners + Smokes (packs/year) + Hormonal Contraceptives (years) + STDs (number)

Optimized Thresholds: Number of sexual partners: 3.4562 | Smokes (packs/year): 0.9050 | Hormonal Contraceptives (years): 9.5744 | STDs (number): 0.3890

Train Sens: 0.654 | Train Spec: 0.643 [TRAIN SCORE: 1.297] || Val Sens: 0.667 | Val Spec: 0.778 [VAL SCORE: 1.444] || Test Sens: 0.500 | Test Spec: 0.696 [TEST SCORE: 1.196]

###### #34: Number of sexual partners + First sexual intercourse* + Smokes (packs/year) + IUD (years) + STDs (number)

Optimized Thresholds: Number of sexual partners: 3.5789 | First sexual intercourse*: 24.5195 | Smokes (packs/year): 0.9296 | IUD (years): 1.5042 | STDs (number): 0.4805

Train Sens: 0.615 | Train Spec: 0.714 [TRAIN SCORE: 1.330] || Val Sens: 0.889 | Val Spec: 0.556 [VAL SCORE: 1.444] || Test Sens: 0.500 | Test Spec: 0.645 [TEST SCORE: 1.145]

###### #35: Num of pregnancies + Smokes (packs/year) + Hormonal Contraceptives (years) + IUD (years)

Optimized Thresholds: Num of pregnancies: 2.4689 | Smokes (packs/year): 0.0300 | Hormonal Contraceptives (years): 12.1588 | IUD (years): 1.4862

Train Sens: 0.538 | Train Spec: 0.607 [TRAIN SCORE: 1.146] || Val Sens: 0.889 | Val Spec: 0.556 [VAL SCORE: 1.444] || Test Sens: 0.700 | Test Spec: 0.536 [TEST SCORE: 1.236]

###### #36: Age + Num of pregnancies + Smokes (packs/year) + Hormonal Contraceptives (years) + IUD (years)

Optimized Thresholds: Age: 34.1887 | Num of pregnancies: 2.4639 | Smokes (packs/year): 0.0285 | Hormonal Contraceptives (years): 11.9072 | IUD (years): 1.2289

Train Sens: 0.577 | Train Spec: 0.607 [TRAIN SCORE: 1.184] || Val Sens: 0.889 | Val Spec: 0.556 [VAL SCORE: 1.444] || Test Sens: 0.800 | Test Spec: 0.488 [TEST SCORE: 1.288]

###### #37: Number of sexual partners* + Num of pregnancies* + Smokes (packs/year) + Hormonal Contraceptives (years)* +

Optimized Thresholds: Number of sexual partners*: 3.8161 | Num of pregnancies*: 3.6970 | Smokes (packs/year): 0.0238 | Hormonal Contraceptives (years)*: 14.5404 | IUD (years): 1.3955

Train Sens: 0.538 | Train Spec: 0.643 [TRAIN SCORE: 1.181] || Val Sens: 0.889 | Val Spec: 0.556 [VAL SCORE: 1.444] || Test Sens: 0.600 | Test Spec: 0.573 [TEST SCORE: 1.173]

###### #38: Age + Number of sexual partners + First sexual intercourse + Num of pregnancies + Smokes (packs/year) + STDs

Optimized Thresholds: Age: 46.0077 | Number of sexual partners: 3.5639 | First sexual intercourse: 24.2906 | Num of pregnancies: 6.5459 | Smokes (packs/year): 0.9121 | STDs (number): 0.4481

Train Sens: 0.615 | Train Spec: 0.714 [TRAIN SCORE: 1.330] || Val Sens: 0.667 | Val Spec: 0.778 [VAL SCORE: 1.444] || Test Sens: 0.400 | Test Spec: 0.683 [TEST SCORE: 1.083]

###### #39: First sexual intercourse + Num of pregnancies

Optimized Thresholds: First sexual intercourse: 22.1934 | Num of pregnancies: 2.6966

Train Sens: 0.500 | Train Spec: 0.607 [TRAIN SCORE: 1.107] || Val Sens: 0.778 | Val Spec: 0.667 [VAL SCORE: 1.444] || Test Sens: 0.500 | Test Spec: 0.611 [TEST SCORE: 1.111]

##### #40: Num of pregnancies + Smokes (packs/year)

Optimized Thresholds: Num of pregnancies: 2.5765 | Smokes (packs/year): 0.0226

Train Sens: 0.462 | Train Spec: 0.607 [TRAIN SCORE: 1.069] || Val Sens: 0.889 | Val Spec: 0.556 [VAL SCORE: 1.444] || Test Sens: 0.600 | Test Spec: 0.568 [TEST SCORE: 1.168]

##### #41: Number of sexual partners + Num of pregnancies* + Smokes (packs/year) + Hormonal Contraceptives (years)*

Optimized Thresholds: Number of sexual partners: 4.0312 | Num of pregnancies*: 3.2667 | Smokes (packs/year): 0.0226 | Hormonal Contraceptives (years)*: 14.1485

Train Sens: 0.462 | Train Spec: 0.714 [TRAIN SCORE: 1.176] || Val Sens: 0.667 | Val Spec: 0.778 [VAL SCORE: 1.444] || Test Sens: 0.400 | Test Spec: 0.660 [TEST SCORE: 1.060]

##### #42: First sexual intercourse + Num of pregnancies + Hormonal Contraceptives (years) + IUD (years)

Optimized Thresholds: First sexual intercourse: 22.0208 | Num of pregnancies: 2.3775 | Hormonal Contraceptives (years): 11.0932 | IUD (years): 1.3955

Train Sens: 0.577 | Train Spec: 0.607 [TRAIN SCORE: 1.184] || Val Sens: 0.778 | Val Spec: 0.667 [VAL SCORE: 1.444] || Test Sens: 0.600 | Test Spec: 0.573 [TEST SCORE: 1.173]

##### #43: Number of sexual partners + First sexual intercourse + Num of pregnancies + Smokes (packs/year) + IUD (years) +

Optimized Thresholds: Number of sexual partners: 3.5209 | First sexual intercourse: 24.2447 | Num of pregnancies: 6.3749 | Smokes (packs/year): 1.0641 | IUD (years): 1.2494 | STDs (number): 0.4818

Train Sens: 0.615 | Train Spec: 0.714 [TRAIN SCORE: 1.330] || Val Sens: 0.889 | Val Spec: 0.556 [VAL SCORE: 1.444] || Test Sens: 0.500 | Test Spec: 0.638 [TEST SCORE: 1.138]

##### #44: Number of sexual partners + First sexual intercourse* + Hormonal Contraceptives (years) + STDs (number)

Optimized Thresholds: Number of sexual partners: 4.0481 | First sexual intercourse*: 25.7162 | Hormonal Contraceptives (years): 7.5151 | STDs (number): 0.4675

Train Sens: 0.577 | Train Spec: 0.679 [TRAIN SCORE: 1.255] || Val Sens: 0.556 | Val Spec: 0.889 [VAL SCORE: 1.444] || Test Sens: 0.300 | Test Spec: 0.744 [TEST SCORE: 1.044]

##### #45: Age + Number of sexual partners + First sexual intercourse + Num of pregnancies + Smokes (packs/year) + IUD (ye

Optimized Thresholds: Age: 46.1826 | Number of sexual partners: 3.1997 | First sexual intercourse: 26.4092 | Num of pregnancies: 6.3991 | Smokes (packs/year): 1.0903 | IUD (years): 1.3614 | STDs (number): 0.3890

Train Sens: 0.654 | Train Spec: 0.714 [TRAIN SCORE: 1.368] || Val Sens: 0.889 | Val Spec: 0.556 [VAL SCORE: 1.444] || Test Sens: 0.500 | Test Spec: 0.645 [TEST SCORE: 1.145]

##### #46: Number of sexual partners + First sexual intercourse* + Num of pregnancies + Smokes (packs/year) + Hormonal C

Optimized Thresholds: Number of sexual partners: 4.4973 | First sexual intercourse*: 24.8840 | Num of pregnancies: 5.7613 | Smokes (packs/year): 1.2424 | Hormonal Contraceptives (years): 9.5809 | STDs (number): 0.5960

Train Sens: 0.654 | Train Spec: 0.714 [TRAIN SCORE: 1.368] || Val Sens: 0.667 | Val Spec: 0.778 [VAL SCORE: 1.444] || Test Sens: 0.300 | Test Spec: 0.713 [TEST SCORE: 1.013]

##### #47: Age + Number of sexual partners + Num of pregnancies + Hormonal Contraceptives (years) + STDs (number)

Optimized Thresholds: Age: 47.4818 | Number of sexual partners: 3.6598 | Num of pregnancies: 6.0291 | Hormonal Contraceptives (years): 8.7337 | STDs (number): 0.5388

Train Sens: 0.615 | Train Spec: 0.607 [TRAIN SCORE: 1.223] || Val Sens: 0.667 | Val Spec: 0.778 [VAL SCORE: 1.444] || Test Sens: 0.500 | Test Spec: 0.703 [TEST SCORE: 1.203]

##### #48: IUD (years) + STDs (number)

Optimized Thresholds: IUD (years): 0.8186 | STDs (number): 0.4303

Train Sens: 0.346 | Train Spec: 0.821 [TRAIN SCORE: 1.168] || Val Sens: 0.778 | Val Spec: 0.667 [VAL SCORE: 1.444] || Test Sens: 0.200 | Test Spec: 0.834 [TEST SCORE: 1.034]

##### #49: First sexual intercourse + Num of pregnancies + IUD (years)*

Optimized Thresholds: First sexual intercourse: 21.8038 | Num of pregnancies: 2.6023 | IUD (years)*: 1.6085

Train Sens: 0.538 | Train Spec: 0.607 [TRAIN SCORE: 1.146] || Val Sens: 0.778 | Val Spec: 0.667 [VAL SCORE: 1.444] || Test Sens: 0.500 | Test Spec: 0.570 [TEST SCORE: 1.070]

##### #50: Age + Number of sexual partners + Smokes (packs/year) + IUD (years) + STDs (number)

Optimized Thresholds: Age: 46.1965 | Number of sexual partners: 3.5899 | Smokes (packs/year): 1.0240 | IUD (years): 1.4603 | STDs (number): 0.4807

Train Sens: 0.615 | Train Spec: 0.714 [TRAIN SCORE: 1.330] || Val Sens: 0.889 | Val Spec: 0.556 [VAL SCORE: 1.444] || Test Sens: 0.500 | Test Spec: 0.664 [TEST SCORE: 1.164]

##### #51: Number of sexual partners + Num of pregnancies + Smokes (packs/year) + Hormonal Contraceptives (years) + STD[illegible]

Optimized Thresholds: Number of sexual partners: 3.3927 | Num of pregnancies: 6.1965 | Smokes (packs/year): 1.3381 | Hormonal Contraceptives (years): 9.4202 | STDs (number): 0.4394

Train Sens: 0.654 | Train Spec: 0.643 [TRAIN SCORE: 1.297] || Val Sens: 0.667 | Val Spec: 0.778 [VAL SCORE: 1.444] || Test Sens: 0.400 | Test Spec: 0.693 [TEST SCORE: 1.093]

##### #52: Age + First sexual intercourse + Num of pregnancies

Optimized Thresholds: Age: 34.0296 | First sexual intercourse: 22.2216 | Num of pregnancies: 2.4437

Train Sens: 0.538 | Train Spec: 0.607 [TRAIN SCORE: 1.146] || Val Sens: 0.778 | Val Spec: 0.667 [VAL SCORE: 1.444] || Test Sens: 0.700 | Test Spec: 0.572 [TEST SCORE: 1.272]

##### #53: Number of sexual partners + Smokes (packs/year) + Hormonal Contraceptives (years) + IUD (years)* + STDs (numb[illegible]

Optimized Thresholds: Number of sexual partners: 3.4858 | Smokes (packs/year): 1.5640 | Hormonal Contraceptives (years): 10.8187 | IUD (years)*: 3.1698 | STDs (number): 0.4816

Train Sens: 0.615 | Train Spec: 0.643 [TRAIN SCORE: 1.258] || Val Sens: 0.778 | Val Spec: 0.556 [VAL SCORE: 1.333] || Test Sens: 0.500 | Test Spec: 0.674 [TEST SCORE: 1.174]

##### #54: First sexual intercourse + Num of pregnancies + STDs (number)

Optimized Thresholds: First sexual intercourse: 21.6627 | Num of pregnancies: 2.4611 | STDs (number): 2.5579

Train Sens: 0.538 | Train Spec: 0.607 [TRAIN SCORE: 1.146] || Val Sens: 0.778 | Val Spec: 0.556 [VAL SCORE: 1.333] || Test Sens: 0.500 | Test Spec: 0.597 [TEST SCORE: 1.097]

##### #55: Number of sexual partners + Num of pregnancies + Hormonal Contraceptives (years) + STDs (number)

Optimized Thresholds: Number of sexual partners: 3.8881 | Num of pregnancies: 5.3579 | Hormonal Contraceptives (years): 7.9810 | STDs (number): 0.4829

Train Sens: 0.615 | Train Spec: 0.607 [TRAIN SCORE: 1.223] || Val Sens: 0.556 | Val Spec: 0.778 [VAL SCORE: 1.333] || Test Sens: 0.500 | Test Spec: 0.672 [TEST SCORE: 1.172]

##### #56: Number of sexual partners* + First sexual intercourse + Num of pregnancies* + Hormonal Contraceptives (years)*

Optimized Thresholds: Number of sexual partners*: 3.7934 | First sexual intercourse: 21.8492 | Num of pregnancies*: 3.7297 | Hormonal Contraceptives (years)*: 14.7549 | IUD (years): 1.5652

Train Sens: 0.500 | Train Spec: 0.643 [TRAIN SCORE: 1.143] || Val Sens: 0.778 | Val Spec: 0.556 [VAL SCORE: 1.333] || Test Sens: 0.500 | Test Spec: 0.597 [TEST SCORE: 1.097]

##### #57: Age + First sexual intercourse + Num of pregnancies + STDs (number)

Optimized Thresholds: Age: 34.0229 | First sexual intercourse: 22.4301 | Num of pregnancies: 2.5862 | STDs (number): 2.5044

Train Sens: 0.577 | Train Spec: 0.607 [TRAIN SCORE: 1.184] || Val Sens: 0.778 | Val Spec: 0.556 [VAL SCORE: 1.333] || Test Sens: 0.700 | Test Spec: 0.567 [TEST SCORE: 1.267]

##### #58: Age + First sexual intercourse + Num of pregnancies + IUD (years) + STDs (number)

Optimized Thresholds: Age: 34.1170 | First sexual intercourse: 21.9586 | Num of pregnancies: 2.4312 | IUD (years): 1.5319 | STDs (number): 2.5091

Train Sens: 0.615 | Train Spec: 0.607 [TRAIN SCORE: 1.223] || Val Sens: 0.778 | Val Spec: 0.556 [VAL SCORE: 1.333] || Test Sens: 0.700 | Test Spec: 0.538 [TEST SCORE: 1.238]

##### #59: Number of sexual partners + Hormonal Contraceptives (years) + STDs (number)

Optimized Thresholds: Number of sexual partners: 3.7016 | Hormonal Contraceptives (years): 8.3813 | STDs (number): 0.3644

Train Sens: 0.577 | Train Spec: 0.607 [TRAIN SCORE: 1.184] || Val Sens: 0.556 | Val Spec: 0.778 [VAL SCORE: 1.333] || Test Sens: 0.500 | Test Spec: 0.718 [TEST SCORE: 1.218]

##### #60: First sexual intercourse + Num of pregnancies + IUD (years) + STDs (number)

Optimized Thresholds: First sexual intercourse: 21.8911 | Num of pregnancies: 2.5597 | IUD (years): 1.7688 | STDs (number): 2.6728

Train Sens: 0.577 | Train Spec: 0.607 [TRAIN SCORE: 1.184] || Val Sens: 0.778 | Val Spec: 0.556 [VAL SCORE: 1.333] || Test Sens: 0.500 | Test Spec: 0.565 [TEST SCORE: 1.065]

##### #61: Number of sexual partners* + First sexual intercourse + Num of pregnancies* + Hormonal Contraceptives (years)*

Optimized Thresholds: Number of sexual partners*: 3.8484 | First sexual intercourse: 22.0196 | Num of pregnancies*: 3.6647 | Hormonal Contraceptives (years)*: 13.6062

Train Sens: 0.500 | Train Spec: 0.643 [TRAIN SCORE: 1.143] || Val Sens: 0.556 | Val Spec: 0.778 [VAL SCORE: 1.333] || Test Sens: 0.400 | Test Spec: 0.642 [TEST SCORE: 1.042]

##### #62: Age + Smokes (packs/year) + IUD (years)* + STDs (number)

Optimized Thresholds: Age: 34.0585 | Smokes (packs/year): 1.0202 | IUD (years)*: 4.7604 | STDs (number): 0.5201

Train Sens: 0.500 | Train Spec: 0.607 [TRAIN SCORE: 1.107] || Val Sens: 0.778 | Val Spec: 0.556 [VAL SCORE: 1.333] || Test Sens: 0.500 | Test Spec: 0.681 [TEST SCORE: 1.181]

##### #63: Age + Number of sexual partners + First sexual intercourse* + Num of pregnancies + Smokes (packs/year) + Horm

Optimized Thresholds: Age: 48.6073 | Number of sexual partners: 4.4422 | First sexual intercourse*: 24.1534 | Num of pregnancies: 5.6930 | Smokes (packs/year): 1.1711 | Hormonal Contraceptives (years): 9.6594 | IUD (years)*: 5.4116 | STDs (number): 0.4120

Train Sens: 0.654 | Train Spec: 0.714 [TRAIN SCORE: 1.368] || Val Sens: 0.778 | Val Spec: 0.556 [VAL SCORE: 1.333] || Test

Sens: 0.400 | Test Spec: 0.679 [TEST SCORE: 1.079]

##### #64: Age + First sexual intercourse + STDs (number)

Optimized Thresholds: Age: 46.1855 | First sexual intercourse: 17.5226 | STDs (number): 0.4732

Train Sens: 0.615 | Train Spec: 0.571 [TRAIN SCORE: 1.187] || Val Sens: 0.778 | Val Spec: 0.556 [VAL SCORE: 1.333] || Test Sens: 0.400 | Test Spec: 0.560 [TEST SCORE: 0.960]

##### #65: Num of pregnancies + Smokes (packs/year) + Hormonal Contraceptives (years) + IUD (years) + STDs (number)

Optimized Thresholds: Num of pregnancies: 3.5627 | Smokes (packs/year): 1.3718 | Hormonal Contraceptives (years): 11.2667 | IUD (years): 4.3349 | STDs (number): 0.5142

Train Sens: 0.577 | Train Spec: 0.643 [TRAIN SCORE: 1.220] || Val Sens: 0.778 | Val Spec: 0.556 [VAL SCORE: 1.333] || Test Sens: 0.500 | Test Spec: 0.689 [TEST SCORE: 1.189]

##### #66: First sexual intercourse + STDs (number)

Optimized Thresholds: First sexual intercourse: 17.5944 | STDs (number): 0.4773

Train Sens: 0.500 | Train Spec: 0.571 [TRAIN SCORE: 1.071] || Val Sens: 0.778 | Val Spec: 0.556 [VAL SCORE: 1.333] || Test Sens: 0.400 | Test Spec: 0.570 [TEST SCORE: 0.970]

##### #67: Number of sexual partners + First sexual intercourse + Smokes (packs/year) + Hormonal Contraceptives (years) +

Optimized Thresholds: Number of sexual partners: 4.5798 | First sexual intercourse: 24.5309 | Smokes (packs/year): 1.0391 | Hormonal Contraceptives (years): 9.5775 | IUD (years): 4.2178 | STDs (number): 0.6071

Train Sens: 0.654 | Train Spec: 0.714 [TRAIN SCORE: 1.368] || Val Sens: 0.778 | Val Spec: 0.556 [VAL SCORE: 1.333] || Test Sens: 0.400 | Test Spec: 0.698 [TEST SCORE: 1.098]

##### #68: Number of sexual partners + Num of pregnancies + Smokes (packs/year) + Hormonal Contraceptives (years) + IUD

Optimized Thresholds: Number of sexual partners: 3.5008 | Num of pregnancies: 5.9655 | Smokes (packs/year): 0.6703 | Hormonal Contraceptives (years): 10.8736 | IUD (years)*: 3.1183 | STDs (number): 0.6104

Train Sens: 0.615 | Train Spec: 0.643 [TRAIN SCORE: 1.258] || Val Sens: 0.778 | Val Spec: 0.556 [VAL SCORE: 1.333] || Test Sens: 0.600 | Test Spec: 0.638 [TEST SCORE: 1.238]

##### #69: Age + Num of pregnancies + IUD (years)

Optimized Thresholds: Age: 32.6013 | Num of pregnancies: 2.4564 | IUD (years): 1.5559

Train Sens: 0.538 | Train Spec: 0.571 [TRAIN SCORE: 1.110] || Val Sens: 0.778 | Val Spec: 0.556 [VAL SCORE: 1.333] || Test Sens: 0.700 | Test Spec: 0.546 [TEST SCORE: 1.246]

##### #70: Age + Num of pregnancies + STDs (number)*

Optimized Thresholds: Age: 33.5429 | Num of pregnancies: 3.1520 | STDs (number)*: 0.9899

Train Sens: 0.538 | Train Spec: 0.571 [TRAIN SCORE: 1.110] || Val Sens: 0.778 | Val Spec: 0.556 [VAL SCORE: 1.333] || Test Sens: 0.600 | Test Spec: 0.657 [TEST SCORE: 1.257]

##### #71: Age + Number of sexual partners + Smokes (packs/year) + Hormonal Contraceptives (years) + IUD (years)* + STDs

Optimized Thresholds: Age: 47.8503 | Number of sexual partners: 3.4541 | Smokes (packs/year): 0.8657 | Hormonal Contraceptives (years): 10.1216 | IUD (years)*: 3.9099 | STDs (number): 0.4882

Train Sens: 0.615 | Train Spec: 0.643 [TRAIN SCORE: 1.258] || Val Sens: 0.778 | Val Spec: 0.556 [VAL SCORE: 1.333] || Test Sens: 0.600 | Test Spec: 0.657 [TEST SCORE: 1.257]

##### #72: First sexual intercourse + Num of pregnancies + Smokes (packs/year) + Hormonal Contraceptives (years) + IUD (ye

Optimized Thresholds: First sexual intercourse: 24.9109 | Num of pregnancies: 3.5005 | Smokes (packs/year): 0.9540 | Hormonal Contraceptives (years): 11.7179 | IUD (years)*: 4.7827 | STDs (number): 0.5614

Train Sens: 0.615 | Train Spec: 0.643 [TRAIN SCORE: 1.258] || Val Sens: 0.778 | Val Spec: 0.556 [VAL SCORE: 1.333] || Test Sens: 0.600 | Test Spec: 0.650 [TEST SCORE: 1.250]

##### #73: Age + Number of sexual partners + First sexual intercourse + Smokes (packs/year) + Hormonal Contraceptives (ye

Optimized Thresholds: Age: 48.0361 | Number of sexual partners: 4.5757 | First sexual intercourse: 24.5992 | Smokes (packs/year): 0.9531 | Hormonal Contraceptives (years): 9.4876 | IUD (years): 5.2853 | STDs (number): 0.5987

Train Sens: 0.654 | Train Spec: 0.714 [TRAIN SCORE: 1.368] || Val Sens: 0.778 | Val Spec: 0.556 [VAL SCORE: 1.333] || Test Sens: 0.400 | Test Spec: 0.691 [TEST SCORE: 1.091]

##### #74: Age + STDs (number)

Optimized Thresholds: Age: 32.3874 | STDs (number): 0.5178

Train Sens: 0.500 | Train Spec: 0.500 [TRAIN SCORE: 1.000] || Val Sens: 0.778 | Val Spec: 0.556 [VAL SCORE: 1.333] || Test Sens: 0.400 | Test Spec: 0.696 [TEST SCORE: 1.096]

##### #75: Age + First sexual intercourse + Num of pregnancies + Smokes (packs/year) + Hormonal Contraceptives (years) +

Optimized Thresholds: Age: 48.2805 | First sexual intercourse: 24.4297 | Num of pregnancies: 3.4352 | Smokes (packs/year): 1.4045 | Hormonal Contraceptives (years): 11.6805 | IUD (years)*: 5.1418 | STDs (number): 0.4103

Train Sens: 0.615 | Train Spec: 0.643 [TRAIN SCORE: 1.258] || Val Sens: 0.778 | Val Spec: 0.556 [VAL SCORE: 1.333] || Test Sens: 0.500 | Test Spec: 0.662 [TEST SCORE: 1.162]

##### #76: First sexual intercourse + Num of pregnancies + Hormonal Contraceptives (years) + IUD (years) + STDs (number)

Optimized Thresholds: First sexual intercourse: 22.3356 | Num of pregnancies: 2.5279 | Hormonal Contraceptives (years): 11.7042 | IUD (years): 1.8280 | STDs (number): 2.4630

Train Sens: 0.615 | Train Spec: 0.607 [TRAIN SCORE: 1.223] || Val Sens: 0.778 | Val Spec: 0.556 [VAL SCORE: 1.333] || Test Sens: 0.600 | Test Spec: 0.568 [TEST SCORE: 1.168]

##### #77: Number of sexual partners + First sexual intercourse* + Num of pregnancies + Smokes (packs/year) + Hormonal C

Optimized Thresholds: Number of sexual partners: 4.5253 | First sexual intercourse*: 23.7780 | Num of pregnancies: 6.0224 | Smokes (packs/year): 1.1860 | Hormonal Contraceptives (years): 9.5494 | IUD (years): 5.8190 | STDs (number): 0.4524

Train Sens: 0.654 | Train Spec: 0.714 [TRAIN SCORE: 1.368] || Val Sens: 0.778 | Val Spec: 0.556 [VAL SCORE: 1.333] || Test Sens: 0.400 | Test Spec: 0.684 [TEST SCORE: 1.084]

##### #78: Age + Num of pregnancies

Optimized Thresholds: Age: 32.5854 | Num of pregnancies: 2.4647

Train Sens: 0.500 | Train Spec: 0.571 [TRAIN SCORE: 1.071] || Val Sens: 0.778 | Val Spec: 0.556 [VAL SCORE: 1.333] || Test Sens: 0.700 | Test Spec: 0.565 [TEST SCORE: 1.265]

##### #79: Age + First sexual intercourse* + Hormonal Contraceptives (years) + STDs (number)

Optimized Thresholds: Age: 48.7256 | First sexual intercourse*: 24.9673 | Hormonal Contraceptives (years): 1.8554 | STDs (number): 0.5674

Train Sens: 0.654 | Train Spec: 0.607 [TRAIN SCORE: 1.261] || Val Sens: 0.667 | Val Spec: 0.667 [VAL SCORE: 1.333] || Test Sens: 0.400 | Test Spec: 0.575 [TEST SCORE: 0.975]

###### #80: Age + First sexual intercourse + Num of pregnancies + Smokes (packs/year)

Optimized Thresholds: Age: 34.1559 | First sexual intercourse: 22.6057 | Num of pregnancies: 2.5115 | Smokes (packs/year): 0.0266

Train Sens: 0.577 | Train Spec: 0.607 [TRAIN SCORE: 1.184] || Val Sens: 0.889 | Val Spec: 0.444 [VAL SCORE: 1.333] || Test Sens: 0.800 | Test Spec: 0.493 [TEST SCORE: 1.293]

###### #81: Age + First sexual intercourse + Num of pregnancies + Smokes (packs/year) + IUD (years) + STDs (number)

Optimized Thresholds: Age: 34.1516 | First sexual intercourse: 21.8453 | Num of pregnancies: 2.5047 | Smokes (packs/year): 0.0236 | IUD (years): 2.2775 | STDs (number): 2.4586

Train Sens: 0.654 | Train Spec: 0.607 [TRAIN SCORE: 1.261] || Val Sens: 0.889 | Val Spec: 0.444 [VAL SCORE: 1.333] || Test Sens: 0.800 | Test Spec: 0.462 [TEST SCORE: 1.262]

###### #82: First sexual intercourse + Num of pregnancies + Smokes (packs/year) + Hormonal Contraceptives (years) + IUD (ye

Optimized Thresholds: First sexual intercourse: 21.9245 | Num of pregnancies: 2.4581 | Smokes (packs/year): 0.0300 | Hormonal Contraceptives (years): 11.1340 | IUD (years): 1.3047

Train Sens: 0.615 | Train Spec: 0.607 [TRAIN SCORE: 1.223] || Val Sens: 0.889 | Val Spec: 0.444 [VAL SCORE: 1.333] || Test Sens: 0.700 | Test Spec: 0.488 [TEST SCORE: 1.188]

###### #83: First sexual intercourse + Num of pregnancies + Smokes (packs/year)

Optimized Thresholds: First sexual intercourse: 21.9770 | Num of pregnancies: 2.3560 | Smokes (packs/year): 0.0251

Train Sens: 0.538 | Train Spec: 0.607 [TRAIN SCORE: 1.146] || Val Sens: 0.889 | Val Spec: 0.444 [VAL SCORE: 1.333] || Test Sens: 0.600 | Test Spec: 0.522 [TEST SCORE: 1.122]

###### #84: Age + First sexual intercourse

Optimized Thresholds: Age: 46.0938 | First sexual intercourse: 17.7745

Train Sens: 0.500 | Train Spec: 0.643 [TRAIN SCORE: 1.143] || Val Sens: 0.667 | Val Spec: 0.667 [VAL SCORE: 1.333] || Test Sens: 0.400 | Test Spec: 0.616 [TEST SCORE: 1.016]

###### #85: First sexual intercourse + Num of pregnancies + Smokes (packs/year) + IUD (years)

Optimized Thresholds: First sexual intercourse: 21.8204 | Num of pregnancies: 2.3703 | Smokes (packs/year): 0.0194 | IUD (years): 1.2568

Train Sens: 0.577 | Train Spec: 0.607 [TRAIN SCORE: 1.184] || Val Sens: 0.889 | Val Spec: 0.444 [VAL SCORE: 1.333] || Test Sens: 0.600 | Test Spec: 0.493 [TEST SCORE: 1.093]

###### #86: First sexual intercourse + Num of pregnancies + Smokes (packs/year) + STDs (number)

Optimized Thresholds: First sexual intercourse: 22.3200 | Num of pregnancies: 2.4532 | Smokes (packs/year): 0.0276 | STDs (number): 2.5235

Train Sens: 0.577 | Train Spec: 0.607 [TRAIN SCORE: 1.184] || Val Sens: 0.889 | Val Spec: 0.444 [VAL SCORE: 1.333] || Test Sens: 0.600 | Test Spec: 0.526 [TEST SCORE: 1.126]

###### #87: First sexual intercourse + Num of pregnancies + Smokes (packs/year) + Hormonal Contraceptives (years)

Optimized Thresholds: First sexual intercourse: 21.9745 | Num of pregnancies: 2.5652 | Smokes (packs/year): 0.0185 | Hormonal Contraceptives (years): 11.4546

Train Sens: 0.577 | Train Spec: 0.607 [TRAIN SCORE: 1.184] || Val Sens: 0.889 | Val Spec: 0.444 [VAL SCORE: 1.333] || Test Sens: 0.700 | Test Spec: 0.514 [TEST SCORE: 1.214]

##### #88: Age + First sexual intercourse + Num of pregnancies + Smokes (packs/year) + STDs (number)

Optimized Thresholds: Age: 34.3602 | First sexual intercourse: 22.2424 | Num of pregnancies: 2.4356 | Smokes (packs/year): 0.0246 | STDs (number): 2.4888

Train Sens: 0.615 | Train Spec: 0.607 [TRAIN SCORE: 1.223] || Val Sens: 0.889 | Val Spec: 0.444 [VAL SCORE: 1.333] || Test Sens: 0.800 | Test Spec: 0.488 [TEST SCORE: 1.288]

##### #89: Age + Hormonal Contraceptives (years) + STDs (number)

Optimized Thresholds: Age: 48.5521 | Hormonal Contraceptives (years): 1.8065 | STDs (number): 0.4273

Train Sens: 0.615 | Train Spec: 0.607 [TRAIN SCORE: 1.223] || Val Sens: 0.667 | Val Spec: 0.667 [VAL SCORE: 1.333] || Test Sens: 0.400 | Test Spec: 0.596 [TEST SCORE: 0.996]

##### #90: First sexual intercourse + Num of pregnancies + Smokes (packs/year) + IUD (years) + STDs (number)

Optimized Thresholds: First sexual intercourse: 22.2226 | Num of pregnancies: 2.4788 | Smokes (packs/year): 0.0244 | IUD (years): 1.2748 | STDs (number): 2.3255

Train Sens: 0.615 | Train Spec: 0.607 [TRAIN SCORE: 1.223] || Val Sens: 0.889 | Val Spec: 0.444 [VAL SCORE: 1.333] || Test Sens: 0.600 | Test Spec: 0.497 [TEST SCORE: 1.097]

##### #91: Age + IUD (years)* + STDs (number)

Optimized Thresholds: Age: 32.4684 | IUD (years)*: 2.6853 | STDs (number): 0.5062

Train Sens: 0.500 | Train Spec: 0.500 [TRAIN SCORE: 1.000] || Val Sens: 0.889 | Val Spec: 0.444 [VAL SCORE: 1.333] || Test Sens: 0.400 | Test Spec: 0.664 [TEST SCORE: 1.064]

##### #92: Number of sexual partners + Num of pregnancies + Hormonal Contraceptives (years) + IUD (years)* + STDs (numb

Optimized Thresholds: Number of sexual partners: 4.1255 | Num of pregnancies: 5.8286 | Hormonal Contraceptives (years): 8.5882 | IUD (years)*: 4.0758 | STDs (number): 0.5199

Train Sens: 0.577 | Train Spec: 0.714 [TRAIN SCORE: 1.291] || Val Sens: 0.667 | Val Spec: 0.667 [VAL SCORE: 1.333] || Test Sens: 0.300 | Test Spec: 0.727 [TEST SCORE: 1.027]

##### #93: Number of sexual partners + First sexual intercourse + Num of pregnancies + Smokes (packs/year) + Hormonal Co

Optimized Thresholds: Number of sexual partners: 3.4356 | First sexual intercourse: 22.2479 | Num of pregnancies: 6.0995 | Smokes (packs/year): 0.0301 | Hormonal Contraceptives (years): 10.4548 | IUD (years): 1.6013

Train Sens: 0.654 | Train Spec: 0.714 [TRAIN SCORE: 1.368] || Val Sens: 0.778 | Val Spec: 0.444 [VAL SCORE: 1.222] || Test Sens: 0.500 | Test Spec: 0.619 [TEST SCORE: 1.119]

##### #94: Age + Number of sexual partners + First sexual intercourse* + Num of pregnancies + Hormonal Contraceptives (ye

Optimized Thresholds: Age: 48.8627 | Number of sexual partners: 3.9396 | First sexual intercourse*: 24.4265 | Num of pregnancies: 5.6125 | Hormonal Contraceptives (years): 8.4420 | IUD (years)*: 3.8488 | STDs (number): 0.5388

Train Sens: 0.654 | Train Spec: 0.607 [TRAIN SCORE: 1.261] || Val Sens: 0.667 | Val Spec: 0.556 [VAL SCORE: 1.222] || Test Sens: 0.500 | Test Spec: 0.626 [TEST SCORE: 1.126]

##### #95: Age + Number of sexual partners + Smokes (packs/year)

Optimized Thresholds: Age: 46.1751 | Number of sexual partners: 2.4833 | Smokes (packs/year): 0.0305

Train Sens: 0.615 | Train Spec: 0.607 [TRAIN SCORE: 1.223] || Val Sens: 0.667 | Val Spec: 0.556 [VAL SCORE: 1.222] || Test Sens: 0.400 | Test Spec: 0.515 [TEST SCORE: 0.915]

##### #96: Age + Number of sexual partners + Num of pregnancies + Smokes (packs/year)

Optimized Thresholds: Age: 46.3640 | Number of sexual partners: 2.4279 | Num of pregnancies: 6.5186 | Smokes (packs/year): 0.0391

Train Sens: 0.615 | Train Spec: 0.607 [TRAIN SCORE: 1.223] || Val Sens: 0.667 | Val Spec: 0.556 [VAL SCORE: 1.222] || Test Sens: 0.400 | Test Spec: 0.514 [TEST SCORE: 0.914]

##### #97: Age + Number of sexual partners + Smokes (packs/year) + Hormonal Contraceptives (years) + IUD (years)

Optimized Thresholds: Age: 46.2992 | Number of sexual partners: 2.5087 | Smokes (packs/year): 0.0208 | Hormonal Contraceptives (years): 21.3113 | IUD (years): 1.8518

Train Sens: 0.654 | Train Spec: 0.607 [TRAIN SCORE: 1.261] || Val Sens: 0.778 | Val Spec: 0.444 [VAL SCORE: 1.222] || Test Sens: 0.500 | Test Spec: 0.485 [TEST SCORE: 0.985]

##### #98: Hormonal Contraceptives (years) + STDs (number)

Optimized Thresholds: Hormonal Contraceptives (years): 2.1210 | STDs (number): 0.3767

Train Sens: 0.577 | Train Spec: 0.607 [TRAIN SCORE: 1.184] || Val Sens: 0.556 | Val Spec: 0.667 [VAL SCORE: 1.222] || Test Sens: 0.400 | Test Spec: 0.660 [TEST SCORE: 1.060]

##### #99: Age + Number of sexual partners + First sexual intercourse + Num of pregnancies + Smokes (packs/year) + Hormo[illegible]

Optimized Thresholds: Age: 45.4026 | Number of sexual partners: 3.1117 | First sexual intercourse: 21.9322 | Num of pregnancies: 6.4546 | Smokes (packs/year): 0.0347 | Hormonal Contraceptives (years): 10.6044 | IUD (years): 1.6613

Train Sens: 0.654 | Train Spec: 0.714 [TRAIN SCORE: 1.368] || Val Sens: 0.778 | Val Spec: 0.444 [VAL SCORE: 1.222] || Test Sens: 0.500 | Test Spec: 0.602 [TEST SCORE: 1.102]

##### #100: Smokes (packs/year) + STDs (number)

Optimized Thresholds: Smokes (packs/year): 1.1512 | STDs (number): 0.5136

Train Sens: 0.346 | Train Spec: 0.857 [TRAIN SCORE: 1.203] || Val Sens: 0.444 | Val Spec: 0.778 [VAL SCORE: 1.222] || Test Sens: 0.200 | Test Spec: 0.855 [TEST SCORE: 1.055]

##### #101: Number of sexual partners* + First sexual intercourse + Hormonal Contraceptives (years)* + IUD (years)

Optimized Thresholds: Number of sexual partners*: 4.0694 | First sexual intercourse: 22.5852 | Hormonal Contraceptives (years)*: 10.9601 | IUD (years): 2.3536

Train Sens: 0.423 | Train Spec: 0.786 [TRAIN SCORE: 1.209] || Val Sens: 0.556 | Val Spec: 0.667 [VAL SCORE: 1.222] || Test Sens: 0.200 | Test Spec: 0.783 [TEST SCORE: 0.983]

##### #102: Smokes (packs/year) + IUD (years)

Optimized Thresholds: Smokes (packs/year): 0.0250 | IUD (years): 0.7937

Train Sens: 0.308 | Train Spec: 0.857 [TRAIN SCORE: 1.165] || Val Sens: 0.667 | Val Spec: 0.556 [VAL SCORE: 1.222] || Test Sens: 0.200 | Test Spec: 0.778 [TEST SCORE: 0.978]

##### #103: Number of sexual partners + First sexual intercourse + Num of pregnancies + Hormonal Contraceptives (years) + [illegible]

Optimized Thresholds: Number of sexual partners: 3.7200 | First sexual intercourse: 25.9010 | Num of pregnancies: 5.8132 | Hormonal Contraceptives (years): 8.6633 | IUD (years)*: 3.3957 | STDs (number): 0.6562

Train Sens: 0.654 | Train Spec: 0.607 [TRAIN SCORE: 1.261] || Val Sens: 0.667 | Val Spec: 0.556 [VAL SCORE: 1.222] || Test Sens: 0.500 | Test Spec: 0.633 [TEST SCORE: 1.133]

##### #104: Age + Number of sexual partners + Num of pregnancies + Smokes (packs/year) + IUD (years)

Optimized Thresholds: Age: 45.9042 | Number of sexual partners: 2.5373 | Num of pregnancies: 6.6016 | Smokes (packs/year): 0.0277 | IUD (years): 1.7150

Train Sens: 0.654 | Train Spec: 0.607 [TRAIN SCORE: 1.261] || Val Sens: 0.778 | Val Spec: 0.444 [VAL SCORE: 1.222] || Test Sens: 0.500 | Test Spec: 0.480 [TEST SCORE: 0.980]

##### #105: Age + Number of sexual partners + Hormonal Contraceptives (years) + IUD (years)* + STDs (number)

Optimized Thresholds: Age: 47.7950 | Number of sexual partners: 3.9187 | Hormonal Contraceptives (years): 8.5741 | IUD (years)*: 3.3044 | STDs (number): 0.5191

Train Sens: 0.615 | Train Spec: 0.607 [TRAIN SCORE: 1.223] || Val Sens: 0.667 | Val Spec: 0.556 [VAL SCORE: 1.222] || Test Sens: 0.500 | Test Spec: 0.671 [TEST SCORE: 1.171]

##### #106: Number of sexual partners + Num of pregnancies + Smokes (packs/year) + IUD (years)

Optimized Thresholds: Number of sexual partners: 2.6364 | Num of pregnancies: 6.5574 | Smokes (packs/year): 0.0176 | IUD (years): 2.2366

Train Sens: 0.615 | Train Spec: 0.607 [TRAIN SCORE: 1.223] || Val Sens: 0.778 | Val Spec: 0.444 [VAL SCORE: 1.222] || Test Sens: 0.500 | Test Spec: 0.490 [TEST SCORE: 0.990]

##### #107: Age + Number of sexual partners + First sexual intercourse + Hormonal Contraceptives (years) + IUD (years)* + S

Optimized Thresholds: Age: 48.3710 | Number of sexual partners: 3.9619 | First sexual intercourse: 25.1012 | Hormonal Contraceptives (years): 8.0110 | IUD (years)*: 3.7008 | STDs (number): 0.4976

Train Sens: 0.654 | Train Spec: 0.607 [TRAIN SCORE: 1.261] || Val Sens: 0.667 | Val Spec: 0.556 [VAL SCORE: 1.222] || Test Sens: 0.500 | Test Spec: 0.652 [TEST SCORE: 1.152]

##### #108: Age + Number of sexual partners + First sexual intercourse + Smokes (packs/year) + Hormonal Contraceptives (y

Optimized Thresholds: Age: 48.2262 | Number of sexual partners: 3.5161 | First sexual intercourse: 22.0129 | Smokes (packs/year): 0.0290 | Hormonal Contraceptives (years): 11.4050 | IUD (years): 1.7213

Train Sens: 0.654 | Train Spec: 0.714 [TRAIN SCORE: 1.368] || Val Sens: 0.778 | Val Spec: 0.444 [VAL SCORE: 1.222] || Test Sens: 0.500 | Test Spec: 0.623 [TEST SCORE: 1.123]

##### #109: First sexual intercourse* + Hormonal Contraceptives (years) + STDs (number)

Optimized Thresholds: First sexual intercourse*: 24.2908 | Hormonal Contraceptives (years): 1.6464 | STDs (number): 0.5645

Train Sens: 0.615 | Train Spec: 0.607 [TRAIN SCORE: 1.223] || Val Sens: 0.556 | Val Spec: 0.667 [VAL SCORE: 1.222] || Test Sens: 0.400 | Test Spec: 0.582 [TEST SCORE: 0.982]

##### #110: Age + Number of sexual partners + Num of pregnancies + Smokes (packs/year) + Hormonal Contraceptives (year

Optimized Thresholds: Age: 45.9171 | Number of sexual partners: 2.3881 | Num of pregnancies: 6.5373 | Smokes (packs/year): 0.0242 | Hormonal Contraceptives (years): 20.8725

Train Sens: 0.615 | Train Spec: 0.607 [TRAIN SCORE: 1.223] || Val Sens: 0.667 | Val Spec: 0.556 [VAL SCORE: 1.222] || Test Sens: 0.400 | Test Spec: 0.510 [TEST SCORE: 0.910]

##### #111: Number of sexual partners + Smokes (packs/year) + Hormonal Contraceptives (years) + IUD (years)

Optimized Thresholds: Number of sexual partners: 2.4582 | Smokes (packs/year): 0.0260 | Hormonal Contraceptives (years): 21.0016 | IUD (years): 2.2384

Train Sens: 0.615 | Train Spec: 0.607 [TRAIN SCORE: 1.223] || Val Sens: 0.778 | Val Spec: 0.444 [VAL SCORE: 1.222] || Test

Sens: 0.500 | Test Spec: 0.493 [TEST SCORE: 0.993]

###### #112: First sexual intercourse + Smokes (packs/year)

Optimized Thresholds: First sexual intercourse: 17.4336 | Smokes (packs/year): 0.0247

Train Sens: 0.538 | Train Spec: 0.571 [TRAIN SCORE: 1.110] || Val Sens: 0.778 | Val Spec: 0.444 [VAL SCORE: 1.222] || Test Sens: 0.500 | Test Spec: 0.522 [TEST SCORE: 1.022]

###### #113: Number of sexual partners + Smokes (packs/year) + IUD (years) + STDs (number)

Optimized Thresholds: Number of sexual partners: 2.3774 | Smokes (packs/year): 0.0195 | IUD (years): 1.8214 | STDs (number): 2.4409

Train Sens: 0.615 | Train Spec: 0.607 [TRAIN SCORE: 1.223] || Val Sens: 0.778 | Val Spec: 0.444 [VAL SCORE: 1.222] || Test Sens: 0.500 | Test Spec: 0.485 [TEST SCORE: 0.985]

###### #114: Number of sexual partners + Hormonal Contraceptives (years) + IUD (years)* + STDs (number)

Optimized Thresholds: Number of sexual partners: 3.7309 | Hormonal Contraceptives (years): 8.4782 | IUD (years)*: 3.8113 | STDs (number): 0.5141

Train Sens: 0.615 | Train Spec: 0.607 [TRAIN SCORE: 1.223] || Val Sens: 0.667 | Val Spec: 0.556 [VAL SCORE: 1.222] || Test Sens: 0.500 | Test Spec: 0.681 [TEST SCORE: 1.181]

###### #115: Age + Smokes (packs/year)* + Hormonal Contraceptives (years) + STDs (number)

Optimized Thresholds: Age: 47.9470 | Smokes (packs/year)*: 3.7297 | Hormonal Contraceptives (years): 2.1326 | STDs (number): 0.4394

Train Sens: 0.615 | Train Spec: 0.571 [TRAIN SCORE: 1.187] || Val Sens: 0.667 | Val Spec: 0.556 [VAL SCORE: 1.222] || Test Sens: 0.400 | Test Spec: 0.638 [TEST SCORE: 1.038]

###### #116: Age + Num of pregnancies + IUD (years) + STDs (number)

Optimized Thresholds: Age: 32.6445 | Num of pregnancies: 2.5291 | IUD (years): 1.4698 | STDs (number): 2.4934

Train Sens: 0.577 | Train Spec: 0.571 [TRAIN SCORE: 1.148] || Val Sens: 0.778 | Val Spec: 0.444 [VAL SCORE: 1.222] || Test Sens: 0.700 | Test Spec: 0.541 [TEST SCORE: 1.241]

###### #117: Number of sexual partners + First sexual intercourse + Hormonal Contraceptives (years) + IUD (years)* + STDs (n

Optimized Thresholds: Number of sexual partners: 3.8060 | First sexual intercourse: 25.9532 | Hormonal Contraceptives (years): 8.7121 | IUD (years)*: 4.3594 | STDs (number): 0.4494

Train Sens: 0.654 | Train Spec: 0.607 [TRAIN SCORE: 1.261] || Val Sens: 0.667 | Val Spec: 0.556 [VAL SCORE: 1.222] || Test Sens: 0.500 | Test Spec: 0.664 [TEST SCORE: 1.164]

###### #118: First sexual intercourse + Smokes (packs/year) + STDs (number)

Optimized Thresholds: First sexual intercourse: 17.4042 | Smokes (packs/year): 0.0303 | STDs (number): 2.4841

Train Sens: 0.577 | Train Spec: 0.571 [TRAIN SCORE: 1.148] || Val Sens: 0.778 | Val Spec: 0.444 [VAL SCORE: 1.222] || Test Sens: 0.500 | Test Spec: 0.519 [TEST SCORE: 1.019]

###### #119: Age + Number of sexual partners + Smokes (packs/year) + Hormonal Contraceptives (years)

Optimized Thresholds: Age: 46.0226 | Number of sexual partners: 2.2921 | Smokes (packs/year): 0.0254 | Hormonal Contraceptives (years): 21.3307

Train Sens: 0.615 | Train Spec: 0.607 [TRAIN SCORE: 1.223] || Val Sens: 0.667 | Val Spec: 0.556 [VAL SCORE: 1.222] || Test

Sens: 0.400 | Test Spec: 0.515  [TEST SCORE: 0.915]

###### #120: Age + Number of sexual partners + Smokes (packs/year) + IUD (years)

Optimized Thresholds: Age: 45.8583 | Number of sexual partners: 2.4420 | Smokes (packs/year): 0.0233 | IUD (years): 2.2190

Train Sens: 0.654 | Train Spec: 0.607  [TRAIN SCORE: 1.261]  ||  Val Sens: 0.778 | Val Spec: 0.444  [VAL SCORE: 1.222]  ||  Test Sens: 0.500 | Test Spec: 0.485  [TEST SCORE: 0.985]

###### #121: Age + Number of sexual partners + Smokes (packs/year) + STDs (number)

Optimized Thresholds: Age: 45.8885 | Number of sexual partners: 2.4082 | Smokes (packs/year): 0.0233 | STDs (number): 2.6335

Train Sens: 0.615 | Train Spec: 0.607  [TRAIN SCORE: 1.223]  ||  Val Sens: 0.667 | Val Spec: 0.556  [VAL SCORE: 1.222]  ||  Test Sens: 0.400 | Test Spec: 0.507  [TEST SCORE: 0.907]

###### #122: Number of sexual partners + Smokes (packs/year) + IUD (years)

Optimized Thresholds: Number of sexual partners: 2.4332 | Smokes (packs/year): 0.0271 | IUD (years): 2.2611

Train Sens: 0.615 | Train Spec: 0.607  [TRAIN SCORE: 1.223]  ||  Val Sens: 0.778 | Val Spec: 0.444  [VAL SCORE: 1.222]  ||  Test Sens: 0.500 | Test Spec: 0.493  [TEST SCORE: 0.993]

###### #123: Age + First sexual intercourse* + Smokes (packs/year) + Hormonal Contraceptives (years) + STDs (number)

Optimized Thresholds: Age: 33.7196 | First sexual intercourse*: 24.7996 | Smokes (packs/year): 0.7909 | Hormonal Contraceptives (years): 11.2427 | STDs (number): 0.6337

Train Sens: 0.577 | Train Spec: 0.607  [TRAIN SCORE: 1.184]  ||  Val Sens: 0.778 | Val Spec: 0.444  [VAL SCORE: 1.222]  ||  Test Sens: 0.500 | Test Spec: 0.659  [TEST SCORE: 1.159]

###### #124: Age + Number of sexual partners + Num of pregnancies + Smokes (packs/year) + Hormonal Contraceptives (year

Optimized Thresholds: Age: 46.5395 | Number of sexual partners: 2.6454 | Num of pregnancies: 6.4625 | Smokes (packs/year): 0.0256 | Hormonal Contraceptives (years): 20.8166 | IUD (years): 2.4027

Train Sens: 0.654 | Train Spec: 0.607  [TRAIN SCORE: 1.261]  ||  Val Sens: 0.778 | Val Spec: 0.444  [VAL SCORE: 1.222]  ||  Test Sens: 0.500 | Test Spec: 0.486  [TEST SCORE: 0.986]

###### #125: Age + First sexual intercourse + Num of pregnancies + Hormonal Contraceptives (years)

Optimized Thresholds: Age: 33.9699 | First sexual intercourse: 22.0269 | Num of pregnancies: 2.4737 | Hormonal Contraceptives (years): 11.3015

Train Sens: 0.577 | Train Spec: 0.607  [TRAIN SCORE: 1.184]  ||  Val Sens: 0.778 | Val Spec: 0.444  [VAL SCORE: 1.222]  ||  Test Sens: 0.700 | Test Spec: 0.563  [TEST SCORE: 1.263]

###### #126: Age + Number of sexual partners + First sexual intercourse + Smokes (packs/year) + Hormonal Contraceptives (y

Optimized Thresholds: Age: 47.5212 | Number of sexual partners: 3.3826 | First sexual intercourse: 22.3251 | Smokes (packs/year): 0.0283 | Hormonal Contraceptives (years): 9.4977

Train Sens: 0.615 | Train Spec: 0.714  [TRAIN SCORE: 1.330]  ||  Val Sens: 0.556 | Val Spec: 0.667  [VAL SCORE: 1.222]  ||  Test Sens: 0.400 | Test Spec: 0.660  [TEST SCORE: 1.060]

###### #127: Age + First sexual intercourse + Smokes (packs/year)

Optimized Thresholds: Age: 46.5310 | First sexual intercourse: 17.4810 | Smokes (packs/year): 0.0263

Train Sens: 0.615 | Train Spec: 0.571  [TRAIN SCORE: 1.187]  ||  Val Sens: 0.778 | Val Spec: 0.444  [VAL SCORE: 1.222]  ||  Test Sens: 0.500 | Test Spec: 0.515  [TEST SCORE: 1.015]

##### #128: Age + First sexual intercourse + Smokes (packs/year) + STDs (number)

Optimized Thresholds: Age: 46.1115 | First sexual intercourse: 17.5389 | Smokes (packs/year): 0.0322 | STDs (number): 2.3930

Train Sens: 0.654 | Train Spec: 0.571 [TRAIN SCORE: 1.225] || Val Sens: 0.778 | Val Spec: 0.444 [VAL SCORE: 1.222] || Test Sens: 0.500 | Test Spec: 0.510 [TEST SCORE: 1.010]

##### #129: Age + First sexual intercourse + Num of pregnancies + Hormonal Contraceptives (years) + IUD (years)

Optimized Thresholds: Age: 33.9999 | First sexual intercourse: 21.9697 | Num of pregnancies: 2.5115 | Hormonal Contraceptives (years): 11.3531 | IUD (years): 1.0689

Train Sens: 0.615 | Train Spec: 0.607 [TRAIN SCORE: 1.223] || Val Sens: 0.778 | Val Spec: 0.444 [VAL SCORE: 1.222] || Test Sens: 0.700 | Test Spec: 0.536 [TEST SCORE: 1.236]

##### #130: Smokes (packs/year) + Hormonal Contraceptives (years) + STDs (number)

Optimized Thresholds: Smokes (packs/year): 2.0793 | Hormonal Contraceptives (years): 1.8626 | STDs (number): 0.4746

Train Sens: 0.615 | Train Spec: 0.571 [TRAIN SCORE: 1.187] || Val Sens: 0.667 | Val Spec: 0.556 [VAL SCORE: 1.222] || Test Sens: 0.400 | Test Spec: 0.582 [TEST SCORE: 0.982]

##### #131: Number of sexual partners + First sexual intercourse + Smokes (packs/year) + Hormonal Contraceptives (years) +

Optimized Thresholds: Number of sexual partners: 3.4764 | First sexual intercourse: 21.9140 | Smokes (packs/year): 0.0275 | Hormonal Contraceptives (years): 10.7009 | IUD (years): 1.7790

Train Sens: 0.654 | Train Spec: 0.714 [TRAIN SCORE: 1.368] || Val Sens: 0.778 | Val Spec: 0.444 [VAL SCORE: 1.222] || Test Sens: 0.500 | Test Spec: 0.618 [TEST SCORE: 1.118]

##### #132: Age + Num of pregnancies + Smokes (packs/year) + STDs (number)

Optimized Thresholds: Age: 33.8500 | Num of pregnancies: 3.2805 | Smokes (packs/year): 0.9042 | STDs (number): 0.6707

Train Sens: 0.577 | Train Spec: 0.571 [TRAIN SCORE: 1.148] || Val Sens: 0.778 | Val Spec: 0.444 [VAL SCORE: 1.222] || Test Sens: 0.700 | Test Spec: 0.619 [TEST SCORE: 1.319]

##### #133: Number of sexual partners + Num of pregnancies + Smokes (packs/year) + IUD (years) + STDs (number)

Optimized Thresholds: Number of sexual partners: 2.6064 | Num of pregnancies: 6.5366 | Smokes (packs/year): 0.0343 | IUD (years): 1.8154 | STDs (number): 2.3563

Train Sens: 0.615 | Train Spec: 0.607 [TRAIN SCORE: 1.223] || Val Sens: 0.778 | Val Spec: 0.444 [VAL SCORE: 1.222] || Test Sens: 0.500 | Test Spec: 0.481 [TEST SCORE: 0.981]

##### #134: Age + Number of sexual partners + First sexual intercourse + Num of pregnancies + Smokes (packs/year) + Horm

Optimized Thresholds: Age: 48.2384 | Number of sexual partners: 3.4304 | First sexual intercourse: 22.0068 | Num of pregnancies: 5.7835 | Smokes (packs/year): 0.0301 | Hormonal Contraceptives (years): 9.4197

Train Sens: 0.615 | Train Spec: 0.714 [TRAIN SCORE: 1.330] || Val Sens: 0.556 | Val Spec: 0.667 [VAL SCORE: 1.222] || Test Sens: 0.400 | Test Spec: 0.643 [TEST SCORE: 1.043]

##### #135: Age + Num of pregnancies + Smokes (packs/year) + Hormonal Contraceptives (years)

Optimized Thresholds: Age: 32.4956 | Num of pregnancies: 2.4676 | Smokes (packs/year): 0.0250 | Hormonal Contraceptives (years): 11.3216

Train Sens: 0.577 | Train Spec: 0.571 [TRAIN SCORE: 1.148] || Val Sens: 0.889 | Val Spec: 0.333 [VAL SCORE: 1.222] || Test Sens: 0.800 | Test Spec: 0.488 [TEST SCORE: 1.288]

##### #136: Age + Num of pregnancies + Smokes (packs/year) + IUD (years)

Optimized Thresholds: Age: 32.5536 | Num of pregnancies: 2.5244 | Smokes (packs/year): 0.0333 | IUD (years): 1.6018

Train Sens: 0.577 | Train Spec: 0.571 [TRAIN SCORE: 1.148] || Val Sens: 0.889 | Val Spec: 0.333 [VAL SCORE: 1.222] || Test Sens: 0.800 | Test Spec: 0.473 [TEST SCORE: 1.273]

##### #137: Age + Num of pregnancies + Smokes (packs/year) + Hormonal Contraceptives (years) + IUD (years) + STDs (num[illegible]

Optimized Thresholds: Age: 33.9832 | Num of pregnancies: 2.3752 | Smokes (packs/year): 0.0290 | Hormonal Contraceptives (years): 11.5081 | IUD (years): 1.6568 | STDs (number): 2.4979

Train Sens: 0.615 | Train Spec: 0.607 [TRAIN SCORE: 1.223] || Val Sens: 0.889 | Val Spec: 0.333 [VAL SCORE: 1.222] || Test Sens: 0.800 | Test Spec: 0.480 [TEST SCORE: 1.280]

##### #138: Age + Num of pregnancies + Smokes (packs/year)

Optimized Thresholds: Age: 32.6669 | Num of pregnancies: 2.4166 | Smokes (packs/year): 0.0347

Train Sens: 0.538 | Train Spec: 0.571 [TRAIN SCORE: 1.110] || Val Sens: 0.889 | Val Spec: 0.333 [VAL SCORE: 1.222] || Test Sens: 0.800 | Test Spec: 0.490 [TEST SCORE: 1.290]

##### #139: Age + Num of pregnancies + Smokes (packs/year) + IUD (years) + STDs (number)

Optimized Thresholds: Age: 33.8326 | Num of pregnancies: 2.5242 | Smokes (packs/year): 0.0291 | IUD (years): 1.6093 | STDs (number): 2.5712

Train Sens: 0.577 | Train Spec: 0.607 [TRAIN SCORE: 1.184] || Val Sens: 0.889 | Val Spec: 0.333 [VAL SCORE: 1.222] || Test Sens: 0.800 | Test Spec: 0.483 [TEST SCORE: 1.283]

##### #140: Age + Number of sexual partners + First sexual intercourse + Smokes (packs/year) + IUD (years)

Optimized Thresholds: Age: 45.7107 | Number of sexual partners: 3.5183 | First sexual intercourse: 22.0374 | Smokes (packs/year): 0.0263 | IUD (years): 1.3814

Train Sens: 0.615 | Train Spec: 0.786 [TRAIN SCORE: 1.401] || Val Sens: 0.667 | Val Spec: 0.444 [VAL SCORE: 1.111] || Test Sens: 0.400 | Test Spec: 0.628 [TEST SCORE: 1.028]

##### #141: Age + Smokes (packs/year) + Hormonal Contraceptives (years) + IUD (years) + STDs (number)

Optimized Thresholds: Age: 47.6780 | Smokes (packs/year): 0.0242 | Hormonal Contraceptives (years): 0.6967 | IUD (years): 2.0903 | STDs (number): 2.6635

Train Sens: 0.654 | Train Spec: 0.571 [TRAIN SCORE: 1.225] || Val Sens: 0.778 | Val Spec: 0.333 [VAL SCORE: 1.111] || Test Sens: 0.400 | Test Spec: 0.427 [TEST SCORE: 0.827]

##### #142: Age + Number of sexual partners + First sexual intercourse + Num of pregnancies + Smokes (packs/year) + IUD ([illegible]

Optimized Thresholds: Age: 45.5954 | Number of sexual partners: 3.4920 | First sexual intercourse: 21.3626 | Num of pregnancies: 6.5819 | Smokes (packs/year): 0.0296 | IUD (years): 1.5686

Train Sens: 0.615 | Train Spec: 0.786 [TRAIN SCORE: 1.401] || Val Sens: 0.667 | Val Spec: 0.444 [VAL SCORE: 1.111] || Test Sens: 0.400 | Test Spec: 0.616 [TEST SCORE: 1.016]

##### #143: Smokes (packs/year) + Hormonal Contraceptives (years) + IUD (years) + STDs (number)

Optimized Thresholds: Smokes (packs/year): 0.0158 | Hormonal Contraceptives (years): 0.7013 | IUD (years): 2.0267 | STDs (number): 2.5039

Train Sens: 0.654 | Train Spec: 0.571 [TRAIN SCORE: 1.225] || Val Sens: 0.778 | Val Spec: 0.333 [VAL SCORE: 1.111] || Test Sens: 0.400 | Test Spec: 0.430 [TEST SCORE: 0.830]

##### #144: Age + Number of sexual partners + Num of pregnancies + Hormonal Contraceptives (years) + IUD (years)

Optimized Thresholds: Age: 45.9571 | Number of sexual partners: 2.3839 | Num of pregnancies: 6.3687 | Hormonal Contraceptives (years): 21.0484 | IUD (years): 1.5186

Train Sens: 0.615 | Train Spec: 0.607 [TRAIN SCORE: 1.223] || Val Sens: 0.556 | Val Spec: 0.556 [VAL SCORE: 1.111] || Test Sens: 0.400 | Test Spec: 0.517 [TEST SCORE: 0.917]

##### #145: Age + First sexual intercourse + Num of pregnancies + Hormonal Contraceptives (years) + IUD (years) + STDs (nu

Optimized Thresholds: Age: 33.9280 | First sexual intercourse: 21.9711 | Num of pregnancies: 2.5506 | Hormonal Contraceptives (years): 11.0282 | IUD (years): 0.9629 | STDs (number): 2.5553

Train Sens: 0.654 | Train Spec: 0.607 [TRAIN SCORE: 1.261] || Val Sens: 0.778 | Val Spec: 0.333 [VAL SCORE: 1.111] || Test Sens: 0.700 | Test Spec: 0.527 [TEST SCORE: 1.227]

##### #146: Age + First sexual intercourse + Num of pregnancies + Smokes (packs/year) + Hormonal Contraceptives (years)

Optimized Thresholds: Age: 33.9250 | First sexual intercourse: 21.8240 | Num of pregnancies: 2.3718 | Smokes (packs/year): 0.0165 | Hormonal Contraceptives (years): 11.7415

Train Sens: 0.615 | Train Spec: 0.607 [TRAIN SCORE: 1.223] || Val Sens: 0.889 | Val Spec: 0.222 [VAL SCORE: 1.111] || Test Sens: 0.800 | Test Spec: 0.476 [TEST SCORE: 1.276]

##### #147: Age + First sexual intercourse + Num of pregnancies + Smokes (packs/year) + IUD (years)

Optimized Thresholds: Age: 33.8001 | First sexual intercourse: 21.9442 | Num of pregnancies: 2.3824 | Smokes (packs/year): 0.0228 | IUD (years): 1.2881

Train Sens: 0.615 | Train Spec: 0.607 [TRAIN SCORE: 1.223] || Val Sens: 0.889 | Val Spec: 0.222 [VAL SCORE: 1.111] || Test Sens: 0.800 | Test Spec: 0.462 [TEST SCORE: 1.262]

##### #148: Age + First sexual intercourse + Num of pregnancies + Hormonal Contraceptives (years) + STDs (number)

Optimized Thresholds: Age: 33.7968 | First sexual intercourse: 22.2994 | Num of pregnancies: 2.5493 | Hormonal Contraceptives (years): 11.5715 | STDs (number): 2.5245

Train Sens: 0.615 | Train Spec: 0.607 [TRAIN SCORE: 1.223] || Val Sens: 0.778 | Val Spec: 0.333 [VAL SCORE: 1.111] || Test Sens: 0.700 | Test Spec: 0.558 [TEST SCORE: 1.258]

##### #149: Age + First sexual intercourse + Num of pregnancies + Smokes (packs/year) + Hormonal Contraceptives (years) +

Optimized Thresholds: Age: 33.9792 | First sexual intercourse: 22.0821 | Num of pregnancies: 2.3864 | Smokes (packs/year): 0.0171 | Hormonal Contraceptives (years): 11.8645 | IUD (years): 0.8573

Train Sens: 0.654 | Train Spec: 0.607 [TRAIN SCORE: 1.261] || Val Sens: 0.889 | Val Spec: 0.222 [VAL SCORE: 1.111] || Test Sens: 0.800 | Test Spec: 0.462 [TEST SCORE: 1.262]

##### #150: Number of sexual partners + First sexual intercourse + Num of pregnancies + Smokes (packs/year) + IUD (years)

Optimized Thresholds: Number of sexual partners: 2.4535 | First sexual intercourse: 22.0916 | Num of pregnancies: 6.4038 | Smokes (packs/year): 0.0268 | IUD (years): 1.7738

Train Sens: 0.692 | Train Spec: 0.607 [TRAIN SCORE: 1.299] || Val Sens: 0.778 | Val Spec: 0.333 [VAL SCORE: 1.111] || Test Sens: 0.500 | Test Spec: 0.456 [TEST SCORE: 0.956]

##### #151: Age + First sexual intercourse + Hormonal Contraceptives (years) + IUD (years)* + STDs (number)

Optimized Thresholds: Age: 48.0807 | First sexual intercourse: 24.0867 | Hormonal Contraceptives (years): 1.9250 | IUD (years)*: 4.7700 | STDs (number): 0.4657

Train Sens: 0.654 | Train Spec: 0.607  [TRAIN SCORE: 1.261]  ||  Val Sens: 0.667 | Val Spec: 0.444  [VAL SCORE: 1.111]  ||  Test Sens: 0.400 | Test Spec: 0.544  [TEST SCORE: 0.944]

##### #152: Number of sexual partners + First sexual intercourse + Num of pregnancies + Smokes (packs/year) + Hormonal C

Optimized Thresholds: Number of sexual partners: 3.5104 | First sexual intercourse: 21.8339 | Num of pregnancies: 5.7626 | Smokes (packs/year): 0.0289 | Hormonal Contraceptives (years): 9.5721

Train Sens: 0.615 | Train Spec: 0.714  [TRAIN SCORE: 1.330]  ||  Val Sens: 0.444 | Val Spec: 0.667  [VAL SCORE: 1.111]  ||  Test Sens: 0.400 | Test Spec: 0.638  [TEST SCORE: 1.038]

##### #153: Age + Number of sexual partners + First sexual intercourse + Num of pregnancies + Smokes (packs/year)

Optimized Thresholds: Age: 46.1930 | Number of sexual partners: 2.7399 | First sexual intercourse: 22.4467 | Num of pregnancies: 6.6186 | Smokes (packs/year): 0.0269

Train Sens: 0.692 | Train Spec: 0.607  [TRAIN SCORE: 1.299]  ||  Val Sens: 0.667 | Val Spec: 0.444  [VAL SCORE: 1.111]  ||  Test Sens: 0.400 | Test Spec: 0.481  [TEST SCORE: 0.881]

##### #154: Age + Number of sexual partners

Optimized Thresholds: Age: 46.0015 | Number of sexual partners: 2.6820

Train Sens: 0.577 | Train Spec: 0.607  [TRAIN SCORE: 1.184]  ||  Val Sens: 0.444 | Val Spec: 0.667  [VAL SCORE: 1.111]  ||  Test Sens: 0.300 | Test Spec: 0.553  [TEST SCORE: 0.853]

##### #155: Number of sexual partners + First sexual intercourse + Smokes (packs/year) + Hormonal Contraceptives (years)

Optimized Thresholds: Number of sexual partners: 3.6094 | First sexual intercourse: 21.9732 | Smokes (packs/year): 0.0256 | Hormonal Contraceptives (years): 9.6771

Train Sens: 0.615 | Train Spec: 0.714  [TRAIN SCORE: 1.330]  ||  Val Sens: 0.444 | Val Spec: 0.667  [VAL SCORE: 1.111]  ||  Test Sens: 0.400 | Test Spec: 0.660  [TEST SCORE: 1.060]

##### #156: Age + Number of sexual partners* + Hormonal Contraceptives (years)* + IUD (years)

Optimized Thresholds: Age: 46.0092 | Number of sexual partners*: 2.8658 | Hormonal Contraceptives (years)*: 18.9323 | IUD (years): 2.0439

Train Sens: 0.615 | Train Spec: 0.571  [TRAIN SCORE: 1.187]  ||  Val Sens: 0.556 | Val Spec: 0.556  [VAL SCORE: 1.111]  ||  Test Sens: 0.400 | Test Spec: 0.524  [TEST SCORE: 0.924]

##### #157: Number of sexual partners + Num of pregnancies + IUD (years)

Optimized Thresholds: Number of sexual partners: 2.5065 | Num of pregnancies: 6.5184 | IUD (years): 1.7787

Train Sens: 0.577 | Train Spec: 0.607  [TRAIN SCORE: 1.184]  ||  Val Sens: 0.556 | Val Spec: 0.556  [VAL SCORE: 1.111]  ||  Test Sens: 0.400 | Test Spec: 0.524  [TEST SCORE: 0.924]

##### #158: Age + Number of sexual partners + Num of pregnancies + IUD (years)

Optimized Thresholds: Age: 46.1474 | Number of sexual partners: 2.5690 | Num of pregnancies: 6.5037 | IUD (years): 1.8648

Train Sens: 0.615 | Train Spec: 0.607  [TRAIN SCORE: 1.223]  ||  Val Sens: 0.556 | Val Spec: 0.556  [VAL SCORE: 1.111]  ||  Test Sens: 0.400 | Test Spec: 0.520  [TEST SCORE: 0.920]

##### #159: Age + Number of sexual partners + Num of pregnancies + Hormonal Contraceptives (years)

Optimized Thresholds: Age: 45.7073 | Number of sexual partners: 2.4259 | Num of pregnancies: 6.4059 | Hormonal Contraceptives (years): 20.8937

Train Sens: 0.577 | Train Spec: 0.607 [TRAIN SCORE: 1.184] || Val Sens: 0.444 | Val Spec: 0.667 [VAL SCORE: 1.111] || Test Sens: 0.300 | Test Spec: 0.548 [TEST SCORE: 0.848]

###### #160: Age + Smokes (packs/year) + Hormonal Contraceptives (years)

Optimized Thresholds: Age: 48.5457 | Smokes (packs/year): 0.0270 | Hormonal Contraceptives (years): 0.6948

Train Sens: 0.615 | Train Spec: 0.571 [TRAIN SCORE: 1.187] || Val Sens: 0.556 | Val Spec: 0.556 [VAL SCORE: 1.111] || Test Sens: 0.400 | Test Spec: 0.462 [TEST SCORE: 0.862]

###### #161: Age + First sexual intercourse + IUD (years)* + STDs (number)

Optimized Thresholds: Age: 45.8027 | First sexual intercourse: 17.3427 | IUD (years)*: 5.3571 | STDs (number): 0.4603

Train Sens: 0.615 | Train Spec: 0.571 [TRAIN SCORE: 1.187] || Val Sens: 0.778 | Val Spec: 0.333 [VAL SCORE: 1.111] || Test Sens: 0.500 | Test Spec: 0.536 [TEST SCORE: 1.036]

###### #162: First sexual intercourse + IUD (years) + STDs (number)

Optimized Thresholds: First sexual intercourse: 17.3598 | IUD (years): 5.0054 | STDs (number): 0.6670

Train Sens: 0.538 | Train Spec: 0.571 [TRAIN SCORE: 1.110] || Val Sens: 0.778 | Val Spec: 0.333 [VAL SCORE: 1.111] || Test Sens: 0.500 | Test Spec: 0.549 [TEST SCORE: 1.049]

###### #163: Age + First sexual intercourse + IUD (years)

Optimized Thresholds: Age: 45.7618 | First sexual intercourse: 17.4900 | IUD (years): 5.3001

Train Sens: 0.500 | Train Spec: 0.643 [TRAIN SCORE: 1.143] || Val Sens: 0.667 | Val Spec: 0.444 [VAL SCORE: 1.111] || Test Sens: 0.500 | Test Spec: 0.590 [TEST SCORE: 1.090]

###### #164: Age + Smokes (packs/year) + Hormonal Contraceptives (years) + IUD (years)

Optimized Thresholds: Age: 48.4059 | Smokes (packs/year): 0.0248 | Hormonal Contraceptives (years): 0.6995 | IUD (years): 2.1203

Train Sens: 0.654 | Train Spec: 0.571 [TRAIN SCORE: 1.225] || Val Sens: 0.778 | Val Spec: 0.333 [VAL SCORE: 1.111] || Test Sens: 0.400 | Test Spec: 0.430 [TEST SCORE: 0.830]

###### #165: Age + Hormonal Contraceptives (years) + IUD (years)* + STDs (number)

Optimized Thresholds: Age: 49.0199 | Hormonal Contraceptives (years): 1.9082 | IUD (years)*: 3.9455 | STDs (number): 0.6455

Train Sens: 0.615 | Train Spec: 0.607 [TRAIN SCORE: 1.223] || Val Sens: 0.667 | Val Spec: 0.444 [VAL SCORE: 1.111] || Test Sens: 0.400 | Test Spec: 0.561 [TEST SCORE: 0.961]

###### #166: Number of sexual partners + Smokes (packs/year) + Hormonal Contraceptives (years)

Optimized Thresholds: Number of sexual partners: 2.5042 | Smokes (packs/year): 0.0210 | Hormonal Contraceptives (years): 20.8557

Train Sens: 0.577 | Train Spec: 0.607 [TRAIN SCORE: 1.184] || Val Sens: 0.556 | Val Spec: 0.556 [VAL SCORE: 1.111] || Test Sens: 0.400 | Test Spec: 0.522 [TEST SCORE: 0.922]

###### #167: Number of sexual partners + Smokes (packs/year) + STDs (number)

Optimized Thresholds: Number of sexual partners: 2.4108 | Smokes (packs/year): 0.0218 | STDs (number): 2.6057

Train Sens: 0.577 | Train Spec: 0.607 [TRAIN SCORE: 1.184] || Val Sens: 0.556 | Val Spec: 0.556 [VAL SCORE: 1.111] || Test Sens: 0.400 | Test Spec: 0.517 [TEST SCORE: 0.917]

###### #168: Age + Number of sexual partners + IUD (years)

Optimized Thresholds: Age: 46.1571 | Number of sexual partners: 2.3215 | IUD (years): 1.8718

Train Sens: 0.615 | Train Spec: 0.607 [TRAIN SCORE: 1.223] || Val Sens: 0.556 | Val Spec: 0.556 [VAL SCORE: 1.111] || Test Sens: 0.400 | Test Spec: 0.522 [TEST SCORE: 0.922]

##### #169: Age + Number of sexual partners* + Hormonal Contraceptives (years)*

Optimized Thresholds: Age: 46.0373 | Number of sexual partners*: 3.0655 | Hormonal Contraceptives (years)*: 16.8196

Train Sens: 0.308 | Train Spec: 0.714 [TRAIN SCORE: 1.022] || Val Sens: 0.222 | Val Spec: 0.889 [VAL SCORE: 1.111] || Test Sens: 0.200 | Test Spec: 0.812 [TEST SCORE: 1.012]

##### #170: Number of sexual partners + First sexual intercourse + Smokes (packs/year) + IUD (years)

Optimized Thresholds: Number of sexual partners: 2.4195 | First sexual intercourse: 21.6128 | Smokes (packs/year): 0.0188 | IUD (years): 1.8180

Train Sens: 0.692 | Train Spec: 0.607 [TRAIN SCORE: 1.299] || Val Sens: 0.778 | Val Spec: 0.333 [VAL SCORE: 1.111] || Test Sens: 0.500 | Test Spec: 0.452 [TEST SCORE: 0.952]

##### #171: Age + Number of sexual partners + First sexual intercourse + Smokes (packs/year)

Optimized Thresholds: Age: 46.0583 | Number of sexual partners: 2.4703 | First sexual intercourse: 22.0741 | Smokes (packs/year): 0.0328

Train Sens: 0.692 | Train Spec: 0.607 [TRAIN SCORE: 1.299] || Val Sens: 0.667 | Val Spec: 0.444 [VAL SCORE: 1.111] || Test Sens: 0.400 | Test Spec: 0.483 [TEST SCORE: 0.883]

##### #172: Age + Number of sexual partners + Num of pregnancies

Optimized Thresholds: Age: 45.9864 | Number of sexual partners: 2.5253 | Num of pregnancies: 6.5192

Train Sens: 0.577 | Train Spec: 0.607 [TRAIN SCORE: 1.184] || Val Sens: 0.444 | Val Spec: 0.667 [VAL SCORE: 1.111] || Test Sens: 0.300 | Test Spec: 0.548 [TEST SCORE: 0.848]

##### #173: Hormonal Contraceptives (years) + IUD (years)* + STDs (number)

Optimized Thresholds: Hormonal Contraceptives (years): 2.2801 | IUD (years)*: 4.2576 | STDs (number): 0.6647

Train Sens: 0.615 | Train Spec: 0.607 [TRAIN SCORE: 1.223] || Val Sens: 0.667 | Val Spec: 0.444 [VAL SCORE: 1.111] || Test Sens: 0.400 | Test Spec: 0.628 [TEST SCORE: 1.028]

##### #174: First sexual intercourse + IUD (years)

Optimized Thresholds: First sexual intercourse: 17.4821 | IUD (years): 0.8039

Train Sens: 0.462 | Train Spec: 0.571 [TRAIN SCORE: 1.033] || Val Sens: 0.667 | Val Spec: 0.444 [VAL SCORE: 1.111] || Test Sens: 0.500 | Test Spec: 0.568 [TEST SCORE: 1.068]

##### #175: Number of sexual partners + IUD (years)

Optimized Thresholds: Number of sexual partners: 2.4753 | IUD (years): 1.8916

Train Sens: 0.577 | Train Spec: 0.607 [TRAIN SCORE: 1.184] || Val Sens: 0.556 | Val Spec: 0.556 [VAL SCORE: 1.111] || Test Sens: 0.400 | Test Spec: 0.529 [TEST SCORE: 0.929]

##### #176: Number of sexual partners + Smokes (packs/year)

Optimized Thresholds: Number of sexual partners: 2.4399 | Smokes (packs/year): 0.0304

Train Sens: 0.577 | Train Spec: 0.607 [TRAIN SCORE: 1.184] || Val Sens: 0.556 | Val Spec: 0.556 [VAL SCORE: 1.111] || Test Sens: 0.400 | Test Spec: 0.522 [TEST SCORE: 0.922]

##### #177: First sexual intercourse + Hormonal Contraceptives (years) + IUD (years) + STDs (number)

Optimized Thresholds: First sexual intercourse: 24.4354 | Hormonal Contraceptives (years): 2.2157 | IUD (years): 5.0927 | STDs (number): 0.6034

Train Sens: 0.654 | Train Spec: 0.607 [TRAIN SCORE: 1.261] || Val Sens: 0.667 | Val Spec: 0.444 [VAL SCORE: 1.111] || Test Sens: 0.400 | Test Spec: 0.606 [TEST SCORE: 1.006]

##### #178: Smokes (packs/year) + Hormonal Contraceptives (years) + IUD (years)

Optimized Thresholds: Smokes (packs/year): 0.0199 | Hormonal Contraceptives (years): 0.7070 | IUD (years): 1.7966

Train Sens: 0.654 | Train Spec: 0.571 [TRAIN SCORE: 1.225] || Val Sens: 0.778 | Val Spec: 0.333 [VAL SCORE: 1.111] || Test Sens: 0.400 | Test Spec: 0.428 [TEST SCORE: 0.828]

##### #179: Number of sexual partners + Num of pregnancies + Smokes (packs/year)

Optimized Thresholds: Number of sexual partners: 2.6520 | Num of pregnancies: 6.4158 | Smokes (packs/year): 0.0311

Train Sens: 0.577 | Train Spec: 0.607 [TRAIN SCORE: 1.184] || Val Sens: 0.556 | Val Spec: 0.556 [VAL SCORE: 1.111] || Test Sens: 0.400 | Test Spec: 0.517 [TEST SCORE: 0.917]

##### #180: Number of sexual partners + Hormonal Contraceptives (years) + IUD (years)

Optimized Thresholds: Number of sexual partners: 5.5658 | Hormonal Contraceptives (years): 0.7090 | IUD (years): 2.3403

Train Sens: 0.615 | Train Spec: 0.607 [TRAIN SCORE: 1.223] || Val Sens: 0.556 | Val Spec: 0.444 [VAL SCORE: 1.000] || Test Sens: 0.400 | Test Spec: 0.481 [TEST SCORE: 0.881]

##### #181: Number of sexual partners + IUD (years) + STDs (number)

Optimized Thresholds: Number of sexual partners: 2.5595 | IUD (years): 2.1254 | STDs (number): 2.4168

Train Sens: 0.577 | Train Spec: 0.607 [TRAIN SCORE: 1.184] || Val Sens: 0.556 | Val Spec: 0.444 [VAL SCORE: 1.000] || Test Sens: 0.400 | Test Spec: 0.524 [TEST SCORE: 0.924]

##### #182: First sexual intercourse + Smokes (packs/year) + Hormonal Contraceptives (years) + IUD (years) + STDs (number

Optimized Thresholds: First sexual intercourse: 21.8633 | Smokes (packs/year): 0.0242 | Hormonal Contraceptives (years): 1.7178 | IUD (years): 1.9997 | STDs (number): 2.3701

Train Sens: 0.615 | Train Spec: 0.607 [TRAIN SCORE: 1.223] || Val Sens: 0.778 | Val Spec: 0.222 [VAL SCORE: 1.000] || Test Sens: 0.300 | Test Spec: 0.488 [TEST SCORE: 0.788]

##### #183: First sexual intercourse + Smokes (packs/year) + IUD (years)

Optimized Thresholds: First sexual intercourse: 17.2829 | Smokes (packs/year): 0.0253 | IUD (years): 4.9694

Train Sens: 0.538 | Train Spec: 0.571 [TRAIN SCORE: 1.110] || Val Sens: 0.778 | Val Spec: 0.222 [VAL SCORE: 1.000] || Test Sens: 0.600 | Test Spec: 0.495 [TEST SCORE: 1.095]

##### #184: Age + Num of pregnancies + Hormonal Contraceptives (years)

Optimized Thresholds: Age: 47.7172 | Num of pregnancies: 5.7337 | Hormonal Contraceptives (years): 0.3437

Train Sens: 0.692 | Train Spec: 0.571 [TRAIN SCORE: 1.264] || Val Sens: 0.444 | Val Spec: 0.556 [VAL SCORE: 1.000] || Test Sens: 0.500 | Test Spec: 0.459 [TEST SCORE: 0.959]

##### #185: Age + Number of sexual partners + First sexual intercourse + Num of pregnancies + Hormonal Contraceptives (ye

Optimized Thresholds: Age: 46.0734 | Number of sexual partners: 2.3268 | First sexual intercourse: 22.0627 | Num of pregnancies: 6.4240 | Hormonal Contraceptives (years): 20.9391 | IUD (years): 2.0518

Train Sens: 0.692 | Train Spec: 0.607 [TRAIN SCORE: 1.299] || Val Sens: 0.556 | Val Spec: 0.444 [VAL SCORE: 1.000] || Test Sens: 0.400 | Test Spec: 0.491 [TEST SCORE: 0.891]

##### #186: Number of sexual partners + First sexual intercourse + IUD (years)

Optimized Thresholds: Number of sexual partners: 2.6779 | First sexual intercourse: 21.9275 | IUD (years): 2.1399

Train Sens: 0.654 | Train Spec: 0.607 [TRAIN SCORE: 1.261] || Val Sens: 0.556 | Val Spec: 0.444 [VAL SCORE: 1.000] || Test Sens: 0.400 | Test Spec: 0.491 [TEST SCORE: 0.891]

##### #187: Number of sexual partners + First sexual intercourse + Smokes (packs/year)

Optimized Thresholds: Number of sexual partners: 2.3297 | First sexual intercourse: 22.1348 | Smokes (packs/year): 0.0226

Train Sens: 0.654 | Train Spec: 0.607 [TRAIN SCORE: 1.261] || Val Sens: 0.556 | Val Spec: 0.444 [VAL SCORE: 1.000] || Test Sens: 0.400 | Test Spec: 0.490 [TEST SCORE: 0.890]

##### #188: Age + Hormonal Contraceptives (years) + IUD (years)

Optimized Thresholds: Age: 47.4635 | Hormonal Contraceptives (years): 0.7188 | IUD (years): 2.4785

Train Sens: 0.615 | Train Spec: 0.607 [TRAIN SCORE: 1.223] || Val Sens: 0.556 | Val Spec: 0.444 [VAL SCORE: 1.000] || Test Sens: 0.400 | Test Spec: 0.488 [TEST SCORE: 0.888]

##### #189: Age + First sexual intercourse* + Hormonal Contraceptives (years)

Optimized Thresholds: Age: 48.8505 | First sexual intercourse*: 23.9833 | Hormonal Contraceptives (years): 0.7686

Train Sens: 0.577 | Train Spec: 0.607 [TRAIN SCORE: 1.184] || Val Sens: 0.333 | Val Spec: 0.667 [VAL SCORE: 1.000] || Test Sens: 0.400 | Test Spec: 0.503 [TEST SCORE: 0.903]

##### #190: Age + Number of sexual partners + STDs (number)

Optimized Thresholds: Age: 45.9324 | Number of sexual partners: 2.5244 | STDs (number): 2.4290

Train Sens: 0.577 | Train Spec: 0.607 [TRAIN SCORE: 1.184] || Val Sens: 0.444 | Val Spec: 0.556 [VAL SCORE: 1.000] || Test Sens: 0.300 | Test Spec: 0.541 [TEST SCORE: 0.841]

##### #191: Age + First sexual intercourse + Smokes (packs/year) + Hormonal Contraceptives (years) + IUD (years) + STDs (n

Optimized Thresholds: Age: 47.7033 | First sexual intercourse: 22.0882 | Smokes (packs/year): 0.0217 | Hormonal Contraceptives (years): 2.2429 | IUD (years): 2.2257 | STDs (number): 2.4719

Train Sens: 0.615 | Train Spec: 0.607 [TRAIN SCORE: 1.223] || Val Sens: 0.778 | Val Spec: 0.222 [VAL SCORE: 1.000] || Test Sens: 0.300 | Test Spec: 0.534 [TEST SCORE: 0.834]

##### #192: Age + Number of sexual partners + First sexual intercourse

Optimized Thresholds: Age: 45.7653 | Number of sexual partners: 2.5709 | First sexual intercourse: 21.6861

Train Sens: 0.654 | Train Spec: 0.607 [TRAIN SCORE: 1.261] || Val Sens: 0.444 | Val Spec: 0.556 [VAL SCORE: 1.000] || Test Sens: 0.300 | Test Spec: 0.507 [TEST SCORE: 0.807]

##### #193: Hormonal Contraceptives (years) + IUD (years)

Optimized Thresholds: Hormonal Contraceptives (years): 0.7098 | IUD (years): 2.6397

Train Sens: 0.615 | Train Spec: 0.607 [TRAIN SCORE: 1.223] || Val Sens: 0.556 | Val Spec: 0.444 [VAL SCORE: 1.000] || Test Sens: 0.400 | Test Spec: 0.495 [TEST SCORE: 0.895]

##### #194: Smokes (packs/year) + Hormonal Contraceptives (years)

Optimized Thresholds: Smokes (packs/year): 0.0210 | Hormonal Contraceptives (years): 0.7224

Train Sens: 0.615 | Train Spec: 0.571 [TRAIN SCORE: 1.187] || Val Sens: 0.444 | Val Spec: 0.556 [VAL SCORE: 1.000] || Test Sens: 0.400 | Test Spec: 0.468 [TEST SCORE: 0.868]

##### #195: Number of sexual partners + Num of pregnancies

Optimized Thresholds: Number of sexual partners: 2.3117 | Num of pregnancies: 6.6075

Train Sens: 0.538 | Train Spec: 0.607 [TRAIN SCORE: 1.146] || Val Sens: 0.333 | Val Spec: 0.667 [VAL SCORE: 1.000] || Test Sens: 0.300 | Test Spec: 0.555 [TEST SCORE: 0.855]

##### #196: Age + Hormonal Contraceptives (years)

Optimized Thresholds: Age: 48.0604 | Hormonal Contraceptives (years): 0.6949

Train Sens: 0.577 | Train Spec: 0.607 [TRAIN SCORE: 1.184] || Val Sens: 0.333 | Val Spec: 0.667 [VAL SCORE: 1.000] || Test Sens: 0.400 | Test Spec: 0.526 [TEST SCORE: 0.926]

##### #197: Age + Number of sexual partners + First sexual intercourse + Num of pregnancies + Hormonal Contraceptives (ye[illegible]

Optimized Thresholds: Age: 45.7756 | Number of sexual partners: 2.6192 | First sexual intercourse: 22.1309 | Num of pregnancies: 6.6508 | Hormonal Contraceptives (years): 20.7731

Train Sens: 0.654 | Train Spec: 0.607 [TRAIN SCORE: 1.261] || Val Sens: 0.444 | Val Spec: 0.556 [VAL SCORE: 1.000] || Test Sens: 0.300 | Test Spec: 0.514 [TEST SCORE: 0.814]

##### #198: First sexual intercourse* + Hormonal Contraceptives (years) + IUD (years)

Optimized Thresholds: First sexual intercourse*: 23.7450 | Hormonal Contraceptives (years): 0.8063 | IUD (years): 2.4587

Train Sens: 0.615 | Train Spec: 0.607 [TRAIN SCORE: 1.223] || Val Sens: 0.556 | Val Spec: 0.444 [VAL SCORE: 1.000] || Test Sens: 0.400 | Test Spec: 0.471 [TEST SCORE: 0.871]

##### #199: Age + First sexual intercourse + Hormonal Contraceptives (years) + IUD (years)

Optimized Thresholds: Age: 47.6014 | First sexual intercourse: 23.0659 | Hormonal Contraceptives (years): 0.7588 | IUD (years): 2.4454

Train Sens: 0.615 | Train Spec: 0.607 [TRAIN SCORE: 1.223] || Val Sens: 0.556 | Val Spec: 0.444 [VAL SCORE: 1.000] || Test Sens: 0.400 | Test Spec: 0.466 [TEST SCORE: 0.866]

##### #200: Age + First sexual intercourse + Smokes (packs/year) + IUD (years)* + STDs (number)

Optimized Thresholds: Age: 46.1260 | First sexual intercourse: 17.5270 | Smokes (packs/year): 0.0226 | IUD (years)*: 4.6370 | STDs (number): 2.4488

Train Sens: 0.654 | Train Spec: 0.571 [TRAIN SCORE: 1.225] || Val Sens: 0.778 | Val Spec: 0.222 [VAL SCORE: 1.000] || Test Sens: 0.600 | Test Spec: 0.480 [TEST SCORE: 1.080]
