## Supplementary material. Outputs TholdStormDX for this study. for "Methodological and Clinical Validation of TholdStormDX v0.0.1: An Advanced Stochastic Engine for the Optimization of Thresholds and Multimarker Panels Applied to Oncology": HCC TholdStormDX_RobustReport_20260401_185100.pdf

### Biomarker: AGE

Processed: 01-Apr-2026 16:43

#### 1. Optimization Results

| MODEL | CUT-OFF | TRAIN (SE/SP) | VAL (SE/SP) | TEST (SE/SP) | R2 SCORE |
|---|---|---|---|---|---|
| Empirical (Exact) | 59.1144 | 0.557 / 0.557 | 0.463 / 0.487 | 0.585 / 0.692 | N/A |
| Logistic 2-Parameter | 59.9459 | 0.543 / 0.543 | 0.463 / 0.487 | 0.585 / 0.692 | 0.9937 |
| **Logistic 4-Parameter (Rec.)** | **59.8701** | **0.542 / 0.542** | **0.463 / 0.487** | **0.585 / 0.692** | **0.9948** |
| ThresholdXpert (Stochastic) | 58.3225 | 0.563 / 0.557 | 0.512 / 0.487 | 0.634 / 0.667 | N/A |

#### 2. Diagnostic Performance Curves (Training)

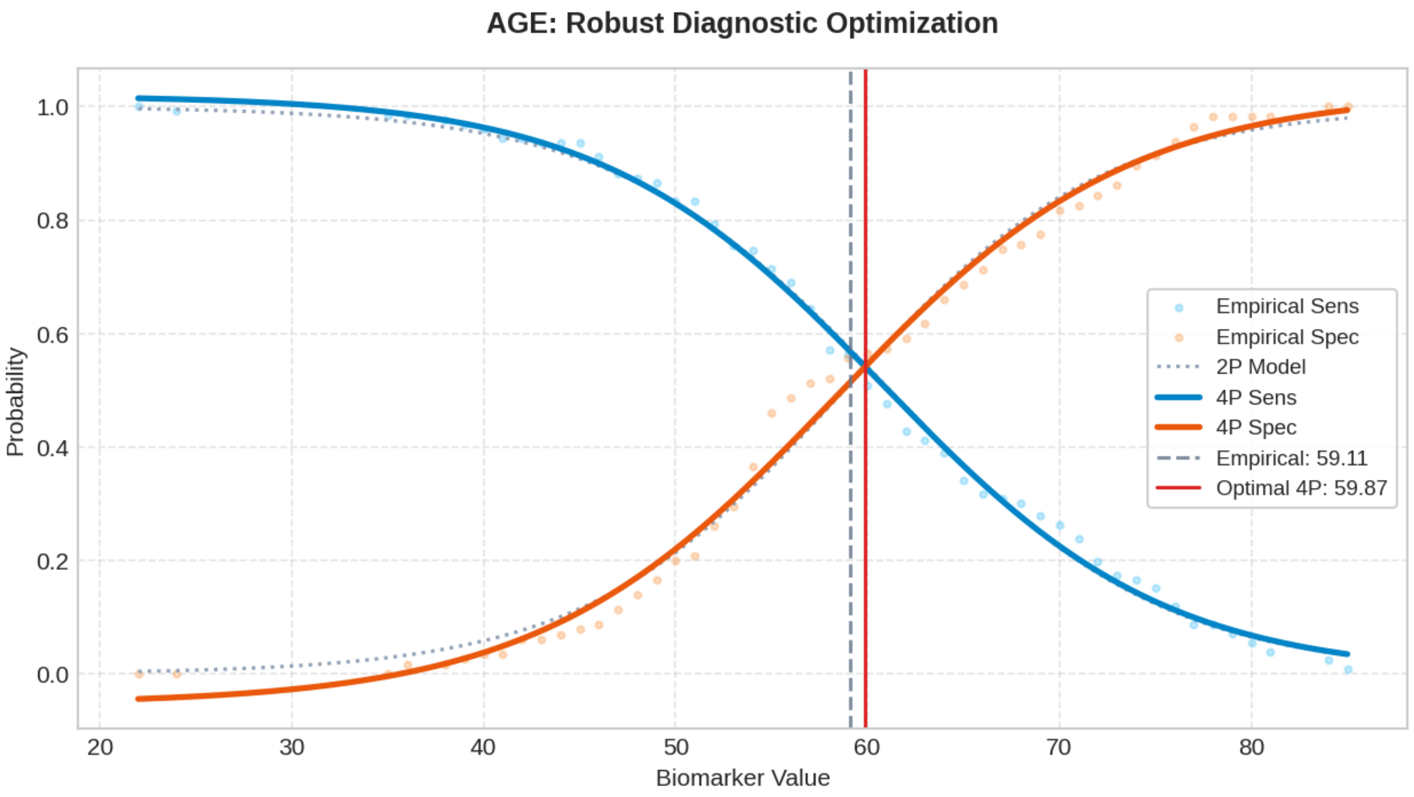

### Biomarker: CTP

Processed: 01-Apr-2026 16:44

#### 1. Optimization Results

| MODEL | CUT-OFF | TRAIN (SE/SP) | VAL (SE/SP) | TEST (SE/SP) | R2 SCORE |
|---|---|---|---|---|---|
| Empirical (Exact) | 5.7790 | 0.549 / 0.549 | 0.293 / 0.564 | 0.366 / 0.718 | N/A |
| Logistic 2-Parameter | 5.8763 | 0.572 / 0.572 | 0.293 / 0.564 | 0.366 / 0.718 | 0.9576 |
| **Logistic 4-Parameter (Rec.)** | **5.8139** | **0.562 / 0.562** | **0.293 / 0.564** | **0.366 / 0.718** | **0.9877** |
| ThresholdXpert (Stochastic) | 5.4267 | 0.421 / 0.704 | 0.293 / 0.564 | 0.366 / 0.718 | N/A |

#### 2. Diagnostic Performance Curves (Training)

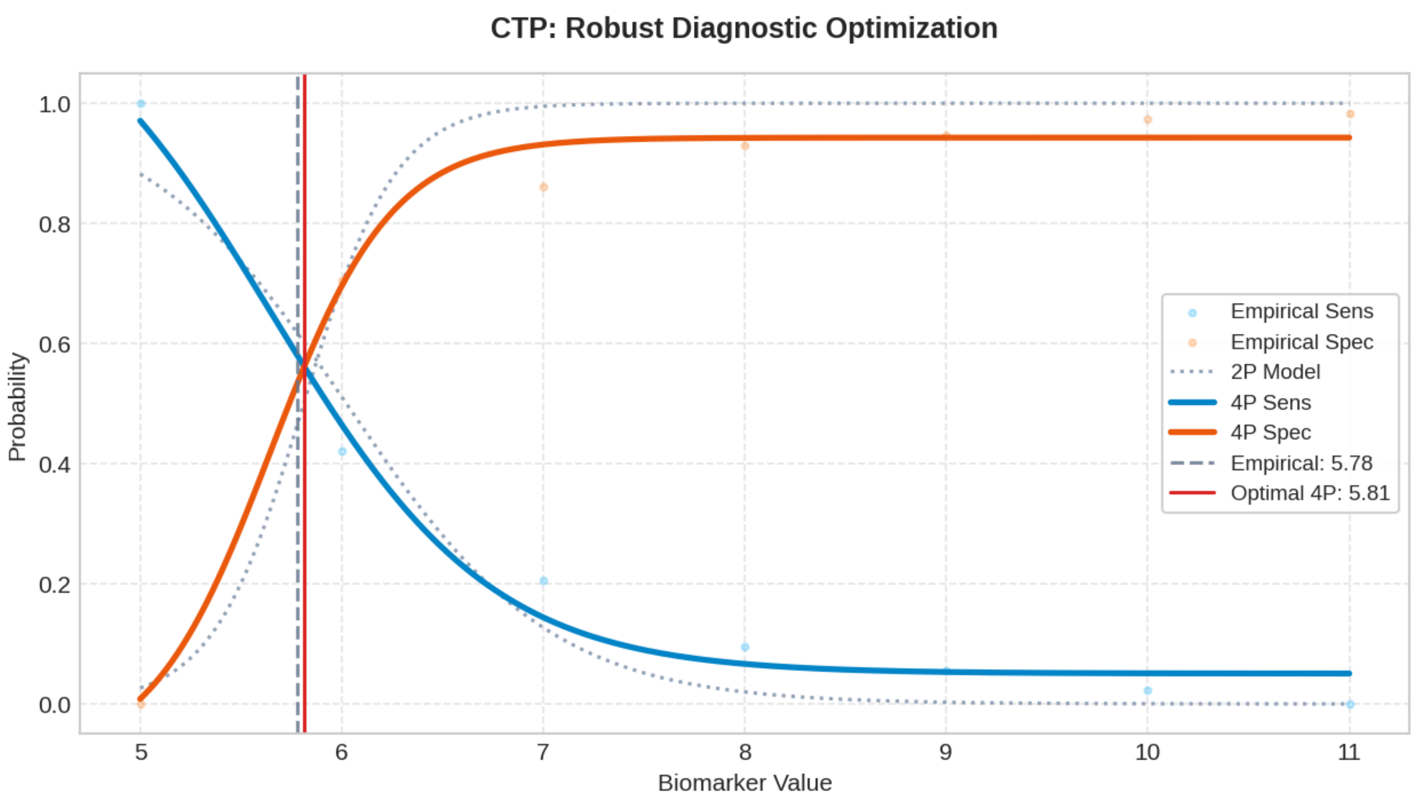

### Biomarker: MELD

Processed: 01-Apr-2026 16:47

#### 1. Optimization Results

| MODEL | CUT-OFF | TRAIN (SE/SP) | VAL (SE/SP) | TEST (SE/SP) | R2 SCORE |
|---|---|---|---|---|---|
| Empirical (Exact) | 6.8424 | 0.356 / 0.356 | 0.366 / 0.205 | 0.439 / 0.282 | N/A |
| Logistic 2-Parameter | 6.7839 | 0.368 / 0.368 | 0.390 / 0.205 | 0.439 / 0.282 | 0.9947 |
| **Logistic 4-Parameter (Rec.)** | **6.7296** | **0.367 / 0.367** | **0.415 / 0.205** | **0.439 / 0.282** | **0.9963** |
| ThresholdXpert (Stochastic) | 6.7691 | 0.381 / 0.357 | 0.390 / 0.205 | 0.439 / 0.282 | N/A |

#### 2. Diagnostic Performance Curves (Training)

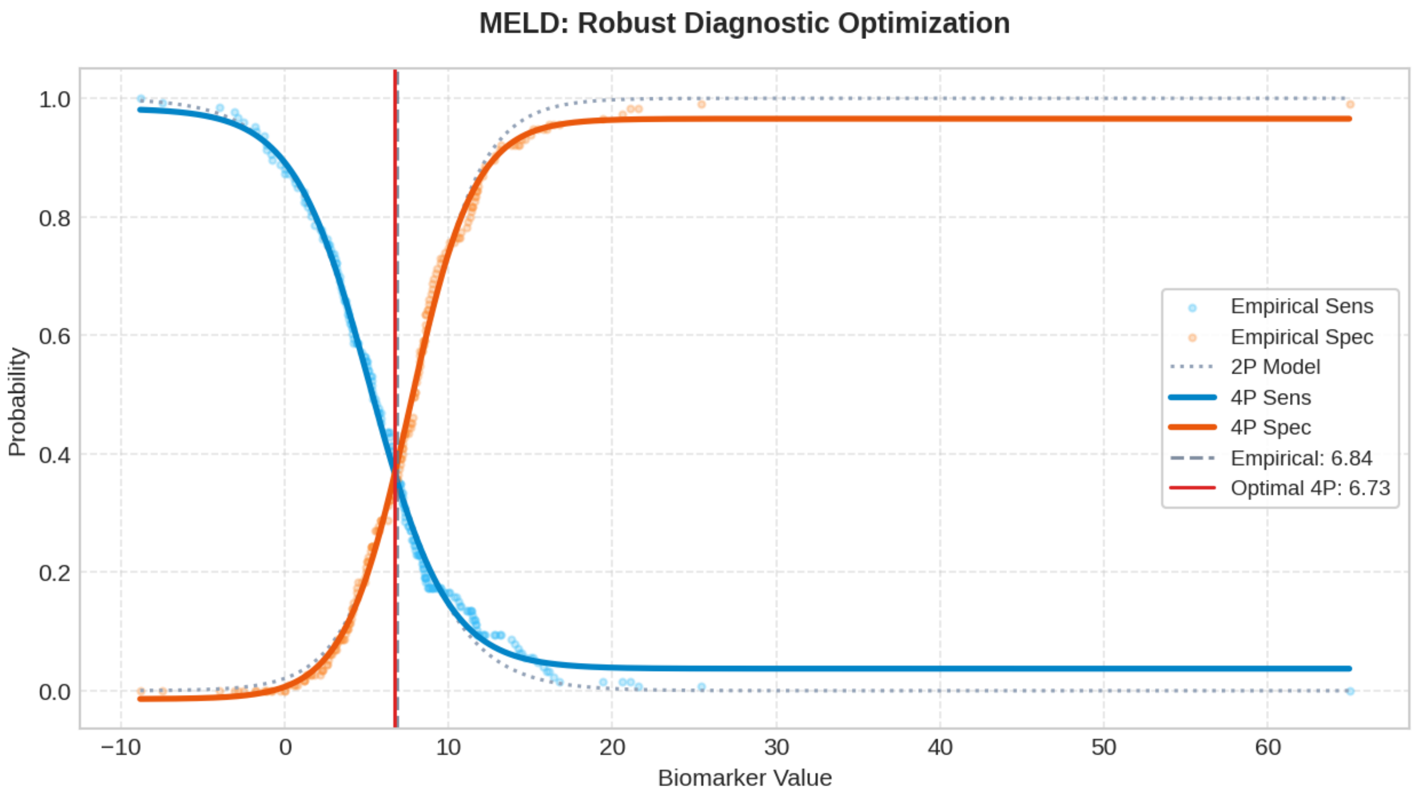

### Biomarker: INV. MELD

Processed: 01-Apr-2026 16:50

#### 1. Optimization Results

| MODEL | CUT-OFF | TRAIN (SE/SP) | VAL (SE/SP) | TEST (SE/SP) | R2 SCORE |
|---|---|---|---|---|---|
| Empirical (Exact) | 0.1305 | 0.603 / 0.603 | 0.659 / 0.744 | 0.610 / 0.667 | N/A |
| Logistic 2-Parameter | 0.1422 | 0.566 / 0.566 | 0.585 / 0.821 | 0.561 / 0.718 | 0.9751 |
| **Logistic 4-Parameter (Rec.)** | **0.1391** | **0.569 / 0.569** | **0.610 / 0.821** | **0.561 / 0.692** | **0.9884** |
| ThresholdXpert (Stochastic) | 0.1343 | 0.595 / 0.574 | 0.634 / 0.795 | 0.561 / 0.667 | N/A |

#### 2. Diagnostic Performance Curves (Training)

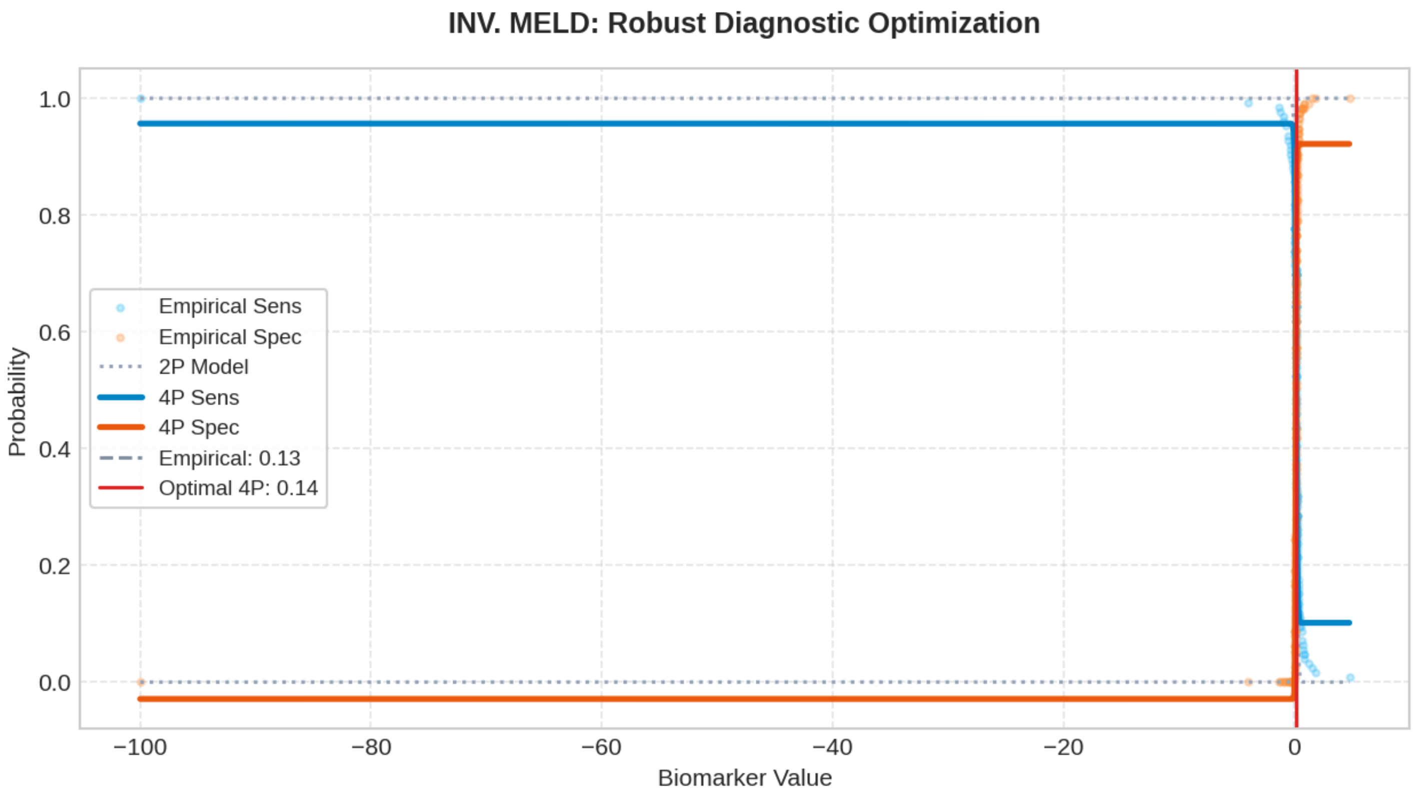

### Biomarker: AFP

Processed: 01-Apr-2026 16:51

#### 1. Optimization Results

| MODEL | CUT-OFF | TRAIN (SE/SP) | VAL (SE/SP) | TEST (SE/SP) | R2 SCORE |
|---|---|---|---|---|---|
| Empirical (Exact) | 19.9738 | 0.668 / 0.668 | 0.585 / 0.974 | 0.512 / 0.846 | N/A |
| Logistic 2-Parameter | 9.8088 | 0.789 / 0.789 | 0.634 / 0.821 | 0.610 / 0.692 | 0.8379 |
| **Logistic 4-Parameter (Rec.)** | **10.0782** | **0.804 / 0.804** | **0.634 / 0.821** | **0.610 / 0.692** | **0.9453** |
| ThresholdXpert (Stochastic) | 9.2875 | 0.746 / 0.748 | 0.634 / 0.821 | 0.634 / 0.615 | N/A |

#### 2. Diagnostic Performance Curves (Training)

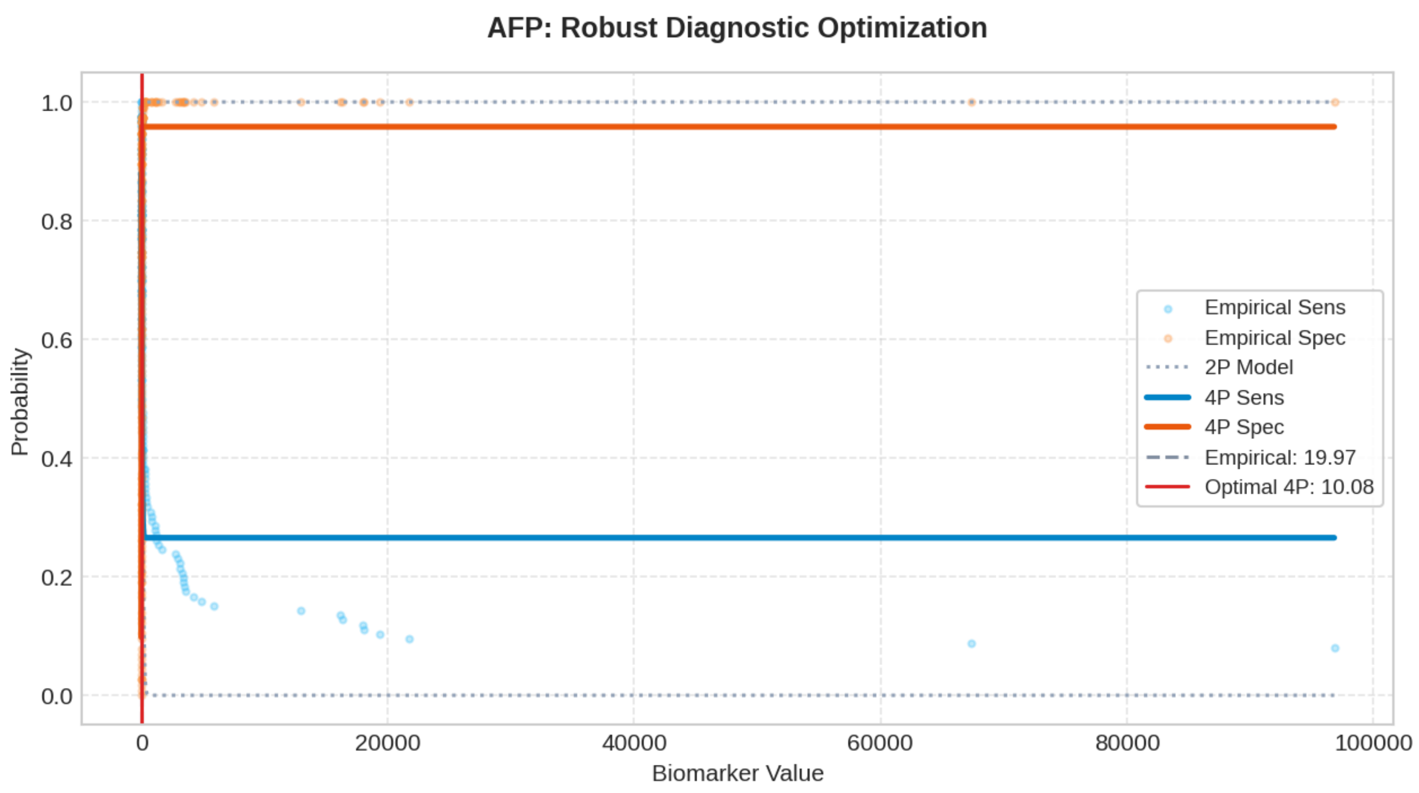

### Biomarker: PIVKA-II

Processed: 01-Apr-2026 16:53

#### 1. Optimization Results

| MODEL | CUT-OFF | TRAIN (SE/SP) | VAL (SE/SP) | TEST (SE/SP) | R2 SCORE |
|---|---|---|---|---|---|
| Empirical (Exact) | 4.1208 | 0.659 / 0.659 | 0.683 / 0.590 | 0.585 / 0.590 | N/A |
| Logistic 2-Parameter | 4.9914 | 0.741 / 0.741 | 0.659 / 0.667 | 0.561 / 0.718 | 0.8182 |
| **Logistic 4-Parameter (Rec.)** | **4.6661** | **0.716 / 0.716** | **0.659 / 0.590** | **0.561 / 0.667** | **0.9488** |
| ThresholdXpert (Stochastic) | 4.3090 | 0.659 / 0.696 | 0.683 / 0.590 | 0.585 / 0.590 | N/A |

#### 2. Diagnostic Performance Curves (Training)

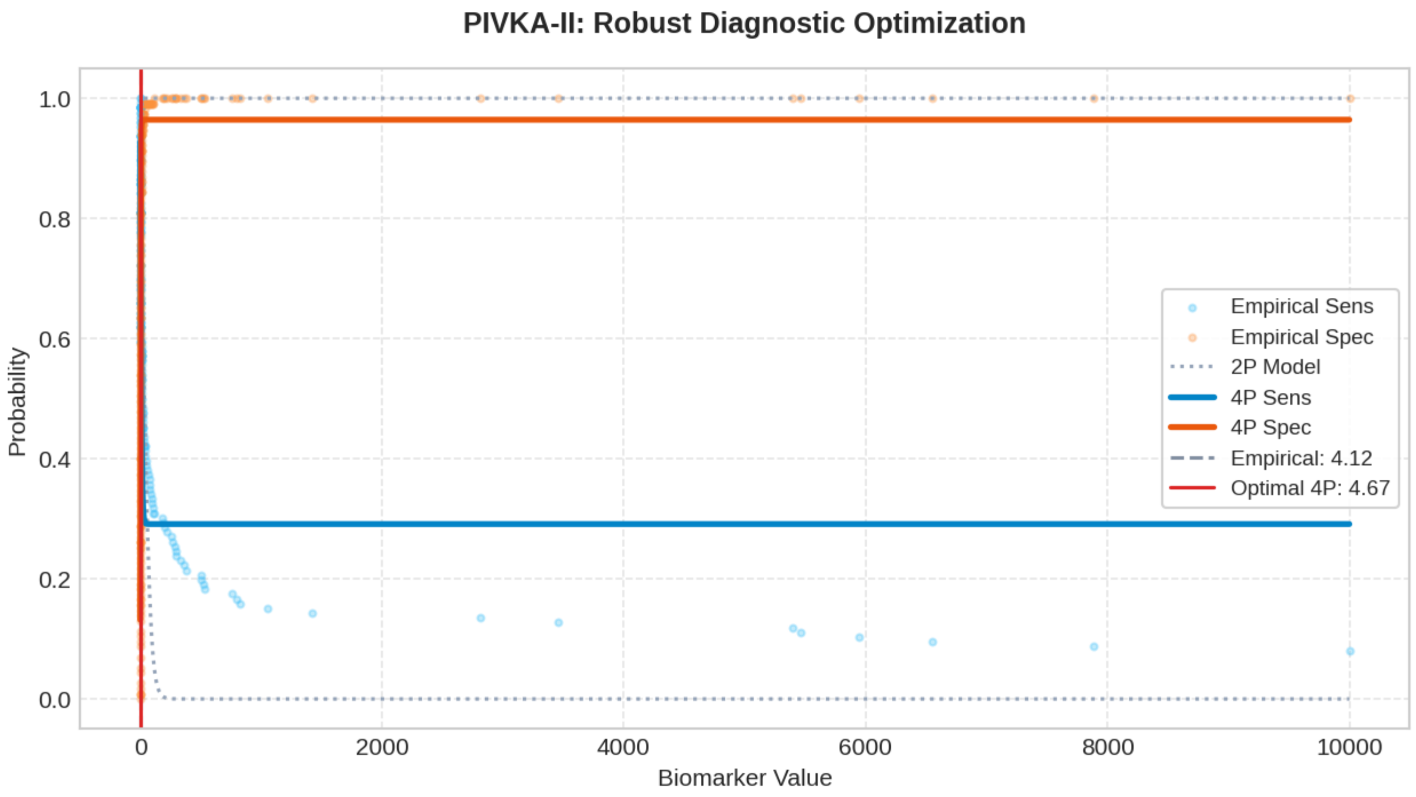

### Biomarker: OPN

Processed: 01-Apr-2026 16:56

#### 1. Optimization Results

| MODEL | CUT-OFF | TRAIN (SE/SP) | VAL (SE/SP) | TEST (SE/SP) | R2 SCORE |
|---|---|---|---|---|---|
| Empirical (Exact) | 79.1821 | 0.628 / 0.628 | 0.634 / 0.590 | 0.659 / 0.615 | N/A |
| Logistic 2-Parameter | 81.9497 | 0.666 / 0.666 | 0.561 / 0.615 | 0.610 / 0.667 | 0.9355 |
| **Logistic 4-Parameter (Rec.)** | **80.1009** | **0.646 / 0.646** | **0.634 / 0.590** | **0.634 / 0.641** | **0.9814** |
| ThresholdXpert (Stochastic) | 78.3407 | 0.635 / 0.626 | 0.634 / 0.590 | 0.659 / 0.564 | N/A |

#### 2. Diagnostic Performance Curves (Training)

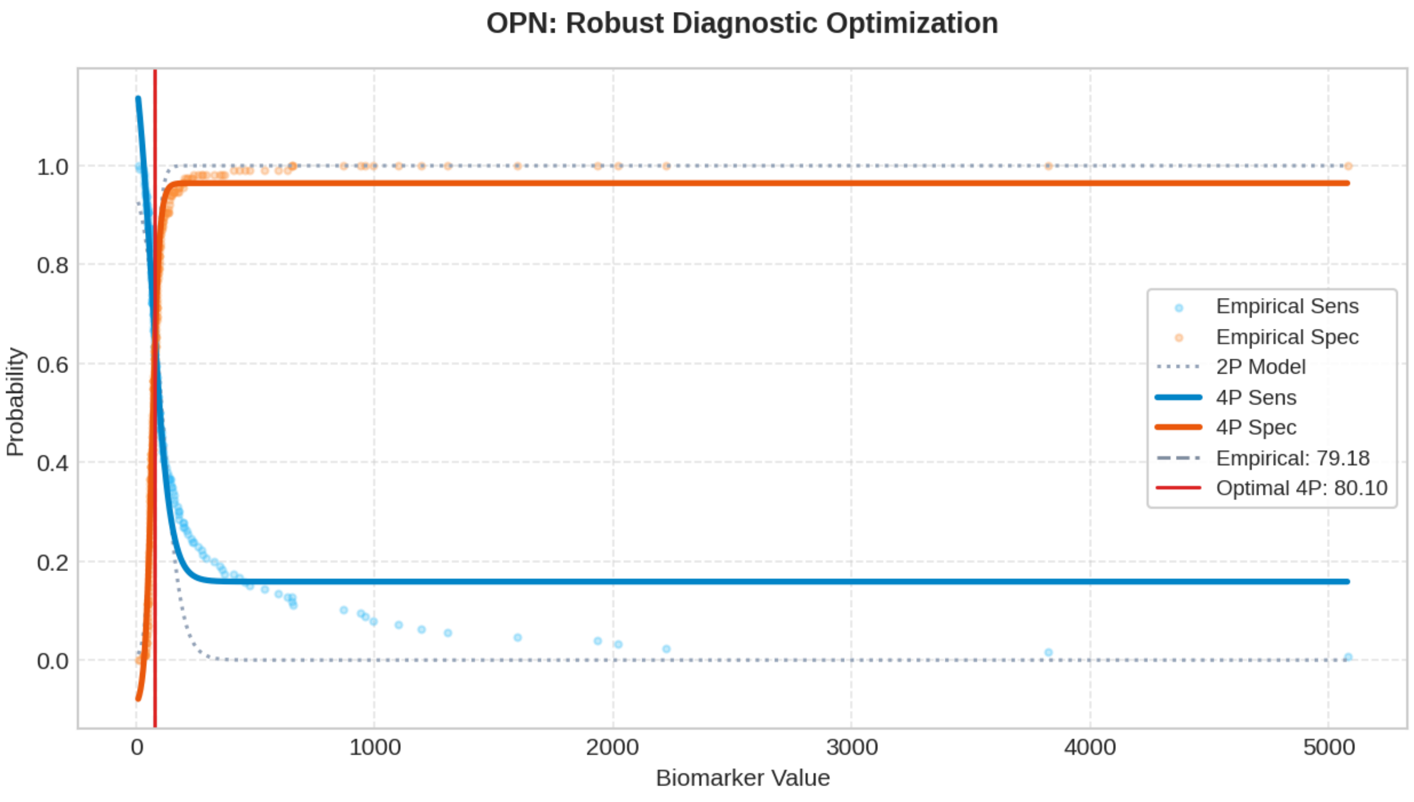

### Biomarker: DKK-1

Processed: 01-Apr-2026 16:58

#### 1. Optimization Results

| MODEL | CUT-OFF | TRAIN (SE/SP) | VAL (SE/SP) | TEST (SE/SP) | R2 SCORE |
|---|---|---|---|---|---|
| Empirical (Exact) | 364.5303 | 0.643 / 0.643 | 0.683 / 0.538 | 0.610 / 0.436 | N/A |
| Logistic 2-Parameter | 390.9040 | 0.672 / 0.672 | 0.585 / 0.538 | 0.488 / 0.564 | 0.9744 |
| **Logistic 4-Parameter (Rec.)** | **380.4725** | **0.657 / 0.657** | **0.634 / 0.538** | **0.537 / 0.487** | **0.9930** |
| ThresholdXpert (Stochastic) | 378.6795 | 0.643 / 0.678 | 0.634 / 0.538 | 0.537 / 0.487 | N/A |

#### 2. Diagnostic Performance Curves (Training)

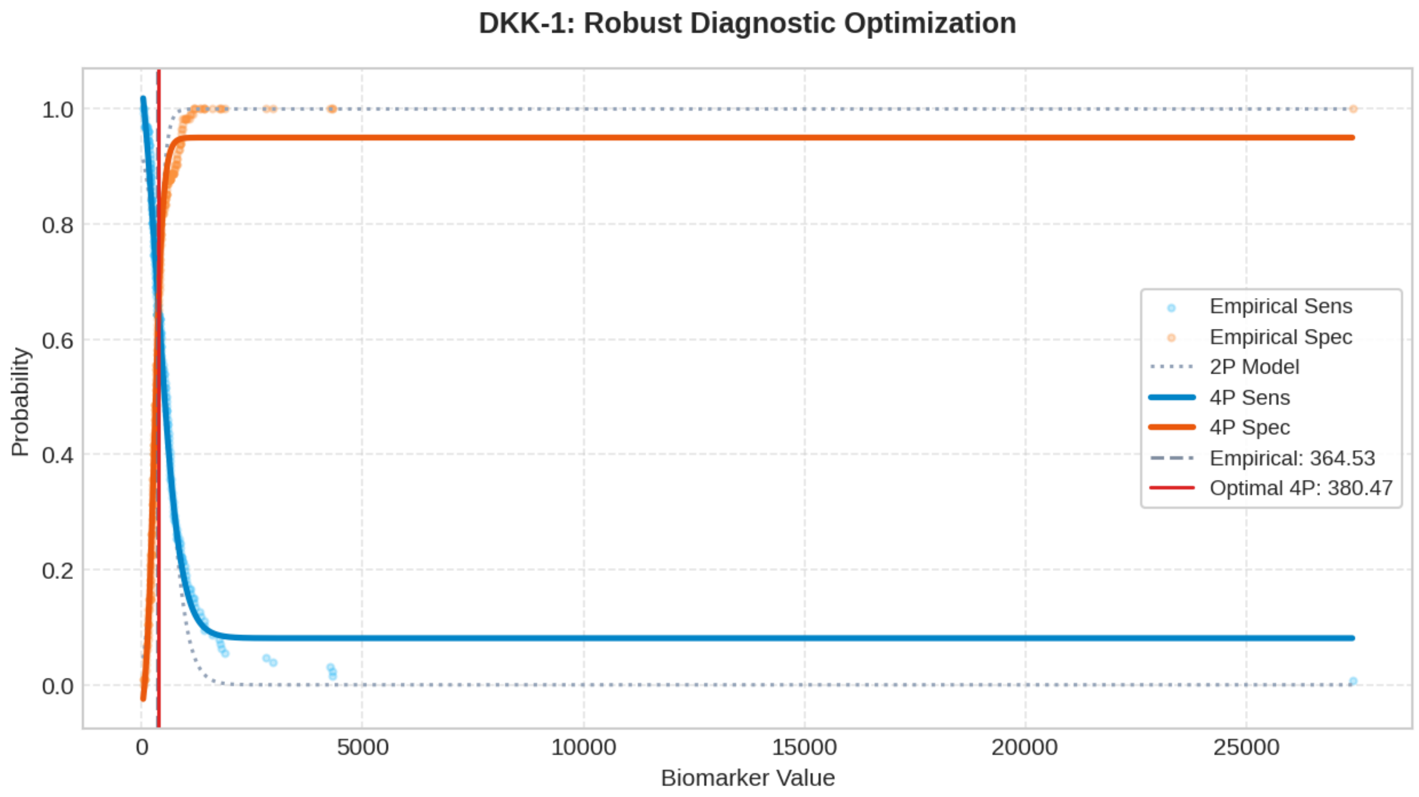

#### Top 200 Combinatorial Panels (ThresholdXpert OR-Logic)

The following multimarker panels have been optimized using high-performance vector-driven Monte Carlo simulations under a Boolean OR-logic framework. The engine employs a Max-Min Balancing logic (0.001 precision) to identify global threshold configurations that maximize the equilibrium between Sensitivity and Specificity across up to 10 million iterations. To ensure clinical robustness, results are sorted strictly by Validation Performance. (* Asterisk indicates an algorithmic threshold instability > 15%, suggesting potential data sparsity or high variance in the stochastic averaging process).

##### #1: AGE + CTP + AFP + PIVKA-II + OPN

Optimized Thresholds: AGE: 74.0994 | CTP: 10.1969 | AFP: 14.9757 | PIVKA-II: 4295.6699 | OPN: 385.8383

Train Sens: 0.754 | Train Spec: 0.757 [TRAIN SCORE: 1.510] || Val Sens: 0.634 | Val Spec: 0.897 [VAL SCORE: 1.532] || Test Sens: 0.683 | Test Spec: 0.769 [TEST SCORE: 1.452]

##### #2: AGE + CTP + MELD + AFP + PIVKA-II*

Optimized Thresholds: AGE: 73.8006 | CTP: 9.2139 | MELD: 58.5109 | AFP: 18.8019 | PIVKA-II*: 2479.0196

Train Sens: 0.746 | Train Spec: 0.783 [TRAIN SCORE: 1.529] || Val Sens: 0.634 | Val Spec: 0.897 [VAL SCORE: 1.532] || Test Sens: 0.634 | Test Spec: 0.795 [TEST SCORE: 1.429]

##### #3: AGE + MELD + AFP

Optimized Thresholds: AGE: 73.9187 | MELD: 24.2720 | AFP: 13.8839

Train Sens: 0.762 | Train Spec: 0.757 [TRAIN SCORE: 1.518] || Val Sens: 0.659 | Val Spec: 0.872 [VAL SCORE: 1.530] || Test Sens: 0.659 | Test Spec: 0.718 [TEST SCORE: 1.376]

##### #4: MELD + INV. MELD + AFP + PIVKA-II

Optimized Thresholds: MELD: 26.9425 | INV. MELD: 0.2747 | AFP: 15.2826 | PIVKA-II: 995.3673

Train Sens: 0.762 | Train Spec: 0.757 [TRAIN SCORE: 1.518] || Val Sens: 0.659 | Val Spec: 0.872 [VAL SCORE: 1.530] || Test Sens: 0.659 | Test Spec: 0.692 [TEST SCORE: 1.351]

##### #5: AGE + AFP + PIVKA-II + OPN

Optimized Thresholds: AGE: 75.3220 | AFP: 12.9880 | PIVKA-II: 6570.9701 | OPN: 381.7924

Train Sens: 0.754 | Train Spec: 0.783 [TRAIN SCORE: 1.537] || Val Sens: 0.659 | Val Spec: 0.872 [VAL SCORE: 1.530] || Test Sens: 0.659 | Test Spec: 0.744 [TEST SCORE: 1.402]

##### #6: MELD + INV. MELD + AFP + PIVKA-II + OPN

Optimized Thresholds: MELD: 26.9425 | INV. MELD: 0.2747 | AFP: 15.2826 | PIVKA-II: 995.3673 | OPN: 4740.9477

Train Sens: 0.770 | Train Spec: 0.757 [TRAIN SCORE: 1.526] || Val Sens: 0.659 | Val Spec: 0.872 [VAL SCORE: 1.530] || Test Sens: 0.659 | Test Spec: 0.692 [TEST SCORE: 1.351]

##### #7: AGE + CTP + MELD + AFP + PIVKA-II + DKK-1

Optimized Thresholds: AGE: 74.7759 | CTP: 7.0201 | MELD: 23.1125 | AFP: 18.3246 | PIVKA-II: 4819.2383 | DKK-1: 15935.5312

Train Sens: 0.754 | Train Spec: 0.757 [TRAIN SCORE: 1.510] || Val Sens: 0.659 | Val Spec: 0.872 [VAL SCORE: 1.530] || Test Sens: 0.659 | Test Spec: 0.769 [TEST SCORE: 1.428]

##### #8: MELD + INV. MELD + AFP + OPN

Optimized Thresholds: MELD: 26.9425 | INV. MELD: 0.2747 | AFP: 15.2826 | OPN: 513.6254

Train Sens: 0.778 | Train Spec: 0.748 [TRAIN SCORE: 1.526] || Val Sens: 0.659 | Val Spec: 0.872 [VAL SCORE: 1.530] || Test Sens: 0.683 | Test Spec: 0.692 [TEST SCORE: 1.375]

##### #9: AGE + CTP + MELD* + AFP + PIVKA-II + OPN*

Optimized Thresholds: AGE: 74.0966 | CTP: 7.8955 | MELD*: 41.8185 | AFP: 20.0538 | PIVKA-II: 4375.7388 | OPN*: 1578.9904

Train Sens: 0.738 | Train Spec: 0.774 [TRAIN SCORE: 1.512] || Val Sens: 0.659 | Val Spec: 0.872 [VAL SCORE: 1.530] || Test Sens: 0.683 | Test Spec: 0.769 [TEST SCORE: 1.452]

##### #10: AGE + CTP + INV. MELD + AFP + PIVKA-II + OPN + DKK-1

Optimized Thresholds: AGE: 74.0825 | CTP: 10.1758 | INV. MELD: 3.5776 | AFP: 13.5831 | PIVKA-II: 2507.1790 | OPN: 3559.2798 | DKK-1: 22275.8298

Train Sens: 0.754 | Train Spec: 0.757 [TRAIN SCORE: 1.510] || Val Sens: 0.659 | Val Spec: 0.872 [VAL SCORE: 1.530] || Test Sens: 0.683 | Test Spec: 0.769 [TEST SCORE: 1.452]

##### #11: AGE + CTP + MELD + AFP + OPN

Optimized Thresholds: AGE: 75.0253 | CTP: 7.7989 | MELD: 58.8988 | AFP: 16.4898 | OPN: 247.2538

Train Sens: 0.754 | Train Spec: 0.757 [TRAIN SCORE: 1.510] || Val Sens: 0.707 | Val Spec: 0.821 [VAL SCORE: 1.528] || Test Sens: 0.683 | Test Spec: 0.769 [TEST SCORE: 1.452]

##### #12: AGE + CTP + AFP

Optimized Thresholds: AGE: 73.4412 | CTP: 9.9219 | AFP: 17.0171

Train Sens: 0.754 | Train Spec: 0.757 [TRAIN SCORE: 1.510] || Val Sens: 0.634 | Val Spec: 0.872 [VAL SCORE: 1.506] || Test Sens: 0.634 | Test Spec: 0.795 [TEST SCORE: 1.429]

##### #13: AGE + CTP + MELD + AFP + OPN + DKK-1

Optimized Thresholds: AGE: 73.1462 | CTP: 10.6209 | MELD: 17.7940 | AFP: 18.8570 | OPN: 429.9948 | DKK-1: 12583.5852

Train Sens: 0.754 | Train Spec: 0.774 [TRAIN SCORE: 1.528] || Val Sens: 0.634 | Val Spec: 0.872 [VAL SCORE: 1.506] || Test Sens: 0.659 | Test Spec: 0.718 [TEST SCORE: 1.376]

##### #14: AGE + MELD + INV. MELD + AFP

Optimized Thresholds: AGE: 73.1989 | MELD: 30.3933 | INV. MELD: 0.9606 | AFP: 13.9309

Train Sens: 0.770 | Train Spec: 0.748 [TRAIN SCORE: 1.518] || Val Sens: 0.659 | Val Spec: 0.846 [VAL SCORE: 1.505] || Test Sens: 0.683 | Test Spec: 0.744 [TEST SCORE: 1.427]

##### #15: AGE + CTP + AFP + PIVKA-II

Optimized Thresholds: AGE: 75.4427 | CTP: 9.7344 | AFP: 13.6761 | PIVKA-II: 7740.5501

Train Sens: 0.738 | Train Spec: 0.774 [TRAIN SCORE: 1.512] || Val Sens: 0.659 | Val Spec: 0.846 [VAL SCORE: 1.505] || Test Sens: 0.659 | Test Spec: 0.769 [TEST SCORE: 1.428]

##### #16: AGE + INV. MELD + AFP

Optimized Thresholds: AGE: 73.2476 | INV. MELD: 1.6093 | AFP: 13.9546

Train Sens: 0.762 | Train Spec: 0.757 [TRAIN SCORE: 1.518] || Val Sens: 0.659 | Val Spec: 0.846 [VAL SCORE: 1.505] || Test Sens: 0.683 | Test Spec: 0.744 [TEST SCORE: 1.427]

##### #17: AGE + CTP + MELD + AFP

Optimized Thresholds: AGE: 72.2333 | CTP: 9.3369 | MELD: 24.8933 | AFP: 19.1411

Train Sens: 0.754 | Train Spec: 0.748 [TRAIN SCORE: 1.502] || Val Sens: 0.659 | Val Spec: 0.846 [VAL SCORE: 1.505] || Test Sens: 0.634 | Test Spec: 0.795 [TEST SCORE: 1.429]

##### #18: AGE + MELD + INV. MELD + AFP + PIVKA-II

Optimized Thresholds: AGE: 76.0647 | MELD: 29.2823 | INV. MELD: 1.0383 | AFP: 13.1778 | PIVKA-II: 4998.4407

Train Sens: 0.754 | Train Spec: 0.800 [TRAIN SCORE: 1.554] || Val Sens: 0.659 | Val Spec: 0.846 [VAL SCORE: 1.505] || Test Sens: 0.659 | Test Spec: 0.769 [TEST SCORE: 1.428]

##### #19: AGE + AFP + PIVKA-II

Optimized Thresholds: AGE: 73.4201 | AFP: 13.2078 | PIVKA-II: 9629.4922

Train Sens: 0.762 | Train Spec: 0.757 [TRAIN SCORE: 1.518] || Val Sens: 0.659 | Val Spec: 0.846 [VAL SCORE: 1.505] || Test Sens: 0.659 | Test Spec: 0.744 [TEST SCORE: 1.402]

##### #20: AGE + AFP

Optimized Thresholds: AGE: 73.9492 | AFP: 13.2093

Train Sens: 0.762 | Train Spec: 0.757 [TRAIN SCORE: 1.518] || Val Sens: 0.659 | Val Spec: 0.846 [VAL SCORE: 1.505] || Test Sens: 0.659 | Test Spec: 0.744 [TEST SCORE: 1.402]

##### #21: AGE + MELD + INV. MELD + AFP + DKK-1

Optimized Thresholds: AGE: 76.0647 | MELD: 29.2823 | INV. MELD: 1.0383 | AFP: 13.1778 | DKK-1: 13713.5291

Train Sens: 0.754 | Train Spec: 0.800 [TRAIN SCORE: 1.554] || Val Sens: 0.659 | Val Spec: 0.846 [VAL SCORE: 1.505] || Test Sens: 0.659 | Test Spec: 0.769 [TEST SCORE: 1.428]

##### #22: AGE + MELD + AFP + PIVKA-II + OPN

Optimized Thresholds: AGE: 71.3666 | MELD: 49.4689 | AFP: 18.8515 | PIVKA-II: 8321.1259 | OPN: 620.0442

Train Sens: 0.762 | Train Spec: 0.757 [TRAIN SCORE: 1.518] || Val Sens: 0.659 | Val Spec: 0.846 [VAL SCORE: 1.505] || Test Sens: 0.634 | Test Spec: 0.821 [TEST SCORE: 1.455]

##### #23: AGE + MELD + INV. MELD + AFP + OPN

Optimized Thresholds: AGE: 76.0647 | MELD: 29.2823 | INV. MELD: 1.0383 | AFP: 13.1778 | OPN: 2542.3994

Train Sens: 0.754 | Train Spec: 0.800 [TRAIN SCORE: 1.554] || Val Sens: 0.659 | Val Spec: 0.846 [VAL SCORE: 1.505] || Test Sens: 0.683 | Test Spec: 0.769 [TEST SCORE: 1.452]

##### #24: AGE + MELD + AFP + DKK-1

Optimized Thresholds: AGE: 76.7750 | MELD: 43.4963 | AFP: 38.1371 | DKK-1: 576.2680

Train Sens: 0.778 | Train Spec: 0.800 [TRAIN SCORE: 1.578] || Val Sens: 0.659 | Val Spec: 0.846 [VAL SCORE: 1.505] || Test Sens: 0.634 | Test Spec: 0.769 [TEST SCORE: 1.403]

##### #25: AGE + INV. MELD + AFP + PIVKA-II + OPN + DKK-1

Optimized Thresholds: AGE: 71.4451 | INV. MELD: 4.2960 | AFP: 19.8976 | PIVKA-II: 304.4115 | OPN: 1874.6784 | DKK-1: 15721.1692

Train Sens: 0.762 | Train Spec: 0.765 [TRAIN SCORE: 1.527] || Val Sens: 0.659 | Val Spec: 0.846 [VAL SCORE: 1.505] || Test Sens: 0.659 | Test Spec: 0.821 [TEST SCORE: 1.479]

###### #26: AGE + MELD + AFP + PIVKA-II

Optimized Thresholds: AGE: 71.9653 | MELD: 50.3423 | AFP: 19.2805 | PIVKA-II: 1631.4592

Train Sens: 0.754 | Train Spec: 0.765 [TRAIN SCORE: 1.519] || Val Sens: 0.659 | Val Spec: 0.846 [VAL SCORE: 1.505] || Test Sens: 0.610 | Test Spec: 0.821 [TEST SCORE: 1.430]

###### #27: AGE + MELD + AFP + PIVKA-II + OPN + DKK-1

Optimized Thresholds: AGE: 82.0877 | MELD: 61.0462 | AFP: 28.7518 | PIVKA-II: 3806.1514 | OPN: 209.9964 | DKK-1: 572.3158

Train Sens: 0.778 | Train Spec: 0.791 [TRAIN SCORE: 1.569] || Val Sens: 0.683 | Val Spec: 0.821 [VAL SCORE: 1.503] || Test Sens: 0.659 | Test Spec: 0.744 [TEST SCORE: 1.402]

###### #28: AGE + INV. MELD + AFP + OPN + DKK-1

Optimized Thresholds: AGE: 77.7087 | INV. MELD: 1.8855 | AFP: 11.9935 | OPN: 182.4645 | DKK-1: 22891.5806

Train Sens: 0.762 | Train Spec: 0.774 [TRAIN SCORE: 1.536] || Val Sens: 0.707 | Val Spec: 0.795 [VAL SCORE: 1.502] || Test Sens: 0.707 | Test Spec: 0.718 [TEST SCORE: 1.425]

###### #29: AGE + CTP + AFP + OPN + DKK-1

Optimized Thresholds: AGE: 77.7087 | CTP: 10.8353 | AFP: 11.9935 | OPN: 182.4645 | DKK-1: 22891.5806

Train Sens: 0.754 | Train Spec: 0.765 [TRAIN SCORE: 1.519] || Val Sens: 0.707 | Val Spec: 0.795 [VAL SCORE: 1.502] || Test Sens: 0.683 | Test Spec: 0.718 [TEST SCORE: 1.401]

###### #30: MELD + INV. MELD + AFP + DKK-1

Optimized Thresholds: MELD: 63.6104 | INV. MELD: 4.1497 | AFP: 25.5772 | DKK-1: 603.8543

Train Sens: 0.762 | Train Spec: 0.800 [TRAIN SCORE: 1.562] || Val Sens: 0.610 | Val Spec: 0.872 [VAL SCORE: 1.482] || Test Sens: 0.634 | Test Spec: 0.821 [TEST SCORE: 1.455]

###### #31: AGE + MELD + INV. MELD + AFP + PIVKA-II + OPN + DKK-1

Optimized Thresholds: AGE: 80.5398 | MELD: 57.0774 | INV. MELD: 0.8137 | AFP: 24.5098 | PIVKA-II: 3680.3343 | OPN: 550.3010 | DKK-1: 609.5601

Train Sens: 0.778 | Train Spec: 0.757 [TRAIN SCORE: 1.534] || Val Sens: 0.634 | Val Spec: 0.846 [VAL SCORE: 1.480] || Test Sens: 0.683 | Test Spec: 0.795 [TEST SCORE: 1.478]

###### #32: AGE + INV. MELD + AFP + PIVKA-II + OPN

Optimized Thresholds: AGE: 81.3537 | INV. MELD: 0.2118 | AFP: 33.8983 | PIVKA-II: 7808.7282 | OPN: 398.9523

Train Sens: 0.778 | Train Spec: 0.757 [TRAIN SCORE: 1.534] || Val Sens: 0.659 | Val Spec: 0.821 [VAL SCORE: 1.479] || Test Sens: 0.659 | Test Spec: 0.795 [TEST SCORE: 1.453]

###### #33: CTP + MELD + AFP + PIVKA-II + OPN + DKK-1

Optimized Thresholds: CTP: 10.7226 | MELD: 61.0462 | AFP: 28.7518 | PIVKA-II: 3806.1514 | OPN: 209.9964 | DKK-1: 572.3158

Train Sens: 0.778 | Train Spec: 0.774 [TRAIN SCORE: 1.552] || Val Sens: 0.659 | Val Spec: 0.821 [VAL SCORE: 1.479] || Test Sens: 0.683 | Test Spec: 0.744 [TEST SCORE: 1.427]

###### #34: AGE + MELD + AFP + OPN

Optimized Thresholds: AGE: 77.6607 | MELD: 21.2187 | AFP: 17.7500 | OPN: 139.0582

Train Sens: 0.762 | Train Spec: 0.791 [TRAIN SCORE: 1.553] || Val Sens: 0.683 | Val Spec: 0.795 [VAL SCORE: 1.478] || Test Sens: 0.659 | Test Spec: 0.769 [TEST SCORE: 1.428]

###### #35: AGE + CTP + MELD + INV. MELD + AFP

Optimized Thresholds: AGE: 77.8657 | CTP: 7.0508 | MELD: 17.5404 | INV. MELD: 0.6293 | AFP: 17.1976

Train Sens: 0.762 | Train Spec: 0.748 [TRAIN SCORE: 1.510] || Val Sens: 0.683 | Val Spec: 0.795 [VAL SCORE: 1.478] || Test Sens: 0.683 | Test Spec: 0.744 [TEST SCORE: 1.427]

###### #36: AGE + CTP + AFP + OPN

Optimized Thresholds: AGE: 77.9659 | CTP: 8.1254 | AFP: 16.5941 | OPN: 167.2147

Train Sens: 0.762 | Train Spec: 0.765 [TRAIN SCORE: 1.527] || Val Sens: 0.683 | Val Spec: 0.795 [VAL SCORE: 1.478] || Test Sens: 0.634 | Test Spec: 0.795 [TEST SCORE: 1.429]

###### #37: AGE + CTP + MELD + INV. MELD + AFP + DKK-1

Optimized Thresholds: AGE: 77.8657 | CTP: 7.0508 | MELD: 17.5404 | INV. MELD: 0.6293 | AFP: 17.1976 | DKK-1: 16197.2365

Train Sens: 0.762 | Train Spec: 0.748 [TRAIN SCORE: 1.510] || Val Sens: 0.683 | Val Spec: 0.795 [VAL SCORE: 1.478] || Test Sens: 0.683 | Test Spec: 0.744 [TEST SCORE: 1.427]

###### #38: AGE + CTP + MELD + INV. MELD + AFP + PIVKA-II

Optimized Thresholds: AGE: 77.8657 | CTP: 7.0508 | MELD: 17.5404 | INV. MELD: 0.6293 | AFP: 17.1976 | PIVKA-II: 5906.0943

Train Sens: 0.762 | Train Spec: 0.748 [TRAIN SCORE: 1.510] || Val Sens: 0.683 | Val Spec: 0.795 [VAL SCORE: 1.478] || Test Sens: 0.683 | Test Spec: 0.744 [TEST SCORE: 1.427]

###### #39: AGE + CTP + MELD + INV. MELD + AFP + OPN

Optimized Thresholds: AGE: 77.8657 | CTP: 7.0508 | MELD: 17.5404 | INV. MELD: 0.6293 | AFP: 17.1976 | OPN: 3002.4019

Train Sens: 0.762 | Train Spec: 0.748 [TRAIN SCORE: 1.510] || Val Sens: 0.683 | Val Spec: 0.795 [VAL SCORE: 1.478] || Test Sens: 0.707 | Test Spec: 0.744 [TEST SCORE: 1.451]

###### #40: AGE + CTP + AFP + PIVKA-II + OPN + DKK-1

Optimized Thresholds: AGE: 82.0877 | CTP: 10.6809 | AFP: 28.7518 | PIVKA-II: 3806.1514 | OPN: 209.9964 | DKK-1: 572.3158

Train Sens: 0.778 | Train Spec: 0.774 [TRAIN SCORE: 1.552] || Val Sens: 0.683 | Val Spec: 0.795 [VAL SCORE: 1.478] || Test Sens: 0.683 | Test Spec: 0.744 [TEST SCORE: 1.427]

###### #41: INV. MELD + AFP + OPN

Optimized Thresholds: INV. MELD: 0.2103 | AFP: 32.0407 | OPN: 2457.0083

Train Sens: 0.770 | Train Spec: 0.765 [TRAIN SCORE: 1.535] || Val Sens: 0.610 | Val Spec: 0.846 [VAL SCORE: 1.456] || Test Sens: 0.659 | Test Spec: 0.795 [TEST SCORE: 1.453]

###### #42: INV. MELD + AFP + PIVKA-II

Optimized Thresholds: INV. MELD: 0.2103 | AFP: 32.0407 | PIVKA-II: 4829.9515

Train Sens: 0.762 | Train Spec: 0.765 [TRAIN SCORE: 1.527] || Val Sens: 0.610 | Val Spec: 0.846 [VAL SCORE: 1.456] || Test Sens: 0.634 | Test Spec: 0.795 [TEST SCORE: 1.429]

###### #43: MELD + INV. MELD + AFP

Optimized Thresholds: MELD: 25.6760 | INV. MELD: 0.2088 | AFP: 40.2954

Train Sens: 0.754 | Train Spec: 0.774 [TRAIN SCORE: 1.528] || Val Sens: 0.610 | Val Spec: 0.846 [VAL SCORE: 1.456] || Test Sens: 0.634 | Test Spec: 0.795 [TEST SCORE: 1.429]

##### #44: AGE + INV. MELD + AFP + DKK-1

Optimized Thresholds: AGE: 83.8389 | INV. MELD: 4.1497 | AFP: 25.5772 | DKK-1: 603.8543

Train Sens: 0.762 | Train Spec: 0.800 [TRAIN SCORE: 1.562] || Val Sens: 0.610 | Val Spec: 0.846 [VAL SCORE: 1.456] || Test Sens: 0.634 | Test Spec: 0.821 [TEST SCORE: 1.455]

##### #45: MELD + INV. MELD + AFP + OPN + DKK-1

Optimized Thresholds: MELD: 60.7004 | INV. MELD: 0.2118 | AFP: 33.8983 | OPN: 3966.6645 | DKK-1: 2140.3410

Train Sens: 0.762 | Train Spec: 0.757 [TRAIN SCORE: 1.518] || Val Sens: 0.610 | Val Spec: 0.846 [VAL SCORE: 1.456] || Test Sens: 0.634 | Test Spec: 0.795 [TEST SCORE: 1.429]

##### #46: CTP + INV. MELD + AFP + DKK-1

Optimized Thresholds: CTP: 10.8894 | INV. MELD: 4.1497 | AFP: 25.5772 | DKK-1: 603.8543

Train Sens: 0.762 | Train Spec: 0.783 [TRAIN SCORE: 1.545] || Val Sens: 0.610 | Val Spec: 0.846 [VAL SCORE: 1.456] || Test Sens: 0.659 | Test Spec: 0.821 [TEST SCORE: 1.479]

##### #47: MELD + INV. MELD + PIVKA-II

Optimized Thresholds: MELD: 62.3317 | INV. MELD: 0.2497 | PIVKA-II: 7.6529

Train Sens: 0.722 | Train Spec: 0.730 [TRAIN SCORE: 1.453] || Val Sens: 0.634 | Val Spec: 0.821 [VAL SCORE: 1.455] || Test Sens: 0.585 | Test Spec: 0.744 [TEST SCORE: 1.329]

##### #48: CTP + INV. MELD + AFP + OPN

Optimized Thresholds: CTP: 9.7503 | INV. MELD: 0.6706 | AFP: 14.4945 | OPN: 264.5499

Train Sens: 0.746 | Train Spec: 0.791 [TRAIN SCORE: 1.537] || Val Sens: 0.634 | Val Spec: 0.821 [VAL SCORE: 1.455] || Test Sens: 0.659 | Test Spec: 0.769 [TEST SCORE: 1.428]

##### #49: MELD* + AFP + PIVKA-II*

Optimized Thresholds: MELD*: 48.0924 | AFP: 9.3314 | PIVKA-II*: 4625.5544

Train Sens: 0.746 | Train Spec: 0.748 [TRAIN SCORE: 1.494] || Val Sens: 0.634 | Val Spec: 0.821 [VAL SCORE: 1.455] || Test Sens: 0.610 | Test Spec: 0.641 [TEST SCORE: 1.251]

##### #50: AFP + PIVKA-II*

Optimized Thresholds: AFP: 9.2837 | PIVKA-II*: 4510.6512

Train Sens: 0.746 | Train Spec: 0.748 [TRAIN SCORE: 1.494] || Val Sens: 0.634 | Val Spec: 0.821 [VAL SCORE: 1.455] || Test Sens: 0.634 | Test Spec: 0.615 [TEST SCORE: 1.250]

##### #51: CTP + INV. MELD + PIVKA-II

Optimized Thresholds: CTP: 10.7854 | INV. MELD: 0.2497 | PIVKA-II: 7.6529

Train Sens: 0.722 | Train Spec: 0.722 [TRAIN SCORE: 1.444] || Val Sens: 0.634 | Val Spec: 0.821 [VAL SCORE: 1.455] || Test Sens: 0.610 | Test Spec: 0.744 [TEST SCORE: 1.353]

##### #52: AGE + INV. MELD + AFP + PIVKA-II + DKK-1

Optimized Thresholds: AGE: 81.3537 | INV. MELD: 0.2118 | AFP: 33.8983 | PIVKA-II: 7808.7282 | DKK-1: 2140.3410

Train Sens: 0.762 | Train Spec: 0.765 [TRAIN SCORE: 1.527] || Val Sens: 0.634 | Val Spec: 0.821 [VAL SCORE: 1.455] || Test Sens: 0.634 | Test Spec: 0.795 [TEST SCORE: 1.429]

##### #53: CTP + MELD + AFP + DKK-1

Optimized Thresholds: CTP: 9.9734 | MELD: 49.0333 | AFP: 26.1236 | DKK-1: 572.1571

Train Sens: 0.778 | Train Spec: 0.774 [TRAIN SCORE: 1.552] || Val Sens: 0.634 | Val Spec: 0.821 [VAL SCORE: 1.455] || Test Sens: 0.634 | Test Spec: 0.744 [TEST SCORE: 1.378]

##### #54: CTP + INV. MELD + AFP + PIVKA-II + OPN

Optimized Thresholds: CTP: 9.2426 | INV. MELD: 0.4428 | AFP: 14.2787 | PIVKA-II: 2653.1585 | OPN: 3858.5341

Train Sens: 0.754 | Train Spec: 0.783 [TRAIN SCORE: 1.537] || Val Sens: 0.634 | Val Spec: 0.821 [VAL SCORE: 1.455] || Test Sens: 0.634 | Test Spec: 0.744 [TEST SCORE: 1.378]

##### #55: CTP + INV. MELD + AFP + OPN + DKK-1

Optimized Thresholds: CTP: 9.2426 | INV. MELD: 0.4428 | AFP: 14.2787 | OPN: 1353.8007 | DKK-1: 20819.7811

Train Sens: 0.754 | Train Spec: 0.783 [TRAIN SCORE: 1.537] || Val Sens: 0.634 | Val Spec: 0.821 [VAL SCORE: 1.455] || Test Sens: 0.659 | Test Spec: 0.744 [TEST SCORE: 1.402]

##### #56: AGE + CTP + MELD + INV. MELD + AFP + OPN + DKK-1

Optimized Thresholds: AGE: 83.6307 | CTP: 8.4812 | MELD: 44.3366 | INV. MELD: 3.9401 | AFP: 10.8458 | OPN: 2296.6784 | DKK-1: 24617.9898

Train Sens: 0.746 | Train Spec: 0.739 [TRAIN SCORE: 1.485] || Val Sens: 0.659 | Val Spec: 0.795 [VAL SCORE: 1.453] || Test Sens: 0.659 | Test Spec: 0.667 [TEST SCORE: 1.325]

##### #57: AGE + CTP + MELD + INV. MELD + AFP + PIVKA-II + DKK-1

Optimized Thresholds: AGE: 83.6307 | CTP: 8.4812 | MELD: 44.3366 | INV. MELD: 3.9401 | AFP: 10.8458 | PIVKA-II: 4513.5966 | DKK-1: 24617.9898

Train Sens: 0.746 | Train Spec: 0.739 [TRAIN SCORE: 1.485] || Val Sens: 0.659 | Val Spec: 0.795 [VAL SCORE: 1.453] || Test Sens: 0.634 | Test Spec: 0.667 [TEST SCORE: 1.301]

##### #58: AGE + CTP + AFP + DKK-1

Optimized Thresholds: AGE: 76.7750 | CTP: 9.2535 | AFP: 38.1371 | DKK-1: 576.2680

Train Sens: 0.786 | Train Spec: 0.774 [TRAIN SCORE: 1.560] || Val Sens: 0.659 | Val Spec: 0.795 [VAL SCORE: 1.453] || Test Sens: 0.659 | Test Spec: 0.744 [TEST SCORE: 1.402]

##### #59: CTP + MELD + INV. MELD + AFP + PIVKA-II + OPN + DKK-1

Optimized Thresholds: CTP: 9.9302 | MELD: 39.4786 | INV. MELD: 0.4911 | AFP: 12.0660 | PIVKA-II: 8406.4128 | OPN: 2783.8294 | DKK-1: 20779.2211

Train Sens: 0.762 | Train Spec: 0.765 [TRAIN SCORE: 1.527] || Val Sens: 0.659 | Val Spec: 0.795 [VAL SCORE: 1.453] || Test Sens: 0.659 | Test Spec: 0.744 [TEST SCORE: 1.402]

##### #60: AGE + CTP + MELD + INV. MELD + AFP + PIVKA-II + OPN

Optimized Thresholds: AGE: 83.6307 | CTP: 8.4812 | MELD: 44.3366 | INV. MELD: 3.9401 | AFP: 10.8458 | PIVKA-II: 4513.5966 | OPN: 4561.9928

Train Sens: 0.746 | Train Spec: 0.739 [TRAIN SCORE: 1.485] || Val Sens: 0.659 | Val Spec: 0.795 [VAL SCORE: 1.453] || Test Sens: 0.634 | Test Spec: 0.667 [TEST SCORE: 1.301]

##### #61: AGE + INV. MELD + PIVKA-II

Optimized Thresholds: AGE: 82.7469 | INV. MELD: 0.2497 | PIVKA-II: 7.6529

Train Sens: 0.730 | Train Spec: 0.739 [TRAIN SCORE: 1.469] || Val Sens: 0.659 | Val Spec: 0.795 [VAL SCORE: 1.453] || Test Sens: 0.585 | Test Spec: 0.744 [TEST SCORE: 1.329]

**#62: AGE + CTP + INV. MELD + AFP**

Optimized Thresholds: AGE: 81.0694 | CTP: 9.0805 | INV. MELD: 1.2580 | AFP: 12.8832

Train Sens: 0.738 | Train Spec: 0.791 [TRAIN SCORE: 1.529] || Val Sens: 0.659 | Val Spec: 0.795 [VAL SCORE: 1.453] || Test Sens: 0.634 | Test Spec: 0.744 [TEST SCORE: 1.378]

**#63: AGE + CTP + MELD + AFP + DKK-1**

Optimized Thresholds: AGE: 81.5973 | CTP: 9.8125 | MELD: 45.1890 | AFP: 37.4491 | DKK-1: 576.4573

Train Sens: 0.762 | Train Spec: 0.800 [TRAIN SCORE: 1.562] || Val Sens: 0.659 | Val Spec: 0.795 [VAL SCORE: 1.453] || Test Sens: 0.634 | Test Spec: 0.744 [TEST SCORE: 1.378]

**#64: MELD + AFP + DKK-1**

Optimized Thresholds: MELD: 57.4688 | AFP: 39.1135 | DKK-1: 489.2357

Train Sens: 0.810 | Train Spec: 0.791 [TRAIN SCORE: 1.601] || Val Sens: 0.732 | Val Spec: 0.718 [VAL SCORE: 1.450] || Test Sens: 0.610 | Test Spec: 0.692 [TEST SCORE: 1.302]

**#65: INV. MELD + AFP + DKK-1**

Optimized Thresholds: INV. MELD: 1.6549 | AFP: 41.8830 | DKK-1: 486.5178

Train Sens: 0.810 | Train Spec: 0.791 [TRAIN SCORE: 1.601] || Val Sens: 0.732 | Val Spec: 0.718 [VAL SCORE: 1.450] || Test Sens: 0.610 | Test Spec: 0.692 [TEST SCORE: 1.302]

**#66: INV. MELD + AFP + PIVKA-II + OPN**

Optimized Thresholds: INV. MELD: 0.2348 | AFP: 38.2365 | PIVKA-II: 3528.8033 | OPN: 1913.4805

Train Sens: 0.770 | Train Spec: 0.809 [TRAIN SCORE: 1.579] || Val Sens: 0.561 | Val Spec: 0.872 [VAL SCORE: 1.433] || Test Sens: 0.634 | Test Spec: 0.821 [TEST SCORE: 1.455]

**#67: INV. MELD + AFP**

Optimized Thresholds: INV. MELD: 0.2348 | AFP: 38.2365

Train Sens: 0.754 | Train Spec: 0.809 [TRAIN SCORE: 1.563] || Val Sens: 0.561 | Val Spec: 0.872 [VAL SCORE: 1.433] || Test Sens: 0.610 | Test Spec: 0.821 [TEST SCORE: 1.430]

**#68: AGE + MELD + INV. MELD + AFP + PIVKA-II + OPN**

Optimized Thresholds: AGE: 80.7117 | MELD: 39.6007 | INV. MELD: 0.2348 | AFP: 38.2365 | PIVKA-II: 3528.8033 | OPN: 1913.4805

Train Sens: 0.770 | Train Spec: 0.791 [TRAIN SCORE: 1.561] || Val Sens: 0.585 | Val Spec: 0.846 [VAL SCORE: 1.432] || Test Sens: 0.659 | Test Spec: 0.821 [TEST SCORE: 1.479]

**#69: AGE + MELD + INV. MELD + AFP + PIVKA-II + DKK-1**

Optimized Thresholds: AGE: 80.7117 | MELD: 39.6007 | INV. MELD: 0.2348 | AFP: 38.2365 | PIVKA-II: 3528.8033 | DKK-1: 10317.7857

Train Sens: 0.762 | Train Spec: 0.791 [TRAIN SCORE: 1.553] || Val Sens: 0.585 | Val Spec: 0.846 [VAL SCORE: 1.432] || Test Sens: 0.634 | Test Spec: 0.821 [TEST SCORE: 1.455]

##### #70: AGE + MELD + INV. MELD + AFP + OPN + DKK-1

Optimized Thresholds: AGE: 80.7117 | MELD: 39.6007 | INV. MELD: 0.2348 | AFP: 38.2365 | OPN: 1797.5811 | DKK-1: 10317.7857

Train Sens: 0.770 | Train Spec: 0.791 [TRAIN SCORE: 1.561] || Val Sens: 0.585 | Val Spec: 0.846 [VAL SCORE: 1.432] || Test Sens: 0.659 | Test Spec: 0.821 [TEST SCORE: 1.479]

##### #71: AGE + INV. MELD + PIVKA-II + DKK-1

Optimized Thresholds: AGE: 79.1672 | INV. MELD: 3.6924 | PIVKA-II: 14.9671 | DKK-1: 549.8327

Train Sens: 0.738 | Train Spec: 0.800 [TRAIN SCORE: 1.538] || Val Sens: 0.610 | Val Spec: 0.821 [VAL SCORE: 1.430] || Test Sens: 0.659 | Test Spec: 0.795 [TEST SCORE: 1.453]

##### #72: CTP + MELD* + AFP + PIVKA-II*

Optimized Thresholds: CTP: 10.5168 | MELD*: 39.8394 | AFP: 9.3601 | PIVKA-II*: 6503.9775

Train Sens: 0.746 | Train Spec: 0.739 [TRAIN SCORE: 1.485] || Val Sens: 0.634 | Val Spec: 0.795 [VAL SCORE: 1.429] || Test Sens: 0.634 | Test Spec: 0.641 [TEST SCORE: 1.275]

##### #73: CTP + MELD + AFP

Optimized Thresholds: CTP: 10.6591 | MELD: 37.1731 | AFP: 9.2962

Train Sens: 0.746 | Train Spec: 0.739 [TRAIN SCORE: 1.485] || Val Sens: 0.634 | Val Spec: 0.795 [VAL SCORE: 1.429] || Test Sens: 0.659 | Test Spec: 0.615 [TEST SCORE: 1.274]

##### #74: CTP + AFP

Optimized Thresholds: CTP: 10.6575 | AFP: 9.3067

Train Sens: 0.746 | Train Spec: 0.739 [TRAIN SCORE: 1.485] || Val Sens: 0.634 | Val Spec: 0.795 [VAL SCORE: 1.429] || Test Sens: 0.659 | Test Spec: 0.615 [TEST SCORE: 1.274]

##### #75: CTP + AFP + PIVKA-II*

Optimized Thresholds: CTP: 10.6662 | AFP: 9.2446 | PIVKA-II*: 4868.7780

Train Sens: 0.746 | Train Spec: 0.739 [TRAIN SCORE: 1.485] || Val Sens: 0.634 | Val Spec: 0.795 [VAL SCORE: 1.429] || Test Sens: 0.659 | Test Spec: 0.615 [TEST SCORE: 1.274]

##### #76: INV. MELD + PIVKA-II

Optimized Thresholds: INV. MELD: 0.2098 | PIVKA-II: 8.7408

Train Sens: 0.738 | Train Spec: 0.730 [TRAIN SCORE: 1.469] || Val Sens: 0.634 | Val Spec: 0.795 [VAL SCORE: 1.429] || Test Sens: 0.659 | Test Spec: 0.692 [TEST SCORE: 1.351]

##### #77: CTP + INV. MELD + AFP

Optimized Thresholds: CTP: 8.8977 | INV. MELD: 0.5283 | AFP: 13.9239

Train Sens: 0.754 | Train Spec: 0.774 [TRAIN SCORE: 1.528] || Val Sens: 0.634 | Val Spec: 0.795 [VAL SCORE: 1.429] || Test Sens: 0.634 | Test Spec: 0.744 [TEST SCORE: 1.378]

##### #78: AGE + AFP + PIVKA-II + OPN + DKK-1

Optimized Thresholds: AGE: 81.5692 | AFP: 28.6681 | PIVKA-II: 4855.4861 | OPN: 3072.6226 | DKK-1: 542.5534

Train Sens: 0.786 | Train Spec: 0.791 [TRAIN SCORE: 1.577] || Val Sens: 0.659 | Val Spec: 0.769 [VAL SCORE: 1.428] || Test Sens: 0.659 | Test Spec: 0.744 [TEST SCORE: 1.402]

###### #79: MELD + AFP + PIVKA-II + DKK-1

Optimized Thresholds: MELD: 49.2204 | AFP: 35.9605 | PIVKA-II: 3267.9285 | DKK-1: 517.6719

Train Sens: 0.802 | Train Spec: 0.800 [TRAIN SCORE: 1.602] || Val Sens: 0.659 | Val Spec: 0.769 [VAL SCORE: 1.428] || Test Sens: 0.634 | Test Spec: 0.744 [TEST SCORE: 1.378]

###### #80: AGE + CTP + INV. MELD + AFP + DKK-1

Optimized Thresholds: AGE: 76.0647 | CTP: 8.0974 | INV. MELD: 1.0383 | AFP: 13.1778 | DKK-1: 13713.5291

Train Sens: 0.770 | Train Spec: 0.765 [TRAIN SCORE: 1.535] || Val Sens: 0.659 | Val Spec: 0.769 [VAL SCORE: 1.428] || Test Sens: 0.683 | Test Spec: 0.744 [TEST SCORE: 1.427]

###### #81: MELD + AFP + OPN + DKK-1

Optimized Thresholds: MELD: 49.2204 | AFP: 35.9605 | OPN: 1665.3687 | DKK-1: 517.6719

Train Sens: 0.810 | Train Spec: 0.800 [TRAIN SCORE: 1.610] || Val Sens: 0.659 | Val Spec: 0.769 [VAL SCORE: 1.428] || Test Sens: 0.634 | Test Spec: 0.744 [TEST SCORE: 1.378]

###### #82: AGE + CTP + INV. MELD + AFP + OPN

Optimized Thresholds: AGE: 76.0647 | CTP: 8.0974 | INV. MELD: 1.0383 | AFP: 13.1778 | OPN: 2542.3994

Train Sens: 0.770 | Train Spec: 0.765 [TRAIN SCORE: 1.535] || Val Sens: 0.659 | Val Spec: 0.769 [VAL SCORE: 1.428] || Test Sens: 0.707 | Test Spec: 0.744 [TEST SCORE: 1.451]

###### #83: AGE + CTP + INV. MELD + AFP + PIVKA-II

Optimized Thresholds: AGE: 76.0647 | CTP: 8.0974 | INV. MELD: 1.0383 | AFP: 13.1778 | PIVKA-II: 4998.4407

Train Sens: 0.770 | Train Spec: 0.765 [TRAIN SCORE: 1.535] || Val Sens: 0.659 | Val Spec: 0.769 [VAL SCORE: 1.428] || Test Sens: 0.683 | Test Spec: 0.744 [TEST SCORE: 1.427]

###### #84: AGE + AFP + OPN

Optimized Thresholds: AGE: 75.6419 | AFP: 13.9824 | OPN: 142.7693

Train Sens: 0.778 | Train Spec: 0.757 [TRAIN SCORE: 1.534] || Val Sens: 0.683 | Val Spec: 0.744 [VAL SCORE: 1.427] || Test Sens: 0.683 | Test Spec: 0.744 [TEST SCORE: 1.427]

###### #85: MELD + PIVKA-II + OPN

Optimized Thresholds: MELD: 26.5590 | PIVKA-II: 7.4956 | OPN: 104.1741

Train Sens: 0.714 | Train Spec: 0.730 [TRAIN SCORE: 1.445] || Val Sens: 0.707 | Val Spec: 0.718 [VAL SCORE: 1.425] || Test Sens: 0.610 | Test Spec: 0.615 [TEST SCORE: 1.225]

###### #86: PIVKA-II + OPN

Optimized Thresholds: PIVKA-II: 7.6345 | OPN: 104.1131

Train Sens: 0.714 | Train Spec: 0.739 [TRAIN SCORE: 1.453] || Val Sens: 0.707 | Val Spec: 0.718 [VAL SCORE: 1.425] || Test Sens: 0.610 | Test Spec: 0.615 [TEST SCORE: 1.225]

###### #87: CTP + AFP + DKK-1

Optimized Thresholds: CTP: 10.3899 | AFP: 39.1135 | DKK-1: 489.2357

Train Sens: 0.810 | Train Spec: 0.774 [TRAIN SCORE: 1.583] || Val Sens: 0.732 | Val Spec: 0.692 [VAL SCORE: 1.424] || Test Sens: 0.634 | Test Spec: 0.692 [TEST SCORE: 1.326]

###### #88: AGE + AFP + DKK-1

Optimized Thresholds: AGE: 83.1315 | AFP: 41.8830 | DKK-1: 486.5178

Train Sens: 0.802 | Train Spec: 0.791 [TRAIN SCORE: 1.593] || Val Sens: 0.732 | Val Spec: 0.692 [VAL SCORE: 1.424] || Test Sens: 0.585 | Test Spec: 0.692 [TEST SCORE: 1.278]

###### #89: MELD + INV. MELD + AFP + PIVKA-II + OPN + DKK-1

Optimized Thresholds: MELD: 29.0977 | INV. MELD: 1.6294 | AFP: 42.0989 | PIVKA-II: 9071.5310 | OPN: 275.1615 | DKK-1: 495.1077

Train Sens: 0.817 | Train Spec: 0.774 [TRAIN SCORE: 1.591] || Val Sens: 0.732 | Val Spec: 0.692 [VAL SCORE: 1.424] || Test Sens: 0.659 | Test Spec: 0.692 [TEST SCORE: 1.351]

###### #90: INV. MELD + AFP + PIVKA-II + OPN + DKK-1

Optimized Thresholds: INV. MELD: 1.6294 | AFP: 42.0989 | PIVKA-II: 9071.5310 | OPN: 275.1615 | DKK-1: 495.1077

Train Sens: 0.817 | Train Spec: 0.774 [TRAIN SCORE: 1.591] || Val Sens: 0.732 | Val Spec: 0.692 [VAL SCORE: 1.424] || Test Sens: 0.659 | Test Spec: 0.692 [TEST SCORE: 1.351]

###### #91: CTP + MELD + INV. MELD + AFP + OPN + DKK-1

Optimized Thresholds: CTP: 10.5916 | MELD: 39.6007 | INV. MELD: 0.2348 | AFP: 38.2365 | OPN: 1797.5811 | DKK-1: 10317.7857

Train Sens: 0.770 | Train Spec: 0.783 [TRAIN SCORE: 1.552] || Val Sens: 0.561 | Val Spec: 0.846 [VAL SCORE: 1.407] || Test Sens: 0.659 | Test Spec: 0.821 [TEST SCORE: 1.479]

###### #92: CTP + MELD + INV. MELD + AFP + PIVKA-II + DKK-1

Optimized Thresholds: CTP: 10.5916 | MELD: 39.6007 | INV. MELD: 0.2348 | AFP: 38.2365 | PIVKA-II: 3528.8033 | DKK-1: 10317.7857

Train Sens: 0.754 | Train Spec: 0.783 [TRAIN SCORE: 1.537] || Val Sens: 0.561 | Val Spec: 0.846 [VAL SCORE: 1.407] || Test Sens: 0.634 | Test Spec: 0.821 [TEST SCORE: 1.455]

###### #93: CTP + MELD + INV. MELD + AFP + PIVKA-II + OPN

Optimized Thresholds: CTP: 10.5916 | MELD: 39.6007 | INV. MELD: 0.2348 | AFP: 38.2365 | PIVKA-II: 3528.8033 | OPN: 1913.4805

Train Sens: 0.770 | Train Spec: 0.783 [TRAIN SCORE: 1.552] || Val Sens: 0.561 | Val Spec: 0.846 [VAL SCORE: 1.407] || Test Sens: 0.659 | Test Spec: 0.821 [TEST SCORE: 1.479]

###### #94: CTP + MELD + INV. MELD + AFP

Optimized Thresholds: CTP: 9.8739 | MELD: 25.6760 | INV. MELD: 0.2088 | AFP: 40.2954

Train Sens: 0.762 | Train Spec: 0.748 [TRAIN SCORE: 1.510] || Val Sens: 0.610 | Val Spec: 0.795 [VAL SCORE: 1.405] || Test Sens: 0.659 | Test Spec: 0.769 [TEST SCORE: 1.428]

###### #95: AGE + INV. MELD + AFP + OPN

Optimized Thresholds: AGE: 69.4701 | INV. MELD: 3.3008 | AFP: 24.3693 | OPN: 4623.9120

Train Sens: 0.754 | Train Spec: 0.757 [TRAIN SCORE: 1.510] || Val Sens: 0.610 | Val Spec: 0.795 [VAL SCORE: 1.405] || Test Sens: 0.659 | Test Spec: 0.795 [TEST SCORE: 1.453]

###### #96: AGE + INV. MELD + AFP + PIVKA-II

Optimized Thresholds: AGE: 69.4701 | INV. MELD: 3.3008 | AFP: 24.3693 | PIVKA-II: 9105.5756

Train Sens: 0.754 | Train Spec: 0.757 [TRAIN SCORE: 1.510] || Val Sens: 0.610 | Val Spec: 0.795 [VAL SCORE: 1.405] || Test

Sens: 0.659 | Test Spec: 0.795 [TEST SCORE: 1.453]

###### #97: CTP + INV. MELD + AFP + PIVKA-II

Optimized Thresholds: CTP: 8.0974 | INV. MELD: 1.0383 | AFP: 13.1778 | PIVKA-II: 4998.4407

Train Sens: 0.738 | Train Spec: 0.783 [TRAIN SCORE: 1.521] || Val Sens: 0.610 | Val Spec: 0.795 [VAL SCORE: 1.405] || Test Sens: 0.610 | Test Spec: 0.744 [TEST SCORE: 1.353]

###### #98: CTP + MELD + INV. MELD + AFP + PIVKA-II

Optimized Thresholds: CTP: 9.8739 | MELD: 25.6760 | INV. MELD: 0.2088 | AFP: 40.2954 | PIVKA-II: 5269.0966

Train Sens: 0.762 | Train Spec: 0.748 [TRAIN SCORE: 1.510] || Val Sens: 0.610 | Val Spec: 0.795 [VAL SCORE: 1.405] || Test Sens: 0.659 | Test Spec: 0.769 [TEST SCORE: 1.428]

###### #99: CTP + MELD + INV. MELD + AFP + DKK-1

Optimized Thresholds: CTP: 9.8739 | MELD: 25.6760 | INV. MELD: 0.2088 | AFP: 40.2954 | DKK-1: 14454.1530

Train Sens: 0.762 | Train Spec: 0.748 [TRAIN SCORE: 1.510] || Val Sens: 0.610 | Val Spec: 0.795 [VAL SCORE: 1.405] || Test Sens: 0.659 | Test Spec: 0.769 [TEST SCORE: 1.428]

###### #100: MELD + INV. MELD + PIVKA-II + OPN + DKK-1

Optimized Thresholds: MELD: 48.7581 | INV. MELD: 1.2069 | PIVKA-II: 10.6324 | OPN: 1328.9453 | DKK-1: 577.2045

Train Sens: 0.738 | Train Spec: 0.791 [TRAIN SCORE: 1.529] || Val Sens: 0.610 | Val Spec: 0.795 [VAL SCORE: 1.405] || Test Sens: 0.585 | Test Spec: 0.795 [TEST SCORE: 1.380]

###### #101: INV. MELD + PIVKA-II + OPN

Optimized Thresholds: INV. MELD: 0.2261 | PIVKA-II: 10.4085 | OPN: 1484.2514

Train Sens: 0.722 | Train Spec: 0.757 [TRAIN SCORE: 1.479] || Val Sens: 0.610 | Val Spec: 0.795 [VAL SCORE: 1.405] || Test Sens: 0.659 | Test Spec: 0.769 [TEST SCORE: 1.428]

###### #102: CTP + MELD + INV. MELD + AFP + OPN

Optimized Thresholds: CTP: 9.8739 | MELD: 25.6760 | INV. MELD: 0.2088 | AFP: 40.2954 | OPN: 2679.5688

Train Sens: 0.770 | Train Spec: 0.748 [TRAIN SCORE: 1.518] || Val Sens: 0.610 | Val Spec: 0.795 [VAL SCORE: 1.405] || Test Sens: 0.683 | Test Spec: 0.769 [TEST SCORE: 1.452]

###### #103: CTP + MELD + AFP + PIVKA-II + OPN

Optimized Thresholds: CTP: 9.1518 | MELD: 45.6422 | AFP: 15.1744 | PIVKA-II: 2608.4946 | OPN: 143.5324

Train Sens: 0.746 | Train Spec: 0.774 [TRAIN SCORE: 1.520] || Val Sens: 0.634 | Val Spec: 0.769 [VAL SCORE: 1.403] || Test Sens: 0.659 | Test Spec: 0.744 [TEST SCORE: 1.402]

###### #104: MELD + AFP

Optimized Thresholds: MELD: 22.6739 | AFP: 9.1927

Train Sens: 0.754 | Train Spec: 0.748 [TRAIN SCORE: 1.502] || Val Sens: 0.634 | Val Spec: 0.769 [VAL SCORE: 1.403] || Test Sens: 0.634 | Test Spec: 0.590 [TEST SCORE: 1.224]

###### #105: CTP + AFP + PIVKA-II + OPN + DKK-1

Optimized Thresholds: CTP: 9.7845 | AFP: 39.3956 | PIVKA-II: 6319.0252 | OPN: 1453.6529 | DKK-1: 547.4681

Train Sens: 0.786 | Train Spec: 0.791 [TRAIN SCORE: 1.577] || Val Sens: 0.634 | Val Spec: 0.769 [VAL SCORE: 1.403] || Test

Sens: 0.585 | Test Spec: 0.718 [TEST SCORE: 1.303]

##### #106: AFP + OPN

Optimized Thresholds: AFP: 16.3857 | OPN: 133.1729

Train Sens: 0.754 | Train Spec: 0.774 [TRAIN SCORE: 1.528] || Val Sens: 0.634 | Val Spec: 0.769 [VAL SCORE: 1.403] || Test Sens: 0.634 | Test Spec: 0.769 [TEST SCORE: 1.403]

##### #107: AGE + MELD + PIVKA-II + OPN

Optimized Thresholds: AGE: 74.0645 | MELD: 20.5544 | PIVKA-II: 9.1361 | OPN: 107.9839

Train Sens: 0.730 | Train Spec: 0.748 [TRAIN SCORE: 1.478] || Val Sens: 0.659 | Val Spec: 0.744 [VAL SCORE: 1.402] || Test Sens: 0.683 | Test Spec: 0.692 [TEST SCORE: 1.375]

##### #108: AFP + PIVKA-II + OPN

Optimized Thresholds: AFP: 15.5447 | PIVKA-II: 1317.2553 | OPN: 118.8853

Train Sens: 0.762 | Train Spec: 0.774 [TRAIN SCORE: 1.536] || Val Sens: 0.659 | Val Spec: 0.744 [VAL SCORE: 1.402] || Test Sens: 0.659 | Test Spec: 0.744 [TEST SCORE: 1.402]

##### #109: CTP + PIVKA-II + OPN

Optimized Thresholds: CTP: 10.2692 | PIVKA-II: 9.7626 | OPN: 95.3618

Train Sens: 0.714 | Train Spec: 0.713 [TRAIN SCORE: 1.427] || Val Sens: 0.683 | Val Spec: 0.718 [VAL SCORE: 1.401] || Test Sens: 0.683 | Test Spec: 0.667 [TEST SCORE: 1.350]

##### #110: AGE + PIVKA-II + OPN

Optimized Thresholds: AGE: 74.0409 | PIVKA-II: 11.0199 | OPN: 96.9789

Train Sens: 0.746 | Train Spec: 0.730 [TRAIN SCORE: 1.476] || Val Sens: 0.683 | Val Spec: 0.718 [VAL SCORE: 1.401] || Test Sens: 0.732 | Test Spec: 0.641 [TEST SCORE: 1.373]

##### #111: MELD + AFP + PIVKA-II + OPN + DKK-1

Optimized Thresholds: MELD: 31.6704 | AFP: 44.0166 | PIVKA-II: 8769.9894 | OPN: 4880.4090 | DKK-1: 480.2634

Train Sens: 0.794 | Train Spec: 0.791 [TRAIN SCORE: 1.585] || Val Sens: 0.732 | Val Spec: 0.667 [VAL SCORE: 1.398] || Test Sens: 0.610 | Test Spec: 0.692 [TEST SCORE: 1.302]

##### #112: AFP + PIVKA-II* + OPN + DKK-1

Optimized Thresholds: AFP: 36.4003 | PIVKA-II*: 5792.1982 | OPN: 1291.4765 | DKK-1: 475.7896

Train Sens: 0.817 | Train Spec: 0.791 [TRAIN SCORE: 1.609] || Val Sens: 0.732 | Val Spec: 0.667 [VAL SCORE: 1.398] || Test Sens: 0.683 | Test Spec: 0.692 [TEST SCORE: 1.375]

##### #113: MELD + INV. MELD + PIVKA-II + OPN

Optimized Thresholds: MELD: 64.9606 | INV. MELD: 0.2406 | PIVKA-II: 11.9378 | OPN: 612.5385

Train Sens: 0.714 | Train Spec: 0.783 [TRAIN SCORE: 1.497] || Val Sens: 0.561 | Val Spec: 0.821 [VAL SCORE: 1.381] || Test Sens: 0.634 | Test Spec: 0.821 [TEST SCORE: 1.455]

##### #114: CTP + INV. MELD + AFP + PIVKA-II + OPN + DKK-1

Optimized Thresholds: CTP: 9.7538 | INV. MELD: 0.2265 | AFP: 39.8410 | PIVKA-II: 2926.8623 | OPN: 3255.7859 | DKK-1: 19668.9969

Train Sens: 0.770 | Train Spec: 0.765 [TRAIN SCORE: 1.535] || Val Sens: 0.585 | Val Spec: 0.795 [VAL SCORE: 1.380] || Test Sens: 0.659 | Test Spec: 0.795 [TEST SCORE: 1.453]

###### #115: AGE + CTP + MELD + INV. MELD + AFP + PIVKA-II + OPN + DKK-1

Optimized Thresholds: AGE: 69.8240 | CTP: 10.1256 | MELD: 35.1264 | INV. MELD: 4.1753 | AFP: 38.8429 | PIVKA-II: 5775.1940 | OPN: 329.3405 | DKK-1: 17805.8913

Train Sens: 0.738 | Train Spec: 0.739 [TRAIN SCORE: 1.477] || Val Sens: 0.585 | Val Spec: 0.795 [VAL SCORE: 1.380] || Test Sens: 0.659 | Test Spec: 0.821 [TEST SCORE: 1.479]

###### #116: CTP + INV. MELD + PIVKA-II + DKK-1

Optimized Thresholds: CTP: 7.8317 | INV. MELD: 4.3700 | PIVKA-II: 11.8320 | DKK-1: 571.1304

Train Sens: 0.762 | Train Spec: 0.739 [TRAIN SCORE: 1.501] || Val Sens: 0.610 | Val Spec: 0.769 [VAL SCORE: 1.379] || Test Sens: 0.659 | Test Spec: 0.692 [TEST SCORE: 1.351]

###### #117: AGE + INV. MELD

Optimized Thresholds: AGE: 77.7920 | INV. MELD: 0.1387

Train Sens: 0.587 | Train Spec: 0.600 [TRAIN SCORE: 1.187] || Val Sens: 0.610 | Val Spec: 0.769 [VAL SCORE: 1.379] || Test Sens: 0.610 | Test Spec: 0.667 [TEST SCORE: 1.276]

###### #118: CTP + INV. MELD + PIVKA-II + OPN + DKK-1

Optimized Thresholds: CTP: 9.6814 | INV. MELD: 1.2069 | PIVKA-II: 10.6324 | OPN: 1328.9453 | DKK-1: 577.2045

Train Sens: 0.738 | Train Spec: 0.765 [TRAIN SCORE: 1.503] || Val Sens: 0.610 | Val Spec: 0.769 [VAL SCORE: 1.379] || Test Sens: 0.610 | Test Spec: 0.769 [TEST SCORE: 1.379]

###### #119: CTP + AFP + OPN

Optimized Thresholds: CTP: 10.1088 | AFP: 13.9824 | OPN: 142.7693

Train Sens: 0.754 | Train Spec: 0.765 [TRAIN SCORE: 1.519] || Val Sens: 0.634 | Val Spec: 0.744 [VAL SCORE: 1.378] || Test Sens: 0.659 | Test Spec: 0.744 [TEST SCORE: 1.402]

###### #120: MELD + AFP + OPN

Optimized Thresholds: MELD: 23.1815 | AFP: 13.2340 | OPN: 137.3397

Train Sens: 0.762 | Train Spec: 0.757 [TRAIN SCORE: 1.518] || Val Sens: 0.659 | Val Spec: 0.718 [VAL SCORE: 1.376] || Test Sens: 0.683 | Test Spec: 0.744 [TEST SCORE: 1.427]

###### #121: MELD + PIVKA-II + DKK-1

Optimized Thresholds: MELD: 52.1374 | PIVKA-II: 11.0199 | DKK-1: 509.8858

Train Sens: 0.778 | Train Spec: 0.774 [TRAIN SCORE: 1.552] || Val Sens: 0.659 | Val Spec: 0.718 [VAL SCORE: 1.376] || Test Sens: 0.634 | Test Spec: 0.718 [TEST SCORE: 1.352]

###### #122: CTP + AFP + OPN + DKK-1

Optimized Thresholds: CTP: 9.7190 | AFP: 35.9605 | OPN: 1665.3687 | DKK-1: 517.6719

Train Sens: 0.810 | Train Spec: 0.774 [TRAIN SCORE: 1.583] || Val Sens: 0.659 | Val Spec: 0.718 [VAL SCORE: 1.376] || Test Sens: 0.659 | Test Spec: 0.718 [TEST SCORE: 1.376]

###### #123: CTP + MELD + AFP + OPN

Optimized Thresholds: CTP: 9.2961 | MELD: 36.1849 | AFP: 13.7262 | OPN: 140.1006

Train Sens: 0.754 | Train Spec: 0.748 [TRAIN SCORE: 1.502] || Val Sens: 0.659 | Val Spec: 0.718 [VAL SCORE: 1.376] || Test Sens: 0.659 | Test Spec: 0.744 [TEST SCORE: 1.402]

##### #124: CTP + AFP + PIVKA-II + DKK-1

Optimized Thresholds: CTP: 9.7190 | AFP: 35.9605 | PIVKA-II: 3267.9285 | DKK-1: 517.6719

Train Sens: 0.810 | Train Spec: 0.774 [TRAIN SCORE: 1.583] || Val Sens: 0.659 | Val Spec: 0.718 [VAL SCORE: 1.376] || Test Sens: 0.659 | Test Spec: 0.718 [TEST SCORE: 1.376]

##### #125: MELD + AFP + PIVKA-II + OPN

Optimized Thresholds: MELD: 46.1249 | AFP: 13.4241 | PIVKA-II: 2741.1701 | OPN: 137.9699

Train Sens: 0.754 | Train Spec: 0.765 [TRAIN SCORE: 1.519] || Val Sens: 0.659 | Val Spec: 0.718 [VAL SCORE: 1.376] || Test Sens: 0.683 | Test Spec: 0.744 [TEST SCORE: 1.427]

##### #126: AGE + CTP + PIVKA-II + OPN

Optimized Thresholds: AGE: 74.0645 | CTP: 7.3875 | PIVKA-II: 9.1361 | OPN: 107.9839

Train Sens: 0.722 | Train Spec: 0.722 [TRAIN SCORE: 1.444] || Val Sens: 0.683 | Val Spec: 0.692 [VAL SCORE: 1.375] || Test Sens: 0.707 | Test Spec: 0.692 [TEST SCORE: 1.400]

##### #127: AGE + MELD + PIVKA-II + DKK-1

Optimized Thresholds: AGE: 79.1809 | MELD: 32.5155 | PIVKA-II: 13.3903 | DKK-1: 489.7092

Train Sens: 0.770 | Train Spec: 0.765 [TRAIN SCORE: 1.535] || Val Sens: 0.707 | Val Spec: 0.667 [VAL SCORE: 1.374] || Test Sens: 0.659 | Test Spec: 0.718 [TEST SCORE: 1.376]

##### #128: CTP + MELD + PIVKA-II + OPN

Optimized Thresholds: CTP: 7.5410 | MELD: 35.9853 | PIVKA-II: 10.9278 | OPN: 97.7823

Train Sens: 0.706 | Train Spec: 0.722 [TRAIN SCORE: 1.428] || Val Sens: 0.707 | Val Spec: 0.667 [VAL SCORE: 1.374] || Test Sens: 0.659 | Test Spec: 0.641 [TEST SCORE: 1.300]

##### #129: AGE + PIVKA-II + DKK-1

Optimized Thresholds: AGE: 81.8663 | PIVKA-II: 10.7664 | DKK-1: 490.7707

Train Sens: 0.778 | Train Spec: 0.765 [TRAIN SCORE: 1.543] || Val Sens: 0.732 | Val Spec: 0.641 [VAL SCORE: 1.373] || Test Sens: 0.634 | Test Spec: 0.667 [TEST SCORE: 1.301]

##### #130: AGE + INV. MELD + PIVKA-II + OPN

Optimized Thresholds: AGE: 84.9920 | INV. MELD: 0.2406 | PIVKA-II: 11.9378 | OPN: 612.5385

Train Sens: 0.714 | Train Spec: 0.791 [TRAIN SCORE: 1.506] || Val Sens: 0.561 | Val Spec: 0.795 [VAL SCORE: 1.356] || Test Sens: 0.634 | Test Spec: 0.821 [TEST SCORE: 1.455]

##### #131: AGE + CTP + INV. MELD + AFP + PIVKA-II + DKK-1

Optimized Thresholds: AGE: 80.7117 | CTP: 8.9366 | INV. MELD: 0.2348 | AFP: 38.2365 | PIVKA-II: 3528.8033 | DKK-1: 10317.7857

Train Sens: 0.778 | Train Spec: 0.748 [TRAIN SCORE: 1.526] || Val Sens: 0.585 | Val Spec: 0.769 [VAL SCORE: 1.355] || Test Sens: 0.659 | Test Spec: 0.769 [TEST SCORE: 1.428]

##### #132: AGE + CTP + INV. MELD + AFP + OPN + DKK-1

Optimized Thresholds: AGE: 80.7117 | CTP: 8.9366 | INV. MELD: 0.2348 | AFP: 38.2365 | OPN: 1797.5811 | DKK-1: 10317.7857

Train Sens: 0.778 | Train Spec: 0.748 [TRAIN SCORE: 1.526] || Val Sens: 0.585 | Val Spec: 0.769 [VAL SCORE: 1.355] || Test Sens: 0.683 | Test Spec: 0.769 [TEST SCORE: 1.452]

**#133: AGE + CTP + INV. MELD + AFP + PIVKA-II + OPN**

Optimized Thresholds: AGE: 80.7117 | CTP: 8.9366 | INV. MELD: 0.2348 | AFP: 38.2365 | PIVKA-II: 3528.8033 | OPN: 1913.4805

Train Sens: 0.778 | Train Spec: 0.748 [TRAIN SCORE: 1.526] || Val Sens: 0.585 | Val Spec: 0.769 [VAL SCORE: 1.355] || Test Sens: 0.683 | Test Spec: 0.769 [TEST SCORE: 1.452]

**#134: AGE + MELD + INV. MELD + PIVKA-II**

Optimized Thresholds: AGE: 69.2953 | MELD: 23.5452 | INV. MELD: 0.6118 | PIVKA-II: 12.3573

Train Sens: 0.730 | Train Spec: 0.757 [TRAIN SCORE: 1.487] || Val Sens: 0.585 | Val Spec: 0.769 [VAL SCORE: 1.355] || Test Sens: 0.585 | Test Spec: 0.821 [TEST SCORE: 1.406]

**#135: INV. MELD + PIVKA-II + DKK-1**

Optimized Thresholds: INV. MELD: 1.0352 | PIVKA-II: 13.0934 | DKK-1: 518.8294

Train Sens: 0.770 | Train Spec: 0.783 [TRAIN SCORE: 1.552] || Val Sens: 0.610 | Val Spec: 0.744 [VAL SCORE: 1.353] || Test Sens: 0.634 | Test Spec: 0.769 [TEST SCORE: 1.403]

**#136: AGE + CTP + INV. MELD + PIVKA-II**

Optimized Thresholds: AGE: 81.2717 | CTP: 8.2374 | INV. MELD: 0.2358 | PIVKA-II: 12.0933

Train Sens: 0.722 | Train Spec: 0.739 [TRAIN SCORE: 1.461] || Val Sens: 0.610 | Val Spec: 0.744 [VAL SCORE: 1.353] || Test Sens: 0.634 | Test Spec: 0.795 [TEST SCORE: 1.429]

**#137: AGE + MELD + INV. MELD + PIVKA-II + OPN**

Optimized Thresholds: AGE: 74.0454 | MELD: 23.0411 | INV. MELD: 0.6039 | PIVKA-II: 13.1209 | OPN: 120.0472

Train Sens: 0.722 | Train Spec: 0.791 [TRAIN SCORE: 1.514] || Val Sens: 0.610 | Val Spec: 0.744 [VAL SCORE: 1.353] || Test Sens: 0.683 | Test Spec: 0.872 [TEST SCORE: 1.555]

**#138: AGE + CTP + PIVKA-II**

Optimized Thresholds: AGE: 69.5654 | CTP: 8.0844 | PIVKA-II: 10.0927

Train Sens: 0.706 | Train Spec: 0.722 [TRAIN SCORE: 1.428] || Val Sens: 0.610 | Val Spec: 0.744 [VAL SCORE: 1.353] || Test Sens: 0.561 | Test Spec: 0.769 [TEST SCORE: 1.330]

**#139: AGE + CTP + MELD + INV. MELD + PIVKA-II + OPN**

Optimized Thresholds: AGE: 69.2493 | CTP: 8.7096 | MELD: 47.9747 | INV. MELD: 2.7565 | PIVKA-II: 9.0251 | OPN: 2960.4202

Train Sens: 0.722 | Train Spec: 0.713 [TRAIN SCORE: 1.435] || Val Sens: 0.610 | Val Spec: 0.744 [VAL SCORE: 1.353] || Test Sens: 0.634 | Test Spec: 0.744 [TEST SCORE: 1.378]

**#140: CTP + MELD + PIVKA-II + OPN + DKK-1**

Optimized Thresholds: CTP: 9.3007 | MELD: 27.6934 | PIVKA-II: 12.4977 | OPN: 2031.1775 | DKK-1: 507.2223

Train Sens: 0.786 | Train Spec: 0.757 [TRAIN SCORE: 1.542] || Val Sens: 0.634 | Val Spec: 0.718 [VAL SCORE: 1.352] || Test Sens: 0.659 | Test Spec: 0.744 [TEST SCORE: 1.402]

**#141: MELD + INV. MELD + AFP + PIVKA-II + DKK-1**

Optimized Thresholds: MELD: 31.6588 | INV. MELD: 1.7937 | AFP: 29859.4810 | PIVKA-II: 10.9539 | DKK-1: 528.9397

Train Sens: 0.770 | Train Spec: 0.774 [TRAIN SCORE: 1.544] || Val Sens: 0.634 | Val Spec: 0.718 [VAL SCORE: 1.352] || Test Sens: 0.634 | Test Spec: 0.718 [TEST SCORE: 1.352]

##### #142: AGE + CTP + MELD + PIVKA-II + OPN + DKK-1

Optimized Thresholds: AGE: 78.1856 | CTP: 10.3377 | MELD: 37.4050 | PIVKA-II: 18.6855 | OPN: 1645.7683 | DKK-1: 510.2517

Train Sens: 0.762 | Train Spec: 0.765 [TRAIN SCORE: 1.527] || Val Sens: 0.634 | Val Spec: 0.718 [VAL SCORE: 1.352] || Test Sens: 0.659 | Test Spec: 0.795 [TEST SCORE: 1.453]

##### #143: CTP + AFP + PIVKA-II + OPN

Optimized Thresholds: CTP: 9.4673 | AFP: 13.4241 | PIVKA-II: 2741.1701 | OPN: 137.9699

Train Sens: 0.754 | Train Spec: 0.757 [TRAIN SCORE: 1.510] || Val Sens: 0.659 | Val Spec: 0.692 [VAL SCORE: 1.351] || Test Sens: 0.683 | Test Spec: 0.744 [TEST SCORE: 1.427]

##### #144: INV. MELD + PIVKA-II + OPN + DKK-1

Optimized Thresholds: INV. MELD: 3.8226 | PIVKA-II: 14.1194 | OPN: 1660.4797 | DKK-1: 488.4382

Train Sens: 0.778 | Train Spec: 0.783 [TRAIN SCORE: 1.560] || Val Sens: 0.659 | Val Spec: 0.692 [VAL SCORE: 1.351] || Test Sens: 0.659 | Test Spec: 0.744 [TEST SCORE: 1.402]

##### #145: CTP + MELD + PIVKA-II + DKK-1

Optimized Thresholds: CTP: 10.5156 | MELD: 22.8360 | PIVKA-II: 13.5346 | DKK-1: 489.0697

Train Sens: 0.770 | Train Spec: 0.765 [TRAIN SCORE: 1.535] || Val Sens: 0.659 | Val Spec: 0.692 [VAL SCORE: 1.351] || Test Sens: 0.659 | Test Spec: 0.718 [TEST SCORE: 1.376]

##### #146: MELD* + PIVKA-II + OPN + DKK-1

Optimized Thresholds: MELD*: 38.3415 | PIVKA-II: 12.5639 | OPN: 1727.6392 | DKK-1: 494.7526

Train Sens: 0.778 | Train Spec: 0.783 [TRAIN SCORE: 1.560] || Val Sens: 0.659 | Val Spec: 0.692 [VAL SCORE: 1.351] || Test Sens: 0.634 | Test Spec: 0.718 [TEST SCORE: 1.352]

##### #147: AGE + CTP + PIVKA-II + DKK-1

Optimized Thresholds: AGE: 78.4477 | CTP: 9.1476 | PIVKA-II: 13.2459 | DKK-1: 490.3488

Train Sens: 0.786 | Train Spec: 0.739 [TRAIN SCORE: 1.525] || Val Sens: 0.707 | Val Spec: 0.641 [VAL SCORE: 1.348] || Test Sens: 0.683 | Test Spec: 0.692 [TEST SCORE: 1.375]

##### #148: AGE + MELD + AFP + OPN + DKK-1

Optimized Thresholds: AGE: 80.5309 | MELD: 30.5986 | AFP: 47.0318 | OPN: 475.7774 | DKK-1: 466.9367

Train Sens: 0.786 | Train Spec: 0.765 [TRAIN SCORE: 1.551] || Val Sens: 0.732 | Val Spec: 0.615 [VAL SCORE: 1.347] || Test Sens: 0.683 | Test Spec: 0.692 [TEST SCORE: 1.375]

##### #149: AFP + DKK-1

Optimized Thresholds: AFP: 38.9975 | DKK-1: 458.1944

Train Sens: 0.810 | Train Spec: 0.791 [TRAIN SCORE: 1.601] || Val Sens: 0.732 | Val Spec: 0.615 [VAL SCORE: 1.347] || Test Sens: 0.659 | Test Spec: 0.692 [TEST SCORE: 1.351]

##### #150: AFP + PIVKA-II + DKK-1

Optimized Thresholds: AFP: 39.6715 | PIVKA-II: 1515.1534 | DKK-1: 458.7658

Train Sens: 0.810 | Train Spec: 0.791 [TRAIN SCORE: 1.601] || Val Sens: 0.732 | Val Spec: 0.615 [VAL SCORE: 1.347] || Test Sens: 0.659 | Test Spec: 0.692 [TEST SCORE: 1.351]

###### #151: AFP + OPN + DKK-1

Optimized Thresholds: AFP: 39.6715 | OPN: 777.0551 | DKK-1: 458.7658

Train Sens: 0.817 | Train Spec: 0.791 [TRAIN SCORE: 1.609] || Val Sens: 0.732 | Val Spec: 0.615 [VAL SCORE: 1.347] || Test Sens: 0.683 | Test Spec: 0.692 [TEST SCORE: 1.375]

###### #152: AGE + MELD + AFP + PIVKA-II + DKK-1

Optimized Thresholds: AGE: 80.5309 | MELD: 30.5986 | AFP: 47.0318 | PIVKA-II: 920.6875 | DKK-1: 466.9367

Train Sens: 0.778 | Train Spec: 0.774 [TRAIN SCORE: 1.552] || Val Sens: 0.732 | Val Spec: 0.615 [VAL SCORE: 1.347] || Test Sens: 0.659 | Test Spec: 0.692 [TEST SCORE: 1.351]

###### #153: CTP + INV. MELD + AFP + PIVKA-II + DKK-1

Optimized Thresholds: CTP: 10.2962 | INV. MELD: 0.7802 | AFP: 50.6570 | PIVKA-II: 7273.7000 | DKK-1: 1284.2800

Train Sens: 0.587 | Train Spec: 0.913 [TRAIN SCORE: 1.500] || Val Sens: 0.415 | Val Spec: 0.923 [VAL SCORE: 1.338] || Test Sens: 0.415 | Test Spec: 1.000 [TEST SCORE: 1.415]

###### #154: AGE + CTP + MELD + INV. MELD + PIVKA-II

Optimized Thresholds: AGE: 70.8001 | CTP: 8.6505 | MELD: 15.9001 | INV. MELD: 4.2681 | PIVKA-II: 11.6862

Train Sens: 0.714 | Train Spec: 0.722 [TRAIN SCORE: 1.436] || Val Sens: 0.610 | Val Spec: 0.718 [VAL SCORE: 1.328] || Test Sens: 0.561 | Test Spec: 0.744 [TEST SCORE: 1.305]

###### #155: AGE + CTP + MELD + PIVKA-II

Optimized Thresholds: AGE: 69.7476 | CTP: 8.7081 | MELD: 19.9471 | PIVKA-II: 9.5052

Train Sens: 0.722 | Train Spec: 0.722 [TRAIN SCORE: 1.444] || Val Sens: 0.610 | Val Spec: 0.718 [VAL SCORE: 1.328] || Test Sens: 0.561 | Test Spec: 0.744 [TEST SCORE: 1.305]

###### #156: AGE + CTP + INV. MELD + PIVKA-II + OPN

Optimized Thresholds: AGE: 74.0454 | CTP: 7.5898 | INV. MELD: 0.6039 | PIVKA-II: 13.1209 | OPN: 120.0472

Train Sens: 0.722 | Train Spec: 0.748 [TRAIN SCORE: 1.470] || Val Sens: 0.634 | Val Spec: 0.692 [VAL SCORE: 1.326] || Test Sens: 0.707 | Test Spec: 0.821 [TEST SCORE: 1.528]

###### #157: CTP + INV. MELD + PIVKA-II + OPN

Optimized Thresholds: CTP: 6.1760 | INV. MELD: 0.2852 | PIVKA-II: 9.3377 | OPN: 966.6345

Train Sens: 0.722 | Train Spec: 0.722 [TRAIN SCORE: 1.444] || Val Sens: 0.634 | Val Spec: 0.692 [VAL SCORE: 1.326] || Test Sens: 0.683 | Test Spec: 0.692 [TEST SCORE: 1.375]

###### #158: AGE + CTP + MELD + AFP + PIVKA-II + OPN + DKK-1

Optimized Thresholds: AGE: 81.7297 | CTP: 10.1031 | MELD: 19.8168 | AFP: 50.2056 | PIVKA-II: 1811.7782 | OPN: 4577.0188 | DKK-1: 531.1837

Train Sens: 0.770 | Train Spec: 0.783 [TRAIN SCORE: 1.552] || Val Sens: 0.634 | Val Spec: 0.692 [VAL SCORE: 1.326] || Test Sens: 0.585 | Test Spec: 0.769 [TEST SCORE: 1.355]

###### #159: CTP + MELD + INV. MELD + PIVKA-II + OPN

Optimized Thresholds: CTP: 6.5219 | MELD: 45.9469 | INV. MELD: 0.2782 | PIVKA-II: 10.2036 | OPN: 1943.8320

Train Sens: 0.714 | Train Spec: 0.713 [TRAIN SCORE: 1.427] || Val Sens: 0.634 | Val Spec: 0.692 [VAL SCORE: 1.326] || Test Sens: 0.659 | Test Spec: 0.667 [TEST SCORE: 1.325]

###### #160: CTP + MELD + INV. MELD + PIVKA-II

Optimized Thresholds: CTP: 6.5219 | MELD: 45.9469 | INV. MELD: 0.2782 | PIVKA-II: 10.2036

Train Sens: 0.706 | Train Spec: 0.713 [TRAIN SCORE: 1.419] || Val Sens: 0.634 | Val Spec: 0.692 [VAL SCORE: 1.326] || Test Sens: 0.634 | Test Spec: 0.667 [TEST SCORE: 1.301]

###### #161: AGE + PIVKA-II + OPN* + DKK-1

Optimized Thresholds: AGE: 83.4645 | PIVKA-II: 13.4274 | OPN*: 3261.2352 | DKK-1: 487.6887

Train Sens: 0.762 | Train Spec: 0.783 [TRAIN SCORE: 1.545] || Val Sens: 0.659 | Val Spec: 0.667 [VAL SCORE: 1.325] || Test Sens: 0.634 | Test Spec: 0.718 [TEST SCORE: 1.352]

###### #162: AGE + CTP + MELD + PIVKA-II + OPN

Optimized Thresholds: AGE: 75.5799 | CTP: 7.7513 | MELD: 15.7870 | PIVKA-II: 10.7439 | OPN: 109.8422

Train Sens: 0.730 | Train Spec: 0.739 [TRAIN SCORE: 1.469] || Val Sens: 0.659 | Val Spec: 0.667 [VAL SCORE: 1.325] || Test Sens: 0.659 | Test Spec: 0.692 [TEST SCORE: 1.351]

###### #163: PIVKA-II + OPN + DKK-1

Optimized Thresholds: PIVKA-II: 12.7814 | OPN: 1470.4473 | DKK-1: 480.3206

Train Sens: 0.778 | Train Spec: 0.783 [TRAIN SCORE: 1.560] || Val Sens: 0.683 | Val Spec: 0.641 [VAL SCORE: 1.324] || Test Sens: 0.659 | Test Spec: 0.718 [TEST SCORE: 1.376]

###### #164: INV. MELD + AFP + OPN + DKK-1

Optimized Thresholds: INV. MELD: 1.6289 | AFP: 44.8452 | OPN: 1486.7806 | DKK-1: 461.2436

Train Sens: 0.802 | Train Spec: 0.791 [TRAIN SCORE: 1.593] || Val Sens: 0.707 | Val Spec: 0.615 [VAL SCORE: 1.323] || Test Sens: 0.659 | Test Spec: 0.692 [TEST SCORE: 1.351]

###### #165: CTP + MELD + AFP + PIVKA-II + DKK-1

Optimized Thresholds: CTP: 10.5744 | MELD: 30.5986 | AFP: 47.0318 | PIVKA-II: 920.6875 | DKK-1: 466.9367

Train Sens: 0.778 | Train Spec: 0.774 [TRAIN SCORE: 1.552] || Val Sens: 0.707 | Val Spec: 0.615 [VAL SCORE: 1.323] || Test Sens: 0.659 | Test Spec: 0.692 [TEST SCORE: 1.351]

###### #166: CTP + MELD + AFP + OPN + DKK-1

Optimized Thresholds: CTP: 10.5744 | MELD: 30.5986 | AFP: 47.0318 | OPN: 475.7774 | DKK-1: 466.9367

Train Sens: 0.786 | Train Spec: 0.765 [TRAIN SCORE: 1.551] || Val Sens: 0.707 | Val Spec: 0.615 [VAL SCORE: 1.323] || Test Sens: 0.683 | Test Spec: 0.692 [TEST SCORE: 1.375]

###### #167: INV. MELD + AFP + PIVKA-II + DKK-1

Optimized Thresholds: INV. MELD: 1.6289 | AFP: 44.8452 | PIVKA-II: 2915.5476 | DKK-1: 461.2436

Train Sens: 0.794 | Train Spec: 0.791 [TRAIN SCORE: 1.585] || Val Sens: 0.707 | Val Spec: 0.615 [VAL SCORE: 1.323] || Test Sens: 0.659 | Test Spec: 0.692 [TEST SCORE: 1.351]

##### #168: AGE + MELD* + PIVKA-II

Optimized Thresholds: AGE: 67.0164 | MELD*: 37.6979 | PIVKA-II: 12.7022

Train Sens: 0.714 | Train Spec: 0.722 [TRAIN SCORE: 1.436] || Val Sens: 0.561 | Val Spec: 0.744 [VAL SCORE: 1.305] || Test Sens: 0.634 | Test Spec: 0.846 [TEST SCORE: 1.480]

##### #169: AGE + MELD + INV. MELD + PIVKA-II + DKK-1

Optimized Thresholds: AGE: 79.0489 | MELD: 23.3688 | INV. MELD: 4.0086 | PIVKA-II: 19.3552 | DKK-1: 539.8450

Train Sens: 0.730 | Train Spec: 0.800 [TRAIN SCORE: 1.530] || Val Sens: 0.585 | Val Spec: 0.718 [VAL SCORE: 1.303] || Test Sens: 0.634 | Test Spec: 0.795 [TEST SCORE: 1.429]

##### #170: AGE + MELD + PIVKA-II + OPN + DKK-1

Optimized Thresholds: AGE: 84.5094 | MELD: 48.4513 | PIVKA-II: 12.8440 | OPN: 3355.8465 | DKK-1: 540.3210

Train Sens: 0.754 | Train Spec: 0.800 [TRAIN SCORE: 1.554] || Val Sens: 0.585 | Val Spec: 0.718 [VAL SCORE: 1.303] || Test Sens: 0.585 | Test Spec: 0.769 [TEST SCORE: 1.355]

##### #171: CTP + PIVKA-II + DKK-1

Optimized Thresholds: CTP: 9.8419 | PIVKA-II: 14.9110 | DKK-1: 483.4017

Train Sens: 0.762 | Train Spec: 0.765 [TRAIN SCORE: 1.527] || Val Sens: 0.659 | Val Spec: 0.641 [VAL SCORE: 1.300] || Test Sens: 0.659 | Test Spec: 0.718 [TEST SCORE: 1.376]

##### #172: CTP + MELD + PIVKA-II

Optimized Thresholds: CTP: 6.4384 | MELD: 15.9620 | PIVKA-II: 6.4482

Train Sens: 0.675 | Train Spec: 0.670 [TRAIN SCORE: 1.344] || Val Sens: 0.683 | Val Spec: 0.615 [VAL SCORE: 1.298] || Test Sens: 0.561 | Test Spec: 0.667 [TEST SCORE: 1.228]

##### #173: PIVKA-II + DKK-1

Optimized Thresholds: PIVKA-II: 12.4254 | DKK-1: 469.2185

Train Sens: 0.778 | Train Spec: 0.774 [TRAIN SCORE: 1.552] || Val Sens: 0.683 | Val Spec: 0.615 [VAL SCORE: 1.298] || Test Sens: 0.683 | Test Spec: 0.718 [TEST SCORE: 1.401]

##### #174: CTP + PIVKA-II + OPN + DKK-1

Optimized Thresholds: CTP: 10.5347 | PIVKA-II: 12.9684 | OPN: 1340.9423 | DKK-1: 465.6697

Train Sens: 0.778 | Train Spec: 0.765 [TRAIN SCORE: 1.543] || Val Sens: 0.683 | Val Spec: 0.615 [VAL SCORE: 1.298] || Test Sens: 0.707 | Test Spec: 0.718 [TEST SCORE: 1.425]

##### #175: AGE + AFP + PIVKA-II + DKK-1

Optimized Thresholds: AGE: 83.1160 | AFP: 44.8452 | PIVKA-II: 2915.5476 | DKK-1: 461.2436

Train Sens: 0.786 | Train Spec: 0.791 [TRAIN SCORE: 1.577] || Val Sens: 0.707 | Val Spec: 0.590 [VAL SCORE: 1.297] || Test Sens: 0.634 | Test Spec: 0.692 [TEST SCORE: 1.326]

##### #176: AGE + CTP + AFP + PIVKA-II + DKK-1

Optimized Thresholds: AGE: 76.4259 | CTP: 10.5244 | AFP: 41058.2239 | PIVKA-II: 16.1185 | DKK-1: 471.1216

Train Sens: 0.762 | Train Spec: 0.748 [TRAIN SCORE: 1.510] || Val Sens: 0.707 | Val Spec: 0.590 [VAL SCORE: 1.297] || Test Sens: 0.732 | Test Spec: 0.744 [TEST SCORE: 1.475]

###### #177: AGE + AFP + OPN + DKK-1

Optimized Thresholds: AGE: 83.1160 | AFP: 44.8452 | OPN: 1486.7806 | DKK-1: 461.2436

Train Sens: 0.794 | Train Spec: 0.791 [TRAIN SCORE: 1.585] || Val Sens: 0.707 | Val Spec: 0.590 [VAL SCORE: 1.297] || Test Sens: 0.634 | Test Spec: 0.692 [TEST SCORE: 1.326]

###### #178: AGE + PIVKA-II

Optimized Thresholds: AGE: 66.3802 | PIVKA-II: 12.4307

Train Sens: 0.722 | Train Spec: 0.722 [TRAIN SCORE: 1.444] || Val Sens: 0.561 | Val Spec: 0.718 [VAL SCORE: 1.279] || Test Sens: 0.634 | Test Spec: 0.846 [TEST SCORE: 1.480]

###### #179: AGE + CTP + PIVKA-II + OPN + DKK-1

Optimized Thresholds: AGE: 84.5094 | CTP: 9.6565 | PIVKA-II: 12.8440 | OPN: 3355.8465 | DKK-1: 540.3210

Train Sens: 0.762 | Train Spec: 0.774 [TRAIN SCORE: 1.536] || Val Sens: 0.585 | Val Spec: 0.692 [VAL SCORE: 1.278] || Test Sens: 0.610 | Test Spec: 0.744 [TEST SCORE: 1.353]

###### #180: AGE + CTP + INV. MELD + PIVKA-II + DKK-1

Optimized Thresholds: AGE: 79.0489 | CTP: 7.6164 | INV. MELD: 4.0086 | PIVKA-II: 19.3552 | DKK-1: 539.8450

Train Sens: 0.746 | Train Spec: 0.730 [TRAIN SCORE: 1.476] || Val Sens: 0.610 | Val Spec: 0.667 [VAL SCORE: 1.276] || Test Sens: 0.707 | Test Spec: 0.718 [TEST SCORE: 1.425]

###### #181: AGE + CTP + MELD + INV. MELD + PIVKA-II + DKK-1

Optimized Thresholds: AGE: 76.9009 | CTP: 9.1475 | MELD: 41.2230 | INV. MELD: 3.7604 | PIVKA-II: 35.1146 | DKK-1: 475.5705

Train Sens: 0.730 | Train Spec: 0.765 [TRAIN SCORE: 1.495] || Val Sens: 0.683 | Val Spec: 0.590 [VAL SCORE: 1.273] || Test Sens: 0.707 | Test Spec: 0.718 [TEST SCORE: 1.425]

###### #182: MELD + PIVKA-II

Optimized Thresholds: MELD: 22.8956 | PIVKA-II: 4.3048

Train Sens: 0.667 | Train Spec: 0.687 [TRAIN SCORE: 1.354] || Val Sens: 0.683 | Val Spec: 0.590 [VAL SCORE: 1.273] || Test Sens: 0.585 | Test Spec: 0.564 [TEST SCORE: 1.149]

###### #183: AGE + CTP + MELD + PIVKA-II + DKK-1

Optimized Thresholds: AGE: 76.4259 | CTP: 10.5244 | MELD: 22.4897 | PIVKA-II: 16.1185 | DKK-1: 471.1216

Train Sens: 0.770 | Train Spec: 0.748 [TRAIN SCORE: 1.518] || Val Sens: 0.707 | Val Spec: 0.564 [VAL SCORE: 1.271] || Test Sens: 0.732 | Test Spec: 0.744 [TEST SCORE: 1.475]

###### #184: MELD + INV. MELD

Optimized Thresholds: MELD: 13.4088 | INV. MELD: 0.1473

Train Sens: 0.587 | Train Spec: 0.574 [TRAIN SCORE: 1.161] || Val Sens: 0.634 | Val Spec: 0.615 [VAL SCORE: 1.250] || Test Sens: 0.634 | Test Spec: 0.564 [TEST SCORE: 1.198]

###### #185: CTP + MELD + INV. MELD + PIVKA-II + OPN + DKK-1

Optimized Thresholds: CTP: 9.6899 | MELD: 20.9421 | INV. MELD: 0.4604 | PIVKA-II: 20.8309 | OPN: 3475.8162 | DKK-1: 493.3207

Train Sens: 0.738 | Train Spec: 0.765 [TRAIN SCORE: 1.503] || Val Sens: 0.634 | Val Spec: 0.615 [VAL SCORE: 1.250] || Test Sens: 0.683 | Test Spec: 0.692 [TEST SCORE: 1.375]

##### #186: MELD + INV. MELD + PIVKA-II + DKK-1

Optimized Thresholds: MELD: 18.3306 | INV. MELD: 3.5590 | PIVKA-II: 16.0040 | DKK-1: 475.5374

Train Sens: 0.754 | Train Spec: 0.765 [TRAIN SCORE: 1.519] || Val Sens: 0.659 | Val Spec: 0.590 [VAL SCORE: 1.248] || Test Sens: 0.707 | Test Spec: 0.667 [TEST SCORE: 1.374]

##### #187: CTP + PIVKA-II

Optimized Thresholds: CTP: 9.5069 | PIVKA-II: 4.3009

Train Sens: 0.667 | Train Spec: 0.670 [TRAIN SCORE: 1.336] || Val Sens: 0.683 | Val Spec: 0.564 [VAL SCORE: 1.247] || Test Sens: 0.585 | Test Spec: 0.590 [TEST SCORE: 1.175]

##### #188: AGE + CTP + INV. MELD + PIVKA-II + OPN + DKK-1

Optimized Thresholds: AGE: 77.5157 | CTP: 10.8846 | INV. MELD: 2.2636 | PIVKA-II: 31.2969 | OPN: 3117.5942 | DKK-1: 451.6052

Train Sens: 0.730 | Train Spec: 0.783 [TRAIN SCORE: 1.513] || Val Sens: 0.683 | Val Spec: 0.564 [VAL SCORE: 1.247] || Test Sens: 0.707 | Test Spec: 0.692 [TEST SCORE: 1.400]

##### #189: CTP + MELD + INV. MELD + DKK-1

Optimized Thresholds: CTP: 7.3538 | MELD: 38.2143 | INV. MELD: 0.2508 | DKK-1: 548.0142

Train Sens: 0.690 | Train Spec: 0.704 [TRAIN SCORE: 1.395] || Val Sens: 0.537 | Val Spec: 0.692 [VAL SCORE: 1.229] || Test Sens: 0.610 | Test Spec: 0.667 [TEST SCORE: 1.276]

##### #190: MELD + OPN

Optimized Thresholds: MELD: 22.2717 | OPN: 79.6800

Train Sens: 0.627 | Train Spec: 0.635 [TRAIN SCORE: 1.262] || Val Sens: 0.634 | Val Spec: 0.590 [VAL SCORE: 1.224] || Test Sens: 0.634 | Test Spec: 0.641 [TEST SCORE: 1.275]

##### #191: CTP + MELD + INV. MELD + PIVKA-II + DKK-1

Optimized Thresholds: CTP: 9.8417 | MELD: 40.6774 | INV. MELD: 1.3417 | PIVKA-II: 17.9478 | DKK-1: 464.2649

Train Sens: 0.754 | Train Spec: 0.765 [TRAIN SCORE: 1.519] || Val Sens: 0.634 | Val Spec: 0.590 [VAL SCORE: 1.224] || Test Sens: 0.707 | Test Spec: 0.692 [TEST SCORE: 1.400]

##### #192: INV. MELD + DKK-1

Optimized Thresholds: INV. MELD: 0.2381 | DKK-1: 441.1822

Train Sens: 0.683 | Train Spec: 0.696 [TRAIN SCORE: 1.378] || Val Sens: 0.659 | Val Spec: 0.564 [VAL SCORE: 1.223] || Test Sens: 0.610 | Test Spec: 0.564 [TEST SCORE: 1.174]

##### #193: CTP + INV. MELD + DKK-1

Optimized Thresholds: CTP: 7.0060 | INV. MELD: 0.2671 | DKK-1: 513.7290

Train Sens: 0.690 | Train Spec: 0.704 [TRAIN SCORE: 1.395] || Val Sens: 0.537 | Val Spec: 0.667 [VAL SCORE: 1.203] || Test Sens: 0.634 | Test Spec: 0.667 [TEST SCORE: 1.301]

##### #194: AGE + INV. MELD + PIVKA-II + OPN + DKK-1

Optimized Thresholds: AGE: 77.5629 | INV. MELD: 0.3574 | PIVKA-II: 6762.9881 | OPN: 156.6308 | DKK-1: 540.5317

Train Sens: 0.738 | Train Spec: 0.739 [TRAIN SCORE: 1.477] || Val Sens: 0.585 | Val Spec: 0.615 [VAL SCORE: 1.201] || Test Sens: 0.659 | Test Spec: 0.795 [TEST SCORE: 1.453]

**#195: AGE + MELD + INV. MELD + OPN**

Optimized Thresholds: AGE: 73.9871 | MELD: 46.8003 | INV. MELD: 0.2282 | OPN: 106.4435

Train Sens: 0.683 | Train Spec: 0.678 [TRAIN SCORE: 1.361] || Val Sens: 0.585 | Val Spec: 0.615 [VAL SCORE: 1.201] || Test Sens: 0.659 | Test Spec: 0.615 [TEST SCORE: 1.274]

**#196: MELD + INV. MELD + DKK-1**

Optimized Thresholds: MELD: 22.5293 | INV. MELD: 0.2357 | DKK-1: 451.6863

Train Sens: 0.690 | Train Spec: 0.704 [TRAIN SCORE: 1.395] || Val Sens: 0.659 | Val Spec: 0.538 [VAL SCORE: 1.197] || Test Sens: 0.610 | Test Spec: 0.590 [TEST SCORE: 1.199]

**#197: AGE + INV. MELD + DKK-1**

Optimized Thresholds: AGE: 84.9107 | INV. MELD: 0.2311 | DKK-1: 428.7505

Train Sens: 0.690 | Train Spec: 0.678 [TRAIN SCORE: 1.369] || Val Sens: 0.683 | Val Spec: 0.513 [VAL SCORE: 1.196] || Test Sens: 0.634 | Test Spec: 0.564 [TEST SCORE: 1.198]

**#198: AGE + CTP + MELD + INV. MELD + PIVKA-II + OPN + DKK-1**

Optimized Thresholds: AGE: 79.2265 | CTP: 10.2185 | MELD: 24.7081 | INV. MELD: 4.1013 | PIVKA-II: 6364.6788 | OPN: 148.0747 | DKK-1: 487.2404

Train Sens: 0.738 | Train Spec: 0.748 [TRAIN SCORE: 1.486] || Val Sens: 0.585 | Val Spec: 0.590 [VAL SCORE: 1.175] || Test Sens: 0.707 | Test Spec: 0.718 [TEST SCORE: 1.425]

**#199: AGE + INV. MELD + OPN**

Optimized Thresholds: AGE: 77.0244 | INV. MELD: 0.2558 | OPN: 90.6169

Train Sens: 0.698 | Train Spec: 0.713 [TRAIN SCORE: 1.411] || Val Sens: 0.610 | Val Spec: 0.564 [VAL SCORE: 1.174] || Test Sens: 0.707 | Test Spec: 0.615 [TEST SCORE: 1.323]

**#200: AGE + CTP* + OPN**

Optimized Thresholds: AGE: 69.5233 | CTP*: 9.1744 | OPN: 95.0493

Train Sens: 0.643 | Train Spec: 0.661 [TRAIN SCORE: 1.304] || Val Sens: 0.512 | Val Spec: 0.641 [VAL SCORE: 1.153] || Test Sens: 0.683 | Test Spec: 0.641 [TEST SCORE: 1.324]
