## Supplementary material. Outputs TholdStormDX for this study. for "Methodological and Clinical Validation of TholdStormDX v0.0.1: An Advanced Stochastic Engine for the Optimization of Thresholds and Multimarker Panels Applied to Oncology": Lung TholdStormDX_RobustReport_20260404_124606.pdf

### Biomarker: Age

Processed: 03-Apr-2026 21:05

#### 1. Optimization Results

| MODEL | CUT-OFF | TRAIN (SE/SP) | VAL (SE/SP) | TEST (SE/SP) | R2 SCORE |
|---|---|---|---|---|---|
| Empirical (Exact) | 58.7658 | 0.598 / 0.598 | 0.674 / 0.624 | 0.605 / 0.484 | N/A |
| Logistic 2-Parameter | 59.2095 | 0.611 / 0.611 | 0.581 / 0.688 | 0.593 / 0.505 | 0.9977 |
| **Logistic 4-Parameter (Rec.)** | **59.3906** | **0.605 / 0.605** | **0.581 / 0.688** | **0.593 / 0.505** | **0.9990** |
| ThresholdXpert (Stochastic) | 58.5840 | 0.582 / 0.612 | 0.674 / 0.624 | 0.605 / 0.484 | N/A |

#### 2. Diagnostic Performance Curves (Training)

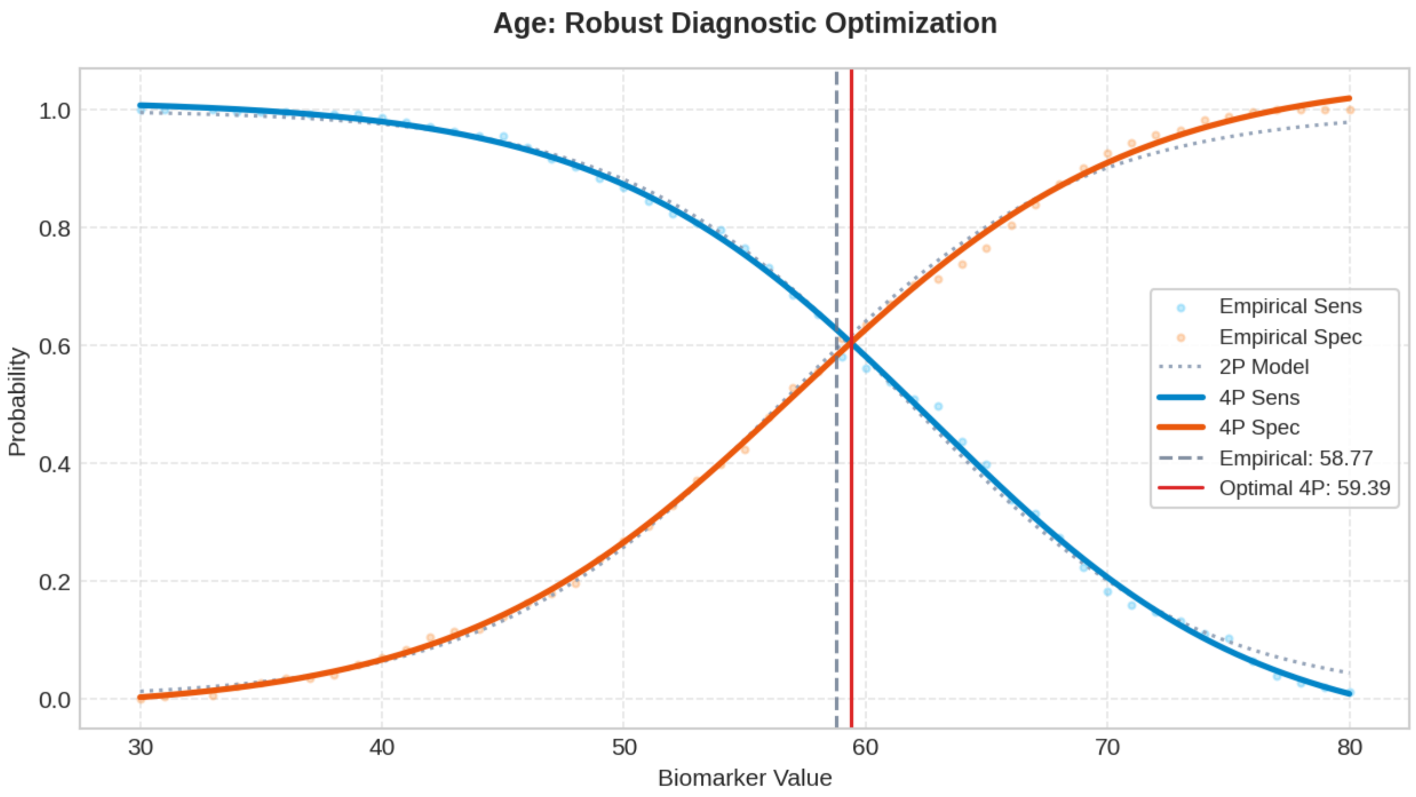

### Biomarker: BMI

Processed: 03-Apr-2026 21:08

#### 1. Optimization Results

| MODEL | CUT-OFF | TRAIN (SE/SP) | VAL (SE/SP) | TEST (SE/SP) | R2 SCORE |
|---|---|---|---|---|---|
| Empirical (Exact) | 24.8753 | 0.524 / 0.524 | 0.616 / 0.516 | 0.535 / 0.495 | N/A |
| Logistic 2-Parameter | 25.0434 | 0.533 / 0.533 | 0.593 / 0.527 | 0.523 / 0.516 | 0.9966 |
| **Logistic 4-Parameter (Rec.)** | **24.9672** | **0.531 / 0.531** | **0.616 / 0.516** | **0.535 / 0.505** | **0.9984** |
| ThresholdXpert (Stochastic) | 24.7485 | 0.542 / 0.521 | 0.628 / 0.462 | 0.535 / 0.473 | N/A |

#### 2. Diagnostic Performance Curves (Training)

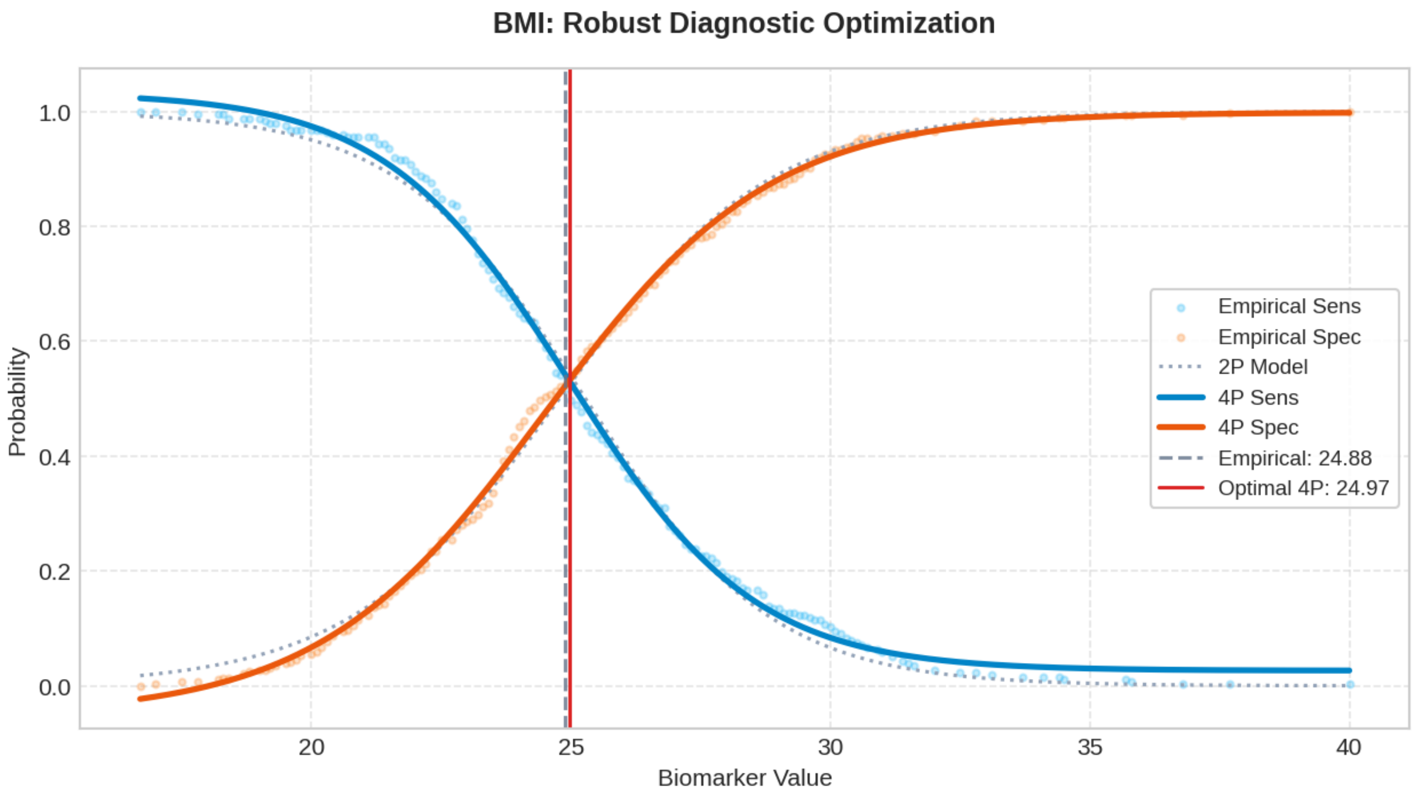

### Biomarker: Nodule Diameter

Processed: 03-Apr-2026 21:10

#### 1. Optimization Results

| MODEL | CUT-OFF | TRAIN (SE/SP) | VAL (SE/SP) | TEST (SE/SP) | R2 SCORE |
|---|---|---|---|---|---|
| Empirical (Exact) | 14.4619 | 0.731 / 0.731 | 0.698 / 0.731 | 0.744 / 0.763 | N/A |
| Logistic 2-Parameter | 14.6493 | 0.752 / 0.752 | 0.698 / 0.731 | 0.744 / 0.763 | 0.9841 |
| **Logistic 4-Parameter (Rec.)** | **14.4003** | **0.744 / 0.744** | **0.698 / 0.731** | **0.744 / 0.763** | **0.9948** |
| ThresholdXpert (Stochastic) | 14.8482 | 0.713 / 0.745 | 0.698 / 0.731 | 0.744 / 0.763 | N/A |

#### 2. Diagnostic Performance Curves (Training)

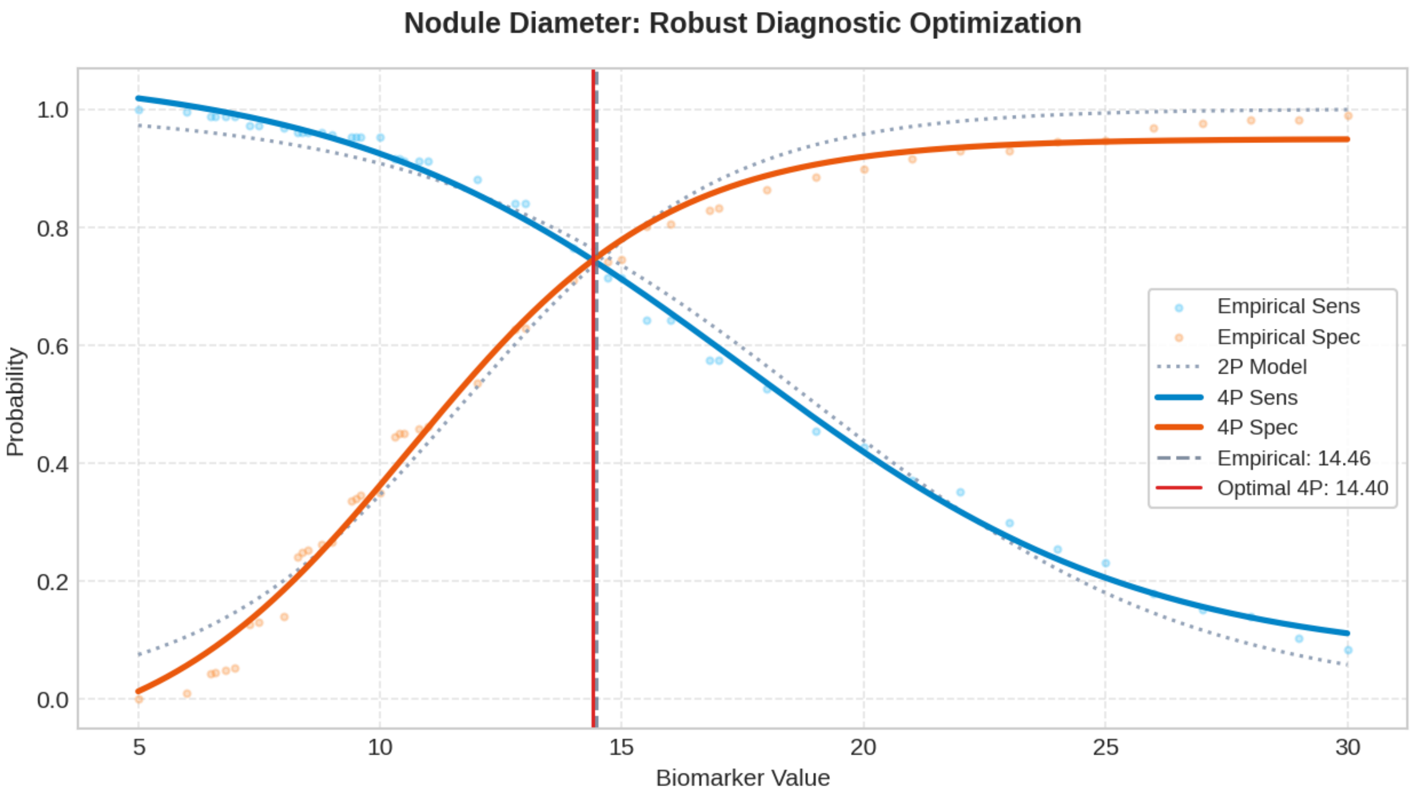

### Biomarker: CT.value

Processed: 03-Apr-2026 21:12

#### 1. Optimization Results

| MODEL | CUT-OFF | TRAIN (SE/SP) | VAL (SE/SP) | TEST (SE/SP) | R2 SCORE |
|---|---|---|---|---|---|
| Empirical (Exact) | -440.8566 | 0.801 / 0.801 | 0.907 / 0.806 | 0.814 / 0.785 | N/A |
| Logistic 2-Parameter | -453.4806 | 0.832 / 0.832 | 0.919 / 0.774 | 0.814 / 0.774 | 0.9838 |
| **Logistic 4-Parameter (Rec.)** | **-459.2655** | **0.824 / 0.824** | **0.930 / 0.774** | **0.826 / 0.774** | **0.9923** |
| ThresholdXpert (Stochastic) | -436.4884 | 0.801 / 0.811 | 0.907 / 0.806 | 0.779 / 0.796 | N/A |

#### 2. Diagnostic Performance Curves (Training)

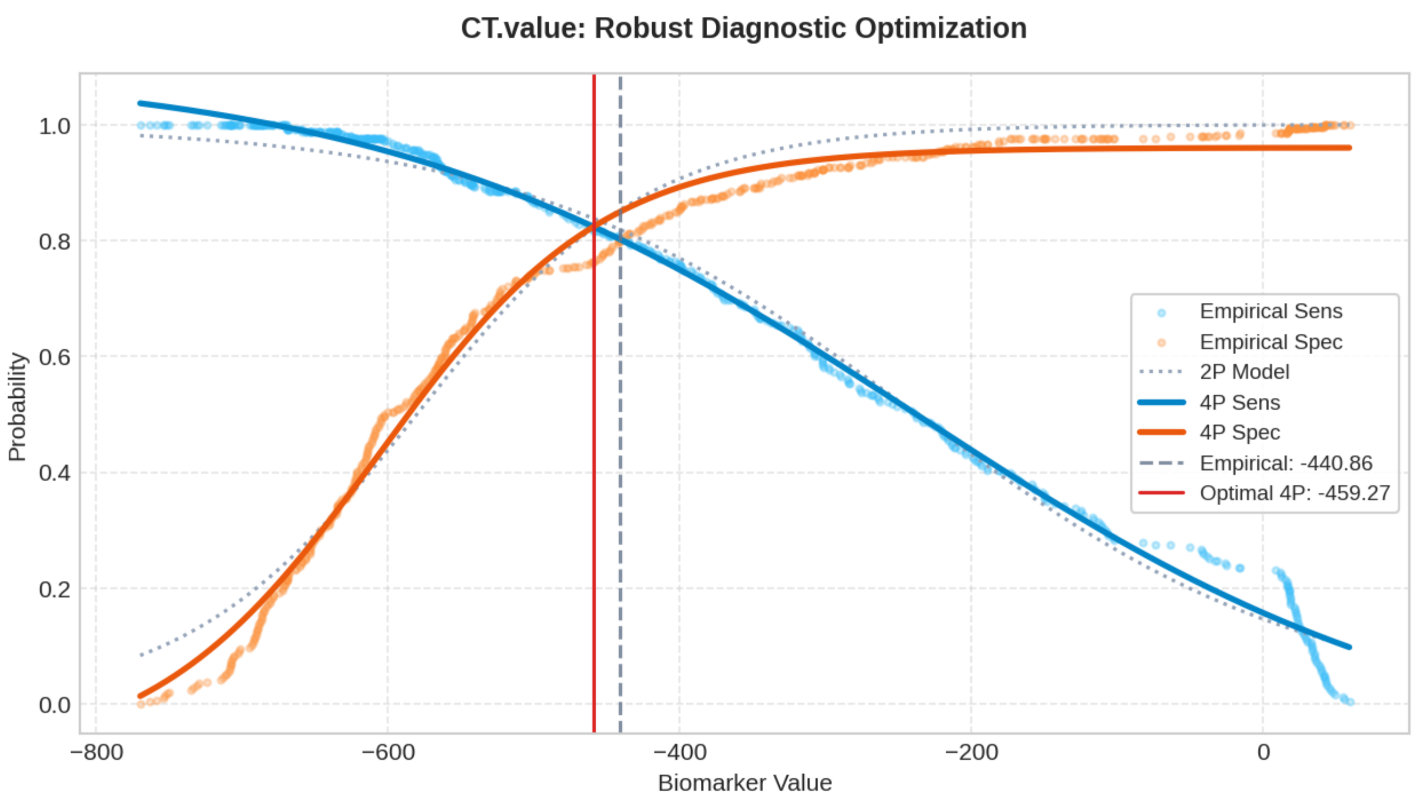

### Biomarker: CTR

Processed: 03-Apr-2026 21:14

#### 1. Optimization Results

| MODEL | CUT-OFF | TRAIN (SE/SP) | VAL (SE/SP) | TEST (SE/SP) | R2 SCORE |
|---|---|---|---|---|---|
| Empirical (Exact) | 0.2164 | 0.816 / 0.816 | 0.930 / 0.774 | 0.791 / 0.806 | N/A |
| Logistic 2-Parameter | 0.2044 | 0.806 / 0.806 | 0.930 / 0.774 | 0.802 / 0.785 | 0.8234 |
| **Logistic 4-Parameter (Rec.)** | **0.1646** | **0.838 / 0.838** | **0.942 / 0.753** | **0.826 / 0.731** | **0.8851** |
| ThresholdXpert (Stochastic) | 0.2144 | 0.813 / 0.822 | 0.930 / 0.774 | 0.791 / 0.806 | N/A |

#### 2. Diagnostic Performance Curves (Training)

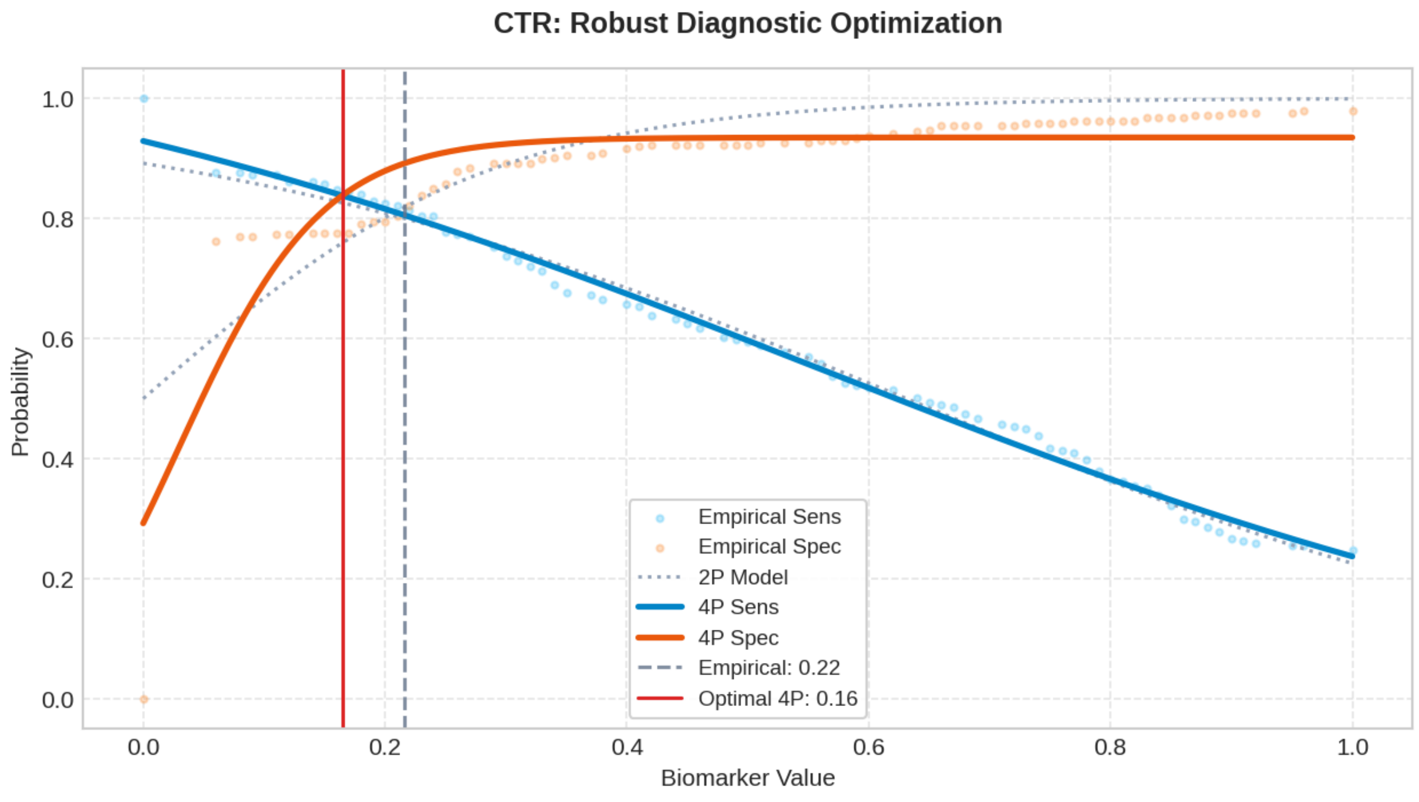

### Biomarker: CEA

Processed: 03-Apr-2026 21:16

#### 1. Optimization Results

| MODEL | CUT-OFF | TRAIN (SE/SP) | VAL (SE/SP) | TEST (SE/SP) | R2 SCORE |
|---|---|---|---|---|---|
| Empirical (Exact) | 2.2670 | 0.575 / 0.575 | 0.651 / 0.613 | 0.581 / 0.581 | N/A |
| Logistic 2-Parameter | 2.3466 | 0.605 / 0.605 | 0.616 / 0.634 | 0.570 / 0.591 | 0.9755 |
| **Logistic 4-Parameter (Rec.)** | **2.2659** | **0.593 / 0.593** | **0.651 / 0.613** | **0.581 / 0.581** | **0.9952** |
| ThresholdXpert (Stochastic) | 2.2257 | 0.590 / 0.587 | 0.651 / 0.602 | 0.593 / 0.570 | N/A |

#### 2. Diagnostic Performance Curves (Training)

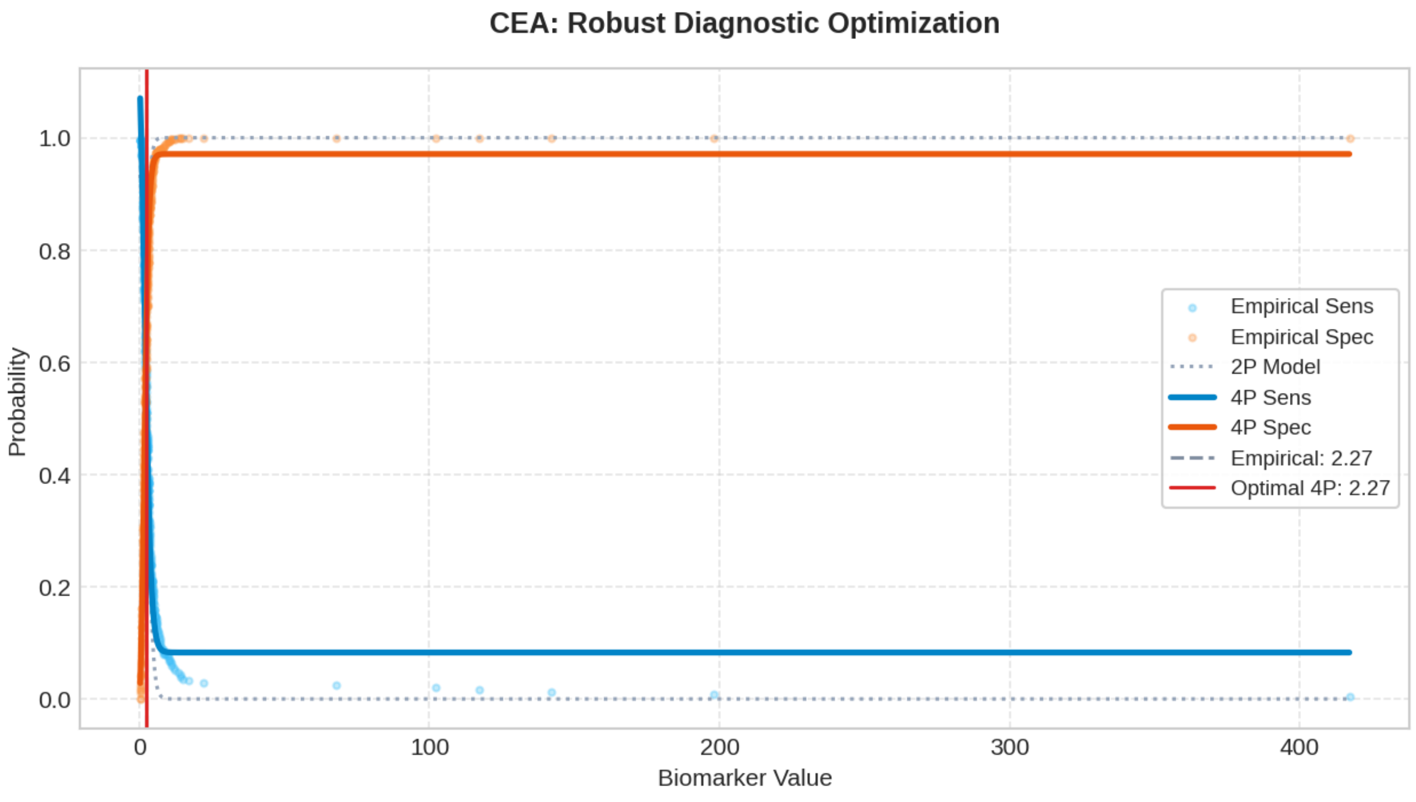

### Biomarker: CYFRA21-1

Processed: 03-Apr-2026 21:18

#### 1. Optimization Results

| MODEL | CUT-OFF | TRAIN (SE/SP) | VAL (SE/SP) | TEST (SE/SP) | R2 SCORE |
|---|---|---|---|---|---|
| Empirical (Exact) | 1.9864 | 0.539 / 0.539 | 0.558 / 0.484 | 0.674 / 0.591 | N/A |
| Logistic 2-Parameter | 2.0142 | 0.542 / 0.542 | 0.558 / 0.495 | 0.651 / 0.591 | 0.9864 |
| **Logistic 4-Parameter (Rec.)** | **1.9707** | **0.542 / 0.542** | **0.558 / 0.473** | **0.674 / 0.591** | **0.9965** |
| ThresholdXpert (Stochastic) | 1.9756 | 0.542 / 0.542 | 0.558 / 0.473 | 0.674 / 0.591 | N/A |

#### 2. Diagnostic Performance Curves (Training)

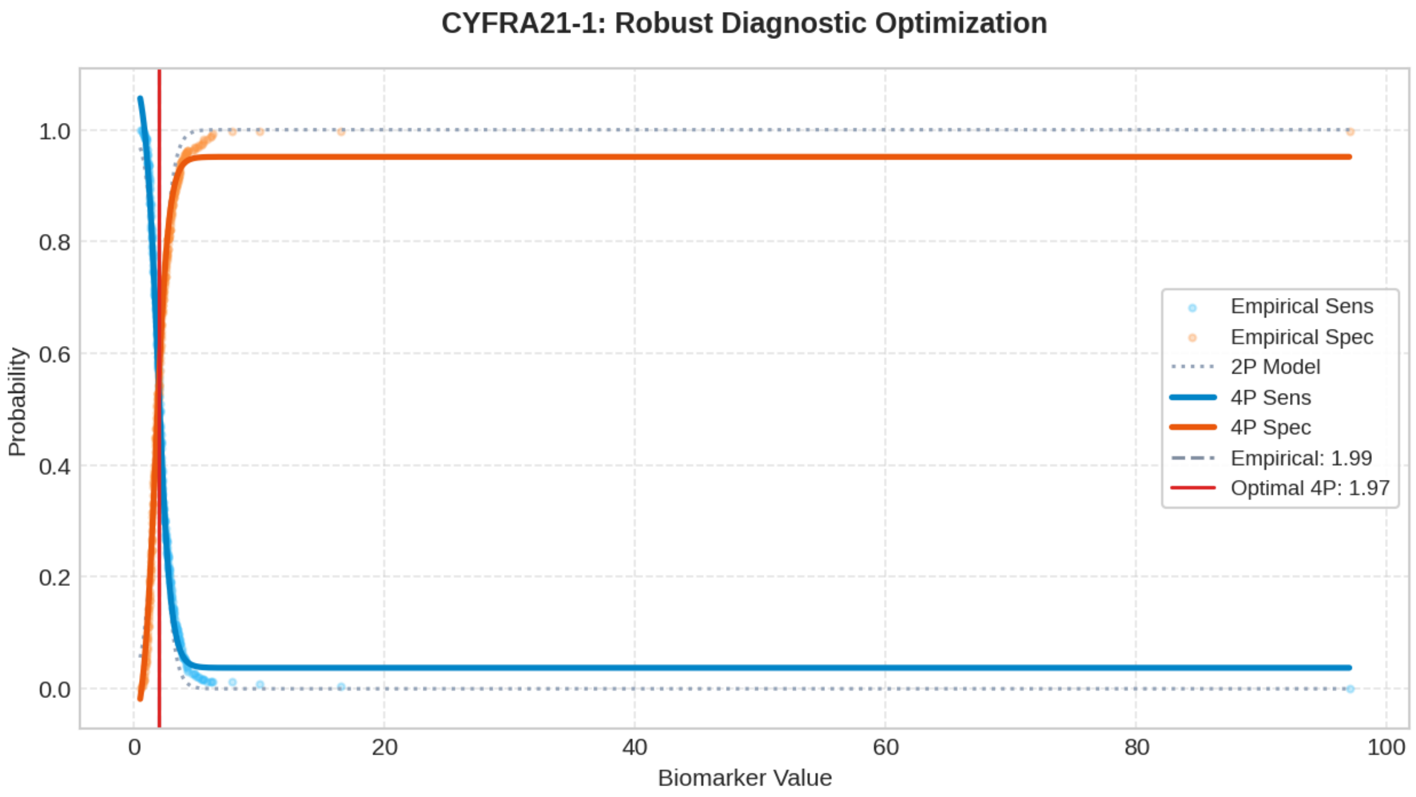

### Biomarker: SCC

Processed: 03-Apr-2026 21:20

#### 1. Optimization Results

| MODEL | CUT-OFF | TRAIN (SE/SP) | VAL (SE/SP) | TEST (SE/SP) | R2 SCORE |
|---|---|---|---|---|---|
| Empirical (Exact) | 0.9400 | 0.486 / 0.486 | 0.407 / 0.473 | 0.558 / 0.559 | N/A |
| Logistic 2-Parameter | 0.9887 | 0.489 / 0.489 | 0.395 / 0.495 | 0.547 / 0.581 | 0.9883 |
| **Logistic 4-Parameter (Rec.)** | **0.9629** | **0.481 / 0.481** | **0.395 / 0.484** | **0.547 / 0.570** | **0.9977** |
| ThresholdXpert (Stochastic) | 0.9357 | 0.486 / 0.486 | 0.407 / 0.452 | 0.570 / 0.548 | N/A |

#### 2. Diagnostic Performance Curves (Training)

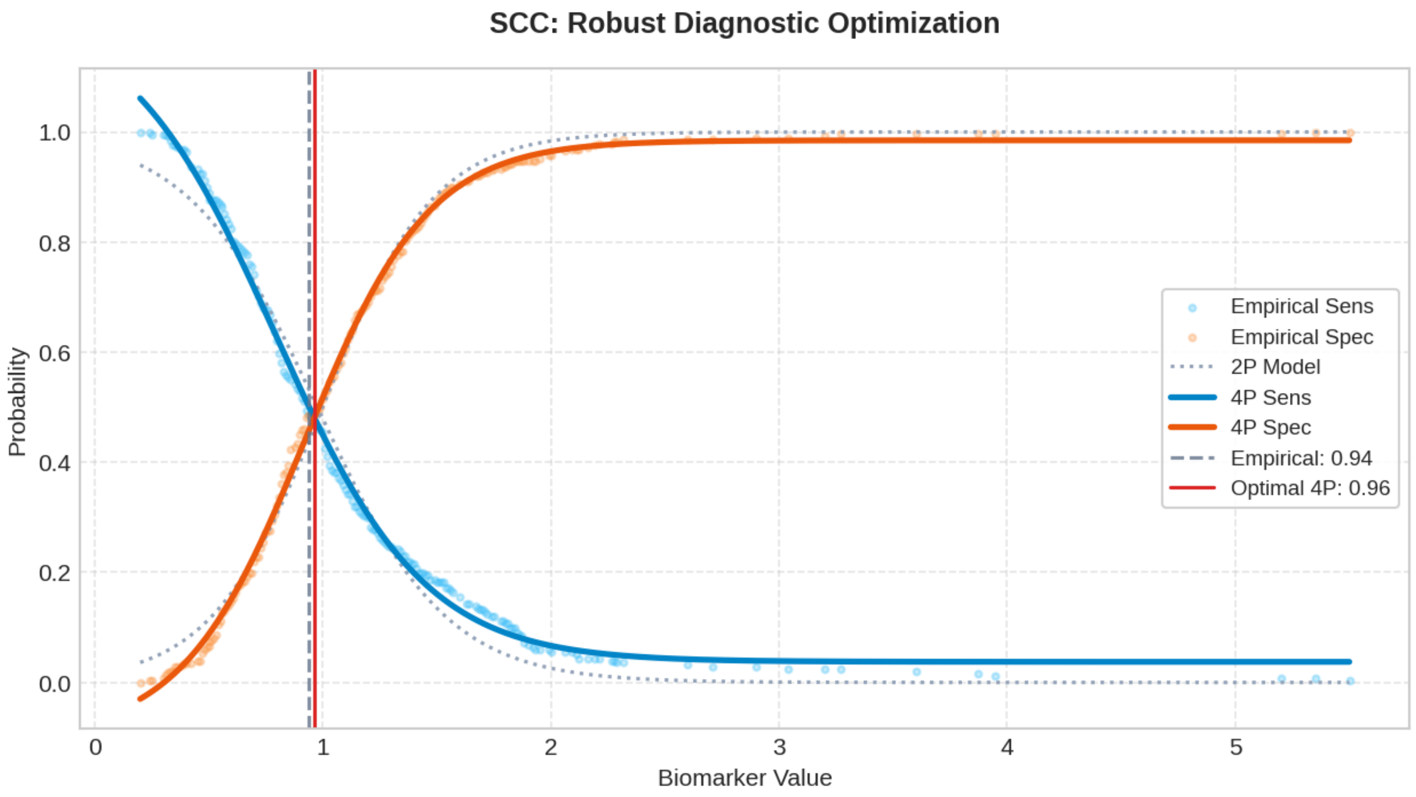

### Biomarker: ProGRP

Processed: 03-Apr-2026 21:23

#### 1. Optimization Results

| MODEL | CUT-OFF | TRAIN (SE/SP) | VAL (SE/SP) | TEST (SE/SP) | R2 SCORE |
|---|---|---|---|---|---|
| Empirical (Exact) | 41.3894 | 0.473 / 0.473 | 0.500 / 0.538 | 0.581 / 0.548 | N/A |
| Logistic 2-Parameter | 41.7453 | 0.482 / 0.482 | 0.465 / 0.538 | 0.570 / 0.591 | 0.9937 |
| **Logistic 4-Parameter (Rec.)** | **41.3501** | **0.479 / 0.479** | **0.500 / 0.527** | **0.581 / 0.548** | **0.9984** |
| ThresholdXpert (Stochastic) | 41.2625 | 0.474 / 0.472 | 0.500 / 0.527 | 0.581 / 0.538 | N/A |

#### 2. Diagnostic Performance Curves (Training)

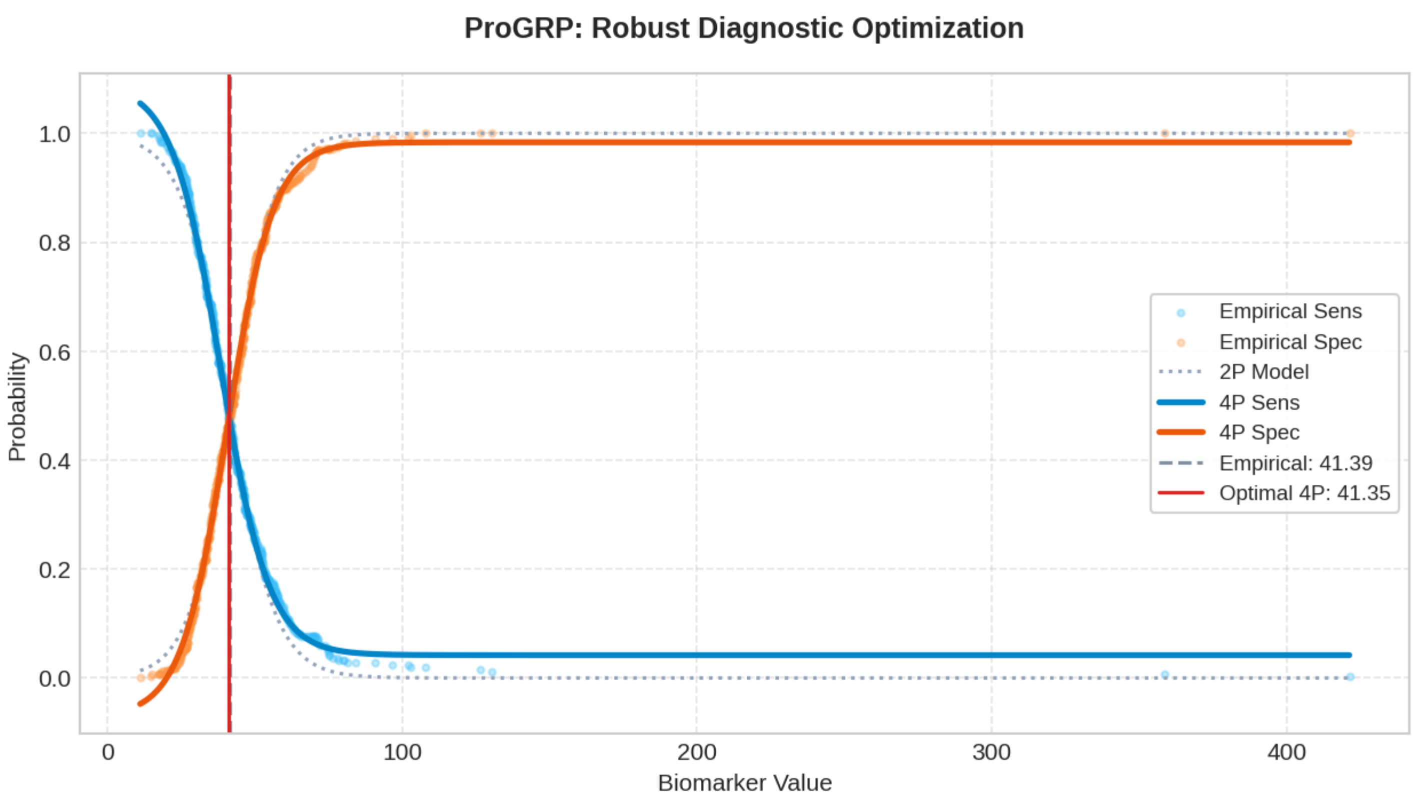

### Biomarker: NSE

Processed: 03-Apr-2026 21:26

#### 1. Optimization Results

| MODEL | CUT-OFF | TRAIN (SE/SP) | VAL (SE/SP) | TEST (SE/SP) | R2 SCORE |
|---|---|---|---|---|---|
| Empirical (Exact) | 18.8496 | 0.502 / 0.502 | 0.488 / 0.516 | 0.477 / 0.559 | N/A |
| Logistic 2-Parameter | 19.2693 | 0.486 / 0.486 | 0.430 / 0.548 | 0.442 / 0.591 | 0.9904 |
| **Logistic 4-Parameter (Rec.)** | **19.0216** | **0.489 / 0.489** | **0.442 / 0.516** | **0.465 / 0.581** | **0.9969** |
| ThresholdXpert (Stochastic) | 18.7055 | 0.510 / 0.500 | 0.500 / 0.516 | 0.488 / 0.548 | N/A |

#### 2. Diagnostic Performance Curves (Training)

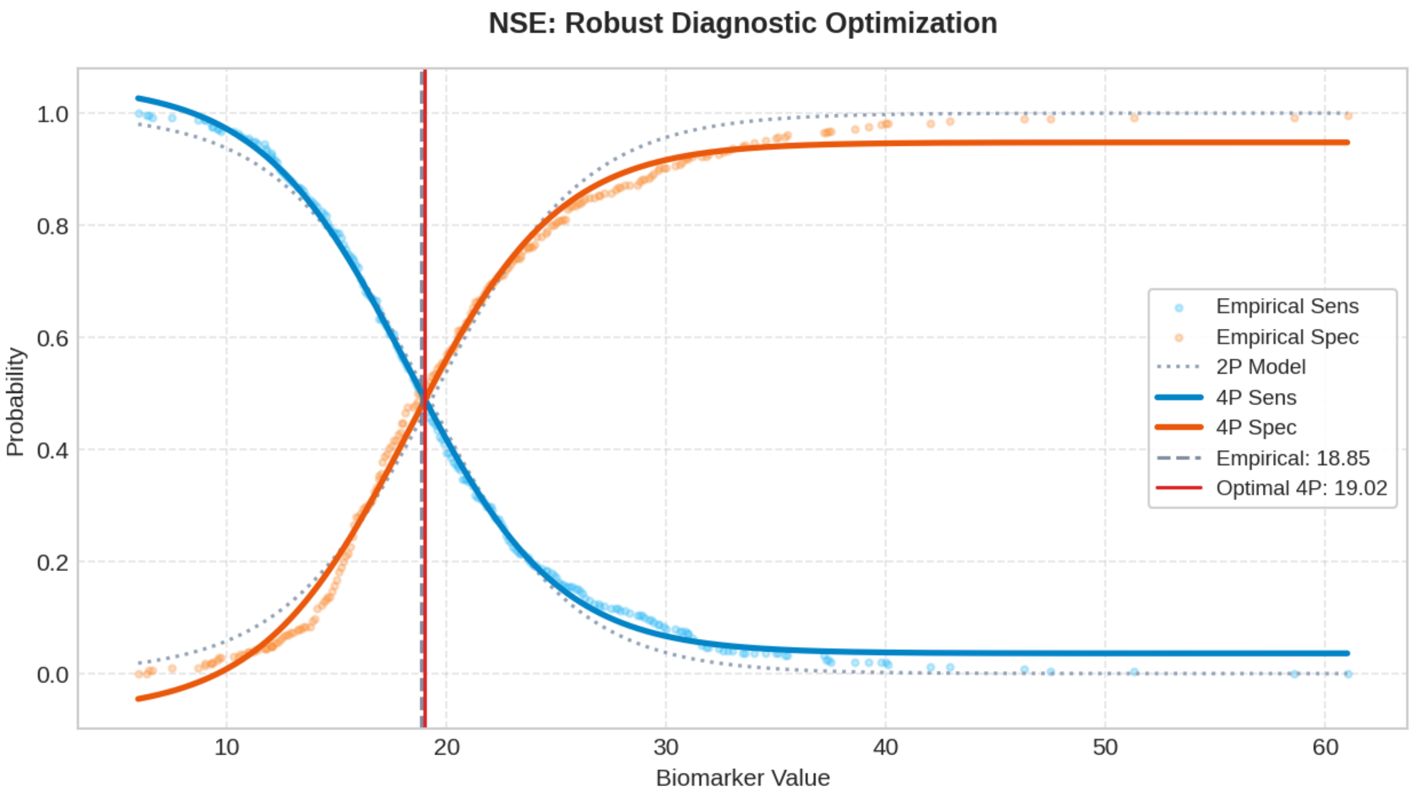

#### Top 200 Combinatorial Panels (ThresholdXpert OR-Logic)

The following multimarker panels have been optimized using high-performance vector-driven Monte Carlo simulations under a Boolean OR-logic framework. The engine employs a Max-Min Balancing logic (0.001 precision) to identify global threshold configurations that maximize the equilibrium between Sensitivity and Specificity across up to 10 million iterations. To ensure clinical robustness, results are sorted strictly by Validation Performance. (* Asterisk indicates an algorithmic threshold instability > 15%, suggesting potential data sparsity or high variance in the stochastic averaging process).

##### #1: Age + BMI + Nodule Diameter + CT.value + CYFRA21-1* + SCC + NSE

Optimized Thresholds: Age: 75.8507 | BMI: 38.7805 | Nodule Diameter: 21.2831 | CT.value: -402.8966 | CYFRA21-1*: 38.5207 | SCC: 4.9115 | NSE: 57.0306

Train Sens: 0.801 | Train Spec: 0.811 [TRAIN SCORE: 1.612] || Val Sens: 0.907 | Val Spec: 0.817 [VAL SCORE: 1.724] || Test Sens: 0.721 | Test Spec: 0.817 [TEST SCORE: 1.538]

##### #2: Age + BMI + Nodule Diameter + CT.value + CEA* + SCC + NSE

Optimized Thresholds: Age: 75.8507 | BMI: 38.7805 | Nodule Diameter: 21.2831 | CT.value: -402.8966 | CEA*: 164.5088 | SCC: 4.9115 | NSE: 57.0306

Train Sens: 0.801 | Train Spec: 0.811 [TRAIN SCORE: 1.612] || Val Sens: 0.907 | Val Spec: 0.817 [VAL SCORE: 1.724] || Test Sens: 0.721 | Test Spec: 0.817 [TEST SCORE: 1.538]

##### #3: BMI + Nodule Diameter + CTR + CYFRA21-1* + ProGRP

Optimized Thresholds: BMI: 38.7625 | Nodule Diameter: 23.4877 | CTR: 0.2359 | CYFRA21-1*: 43.9635 | ProGRP: 264.2898

Train Sens: 0.825 | Train Spec: 0.829 [TRAIN SCORE: 1.653] || Val Sens: 0.930 | Val Spec: 0.785 [VAL SCORE: 1.715] || Test Sens: 0.779 | Test Spec: 0.796 [TEST SCORE: 1.575]

##### #4: BMI + Nodule Diameter + CTR

Optimized Thresholds: BMI: 38.8794 | Nodule Diameter: 23.5535 | CTR: 0.2334

Train Sens: 0.825 | Train Spec: 0.832 [TRAIN SCORE: 1.657] || Val Sens: 0.930 | Val Spec: 0.785 [VAL SCORE: 1.715] || Test Sens: 0.779 | Test Spec: 0.796 [TEST SCORE: 1.575]

##### #5: Age + Nodule Diameter + CT.value + CTR + CEA* + ProGRP*

Optimized Thresholds: Age: 76.9567 | Nodule Diameter: 23.5744 | CT.value: -297.7185 | CTR: 0.2372 | CEA*: 269.8499 | ProGRP*: 293.9106

Train Sens: 0.833 | Train Spec: 0.832 [TRAIN SCORE: 1.665] || Val Sens: 0.930 | Val Spec: 0.785 [VAL SCORE: 1.715] || Test Sens: 0.779 | Test Spec: 0.785 [TEST SCORE: 1.564]

##### #6: BMI + Nodule Diameter + CT.value + CTR + CEA*

Optimized Thresholds: BMI: 38.5921 | Nodule Diameter: 23.4929 | CT.value: -295.5174 | CTR: 0.2344 | CEA*: 168.5009

Train Sens: 0.833 | Train Spec: 0.832 [TRAIN SCORE: 1.665] || Val Sens: 0.930 | Val Spec: 0.785 [VAL SCORE: 1.715] || Test Sens: 0.779 | Test Spec: 0.785 [TEST SCORE: 1.564]

##### #7: Age + BMI + Nodule Diameter + CT.value + CTR + CEA* + ProGRP*

Optimized Thresholds: Age: 78.3235 | BMI: 38.6463 | Nodule Diameter: 23.2980 | CT.value: -297.5549 | CTR: 0.2339 | CEA*: 116.8063 | ProGRP*: 221.7999

Train Sens: 0.833 | Train Spec: 0.832 [TRAIN SCORE: 1.665] || Val Sens: 0.930 | Val Spec: 0.785 [VAL SCORE: 1.715] || Test

Sens: 0.779 | Test Spec: 0.785 [TEST SCORE: 1.564]

###### #8: BMI + Nodule Diameter + CT.value + CTR + CEA* + ProGRP*

Optimized Thresholds: BMI: 38.9217 | Nodule Diameter: 23.4463 | CT.value: -297.6593 | CTR: 0.2363 | CEA*: 208.7685 | ProGRP*: 258.6997

Train Sens: 0.833 | Train Spec: 0.832 [TRAIN SCORE: 1.665] || Val Sens: 0.930 | Val Spec: 0.785 [VAL SCORE: 1.715] || Test Sens: 0.779 | Test Spec: 0.785 [TEST SCORE: 1.564]

###### #9: Nodule Diameter + CTR + CEA* + NSE

Optimized Thresholds: Nodule Diameter: 23.6318 | CTR: 0.2354 | CEA*: 208.8746 | NSE: 59.6171

Train Sens: 0.825 | Train Spec: 0.829 [TRAIN SCORE: 1.653] || Val Sens: 0.930 | Val Spec: 0.785 [VAL SCORE: 1.715] || Test Sens: 0.779 | Test Spec: 0.796 [TEST SCORE: 1.575]

###### #10: Nodule Diameter + CTR + CYFRA21-1* + ProGRP*

Optimized Thresholds: Nodule Diameter: 23.6640 | CTR: 0.2345 | CYFRA21-1*: 48.8235 | ProGRP*: 270.2984

Train Sens: 0.825 | Train Spec: 0.829 [TRAIN SCORE: 1.653] || Val Sens: 0.930 | Val Spec: 0.785 [VAL SCORE: 1.715] || Test Sens: 0.779 | Test Spec: 0.796 [TEST SCORE: 1.575]

###### #11: BMI + Nodule Diameter + CTR + CEA* + CYFRA21-1* + ProGRP* + NSE

Optimized Thresholds: BMI: 38.5714 | Nodule Diameter: 23.4137 | CTR: 0.2350 | CEA*: 195.9328 | CYFRA21-1*: 52.1903 | ProGRP*: 263.4385 | NSE: 59.9487

Train Sens: 0.825 | Train Spec: 0.825 [TRAIN SCORE: 1.650] || Val Sens: 0.930 | Val Spec: 0.785 [VAL SCORE: 1.715] || Test Sens: 0.779 | Test Spec: 0.796 [TEST SCORE: 1.575]

###### #12: Nodule Diameter + CTR + CEA* + CYFRA21-1* + ProGRP* + NSE

Optimized Thresholds: Nodule Diameter: 23.4999 | CTR: 0.2344 | CEA*: 187.5053 | CYFRA21-1*: 56.8616 | ProGRP*: 278.1440 | NSE: 59.5292

Train Sens: 0.825 | Train Spec: 0.825 [TRAIN SCORE: 1.650] || Val Sens: 0.930 | Val Spec: 0.785 [VAL SCORE: 1.715] || Test Sens: 0.779 | Test Spec: 0.796 [TEST SCORE: 1.575]

###### #13: BMI + Nodule Diameter + CTR + ProGRP* + NSE

Optimized Thresholds: BMI: 39.1373 | Nodule Diameter: 23.5729 | CTR: 0.2345 | ProGRP*: 256.4631 | NSE: 59.5310

Train Sens: 0.825 | Train Spec: 0.829 [TRAIN SCORE: 1.653] || Val Sens: 0.930 | Val Spec: 0.785 [VAL SCORE: 1.715] || Test Sens: 0.779 | Test Spec: 0.796 [TEST SCORE: 1.575]

###### #14: Nodule Diameter + CTR + ProGRP* + NSE

Optimized Thresholds: Nodule Diameter: 23.6418 | CTR: 0.2352 | ProGRP*: 276.6925 | NSE: 59.9493

Train Sens: 0.825 | Train Spec: 0.829 [TRAIN SCORE: 1.653] || Val Sens: 0.930 | Val Spec: 0.785 [VAL SCORE: 1.715] || Test Sens: 0.779 | Test Spec: 0.796 [TEST SCORE: 1.575]

###### #15: Nodule Diameter + CTR + CEA* + CYFRA21-1* + NSE

Optimized Thresholds: Nodule Diameter: 23.5020 | CTR: 0.2358 | CEA*: 208.7065 | CYFRA21-1*: 60.8660 | NSE: 60.0861

Train Sens: 0.825 | Train Spec: 0.825 [TRAIN SCORE: 1.650] || Val Sens: 0.930 | Val Spec: 0.785 [VAL SCORE: 1.715] || Test Sens: 0.779 | Test Spec: 0.796 [TEST SCORE: 1.575]

##### #16: Age + BMI + Nodule Diameter + CT.value + CTR

Optimized Thresholds: Age: 77.2405 | BMI: 38.9235 | Nodule Diameter: 23.7691 | CT.value: -294.4861 | CTR: 0.2359

Train Sens: 0.833 | Train Spec: 0.832 [TRAIN SCORE: 1.665] || Val Sens: 0.930 | Val Spec: 0.785 [VAL SCORE: 1.715] || Test Sens: 0.779 | Test Spec: 0.785 [TEST SCORE: 1.564]

##### #17: Age + Nodule Diameter + CTR + ProGRP* + NSE

Optimized Thresholds: Age: 77.7791 | Nodule Diameter: 23.6260 | CTR: 0.2352 | ProGRP*: 242.3186 | NSE: 59.6873

Train Sens: 0.825 | Train Spec: 0.829 [TRAIN SCORE: 1.653] || Val Sens: 0.930 | Val Spec: 0.785 [VAL SCORE: 1.715] || Test Sens: 0.779 | Test Spec: 0.796 [TEST SCORE: 1.575]

##### #18: Nodule Diameter + CTR + CYFRA21-1* + ProGRP* + NSE

Optimized Thresholds: Nodule Diameter: 23.4218 | CTR: 0.2345 | CYFRA21-1*: 51.0535 | ProGRP*: 282.8396 | NSE: 60.1498

Train Sens: 0.825 | Train Spec: 0.825 [TRAIN SCORE: 1.650] || Val Sens: 0.930 | Val Spec: 0.785 [VAL SCORE: 1.715] || Test Sens: 0.779 | Test Spec: 0.796 [TEST SCORE: 1.575]

##### #19: Age + Nodule Diameter + CTR

Optimized Thresholds: Age: 76.6611 | Nodule Diameter: 23.5520 | CTR: 0.2350

Train Sens: 0.825 | Train Spec: 0.832 [TRAIN SCORE: 1.657] || Val Sens: 0.930 | Val Spec: 0.785 [VAL SCORE: 1.715] || Test Sens: 0.779 | Test Spec: 0.796 [TEST SCORE: 1.575]

##### #20: Age + Nodule Diameter + CTR + CYFRA21-1* + NSE

Optimized Thresholds: Age: 77.6477 | Nodule Diameter: 23.7013 | CTR: 0.2355 | CYFRA21-1*: 54.5150 | NSE: 59.8234

Train Sens: 0.825 | Train Spec: 0.825 [TRAIN SCORE: 1.650] || Val Sens: 0.930 | Val Spec: 0.785 [VAL SCORE: 1.715] || Test Sens: 0.779 | Test Spec: 0.796 [TEST SCORE: 1.575]

##### #21: Age + Nodule Diameter + CTR + CYFRA21-1* + ProGRP*

Optimized Thresholds: Age: 75.7831 | Nodule Diameter: 23.4957 | CTR: 0.2345 | CYFRA21-1*: 54.7283 | ProGRP*: 256.9869

Train Sens: 0.825 | Train Spec: 0.829 [TRAIN SCORE: 1.653] || Val Sens: 0.930 | Val Spec: 0.785 [VAL SCORE: 1.715] || Test Sens: 0.779 | Test Spec: 0.796 [TEST SCORE: 1.575]

##### #22: Age + Nodule Diameter + CTR + CEA* + CYFRA21-1* + ProGRP* + NSE

Optimized Thresholds: Age: 77.2499 | Nodule Diameter: 23.5205 | CTR: 0.2339 | CEA*: 197.8794 | CYFRA21-1*: 55.1322 | ProGRP*: 253.4116 | NSE: 59.9243

Train Sens: 0.825 | Train Spec: 0.825 [TRAIN SCORE: 1.650] || Val Sens: 0.930 | Val Spec: 0.785 [VAL SCORE: 1.715] || Test Sens: 0.779 | Test Spec: 0.796 [TEST SCORE: 1.575]

##### #23: BMI + Nodule Diameter + CT.value + CTR + ProGRP*

Optimized Thresholds: BMI: 38.5556 | Nodule Diameter: 23.4776 | CT.value: -293.4542 | CTR: 0.2339 | ProGRP*: 251.6365

Train Sens: 0.833 | Train Spec: 0.832 [TRAIN SCORE: 1.665] || Val Sens: 0.930 | Val Spec: 0.785 [VAL SCORE: 1.715] || Test Sens: 0.779 | Test Spec: 0.785 [TEST SCORE: 1.564]

##### #24: BMI + Nodule Diameter + CTR + NSE

Optimized Thresholds: BMI: 38.4692 | Nodule Diameter: 23.5325 | CTR: 0.2369 | NSE: 59.8691

Train Sens: 0.825 | Train Spec: 0.829 [TRAIN SCORE: 1.653] || Val Sens: 0.930 | Val Spec: 0.785 [VAL SCORE: 1.715] || Test Sens: 0.779 | Test Spec: 0.796 [TEST SCORE: 1.575]

##### #25: Nodule Diameter + CTR + CYFRA21-1*

Optimized Thresholds: Nodule Diameter: 23.5648 | CTR: 0.2354 | CYFRA21-1*: 41.3900

Train Sens: 0.825 | Train Spec: 0.829 [TRAIN SCORE: 1.653] || Val Sens: 0.930 | Val Spec: 0.785 [VAL SCORE: 1.715] || Test Sens: 0.779 | Test Spec: 0.796 [TEST SCORE: 1.575]

##### #26: Nodule Diameter + CT.value + CTR + CEA*

Optimized Thresholds: Nodule Diameter: 23.5666 | CT.value: -301.0754 | CTR: 0.2353 | CEA*: 215.1485

Train Sens: 0.833 | Train Spec: 0.832 [TRAIN SCORE: 1.665] || Val Sens: 0.930 | Val Spec: 0.785 [VAL SCORE: 1.715] || Test Sens: 0.779 | Test Spec: 0.785 [TEST SCORE: 1.564]

##### #27: Age + Nodule Diameter + CT.value + CTR

Optimized Thresholds: Age: 77.7392 | Nodule Diameter: 23.6525 | CT.value: -302.1738 | CTR: 0.2341

Train Sens: 0.833 | Train Spec: 0.832 [TRAIN SCORE: 1.665] || Val Sens: 0.930 | Val Spec: 0.785 [VAL SCORE: 1.715] || Test Sens: 0.779 | Test Spec: 0.785 [TEST SCORE: 1.564]

##### #28: Age + BMI + Nodule Diameter + CTR + CEA* + CYFRA21-1* + ProGRP*

Optimized Thresholds: Age: 76.8186 | BMI: 39.0693 | Nodule Diameter: 23.5867 | CTR: 0.2357 | CEA*: 248.5382 | CYFRA21-1*: 58.1197 | ProGRP*: 285.5726

Train Sens: 0.825 | Train Spec: 0.829 [TRAIN SCORE: 1.653] || Val Sens: 0.930 | Val Spec: 0.785 [VAL SCORE: 1.715] || Test Sens: 0.779 | Test Spec: 0.796 [TEST SCORE: 1.575]

##### #29: Age + BMI + Nodule Diameter + CTR + CEA + CYFRA21-1 + NSE

Optimized Thresholds: Age: 75.2145 | BMI: 39.7267 | Nodule Diameter: 23.8040 | CTR: 0.2364 | CEA: 27.0915 | CYFRA21-1: 60.0304 | NSE: 60.8253

Train Sens: 0.825 | Train Spec: 0.825 [TRAIN SCORE: 1.650] || Val Sens: 0.930 | Val Spec: 0.785 [VAL SCORE: 1.715] || Test Sens: 0.779 | Test Spec: 0.796 [TEST SCORE: 1.575]

##### #30: Age + Nodule Diameter + CTR + CYFRA21-1* + ProGRP + NSE

Optimized Thresholds: Age: 76.6651 | Nodule Diameter: 23.6128 | CTR: 0.2348 | CYFRA21-1*: 49.4417 | ProGRP: 243.2088 | NSE: 59.7720

Train Sens: 0.825 | Train Spec: 0.825 [TRAIN SCORE: 1.650] || Val Sens: 0.930 | Val Spec: 0.785 [VAL SCORE: 1.715] || Test Sens: 0.779 | Test Spec: 0.796 [TEST SCORE: 1.575]

##### #31: Nodule Diameter + CTR + NSE

Optimized Thresholds: Nodule Diameter: 23.4887 | CTR: 0.2344 | NSE: 59.7937

Train Sens: 0.825 | Train Spec: 0.829 [TRAIN SCORE: 1.653] || Val Sens: 0.930 | Val Spec: 0.785 [VAL SCORE: 1.715] || Test Sens: 0.779 | Test Spec: 0.796 [TEST SCORE: 1.575]

##### #32: Nodule Diameter + CTR + ProGRP*

Optimized Thresholds: Nodule Diameter: 23.4294 | CTR: 0.2357 | ProGRP*: 289.4592

Train Sens: 0.825 | Train Spec: 0.832 [TRAIN SCORE: 1.657] || Val Sens: 0.930 | Val Spec: 0.785 [VAL SCORE: 1.715] || Test Sens: 0.779 | Test Spec: 0.796 [TEST SCORE: 1.575]

##### #33: BMI + Nodule Diameter + CTR + CYFRA21-1* + NSE

Optimized Thresholds: BMI: 39.1533 | Nodule Diameter: 23.7506 | CTR: 0.2360 | CYFRA21-1*: 61.8578 | NSE: 59.5812

Train Sens: 0.825 | Train Spec: 0.825 [TRAIN SCORE: 1.650] || Val Sens: 0.930 | Val Spec: 0.785 [VAL SCORE: 1.715] || Test Sens: 0.779 | Test Spec: 0.796 [TEST SCORE: 1.575]

###### #34: Age + BMI + Nodule Diameter + CTR + CEA + ProGRP + NSE

Optimized Thresholds: Age: 75.2145 | BMI: 39.7267 | Nodule Diameter: 23.8040 | CTR: 0.2364 | CEA: 27.0915 | ProGRP: 264.1889 | NSE: 60.8253

Train Sens: 0.825 | Train Spec: 0.829 [TRAIN SCORE: 1.653] || Val Sens: 0.930 | Val Spec: 0.785 [VAL SCORE: 1.715] || Test Sens: 0.779 | Test Spec: 0.796 [TEST SCORE: 1.575]

###### #35: Age + Nodule Diameter + CTR + CEA* + CYFRA21-1* + NSE

Optimized Thresholds: Age: 76.9643 | Nodule Diameter: 23.5563 | CTR: 0.2345 | CEA*: 235.6567 | CYFRA21-1*: 48.4851 | NSE: 59.6417

Train Sens: 0.825 | Train Spec: 0.825 [TRAIN SCORE: 1.650] || Val Sens: 0.930 | Val Spec: 0.785 [VAL SCORE: 1.715] || Test Sens: 0.779 | Test Spec: 0.796 [TEST SCORE: 1.575]

###### #36: Age + Nodule Diameter + CTR + CEA* + ProGRP + NSE

Optimized Thresholds: Age: 76.6651 | Nodule Diameter: 23.6128 | CTR: 0.2348 | CEA*: 211.6616 | ProGRP: 243.2088 | NSE: 59.7720

Train Sens: 0.825 | Train Spec: 0.829 [TRAIN SCORE: 1.653] || Val Sens: 0.930 | Val Spec: 0.785 [VAL SCORE: 1.715] || Test Sens: 0.779 | Test Spec: 0.796 [TEST SCORE: 1.575]

###### #37: Age + Nodule Diameter + CTR + CEA*

Optimized Thresholds: Age: 77.4152 | Nodule Diameter: 23.6056 | CTR: 0.2345 | CEA*: 234.9320

Train Sens: 0.825 | Train Spec: 0.832 [TRAIN SCORE: 1.657] || Val Sens: 0.930 | Val Spec: 0.785 [VAL SCORE: 1.715] || Test Sens: 0.779 | Test Spec: 0.796 [TEST SCORE: 1.575]

###### #38: Age + Nodule Diameter + CTR + CYFRA21-1*

Optimized Thresholds: Age: 77.3822 | Nodule Diameter: 23.6234 | CTR: 0.2350 | CYFRA21-1*: 53.5147

Train Sens: 0.825 | Train Spec: 0.829 [TRAIN SCORE: 1.653] || Val Sens: 0.930 | Val Spec: 0.785 [VAL SCORE: 1.715] || Test Sens: 0.779 | Test Spec: 0.796 [TEST SCORE: 1.575]

###### #39: Nodule Diameter + CT.value + CTR

Optimized Thresholds: Nodule Diameter: 23.4267 | CT.value: -303.3285 | CTR: 0.2354

Train Sens: 0.833 | Train Spec: 0.832 [TRAIN SCORE: 1.665] || Val Sens: 0.930 | Val Spec: 0.785 [VAL SCORE: 1.715] || Test Sens: 0.779 | Test Spec: 0.785 [TEST SCORE: 1.564]

###### #40: BMI + Nodule Diameter + CTR + CEA* + NSE

Optimized Thresholds: BMI: 38.9598 | Nodule Diameter: 23.7205 | CTR: 0.2359 | CEA*: 239.2627 | NSE: 59.6865

Train Sens: 0.825 | Train Spec: 0.829 [TRAIN SCORE: 1.653] || Val Sens: 0.930 | Val Spec: 0.785 [VAL SCORE: 1.715] || Test Sens: 0.779 | Test Spec: 0.796 [TEST SCORE: 1.575]

###### #41: Nodule Diameter + CT.value + CTR + ProGRP*

Optimized Thresholds: Nodule Diameter: 23.5928 | CT.value: -293.5507 | CTR: 0.2343 | ProGRP*: 233.8361

Train Sens: 0.833 | Train Spec: 0.832 [TRAIN SCORE: 1.665] || Val Sens: 0.930 | Val Spec: 0.785 [VAL SCORE: 1.715] || Test Sens: 0.779 | Test Spec: 0.785 [TEST SCORE: 1.564]

##### #42: Age + Nodule Diameter + CTR + ProGRP*

Optimized Thresholds: Age: 76.9189 | Nodule Diameter: 23.3962 | CTR: 0.2357 | ProGRP*: 270.1808

Train Sens: 0.825 | Train Spec: 0.832 [TRAIN SCORE: 1.657] || Val Sens: 0.930 | Val Spec: 0.785 [VAL SCORE: 1.715] || Test Sens: 0.779 | Test Spec: 0.796 [TEST SCORE: 1.575]

##### #43: Age + Nodule Diameter + CTR + NSE

Optimized Thresholds: Age: 77.2301 | Nodule Diameter: 23.4836 | CTR: 0.2375 | NSE: 59.7618

Train Sens: 0.825 | Train Spec: 0.829 [TRAIN SCORE: 1.653] || Val Sens: 0.930 | Val Spec: 0.785 [VAL SCORE: 1.715] || Test Sens: 0.779 | Test Spec: 0.796 [TEST SCORE: 1.575]

##### #44: Age + Nodule Diameter + CTR + CEA* + NSE

Optimized Thresholds: Age: 77.9369 | Nodule Diameter: 23.5712 | CTR: 0.2343 | CEA*: 211.0228 | NSE: 59.7783

Train Sens: 0.825 | Train Spec: 0.829 [TRAIN SCORE: 1.653] || Val Sens: 0.930 | Val Spec: 0.785 [VAL SCORE: 1.715] || Test Sens: 0.779 | Test Spec: 0.796 [TEST SCORE: 1.575]

##### #45: Nodule Diameter + CTR + CYFRA21-1* + NSE

Optimized Thresholds: Nodule Diameter: 23.5083 | CTR: 0.2356 | CYFRA21-1*: 40.3586 | NSE: 59.9065

Train Sens: 0.825 | Train Spec: 0.825 [TRAIN SCORE: 1.650] || Val Sens: 0.930 | Val Spec: 0.785 [VAL SCORE: 1.715] || Test Sens: 0.779 | Test Spec: 0.796 [TEST SCORE: 1.575]

##### #46: BMI + Nodule Diameter + CTR + CEA* + CYFRA21-1* + NSE

Optimized Thresholds: BMI: 38.4718 | Nodule Diameter: 23.5379 | CTR: 0.2337 | CEA*: 262.4081 | CYFRA21-1*: 47.1402 | NSE: 59.6187

Train Sens: 0.825 | Train Spec: 0.825 [TRAIN SCORE: 1.650] || Val Sens: 0.930 | Val Spec: 0.785 [VAL SCORE: 1.715] || Test Sens: 0.779 | Test Spec: 0.796 [TEST SCORE: 1.575]

##### #47: Age + BMI + Nodule Diameter + CTR + ProGRP + NSE

Optimized Thresholds: Age: 77.1054 | BMI: 39.1947 | Nodule Diameter: 23.2849 | CTR: 0.2368 | ProGRP: 282.9563 | NSE: 60.0323

Train Sens: 0.825 | Train Spec: 0.829 [TRAIN SCORE: 1.653] || Val Sens: 0.930 | Val Spec: 0.785 [VAL SCORE: 1.715] || Test Sens: 0.779 | Test Spec: 0.796 [TEST SCORE: 1.575]

##### #48: Nodule Diameter + CTR

Optimized Thresholds: Nodule Diameter: 23.5428 | CTR: 0.2343

Train Sens: 0.825 | Train Spec: 0.832 [TRAIN SCORE: 1.657] || Val Sens: 0.930 | Val Spec: 0.785 [VAL SCORE: 1.715] || Test Sens: 0.779 | Test Spec: 0.796 [TEST SCORE: 1.575]

##### #49: BMI + Nodule Diameter + CTR + CEA* + ProGRP + NSE

Optimized Thresholds: BMI: 38.4370 | Nodule Diameter: 23.6254 | CTR: 0.2343 | CEA*: 238.4979 | ProGRP: 254.3901 | NSE: 59.7666

Train Sens: 0.825 | Train Spec: 0.829 [TRAIN SCORE: 1.653] || Val Sens: 0.930 | Val Spec: 0.785 [VAL SCORE: 1.715] || Test Sens: 0.779 | Test Spec: 0.796 [TEST SCORE: 1.575]

##### #50: Age + BMI + Nodule Diameter + CTR + CYFRA21-1 + NSE

Optimized Thresholds: Age: 77.1054 | BMI: 39.1947 | Nodule Diameter: 23.2849 | CTR: 0.2368 | CYFRA21-1: 64.4466 | NSE: 60.0323

Train Sens: 0.825 | Train Spec: 0.825 [TRAIN SCORE: 1.650] || Val Sens: 0.930 | Val Spec: 0.785 [VAL SCORE: 1.715] || Test Sens: 0.779 | Test Spec: 0.796 [TEST SCORE: 1.575]

###### #51: Age + BMI + Nodule Diameter + CTR + CEA* + ProGRP*

Optimized Thresholds: Age: 76.6482 | BMI: 38.6101 | Nodule Diameter: 23.5509 | CTR: 0.2352 | CEA*: 225.0441 | ProGRP*: 244.8072

Train Sens: 0.825 | Train Spec: 0.832 [TRAIN SCORE: 1.657] || Val Sens: 0.930 | Val Spec: 0.785 [VAL SCORE: 1.715] || Test Sens: 0.779 | Test Spec: 0.796 [TEST SCORE: 1.575]

###### #52: Age + BMI + Nodule Diameter + CTR + CYFRA21-1* + ProGRP*

Optimized Thresholds: Age: 77.0439 | BMI: 38.7299 | Nodule Diameter: 23.4913 | CTR: 0.2350 | CYFRA21-1*: 45.5448 | ProGRP*: 254.2532

Train Sens: 0.825 | Train Spec: 0.829 [TRAIN SCORE: 1.653] || Val Sens: 0.930 | Val Spec: 0.785 [VAL SCORE: 1.715] || Test Sens: 0.779 | Test Spec: 0.796 [TEST SCORE: 1.575]

###### #53: BMI + Nodule Diameter + CTR + ProGRP*

Optimized Thresholds: BMI: 38.7399 | Nodule Diameter: 23.4088 | CTR: 0.2359 | ProGRP*: 211.0332

Train Sens: 0.825 | Train Spec: 0.832 [TRAIN SCORE: 1.657] || Val Sens: 0.930 | Val Spec: 0.785 [VAL SCORE: 1.715] || Test Sens: 0.779 | Test Spec: 0.796 [TEST SCORE: 1.575]

###### #54: Age + Nodule Diameter + CT.value + CTR + CEA*

Optimized Thresholds: Age: 76.4947 | Nodule Diameter: 23.7474 | CT.value: -303.7885 | CTR: 0.2344 | CEA*: 230.2776

Train Sens: 0.833 | Train Spec: 0.832 [TRAIN SCORE: 1.665] || Val Sens: 0.930 | Val Spec: 0.785 [VAL SCORE: 1.715] || Test Sens: 0.779 | Test Spec: 0.785 [TEST SCORE: 1.564]

###### #55: Age + BMI + Nodule Diameter + CTR + CEA* + CYFRA21-1*

Optimized Thresholds: Age: 75.9311 | BMI: 38.8286 | Nodule Diameter: 23.5368 | CTR: 0.2359 | CEA*: 258.8094 | CYFRA21-1*: 53.4993

Train Sens: 0.825 | Train Spec: 0.829 [TRAIN SCORE: 1.653] || Val Sens: 0.930 | Val Spec: 0.785 [VAL SCORE: 1.715] || Test Sens: 0.779 | Test Spec: 0.796 [TEST SCORE: 1.575]

###### #56: Age + BMI + Nodule Diameter + CTR + CEA + CYFRA21-1 + ProGRP + NSE

Optimized Thresholds: Age: 76.2860 | BMI: 39.0764 | Nodule Diameter: 23.4906 | CTR: 0.2349 | CEA: 367.1128 | CYFRA21-1: 61.0056 | ProGRP: 192.1620 | NSE: 59.6230

Train Sens: 0.825 | Train Spec: 0.825 [TRAIN SCORE: 1.650] || Val Sens: 0.930 | Val Spec: 0.785 [VAL SCORE: 1.715] || Test Sens: 0.779 | Test Spec: 0.796 [TEST SCORE: 1.575]

###### #57: Nodule Diameter + CT.value + CTR + CEA* + ProGRP

Optimized Thresholds: Nodule Diameter: 23.4260 | CT.value: -293.6584 | CTR: 0.2352 | CEA*: 216.5564 | ProGRP: 222.9988

Train Sens: 0.833 | Train Spec: 0.832 [TRAIN SCORE: 1.665] || Val Sens: 0.930 | Val Spec: 0.785 [VAL SCORE: 1.715] || Test Sens: 0.779 | Test Spec: 0.785 [TEST SCORE: 1.564]

###### #58: BMI + Nodule Diameter + CTR + CYFRA21-1*

Optimized Thresholds: BMI: 38.9517 | Nodule Diameter: 23.5641 | CTR: 0.2357 | CYFRA21-1*: 51.0250

Train Sens: 0.825 | Train Spec: 0.829 [TRAIN SCORE: 1.653] || Val Sens: 0.930 | Val Spec: 0.785 [VAL SCORE: 1.715] || Test

Sens: 0.779 | Test Spec: 0.796 [TEST SCORE: 1.575]

**#59: Age + BMI + Nodule Diameter + CTR + NSE**

Optimized Thresholds: Age: 77.4393 | BMI: 38.7056 | Nodule Diameter: 23.8837 | CTR: 0.2363 | NSE: 59.6602

Train Sens: 0.825 | Train Spec: 0.829 [TRAIN SCORE: 1.653] || Val Sens: 0.930 | Val Spec: 0.785 [VAL SCORE: 1.715] || Test Sens: 0.779 | Test Spec: 0.796 [TEST SCORE: 1.575]

**#60: Age + BMI + Nodule Diameter + CTR + ProGRP***

Optimized Thresholds: Age: 76.6601 | BMI: 38.2769 | Nodule Diameter: 23.4401 | CTR: 0.2354 | ProGRP*: 290.9265

Train Sens: 0.825 | Train Spec: 0.832 [TRAIN SCORE: 1.657] || Val Sens: 0.930 | Val Spec: 0.785 [VAL SCORE: 1.715] || Test Sens: 0.779 | Test Spec: 0.796 [TEST SCORE: 1.575]

**#61: BMI + Nodule Diameter + CT.value + CTR**

Optimized Thresholds: BMI: 39.2994 | Nodule Diameter: 23.6045 | CT.value: -302.4849 | CTR: 0.2342

Train Sens: 0.833 | Train Spec: 0.832 [TRAIN SCORE: 1.665] || Val Sens: 0.930 | Val Spec: 0.785 [VAL SCORE: 1.715] || Test Sens: 0.779 | Test Spec: 0.785 [TEST SCORE: 1.564]

**#62: Age + BMI + Nodule Diameter + CTR**

Optimized Thresholds: Age: 76.9932 | BMI: 38.6034 | Nodule Diameter: 23.6150 | CTR: 0.2358

Train Sens: 0.825 | Train Spec: 0.832 [TRAIN SCORE: 1.657] || Val Sens: 0.930 | Val Spec: 0.785 [VAL SCORE: 1.715] || Test Sens: 0.779 | Test Spec: 0.796 [TEST SCORE: 1.575]

**#63: Age + Nodule Diameter + CT.value + CTR + ProGRP***

Optimized Thresholds: Age: 76.8363 | Nodule Diameter: 23.4423 | CT.value: -293.0694 | CTR: 0.2348 | ProGRP*: 215.4109

Train Sens: 0.833 | Train Spec: 0.832 [TRAIN SCORE: 1.665] || Val Sens: 0.930 | Val Spec: 0.785 [VAL SCORE: 1.715] || Test Sens: 0.779 | Test Spec: 0.785 [TEST SCORE: 1.564]

**#64: Age + BMI + Nodule Diameter + CTR + CEA + NSE**

Optimized Thresholds: Age: 77.1054 | BMI: 39.1947 | Nodule Diameter: 23.2849 | CTR: 0.2368 | CEA: 276.4467 | NSE: 60.0323

Train Sens: 0.825 | Train Spec: 0.829 [TRAIN SCORE: 1.653] || Val Sens: 0.930 | Val Spec: 0.785 [VAL SCORE: 1.715] || Test Sens: 0.779 | Test Spec: 0.796 [TEST SCORE: 1.575]

**#65: Age + BMI + Nodule Diameter + CTR + CYFRA21-1***

Optimized Thresholds: Age: 77.2176 | BMI: 38.7003 | Nodule Diameter: 23.4771 | CTR: 0.2352 | CYFRA21-1*: 50.9371

Train Sens: 0.825 | Train Spec: 0.829 [TRAIN SCORE: 1.653] || Val Sens: 0.930 | Val Spec: 0.785 [VAL SCORE: 1.715] || Test Sens: 0.779 | Test Spec: 0.796 [TEST SCORE: 1.575]

**#66: BMI + Nodule Diameter + CTR + CYFRA21-1* + ProGRP + NSE**

Optimized Thresholds: BMI: 38.4370 | Nodule Diameter: 23.6254 | CTR: 0.2343 | CYFRA21-1*: 55.6573 | ProGRP: 254.3901 | NSE: 59.7666

Train Sens: 0.825 | Train Spec: 0.825 [TRAIN SCORE: 1.650] || Val Sens: 0.930 | Val Spec: 0.785 [VAL SCORE: 1.715] || Test Sens: 0.779 | Test Spec: 0.796 [TEST SCORE: 1.575]

**#67: Age + BMI + Nodule Diameter + CTR + CEA***

Optimized Thresholds: Age: 77.2176 | BMI: 38.7003 | Nodule Diameter: 23.4771 | CTR: 0.2352 | CEA*: 218.1183

Train Sens: 0.825 | Train Spec: 0.832 [TRAIN SCORE: 1.657] || Val Sens: 0.930 | Val Spec: 0.785 [VAL SCORE: 1.715] || Test Sens: 0.779 | Test Spec: 0.796 [TEST SCORE: 1.575]

##### #68: BMI + Nodule Diameter + CT.value + CTR + NSE

Optimized Thresholds: BMI: 38.8017 | Nodule Diameter: 25.0177 | CT.value: -295.9235 | CTR: 0.2326 | NSE: 60.2730

Train Sens: 0.829 | Train Spec: 0.839 [TRAIN SCORE: 1.668] || Val Sens: 0.919 | Val Spec: 0.796 [VAL SCORE: 1.714] || Test Sens: 0.779 | Test Spec: 0.796 [TEST SCORE: 1.575]

##### #69: Age + Nodule Diameter + CT.value + CTR + CEA + ProGRP + NSE

Optimized Thresholds: Age: 78.7141 | Nodule Diameter: 25.5355 | CT.value: -286.7534 | CTR: 0.2383 | CEA: 297.5691 | ProGRP: 290.3748 | NSE: 60.5205

Train Sens: 0.829 | Train Spec: 0.839 [TRAIN SCORE: 1.668] || Val Sens: 0.919 | Val Spec: 0.796 [VAL SCORE: 1.714] || Test Sens: 0.779 | Test Spec: 0.796 [TEST SCORE: 1.575]

##### #70: Age + Nodule Diameter + CT.value + CTR + CYFRA21-1 + ProGRP + NSE

Optimized Thresholds: Age: 78.7141 | Nodule Diameter: 25.5355 | CT.value: -286.7534 | CTR: 0.2383 | CYFRA21-1: 69.3387 | ProGRP: 290.3748 | NSE: 60.5205

Train Sens: 0.829 | Train Spec: 0.836 [TRAIN SCORE: 1.664] || Val Sens: 0.919 | Val Spec: 0.796 [VAL SCORE: 1.714] || Test Sens: 0.779 | Test Spec: 0.796 [TEST SCORE: 1.575]

##### #71: Age + Nodule Diameter + CT.value + CTR + ProGRP + NSE

Optimized Thresholds: Age: 77.0153 | Nodule Diameter: 25.2189 | CT.value: -291.9868 | CTR: 0.2364 | ProGRP: 278.0449 | NSE: 59.7736

Train Sens: 0.829 | Train Spec: 0.839 [TRAIN SCORE: 1.668] || Val Sens: 0.919 | Val Spec: 0.796 [VAL SCORE: 1.714] || Test Sens: 0.779 | Test Spec: 0.796 [TEST SCORE: 1.575]

##### #72: Age + Nodule Diameter + CT.value + CTR + CEA + CYFRA21-1 + NSE

Optimized Thresholds: Age: 78.7141 | Nodule Diameter: 25.5355 | CT.value: -286.7534 | CTR: 0.2383 | CEA: 297.5691 | CYFRA21-1: 66.1922 | NSE: 60.5205

Train Sens: 0.829 | Train Spec: 0.836 [TRAIN SCORE: 1.664] || Val Sens: 0.919 | Val Spec: 0.796 [VAL SCORE: 1.714] || Test Sens: 0.779 | Test Spec: 0.796 [TEST SCORE: 1.575]

##### #73: Age + Nodule Diameter + CT.value + CTR + CEA* + CYFRA21-1* + ProGRP*

Optimized Thresholds: Age: 77.6226 | Nodule Diameter: 25.4566 | CT.value: -290.8026 | CTR: 0.2343 | CEA*: 230.9050 | CYFRA21-1*: 55.5732 | ProGRP*: 277.0486

Train Sens: 0.829 | Train Spec: 0.839 [TRAIN SCORE: 1.668] || Val Sens: 0.919 | Val Spec: 0.796 [VAL SCORE: 1.714] || Test Sens: 0.779 | Test Spec: 0.796 [TEST SCORE: 1.575]

##### #74: Age + BMI + Nodule Diameter + CT.value + CTR + CEA + ProGRP + NSE

Optimized Thresholds: Age: 75.5512 | BMI: 37.0254 | Nodule Diameter: 25.4884 | CT.value: -283.1387 | CTR: 0.2338 | CEA: 211.5828 | ProGRP: 238.1582 | NSE: 59.2064

Train Sens: 0.829 | Train Spec: 0.836 [TRAIN SCORE: 1.664] || Val Sens: 0.919 | Val Spec: 0.796 [VAL SCORE: 1.714] || Test Sens: 0.779 | Test Spec: 0.796 [TEST SCORE: 1.575]

##### #75: Nodule Diameter + CT.value + CTR + NSE

Optimized Thresholds: Nodule Diameter: 25.4963 | CT.value: -307.3048 | CTR: 0.2336 | NSE: 59.7563

Train Sens: 0.829 | Train Spec: 0.839 [TRAIN SCORE: 1.668] || Val Sens: 0.919 | Val Spec: 0.796 [VAL SCORE: 1.714] || Test Sens: 0.779 | Test Spec: 0.796 [TEST SCORE: 1.575]

##### #76: BMI + Nodule Diameter + CT.value + CTR + CEA + CYFRA21-1* + ProGRP* + NSE

Optimized Thresholds: BMI: 39.3142 | Nodule Diameter: 25.5530 | CT.value: -305.2379 | CTR: 0.2346 | CEA: 68.6950 | CYFRA21-1*: 51.5090 | ProGRP*: 236.4806 | NSE: 59.3923

Train Sens: 0.829 | Train Spec: 0.836 [TRAIN SCORE: 1.664] || Val Sens: 0.919 | Val Spec: 0.796 [VAL SCORE: 1.714] || Test Sens: 0.779 | Test Spec: 0.796 [TEST SCORE: 1.575]

##### #77: Age + BMI + Nodule Diameter + CT.value + CTR + CYFRA21-1* + ProGRP*

Optimized Thresholds: Age: 79.2745 | BMI: 38.6732 | Nodule Diameter: 25.5173 | CT.value: -298.5627 | CTR: 0.2330 | CYFRA21-1*: 57.1232 | ProGRP*: 224.4671

Train Sens: 0.829 | Train Spec: 0.839 [TRAIN SCORE: 1.668] || Val Sens: 0.919 | Val Spec: 0.796 [VAL SCORE: 1.714] || Test Sens: 0.779 | Test Spec: 0.796 [TEST SCORE: 1.575]

##### #78: BMI + Nodule Diameter + CT.value + CTR + ProGRP + NSE

Optimized Thresholds: BMI: 38.6091 | Nodule Diameter: 25.2189 | CT.value: -291.9868 | CTR: 0.2364 | ProGRP: 278.0449 | NSE: 59.7736

Train Sens: 0.829 | Train Spec: 0.839 [TRAIN SCORE: 1.668] || Val Sens: 0.919 | Val Spec: 0.796 [VAL SCORE: 1.714] || Test Sens: 0.779 | Test Spec: 0.796 [TEST SCORE: 1.575]

##### #79: Age + Nodule Diameter + CT.value + CTR + CEA + CYFRA21-1* + ProGRP* + NSE

Optimized Thresholds: Age: 78.5284 | Nodule Diameter: 25.5530 | CT.value: -305.2379 | CTR: 0.2346 | CEA: 68.6950 | CYFRA21-1*: 51.5090 | ProGRP*: 236.4806 | NSE: 59.3923

Train Sens: 0.829 | Train Spec: 0.836 [TRAIN SCORE: 1.664] || Val Sens: 0.919 | Val Spec: 0.796 [VAL SCORE: 1.714] || Test Sens: 0.779 | Test Spec: 0.796 [TEST SCORE: 1.575]

##### #80: Age + BMI + Nodule Diameter + CT.value + CTR + NSE

Optimized Thresholds: Age: 77.2585 | BMI: 35.1169 | Nodule Diameter: 25.0734 | CT.value: -321.1286 | CTR: 0.2365 | NSE: 60.9835

Train Sens: 0.829 | Train Spec: 0.832 [TRAIN SCORE: 1.661] || Val Sens: 0.919 | Val Spec: 0.796 [VAL SCORE: 1.714] || Test Sens: 0.779 | Test Spec: 0.796 [TEST SCORE: 1.575]

##### #81: Age + BMI + Nodule Diameter + CT.value + CTR + ProGRP

Optimized Thresholds: Age: 79.9619 | BMI: 39.0520 | Nodule Diameter: 25.5325 | CT.value: -305.4893 | CTR: 0.2313 | ProGRP: 165.6779

Train Sens: 0.829 | Train Spec: 0.843 [TRAIN SCORE: 1.671] || Val Sens: 0.919 | Val Spec: 0.796 [VAL SCORE: 1.714] || Test Sens: 0.779 | Test Spec: 0.796 [TEST SCORE: 1.575]

##### #82: Age + BMI + Nodule Diameter + CT.value + CTR + CEA* + CYFRA21-1* + ProGRP* + NSE

Optimized Thresholds: Age: 76.4780 | BMI: 37.7224 | Nodule Diameter: 25.7204 | CT.value: -283.0163 | CTR: 0.2304 | CEA*: 292.0468 | CYFRA21-1*: 70.9110 | ProGRP*: 299.9279 | NSE: 57.8111

Train Sens: 0.829 | Train Spec: 0.832 [TRAIN SCORE: 1.661] || Val Sens: 0.919 | Val Spec: 0.796 [VAL SCORE: 1.714] || Test Sens: 0.779 | Test Spec: 0.796 [TEST SCORE: 1.575]

##### #83: Age + BMI + Nodule Diameter + CT.value + CTR + CYFRA21-1

Optimized Thresholds: Age: 79.9619 | BMI: 39.0520 | Nodule Diameter: 25.5325 | CT.value: -305.4893 | CTR: 0.2313 | CYFRA21-1:

36.8500

Train Sens: 0.829 | Train Spec: 0.839 [TRAIN SCORE: 1.668] || Val Sens: 0.919 | Val Spec: 0.796 [VAL SCORE: 1.714] || Test Sens: 0.779 | Test Spec: 0.796 [TEST SCORE: 1.575]

###### #84: Nodule Diameter + CT.value + CTR + CYFRA21-1* + ProGRP*

Optimized Thresholds: Nodule Diameter: 25.5908 | CT.value: -300.3797 | CTR: 0.2360 | CYFRA21-1*: 42.4465 | ProGRP*: 228.3979

Train Sens: 0.829 | Train Spec: 0.839 [TRAIN SCORE: 1.668] || Val Sens: 0.919 | Val Spec: 0.796 [VAL SCORE: 1.714] || Test Sens: 0.779 | Test Spec: 0.796 [TEST SCORE: 1.575]

###### #85: Nodule Diameter + CT.value + CTR + CYFRA21-1*

Optimized Thresholds: Nodule Diameter: 25.4721 | CT.value: -302.1247 | CTR: 0.2372 | CYFRA21-1*: 48.7755

Train Sens: 0.829 | Train Spec: 0.839 [TRAIN SCORE: 1.668] || Val Sens: 0.919 | Val Spec: 0.796 [VAL SCORE: 1.714] || Test Sens: 0.779 | Test Spec: 0.796 [TEST SCORE: 1.575]

###### #86: Age + BMI + Nodule Diameter + CT.value + CTR + CYFRA21-1 + NSE

Optimized Thresholds: Age: 77.8688 | BMI: 38.5332 | Nodule Diameter: 25.7151 | CT.value: -273.0064 | CTR: 0.2335 | CYFRA21-1: 64.2459 | NSE: 60.5622

Train Sens: 0.829 | Train Spec: 0.836 [TRAIN SCORE: 1.664] || Val Sens: 0.919 | Val Spec: 0.796 [VAL SCORE: 1.714] || Test Sens: 0.779 | Test Spec: 0.796 [TEST SCORE: 1.575]

###### #87: Nodule Diameter + CT.value + CTR + CEA* + NSE

Optimized Thresholds: Nodule Diameter: 25.5537 | CT.value: -293.6683 | CTR: 0.2358 | CEA*: 189.7443 | NSE: 59.8533

Train Sens: 0.829 | Train Spec: 0.839 [TRAIN SCORE: 1.668] || Val Sens: 0.919 | Val Spec: 0.796 [VAL SCORE: 1.714] || Test Sens: 0.779 | Test Spec: 0.796 [TEST SCORE: 1.575]

###### #88: BMI + Nodule Diameter + CT.value + CTR + CEA* + CYFRA21-1*

Optimized Thresholds: BMI: 39.1614 | Nodule Diameter: 25.5828 | CT.value: -289.6099 | CTR: 0.2361 | CEA*: 208.7118 | CYFRA21-1*: 51.4357

Train Sens: 0.829 | Train Spec: 0.839 [TRAIN SCORE: 1.668] || Val Sens: 0.919 | Val Spec: 0.796 [VAL SCORE: 1.714] || Test Sens: 0.779 | Test Spec: 0.796 [TEST SCORE: 1.575]

###### #89: Age + Nodule Diameter + CT.value + CTR + CYFRA21-1* + ProGRP*

Optimized Thresholds: Age: 77.0260 | Nodule Diameter: 25.6267 | CT.value: -299.2310 | CTR: 0.2365 | CYFRA21-1*: 57.5908 | ProGRP*: 262.6122

Train Sens: 0.829 | Train Spec: 0.839 [TRAIN SCORE: 1.668] || Val Sens: 0.919 | Val Spec: 0.796 [VAL SCORE: 1.714] || Test Sens: 0.779 | Test Spec: 0.796 [TEST SCORE: 1.575]

###### #90: Age + BMI + Nodule Diameter + CT.value + CTR + ProGRP + NSE

Optimized Thresholds: Age: 77.8688 | BMI: 38.5332 | Nodule Diameter: 25.7151 | CT.value: -273.0064 | CTR: 0.2335 | ProGRP: 282.1035 | NSE: 60.5622

Train Sens: 0.829 | Train Spec: 0.839 [TRAIN SCORE: 1.668] || Val Sens: 0.919 | Val Spec: 0.796 [VAL SCORE: 1.714] || Test Sens: 0.779 | Test Spec: 0.796 [TEST SCORE: 1.575]

###### #91: BMI + Nodule Diameter + CT.value + CTR + CYFRA21-1 + NSE

Optimized Thresholds: BMI: 38.6091 | Nodule Diameter: 25.2189 | CT.value: -291.9868 | CTR: 0.2364 | CYFRA21-1: 63.2909 | NSE: 59.7736

Train Sens: 0.829 | Train Spec: 0.836 [TRAIN SCORE: 1.664] || Val Sens: 0.919 | Val Spec: 0.796 [VAL SCORE: 1.714] || Test Sens: 0.779 | Test Spec: 0.796 [TEST SCORE: 1.575]

###### #92: Age + Nodule Diameter + CT.value + CTR + CYFRA21-1*

Optimized Thresholds: Age: 77.2846 | Nodule Diameter: 25.3454 | CT.value: -290.5546 | CTR: 0.2350 | CYFRA21-1*: 47.6589

Train Sens: 0.829 | Train Spec: 0.839 [TRAIN SCORE: 1.668] || Val Sens: 0.919 | Val Spec: 0.796 [VAL SCORE: 1.714] || Test Sens: 0.779 | Test Spec: 0.796 [TEST SCORE: 1.575]

###### #93: Nodule Diameter + CT.value + CTR + CEA* + CYFRA21-1* + ProGRP* + NSE

Optimized Thresholds: Nodule Diameter: 25.5323 | CT.value: -297.0636 | CTR: 0.2343 | CEA*: 248.7150 | CYFRA21-1*: 62.8519 | ProGRP*: 287.5762 | NSE: 59.6865

Train Sens: 0.829 | Train Spec: 0.836 [TRAIN SCORE: 1.664] || Val Sens: 0.919 | Val Spec: 0.796 [VAL SCORE: 1.714] || Test Sens: 0.779 | Test Spec: 0.796 [TEST SCORE: 1.575]

###### #94: Nodule Diameter + CT.value + CTR + ProGRP* + NSE

Optimized Thresholds: Nodule Diameter: 25.5523 | CT.value: -291.6728 | CTR: 0.2364 | ProGRP*: 222.5539 | NSE: 59.8138

Train Sens: 0.829 | Train Spec: 0.839 [TRAIN SCORE: 1.668] || Val Sens: 0.919 | Val Spec: 0.796 [VAL SCORE: 1.714] || Test Sens: 0.779 | Test Spec: 0.796 [TEST SCORE: 1.575]

###### #95: Nodule Diameter + CT.value + CTR + CEA* + CYFRA21-1* + ProGRP*

Optimized Thresholds: Nodule Diameter: 25.3334 | CT.value: -301.6804 | CTR: 0.2358 | CEA*: 234.2124 | CYFRA21-1*: 27.4472 | ProGRP*: 229.1701

Train Sens: 0.829 | Train Spec: 0.839 [TRAIN SCORE: 1.668] || Val Sens: 0.919 | Val Spec: 0.796 [VAL SCORE: 1.714] || Test Sens: 0.779 | Test Spec: 0.796 [TEST SCORE: 1.575]

###### #96: BMI + Nodule Diameter + CT.value + CTR + CEA + NSE

Optimized Thresholds: BMI: 38.6091 | Nodule Diameter: 25.2189 | CT.value: -291.9868 | CTR: 0.2364 | CEA: 271.4568 | NSE: 59.7736

Train Sens: 0.829 | Train Spec: 0.839 [TRAIN SCORE: 1.668] || Val Sens: 0.919 | Val Spec: 0.796 [VAL SCORE: 1.714] || Test Sens: 0.779 | Test Spec: 0.796 [TEST SCORE: 1.575]

###### #97: Nodule Diameter + CT.value + CTR + CYFRA21-1* + NSE

Optimized Thresholds: Nodule Diameter: 25.5537 | CT.value: -293.6683 | CTR: 0.2358 | CYFRA21-1*: 44.3655 | NSE: 59.8533

Train Sens: 0.829 | Train Spec: 0.836 [TRAIN SCORE: 1.664] || Val Sens: 0.919 | Val Spec: 0.796 [VAL SCORE: 1.714] || Test Sens: 0.779 | Test Spec: 0.796 [TEST SCORE: 1.575]

###### #98: Age + Nodule Diameter + CT.value + CTR + CYFRA21-1 + NSE

Optimized Thresholds: Age: 77.0153 | Nodule Diameter: 25.2189 | CT.value: -291.9868 | CTR: 0.2364 | CYFRA21-1: 63.2909 | NSE: 59.7736

Train Sens: 0.829 | Train Spec: 0.836 [TRAIN SCORE: 1.664] || Val Sens: 0.919 | Val Spec: 0.796 [VAL SCORE: 1.714] || Test Sens: 0.779 | Test Spec: 0.796 [TEST SCORE: 1.575]

###### #99: Nodule Diameter + CT.value + CTR + CEA* + CYFRA21-1* + NSE

Optimized Thresholds: Nodule Diameter: 25.3383 | CT.value: -293.2433 | CTR: 0.2340 | CEA*: 230.3402 | CYFRA21-1*: 51.4209 | NSE: 60.3292

Train Sens: 0.829 | Train Spec: 0.836 [TRAIN SCORE: 1.664] || Val Sens: 0.919 | Val Spec: 0.796 [VAL SCORE: 1.714] || Test Sens: 0.779 | Test Spec: 0.796 [TEST SCORE: 1.575]

**#100: Age + Nodule Diameter + CT.value + CTR + CEA* + CYFRA21-1***

Optimized Thresholds: Age: 77.6478 | Nodule Diameter: 25.5383 | CT.value: -293.1888 | CTR: 0.2357 | CEA*: 191.3590 | CYFRA21-1*: 53.9375

Train Sens: 0.829 | Train Spec: 0.839 [TRAIN SCORE: 1.668] || Val Sens: 0.919 | Val Spec: 0.796 [VAL SCORE: 1.714] || Test Sens: 0.779 | Test Spec: 0.796 [TEST SCORE: 1.575]

**#101: Age + BMI + Nodule Diameter + CT.value + CTR + CEA* + CYFRA21-1***

Optimized Thresholds: Age: 79.2745 | BMI: 38.6732 | Nodule Diameter: 25.5173 | CT.value: -298.5627 | CTR: 0.2330 | CEA*: 244.8271 | CYFRA21-1*: 50.6835

Train Sens: 0.829 | Train Spec: 0.839 [TRAIN SCORE: 1.668] || Val Sens: 0.919 | Val Spec: 0.796 [VAL SCORE: 1.714] || Test Sens: 0.779 | Test Spec: 0.796 [TEST SCORE: 1.575]

**#102: BMI + Nodule Diameter + CT.value + CTR + CYFRA21-1 + ProGRP + NSE**

Optimized Thresholds: BMI: 39.4008 | Nodule Diameter: 25.5355 | CT.value: -286.7534 | CTR: 0.2383 | CYFRA21-1: 69.3387 | ProGRP: 290.3748 | NSE: 60.5205

Train Sens: 0.829 | Train Spec: 0.836 [TRAIN SCORE: 1.664] || Val Sens: 0.919 | Val Spec: 0.796 [VAL SCORE: 1.714] || Test Sens: 0.779 | Test Spec: 0.796 [TEST SCORE: 1.575]

**#103: Nodule Diameter + CT.value + CTR + CEA* + CYFRA21-1***

Optimized Thresholds: Nodule Diameter: 25.4625 | CT.value: -291.3719 | CTR: 0.2334 | CEA*: 195.2023 | CYFRA21-1*: 50.6590

Train Sens: 0.829 | Train Spec: 0.839 [TRAIN SCORE: 1.668] || Val Sens: 0.919 | Val Spec: 0.796 [VAL SCORE: 1.714] || Test Sens: 0.779 | Test Spec: 0.796 [TEST SCORE: 1.575]

**#104: Nodule Diameter + CT.value + CTR + CYFRA21-1* + ProGRP* + NSE**

Optimized Thresholds: Nodule Diameter: 25.2942 | CT.value: -298.3655 | CTR: 0.2350 | CYFRA21-1*: 50.6742 | ProGRP*: 293.7717 | NSE: 60.6512

Train Sens: 0.829 | Train Spec: 0.836 [TRAIN SCORE: 1.664] || Val Sens: 0.919 | Val Spec: 0.796 [VAL SCORE: 1.714] || Test Sens: 0.779 | Test Spec: 0.796 [TEST SCORE: 1.575]

**#105: BMI + Nodule Diameter + CT.value + CTR + CEA + ProGRP + NSE**

Optimized Thresholds: BMI: 39.4008 | Nodule Diameter: 25.5355 | CT.value: -286.7534 | CTR: 0.2383 | CEA: 297.5691 | ProGRP: 290.3748 | NSE: 60.5205

Train Sens: 0.829 | Train Spec: 0.839 [TRAIN SCORE: 1.668] || Val Sens: 0.919 | Val Spec: 0.796 [VAL SCORE: 1.714] || Test Sens: 0.779 | Test Spec: 0.796 [TEST SCORE: 1.575]

**#106: BMI + Nodule Diameter + CT.value + CTR + CEA + CYFRA21-1 + NSE**

Optimized Thresholds: BMI: 39.4008 | Nodule Diameter: 25.5355 | CT.value: -286.7534 | CTR: 0.2383 | CEA: 297.5691 | CYFRA21-1: 66.1922 | NSE: 60.5205

Train Sens: 0.829 | Train Spec: 0.836 [TRAIN SCORE: 1.664] || Val Sens: 0.919 | Val Spec: 0.796 [VAL SCORE: 1.714] || Test Sens: 0.779 | Test Spec: 0.796 [TEST SCORE: 1.575]

**#107: Age + BMI + Nodule Diameter + CT.value + CTR + CYFRA21-1 + ProGRP + NSE**

Optimized Thresholds: Age: 75.5512 | BMI: 37.0254 | Nodule Diameter: 25.4884 | CT.value: -283.1387 | CTR: 0.2338 | CYFRA21-1:

49.4235 | ProGRP: 238.1582 | NSE: 59.2064

Train Sens: 0.829 | Train Spec: 0.832 [TRAIN SCORE: 1.661] || Val Sens: 0.919 | Val Spec: 0.796 [VAL SCORE: 1.714] || Test Sens: 0.779 | Test Spec: 0.796 [TEST SCORE: 1.575]

**#108: BMI + Nodule Diameter + CT.value + CTR + CEA* + CYFRA21-1* + ProGRP***

Optimized Thresholds: BMI: 38.8921 | Nodule Diameter: 25.4566 | CT.value: -290.8026 | CTR: 0.2343 | CEA*: 230.9050 | CYFRA21-1*: 55.5732 | ProGRP*: 277.0486

Train Sens: 0.829 | Train Spec: 0.839 [TRAIN SCORE: 1.668] || Val Sens: 0.919 | Val Spec: 0.796 [VAL SCORE: 1.714] || Test Sens: 0.779 | Test Spec: 0.796 [TEST SCORE: 1.575]

**#109: BMI + Nodule Diameter + CT.value + CTR + CYFRA21-1***

Optimized Thresholds: BMI: 38.7346 | Nodule Diameter: 25.3454 | CT.value: -290.5546 | CTR: 0.2350 | CYFRA21-1*: 47.6589

Train Sens: 0.829 | Train Spec: 0.839 [TRAIN SCORE: 1.668] || Val Sens: 0.919 | Val Spec: 0.796 [VAL SCORE: 1.714] || Test Sens: 0.779 | Test Spec: 0.796 [TEST SCORE: 1.575]

**#110: Age + BMI + Nodule Diameter + CT.value + CTR + CEA + CYFRA21-1 + ProGRP**

Optimized Thresholds: Age: 79.9930 | BMI: 38.4342 | Nodule Diameter: 25.3042 | CT.value: -317.1923 | CTR: 0.2344 | CEA: 301.6058 | CYFRA21-1: 30.3114 | ProGRP: 145.4684

Train Sens: 0.829 | Train Spec: 0.839 [TRAIN SCORE: 1.668] || Val Sens: 0.919 | Val Spec: 0.796 [VAL SCORE: 1.714] || Test Sens: 0.779 | Test Spec: 0.796 [TEST SCORE: 1.575]

**#111: Age + Nodule Diameter + CT.value + CTR + CEA + NSE**

Optimized Thresholds: Age: 77.0153 | Nodule Diameter: 25.2189 | CT.value: -291.9868 | CTR: 0.2364 | CEA: 271.4568 | NSE: 59.7736

Train Sens: 0.829 | Train Spec: 0.839 [TRAIN SCORE: 1.668] || Val Sens: 0.919 | Val Spec: 0.796 [VAL SCORE: 1.714] || Test Sens: 0.779 | Test Spec: 0.796 [TEST SCORE: 1.575]

**#112: Age + BMI + Nodule Diameter + CT.value + CTR + CEA + NSE**

Optimized Thresholds: Age: 77.8688 | BMI: 38.5332 | Nodule Diameter: 25.7151 | CT.value: -273.0064 | CTR: 0.2335 | CEA: 275.5803 | NSE: 60.5622

Train Sens: 0.829 | Train Spec: 0.839 [TRAIN SCORE: 1.668] || Val Sens: 0.919 | Val Spec: 0.796 [VAL SCORE: 1.714] || Test Sens: 0.779 | Test Spec: 0.796 [TEST SCORE: 1.575]

**#113: Age + BMI + Nodule Diameter + CT.value + CTR + CEA + CYFRA21-1 + NSE**

Optimized Thresholds: Age: 75.5512 | BMI: 37.0254 | Nodule Diameter: 25.4884 | CT.value: -283.1387 | CTR: 0.2338 | CEA: 211.5828 | CYFRA21-1: 53.9052 | NSE: 59.2064

Train Sens: 0.829 | Train Spec: 0.832 [TRAIN SCORE: 1.661] || Val Sens: 0.919 | Val Spec: 0.796 [VAL SCORE: 1.714] || Test Sens: 0.779 | Test Spec: 0.796 [TEST SCORE: 1.575]

**#114: Nodule Diameter + CT.value + CTR + CEA* + ProGRP* + NSE**

Optimized Thresholds: Nodule Diameter: 25.2942 | CT.value: -298.3655 | CTR: 0.2350 | CEA*: 216.9829 | ProGRP*: 293.7717 | NSE: 60.6512

Train Sens: 0.829 | Train Spec: 0.839 [TRAIN SCORE: 1.668] || Val Sens: 0.919 | Val Spec: 0.796 [VAL SCORE: 1.714] || Test Sens: 0.779 | Test Spec: 0.796 [TEST SCORE: 1.575]

**#115: Age + Nodule Diameter + CT.value + CTR + NSE**

Optimized Thresholds: Age: 77.4285 | Nodule Diameter: 25.0177 | CT.value: -295.9235 | CTR: 0.2326 | NSE: 60.2730

Train Sens: 0.829 | Train Spec: 0.839 [TRAIN SCORE: 1.668] || Val Sens: 0.919 | Val Spec: 0.796 [VAL SCORE: 1.714] || Test Sens: 0.779 | Test Spec: 0.796 [TEST SCORE: 1.575]

##### #116: BMI + Nodule Diameter + CT.value + CTR + CYFRA21-1* + ProGRP*

Optimized Thresholds: BMI: 38.8794 | Nodule Diameter: 25.3799 | CT.value: -293.6420 | CTR: 0.2358 | CYFRA21-1*: 56.5411 | ProGRP*: 241.5717

Train Sens: 0.829 | Train Spec: 0.839 [TRAIN SCORE: 1.668] || Val Sens: 0.919 | Val Spec: 0.796 [VAL SCORE: 1.714] || Test Sens: 0.779 | Test Spec: 0.796 [TEST SCORE: 1.575]

##### #117: BMI + CT.value + CEA* + ProGRP* + NSE

Optimized Thresholds: BMI: 38.7160 | CT.value: -436.3479 | CEA*: 215.5425 | ProGRP*: 237.0194 | NSE: 59.9618

Train Sens: 0.801 | Train Spec: 0.808 [TRAIN SCORE: 1.608] || Val Sens: 0.907 | Val Spec: 0.806 [VAL SCORE: 1.713] || Test Sens: 0.779 | Test Spec: 0.796 [TEST SCORE: 1.575]

##### #118: CT.value + ProGRP* + NSE

Optimized Thresholds: CT.value: -436.3499 | ProGRP*: 253.3647 | NSE: 59.2903

Train Sens: 0.801 | Train Spec: 0.808 [TRAIN SCORE: 1.608] || Val Sens: 0.907 | Val Spec: 0.806 [VAL SCORE: 1.713] || Test Sens: 0.779 | Test Spec: 0.796 [TEST SCORE: 1.575]

##### #119: BMI + CT.value + CEA* + CYFRA21-1* + NSE

Optimized Thresholds: BMI: 38.5330 | CT.value: -437.5104 | CEA*: 220.9080 | CYFRA21-1*: 47.4229 | NSE: 59.9344

Train Sens: 0.801 | Train Spec: 0.804 [TRAIN SCORE: 1.605] || Val Sens: 0.907 | Val Spec: 0.806 [VAL SCORE: 1.713] || Test Sens: 0.779 | Test Spec: 0.796 [TEST SCORE: 1.575]

##### #120: CT.value + CEA* + CYFRA21-1*

Optimized Thresholds: CT.value: -437.8454 | CEA*: 206.9475 | CYFRA21-1*: 37.9656

Train Sens: 0.801 | Train Spec: 0.808 [TRAIN SCORE: 1.608] || Val Sens: 0.907 | Val Spec: 0.806 [VAL SCORE: 1.713] || Test Sens: 0.779 | Test Spec: 0.796 [TEST SCORE: 1.575]

##### #121: BMI + CT.value + ProGRP* + NSE

Optimized Thresholds: BMI: 38.7094 | CT.value: -436.3632 | ProGRP*: 264.3785 | NSE: 59.8420

Train Sens: 0.801 | Train Spec: 0.808 [TRAIN SCORE: 1.608] || Val Sens: 0.907 | Val Spec: 0.806 [VAL SCORE: 1.713] || Test Sens: 0.779 | Test Spec: 0.796 [TEST SCORE: 1.575]

##### #122: BMI + CT.value + CYFRA21-1* + ProGRP* + NSE

Optimized Thresholds: BMI: 38.7629 | CT.value: -437.9845 | CYFRA21-1*: 30.9855 | ProGRP*: 313.9697 | NSE: 59.7224

Train Sens: 0.801 | Train Spec: 0.804 [TRAIN SCORE: 1.605] || Val Sens: 0.907 | Val Spec: 0.806 [VAL SCORE: 1.713] || Test Sens: 0.791 | Test Spec: 0.796 [TEST SCORE: 1.586]

##### #123: BMI + CT.value + CEA* + CYFRA21-1* + ProGRP* + NSE

Optimized Thresholds: BMI: 38.6863 | CT.value: -437.4870 | CEA*: 207.7215 | CYFRA21-1*: 58.3495 | ProGRP*: 290.8929 | NSE: 59.9748

Train Sens: 0.801 | Train Spec: 0.804 [TRAIN SCORE: 1.605] || Val Sens: 0.907 | Val Spec: 0.806 [VAL SCORE: 1.713] || Test Sens: 0.779 | Test Spec: 0.796 [TEST SCORE: 1.575]

###### #124: BMI + CT.value + CEA* + NSE

Optimized Thresholds: BMI: 38.5504 | CT.value: -436.5730 | CEA*: 230.1992 | NSE: 59.9155

Train Sens: 0.801 | Train Spec: 0.808 [TRAIN SCORE: 1.608] || Val Sens: 0.907 | Val Spec: 0.806 [VAL SCORE: 1.713] || Test Sens: 0.779 | Test Spec: 0.796 [TEST SCORE: 1.575]

###### #125: BMI + CT.value + CEA* + ProGRP*

Optimized Thresholds: BMI: 38.7330 | CT.value: -436.5259 | CEA*: 203.4530 | ProGRP*: 249.3524

Train Sens: 0.801 | Train Spec: 0.811 [TRAIN SCORE: 1.612] || Val Sens: 0.907 | Val Spec: 0.806 [VAL SCORE: 1.713] || Test Sens: 0.779 | Test Spec: 0.796 [TEST SCORE: 1.575]

###### #126: CT.value + CEA* + ProGRP*

Optimized Thresholds: CT.value: -436.3851 | CEA*: 258.7761 | ProGRP*: 207.8105

Train Sens: 0.801 | Train Spec: 0.811 [TRAIN SCORE: 1.612] || Val Sens: 0.907 | Val Spec: 0.806 [VAL SCORE: 1.713] || Test Sens: 0.779 | Test Spec: 0.796 [TEST SCORE: 1.575]

###### #127: CT.value + CEA* + NSE

Optimized Thresholds: CT.value: -436.5642 | CEA*: 137.9174 | NSE: 59.5402

Train Sens: 0.801 | Train Spec: 0.808 [TRAIN SCORE: 1.608] || Val Sens: 0.907 | Val Spec: 0.806 [VAL SCORE: 1.713] || Test Sens: 0.779 | Test Spec: 0.796 [TEST SCORE: 1.575]

###### #128: CT.value + CYFRA21-1* + ProGRP*

Optimized Thresholds: CT.value: -437.7815 | CYFRA21-1*: 63.6701 | ProGRP*: 302.1635

Train Sens: 0.801 | Train Spec: 0.808 [TRAIN SCORE: 1.608] || Val Sens: 0.907 | Val Spec: 0.806 [VAL SCORE: 1.713] || Test Sens: 0.779 | Test Spec: 0.796 [TEST SCORE: 1.575]

###### #129: CT.value + CYFRA21-1* + NSE

Optimized Thresholds: CT.value: -437.7977 | CYFRA21-1*: 46.7621 | NSE: 59.3912

Train Sens: 0.801 | Train Spec: 0.804 [TRAIN SCORE: 1.605] || Val Sens: 0.907 | Val Spec: 0.806 [VAL SCORE: 1.713] || Test Sens: 0.779 | Test Spec: 0.796 [TEST SCORE: 1.575]

###### #130: BMI + CT.value + CEA* + CYFRA21-1* + ProGRP*

Optimized Thresholds: BMI: 38.8929 | CT.value: -437.9631 | CEA*: 239.0423 | CYFRA21-1*: 58.9403 | ProGRP*: 267.6127

Train Sens: 0.801 | Train Spec: 0.808 [TRAIN SCORE: 1.608] || Val Sens: 0.907 | Val Spec: 0.806 [VAL SCORE: 1.713] || Test Sens: 0.791 | Test Spec: 0.796 [TEST SCORE: 1.586]

###### #131: BMI + CT.value + CEA*

Optimized Thresholds: BMI: 38.7777 | CT.value: -436.4578 | CEA*: 230.8561

Train Sens: 0.801 | Train Spec: 0.811 [TRAIN SCORE: 1.612] || Val Sens: 0.907 | Val Spec: 0.806 [VAL SCORE: 1.713] || Test Sens: 0.779 | Test Spec: 0.796 [TEST SCORE: 1.575]

###### #132: CT.value + CEA* + CYFRA21-1* + ProGRP* + NSE

Optimized Thresholds: CT.value: -437.5864 | CEA*: 133.2510 | CYFRA21-1*: 42.2924 | ProGRP*: 264.6457 | NSE: 59.7786

Train Sens: 0.801 | Train Spec: 0.804 [TRAIN SCORE: 1.605] || Val Sens: 0.907 | Val Spec: 0.806 [VAL SCORE: 1.713] || Test Sens: 0.779 | Test Spec: 0.796 [TEST SCORE: 1.575]

##### #133: BMI + CT.value + CYFRA21-1*

Optimized Thresholds: BMI: 38.7575 | CT.value: -437.6153 | CYFRA21-1*: 57.2101

Train Sens: 0.801 | Train Spec: 0.808 [TRAIN SCORE: 1.608] || Val Sens: 0.907 | Val Spec: 0.806 [VAL SCORE: 1.713] || Test Sens: 0.779 | Test Spec: 0.796 [TEST SCORE: 1.575]

##### #134: CT.value + CYFRA21-1* + ProGRP* + NSE

Optimized Thresholds: CT.value: -438.1573 | CYFRA21-1*: 35.0891 | ProGRP*: 261.8695 | NSE: 59.8479

Train Sens: 0.801 | Train Spec: 0.804 [TRAIN SCORE: 1.605] || Val Sens: 0.907 | Val Spec: 0.806 [VAL SCORE: 1.713] || Test Sens: 0.791 | Test Spec: 0.796 [TEST SCORE: 1.586]

##### #135: CT.value + CEA* + ProGRP* + NSE

Optimized Thresholds: CT.value: -436.4563 | CEA*: 209.4362 | ProGRP*: 224.0379 | NSE: 59.8639

Train Sens: 0.801 | Train Spec: 0.808 [TRAIN SCORE: 1.608] || Val Sens: 0.907 | Val Spec: 0.806 [VAL SCORE: 1.713] || Test Sens: 0.779 | Test Spec: 0.796 [TEST SCORE: 1.575]

##### #136: CT.value + CEA* + CYFRA21-1* + NSE

Optimized Thresholds: CT.value: -436.8275 | CEA*: 208.2730 | CYFRA21-1*: 58.8241 | NSE: 59.5435

Train Sens: 0.801 | Train Spec: 0.804 [TRAIN SCORE: 1.605] || Val Sens: 0.907 | Val Spec: 0.806 [VAL SCORE: 1.713] || Test Sens: 0.779 | Test Spec: 0.796 [TEST SCORE: 1.575]

##### #137: CT.value + CEA* + CYFRA21-1* + ProGRP*

Optimized Thresholds: CT.value: -437.3049 | CEA*: 285.0557 | CYFRA21-1*: 44.4064 | ProGRP*: 223.4876

Train Sens: 0.801 | Train Spec: 0.808 [TRAIN SCORE: 1.608] || Val Sens: 0.907 | Val Spec: 0.806 [VAL SCORE: 1.713] || Test Sens: 0.779 | Test Spec: 0.796 [TEST SCORE: 1.575]

##### #138: CT.value + CEA*

Optimized Thresholds: CT.value: -436.2855 | CEA*: 182.9960

Train Sens: 0.801 | Train Spec: 0.811 [TRAIN SCORE: 1.612] || Val Sens: 0.907 | Val Spec: 0.806 [VAL SCORE: 1.713] || Test Sens: 0.779 | Test Spec: 0.796 [TEST SCORE: 1.575]

##### #139: CT.value + CYFRA21-1*

Optimized Thresholds: CT.value: -437.1999 | CYFRA21-1*: 51.8972

Train Sens: 0.801 | Train Spec: 0.808 [TRAIN SCORE: 1.608] || Val Sens: 0.907 | Val Spec: 0.806 [VAL SCORE: 1.713] || Test Sens: 0.779 | Test Spec: 0.796 [TEST SCORE: 1.575]

##### #140: CT.value + ProGRP*

Optimized Thresholds: CT.value: -436.5188 | ProGRP*: 186.6477

Train Sens: 0.801 | Train Spec: 0.811 [TRAIN SCORE: 1.612] || Val Sens: 0.907 | Val Spec: 0.806 [VAL SCORE: 1.713] || Test Sens: 0.779 | Test Spec: 0.796 [TEST SCORE: 1.575]

##### #141: BMI + CT.value + CYFRA21-1* + ProGRP*

Optimized Thresholds: BMI: 39.1139 | CT.value: -437.0438 | CYFRA21-1*: 54.3732 | ProGRP*: 284.6751

Train Sens: 0.801 | Train Spec: 0.808 [TRAIN SCORE: 1.608] || Val Sens: 0.907 | Val Spec: 0.806 [VAL SCORE: 1.713] || Test Sens: 0.779 | Test Spec: 0.796 [TEST SCORE: 1.575]

###### #142: BMI + CT.value + CYFRA21-1* + NSE

Optimized Thresholds: BMI: 39.0958 | CT.value: -437.4537 | CYFRA21-1*: 56.7476 | NSE: 59.9022

Train Sens: 0.801 | Train Spec: 0.804 [TRAIN SCORE: 1.605] || Val Sens: 0.907 | Val Spec: 0.806 [VAL SCORE: 1.713] || Test Sens: 0.779 | Test Spec: 0.796 [TEST SCORE: 1.575]

###### #143: BMI + CT.value + NSE

Optimized Thresholds: BMI: 38.8730 | CT.value: -436.2695 | NSE: 59.9292

Train Sens: 0.801 | Train Spec: 0.808 [TRAIN SCORE: 1.608] || Val Sens: 0.907 | Val Spec: 0.806 [VAL SCORE: 1.713] || Test Sens: 0.779 | Test Spec: 0.796 [TEST SCORE: 1.575]

###### #144: BMI + CT.value + ProGRP*

Optimized Thresholds: BMI: 38.5678 | CT.value: -436.5775 | ProGRP*: 246.3711

Train Sens: 0.801 | Train Spec: 0.811 [TRAIN SCORE: 1.612] || Val Sens: 0.907 | Val Spec: 0.806 [VAL SCORE: 1.713] || Test Sens: 0.779 | Test Spec: 0.796 [TEST SCORE: 1.575]

###### #145: BMI + CT.value + CEA* + CYFRA21-1*

Optimized Thresholds: BMI: 39.0583 | CT.value: -437.5526 | CEA*: 205.3425 | CYFRA21-1*: 53.5690

Train Sens: 0.801 | Train Spec: 0.808 [TRAIN SCORE: 1.608] || Val Sens: 0.907 | Val Spec: 0.806 [VAL SCORE: 1.713] || Test Sens: 0.779 | Test Spec: 0.796 [TEST SCORE: 1.575]

###### #146: Age + BMI + Nodule Diameter + CT.value + CYFRA21-1* + ProGRP* + NSE

Optimized Thresholds: Age: 74.8926 | BMI: 38.6822 | Nodule Diameter: 21.1290 | CT.value: -401.5252 | CYFRA21-1*: 46.4067 | ProGRP*: 254.6757 | NSE: 56.0752

Train Sens: 0.805 | Train Spec: 0.815 [TRAIN SCORE: 1.619] || Val Sens: 0.895 | Val Spec: 0.817 [VAL SCORE: 1.713] || Test Sens: 0.721 | Test Spec: 0.828 [TEST SCORE: 1.549]

###### #147: BMI + Nodule Diameter + CT.value + CEA + SCC* + ProGRP* + NSE

Optimized Thresholds: BMI: 39.0219 | Nodule Diameter: 21.4008 | CT.value: -406.6104 | CEA: 100.9097 | SCC*: 4.6073 | ProGRP*: 301.0910 | NSE: 55.5139

Train Sens: 0.805 | Train Spec: 0.811 [TRAIN SCORE: 1.616] || Val Sens: 0.895 | Val Spec: 0.817 [VAL SCORE: 1.713] || Test Sens: 0.733 | Test Spec: 0.817 [TEST SCORE: 1.550]

###### #148: Age + BMI + Nodule Diameter + CT.value + CEA* + CYFRA21-1* + SCC + NSE

Optimized Thresholds: Age: 75.8461 | BMI: 39.3048 | Nodule Diameter: 21.0193 | CT.value: -400.5013 | CEA*: 173.1869 | CYFRA21-1*: 51.4606 | SCC: 4.9890 | NSE: 52.0164

Train Sens: 0.797 | Train Spec: 0.815 [TRAIN SCORE: 1.611] || Val Sens: 0.895 | Val Spec: 0.817 [VAL SCORE: 1.713] || Test Sens: 0.721 | Test Spec: 0.839 [TEST SCORE: 1.560]

###### #149: Age + BMI + Nodule Diameter + CT.value + CEA* + NSE

Optimized Thresholds: Age: 76.2898 | BMI: 39.0457 | Nodule Diameter: 21.0071 | CT.value: -401.6333 | CEA*: 251.6149 | NSE: 54.6605

Train Sens: 0.801 | Train Spec: 0.815 [TRAIN SCORE: 1.615] || Val Sens: 0.884 | Val Spec: 0.828 [VAL SCORE: 1.712] || Test Sens: 0.721 | Test Spec: 0.828 [TEST SCORE: 1.549]

###### #150: BMI + Nodule Diameter + CT.value + CTR + SCC* + ProGRP*

Optimized Thresholds: BMI: 38.6657 | Nodule Diameter: 23.8592 | CT.value: -300.0397 | CTR: 0.2362 | SCC*: 4.1464 | ProGRP*: 302.3957

Train Sens: 0.833 | Train Spec: 0.832  [TRAIN SCORE: 1.665]  ||  Val Sens: 0.930 | Val Spec: 0.774  [VAL SCORE: 1.704]  ||  Test Sens: 0.779 | Test Spec: 0.785  [TEST SCORE: 1.564]

###### #151: Age + Nodule Diameter + CTR + CEA + CYFRA21-1 + ProGRP

Optimized Thresholds: Age: 77.5812 | Nodule Diameter: 25.3289 | CTR: 0.2361 | CEA: 7.0321 | CYFRA21-1: 54.8059 | ProGRP: 367.3880

Train Sens: 0.829 | Train Spec: 0.825  [TRAIN SCORE: 1.654]  ||  Val Sens: 0.930 | Val Spec: 0.774  [VAL SCORE: 1.704]  ||  Test Sens: 0.791 | Test Spec: 0.796  [TEST SCORE: 1.586]

###### #152: Age + BMI + Nodule Diameter + CTR + CYFRA21-1* + SCC

Optimized Thresholds: Age: 76.9092 | BMI: 39.2360 | Nodule Diameter: 23.4229 | CTR: 0.2341 | CYFRA21-1*: 54.9011 | SCC: 4.1598

Train Sens: 0.825 | Train Spec: 0.829  [TRAIN SCORE: 1.653]  ||  Val Sens: 0.930 | Val Spec: 0.774  [VAL SCORE: 1.704]  ||  Test Sens: 0.779 | Test Spec: 0.796  [TEST SCORE: 1.575]

###### #153: Age + Nodule Diameter + CTR + CEA* + SCC + NSE

Optimized Thresholds: Age: 77.1093 | Nodule Diameter: 23.6330 | CTR: 0.2350 | CEA*: 209.3945 | SCC: 4.2590 | NSE: 59.9786

Train Sens: 0.825 | Train Spec: 0.829  [TRAIN SCORE: 1.653]  ||  Val Sens: 0.930 | Val Spec: 0.774  [VAL SCORE: 1.704]  ||  Test Sens: 0.779 | Test Spec: 0.796  [TEST SCORE: 1.575]

###### #154: Age + Nodule Diameter + CT.value + CTR + SCC* + ProGRP*

Optimized Thresholds: Age: 76.9297 | Nodule Diameter: 23.9280 | CT.value: -299.4033 | CTR: 0.2363 | SCC*: 4.0849 | ProGRP*: 286.3938

Train Sens: 0.833 | Train Spec: 0.832  [TRAIN SCORE: 1.665]  ||  Val Sens: 0.930 | Val Spec: 0.774  [VAL SCORE: 1.704]  ||  Test Sens: 0.779 | Test Spec: 0.785  [TEST SCORE: 1.564]

###### #155: Age + Nodule Diameter + CTR + CYFRA21-1* + SCC + ProGRP*

Optimized Thresholds: Age: 77.1235 | Nodule Diameter: 23.4425 | CTR: 0.2337 | CYFRA21-1*: 66.2783 | SCC: 4.5678 | ProGRP*: 273.9138

Train Sens: 0.825 | Train Spec: 0.829  [TRAIN SCORE: 1.653]  ||  Val Sens: 0.930 | Val Spec: 0.774  [VAL SCORE: 1.704]  ||  Test Sens: 0.779 | Test Spec: 0.796  [TEST SCORE: 1.575]

###### #156: Age + BMI + Nodule Diameter + CTR + SCC* + ProGRP*

Optimized Thresholds: Age: 77.8692 | BMI: 38.8864 | Nodule Diameter: 23.6763 | CTR: 0.2357 | SCC*: 3.9378 | ProGRP*: 271.5612

Train Sens: 0.825 | Train Spec: 0.832  [TRAIN SCORE: 1.657]  ||  Val Sens: 0.930 | Val Spec: 0.774  [VAL SCORE: 1.704]  ||  Test Sens: 0.779 | Test Spec: 0.796  [TEST SCORE: 1.575]

###### #157: Age + BMI + CTR + ProGRP*

Optimized Thresholds: Age: 76.8201 | BMI: 38.9391 | CTR: 0.2161 | ProGRP*: 286.1727

Train Sens: 0.813 | Train Spec: 0.822  [TRAIN SCORE: 1.634]  ||  Val Sens: 0.930 | Val Spec: 0.774  [VAL SCORE: 1.704]  ||  Test Sens: 0.791 | Test Spec: 0.806  [TEST SCORE: 1.597]

###### #158: Age + Nodule Diameter + CTR + SCC + ProGRP* + NSE

Optimized Thresholds: Age: 76.9645 | Nodule Diameter: 23.5786 | CTR: 0.2337 | SCC: 3.9992 | ProGRP*: 279.3688 | NSE: 59.8848

Train Sens: 0.825 | Train Spec: 0.829 [TRAIN SCORE: 1.653] || Val Sens: 0.930 | Val Spec: 0.774 [VAL SCORE: 1.704] || Test Sens: 0.779 | Test Spec: 0.796 [TEST SCORE: 1.575]

###### #159: Age + BMI + Nodule Diameter + CTR + CEA* + SCC

Optimized Thresholds: Age: 76.9092 | BMI: 39.2360 | Nodule Diameter: 23.4229 | CTR: 0.2341 | CEA*: 235.2329 | SCC: 4.1598

Train Sens: 0.825 | Train Spec: 0.832 [TRAIN SCORE: 1.657] || Val Sens: 0.930 | Val Spec: 0.774 [VAL SCORE: 1.704] || Test Sens: 0.779 | Test Spec: 0.796 [TEST SCORE: 1.575]

###### #160: Age + BMI + CTR + CYFRA21-1*

Optimized Thresholds: Age: 76.4085 | BMI: 39.4721 | CTR: 0.2158 | CYFRA21-1*: 42.8819

Train Sens: 0.813 | Train Spec: 0.818 [TRAIN SCORE: 1.631] || Val Sens: 0.930 | Val Spec: 0.774 [VAL SCORE: 1.704] || Test Sens: 0.791 | Test Spec: 0.806 [TEST SCORE: 1.597]

###### #161: Age + BMI + Nodule Diameter + CT.value + CTR + CEA

Optimized Thresholds: Age: 78.3992 | BMI: 39.5804 | Nodule Diameter: 23.2052 | CT.value: -321.2375 | CTR: 0.2386 | CEA: 25.1596

Train Sens: 0.833 | Train Spec: 0.832 [TRAIN SCORE: 1.665] || Val Sens: 0.930 | Val Spec: 0.774 [VAL SCORE: 1.704] || Test Sens: 0.779 | Test Spec: 0.785 [TEST SCORE: 1.564]

###### #162: Age + Nodule Diameter + CTR + CYFRA21-1* + SCC + NSE

Optimized Thresholds: Age: 77.1093 | Nodule Diameter: 23.6330 | CTR: 0.2350 | CYFRA21-1*: 48.9166 | SCC: 4.2590 | NSE: 59.9786

Train Sens: 0.825 | Train Spec: 0.825 [TRAIN SCORE: 1.650] || Val Sens: 0.930 | Val Spec: 0.774 [VAL SCORE: 1.704] || Test Sens: 0.779 | Test Spec: 0.796 [TEST SCORE: 1.575]

###### #163: Age + CTR + CYFRA21-1* + ProGRP*

Optimized Thresholds: Age: 77.4291 | CTR: 0.2161 | CYFRA21-1*: 36.4802 | ProGRP*: 281.7077

Train Sens: 0.813 | Train Spec: 0.818 [TRAIN SCORE: 1.631] || Val Sens: 0.930 | Val Spec: 0.774 [VAL SCORE: 1.704] || Test Sens: 0.791 | Test Spec: 0.806 [TEST SCORE: 1.597]

###### #164: Age + BMI + Nodule Diameter + CTR + SCC + NSE

Optimized Thresholds: Age: 78.0235 | BMI: 39.0038 | Nodule Diameter: 23.1776 | CTR: 0.2357 | SCC: 4.4222 | NSE: 60.5939

Train Sens: 0.825 | Train Spec: 0.829 [TRAIN SCORE: 1.653] || Val Sens: 0.930 | Val Spec: 0.774 [VAL SCORE: 1.704] || Test Sens: 0.779 | Test Spec: 0.796 [TEST SCORE: 1.575]

###### #165: BMI + Nodule Diameter + CT.value + CTR + CEA* + SCC

Optimized Thresholds: BMI: 38.9810 | Nodule Diameter: 23.3804 | CT.value: -301.3300 | CTR: 0.2361 | CEA*: 288.6647 | SCC: 4.4709

Train Sens: 0.833 | Train Spec: 0.832 [TRAIN SCORE: 1.665] || Val Sens: 0.930 | Val Spec: 0.774 [VAL SCORE: 1.704] || Test Sens: 0.779 | Test Spec: 0.785 [TEST SCORE: 1.564]

###### #166: Age + BMI + CT.value + CTR + CEA* + NSE

Optimized Thresholds: Age: 77.7405 | BMI: 39.0500 | CT.value: -297.4592 | CTR: 0.2180 | CEA*: 182.8471 | NSE: 56.5303

Train Sens: 0.821 | Train Spec: 0.815 [TRAIN SCORE: 1.635] || Val Sens: 0.930 | Val Spec: 0.774 [VAL SCORE: 1.704] || Test Sens: 0.791 | Test Spec: 0.796 [TEST SCORE: 1.586]

###### #167: Age + BMI + CT.value + CTR + CYFRA21-1* + ProGRP*

Optimized Thresholds: Age: 77.0477 | BMI: 38.8329 | CT.value: -298.6583 | CTR: 0.2130 | CYFRA21-1*: 61.5288 | ProGRP*: 273.4820

Train Sens: 0.821 | Train Spec: 0.818 [TRAIN SCORE: 1.639] || Val Sens: 0.930 | Val Spec: 0.774 [VAL SCORE: 1.704] || Test Sens: 0.791 | Test Spec: 0.796 [TEST SCORE: 1.586]

###### #168: Age + BMI + CT.value + CTR + ProGRP* + NSE

Optimized Thresholds: Age: 77.4894 | BMI: 39.0671 | CT.value: -304.2488 | CTR: 0.2165 | ProGRP*: 289.5347 | NSE: 59.9276

Train Sens: 0.821 | Train Spec: 0.818 [TRAIN SCORE: 1.639] || Val Sens: 0.930 | Val Spec: 0.774 [VAL SCORE: 1.704] || Test Sens: 0.791 | Test Spec: 0.796 [TEST SCORE: 1.586]

###### #169: Nodule Diameter + CTR + SCC* + ProGRP* + NSE

Optimized Thresholds: Nodule Diameter: 23.8875 | CTR: 0.2335 | SCC*: 3.9926 | ProGRP*: 263.0152 | NSE: 58.9011

Train Sens: 0.825 | Train Spec: 0.829 [TRAIN SCORE: 1.653] || Val Sens: 0.930 | Val Spec: 0.774 [VAL SCORE: 1.704] || Test Sens: 0.779 | Test Spec: 0.796 [TEST SCORE: 1.575]

###### #170: Nodule Diameter + CTR + CEA + ProGRP + NSE

Optimized Thresholds: Nodule Diameter: 25.4083 | CTR: 0.2375 | CEA: 6.7069 | ProGRP: 345.9276 | NSE: 60.6902

Train Sens: 0.829 | Train Spec: 0.825 [TRAIN SCORE: 1.654] || Val Sens: 0.930 | Val Spec: 0.774 [VAL SCORE: 1.704] || Test Sens: 0.791 | Test Spec: 0.796 [TEST SCORE: 1.586]

###### #171: Nodule Diameter + CTR + CEA + CYFRA21-1* + ProGRP*

Optimized Thresholds: Nodule Diameter: 25.4001 | CTR: 0.2349 | CEA: 6.8670 | CYFRA21-1*: 52.3901 | ProGRP*: 290.1765

Train Sens: 0.829 | Train Spec: 0.825 [TRAIN SCORE: 1.654] || Val Sens: 0.930 | Val Spec: 0.774 [VAL SCORE: 1.704] || Test Sens: 0.791 | Test Spec: 0.796 [TEST SCORE: 1.586]

###### #172: Nodule Diameter + CT.value + CTR + SCC + ProGRP*

Optimized Thresholds: Nodule Diameter: 23.4878 | CT.value: -298.8685 | CTR: 0.2352 | SCC: 3.9991 | ProGRP*: 214.8786

Train Sens: 0.833 | Train Spec: 0.832 [TRAIN SCORE: 1.665] || Val Sens: 0.930 | Val Spec: 0.774 [VAL SCORE: 1.704] || Test Sens: 0.779 | Test Spec: 0.785 [TEST SCORE: 1.564]

###### #173: BMI + CT.value + CTR + CYFRA21-1*

Optimized Thresholds: BMI: 38.6612 | CT.value: -299.0683 | CTR: 0.2136 | CYFRA21-1*: 56.6156

Train Sens: 0.821 | Train Spec: 0.818 [TRAIN SCORE: 1.639] || Val Sens: 0.930 | Val Spec: 0.774 [VAL SCORE: 1.704] || Test Sens: 0.791 | Test Spec: 0.796 [TEST SCORE: 1.586]

###### #174: Age + CT.value + CTR + ProGRP*

Optimized Thresholds: Age: 76.6160 | CT.value: -304.7293 | CTR: 0.2138 | ProGRP*: 316.4209

Train Sens: 0.821 | Train Spec: 0.822 [TRAIN SCORE: 1.642] || Val Sens: 0.930 | Val Spec: 0.774 [VAL SCORE: 1.704] || Test Sens: 0.791 | Test Spec: 0.796 [TEST SCORE: 1.586]

###### #175: Age + CT.value + CTR + CYFRA21-1*

Optimized Thresholds: Age: 76.4844 | CT.value: -301.0099 | CTR: 0.2134 | CYFRA21-1*: 63.9337

Train Sens: 0.821 | Train Spec: 0.818 [TRAIN SCORE: 1.639] || Val Sens: 0.930 | Val Spec: 0.774 [VAL SCORE: 1.704] || Test Sens: 0.791 | Test Spec: 0.796 [TEST SCORE: 1.586]

##### #176: Age + Nodule Diameter + CT.value + CTR + CEA* + SCC

Optimized Thresholds: Age: 76.9261 | Nodule Diameter: 23.5813 | CT.value: -302.2441 | CTR: 0.2364 | CEA*: 281.0277 | SCC: 4.4834

Train Sens: 0.833 | Train Spec: 0.832 [TRAIN SCORE: 1.665] || Val Sens: 0.930 | Val Spec: 0.774 [VAL SCORE: 1.704] || Test Sens: 0.779 | Test Spec: 0.785 [TEST SCORE: 1.564]

##### #177: BMI + CT.value + CTR + ProGRP*

Optimized Thresholds: BMI: 38.9761 | CT.value: -301.3084 | CTR: 0.2150 | ProGRP*: 285.9718

Train Sens: 0.821 | Train Spec: 0.822 [TRAIN SCORE: 1.642] || Val Sens: 0.930 | Val Spec: 0.774 [VAL SCORE: 1.704] || Test Sens: 0.791 | Test Spec: 0.796 [TEST SCORE: 1.586]

##### #178: Nodule Diameter + CTR + CYFRA21-1* + SCC* + NSE

Optimized Thresholds: Nodule Diameter: 23.6840 | CTR: 0.2355 | CYFRA21-1*: 51.8441 | SCC*: 4.0685 | NSE: 59.7345

Train Sens: 0.825 | Train Spec: 0.825 [TRAIN SCORE: 1.650] || Val Sens: 0.930 | Val Spec: 0.774 [VAL SCORE: 1.704] || Test Sens: 0.779 | Test Spec: 0.796 [TEST SCORE: 1.575]

##### #179: BMI + Nodule Diameter + CTR + CEA

Optimized Thresholds: BMI: 37.0258 | Nodule Diameter: 25.6665 | CTR: 0.2309 | CEA: 6.9920

Train Sens: 0.829 | Train Spec: 0.825 [TRAIN SCORE: 1.654] || Val Sens: 0.930 | Val Spec: 0.774 [VAL SCORE: 1.704] || Test Sens: 0.791 | Test Spec: 0.796 [TEST SCORE: 1.586]

##### #180: BMI + Nodule Diameter + CTR + CEA + CYFRA21-1 + ProGRP

Optimized Thresholds: BMI: 38.8728 | Nodule Diameter: 25.3289 | CTR: 0.2361 | CEA: 7.0321 | CYFRA21-1: 54.8059 | ProGRP: 367.3880

Train Sens: 0.829 | Train Spec: 0.825 [TRAIN SCORE: 1.654] || Val Sens: 0.930 | Val Spec: 0.774 [VAL SCORE: 1.704] || Test Sens: 0.791 | Test Spec: 0.796 [TEST SCORE: 1.586]

##### #181: Age + BMI + CT.value + CTR

Optimized Thresholds: Age: 77.3784 | BMI: 38.9339 | CT.value: -295.1573 | CTR: 0.2154

Train Sens: 0.821 | Train Spec: 0.822 [TRAIN SCORE: 1.642] || Val Sens: 0.930 | Val Spec: 0.774 [VAL SCORE: 1.704] || Test Sens: 0.791 | Test Spec: 0.796 [TEST SCORE: 1.586]

##### #182: Age + BMI + Nodule Diameter + CTR + CEA* + CYFRA21-1* + SCC + ProGRP*

Optimized Thresholds: Age: 76.4834 | BMI: 38.9764 | Nodule Diameter: 23.6491 | CTR: 0.2344 | CEA*: 245.2701 | CYFRA21-1*: 49.8538 | SCC: 3.6288 | ProGRP*: 250.3308

Train Sens: 0.825 | Train Spec: 0.829 [TRAIN SCORE: 1.653] || Val Sens: 0.930 | Val Spec: 0.774 [VAL SCORE: 1.704] || Test Sens: 0.779 | Test Spec: 0.796 [TEST SCORE: 1.575]

##### #183: Age + Nodule Diameter + CT.value + CTR + SCC*

Optimized Thresholds: Age: 76.7997 | Nodule Diameter: 23.8823 | CT.value: -290.2897 | CTR: 0.2346 | SCC*: 3.7416

Train Sens: 0.833 | Train Spec: 0.832 [TRAIN SCORE: 1.665] || Val Sens: 0.930 | Val Spec: 0.774 [VAL SCORE: 1.704] || Test Sens: 0.779 | Test Spec: 0.785 [TEST SCORE: 1.564]

##### #184: Age + Nodule Diameter + CTR + CEA + CYFRA21-1

Optimized Thresholds: Age: 77.1416 | Nodule Diameter: 25.4435 | CTR: 0.2382 | CEA: 6.7265 | CYFRA21-1: 53.8939

Train Sens: 0.829 | Train Spec: 0.825 [TRAIN SCORE: 1.654] || Val Sens: 0.930 | Val Spec: 0.774 [VAL SCORE: 1.704] || Test Sens: 0.791 | Test Spec: 0.796 [TEST SCORE: 1.586]

##### #185: Age + Nodule Diameter + CTR + CEA + ProGRP

Optimized Thresholds: Age: 77.1416 | Nodule Diameter: 25.4435 | CTR: 0.2382 | CEA: 6.7265 | ProGRP: 238.1101

Train Sens: 0.829 | Train Spec: 0.829 [TRAIN SCORE: 1.657] || Val Sens: 0.930 | Val Spec: 0.774 [VAL SCORE: 1.704] || Test Sens: 0.791 | Test Spec: 0.796 [TEST SCORE: 1.586]

##### #186: Age + BMI + CTR

Optimized Thresholds: Age: 76.6826 | BMI: 39.2601 | CTR: 0.2132

Train Sens: 0.813 | Train Spec: 0.822 [TRAIN SCORE: 1.634] || Val Sens: 0.930 | Val Spec: 0.774 [VAL SCORE: 1.704] || Test Sens: 0.791 | Test Spec: 0.806 [TEST SCORE: 1.597]

##### #187: Nodule Diameter + CTR + CEA* + CYFRA21-1* + SCC + ProGRP + NSE

Optimized Thresholds: Nodule Diameter: 23.4460 | CTR: 0.2341 | CEA*: 211.4891 | CYFRA21-1*: 51.2637 | SCC: 3.9532 | ProGRP: 313.8180 | NSE: 59.7921

Train Sens: 0.825 | Train Spec: 0.825 [TRAIN SCORE: 1.650] || Val Sens: 0.930 | Val Spec: 0.774 [VAL SCORE: 1.704] || Test Sens: 0.779 | Test Spec: 0.796 [TEST SCORE: 1.575]

##### #188: Age + Nodule Diameter + CTR + SCC + NSE

Optimized Thresholds: Age: 78.2216 | Nodule Diameter: 23.5444 | CTR: 0.2353 | SCC: 4.7248 | NSE: 59.5015

Train Sens: 0.825 | Train Spec: 0.829 [TRAIN SCORE: 1.653] || Val Sens: 0.930 | Val Spec: 0.774 [VAL SCORE: 1.704] || Test Sens: 0.779 | Test Spec: 0.796 [TEST SCORE: 1.575]

##### #189: Age + CT.value + CTR + CYFRA21-1* + ProGRP*

Optimized Thresholds: Age: 76.8656 | CT.value: -293.6139 | CTR: 0.2157 | CYFRA21-1*: 45.2699 | ProGRP*: 279.5291

Train Sens: 0.821 | Train Spec: 0.818 [TRAIN SCORE: 1.639] || Val Sens: 0.930 | Val Spec: 0.774 [VAL SCORE: 1.704] || Test Sens: 0.791 | Test Spec: 0.796 [TEST SCORE: 1.586]

##### #190: BMI + Nodule Diameter + CTR + CYFRA21-1* + SCC + ProGRP* + NSE

Optimized Thresholds: BMI: 38.2128 | Nodule Diameter: 23.5670 | CTR: 0.2343 | CYFRA21-1*: 54.6227 | SCC: 4.4161 | ProGRP*: 276.9032 | NSE: 59.7633

Train Sens: 0.825 | Train Spec: 0.825 [TRAIN SCORE: 1.650] || Val Sens: 0.930 | Val Spec: 0.774 [VAL SCORE: 1.704] || Test Sens: 0.779 | Test Spec: 0.796 [TEST SCORE: 1.575]

##### #191: Age + CT.value + CTR

Optimized Thresholds: Age: 77.7264 | CT.value: -297.7504 | CTR: 0.2160

Train Sens: 0.821 | Train Spec: 0.822 [TRAIN SCORE: 1.642] || Val Sens: 0.930 | Val Spec: 0.774 [VAL SCORE: 1.704] || Test Sens: 0.791 | Test Spec: 0.796 [TEST SCORE: 1.586]

##### #192: Age + CT.value + CTR + ProGRP* + NSE*

Optimized Thresholds: Age: 76.4327 | CT.value: -291.0843 | CTR: 0.2180 | ProGRP*: 251.4646 | NSE*: 55.3011

Train Sens: 0.821 | Train Spec: 0.815 [TRAIN SCORE: 1.635] || Val Sens: 0.930 | Val Spec: 0.774 [VAL SCORE: 1.704] || Test Sens: 0.791 | Test Spec: 0.796 [TEST SCORE: 1.586]

###### #193: BMI + Nodule Diameter + CTR + CEA* + SCC + ProGRP* + NSE

Optimized Thresholds: BMI: 38.3094 | Nodule Diameter: 23.5909 | CTR: 0.2348 | CEA*: 210.6870 | SCC: 4.3902 | ProGRP*: 277.2977 | NSE: 59.8735

Train Sens: 0.825 | Train Spec: 0.829 [TRAIN SCORE: 1.653] || Val Sens: 0.930 | Val Spec: 0.774 [VAL SCORE: 1.704] || Test Sens: 0.779 | Test Spec: 0.796 [TEST SCORE: 1.575]

###### #194: Nodule Diameter + CTR + SCC* + NSE

Optimized Thresholds: Nodule Diameter: 23.7543 | CTR: 0.2350 | SCC*: 4.0917 | NSE: 58.8928

Train Sens: 0.825 | Train Spec: 0.829 [TRAIN SCORE: 1.653] || Val Sens: 0.930 | Val Spec: 0.774 [VAL SCORE: 1.704] || Test Sens: 0.779 | Test Spec: 0.796 [TEST SCORE: 1.575]

###### #195: Age + CTR + CYFRA21-1*

Optimized Thresholds: Age: 78.0266 | CTR: 0.2166 | CYFRA21-1*: 39.6294

Train Sens: 0.813 | Train Spec: 0.818 [TRAIN SCORE: 1.631] || Val Sens: 0.930 | Val Spec: 0.774 [VAL SCORE: 1.704] || Test Sens: 0.791 | Test Spec: 0.806 [TEST SCORE: 1.597]

###### #196: BMI + Nodule Diameter + CTR + CEA* + CYFRA21-1* + SCC + NSE

Optimized Thresholds: BMI: 38.6428 | Nodule Diameter: 23.3966 | CTR: 0.2358 | CEA*: 170.0762 | CYFRA21-1*: 54.7475 | SCC: 4.2470 | NSE: 59.9827

Train Sens: 0.825 | Train Spec: 0.825 [TRAIN SCORE: 1.650] || Val Sens: 0.930 | Val Spec: 0.774 [VAL SCORE: 1.704] || Test Sens: 0.779 | Test Spec: 0.796 [TEST SCORE: 1.575]

###### #197: Age + BMI + Nodule Diameter + CTR + CEA* + CYFRA21-1* + SCC* + NSE

Optimized Thresholds: Age: 76.2433 | BMI: 38.8855 | Nodule Diameter: 23.3041 | CTR: 0.2319 | CEA*: 205.8196 | CYFRA21-1*: 43.6402 | SCC*: 4.0967 | NSE: 56.4784

Train Sens: 0.825 | Train Spec: 0.822 [TRAIN SCORE: 1.646] || Val Sens: 0.930 | Val Spec: 0.774 [VAL SCORE: 1.704] || Test Sens: 0.779 | Test Spec: 0.796 [TEST SCORE: 1.575]

###### #198: CTR + ProGRP*

Optimized Thresholds: CTR: 0.2153 | ProGRP*: 246.9164

Train Sens: 0.813 | Train Spec: 0.822 [TRAIN SCORE: 1.634] || Val Sens: 0.930 | Val Spec: 0.774 [VAL SCORE: 1.704] || Test Sens: 0.791 | Test Spec: 0.806 [TEST SCORE: 1.597]

###### #199: Age + BMI + Nodule Diameter + CTR + CEA + SCC + ProGRP + NSE

Optimized Thresholds: Age: 76.2860 | BMI: 39.0764 | Nodule Diameter: 23.4906 | CTR: 0.2349 | CEA: 367.1128 | SCC: 3.5231 | ProGRP: 192.1620 | NSE: 59.6230

Train Sens: 0.825 | Train Spec: 0.829 [TRAIN SCORE: 1.653] || Val Sens: 0.930 | Val Spec: 0.774 [VAL SCORE: 1.704] || Test Sens: 0.779 | Test Spec: 0.796 [TEST SCORE: 1.575]

###### #200: BMI + Nodule Diameter + CTR + CEA* + CYFRA21-1 + SCC* + ProGRP* + NSE

Optimized Thresholds: BMI: 38.9965 | Nodule Diameter: 23.4810 | CTR: 0.2339 | CEA*: 183.8673 | CYFRA21-1: 68.1194 | SCC*: 4.3066 | ProGRP*: 294.8634 | NSE: 60.1735

Train Sens: 0.825 | Train Spec: 0.825 [TRAIN SCORE: 1.650] || Val Sens: 0.930 | Val Spec: 0.774 [VAL SCORE: 1.704] || Test Sens: 0.779 | Test Spec: 0.796 [TEST SCORE: 1.575]
